# Novel Driver Strength Index highlights important cancer genes in TCGA PanCanAtlas patients

**DOI:** 10.1101/2021.08.01.21261447

**Authors:** Aleksey V. Belikov, Danila V. Otnyukov, Alexey D. Vyatkin, Sergey V. Leonov

## Abstract

Elucidating crucial driver genes is paramount for understanding the cancer origins and mechanisms of progression, as well as selecting targets for molecular therapy. Cancer genes are usually ranked by the frequency of mutation, which, however, does not necessarily reflect their driver strength. Here we hypothesize that driver strength is higher for genes that are preferentially mutated in patients with few driver mutations overall, because these few mutations should be strong enough to initiate cancer. We propose a formula to calculate the corresponding Driver Strength Index (DSI), as well as the Normalized Driver Strength Index (NDSI), the latter completely independent of the overall gene mutation frequency. We validate these indices using the largest database of human cancer mutations – TCGA PanCanAtlas, multiple established algorithms for cancer driver prediction (2020plus, CHASMplus, CompositeDriver, dNdScv, HotMAPS, OncodriveCLUSTL, OncodriveFML) and four custom computational pipelines that integrate driver contributions from SNA, CNA and aneuploidy at the patient-level resolution. We demonstrate that DSI and especially NDSI provide substantially different rankings of genes as compared to frequency approach. For example, NDSI prioritized members of specific protein families, including G proteins *GNAQ, GNA11* and *GNAS*, isocitrate dehydrogenases *IDH1* and *IDH2*, and fibroblast growth factor receptors *FGFR2* and *FGFR3*. KEGG analysis shows that top NDSI-ranked genes comprise *EGFR/FGFR2/GNAQ/GNA11* – *NRAS/HRAS/KRAS* – *BRAF* pathway, *AKT1* – *MTOR* pathway, and *TCEB1* – *VHL* – *HIF1A* pathway. NDSI does not seem to correlate with the number of protein-protein interactions. We share our software to enable calculation of DSI and NDSI for outputs of any third-party driver prediction algorithms or their combinations.

## Introduction

Most cancer driver prediction algorithms answer one question – what is a probability of a given gene being a driver. This is definitely a crucial question and the answers are very valuable. However, high confidence that a gene is a driver does not translate to the statement that this gene is a strong driver. We can imagine a gene that is mutated in the majority of cancer patients (e.g. because it has multiple suitable sites for a driver mutation) but has a very weak contribution to cancer progression in each of these patients (e.g. because this gene is redundant). We can also imagine a gene that is mutated rarely (e.g. because it has only one suitable site for a driver mutation) but if the mutation does occur it immediately leads to cancer (e.g. because this gene is in a key position to control cell growth). The former would be an example of high confidence but weak driver, whereas the latter would be low confidence but strong driver. Overall, algorithms based on mutation recurrence cannot determine driver strength.

Some algorithms try to predict driver strength based on data from protein interaction networks (1)(2)(3). The idea is that a gene having multiple connections with other genes, i.e. playing the role of a network hub, will have more dramatic influence on the cell in case of mutation (4). This seems like a great idea at first sight, but a more detailed look shows that this is not the case. Yes, mutations in network hubs are likely to cause more disturbance in the cell, but what are the reasons to believe that all (or any of) these perturbations would be beneficial for cancer progression? In fact, mutations in network hubs are more likely to lead to cell death than to oncogenic transformation.

Here, we propose another approach. We reason that a few strong drivers are sufficient to initiate cancer, and there would be no need to accumulate additional drivers. On the other hand, weak drivers would need to accumulate in much higher quantity, until their combined strength would become sufficient to initiate cancer. Therefore, it should be statistically more likely to find strong drivers in patients that have only few driver mutations in their tumors, and less likely to find them in patients with multiple drivers per tumor. Likewise, it should be statistically less likely to find weak drivers in patients that have only few driver mutations in their tumors, and more likely to find them in patients with multiple drivers per tumor. Hence, we propose the Driver Strength Index (DSI) that takes into account the frequencies of mutation of a given driver gene in groups of patients with different total number of driver mutations, and gives priority weights to groups with fewer mutations. We also propose a modification of this index that is completely independent of the overall frequency of mutation of a given driver gene – the Normalized Driver Strength Index (NDSI).

Calculating these indices requires data on the number of driver mutations in each individual patient. The majority of existing driver prediction algorithms work at the cohort level, i.e. they predict driver genes for large groups of patients, usually having a particular cancer type. This does not allow to look at the composition of driver mutations in individual patients. We wrote specific scripts to convert cohort-level predictions into patient-level events, which also allowed seamless integration of the results from various third-party algorithms, including 2020plus (5), CHASMplus (6), CompositeDriver (7), dNdScv (8), DriverNet (9), HotMAPS (10), OncodriveCLUSTL (11), and OncodriveFML (12). This is useful, as each individual driver prediction algorithm has its own strengths and shortcomings, and combining results from multiple algorithms allows to obtain more complete and balanced picture, ensuring that less driver mutations have been missed.

In addition to these existing driver prediction algorithms, we decided to create our own, using clear and simple rules to have an internal reference standard. We called this algorithm SNADRIF – Single Nucleotide Alteration DRIver Finder. It predicts cancer driver genes from the TCGA PanCanAtlas SNA data and classifies them into oncogenes and tumor suppressors. Driver prediction is based on calculating the ratio of nonsynonymous SNAs to silent SNAs (8), whereas driver classification is based on calculating the ratio of hyperactivating SNAs to inactivating SNAs (13). Bootstrapping is used to calculate statistical significance and Benjamini–Hochberg procedure is used to keep false discovery rate under 5%.

Copy-number alterations (CNA) usually involve large chunks of DNA containing tens or hundreds of genes, which makes CNA data not very useful for uncovering individual driver genes. Nevertheless, it is an important source of information about amplifications and deletions of driver genes predicted from SNA data. However, due to CNA data coarseness, we wanted to clarify the actual copy number status of individual genes using mRNA and miRNA expression data available at TCGA PanCanAtlas. For this purpose, we created another pipeline called GECNAV - Gene Expression-based CNA Validator. CNA validation is based on comparing the CNA status of a given gene in a given patient to expression of this gene in this patient relative to the median expression of this gene across all patients.

Aneuploidy – chromosome arm and full chromosome gains and losses – makes a substantial contribution to the number of driver alterations per tumor, and thus we needed to take it into account when calculating our indices. However, there are no existing algorithms to differentiate driver aneuploidies from passenger ones. Therefore, we built our own pipeline called ANDRIF - ANeuploidy DRIver Finder. Driver prediction is based on calculating the average alteration status for each arm or chromosome in each cancer type. Bootstrapping is then used to obtain the realistic distribution of the average alteration statuses under the null hypothesis and Benjamini–Hochberg procedure is performed to keep the false discovery rate under 5%.

Finally, we needed an algorithm to integrate all data on driver mutations from different algorithms - our own and third-party. We called this algorithm PALDRIC - PAtient-Level DRIver Classifier. It translates cohort-level lists of driver genes or mutations to the patient level, classifies driver events according to the molecular causes and functional consequences, and presents comprehensive statistics on various kinds of driver events in various demographic and clinical groups of patients. Moreover, we developed a modification of PALDRIC that allows analysis and ranking of individual genes, chromosome arms and full chromosomes according to their frequency of occurrence, DSI, and NDSI.

Our overall workflow can be seen in **Fig 1**.

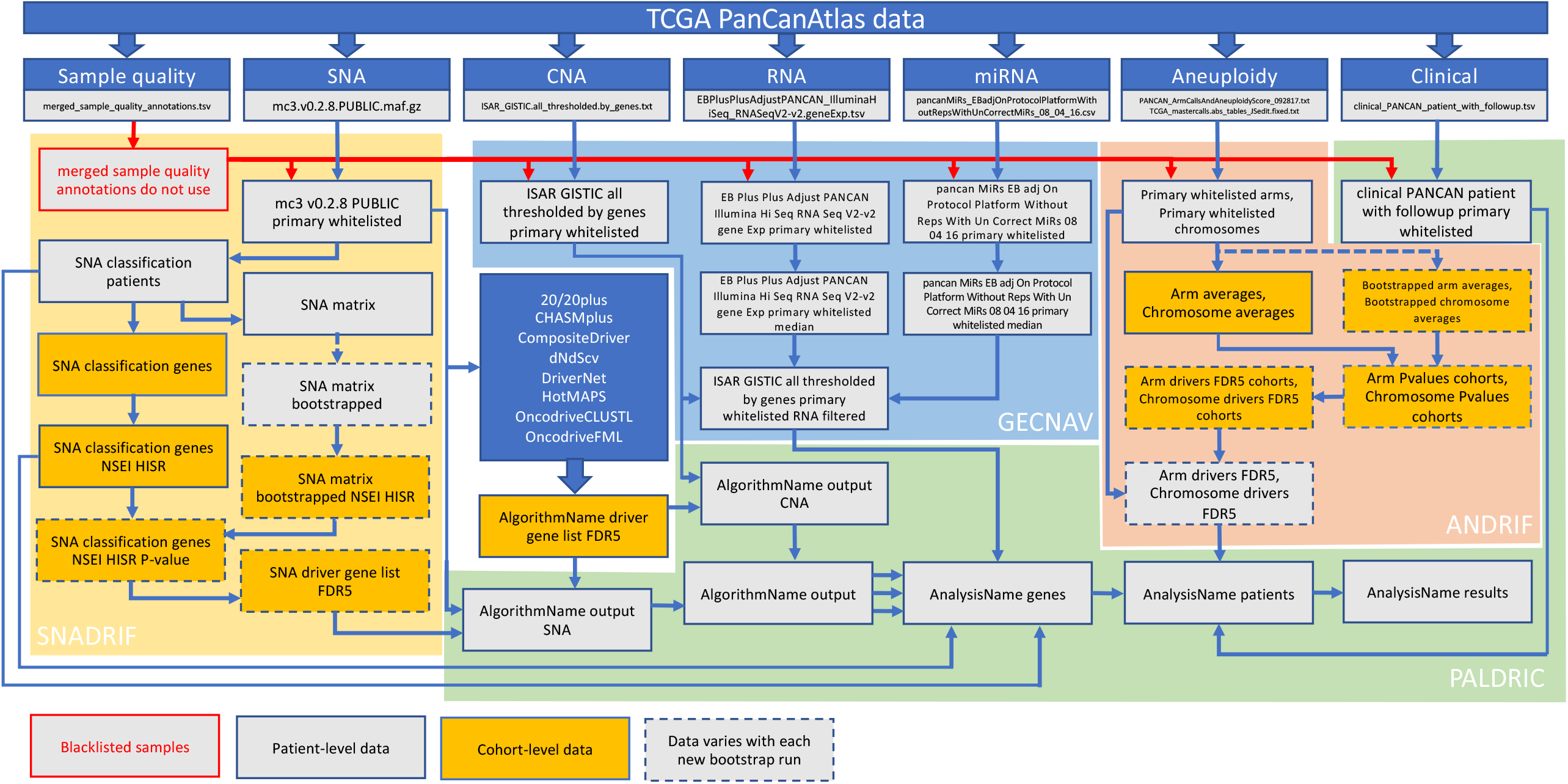

## Results

To get two different perspectives on the number and composition of driver events, we performed two different analyses. In the first one we used the combination of results from SNADRIF, several third-party algorithms - 2020plus, CHASMplus, CompositeDriver, HotMAPS, OncodriveFML, and a consensus driver gene list from 26 algorithms (7), each of them applied to the whole TCGA PanCanAtlas dataset. We also used a list of COSMIC Cancer Gene Census Tier 1 genes affected by somatic SNAs and CNAs, as it represents the current gold standard of verified cancer drivers (14). To minimize false positives, we used only genes that were predicted as drivers by at least two of our sources, including CGC and a list from (7). Unfortunately, application of driver gene lists discovered through pancan analysis equally to every cancer type results in unrealistically high numbers of driver events per patient, which is to be expected as this approach ignores tissue specificity of driver genes. Therefore, we present this analysis only as **Additional file 1**.

In the second analysis we used the combination of results from 2020plus, CHASMplus, CompositeDriver, dNdScv, DriverNet, HotMAPS, OncodriveCLUSTL, OncodriveFML, the consensus list from (7) and a list of Cancer Gene Census Tier 1 genes affected by somatic SNAs and CNAs, applied separately to each cancer cohort of TCGA PanCanAtlas. Applying algorithms to individual cohorts allows to discover cancer type-specific drivers and avoid contamination by false positives, i.e. driver genes discovered during Pancan analysis that do not in reality play any role in a given cancer type. On the other hand, much fewer patients are available for cohort-specific analysis, and this decreases statistical power to discover new driver genes. Of note, our SNADRIF algorithm works best for pancan analysis and struggles with small cohorts, due to scarcity of point mutations. However, when a combination of driver prediction algorithms is used, there are lower chances of missing an important driver gene even in a cohort-specific analysis, as algorithms based on differing principles complement each other. Similar to the first approach, to minimize false positives we used only driver gene-cohort pairs that were predicted by at least two of our sources, including CGC and a list from (7). The results of this analysis would be presented in the following paragraphs.

We calculated the number of various types of driver events in individual genes, chromosome arms or full chromosomes for each cancer type, tumor stage, age group, as well as for patients with each total number of driver events from 1 to 100. We performed the analyses for total population and for males and females separately, and, for each group, plotted the histograms of top 10 driver events in each class and overall (for data and graphs see **Additional file 2**). In **Fig 2** we present the overall ranking of genes for all TCGA PanCanAtlas cohorts combined. It can be seen that *PIK3CA* is the oncogene with the highest number of SNAs, as well as the highest number of simultaneous occurrences of SNAs and gene amplifications. *MYC* is the oncogene with the highest number of amplifications. *TP53* is the tumor suppressor with the highest number of SNAs, as well as the highest number of instances of simultaneous occurrences of an SNA in one allele and a deletion of the other allele. It is also the top mutated gene when driver events of all classes are counted. *CDKN2A* and *PTEN* are tumor suppressors with the highest number of deletions. Losses of chromosomes 13 and 22 are the most frequent cancer-promoting chromosome losses, whereas gains of chromosomes 7 and 20 are the most frequent cancer-promoting chromosome gains. Losses of 8p and 17p arms are the most frequent cancer-promoting chromosome arm losses, whereas gains of 1q and 8q arms are the most frequent cancer-promoting chromosome arm gains. Overall, these results are expected and indicate that our analytic pipelines work as they should.

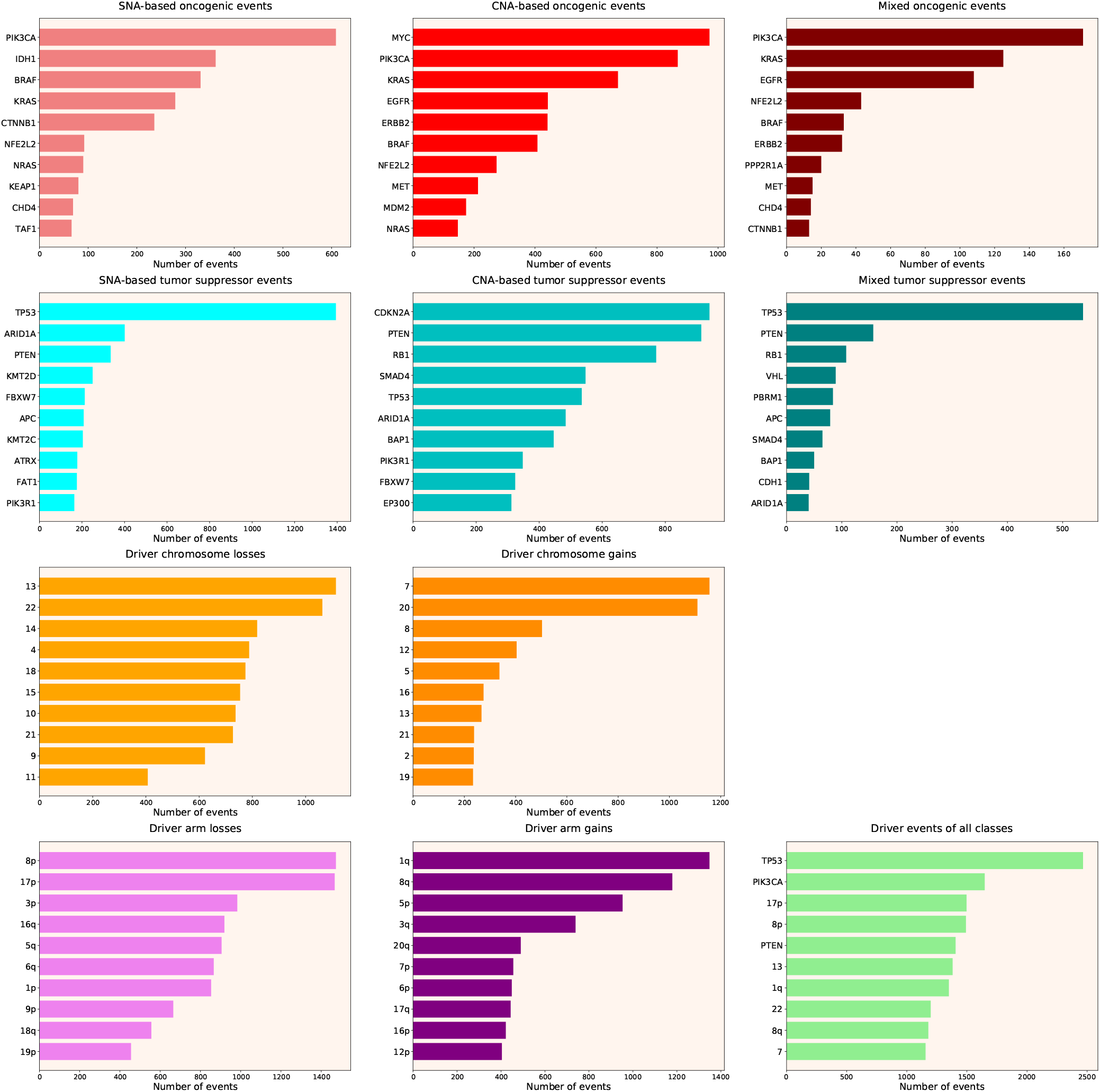

Next, we calculated the Driver Strength Index (DSI)

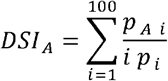

where *p*_*A i*_ is a number of patients with a driver event in the gene/chromosome *A* amongst patients with *i* driver events in total; *p*_*i*_ is a number of patients with i driver events in total. Surprisingly, we do not see much change compared to the simple frequency-of-mutation approach (**Fig 3**). The only dramatic difference is that *BRAF* became the top SNA-based oncogene according to DSI, whereas *PIK3CA* dropped to the second place, lagging behind by a wide margin. Also, *PIK3CA* overtook *MYC* as the top CNA-based oncogene, and *PTEN* displaced *CDKN2A* from the top CNA-based tumor suppressor spot. Moreover, members of several gene families appeared in the top 10 lists, such as *KRAS, NRAS* and *HRAS* in the SNA-based oncogenic events list, histones *HIST2H2BE* and *HIST1H3B* in the CNA-based oncogenic events list, or lysine methyltransferases *KMT2C* and *KMT2D* in the SNA-based tumor suppressor events list. This indicates that our approach is indeed meaningfully selecting for some biological attributes, which are not selected by simple frequency sorting. Finally, multiple small changes of ranking positions happened, nevertheless not affecting the overall picture. We think the reason for the limited effect of changes is that DSI is still very much affected by the overall frequency of gene mutation, due to 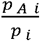 component. Hence, to uncover the true driver strength unrelated to the mutation frequency, further normalization is required.

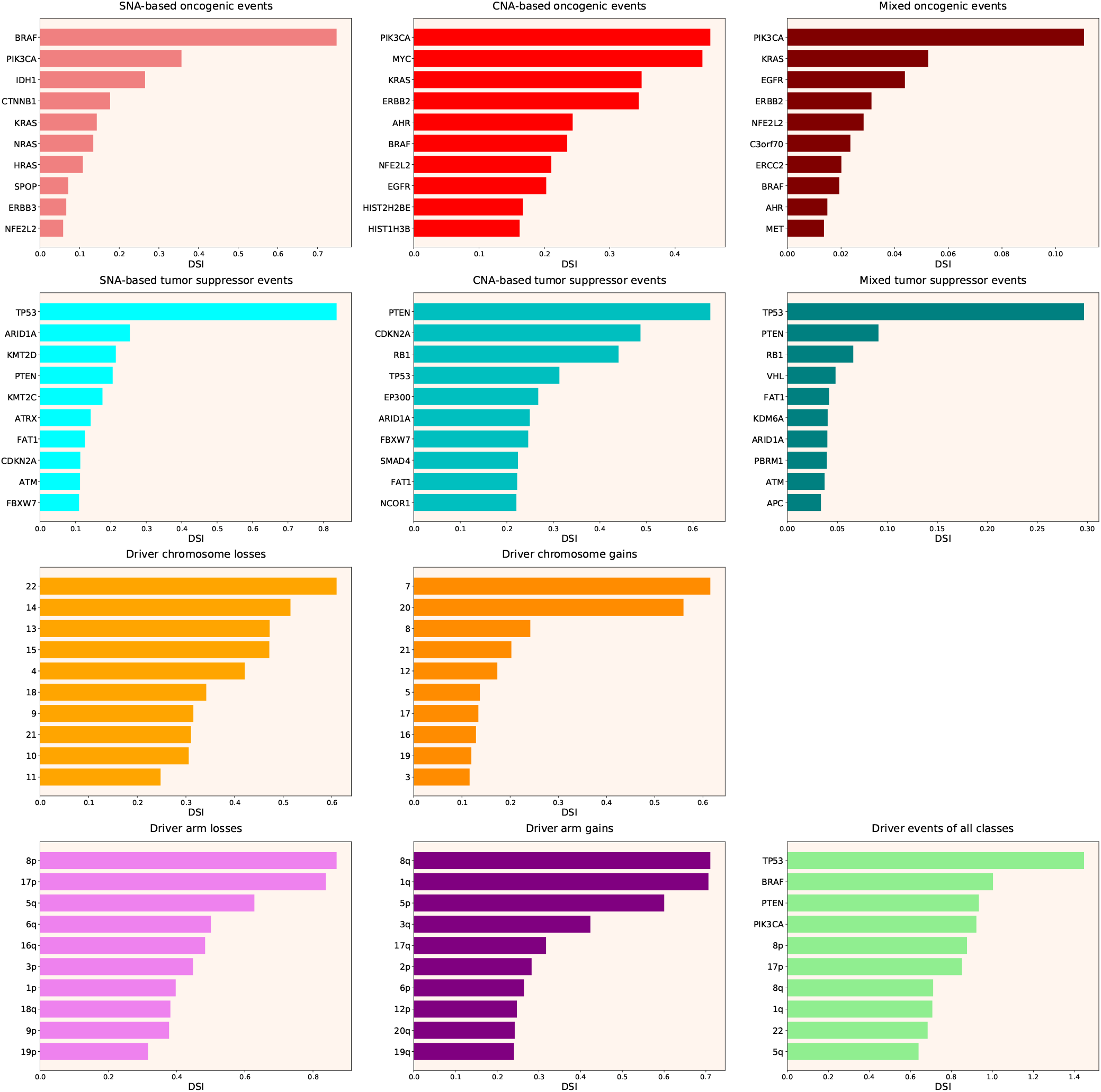

Therefore, we propose Normalized Driver Strength Index (NDSI)

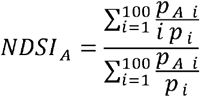

that corrects for the effects of mutation frequencies. As can be seen in **Fig 4**, this time the rankings are completely different from both DSI and frequency-based approaches. *GTF2I* conquers the top spot amongst SNA-based oncogenes and overall, *SPOP* becomes number one CNA-based oncogene, and *MET* occupies the first line of mixed oncogene rating. *ATRX, CSDE1* and *NF2* become the top SNA-based, CNA-based and mixed tumor suppressors, respectively. NDSI reveals the losses of chromosomes 12 and 3 as the strongest cancer-promoting chromosome losses, whereas the gain of chromosome 17 as the strongest cancer-promoting chromosome gain. NDSI shows that the losses of 19q and 12p arms are the strongest cancer-promoting chromosome arm losses, whereas the gain of 5q arm is the strongest cancer-promoting chromosome arm gain.

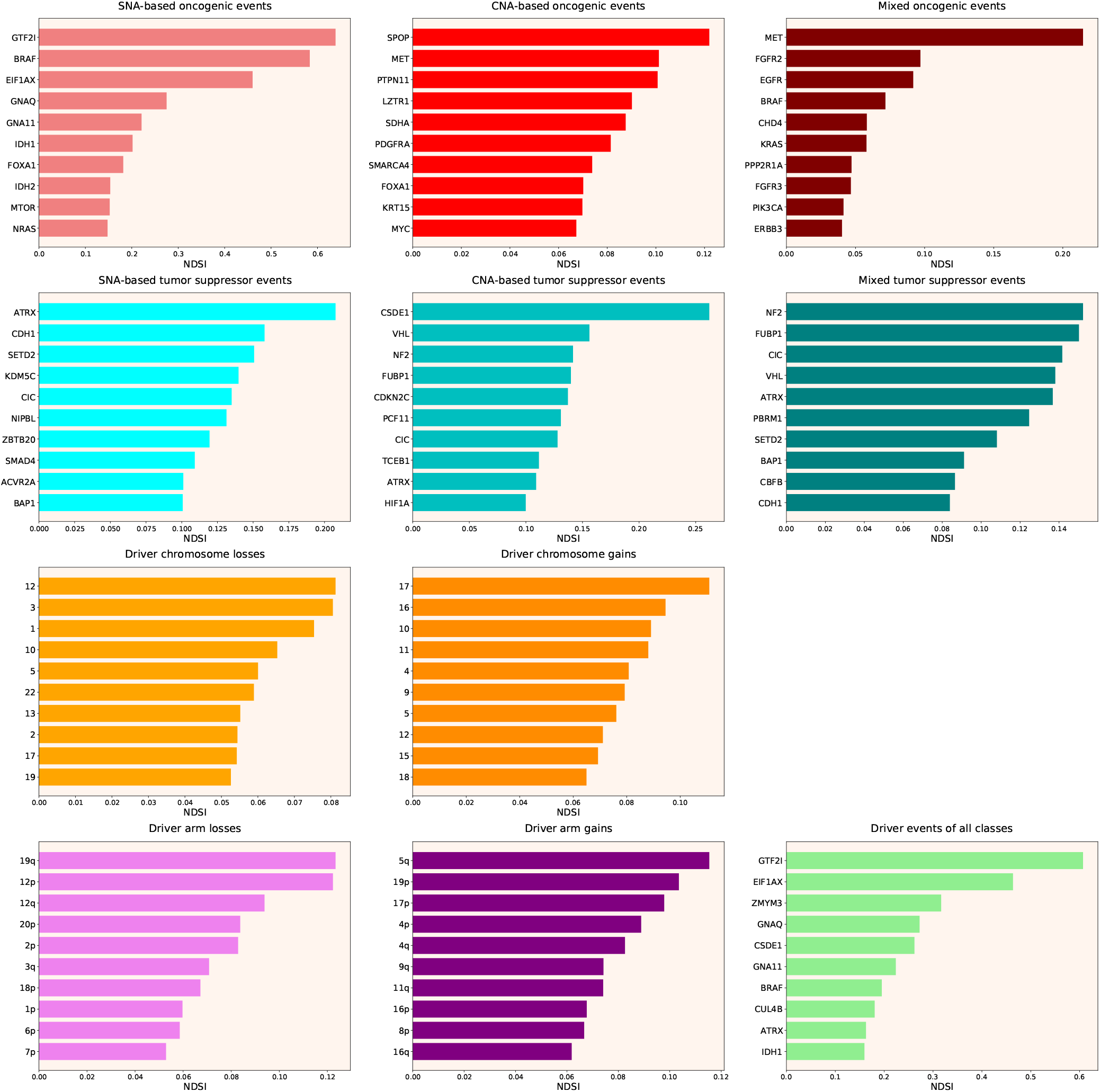

Like DSI, NDSI is able to select for specific gene families. Two members of the guanine nucleotide-binding protein family, *GNAQ* and *GNA11*, appeared on the top 10 SNA-based oncogenic events and top 10 driver events of all classes lists (**Fig 4**). Additionally, one more G protein, *GNAS*, appeared on the top 50 NDSI-ranked driver list (**Table 1**). Of note, no members of this family are present on the top 50 DSI-ranked driver list (**Table 1**). Two members of isocitrate dehydrogenase family, *IDH1* and *IDH2*, appeared on the top 10 SNA-based oncogenic events list, whereas fibroblast growth factor receptors *FGFR2* and *FGFR3* appeared on the top 10 mixed oncogenic events list (**Fig 4**). The ability of NDSI to prioritize members of specific protein families suggests that this index has actual biological meaning.

**Table 1.** Top 50 DSI- and NDSI-ranked genes.

| Rank | Entrez ID | Symbol | DSI | Rank | Entrez ID | Symbol | NDSI |
| --- | --- | --- | --- | --- | --- | --- | --- |
| 1 | 7157 | TP53 | 1.44664 | 1 | 100093631 | GTF2I | 0.6389 |
| 2 | 673 | BRAF | 1.00268 | 2 | 673 | BRAF | 0.58335 |
| 3 | 5728 | PTEN | 0.93317 | 3 | 1964 | EIF1AX | 0.46362 |
| 4 | 5290 | PIK3CA | 0.9215 | 4 | 9203 | ZMYM3 | 0.31661 |
| 5 | 1029 | CDKN2A | 0.60404 | 5 | 2776 | GNAQ | 0.27502 |
| 6 | 5925 | RB1 | 0.57932 | 6 | 7812 | CSDE1 | 0.26198 |
| 7 | 3845 | KRAS | 0.54447 | 7 | 2767 | GNA11 | 0.22382 |
| 8 | 8289 | ARID1A | 0.54286 | 8 | 8731 | MET | 0.21464 |
| 9 | 4609 | MYC | 0.46527 | 9 | 546 | ATRX | 0.20772 |
| 10 | 2064 | ERBB2 | 0.41563 | 10 | 3417 | IDH1 | 0.20172 |
| 11 | 2033 | EP300 | 0.39411 | 11 | 3169 | FOXA1 | 0.18161 |
| 12 | 2195 | FAT1 | 0.39042 | 12 | 8450 | CUL4B | 0.18057 |
| 13 | 55294 | FBXW7 | 0.37252 | 13 | 51343 | CDH1 | 0.158 |
| 14 | 472 | ATM | 0.35055 | 14 | 7428 | VHL | 0.15612 |
| 15 | 3417 | IDH1 | 0.29994 | 15 | 3418 | IDH2 | 0.15364 |
| 16 | 4780 | NFE2L2 | 0.2974 | 16 | 2475 | MTOR | 0.15247 |
| 17 | 1499 | CTNNB1 | 0.29 | 17 | 4771 | NF2 | 0.15232 |
| 18 | 4893 | NRAS | 0.27847 | 18 | 29072 | SETD2 | 0.1507 |
| 19 | 1387 | CREBBP | 0.2738 | 19 | 8880 | FUBP1 | 0.15022 |
| 20 | 5295 | PIK3R1 | 0.27316 | 20 | 4893 | NRAS | 0.14785 |
| 21 | 4089 | SMAD4 | 0.26931 | 21 | 207 | AKT1 | 0.1468 |
| 22 | 196 | AHR | 0.26508 | 22 | 4615 | MYD88 | 0.14625 |
| 23 | 9611 | NCOR1 | 0.25715 | 23 | 23152 | CIC | 0.14165 |
| 24 | 1956 | EGFR | 0.25138 | 24 | 51585 | PCF11 | 0.14083 |
| 25 | 7403 | KDM6A | 0.24971 | 25 | 8242 | KDM5C | 0.13977 |
| 26 | 9223 | BAP1 | 0.24578 | 26 | 1031 | CDKN2C | 0.13712 |
| 27 | 8085 | KMT2D | 0.24363 | 27 | 6597 | SMARCA4 | 0.13643 |
| 28 | 4297 | MLL | 0.23565 | 28 | 25836 | NIPBL | 0.13291 |
| 29 | 5624 | APC | 0.22422 | 29 | 7114 | TMSB4X | 0.12846 |
| 30 | 546 | ATRX | 0.21608 | 30 | 2778 | GNAS | 0.12708 |
| 31 | 2068 | ERCC2 | 0.21513 | 31 | 26137 | ZBTB20 | 0.1269 |
| 32 | 9757 | MLL2 | 0.20883 | 32 | 84433 | CARD11 | 0.12676 |
| 33 | 196528 | ARID2 | 0.20073 | 33 | 3265 | HRAS | 0.12612 |
| 34 | 58508 | MLL3 | 0.1921 | 34 | 55193 | PBRM1 | 0.12463 |
| 35 | 58508 | KMT2C | 0.18892 | 35 | 8405 | SPOP | 0.12197 |
| 36 | 3265 | HRAS | 0.17947 | 36 | 6921 | TCEB1 | 0.1115 |
| 37 | 1105 | CHD1 | 0.17591 | 37 | 4089 | SMAD4 | 0.10913 |
| 38 | 2065 | ERBB3 | 0.17366 | 38 | 5781 | PTPN11 | 0.10708 |
| 39 | 11168 | PSIP1 | 0.17344 | 39 | 92 | ACVR2A | 0.10109 |
| 40 | 8349 | HIST2H2BE | 0.16951 | 40 | 3091 | HIF1A | 0.10082 |
| 41 | 5934 | RBL2 | 0.16701 | 41 | 9223 | BAP1 | 0.10069 |
| 42 | 8358 | HIST1H3B | 0.16666 | 42 | 4221 | MEN1 | 0.10049 |
| 43 | 23019 | CNOT1 | 0.1655 | 43 | 3845 | KRAS | 0.09984 |
| 44 | 8454 | CUL1 | 0.16319 | 44 | 2625 | GATA3 | 0.09885 |
| 45 | 55193 | PBRM1 | 0.15965 | 45 | 841 | CASP8 | 0.09882 |
| 46 | 677 | ZFP36L1 | 0.15549 | 46 | 9361 | PIM1 | 0.09702 |
| 47 | 10735 | STAG2 | 0.15149 | 47 | 2263 | FGFR2 | 0.09696 |
| 48 | 285382 | C3orf70 | 0.14784 | 48 | 2559 | GABRA6 | 0.09553 |
| 49 | 2261 | FGFR3 | 0.14772 | 49 | 1956 | EGFR | 0.09175 |
| 50 | 4763 | NF1 | 0.14239 | 50 | 54894 | RNF43 | 0.09159 |

Next, we wanted to analyze top DSI- and NDSI-ranked genes using several common gene list analysis tools. To this aim, we combined the lists of drivers from various classes. If the same gene was affected by more than one kind of alteration, we chose the alteration type with the highest DSI or NDSI, depending on the analysis. Also, we removed the data on chromosome arms and full chromosomes, as external pathway and network analysis tools can work only with genes. Then, we selected top 50 DSI- and NDSI-ranked genes. The resulting lists can be seen in **Table 1**.

First, we uploaded the resulting lists to “Reactome v77 Analyse gene list” tool and studied affected Reactome pathways on Voronoi visualizations (Reacfoam). It can be seen in **Fig 5** that top 50 DSI-ranked genes are significantly overrepresented in such categories as signaling by NOTCH, signaling by PTK6, ESR-mediated signaling, PIP3 activates AKT signaling, signaling by receptor tyrosine kinases, signaling by WNT, signaling by erythropoietin, RAF/MAP kinase cascade, signaling by TGF-beta receptor complex, mitotic cell cycle, meiosis, cell cycle checkpoints, DNA double-stand break repair, generic transcription pathway, epigenetic regulation of gene expression, RNA polymerase I transcription, circadian clock, chromatin modifying enzymes, diseases of signal transduction by growth factor receptors and second messengers, diseases of cellular senescence, diseases of programmed cell death, cellular responses to stress, activation of HOX genes during differentiation, and transcriptional regulation of granulopoiesis. Top 50 NDSI-ranked genes are significantly overrepresented in even fewer categories **(Fig 6)** – signaling by PTK6, extra-nuclear estrogen signaling, negative regulation of PI3K/AKT signaling, signaling by receptor tyrosine kinases, GPCR downstream signaling, erythropoietin activates RAS, RAF/MAP kinase cascade, cytokine signaling in immune system, adaptive immune system, hemostasis, generic transcription pathway, chromatin modifying enzymes, and diseases of signal transduction by growth factor receptors and second messengers. Surprisingly, several large categories often deemed important for cancer – cell cycle, DNA replication, DNA repair, autophagy, cellular responses to stress, programmed cell death, cell-cell communication and metabolism – are not affected.

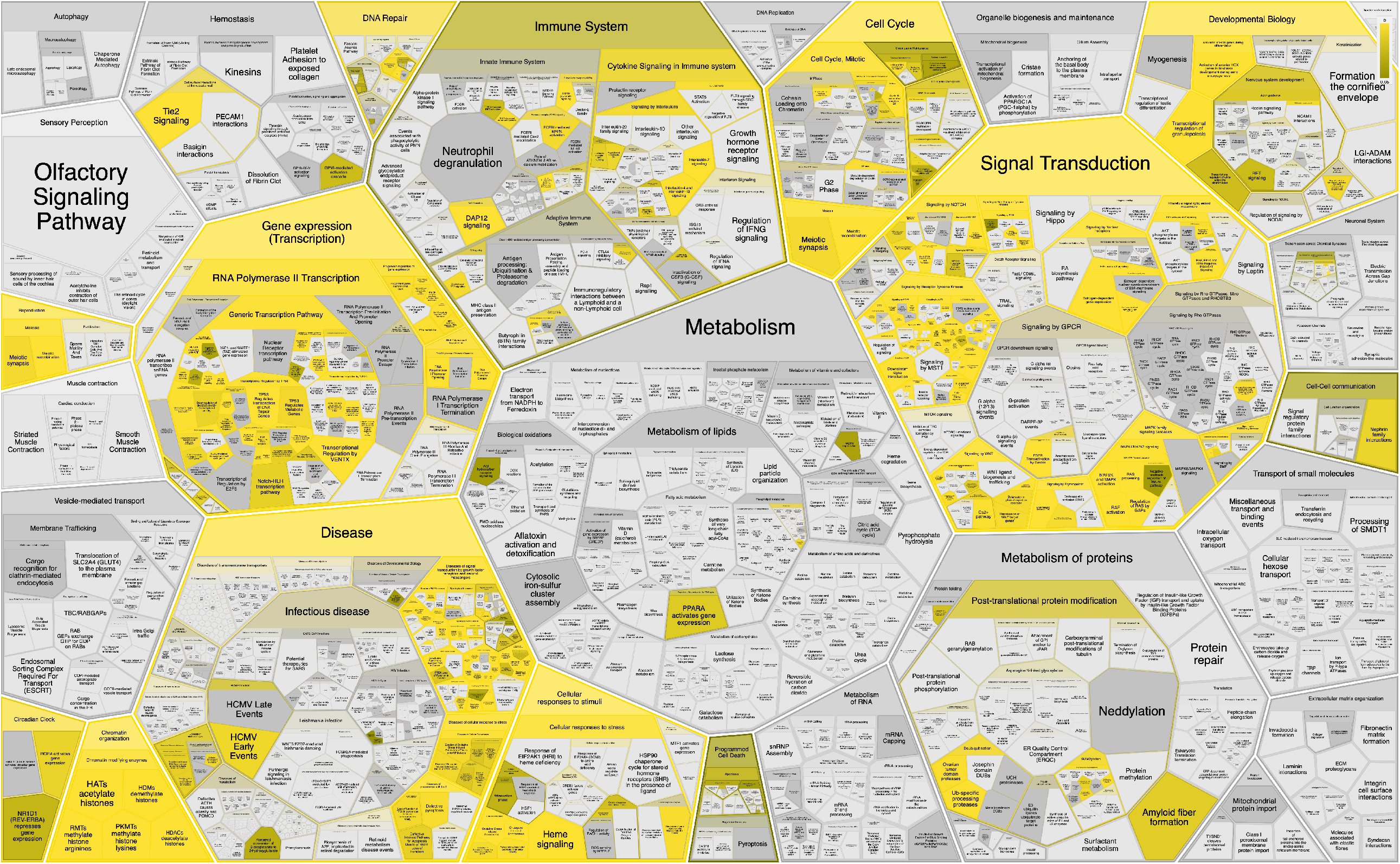

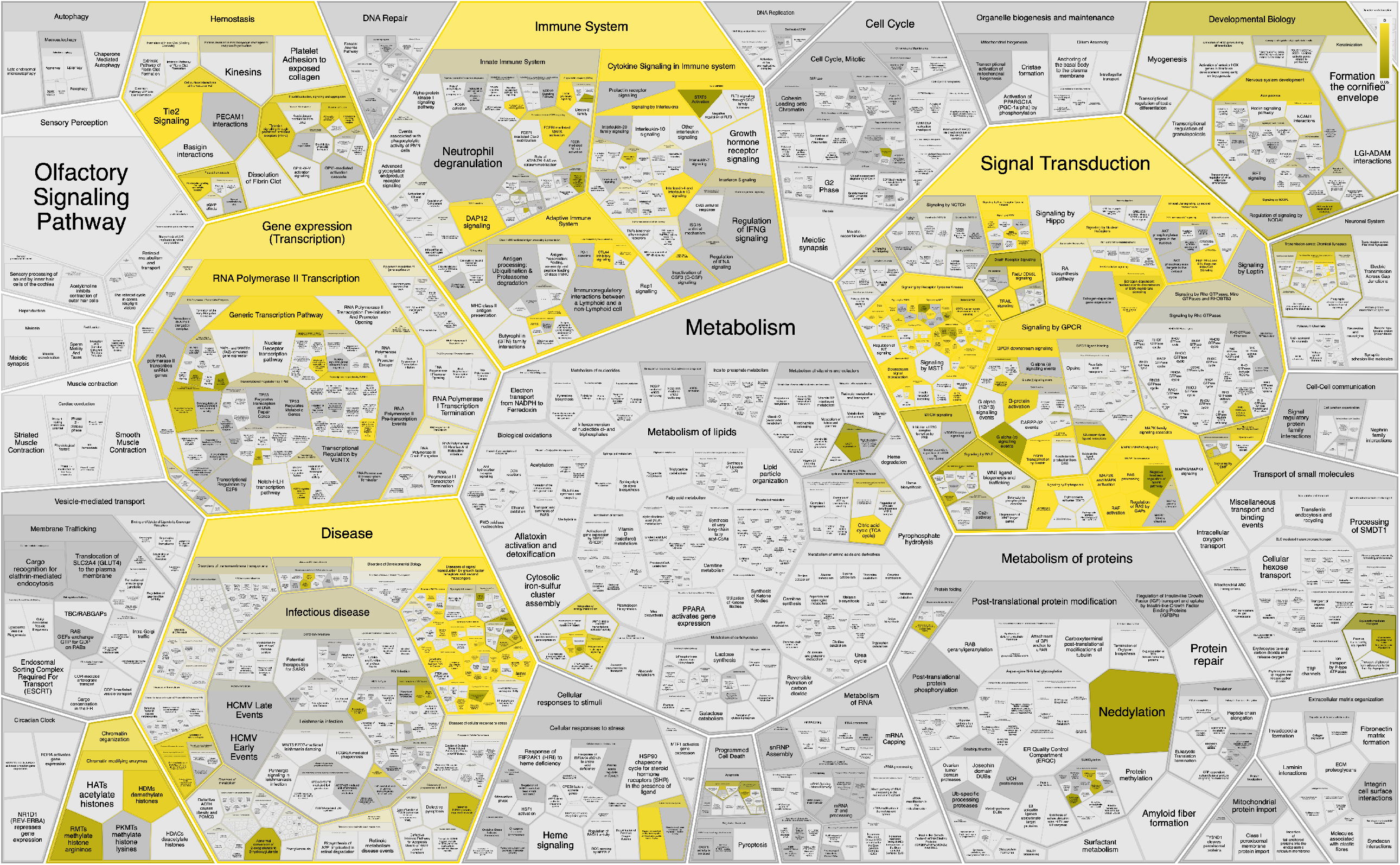

Next, we uploaded the resulting lists to “KEGG Mapper –Color” tool and mapped them to “Pathways in cancer - Homo sapiens (human)” (hsa05200) KEGG pathway map. **Fig 7** and **Table 1** together suggest that top DSI-ranked genes comprise *EGFR*/ERBB2/FGFR3-KRAS/NRAS/HRAS-BRAF-MYC pathway, PIK3CA-PTEN pathway, *CTNNB1*-MYC pathway, *TP53*-CDKN2A-RB1 pathway and MYC-CUL1-RB1 pathway. **Fig 8** and **Table 1** together suggest that top NDSI-ranked genes comprise *EGFR*/FGFR2/GNAQ/GNA11-NRAS/HRAS/KRAS-BRAF pathway, *AKT1*-MTOR pathway, and TCEB1-VHL-HIF1A pathway.

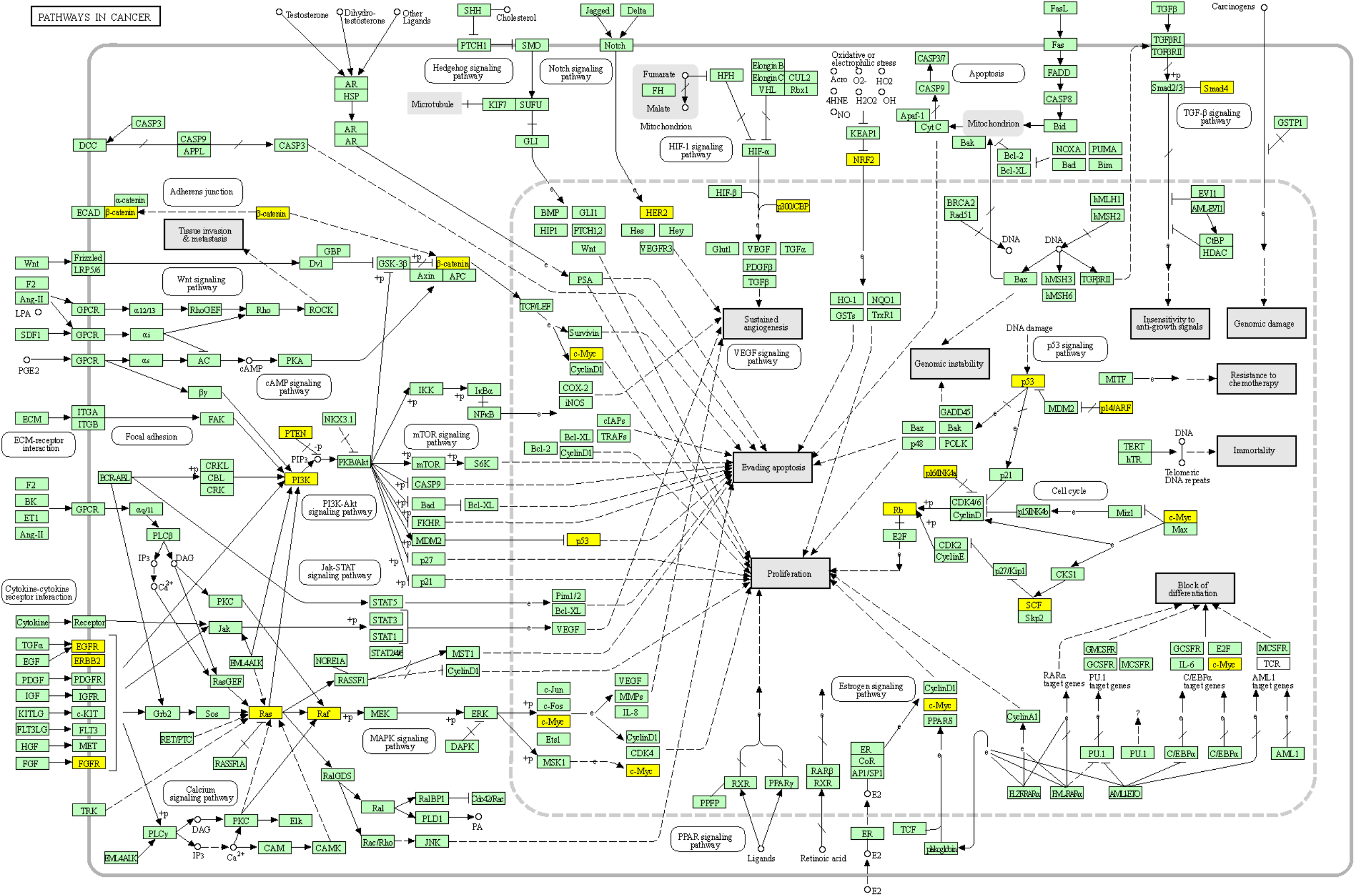

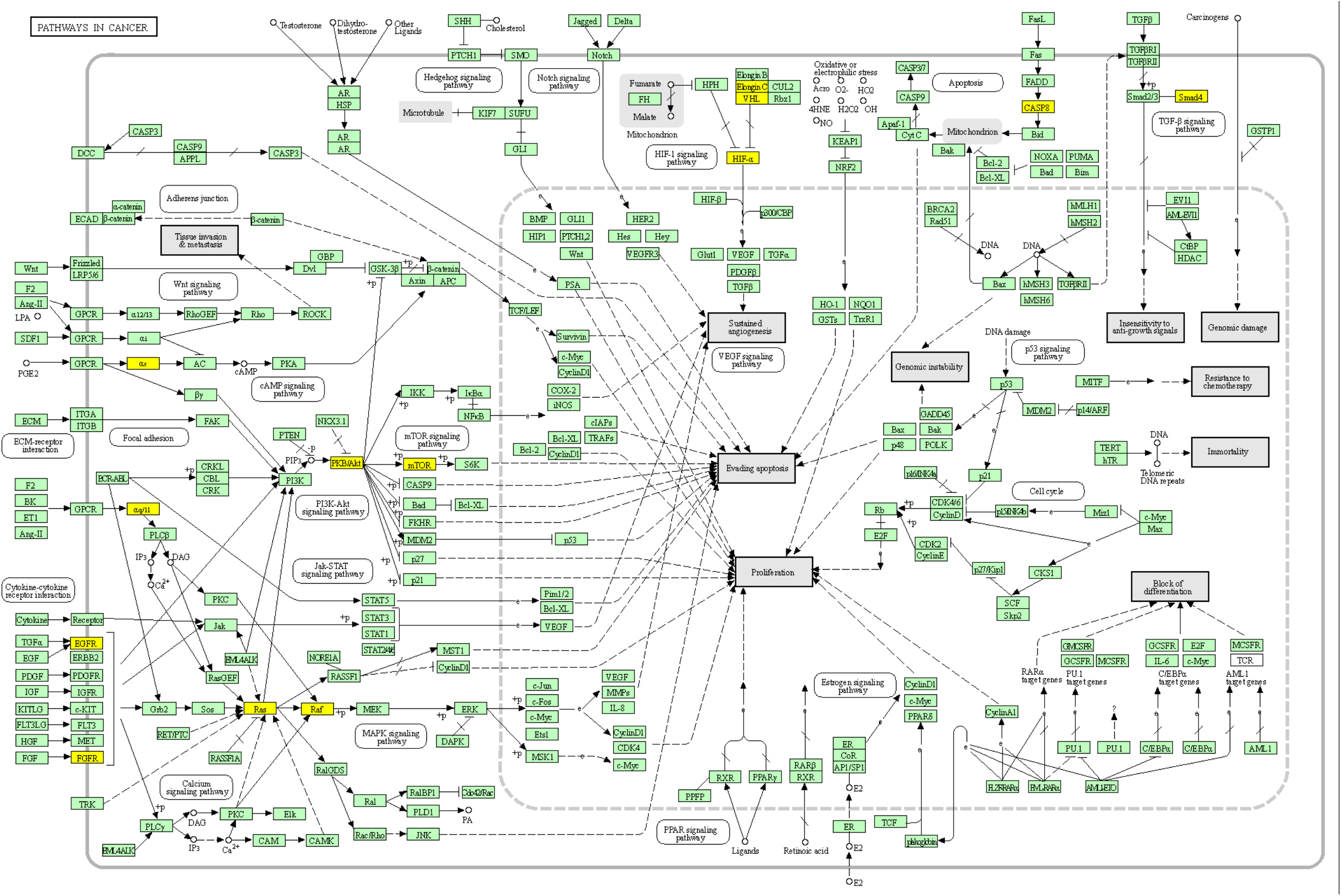

Finally, we analyzed the data in Cytoscape 3.8.2. We imported BioGRID: Protein-Protein Interactions (H. sapiens) network, appended (N)DSI values from the top 50 (N)DSI-ranked driver list, and mapped node color to (N)DSI values, whereas node size to the degree of connectedness. **Fig 9** shows that although *CTNNB1* and *EGFR* are the biggest hubs of the top-DSI-ranked gene network, their DSI values are much lower than those of *BRAF* and *PTEN*, which have fewer connections. Notably, *TP53* exhibited the highest DSI value and second-highest connectedness. Similarly, **Fig 10** shows that although *EGFR, AKT1* and *HRAS* are the biggest, centrally located hubs of the top-NDSI-ranked gene network, their NDSI values are much lower than those of *GTF2I, BRAF* and *ZMYM3*, located on the periphery of the network. Moreover, the top-NDSI-ranked gene network has much fewer edges than the top-DSI-ranked gene network, despite containing presumably stronger drivers. All of this supports our initially proposed notion that network centrality does not equal driver strength.

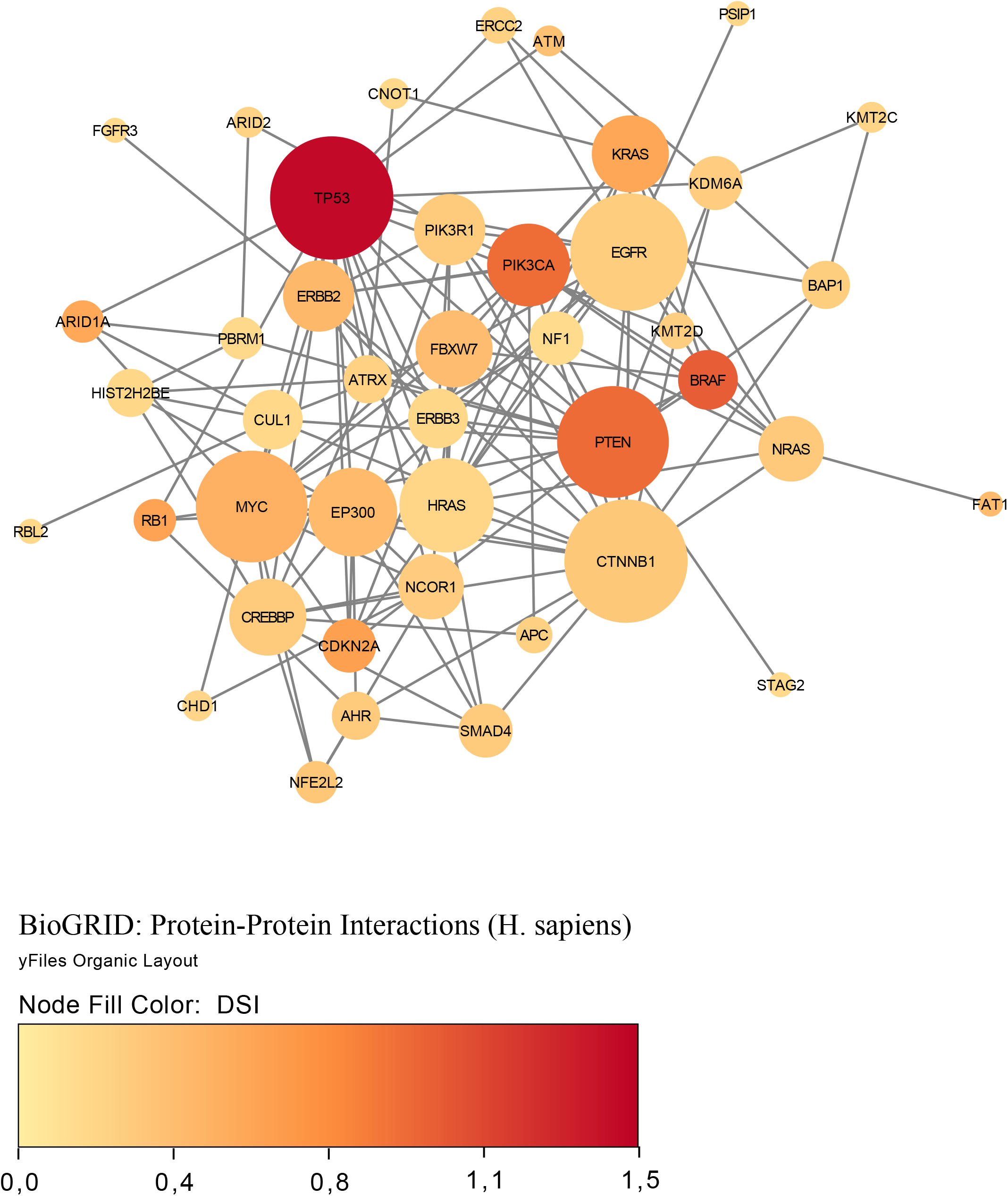

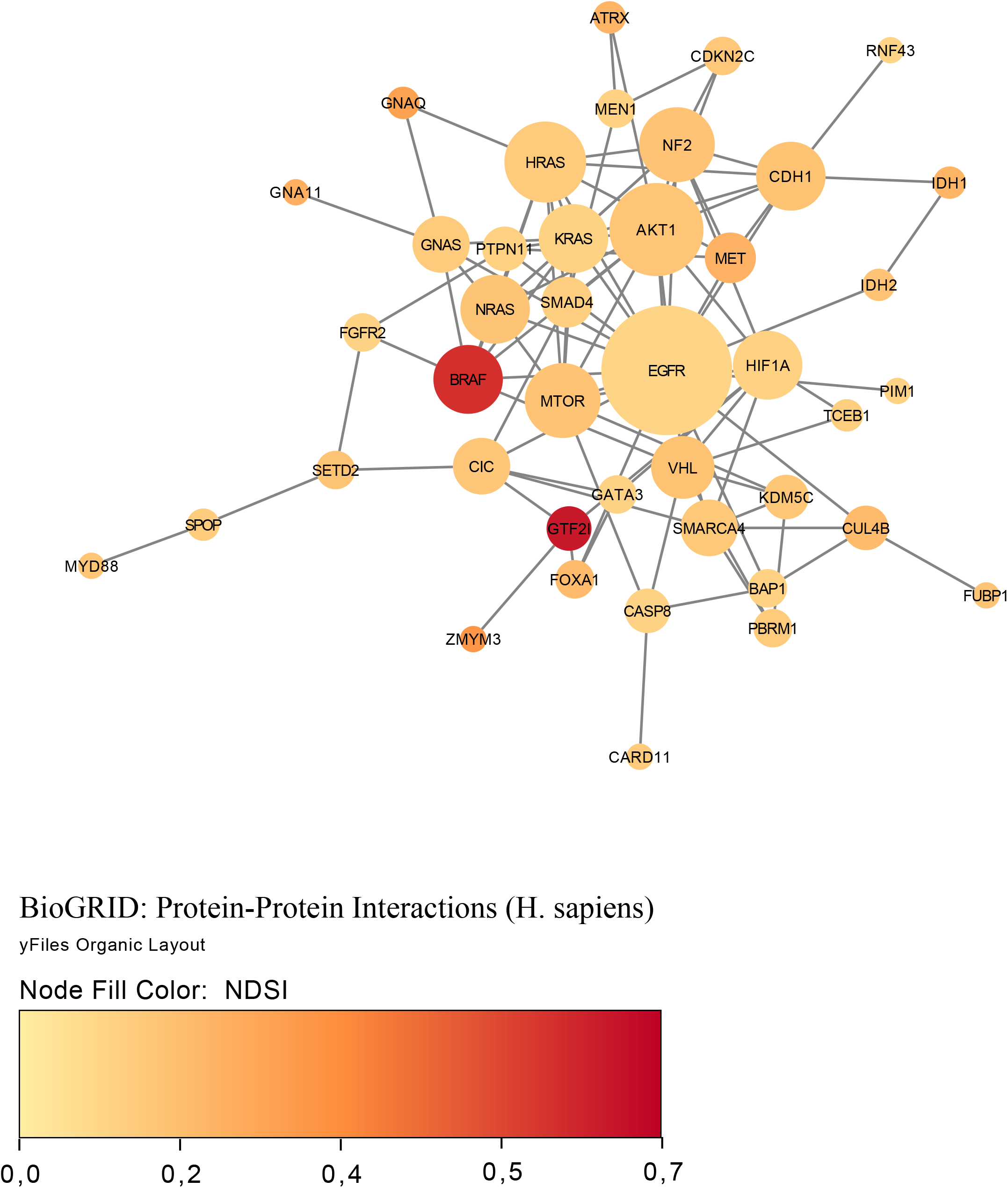

## Discussion

DSI places *BRAF* on the top spot amongst SNA-based oncogenes and on the second place amongst drivers of all classes. *BRAF* encodes a protein belonging to the RAF family of serine/threonine protein kinases. *BRAF* plays a role in regulating the MAP kinase/ERK signaling pathway, which affects cell division and differentiation. Mutations in *BRAF*, most commonly the V600E mutation, are the most frequently identified cancer-causing mutations in melanoma, and have been identified in various other cancers as well, including non-Hodgkin lymphoma, colorectal cancer, thyroid carcinoma, non-small cell lung carcinoma, hairy cell leukemia and adenocarcinoma of lung (15). Our analysis shows frequent mutations and amplifications of *BRAF* in TCGA COAD, GBM, LGG, LUAD, LUSC, PRAD, SKCM and THCA cohorts. From recent studies, a new classification system is emerging for *BRAF* mutations based on biochemical and signaling mechanisms associated with these mutants. Class I *BRAF* mutations affect amino acid V600 and lead to *BRAF* protein signaling as RAS-independent active monomer, class II mutations make *BRAF* proteins function as RAS-independent activated dimers, and class III mutations impair *BRAF* kinase activity but increase signaling through the MAPK pathway due to enhanced RAS binding and subsequent CRAF activation (16). It would be interesting to rate the strength of these *BRAF* mutation classes using NDSI.

DSI prioritizes *KRAS, NRAS* and *HRAS* amongst SNA-based oncogenes. These genes belong to the RAS oncogene family, whose members are related to the transforming genes of mammalian sarcoma retroviruses. The products encoded by these genes function in signal transduction pathways. These proteins can bind GTP and GDP, and they have intrinsic GTPase activity. Mutations in the RAS family of proteins have frequently been observed across cancer types, including lung adenocarcinoma, ductal carcinoma of the pancreas, colorectal carcinoma, follicular thyroid cancer, juvenile myelomonocytic leukemia, bladder cancer, and oral squamous cell carcinoma (17). Our analysis shows frequent mutations and amplifications of *KRAS, NRAS* and *HRAS* in TCGA BLCA, BRCA, CESC, COAD, ESCA, HNSC, KIRP, LGG, LIHC, LUAD, LUSC, OV, PAAD, PRAD, READ, SKCM, STAD, TGCT, THCA, THYM, UCEC and UCS cohorts. Gain-of-function missense mutations, mostly located at codons 12, 13, and 61, constitutively activate RAS proteins, however, each isoform exhibits distinctive mutation frequency at each codon, supporting the hypothesis that different RAS mutants may lead to distinct biologic manifestations (18).

DSI prioritizes *HIST2H2BE* and *HIST1H3B* amongst CNA-based oncogenes. These genes encode replication-dependent histones that are members of the histone H2B and H3.1 families. *HIST2H2BE* is a direct target of the tumor suppressor *IRX1* in gastric cancer (19). HIST2H2BE is one of the few surface proteins of exosomes from pancreatic ductal adenocarcinoma (20). Elevated HIST2H2BE has recently been shown to promote progression of breast invasive ductal carcinoma (21). p.Lys27Met mutation in HIST1H3B is found in the majority of pediatric diffuse intrinsic pontine gliomas and many non-brainstem pediatric glioblastomas (22). HIST1H3B K27M-mutated tumors exhibit a mesenchymal/astrocytic phenotype and a pro-angiogenic/hypoxic signature (23). *HIST1H3B* mutations are also identified in cell-free circulating tumor DNA in the cerebrospinal fluid of patients with diffuse gliomas (24). *HIST1H3B* is amplified and overexpressed in 19% of liver cancer patients (25). Autoantibodies to HIST1H3B have been recently found in high quantities in both early and advanced stage lung cancer patients (26). Cancer-associated mutations of histones H2B and H3.1 affect the structure and stability of the nucleosome (27). Our analysis shows frequent amplifications of *HIST2H2BE* and *HIST1H3B* in TCGA BLCA (bladder urothelial carcinoma) cohort.

DSI prioritizes *KMT2C* and *KMT2D* amongst SNA-based tumor suppressors. The proteins encoded by these genes are histone methyltransferases that methylate the Lys-4 position of histone H3 (H3K4me). H3K4me represents a specific tag for epigenetic transcriptional activation. The encoded proteins are part of a large protein complex called ASCOM, which is a transcriptional regulator of the beta-globin and estrogen receptor genes. Whereas *KMT2C* loss disrupts estrogen-driven proliferation, it conversely promotes tumor outgrowth under hormone-depleted conditions (28). In accordance, *KMT2C* is one of the most frequently mutated genes in estrogen receptor-positive breast cancer with *KMT2C* deletion correlating with significantly shorter progression-free survival on anti-estrogen therapy (28). *KMT2D* is among the most highly inactivated epigenetic modifiers in lung cancer (29). Recently, it has been shown that *KMT2D* loss widely impairs epigenomic signals for super-enhancers/enhancers, including the super-enhancer for the circadian rhythm repressor *PER2* (29). Loss of *KMT2D* decreases expression of *PER2*, leading to increase in glycolytic gene expression (29). The role of *KMT2C* and *KMT2D* in cancer has been recently reviewed (30). Our analysis shows frequent mutations and deletions of *KMT2C* and *KMT2D* in TCGA BLCA, BRCA, CESC, DLBC, ESCA, HNSC, LIHC, LUSC, PRAD, STAD and UCEC cohorts.

NDSI places *GTF2I* on the top spot both amongst the strongest SNA-based oncogenes and amongst the strongest drivers averaged across all classes. *GTF2I* encoded protein binds to the initiator element (Inr) and E-box element in promoters and functions as a regulator of transcription. *GTF2I* c.74146970 T>A mutation was detected in 82% of type A and 74% of type AB thymomas (31). *GTF2I* β and δ isoforms are expressed in thymomas, and both mutant isoforms are able to stimulate cell proliferation in vitro (31). Recently, it has been shown that expression of mutant *GTF2I* alters the transcriptome of normal thymic epithelial cells and upregulates several oncogenic genes (32). *GTF2I* L424H knockin cells exhibit cell transformation, aneuploidy, and increased tumor growth and survival under glucose deprivation or DNA damage (32). Our analysis also shows frequent mutations of *GTF2I* in TCGA THYM (thymoma) cohort. *GTF2I* has been recently named gene of the month and its role in cancer reviewed (33).

*SPOP* is categorized by NDSI as the strongest CNA-based oncogene. *SPOP* encodes a protein that is a component of a cullin-RING-based E3 ubiquitin-protein ligase complex that mediates the ubiquitination of target proteins, leading most often to their proteasomal degradation. *SPOP* is the most commonly mutated gene in primary prostate cancer (34). *SPOP* mutations in prostate cancer result in impaired homology-directed repair of double strand breaks and are associated with genomic instability (35). As most cancer-associated mutations in *SPOP* are missense and almost none are frameshift or nonsense, PALDRIC classifies it as an oncogene. However, *SPOP* is usually viewed as a tumor suppressor (36). Recently it has been discussed that SPOP actually has a dual role, and while being a tumor suppressor in prostate cancer it performs as an oncogene in kidney cancer (37). Indeed, cytoplasmic accumulation of SPOP leads to the ubiquitination and degradation of multiple regulators of cellular proliferation and apoptosis, including the tumor suppressor PTEN, ERK phosphatases, the proapoptotic molecule DAXX, and the Hedgehog pathway transcription factor GLI2 and is sufficient to induce tumorigenesis in clear cell renal cell carcinoma (38). Our analysis shows frequent mutations and amplifications of *SPOP* in TCGA PRAD (prostate adenocarcinoma) and UCEC (uterine corpus endometrial carcinoma) cohorts.

*MET* is the strongest mixed (SNA+CNA) oncogene and the second-strongest CNA-based oncogene, according to NDSI rating. *MET* encodes a receptor tyrosine kinase that transduces signals from the extracellular space into the cytoplasm by binding to hepatocyte growth factor ligand. MET regulates many physiological processes including proliferation, morphogenesis and survival. Ligand binding at the cell surface induces dimerization and autophosphorylation of MET on its intracellular domain that provides docking sites for downstream signaling molecules. Following activation by its ligand, MET interacts with the PI3-kinase subunit PIK3R1, PLCG1, SRC, GRB2, STAT3 or the adapter GAB1. Recruitment of these downstream effectors by MET leads to the activation of several signaling cascades including the RAS-ERK, PI3K-AKT, or PLCG-PKC. Mutations in *MET* are associated with papillary renal cell carcinoma, hepatocellular carcinoma, and various head and neck cancers. Amplification and overexpression of this gene are also associated with multiple human cancers (39,40). Our analysis shows frequent mutations and amplifications of *MET* in TCGA KIRP (kidney renal papillary cell carcinoma) and LUAD (lung adenocarcinoma) cohorts.

*ATRX* is ranked by NDSI as the strongest SNA-based tumor suppressor, 9^th^ strongest CNA-based tumor suppressor, 5^th^ strongest mixed (SNA+CNA) tumor suppressor and 9^th^ strongest driver averaged across all classes. *ATRX* (Alpha-Thalassemia/Mental Retardation Syndrome, X-Linked) encodes a protein that contains an ATPase/helicase domain, and thus it belongs to the SWI/SNF family of chromatin remodeling proteins. *ATRX* together with *DAXX* encode a complex that deposits histone variant H3.3 into repetitive heterochromatin, including retrotransposons, pericentric heterochromatin, and telomeres, the latter of which show deregulation in *ATRX*/*DAXX*-mutant tumors (41,42). *ATRX* loss induces extensive changes in chromatin accessibility in both repetitive DNA regions and non-repetitive regulatory regions (43). These changes are highly correlated with changes in transcription, which lead to alterations in cancer-related signaling pathways, such as upregulation of the TGF-β pathway and downregulation of the cadherin family of proteins (43). Our analysis shows frequent mutations and deletions of *ATRX* in TCGA ACC, GBM, LGG and SARC cohorts.

*CSDE1* is ranked by NDSI as the strongest CNA-based tumor suppressor and 5^th^ strongest driver averaged across all classes. *CSDE1* encodes for an RNA-binding protein involved in translationally coupled mRNA turnover. CSDE1 not only promotes and represses the translation of RNAs but also increases and decreases the abundance of RNAs (44). *CSDE1* loss-of-function mutations and deletions define a Wnt-altered subtype of pheochromocytomas and paragangliomas (45). Our analysis also shows frequent deletions of *CSDE1* in TCGA PCPG (pheochromocytoma and paraganglioma) cohort.

*NF2* is ranked by NDSI as the strongest mixed tumor suppressor and 3^rd^ strongest CNA-based tumor suppressor. *NF2* encodes Merlin (Moesin-ezrin-radixin-like protein), also known as Neurofibromin 2 and Schwannomin. Merlin is a tumor suppressor classically known for its ability to induce contact-dependent growth inhibition (46). Loss-of-function mutations or deletions in *NF2* cause neurofibromatosis type 2, a multiple tumor forming disease of the nervous system, characterized by the development of bilateral schwannomas, as well as meningiomas and ependymomas (47). *NF2* is also mutated and deleted in mesotheliomas (48), clear cell renal cell carcinomas (49), collecting duct carcinomas of the kidney (50), and renal cell carcinomas with sarcomatoid dedifferentiation (51). Our analysis shows frequent mutations and deletions of *NF2* in TCGA KIRC (kidney renal clear cell carcinoma), KIRP (kidney renal papillary cell carcinoma) and MESO (mesothelioma) cohorts.

Interestingly, NDSI prioritized three members of guanine nucleotide-binding protein (G protein) family: *GNAQ, GNA11*, and *GNAS*. Guanine nucleotide-binding proteins function as transducers downstream of G protein-coupled receptors (GPCRs) in numerous signaling cascades. The alpha chain contains the guanine nucleotide binding site and alternates between an active, GTP-bound state and an inactive, GDP-bound state. Signaling by an activated GPCR promotes GDP release and GTP binding. The alpha subunit has a low GTPase activity that converts bound GTP to GDP, thereby terminating the signal. *GNAQ*-encoded protein, an α subunit in the Gq class, couples a seven-transmembrane-domain receptor to activation of PLCβ. Some *GNAQ* cancer mutants display normal basal activity and GPCR-mediated activation, but deactivate slowly due to GTPase activating protein (GAP) insensitivity (52). *GNAQ* mutations occur in about half of uveal melanomas, representing the most common known oncogenic mutation in this cancer (53). The presence of this mutation in tumors at all stages of malignant progression suggests that it is an early event in uveal melanoma (53). Mutations affecting Q209 in *GNAQ* were present in 45% of primary uveal melanomas and 22% of uveal melanoma metastases (54). Our analysis also shows frequent mutations of *GNAQ* in TCGA UVM (uveal melanoma) cohort. Recently, of the 11111 patients screened, 117 patients have been found to harbor *GNAQ*/*GNA11* mutations, in melanoma, colorectal, liver, glioma, lung, bile duct and gastric cancers (55). *GNA11* encodes subunit α-11 in the Gq class and acts as an activator of PLC. Mutations affecting Q209 in *GNA11* were present in 32% of primary uveal melanomas and 57% of uveal melanoma metastases (54). Our analysis also shows frequent mutations of *GNA11* in TCGA UVM (uveal melanoma) cohort. *GNAS* encodes subunit α in the Gs class and participates in the activation of adenylyl cyclases, resulting in increased levels of the signaling molecule cAMP. GNAS functions downstream of several GPCRs, including beta-adrenergic receptors. *GNAS* mutations are found in 67% of intraductal papillary mucinous neoplasms and many associated pancreatic ductal adenocarcinomas (56). High GNAS expression in breast tumor tissue showed a close correlation with a reduced overall survival, frequent distal metastasis, advanced clinical stage, stronger cell proliferation and enhanced cancer cell migration (57). Recently, it has been shown that GNAS promotes the development of small cell lung cancer via PKA (58). Our analysis shows frequent mutations and amplifications of *GNAS* in TCGA COAD (colon adenocarcinoma), LIHC (liver hepatocellular carcinoma) and READ (rectum adenocarcinoma) cohorts. The current knowledge on cancer-associated alterations of GPCRs and G proteins has been recently reviewed (59). Strikingly, approximately 36% of all drugs approved by the US Food and Drug Administration during the past three decades target GPCRs (60).

Two members of isocitrate dehydrogenase family, *IDH1* and *IDH2*, appeared on the top 10 SNA-based oncogenic events list as ranked by NDSI. The protein encoded by *IDH1* is the NADP^+^-dependent isocitrate dehydrogenase found in the cytoplasm and peroxisomes. The cytoplasmic enzyme serves a significant role in cytoplasmic NADPH production. The protein encoded by *IDH2* is the NADP^+^-dependent isocitrate dehydrogenase found in the mitochondria. It plays a role in intermediary metabolism and energy production. The most frequent mutations R132 (*IDH1*) and R172 (*IDH2*) involve the active site and result in simultaneous loss of normal catalytic activity, the production of α-ketoglutarate, and gain of a new function, the production of 2-hydroxyglutarate (61–64). 2-hydroxyglutarate is structurally similar to α-ketoglutarate, and acts as an α-ketoglutarate antagonist to competitively inhibit multiple α-ketoglutarate– dependent dioxygenases, including both lysine histone demethylases and the ten-eleven translocation family of DNA hydroxylases (64). Abnormal histone and DNA methylation are emerging as a common feature of tumors with *IDH1* and *IDH2* mutations and may cause altered stem cell differentiation and eventual tumorigenesis (64). In acute myeloid leukemia, *IDH1* and *IDH2* mutations have been associated with worse outcome, shorter overall survival, and normal karyotype (65). All the 1p19q co-deleted gliomas are mutated on *IDH1* or *IDH2* (66). Our analysis shows frequent mutations of *IDH1* and *IDH2* in TCGA LGG (lower grade glioma) cohort and frequent amplifications of *IDH1* in LIHC (liver hepatocellular carcinoma), as well as less frequent mutations and amplifications of *IDH1* in CHOL, GBM, PRAD and SKCM.

Two fibroblast growth factor receptors *FGFR2* and *FGFR3* appeared on the top 10 mixed oncogenic events list as ranked by NDSI. The extracellular region of these proteins, composed of three immunoglobulin-like domains, interacts with fibroblast growth factors, leading to the activation of a cytoplasmic tyrosine kinase domain that phosphorylates PLCG1, FRS2 and other proteins. This sets in motion a cascade of downstream signals, including RAS-MAPK and PI3K-AKT pathways, ultimately influencing cell proliferation, differentiation, migration and apoptosis. FGFR aberrations were found in 7.1% of cancers, with the majority being gene amplification (66% of the aberrations), followed by mutations (26%) and rearrangements (8%) (67). FGFR1 was affected in 3.5% of 4,853 patients; *FGFR2* in 1.5%; *FGFR3* in 2.0%; and FGFR4 in 0.5% (67). The cancers most commonly affected were urothelial (32% FGFR-aberrant); breast (18%); endometrial (∼13%), squamous lung cancers (∼13%), and ovarian cancer (∼9%) (67). Our analysis also shows frequent mutations and amplifications of *FGFR2* in TCGA LUSC (lung squamous cell carcinoma) and UCEC (uterine corpus endometrial carcinoma) cohorts, as well as frequent mutations and amplifications of *FGFR3* in BLCA (bladder urothelial carcinoma), HNSC (head and neck squamous cell carcinoma) and LUSC (lung squamous cell carcinoma) cohorts.

While both DSI- and NDSI-ranked top 50 genes are significantly overrepresented in such Reactome categories as signaling by PTK6, ESR-mediated signaling, PIP3 activates AKT signaling, signaling by receptor tyrosine kinases, signaling by erythropoietin, RAF/MAP kinase cascade, generic transcription pathway, chromatin modifying enzymes, and diseases of signal transduction by growth factor receptors and second messengers, there are also multiple differences. Top 50 DSI-ranked genes are additionally overrepresented in signaling by NOTCH, signaling by WNT, signaling by TGF-beta receptor complex, mitotic cell cycle, meiosis, cell cycle checkpoints, DNA double-stand break repair, epigenetic regulation of gene expression, RNA polymerase I transcription, circadian clock, diseases of cellular senescence, diseases of programmed cell death, and cellular responses to stress. This suggests that although these pathways are frequently mutated in cancer, none of them possesses strong tumor-promoting activity on its own. On the other hand, top 50 NDSI-ranked genes are additionally overrepresented in GPCR downstream signaling, which suggests that although this pathway is mutated more rarely in cancer, it nevertheless has very strong tumor-promoting activity. It is also peculiar why neither DSI-nor NDSI-ranked top 50 genes are significantly overrepresented in DNA replication, autophagy, and metabolism categories. This may indicate that the role of these processes in oncogenesis is overestimated.

The major signaling pathway activated by mutations in both top DSI- and top NDSI-ranked driver genes is the RAS-RAF pathway. Although the pathway can be triggered via mutations in *EGFR, FGFR, NRAS, HRAS, KRAS* and *BRAF* genes, all of which are in top 50 of both DSI and NDSI rankings, it can be additionally engaged through mutations in top DSI-ranked driver *ERBB2* and top NDSI-ranked drivers *GNAQ* and *GNA11*. This suggests that *ERBB2* driver mutations occur more frequently but are weaker than *GNAQ* and *GNA11* driver mutations. Also, top DSI-ranked driver mutations affect the upper part of the PI3K-AKT-MTOR pathway via constitutive *PIK3CA* activation or PTEN inactivation, whereas top NDSI-ranked mutations affect the lower part of the pathway by activating AKT1 and MTOR. Similarly, this suggests that *PIK3CA* and *PTEN* driver mutations occur more frequently but are weaker than *AKT1* and *MTOR* driver mutations. Moreover, CTNNB1-MYC pathway, TP53-CDKN2A-RB1 pathway and *MYC*-CUL1-RB1 pathway are engaged only by top DSI-ranked drivers, indicating their relative weakness in cancer promotion despite high frequency of mutation, whereas TCEB1-VHL-HIF1A pathway - only by top NDSI-ranked drivers, suggesting that this pathway has very strong tumor-promoting potential whilst being mutated more rarely.

Overall, we presented a comprehensive overview on the landscape of cancer driver genes and chromosomes in TCGA PanCanAtlas patients and highlighted particular genes, gene families and pathways deemed strong drivers according to our Normalized Driver Strength Index. A puzzling question that remains in cancer genomics is why mutations in a given driver gene are typically confined to one or a few cancer types, resulting in each cancer type having its own unique set of driver genes (68)? As mutations are supposed to happen randomly as a result of stochastic mutagenesis processes (69,70), it is logical to suggest that mutations in different tissues can affect the same genes. However, the same mutation can be selected for in some tissues and selected against in others (71). This selection most likely depends on the tissue-specific epigenetic profiles and microenvironments of the cancer-initiating stem or progenitor cells (72,73). Thus, investigating the interplay between stem cell mutations, epigenetic profiles and microenvironments in various tissues appears to be a promising and exciting avenue for future research.

## Methods

### Source files and initial filtering

TCGA PanCanAtlas data were used. Files “Analyte level annotations - merged_sample_quality_annotations.tsv”, “ABSOLUTE purity/ploidy file - TCGA_mastercalls.abs_tables_JSedit.fixed.txt”, “Aneuploidy scores and arm calls file - PANCAN_ArmCallsAndAneuploidyScore_092817.txt”, “Public mutation annotation file - mc3.v0.2.8.PUBLIC.maf.gz”, “gzipped ISAR-corrected GISTIC2.0 all_thresholded.by_genes file - ISAR_GISTIC.all_thresholded.by_genes.txt”, “RNA batch corrected matrix - EBPlusPlusAdjustPANCAN_IlluminaHiSeq_RNASeqV2.geneExp.tsv”, “miRNA batch corrected matrix - pancanMiRs_EBadjOnProtocolPlatformWithoutRepsWithUnCorrectMiRs_08_04_16.csv”, were downloaded from https://gdc.cancer.gov/about-data/publications/PanCan-CellOfOrigin.

Using TCGA barcodes (see https://docs.gdc.cancer.gov/Encyclopedia/pages/TCGA_Barcode/ and https://gdc.cancer.gov/resources-tcga-users/tcga-code-tables/sample-type-codes), all samples except primary tumors (barcoded 01, 03, 09) were removed from all files. Based on the information in the column “Do_not_use” in the file “Analyte level annotations - merged_sample_quality_annotations.tsv”, all samples with “True” value were removed from all files. All samples with “Cancer DNA fraction” <0.5 or unknown or with “Subclonal genome fraction” >0.5 or unknown in the file “TCGA_mastercalls.abs_tables_JSedit.fixed.txt” were removed from the file “PANCAN_ArmCallsAndAneuploidyScore_092817.txt”. Moreover, all samples without “PASS” value in the column “FILTER” were removed from the file “mc3.v0.2.8.PUBLIC.maf.gz” and zeros in the column “Entrez_Gene_Id” were replaced with actual Entrez gene IDs, determined from the corresponding ENSEMBL gene IDs in the column “Gene” and external database ftp://ftp.ncbi.nih.gov/gene/DATA/GENE_INFO/Mammalia/Homo_sapiens.gene_info.gz. Filtered files were saved as “Primary_whitelisted_arms.tsv”, “mc3.v0.2.8.PUBLIC_primary_whitelisted_Entrez.tsv”, “ISAR_GISTIC.all_thresholded.by_genes_primary_whitelisted.tsv”, “EBPlusPlusAdjustPANCAN_IlluminaHiSeq_RNASeqV2-v2.geneExp_primary_whitelisted.tsv”, “pancanMiRs_EBadjOnProtocolPlatformWithoutRepsWithUnCorrectMiRs_08_04_16_primary_whitelisted.tsv”.

### RNA filtering of CNAs

Using the file “EBPlusPlusAdjustPANCAN_IlluminaHiSeq_RNASeqV2-v2.geneExp_primary_whitelisted.tsv”, the median expression level for each gene across patients was determined. If the expression for a given gene in a given patient was below 0.05x median value, it was encoded as “-2”, if between 0.05x and 0.75x median value, it was encoded as “-1”, if between 1.25x and 1.75x median value, it was encoded as “1”, if above 1.75x median value, it was encoded as “2”, otherwise it was encoded as “0”. The file was saved as “EBPlusPlusAdjustPANCAN_IlluminaHiSeq_RNASeqV2-v2.geneExp_primary_whitelisted_median.tsv.” The same operations were performed with the file “pancanMiRs_EBadjOnProtocolPlatformWithoutRepsWithUnCorrectMiRs_08_04_16_primary_whitelisted.tsv”, which was saved as “pancanMiRs_EBadjOnProtocolPlatformWithoutRepsWithUnCorrectMiRs_08_04_16_primary_ whitelisted_median.tsv” Next, the file “ISAR_GISTIC.all_thresholded.by_genes_primary_whitelisted.tsv” was processed according to the following rules: if the gene CNA status in a given patient was not zero and had the same sign as the gene expression status in the same patient (file “EBPlusPlusAdjustPANCAN_IlluminaHiSeq_RNASeqV2-v2.geneExp_primary_whitelisted_median.tsv” or “pancanMiRs_EBadjOnProtocolPlatformWithoutRepsWithUnCorrectMiRs_08_04_16_primary_ whitelisted_median.tsv” for miRNA genes), then the CNA status value was replaced with the gene expression status value, otherwise it was replaced by zero. If the corresponding expression status for a given gene was not found then its CNA status was not changed. The resulting file was saved as “ISAR_GISTIC.all_thresholded.by_genes_primary_whitelisted_RNAfiltered.tsv”

We named this algorithm GECNAV (Gene Expression-based CNA Validator) and created a Github repository: https://github.com/belikov-av/GECNAV. The package used to generate data in this article is available as **Additional file 3**.

### Aneuploidy driver prediction

Using the file “Primary_whitelisted_arms.tsv”, the average alteration status of each chromosomal arm was calculated for each cancer type and saved as a matrix file “Arm_averages.tsv”. By drawing statuses randomly with replacement (bootstrapping) from any cell of “Primary_whitelisted_arms.tsv”, for each cancer type the number of statuses corresponding to the number of patients in that cancer type were generated and their average was calculated. The procedure was repeated 10000 times, the median for each cancer type was calculated and the results were saved as a matrix file “Bootstrapped_arm_averages.tsv”.

P-value for each arm alteration status was calculated for each cancer type. To do this, first the alteration status for a given cancer type and a given arm in “Arm_averages.tsv” was compared to the median bootstrapped arm alteration status for this cancer type in “Bootstrapped_arm_averages.tsv”. If the status in “Arm_averages.tsv” was higher than zero and the median in “Bootstrapped_arm_averages.tsv”, the number of statuses for this cancer type in “Bootstrapped_arm_averages.tsv” that are higher than the status in “Arm_averages.tsv” was counted and divided by 5000. If the status in “Arm_averages.tsv” was lower than zero and the median in “Bootstrapped_arm_averages.tsv”, the number of statuses for this cancer type in “Bootstrapped_arm_averages.tsv” that are lower than the status in “Arm_averages.tsv” was counted and divided by 5000, and marked with minus to indicate arm loss. Other values were ignored (cells left empty). The results were saved as a matrix file “Arm_Pvalues_cohorts.tsv”.

For each cancer type, Benjamini–Hochberg procedure with FDR=5% was applied to P-values in “Arm_Pvalues_cohorts.tsv” and passing P-values were encoded as “DAG” (Driver arm gain) or “DAL” (Driver arm loss) if marked with minus. The other cells were made empty and the results were saved as a matrix file “Arm_drivers_FDR5_cohorts.tsv”.

Alterations were classified according to the following rules: if the arm status in a given patient (file “Primary_whitelisted_arms.tsv”) was “-1” and the average alteration status of a given arm in the same cancer type (file “Arm_drivers_FDR5_cohorts.tsv”) was “DAL”, then the alteration in the patient was classified as “DAL”. If the arm status in a given patient was “1” and the average alteration status of a given arm in the same cancer type was “DAG”, then the alteration in the patient was classified as “DAG”. In all other cases an empty cell was written. The total number of DALs and DAGs was calculated, patients with zero drivers were removed, and the results were saved as a matrix file “Arm_drivers_FDR5.tsv”.

Using the file “Primary_whitelisted_arms.tsv”, the values for the whole chromosomes were calculated using the following rules: if both p- and q-arm statuses were “1” then the chromosome status was written as “1”; if both p- and q-arm statuses were “-1” then the chromosome status was written as “-1”; if at least one arm status was not known (empty cell) then the chromosome status was written as empty cell; in all other cases the chromosome status was written as “0”. For one-arm chromosomes (13, 14, 15, 21, 22), their status equals the status of the arm. The resulting file was saved as “Primary_whitelisted_chromosomes.tsv”.

The same procedures as described above for chromosomal arms were repeated for the whole chromosomes, with the resulting file “Chromosome_drivers_FDR5.tsv”. Chromosome drivers were considered to override arm drivers, so if a chromosome had “DCL” (Driver chromosome loss) or “DCG” (Driver chromosome gain), no alterations were counted on the arm level, to prevent triple counting of the same event.

We named this algorithm ANDRIF (ANeuploidy DRIver Finder) and created a Github repository: https://github.com/belikov-av/ANDRIF. The package used to generate data in this article is available as **Additional file 4**.

### SNA driver prediction

Using the file “mc3.v0.2.8.PUBLIC_primary_whitelisted_Entrez.tsv” all SNAs were classified according to the column “Variant_Classification”. “Frame_Shift_Del”, “Frame_Shift_Ins”, “Nonsense_Mutation”, “Nonstop_Mutation” and “Translation_Start_Site” were considered potentially inactivating; “De_novo_Start_InFrame”, “In_Frame_Del”, “In_Frame_Ins” and “Missense_Mutation” were considered potentially hyperactivating; “De_novo_Start_OutOfFrame” and “Silent” were considered passengers; the rest were considered unclear. The classification results were saved as the file “SNA_classification_patients.tsv”, with columns “Tumor_Sample_Barcode”, “Hugo_Symbol”, “Entrez_Gene_Id”, “Gene”, “Number of hyperactivating SNAs”, “Number of inactivating SNAs”, “Number of SNAs with unclear role”, “Number of passenger SNAs”.

Using this file, the sum of all alterations in all patients was calculated for each gene. Genes containing only SNAs with unclear role (likely, noncoding genes) were removed, also from “SNA_classification_patients.tsv”. Next, the Nonsynonymous SNA Enrichment Index (NSEI) was calculated for each gene as

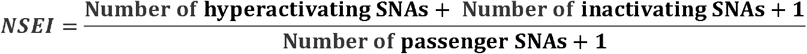

and the Hyperactivating to Inactivating SNA Ratio (HISR) was calculated for each gene as

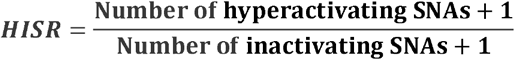

Genes for which the sum of hyperactivating, inactivating and passenger SNAs was less than 10 were removed to ensure sufficient precision of NSEI and HISR calculation, and the results were saved as “SNA_classification_genes_NSEI_HISR.tsv”.

Using the file “SNA_classification_patients.tsv”, the gene-patient matrix “SNA_matrix.tsv” was constructed, encoding the “Number of hyperactivating SNAs”, “Number of inactivating SNAs”, “Number of SNAs with unclear role” and “Number of passenger SNAs” as one number separated by dots (e.g. “2.0.1.1”). If data for a given gene were absent in a given patient, it was encoded as “0.0.0.0”. By drawing statuses randomly with replacement (bootstrapping) from any cell of “SNA_matrix.tsv” 10000 times for each patient, the matrix file “SNA_matrix_bootstrapped.tsv” was created. The sums of statuses in “SNA_matrix_bootstrapped.tsv” were calculated for each iteration separately, and then the corresponding NSEI and HISR indices were calculated and the results were saved as “SNA_bootstrapped_NSEI_HISR.tsv”. Null hypothesis P-values were calculated for each iteration as the number of NSEI values higher than a given iteration’s NSEI value and divided by 10000. The histogram of bootstrapped p-values was plotted to check for the uniformity of null hypothesis p-value distribution.

P-value for each gene was calculated as the number of NSEI values in “SNA_bootstrapped_NSEI_HISR.tsv” higher than its NSEI value in “SNA_classification_genes_NSEI_HISR.tsv” and divided by 10000. The results were saved as “SNA_classification_genes_NSEI_HISR_Pvalues.tsv”. Benjamini–Hochberg procedure with FDR(Q)=5% was applied to P-values in “SNA_classification_genes_NSEI_HISR_Pvalues.tsv”, and genes that pass were saved as “SNA_driver_gene_list_FDR5.tsv”.

We named this algorithm SNADRIF (SNA DRIver Finder) and created a Github repository: https://github.com/belikov-av/SNADRIF. The package used to generate data in this article is available as **Additional file 5**.

### Driver prediction algorithms sources and benchmarking

Lists of driver genes and mutations predicted by various algorithms (**Table 2**) applied to PanCanAtlas data were downloaded from https://gdc.cancer.gov/about-data/publications/pancan-driver (2020plus, CompositeDriver, DriverNet, HotMAPS, OncodriveFML), https://karchinlab.github.io/CHASMplus (CHASMplus), as well as received by personal communication from Francisco Martínez-Jiménez, Institute for Research in Biomedicine, Barcelona, (dNdScv, OncodriveCLUSTL, OncodriveFML). All genes and mutations with q-value > 0.05 were removed. Additionally, a consensus driver gene list from 26 algorithms applied to PanCanAtlas data (7) was downloaded from https://www.cell.com/cell/fulltext/S0092-8674(18)30237-X and a COSMIC Cancer Gene Census (CGC) Tier 1 gene list (14) was downloaded from https://cancer.sanger.ac.uk/cosmic/census?tier=1. Only genes affected by somatic SNAs and CNAs present in the TCGA cancer types were used for further analyses from the CGC list. Cancer type names in the CGC list were manually converted to the closest possible TCGA cancer type abbreviation. Entrez Gene IDs were identified for each gene using HUGO Symbol and external database ftp://ftp.ncbi.nih.gov/gene/DATA/GENE_INFO/Mammalia/Homo_sapiens.gene_info.gz.

**Table 2.**
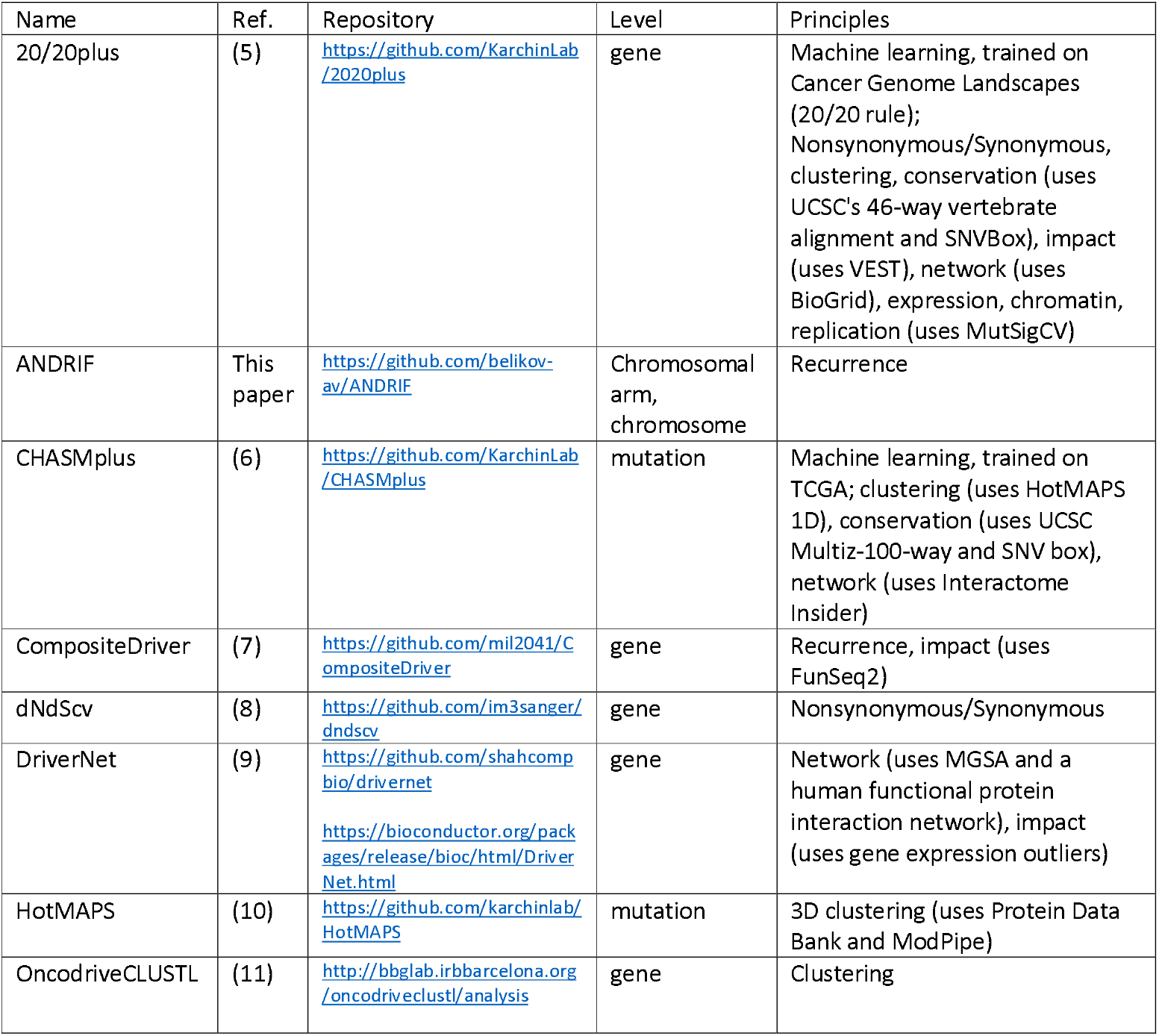
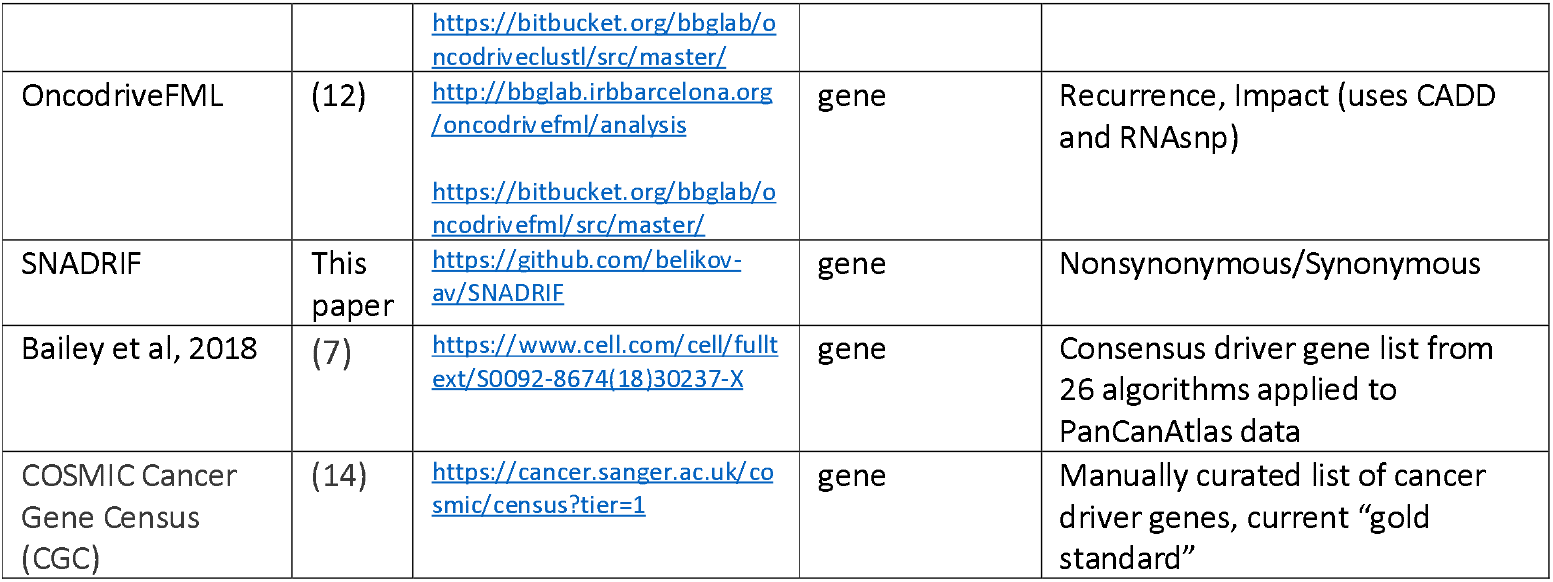
Driver prediction algorithms.

The sensitivity of algorithms was assessed as the percentage of genes in a positive control list that were predicted as drivers by an algorithm, because Sensitivity=True positives/(True positives + False negatives). Three positive control lists were used – CGC Tier 1 genes affected by somatic SNAs or CNAs in TCGA cancer types, a list of genes identified by at least two of all our sources (including CGC and Bailey), and a list of genes identified by at least three of all our sources (including CGC and Bailey). Sensitivity was assessed separately for algorithms applied to individual cancer types as the percentage of gene-cohort pairs in a positive control list that were matched by gene-cohort pairs predicted by an algorithm.

### Conversion of population-level data to patient-level data

For lists of driver genes, all entries from the file “mc3.v0.2.8.PUBLIC_primary_whitelisted_Entrez.tsv” were removed except those that satisfied the following conditions simultaneously: “Entrez Gene ID” matches the one in the driver list; cancer type (identified by matching “Tumor_Sample_Barcode” with “bcr_patient_barcode” and “acronym” in “clinical_PANCAN_patient_with_followup.tsv”) matches “cohort” in the driver list or the driver list is for pancancer analysis; “Variant_Classification” column contains one of the following values: “De_novo_Start_InFrame”, “Frame_Shift_Del”, “Frame_Shift_Ins”, “In_Frame_Del”, “In_Frame_Ins”, “Missense_Mutation”, “Nonsense_Mutation”, “Nonstop_Mutation”, “Translation_Start_Site”.

For lists of driver mutations, the procedures were the same, except that Ensembl Transcript ID and nucleotide/amino acid substitution were used for matching instead of Entrez Gene ID. These data (only columns “TCGA Barcode”, “HUGO Symbol”, “Entrez Gene ID”) were saved as “AlgorithmName_output_SNA.tsv”.

Additionally, all entries from the file

“ISAR_GISTIC.all_thresholded.by_genes_primary_whitelisted.tsv” were removed except those that satisfied the following conditions simultaneously: “Locus ID” matches “Entrez Gene ID” in the driver list; cancer type (identified by matching Tumor Sample Barcode with “bcr_patient_barcode” and “acronym” in “clinical_PANCAN_patient_with_followup.tsv”) matches “cohort” in the driver list or the driver list is for pancancer analysis; CNA values are “2”, “1”, “-1” or “-2”. These data were converted from the matrix to a list format (with columns “TCGA Barcode”, “HUGO Symbol”, “Entrez Gene ID”) and saved as “AlgorithmName_output_CNA.tsv”.

Finally, the files “AlgorithmName_output_SNA.tsv” and “AlgorithmName_output_CNA.tsv” were combined, duplicate TCGA Barcode-Entrez Gene ID pairs were removed, and the results saved as “AlgorithmName_output.tsv”.

### Driver event classification and analysis

The file “Clinical with Follow-up - clinical_PANCAN_patient_with_followup.tsv” was downloaded from https://gdc.cancer.gov/node/905/. All patients with “icd_o_3_histology” different from XXXX/3 (primary malignant neoplasm) were removed, as well as all patients not simultaneously present in the following three files: “mc3.v0.2.8.PUBLIC_primary_whitelisted_Entrez.tsv”, “ISAR_GISTIC.all_thresholded.by_genes_primary_whitelisted.tsv” and “Primary_whitelisted_arms.tsv”. The resulting file was saved as “clinical_PANCAN_patient_with_followup_primary_whitelisted.tsv”.

Several chosen “AlgorithmName_output.tsv” files were combined and all TCGA Barcode-Entrez Gene ID pairs not present in at least two output files were removed. Entries with TCGA Barcodes not present in “clinical_PANCAN_patient_with_followup_primary_whitelisted.tsv” were removed as well. Matching “Number of hyperactivating SNAs” and “Number of inactivating SNAs” for each TCGA Barcode-Entrez Gene ID pair were taken from the “SNA_classification_patients.tsv” file, in case of no match zeros were written. Matching HISR value was taken from “SNA_classification_genes_NSEI_HISR.tsv” for each Entrez Gene ID, in case of no match empty cell was left. Matching CNA status was taken from “ISAR_GISTIC.all_thresholded.by_genes_primary_whitelisted_RNAfiltered.tsv” for each TCGA Barcode-Entrez Gene ID pair, in case of no match zero was written.

Each TCGA Barcode-Entrez Gene ID pair was classified according to the **Table 3**:

**Table 3.** Driver event classification rules.

| Driver type | Number of nonsynonymous SNAs | Number of inactivating SNAs | HISR | CNA status | Count as ... driver event(s) |
| --- | --- | --- | --- | --- | --- |
| SNA-based oncogene | ≥1 | 0 | >5 | 0 | 1 |
| CNA-based oncogene | 0 | 0 | >5 | 1 or 2 | 1 |
| Mixed oncogene | ≥1 | 0 | >5 | 1 or 2 | 1 |
| **SNA-based tumor suppressor** | $\geq 1$ | $\geq 0$ | $\leq 5$ | 0 | 1 |
| **CNA-based tumor suppressor** | 0 | 0 | $\leq 5$ | -1 or -2 | 1 |
| **Mixed tumor suppressor** | $\geq 1$ | $\geq 0$ | $\leq 5$ | -1 or -2 | 1 |
| **Passenger** | 0 | 0 |  | 0 | 0 |
| **Low-probability driver** | All the rest |  |  |  | 0 |

Results of this classification were saved as “AnalysisName_genes_level2.tsv”.

Using this file, the number of driver events of each type was counted for each patient. Information on the number of driver chromosome and arm losses and gains for each patient was taken from the files “Chromosome_drivers_FDR5.tsv” and “Arm_drivers_FDR5.tsv”. All patients not present in the files “AnalysisName_genes_level2.tsv”, “Chromosome_drivers_FDR5.tsv” and “Arm_drivers_FDR5.tsv”, but present in the file “clinical_PANCAN_patient_with_followup_primary_whitelisted.tsv”, were added with zero values for the numbers of driver events. Information on the cancer type (“acronym”), gender (“gender”), age (“age_at_initial_pathologic_diagnosis”) and tumor stage (“pathologic_stage”, if no data then “clinical_stage”, if no data then “pathologic_T”, if no data then “clinical_T”) was taken from the file “clinical_PANCAN_patient_with_followup_primary_whitelisted.tsv”. The results were saved as “AnalysisName_patients.tsv”.

Using the file “AnalysisName_patients.tsv”, the number of patients with each integer total number of driver events from 0 to 100 was counted for each cancer type, also for males and females separately, and cumulative histograms were plotted. Using the same file “AnalysisName_patients.tsv”, the average number of various types of driver events was calculated for each cancer type, tumor stage, age group, as well as for patients with each total number of driver events from 1 to 100. Analyses were performed for total population and for males and females separately, and cumulative histograms were plotted for each file.

We named this algorithm PALDRIC (PAtient-Level DRIver Classifier) and created a Github repository: https://github.com/belikov-av/PALDRIC

We later developed a modification of PALDRIC that allows analysis and ranking of individual genes, chromosome arms and full chromosomes – PALDRIC GENE - and created a Github repository: https://github.com/belikov-av/PALDRIC_GENE. The package used to generate data in this article is available as **Additional file 6**.

Using the files “AnalysisName_genes_level2.tsv”, “Chromosome_drivers_FDR5.tsv” and “Arm_drivers_FDR5.tsv”, the names of individual genes, chromosome arms or full chromosomes affected by driver events of each type were catalogued for each patient. Information on the cancer type, gender, age and tumor stage was taken from the file “clinical_PANCAN_patient_with_followup_primary_whitelisted.tsv”. The results were saved as “AnalysisName_patients_genes.tsv”.

Using the file “AnalysisName_patients_genes.tsv”, the number of various types of driver events in individual genes, chromosome arms or full chromosomes was calculated for each cancer type, tumor stage, age group, as well as for patients with each total number of driver events from 1 to 100. Analyses were performed for total population and for males and females separately, and histograms of top 10 driver events in each class and overall were plotted for each group.

Driver Strength Index (DSI)

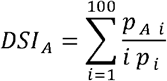

and Normalized Driver Strength Index (NDSI)

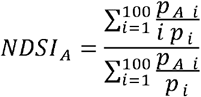

were calculated, where *p*_*A i*_ is a number of patients with a driver event in the gene/chromosome A amongst patients with i driver events in total; *p*_*i*_ is a number of patients with *i* driver events in total. To avoid contamination of NDSI-ranked driver event lists with very rare driver events and to increase precision of the index calculation, all events that were present in less than 10 patients in each driver event class were removed. To compose the top-(N)DSI-ranked driver list, the lists of drivers from various classes were combined, and drivers with lower (N)DSI in case of duplicates and all drivers with NDSI<0.05 were removed.

### Pathway and network analysis of top-(N)DSI-ranked driver genes

First, the chromosome arms and full chromosomes were removed from the top-(N)DSI-ranked driver lists, as external pathway and network analysis services can work only with genes. Next, top 50 DSI-ranked genes and top 50 NDSI-ranked genes were selected, to facilitate proper comparison.

The resulting lists were uploaded as Entrez Gene IDs to “Reactome v77 Analyse gene list” tool (https://reactome.org/PathwayBrowser/#TOOL=AT). Voronoi visualizations (Reacfoam) were exported as jpg files.

The resulting lists were also uploaded as Entrez Gene IDs to “KEGG Mapper –Color” tool (https://www.genome.jp/kegg/mapper/color.html), “hsa” Search mode was selected, default bgcolor assigned to “yellow”, search executed and the top result - “Pathways in cancer - Homo sapiens (human)” (hsa05200) was selected for mapping. The resulting images were exported as png files.

The data were also analyzed in Cytoscape 3.8.2 (https://cytoscape.org). BioGRID: Protein-Protein Interactions (H. sapiens) network was imported and then (N)DSI values appended from the top 50 (N)DSI-ranked driver list. First, Degree Sorted Circle Layout was selected and genes not within the circle were removed. Node Fill Color was mapped to (N)DSI values with Continuous Mapping and Node Height and Width were mapped to degree.layout parameter (number of connections) with Continuous Mapping. Then, yFiles Organic Layout was selected and legend appended. The resulting images were exported as pdf files.

## Data Availability

All data are available as supplementary files

## Acknowledgements

AVB acknowledges MIPT 5-100 program support for early career researchers.

