## Supplementary figures and images for "Novel Driver Strength Index highlights important cancer genes in TCGA PanCanAtlas patients"

### 2021_8_16_14_9_BLCA_FEMALE.pdf

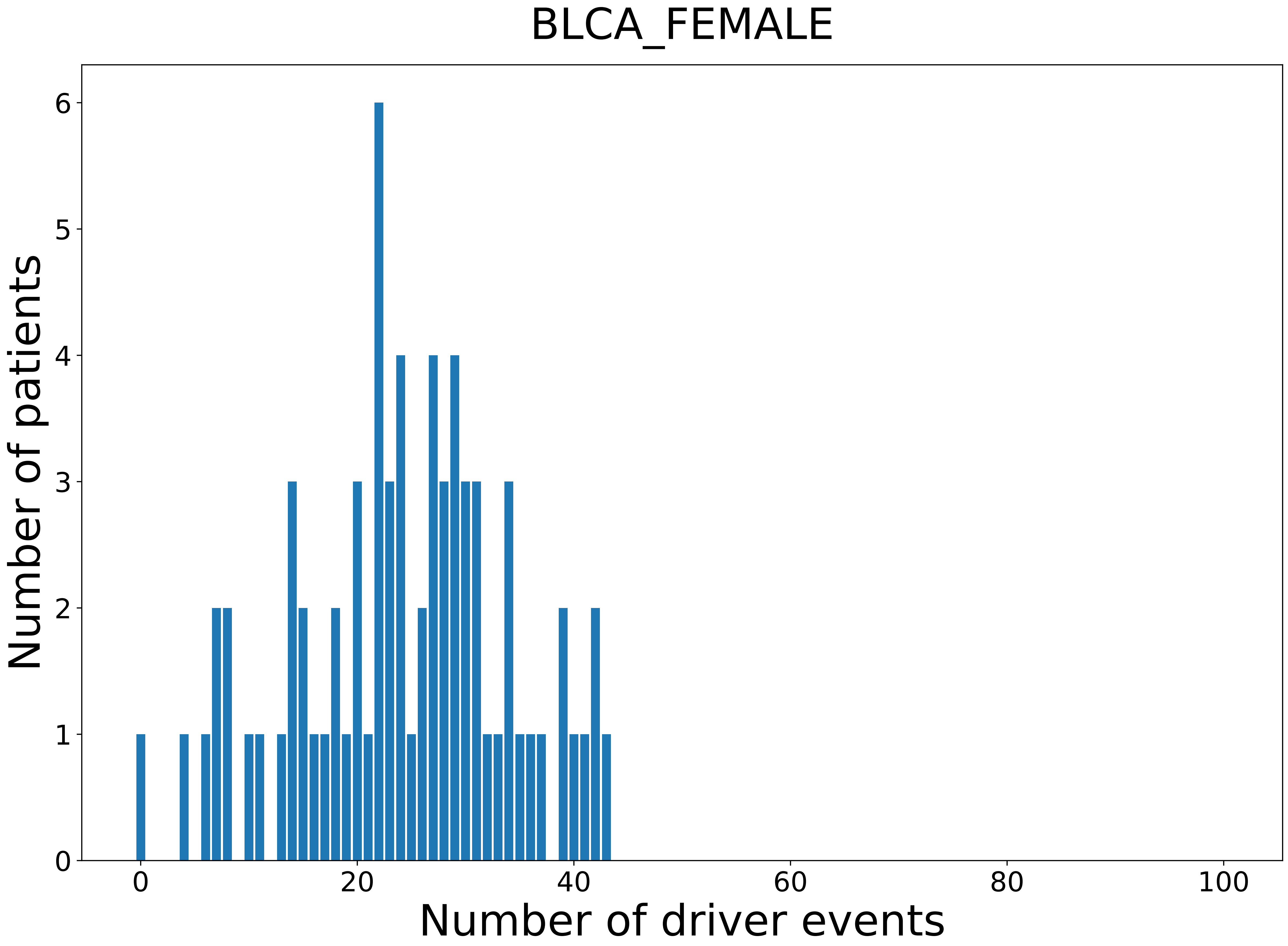

### 2021_8_16_14_9_BRCA.pdf

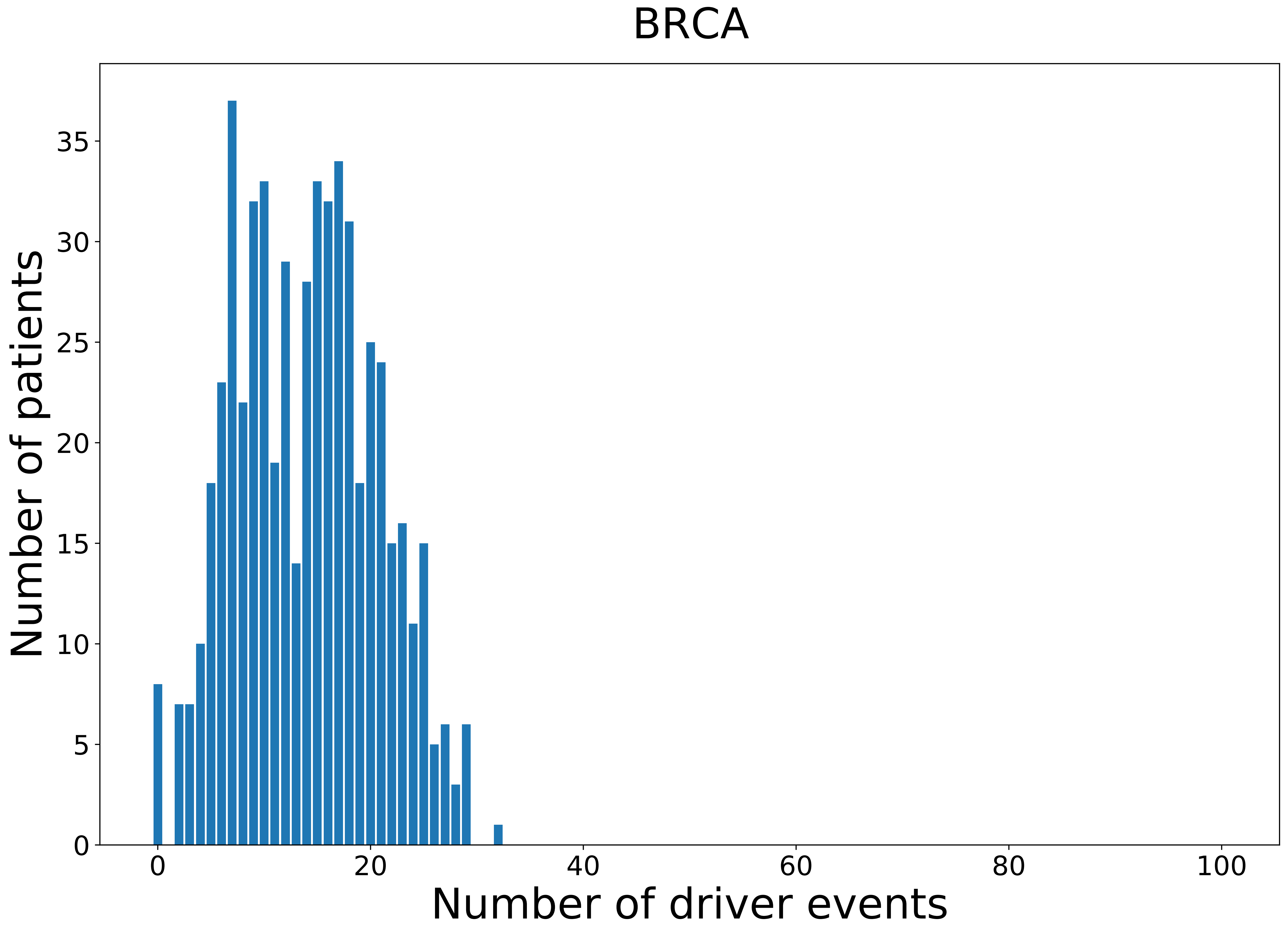

### 2021_8_16_14_9_BRCA_FEMALE.pdf

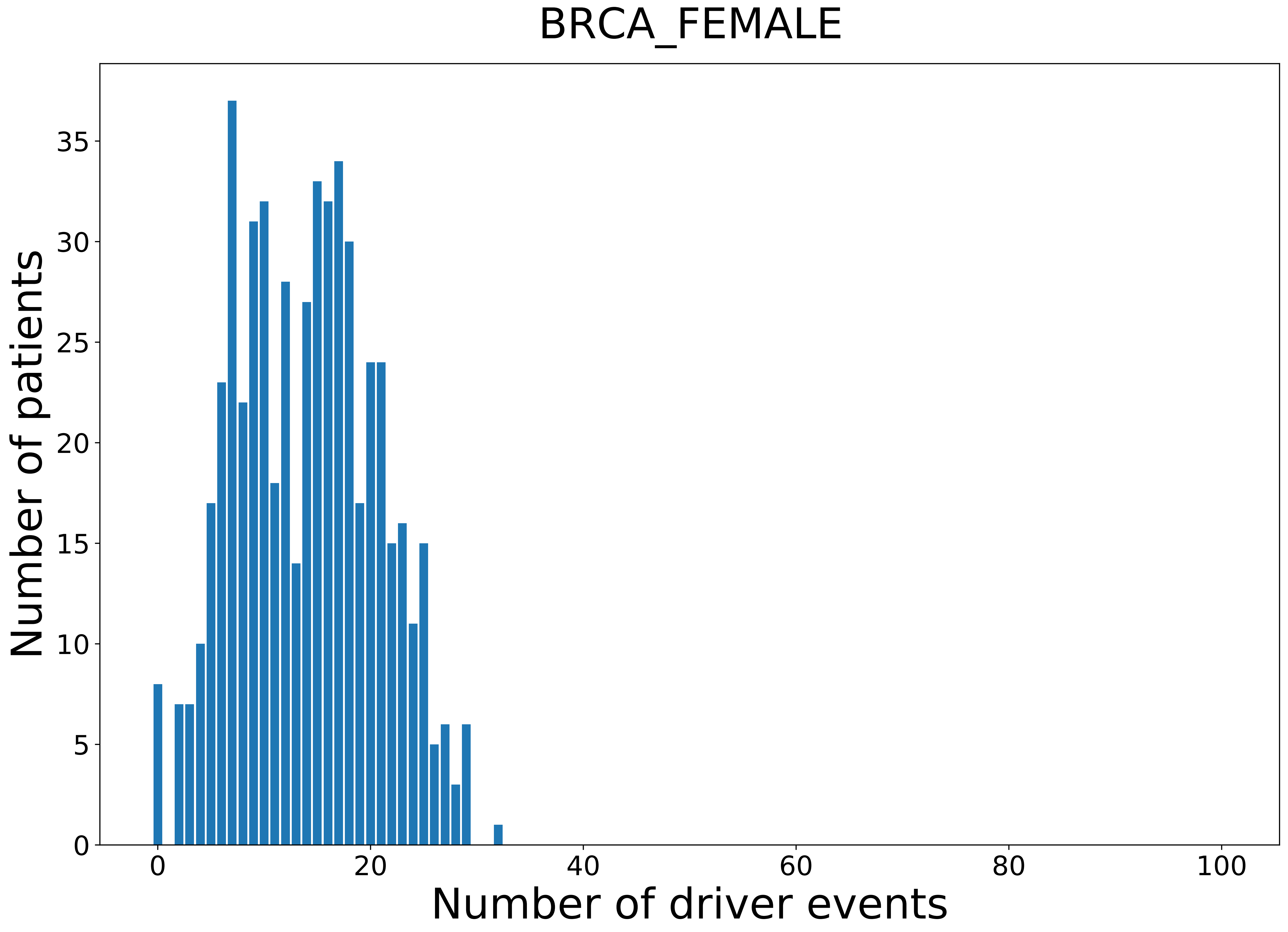

### 2021_8_16_14_9_CHOL.pdf

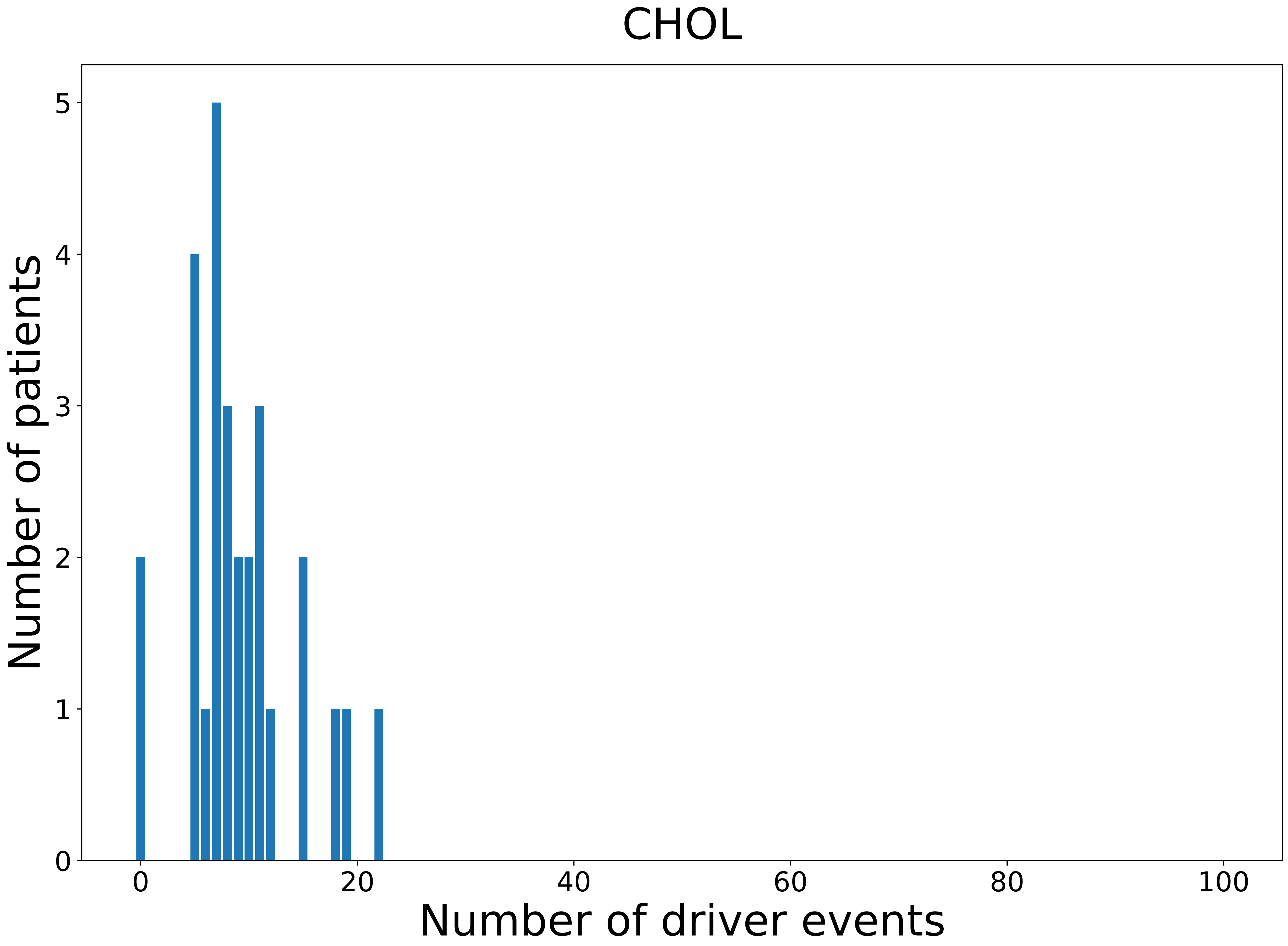

### 2021_8_16_14_9_CHOL_FEMALE.pdf

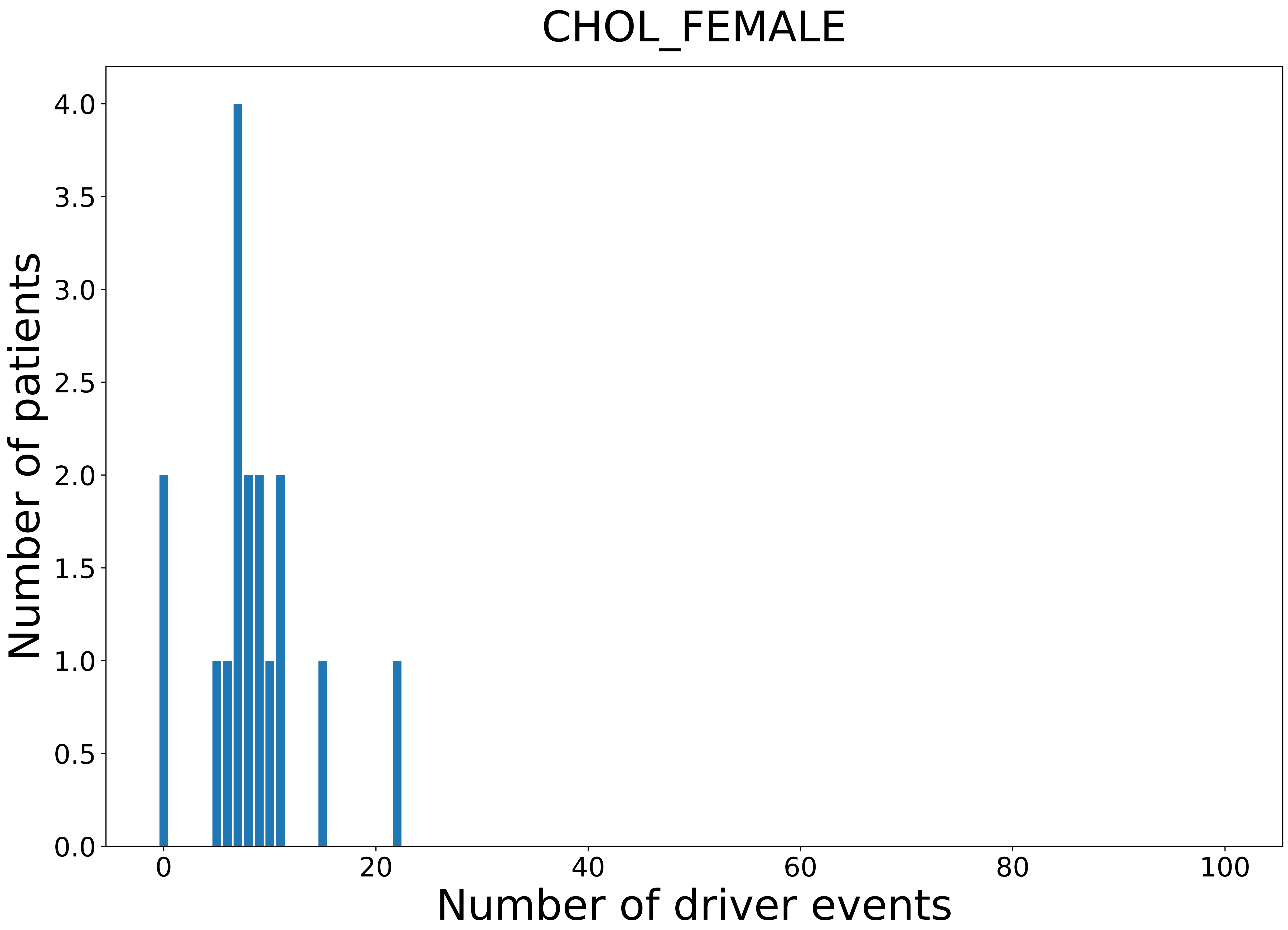

### 2021_8_16_14_9_COAD.pdf

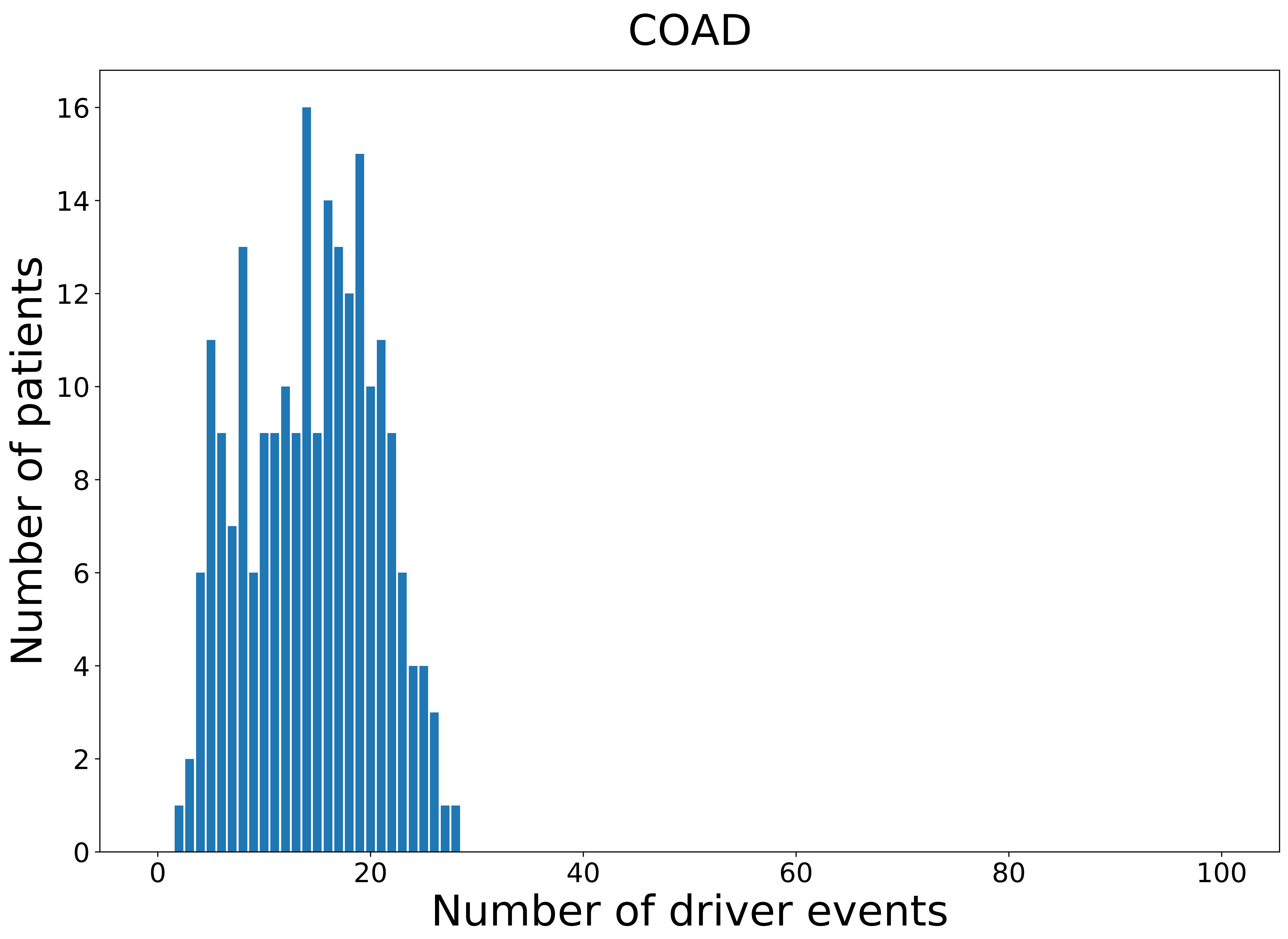

### 2021_8_16_14_9_distribution_age.pdf

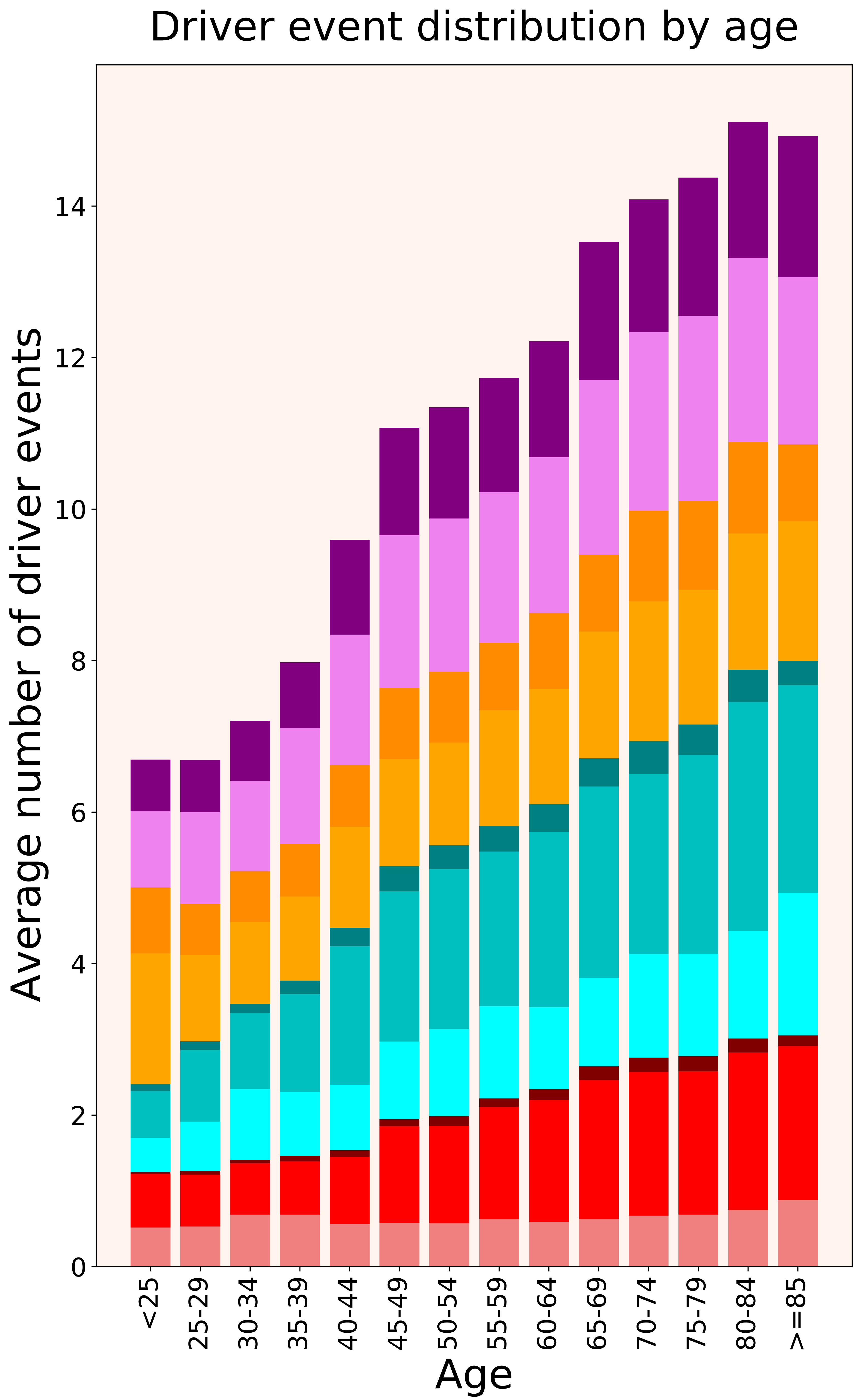

### 2021_8_16_14_9_distribution_age_females.pdf

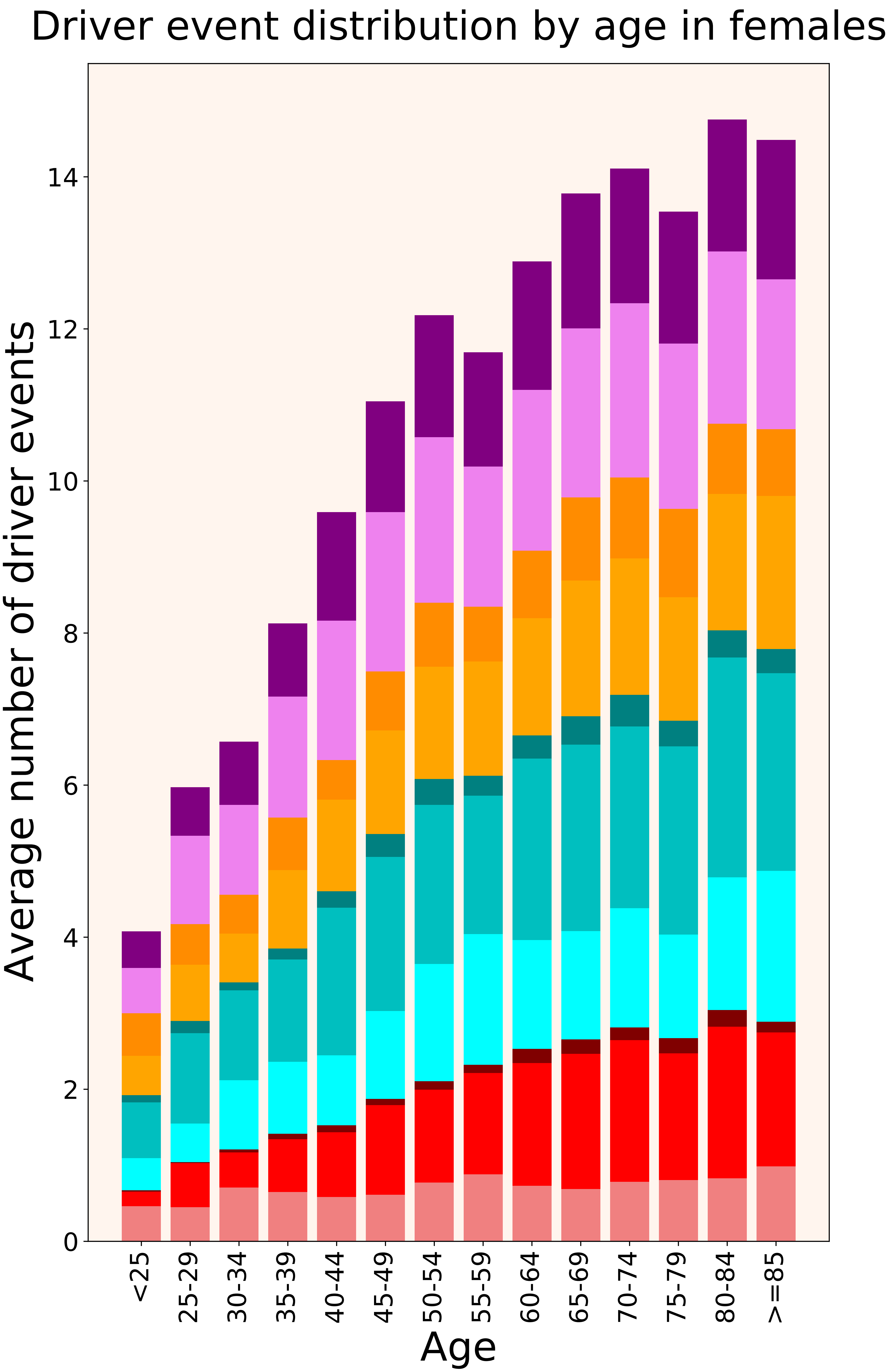

### 2021_8_16_14_9_distribution_age_males.pdf

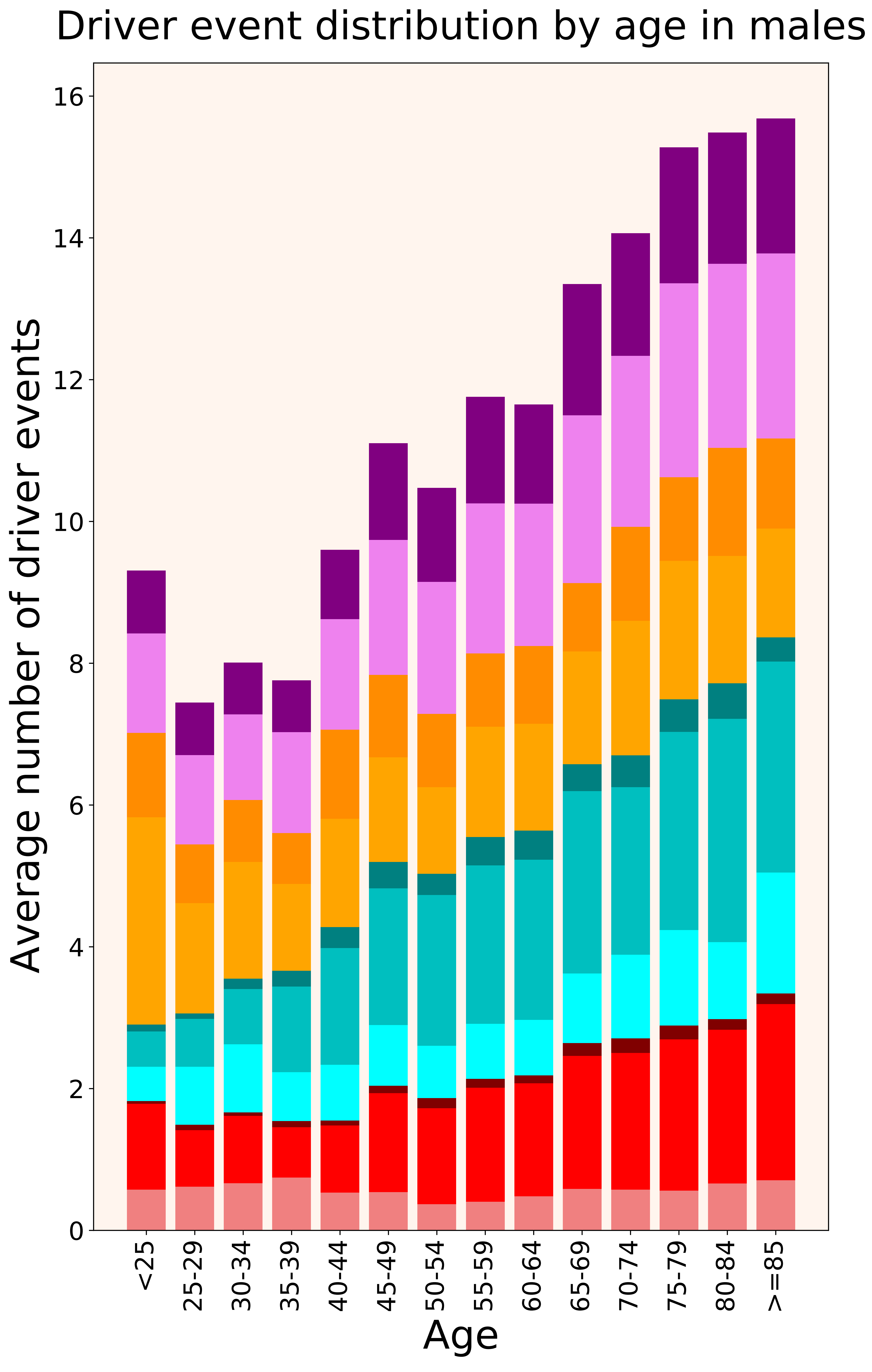

### 2021_8_16_14_9_distribution_cohorts.pdf

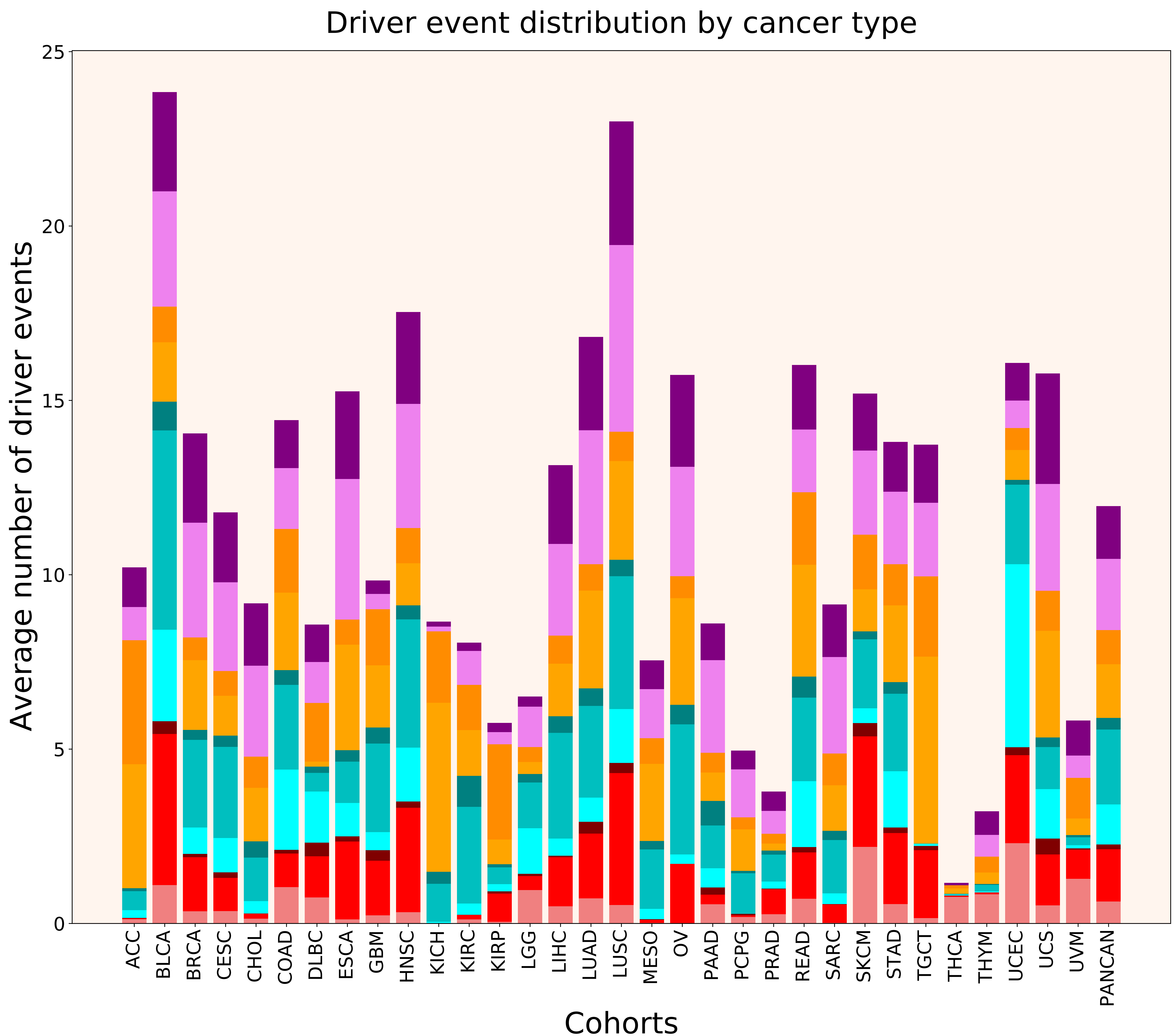

### 2021_8_16_14_9_distribution_cohorts_females.pdf

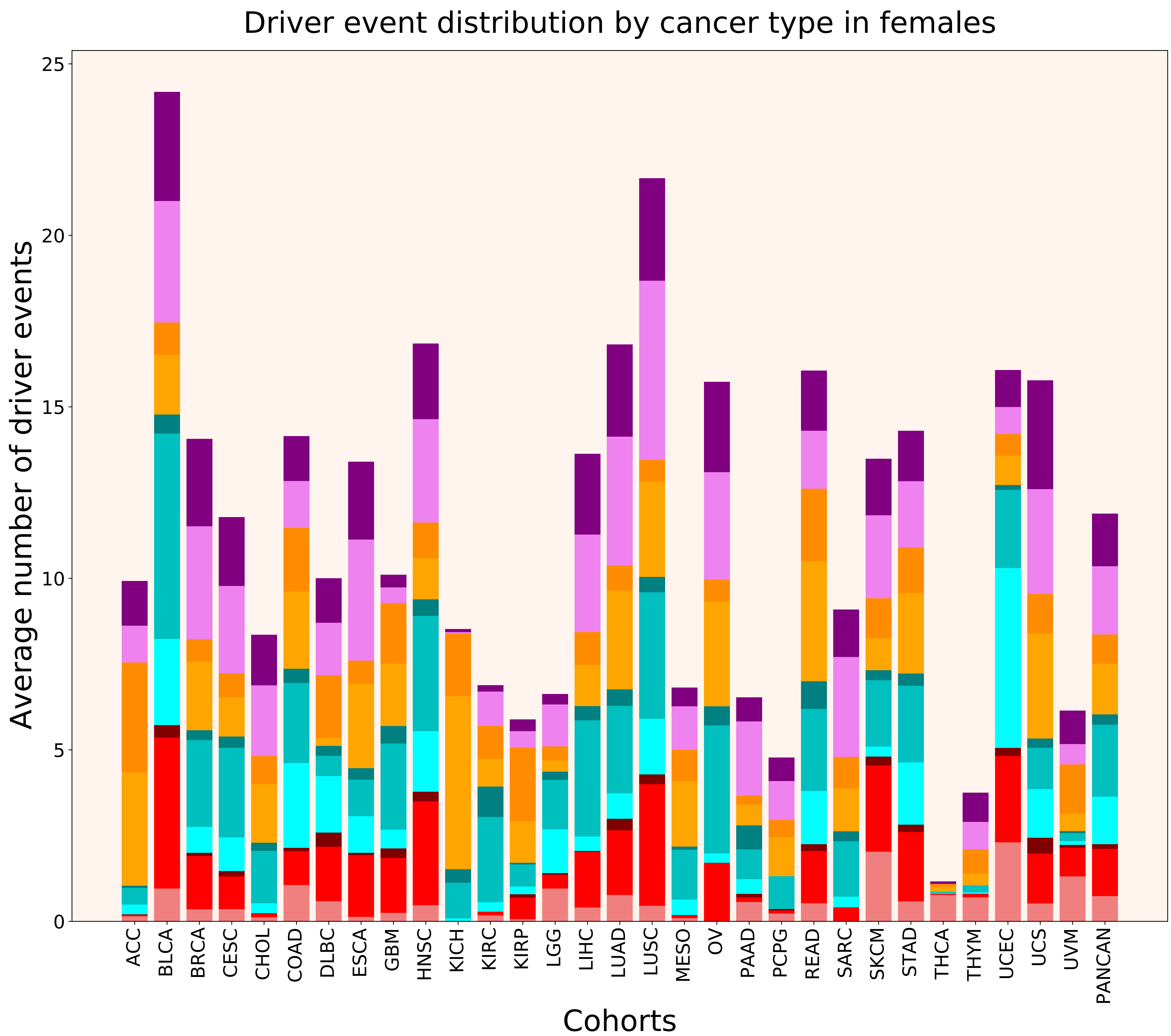

### 2021_8_16_14_9_distribution_cohorts_males.pdf

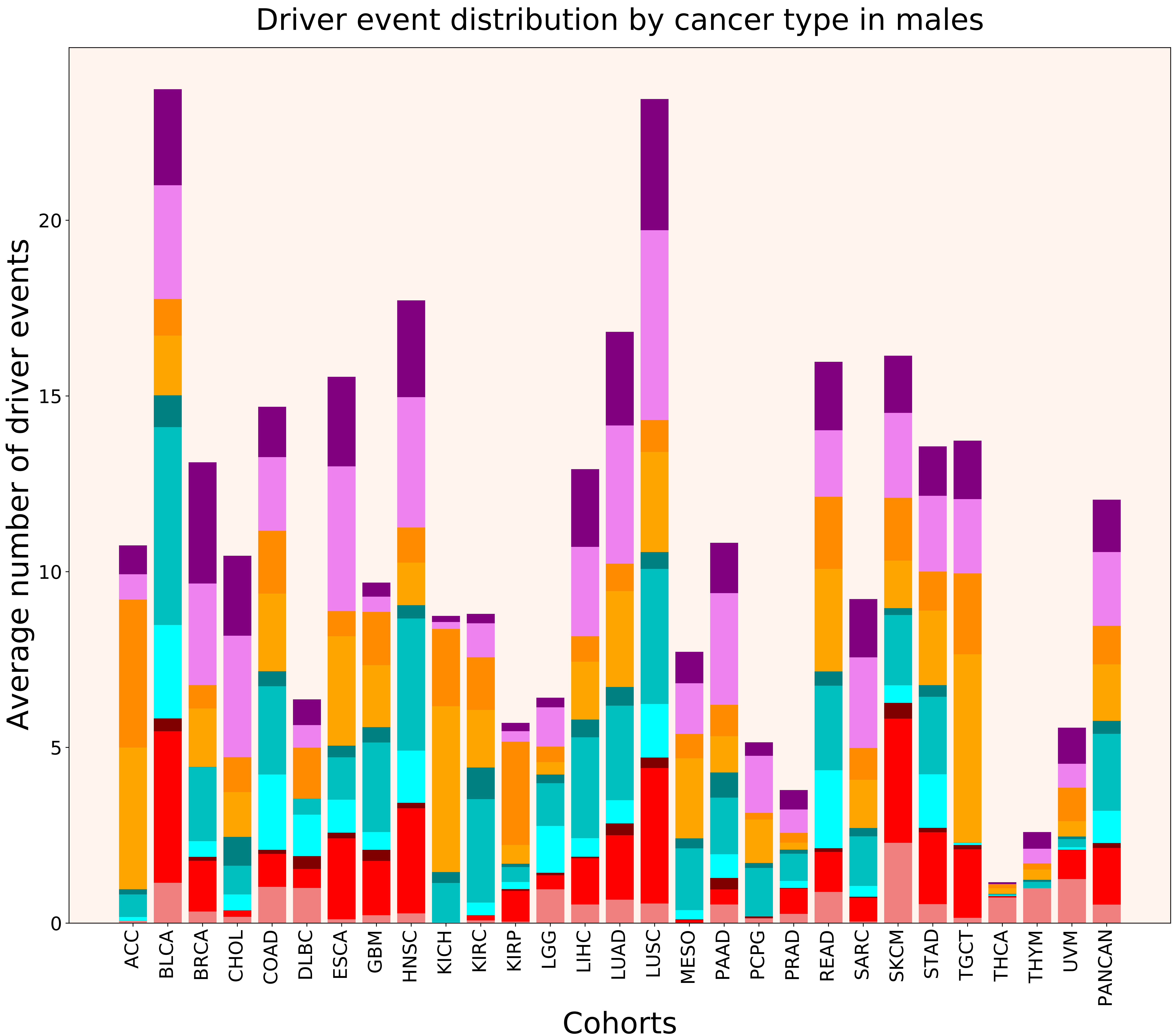

### 2021_8_16_14_9_distribution_events_detailed.pdf

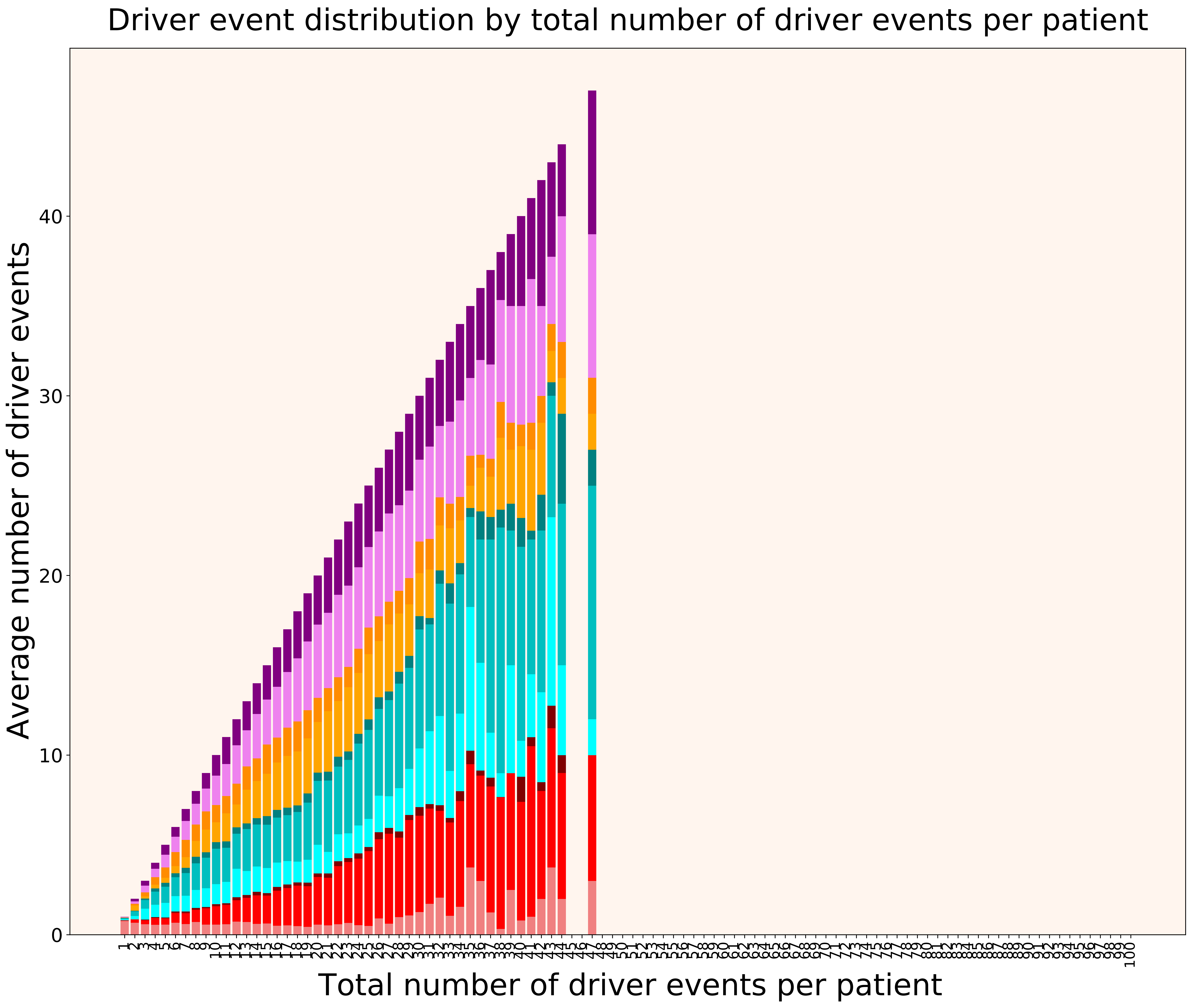

### 2021_8_16_14_9_distribution_events_detailed_females.pdf

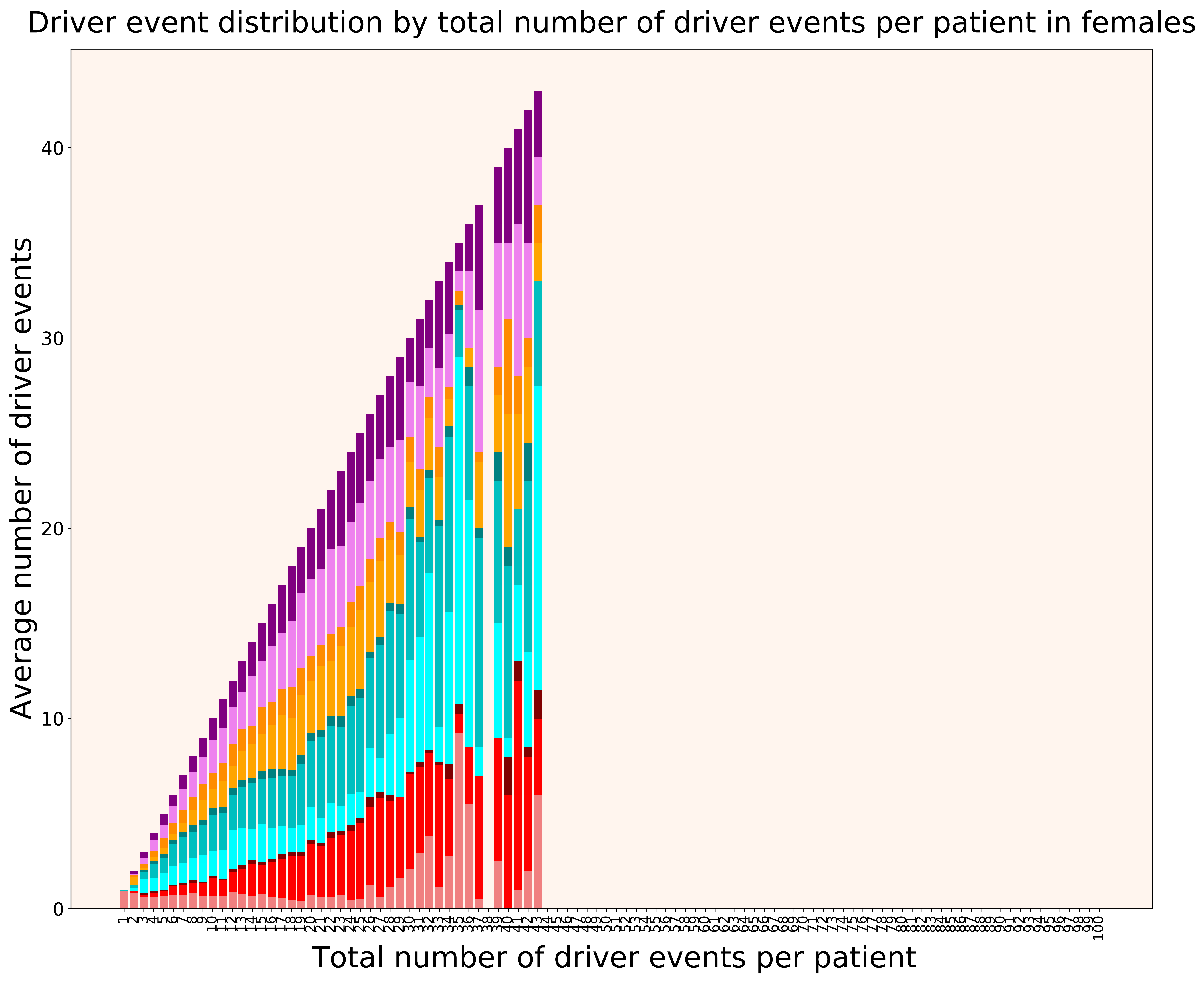

### 2021_8_16_14_9_distribution_events_detailed_males.pdf

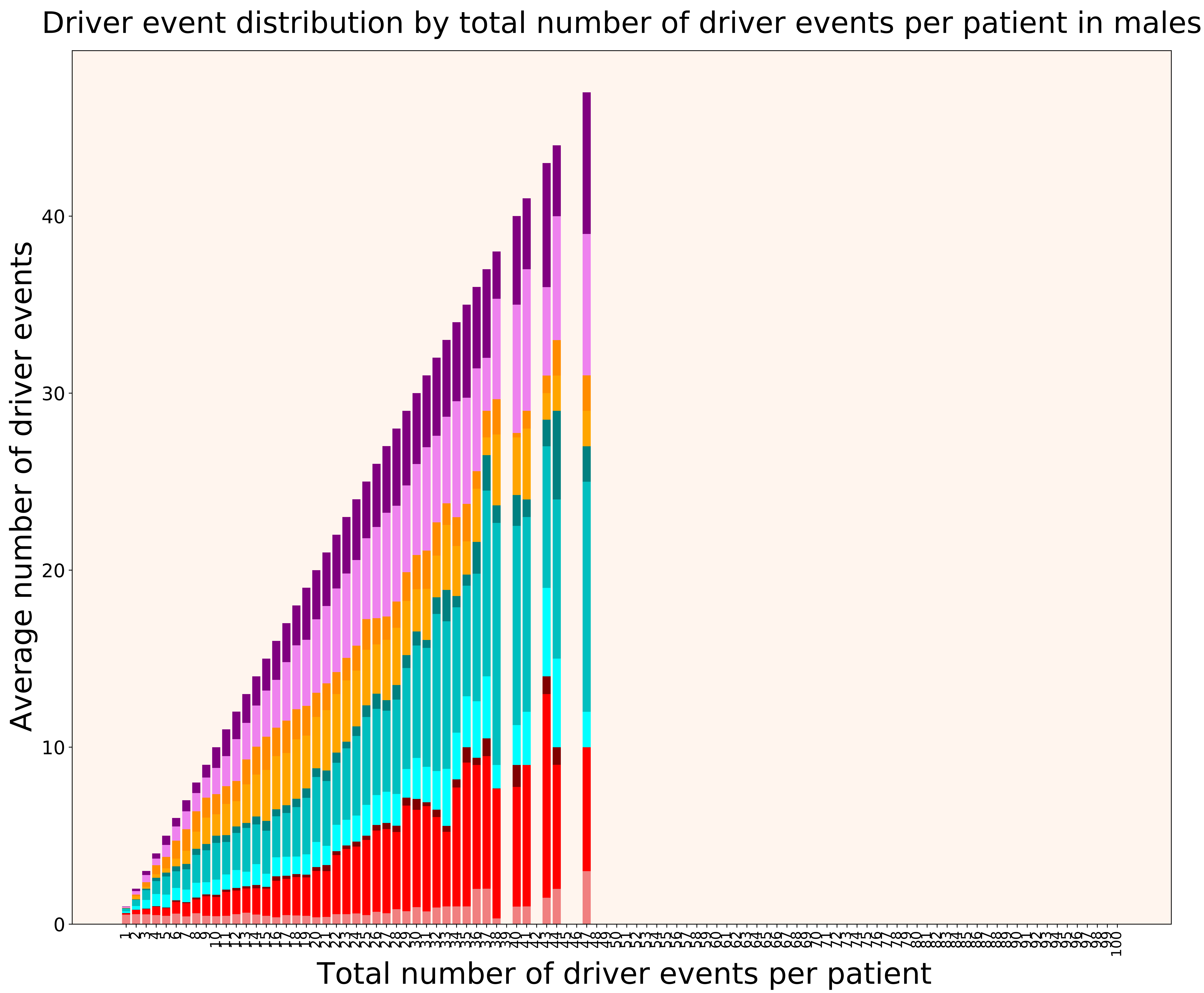

### 2021_8_16_14_9_distribution_gender.pdf

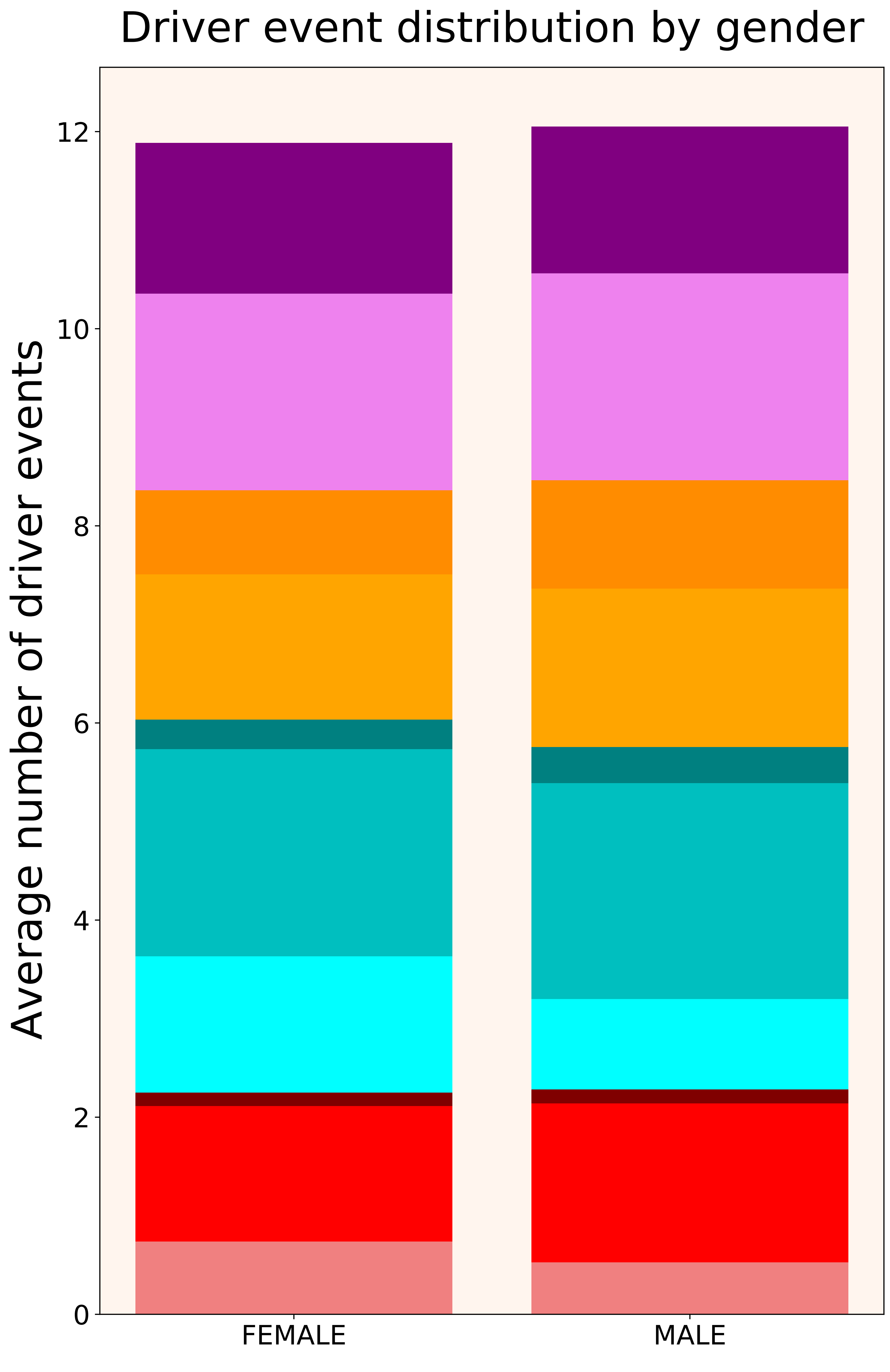

### 2021_8_16_14_9_distribution_stages.pdf

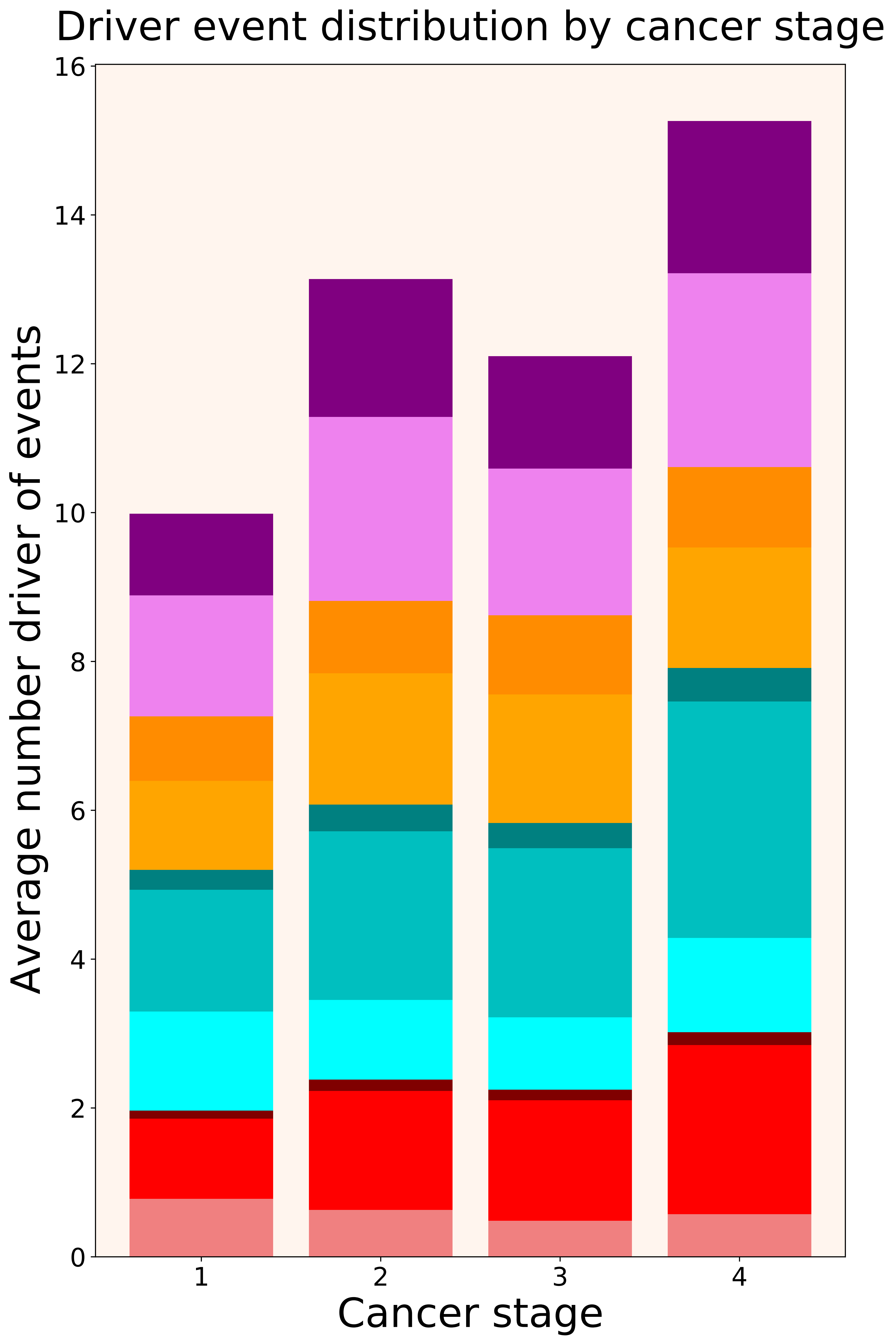

### 2021_8_16_14_9_distribution_stages_females.pdf

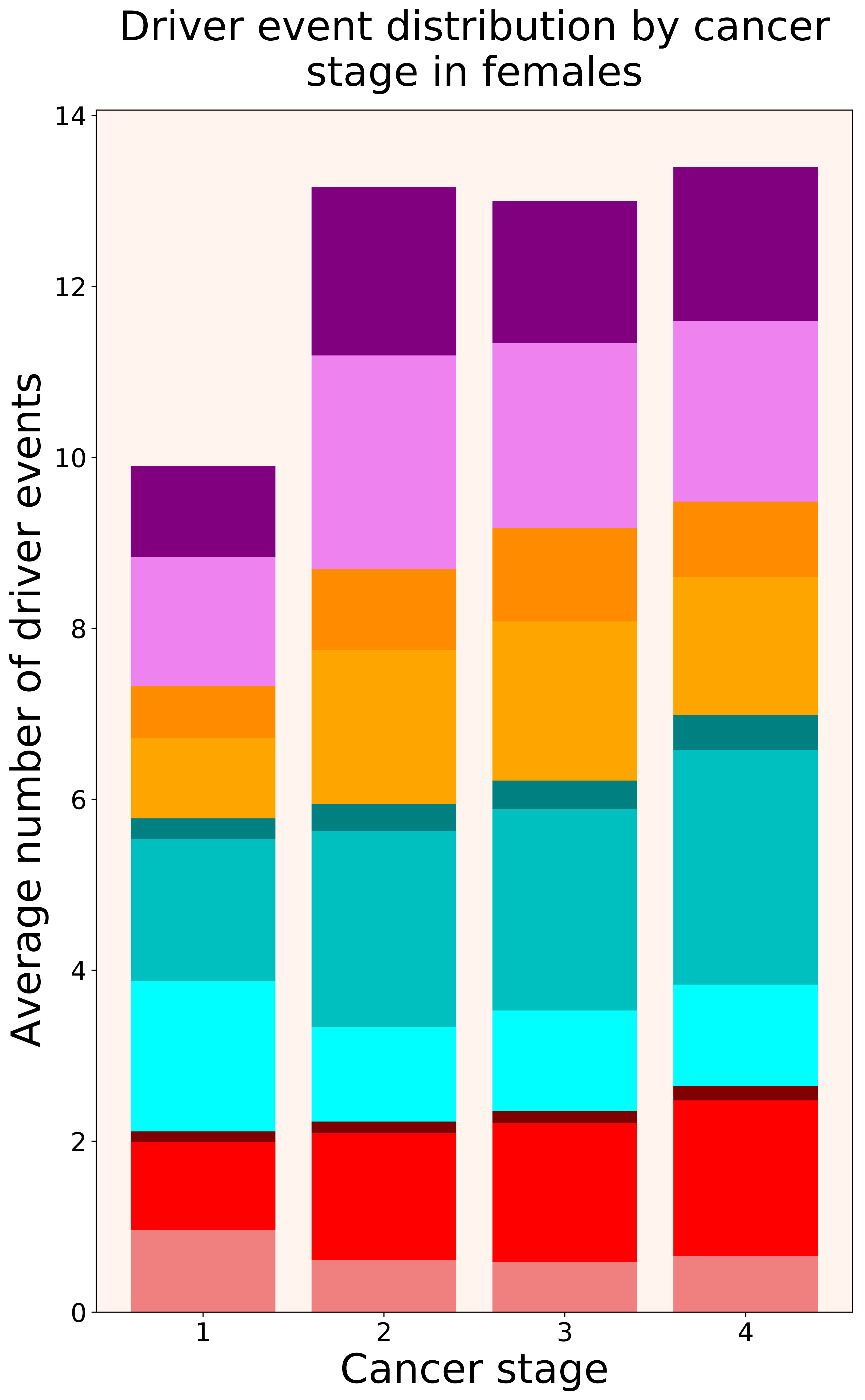

### 2021_8_16_14_9_distribution_stages_males.pdf

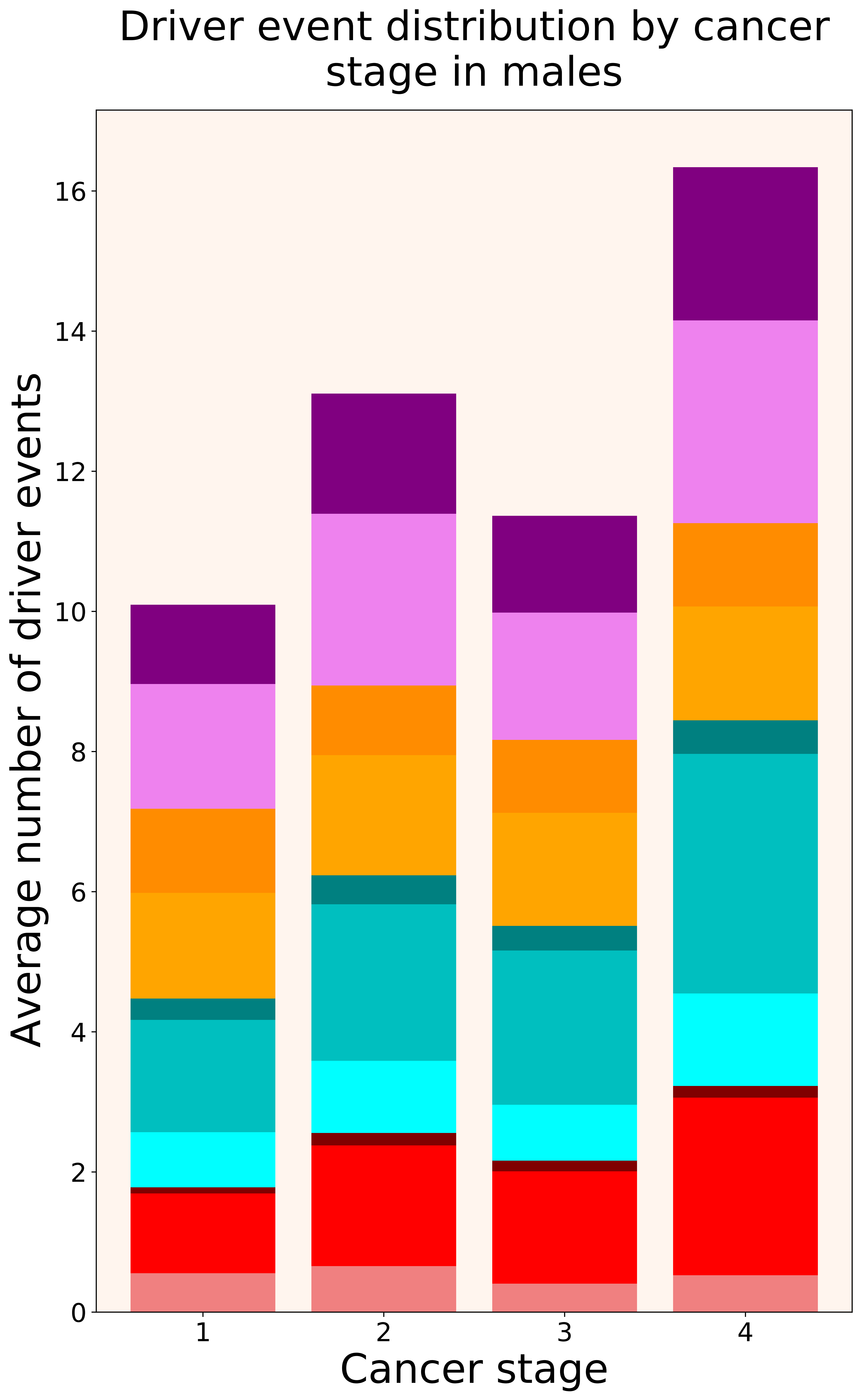

### 2021_8_16_14_9_DLBC.pdf

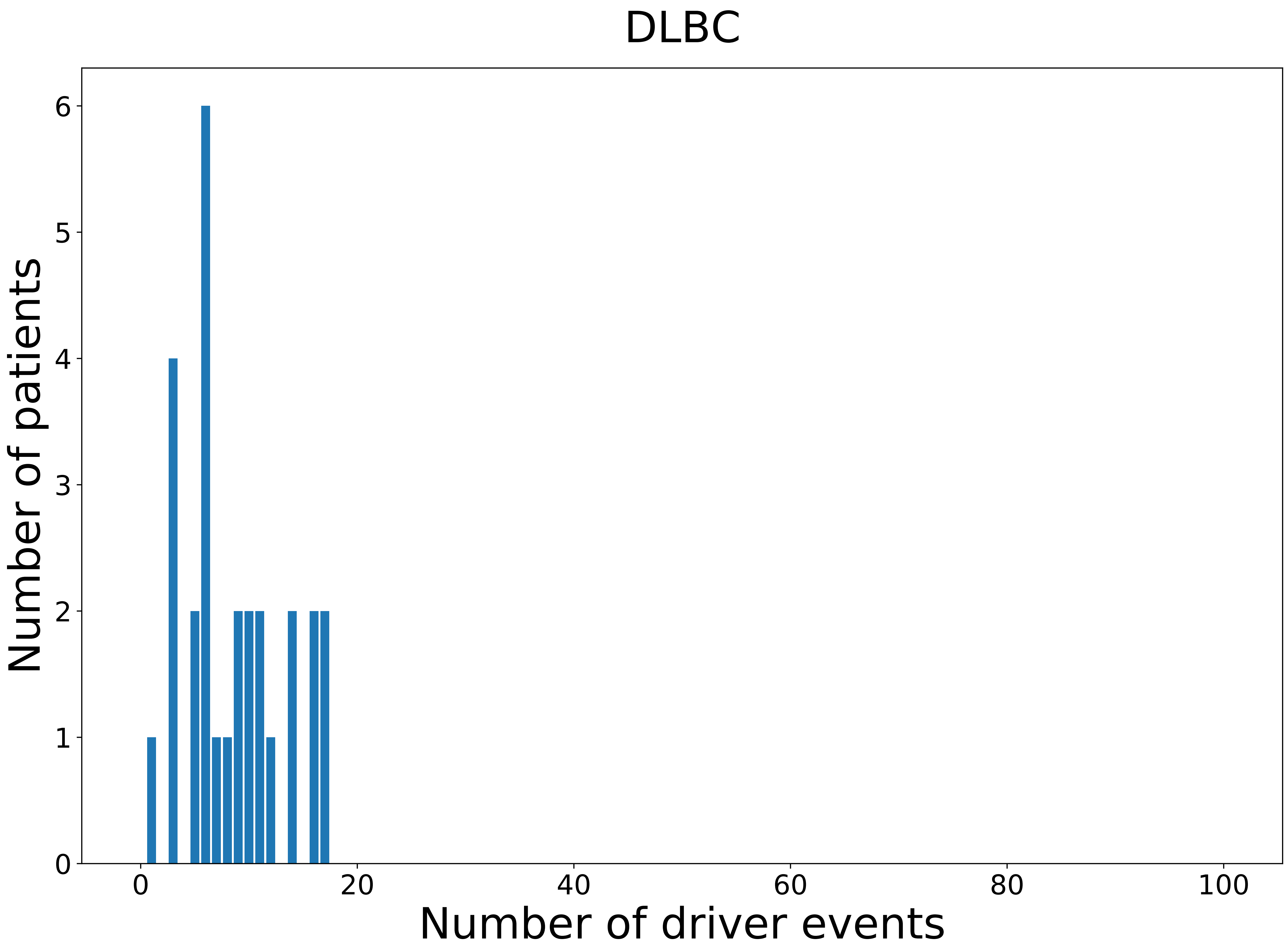

### 2021_8_16_14_9_DLBC_FEMALE.pdf

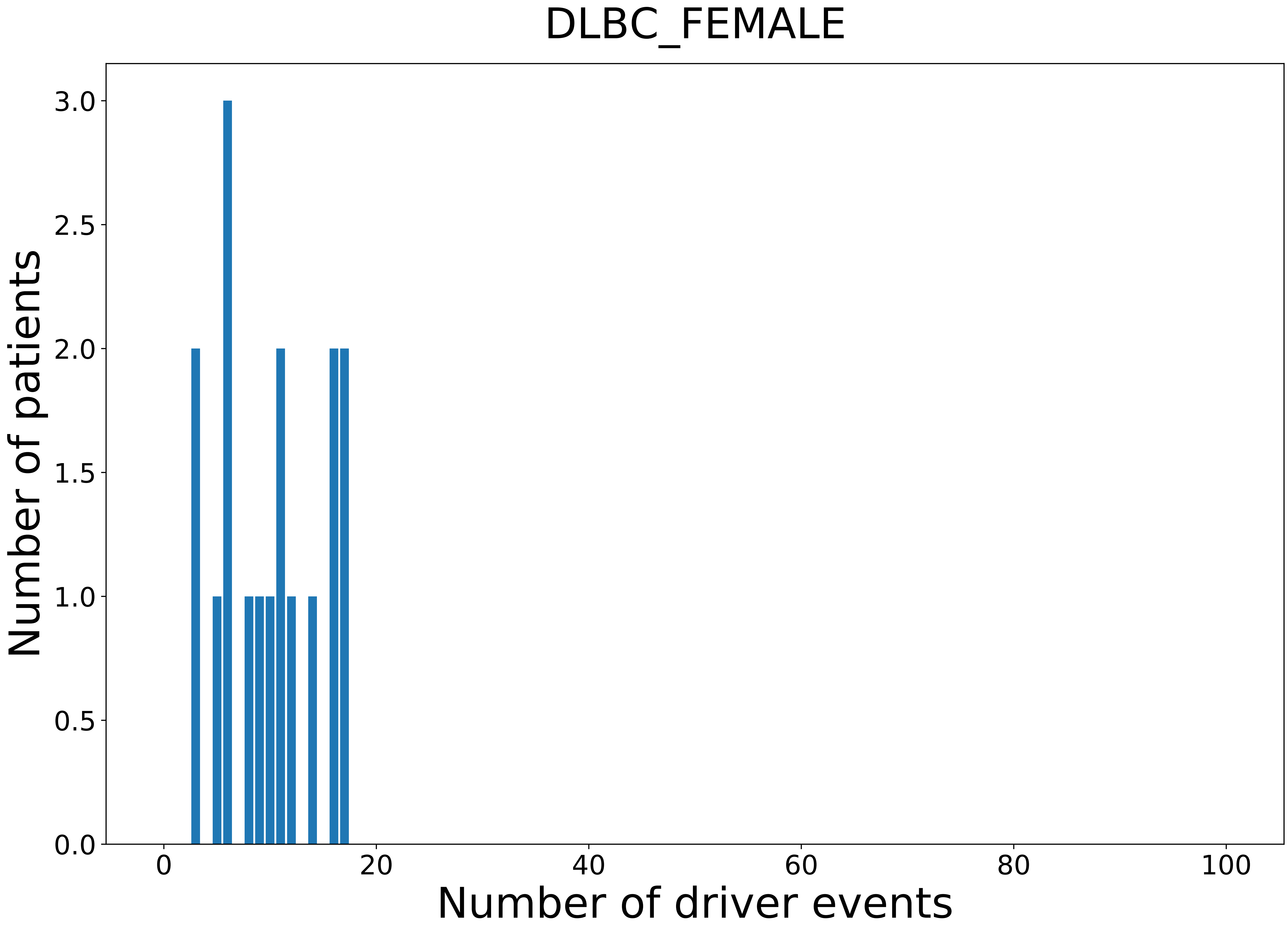

### 2021_8_16_14_9_DLBC_MALE.pdf

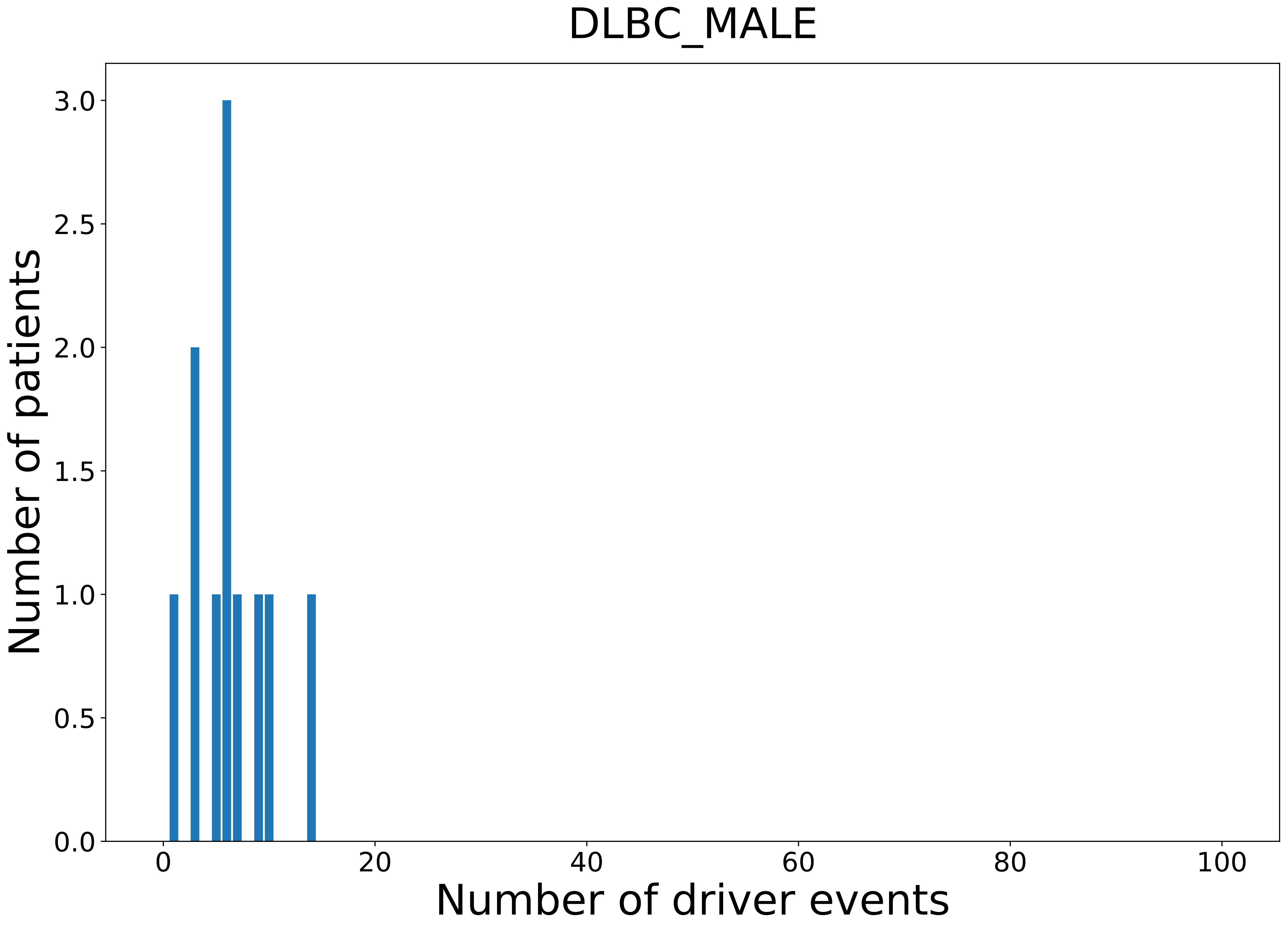

### 2021_8_16_14_9_ESCA_FEMALE.pdf

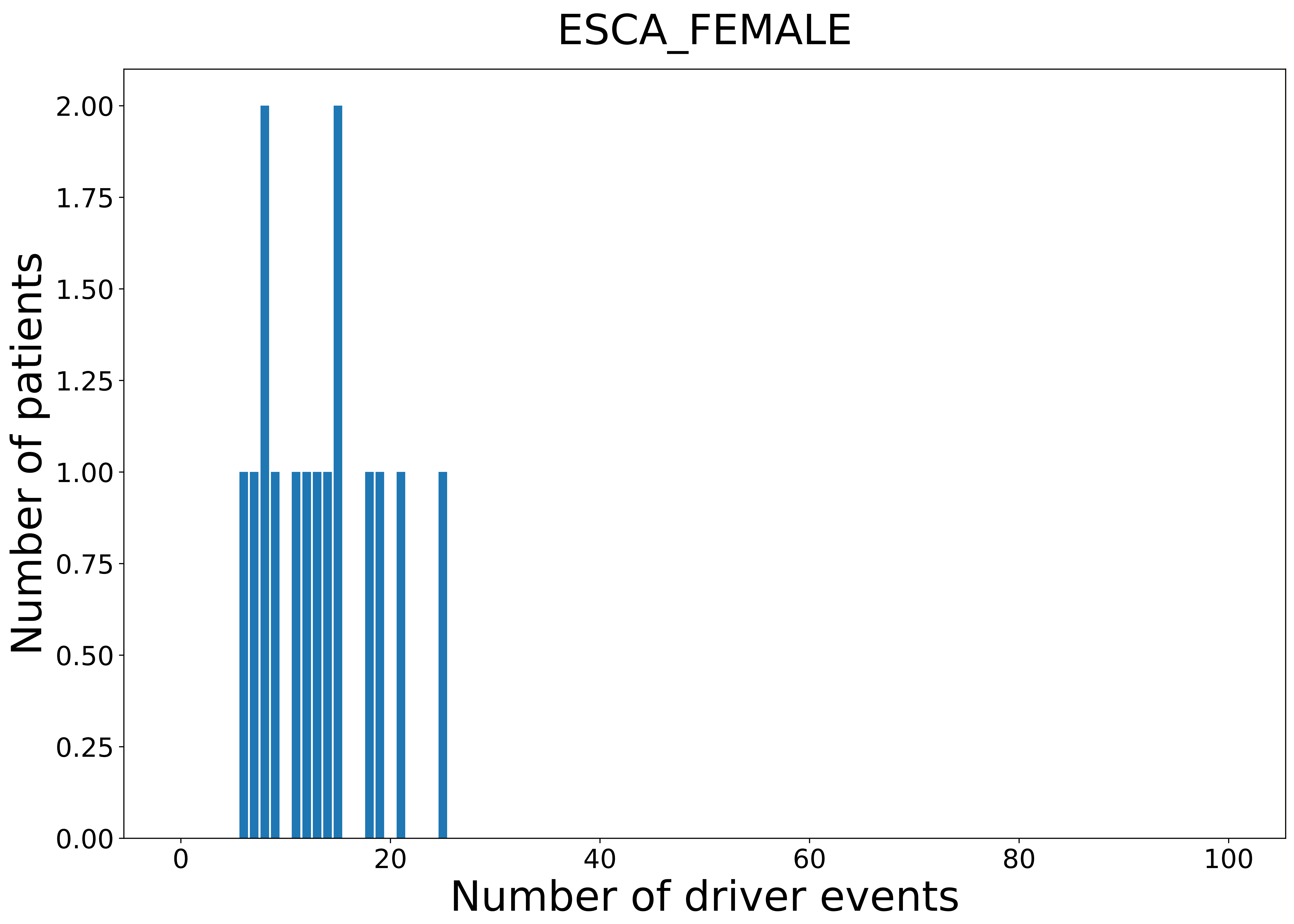

### 2021_8_16_14_9_GBM_MALE.pdf

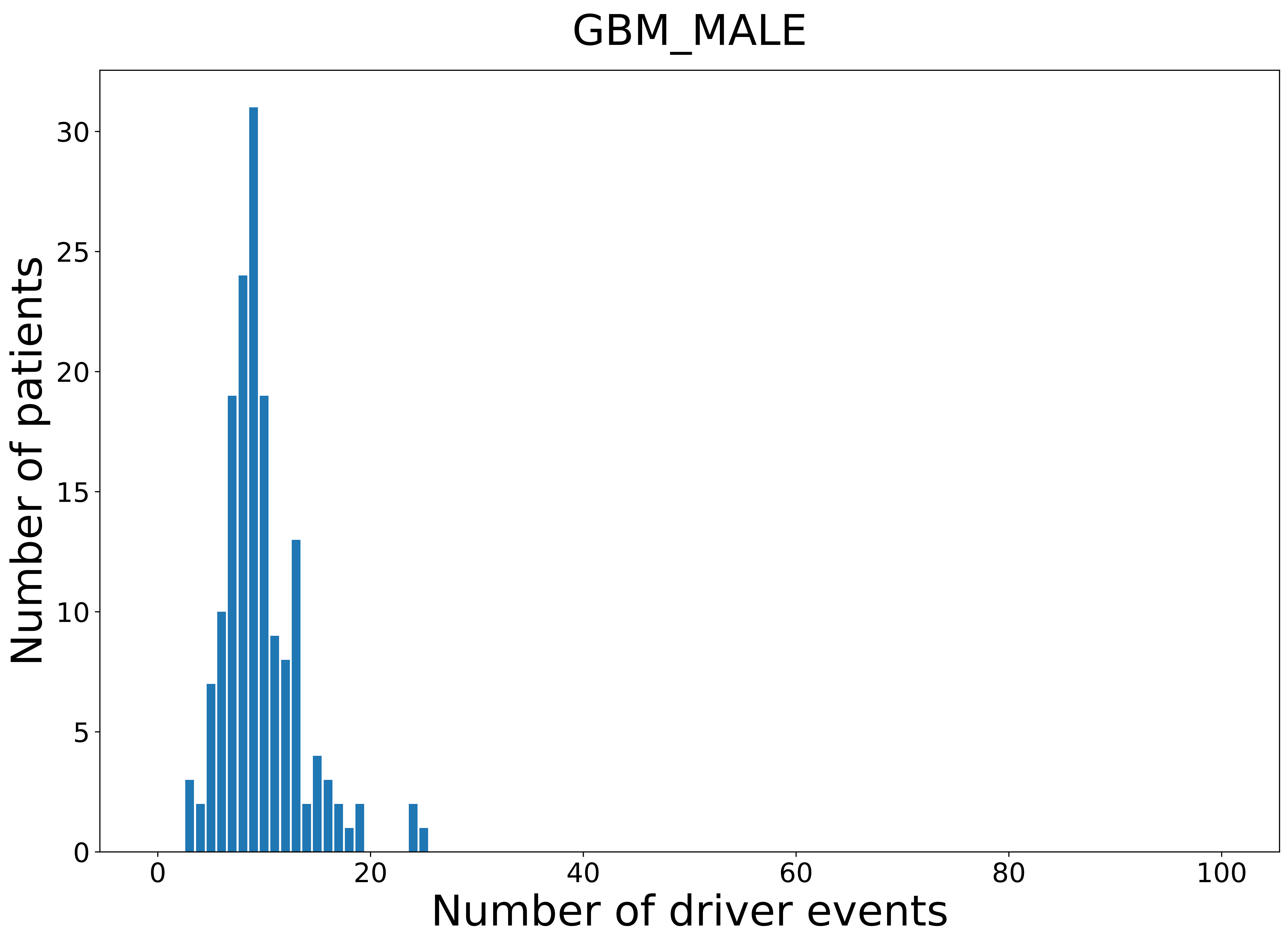

### 2021_8_16_14_9_KICH.pdf

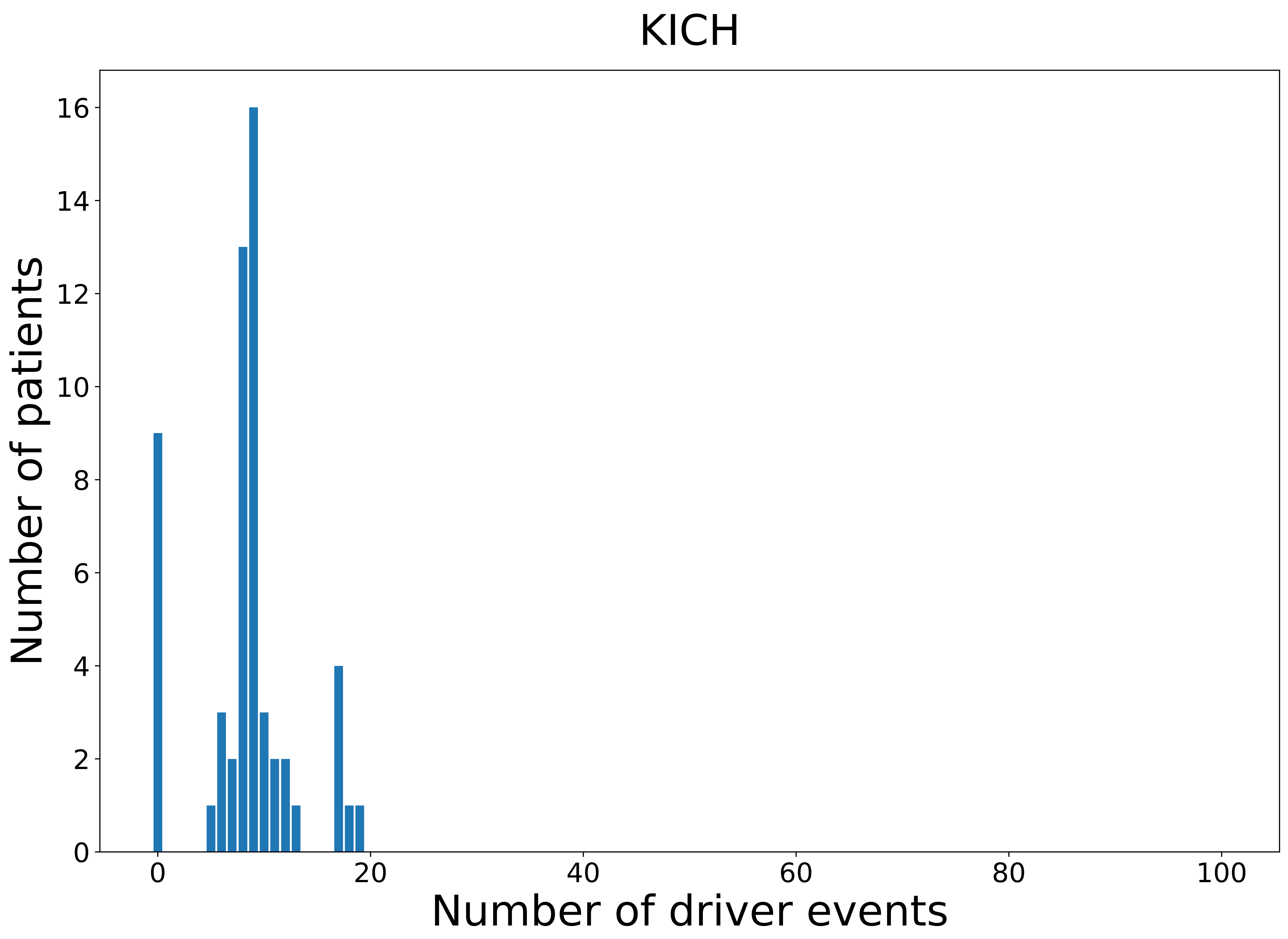

### 2021_8_16_14_9_KICH_FEMALE.pdf

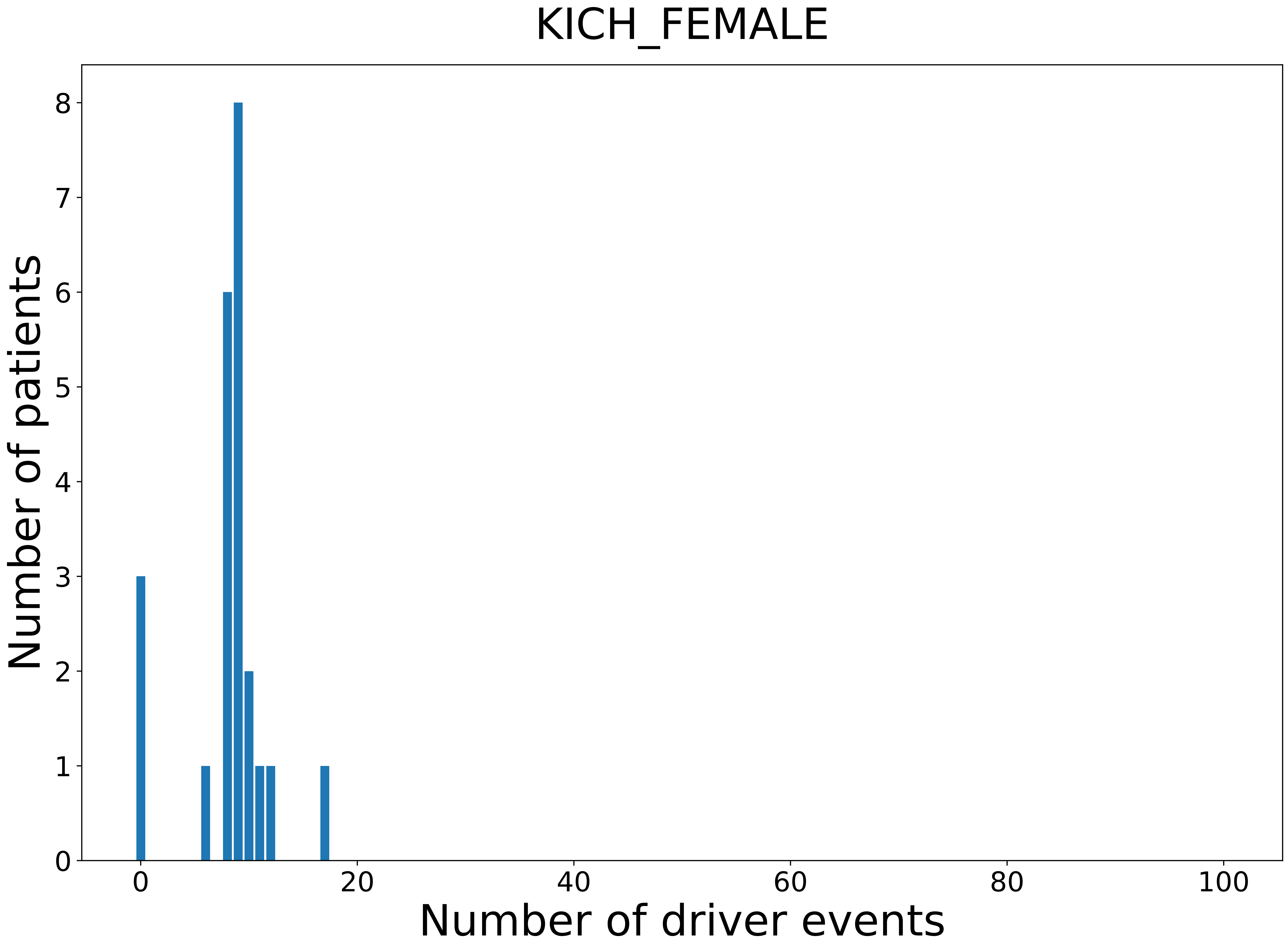

### 2021_8_16_14_9_KIRC_FEMALE.pdf

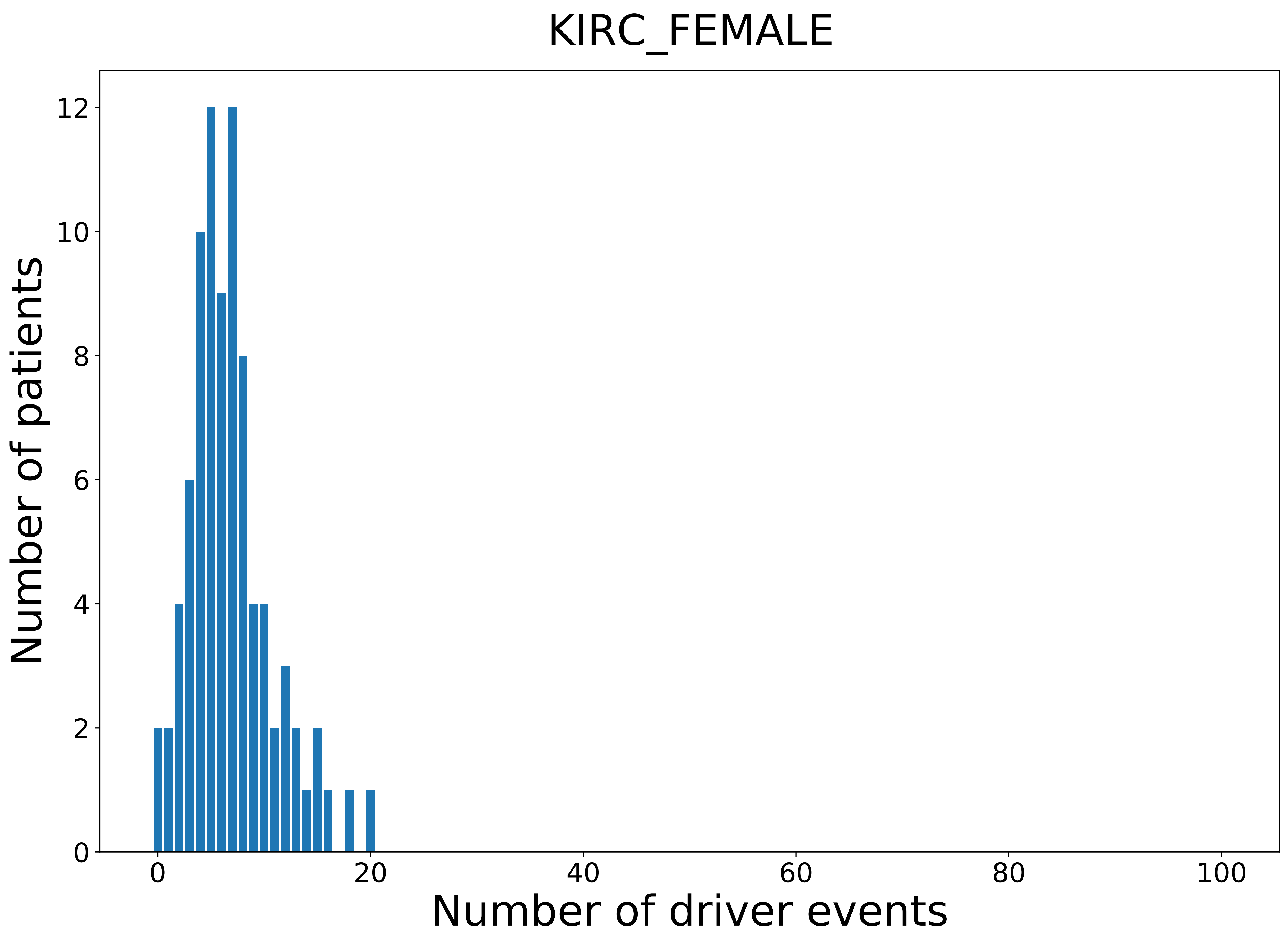

### 2021_8_16_14_9_KIRP.pdf

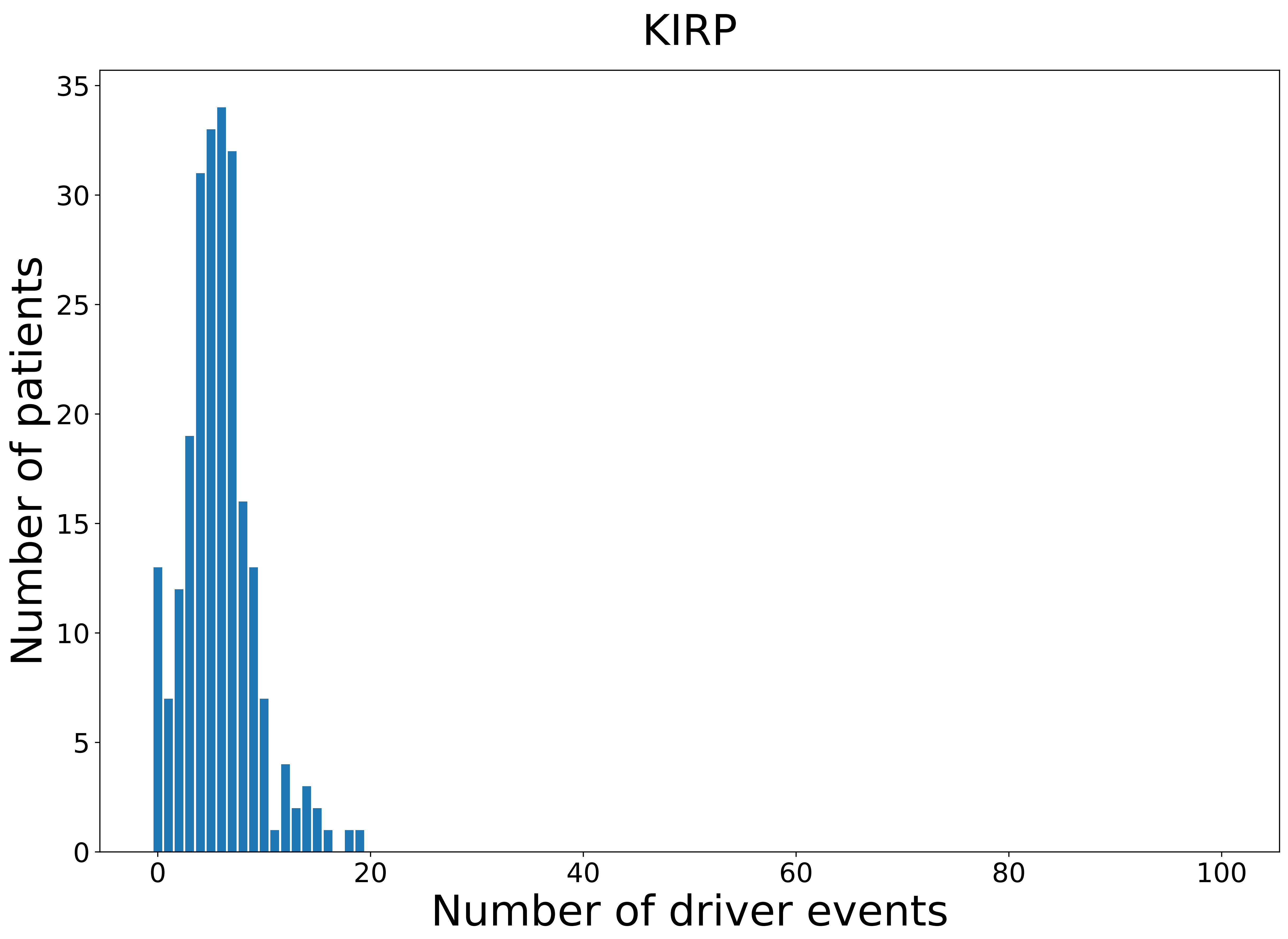

### 2021_8_16_14_9_KIRP_FEMALE.pdf

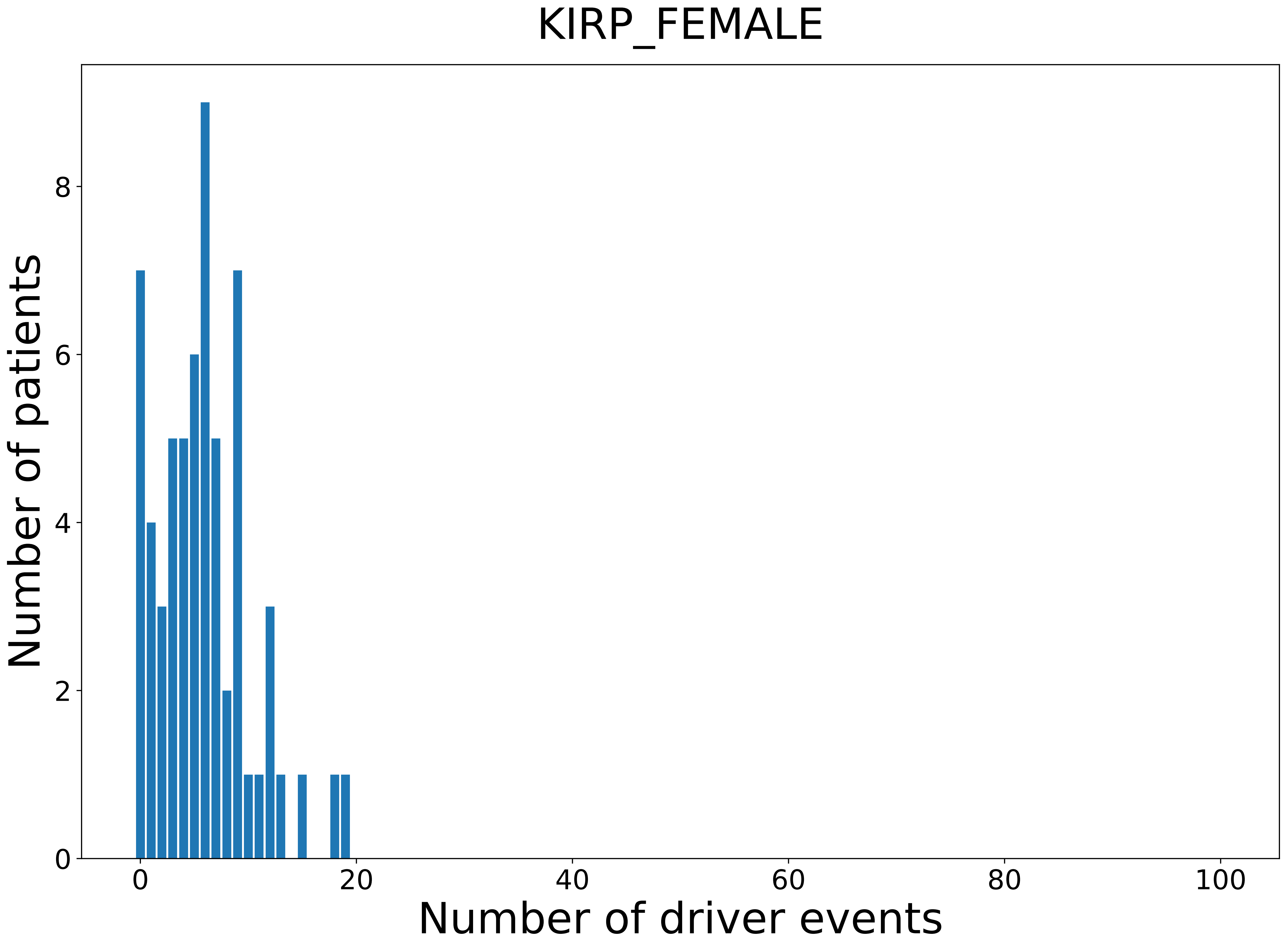

### 2021_8_16_14_9_LGG_MALE.pdf

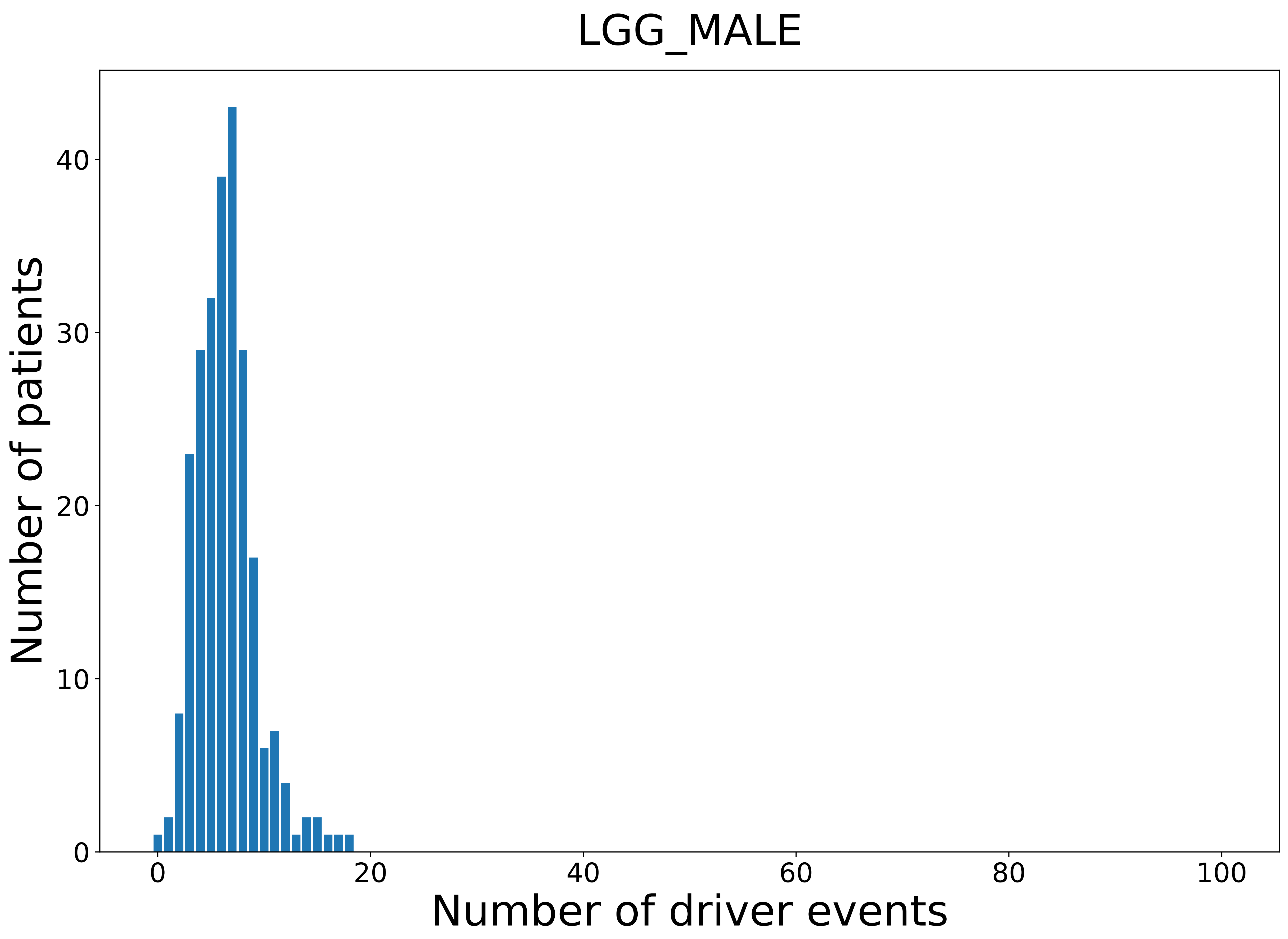

### 2021_8_16_14_9_LIHC_FEMALE.pdf

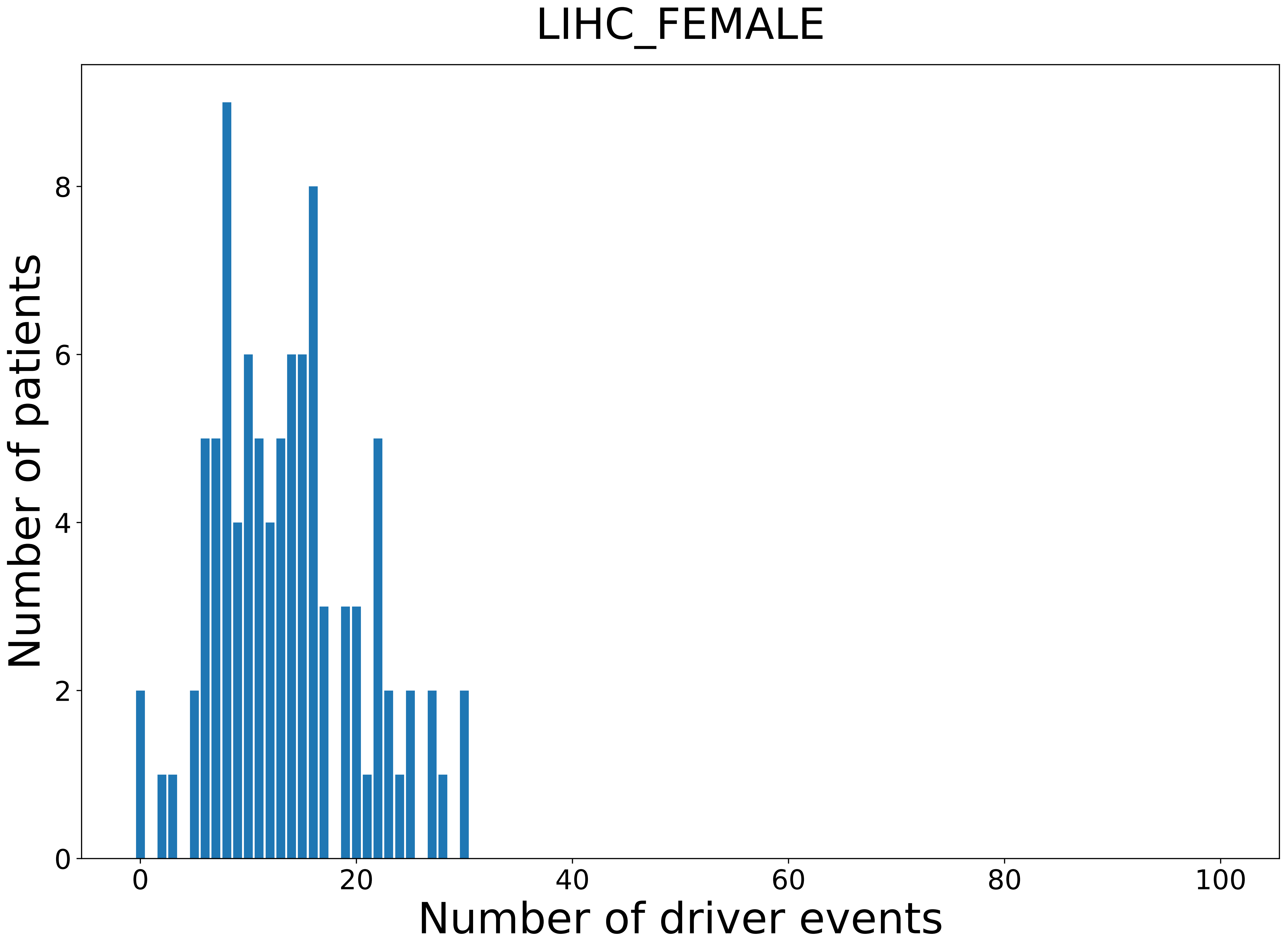

### 2021_8_16_14_9_LUAD_MALE.pdf

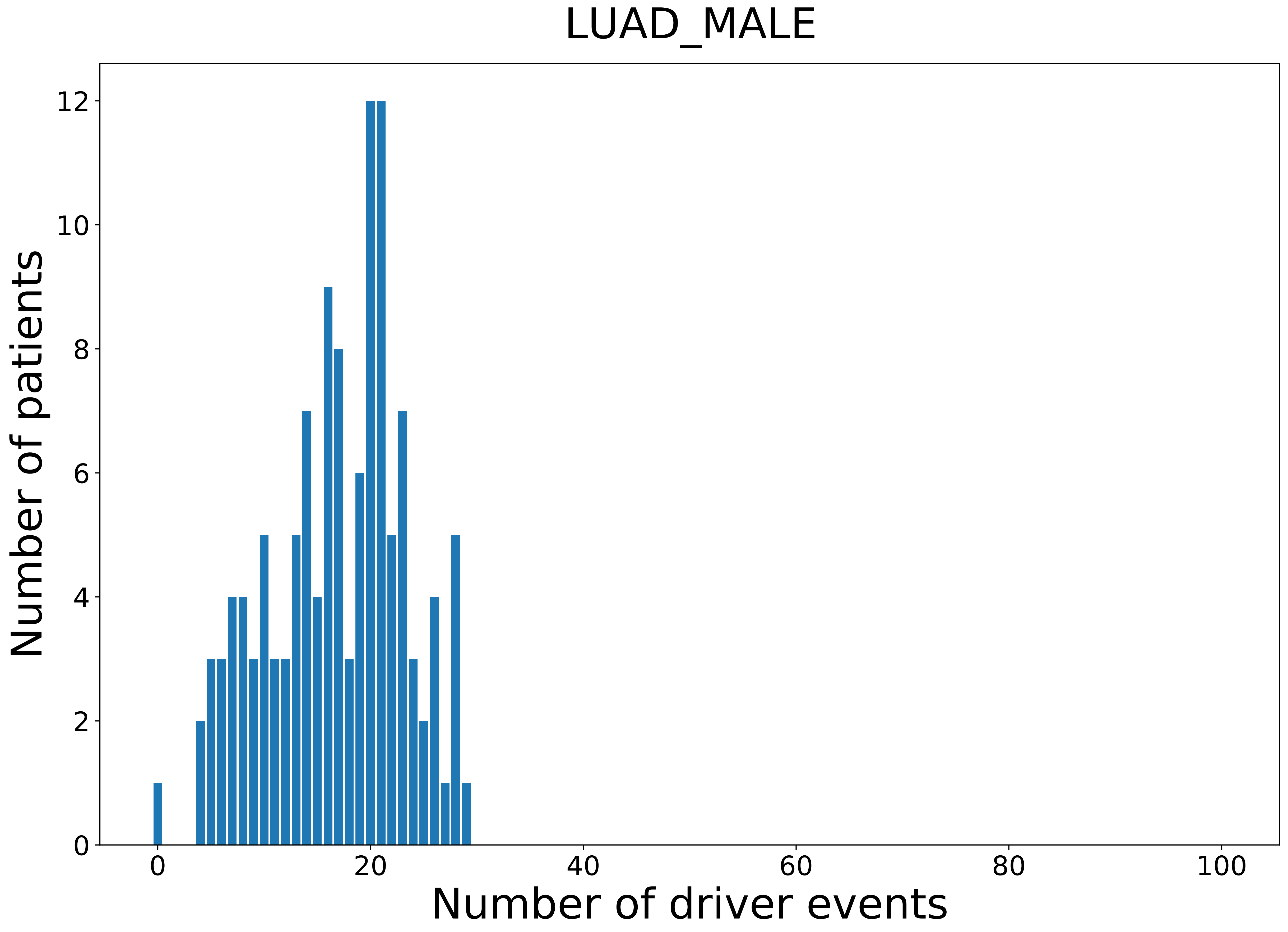

### 2021_8_16_14_9_LUSC_FEMALE.pdf

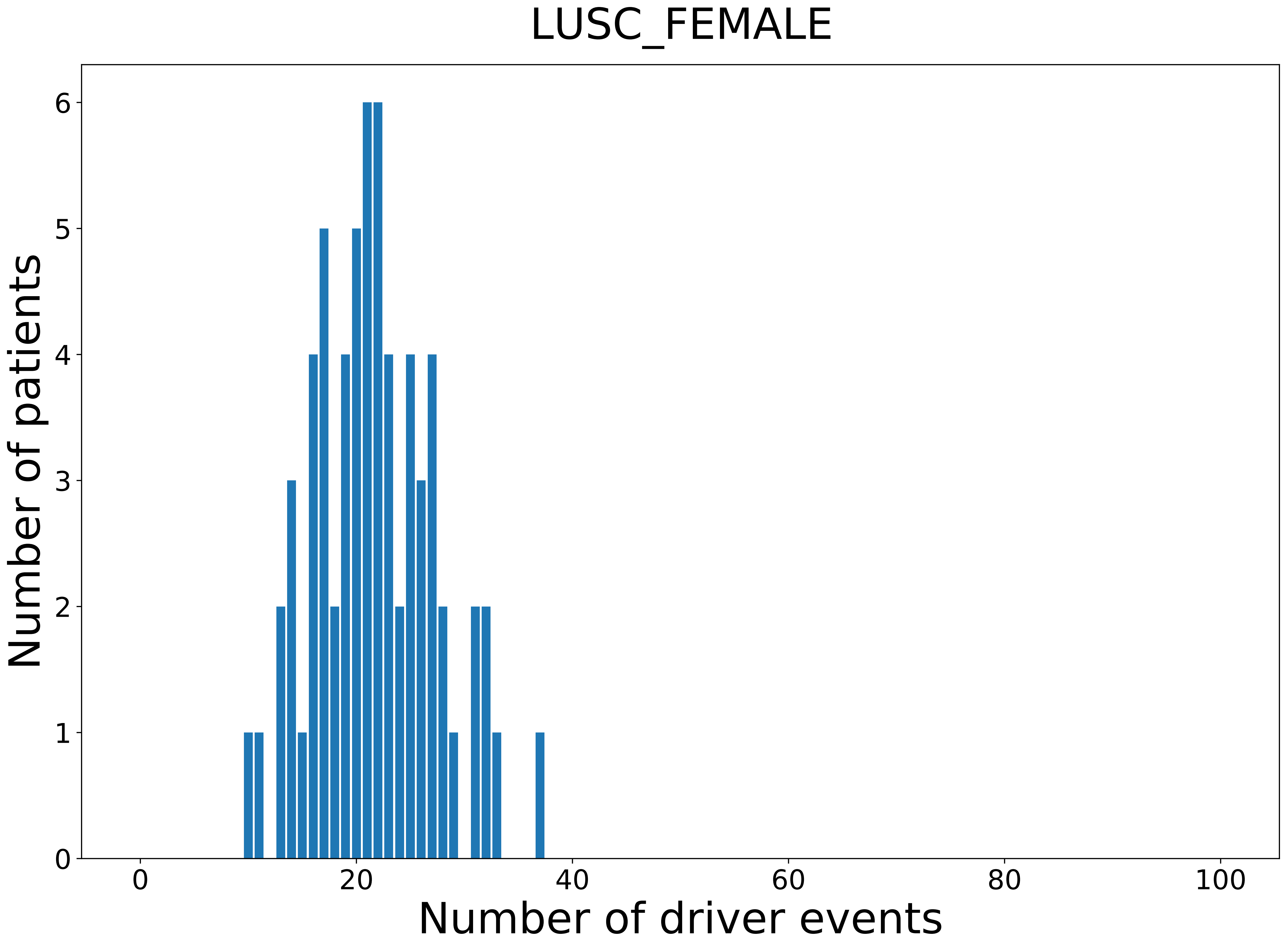

### 2021_8_16_14_9_LUSC_MALE.pdf

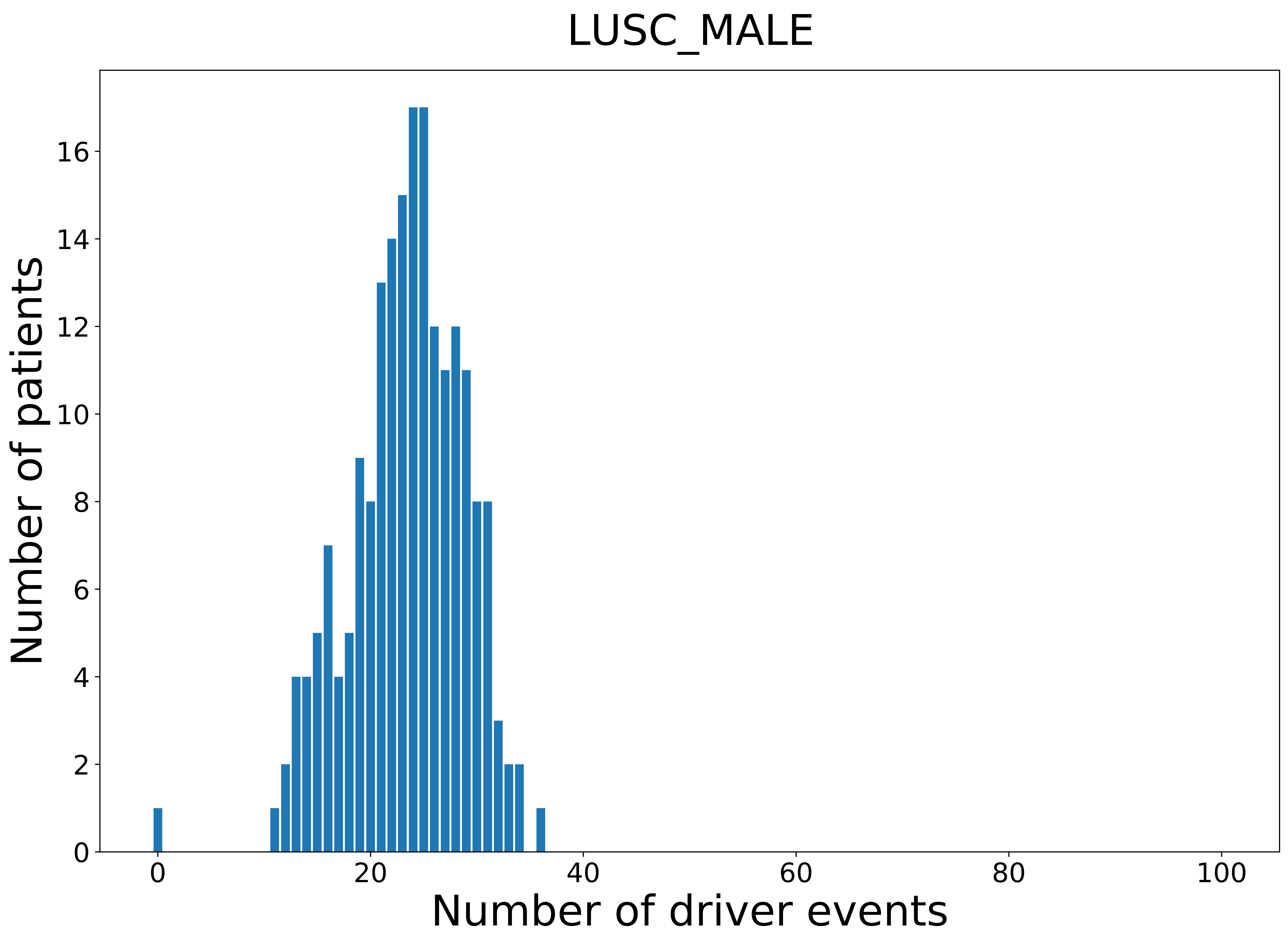

### 2021_8_16_14_9_MESO.pdf

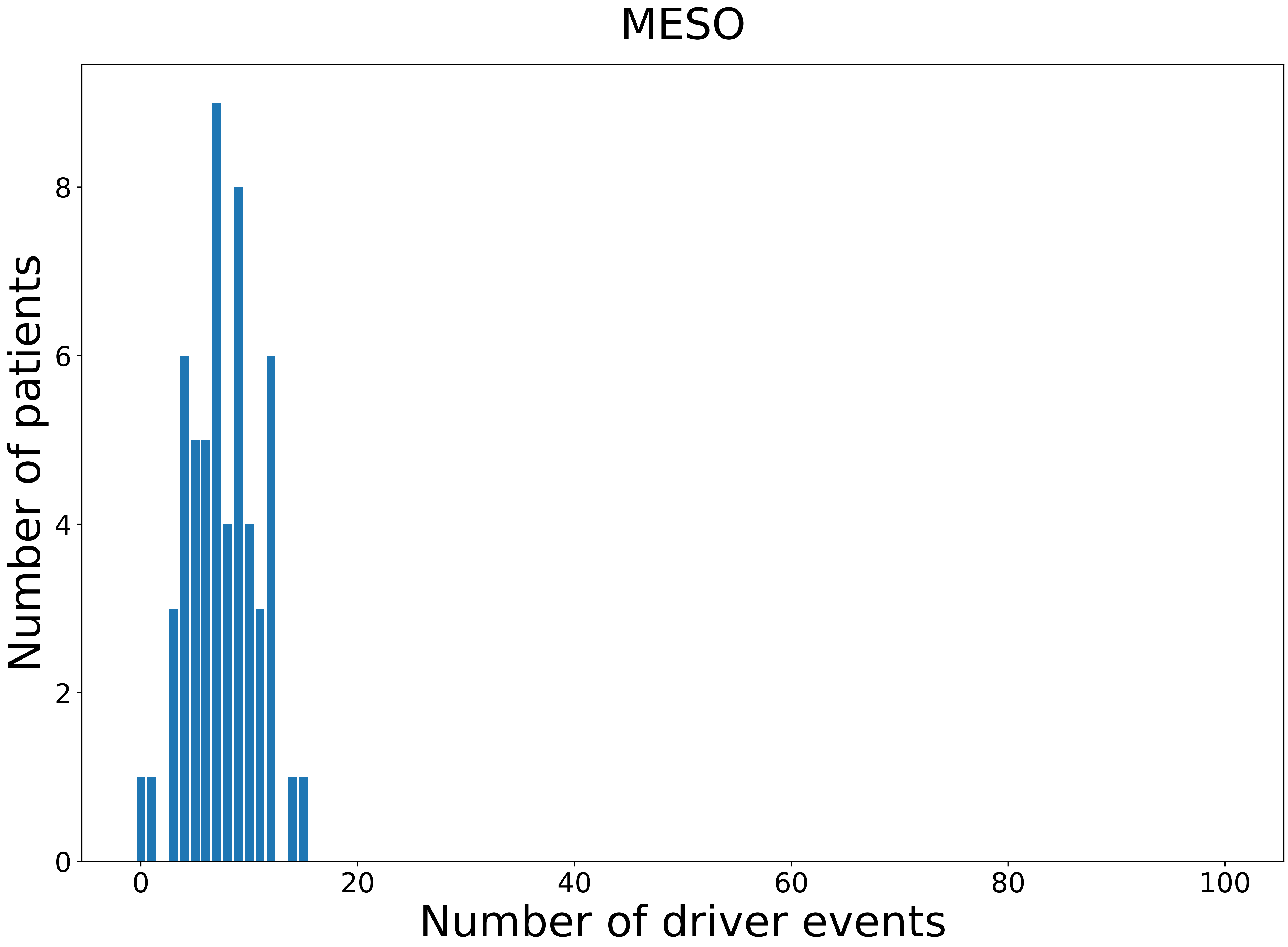

### 2021_8_16_14_9_MESO_MALE.pdf

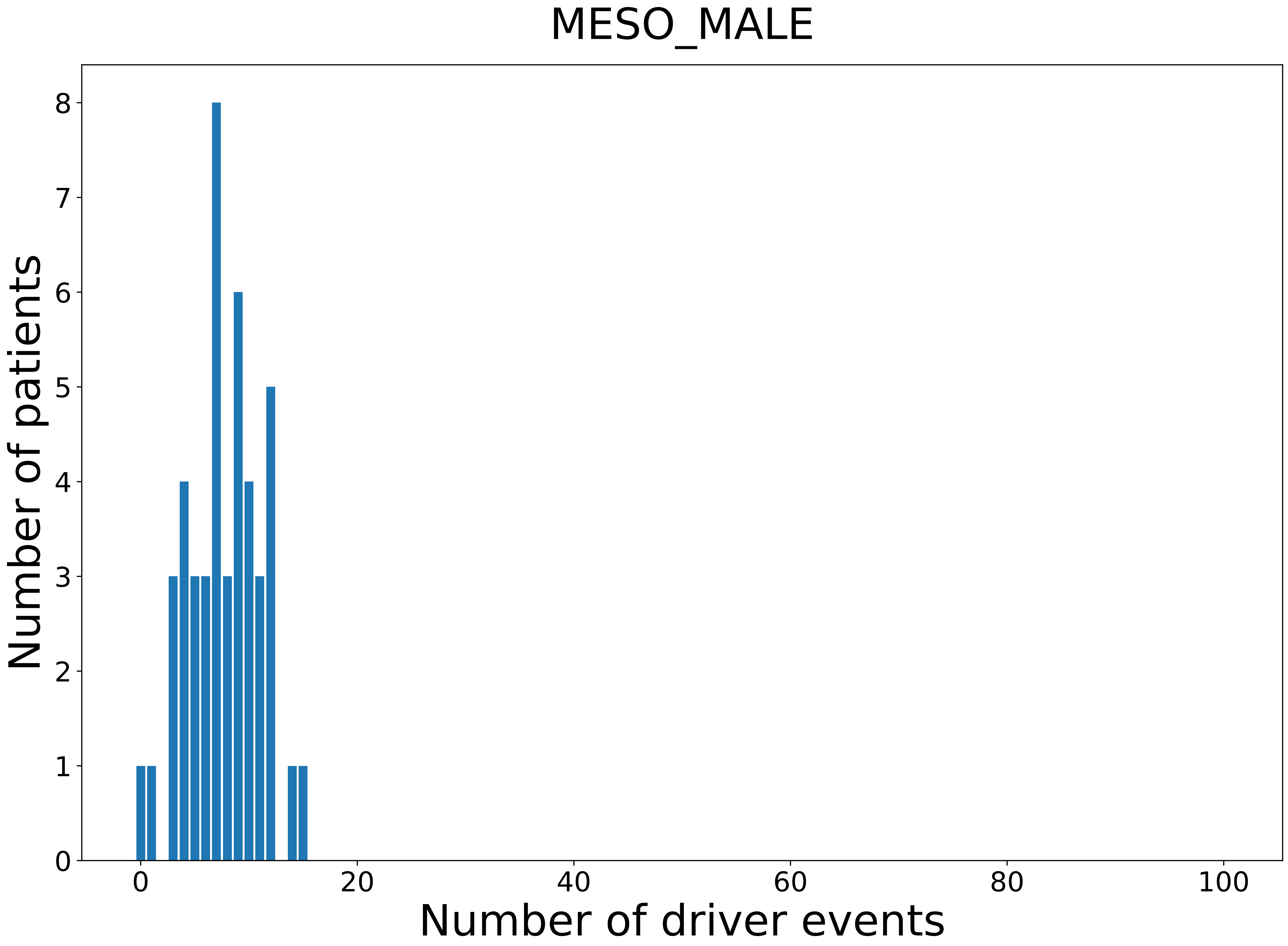

### 2021_8_16_14_9_OV.pdf

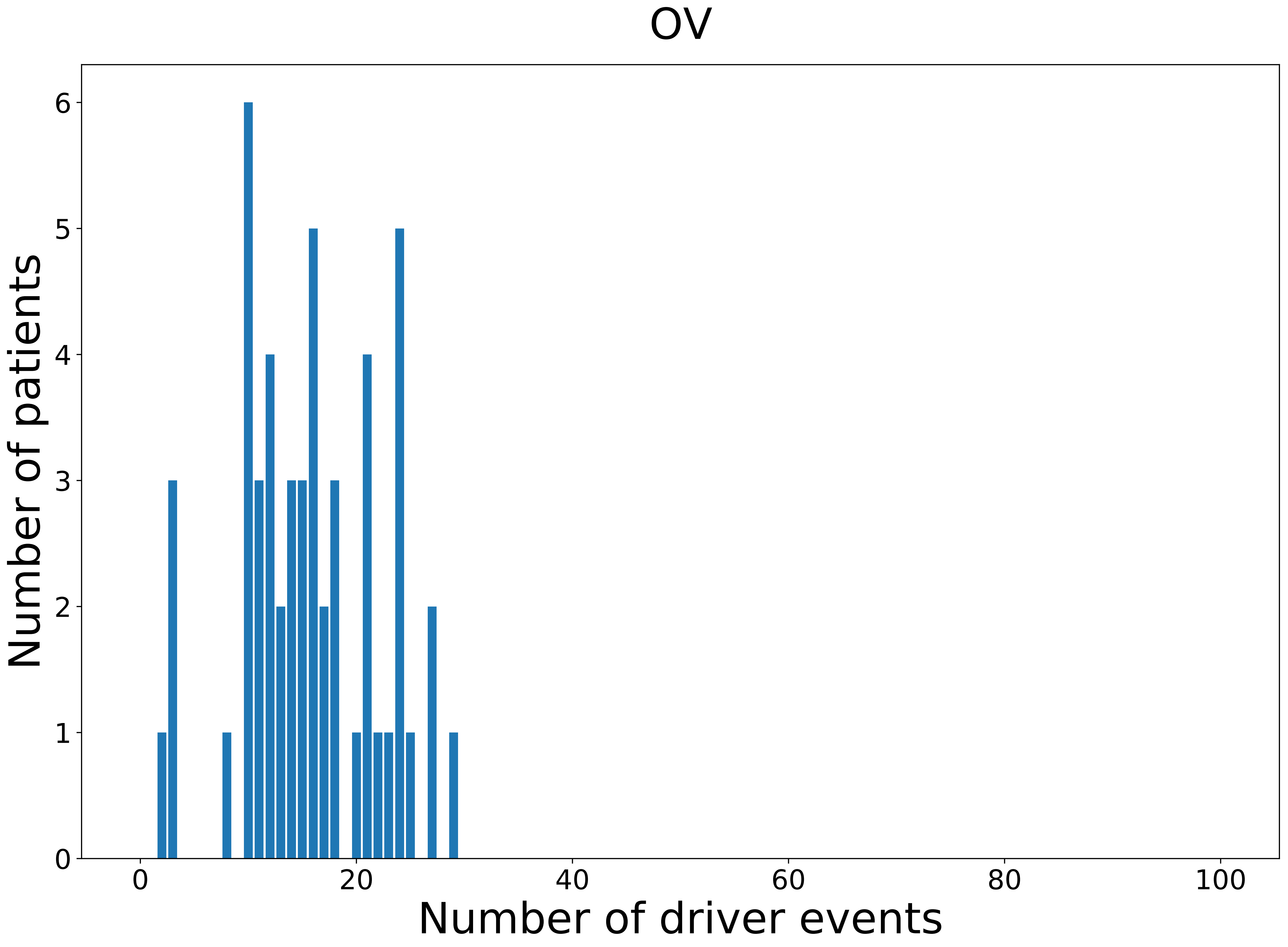

### 2021_8_16_14_9_PAAD_MALE.pdf

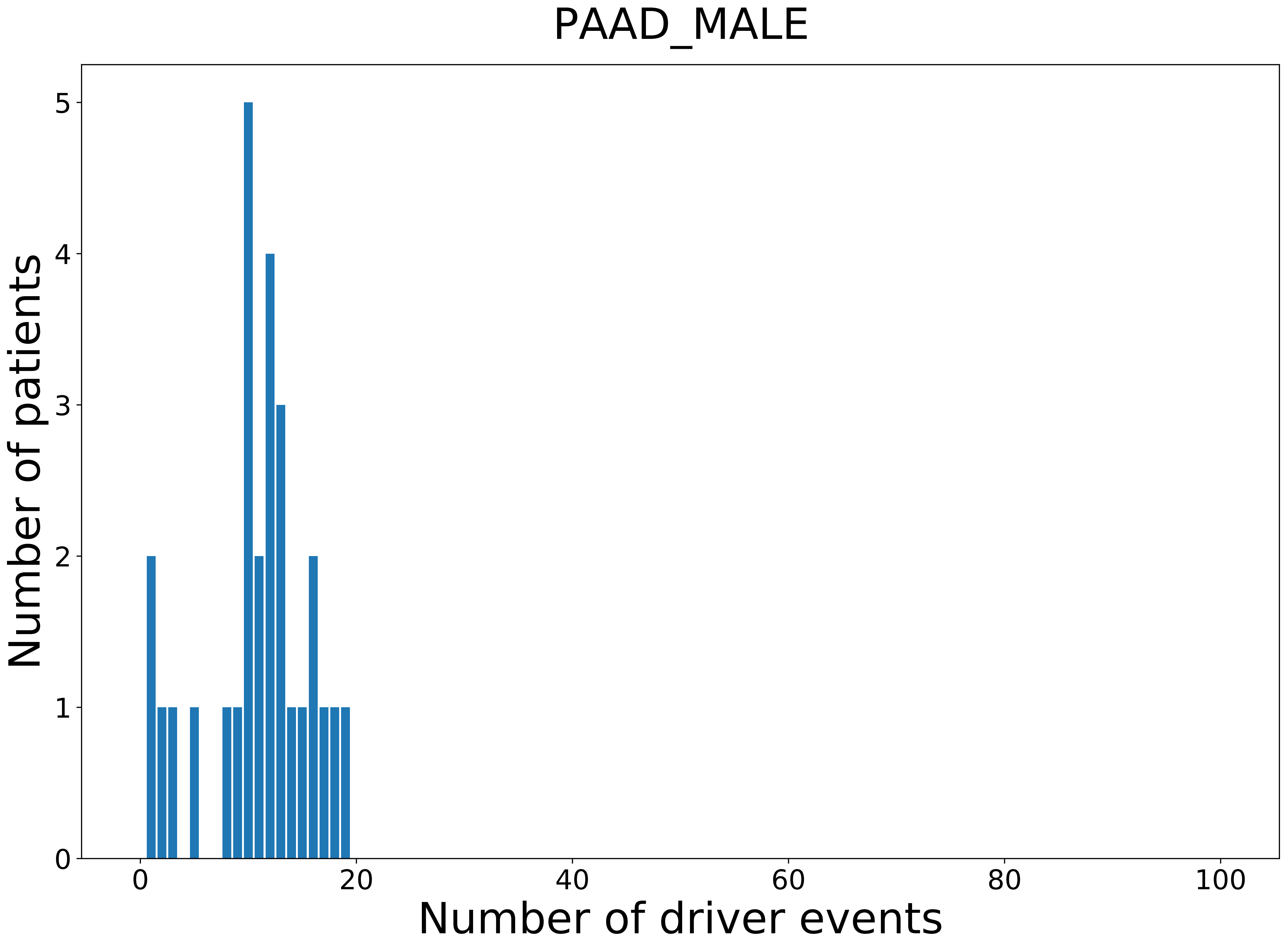

### 2021_8_16_14_9_PRAD_MALE.pdf

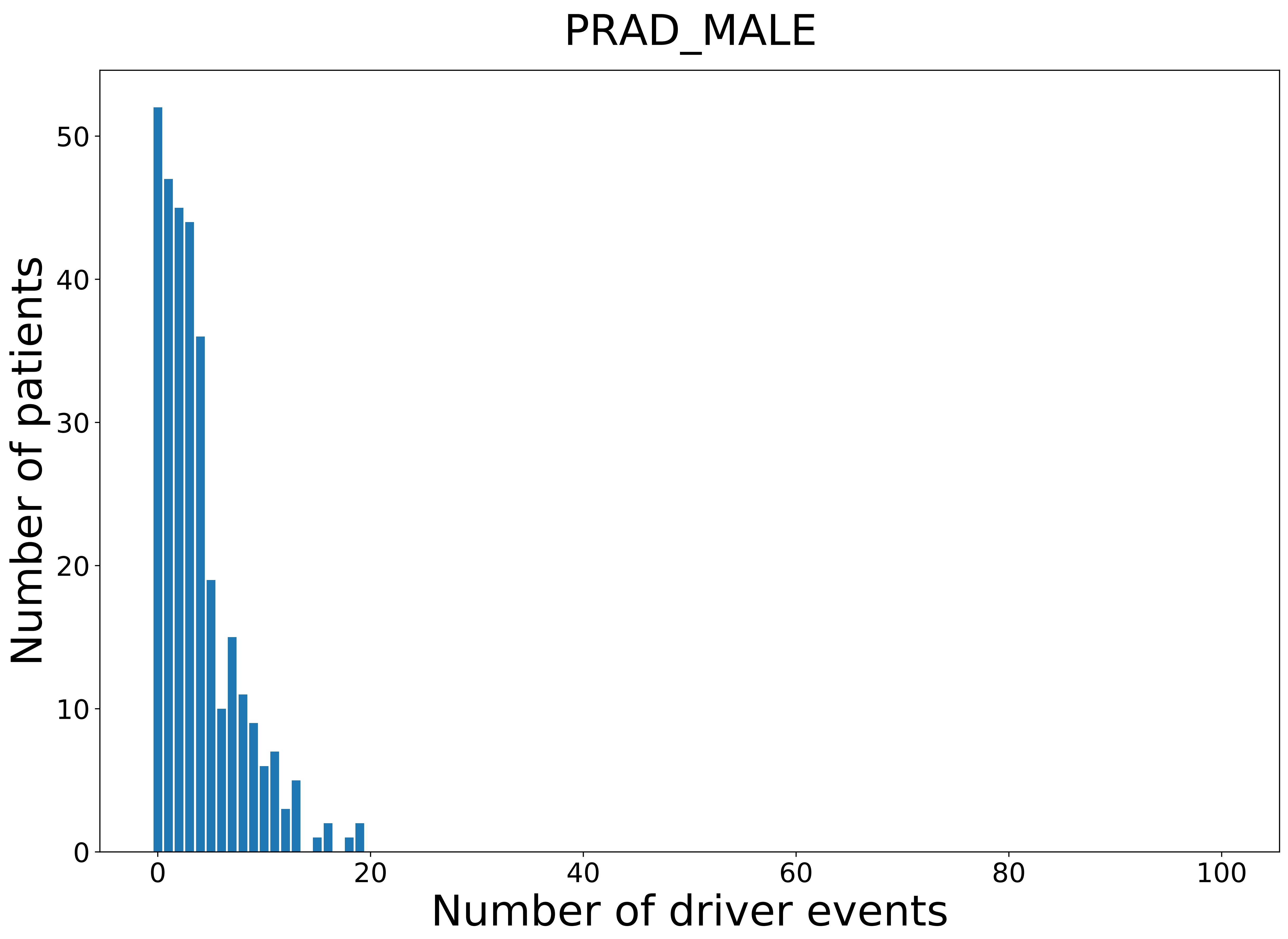

### 2021_8_16_14_9_SARC_MALE.pdf

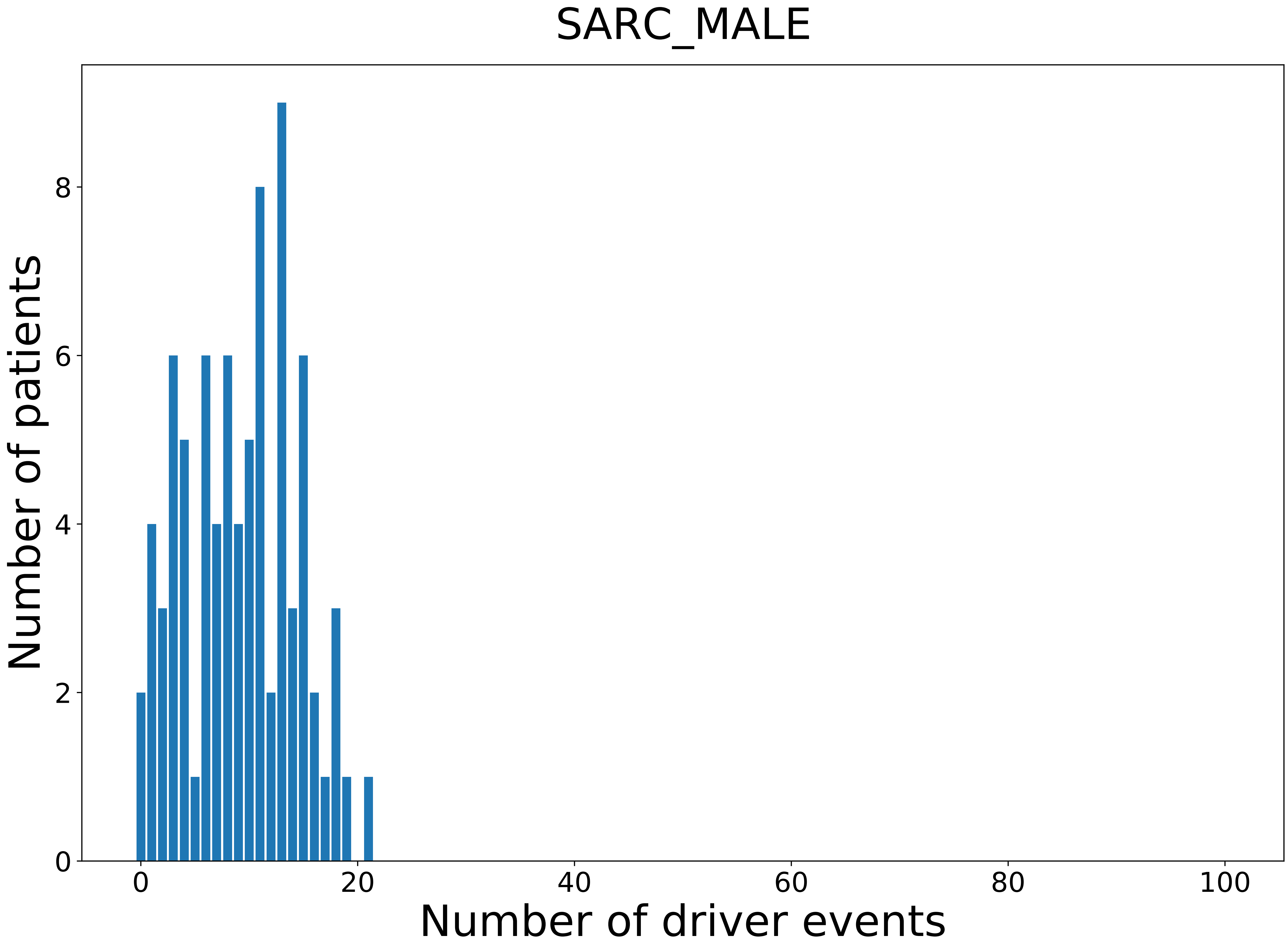

### 2021_8_16_14_9_SKCM.pdf

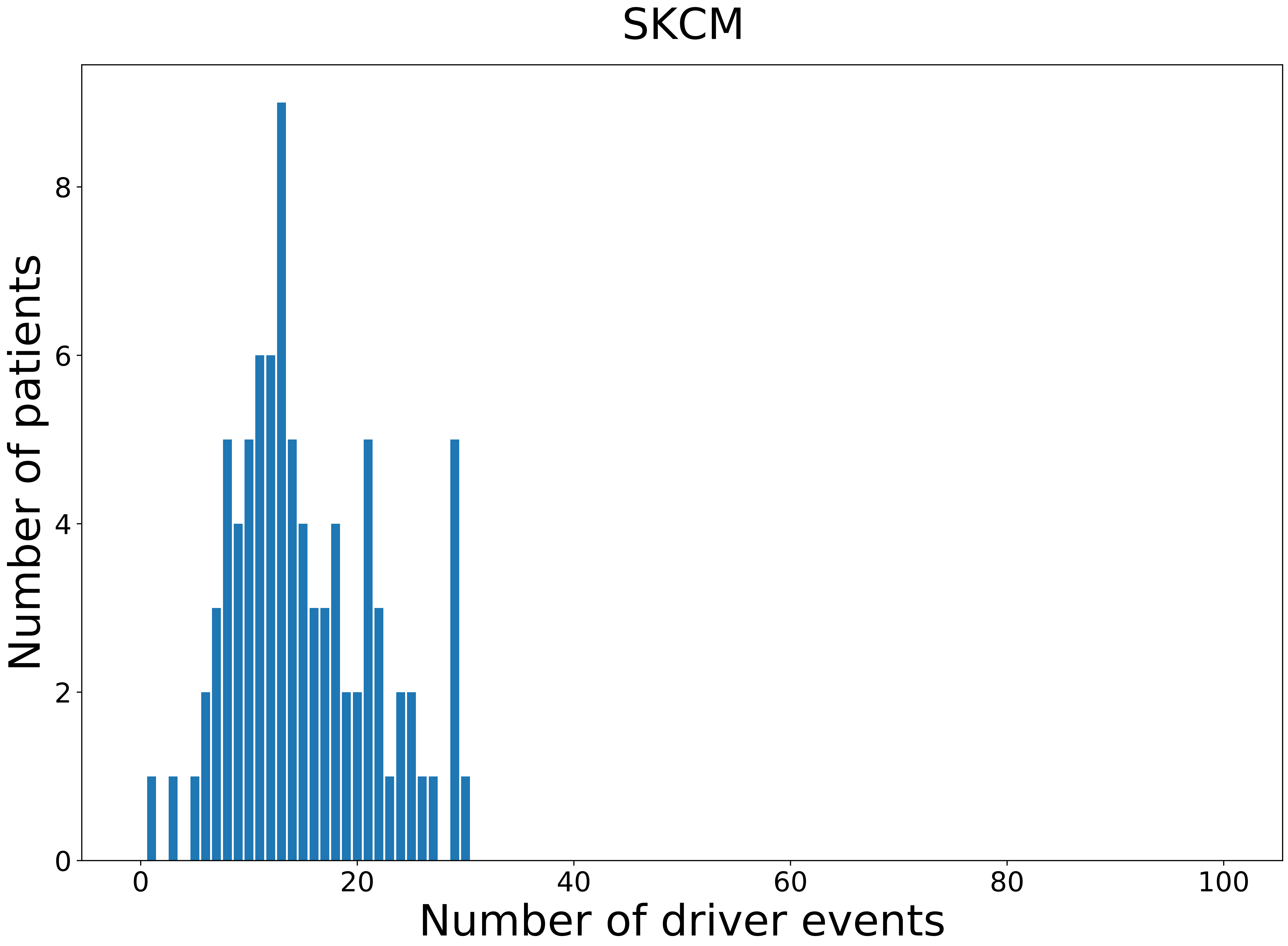

### 2021_8_16_14_9_SKCM_MALE.pdf

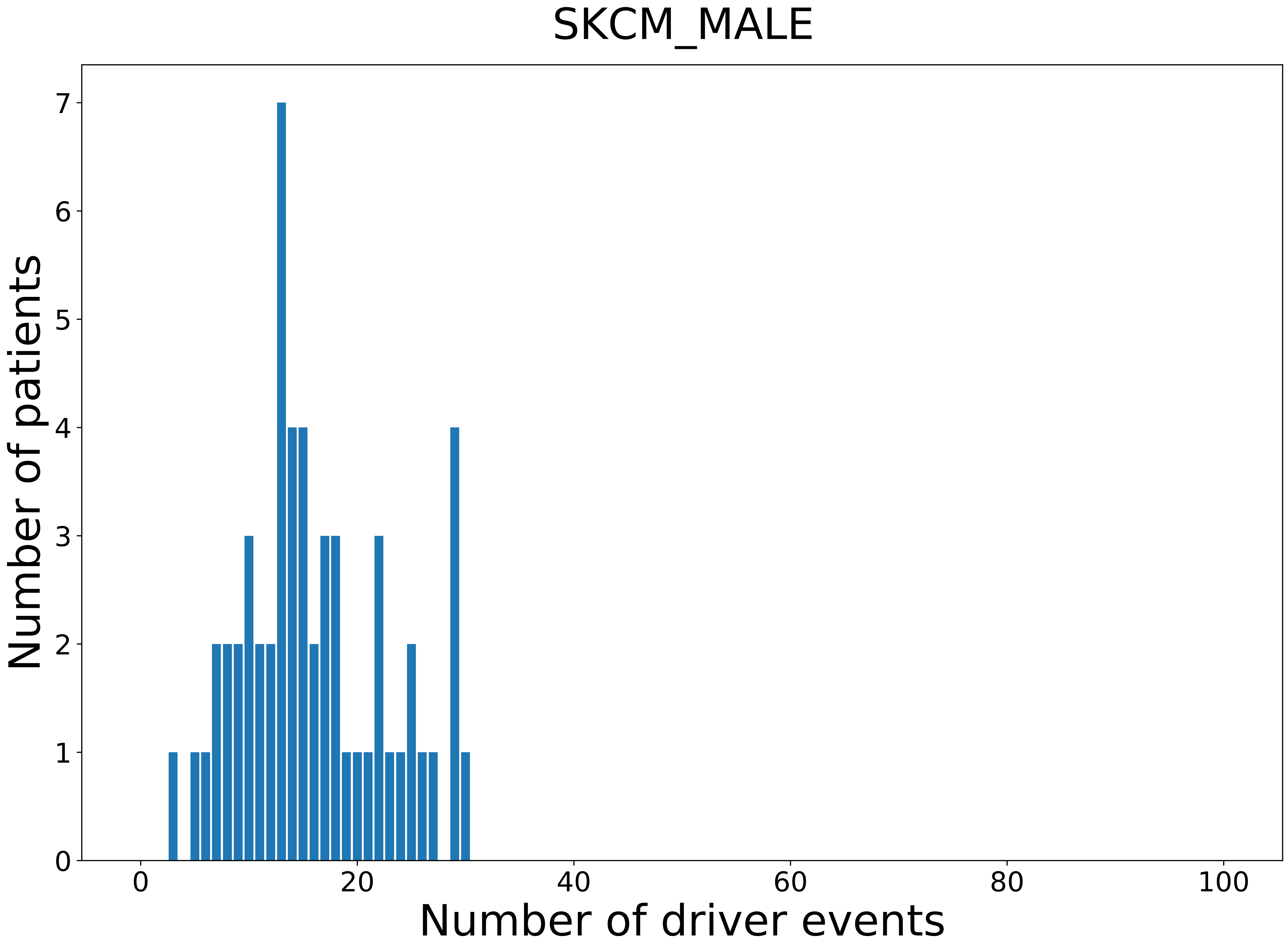

### 2021_8_16_14_9_STAD.pdf

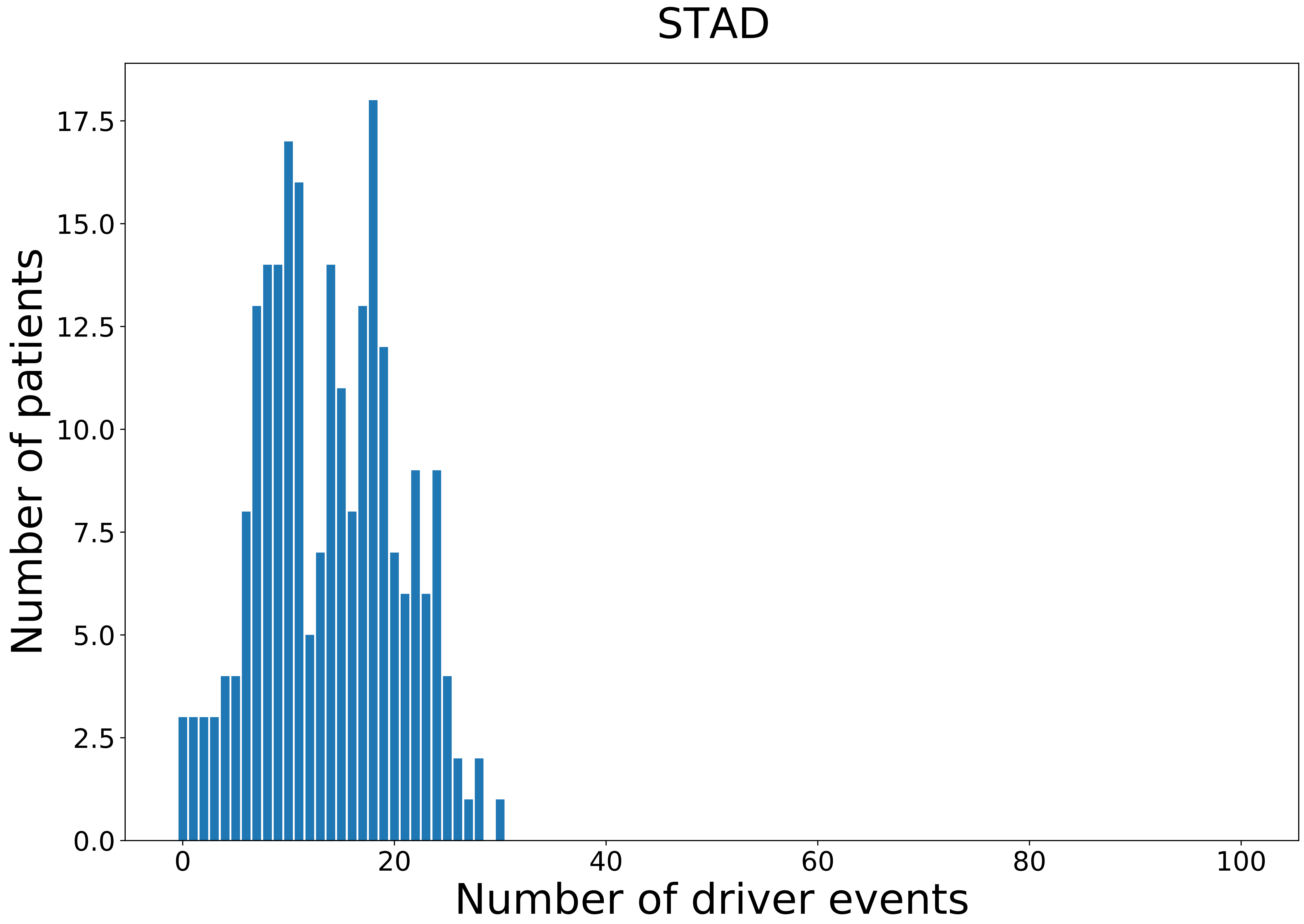

### 2021_8_16_14_9_TGCT_MALE.pdf

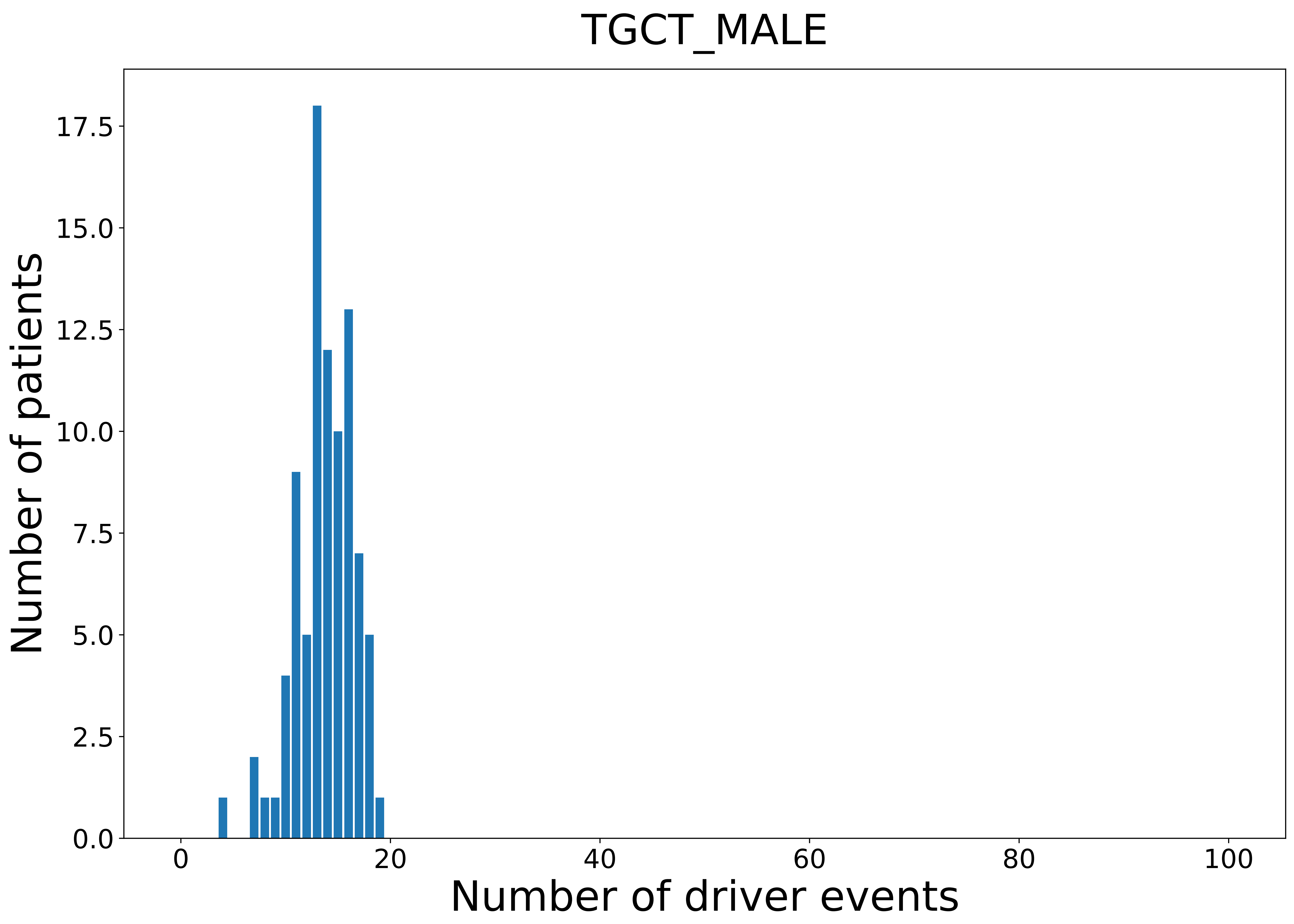

### 2021_8_16_14_9_UCEC.pdf

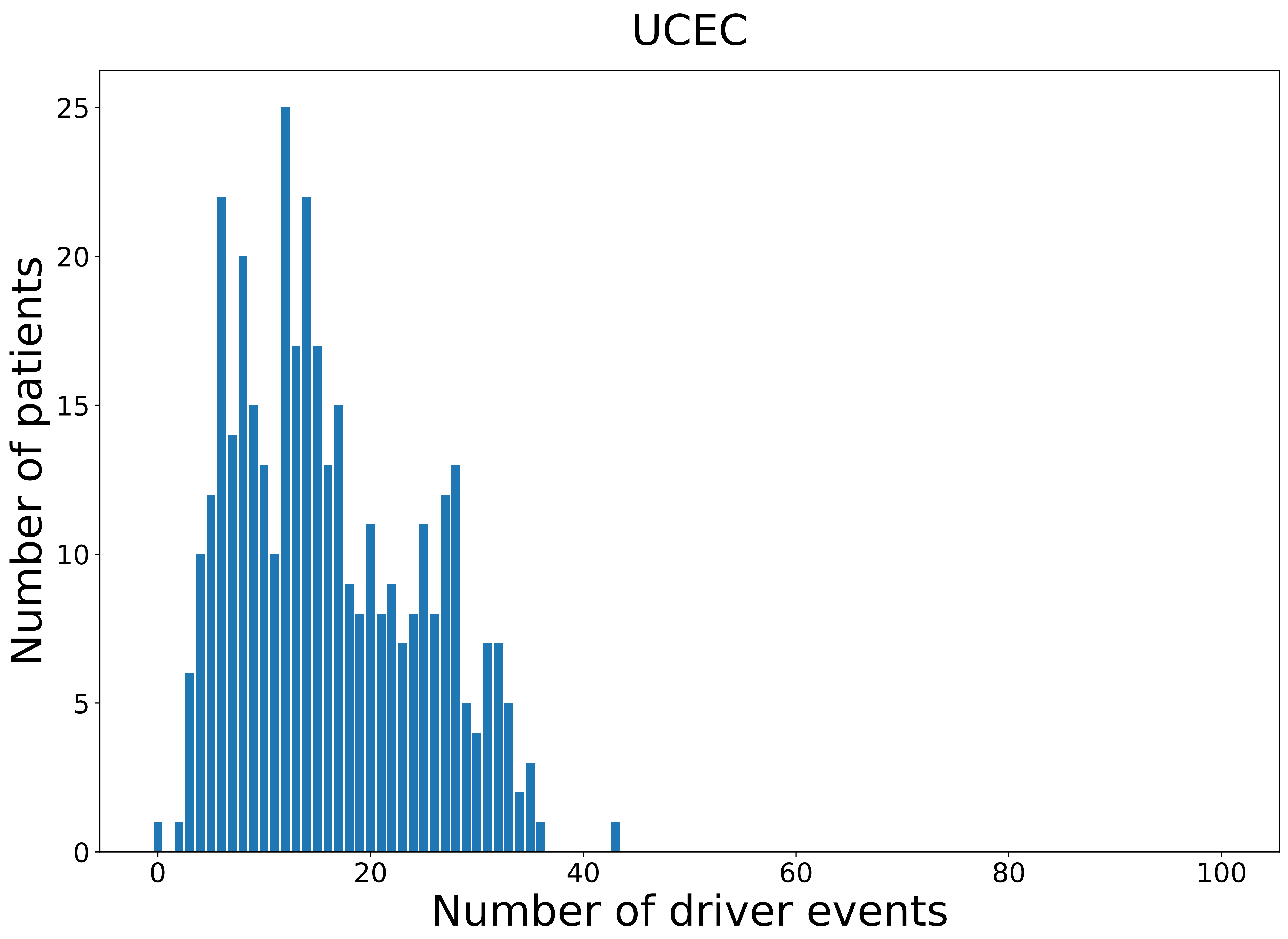

### 2021_8_16_14_9_UVM.pdf

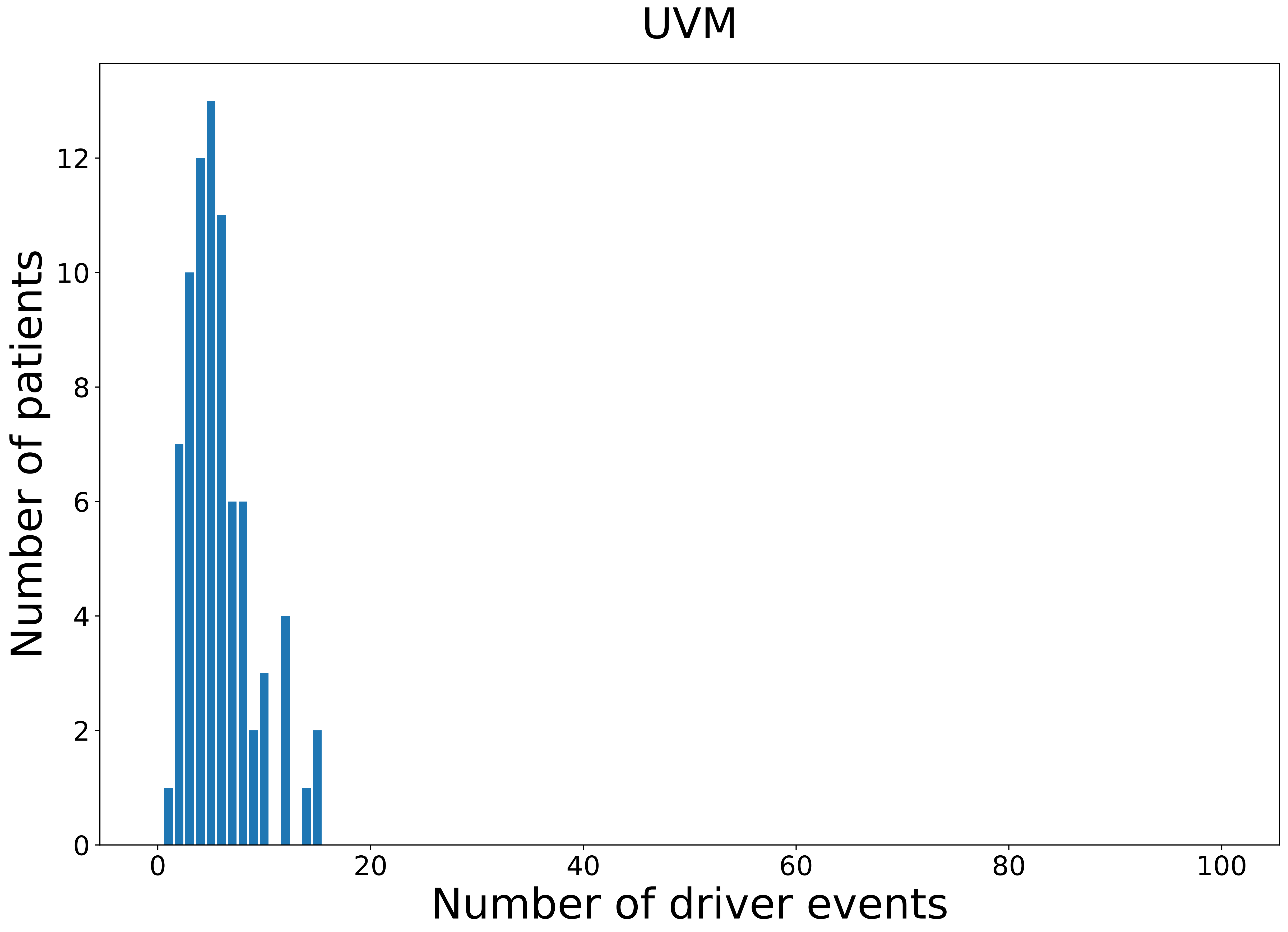

### 2021_8_16_14_9_UVM_FEMALE.pdf

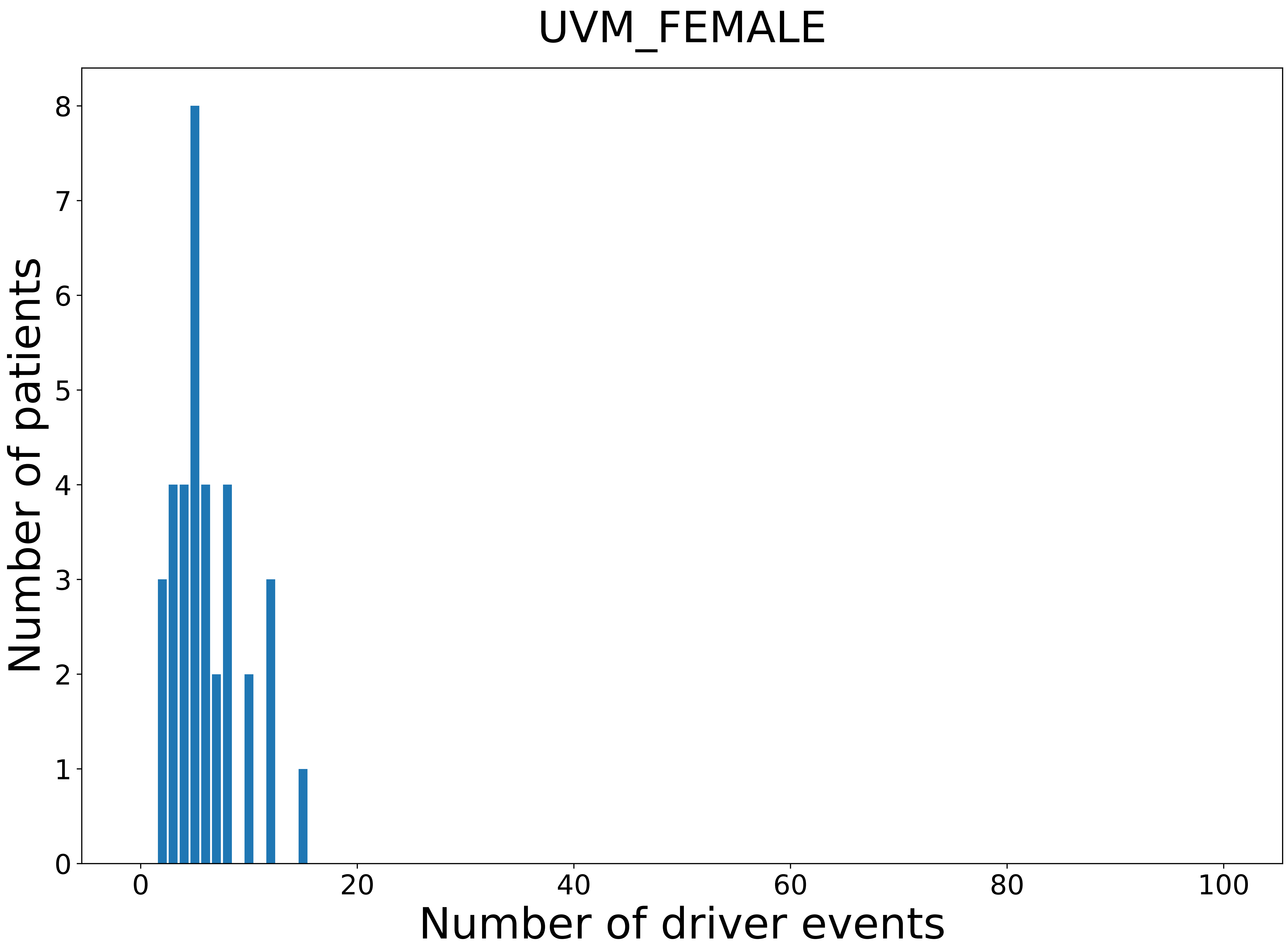

### 2021_8_16_20_32_distribution_events_detailed_1.pdf

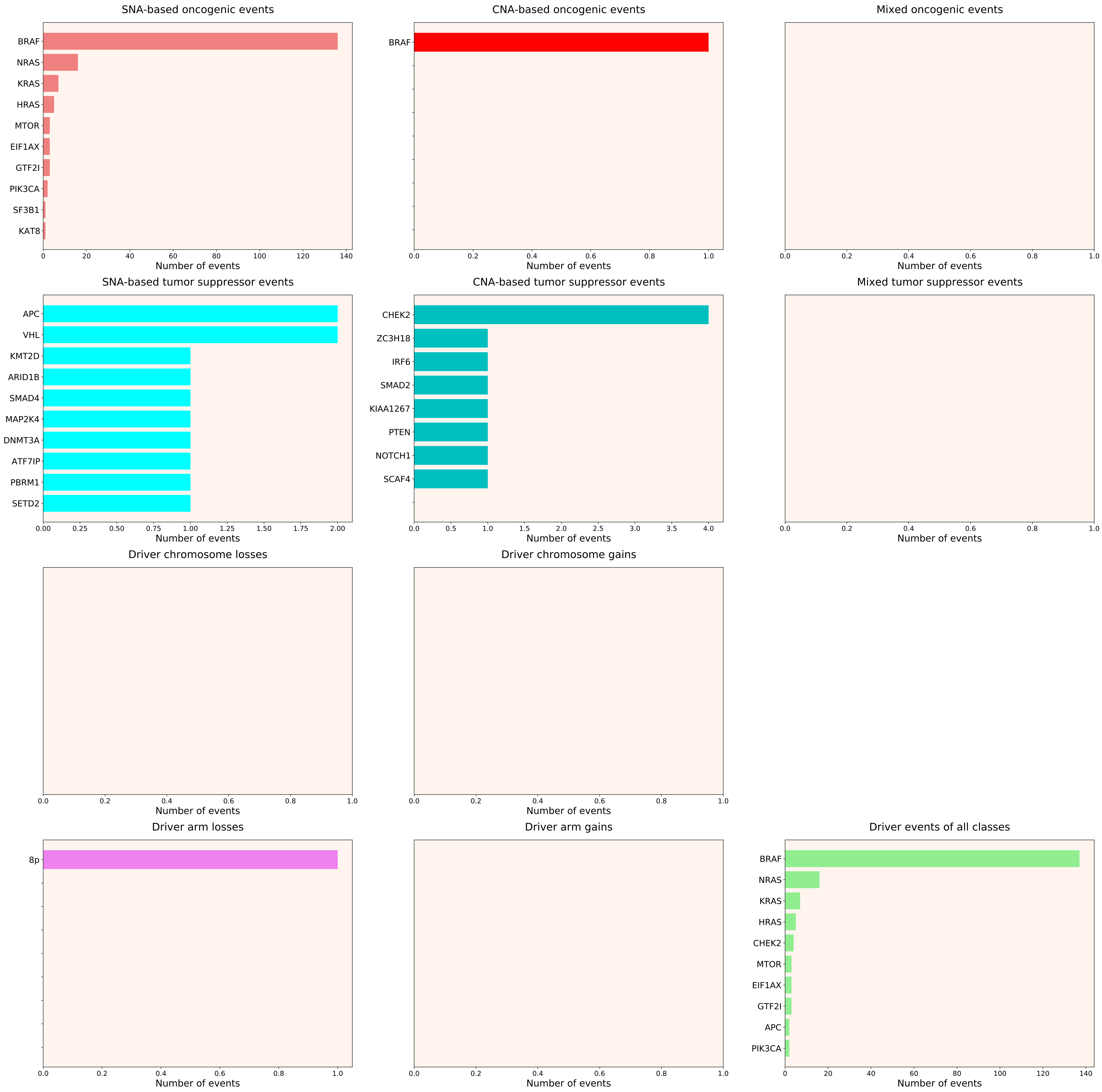

### 2021_8_16_20_32_distribution_events_detailed_2.pdf

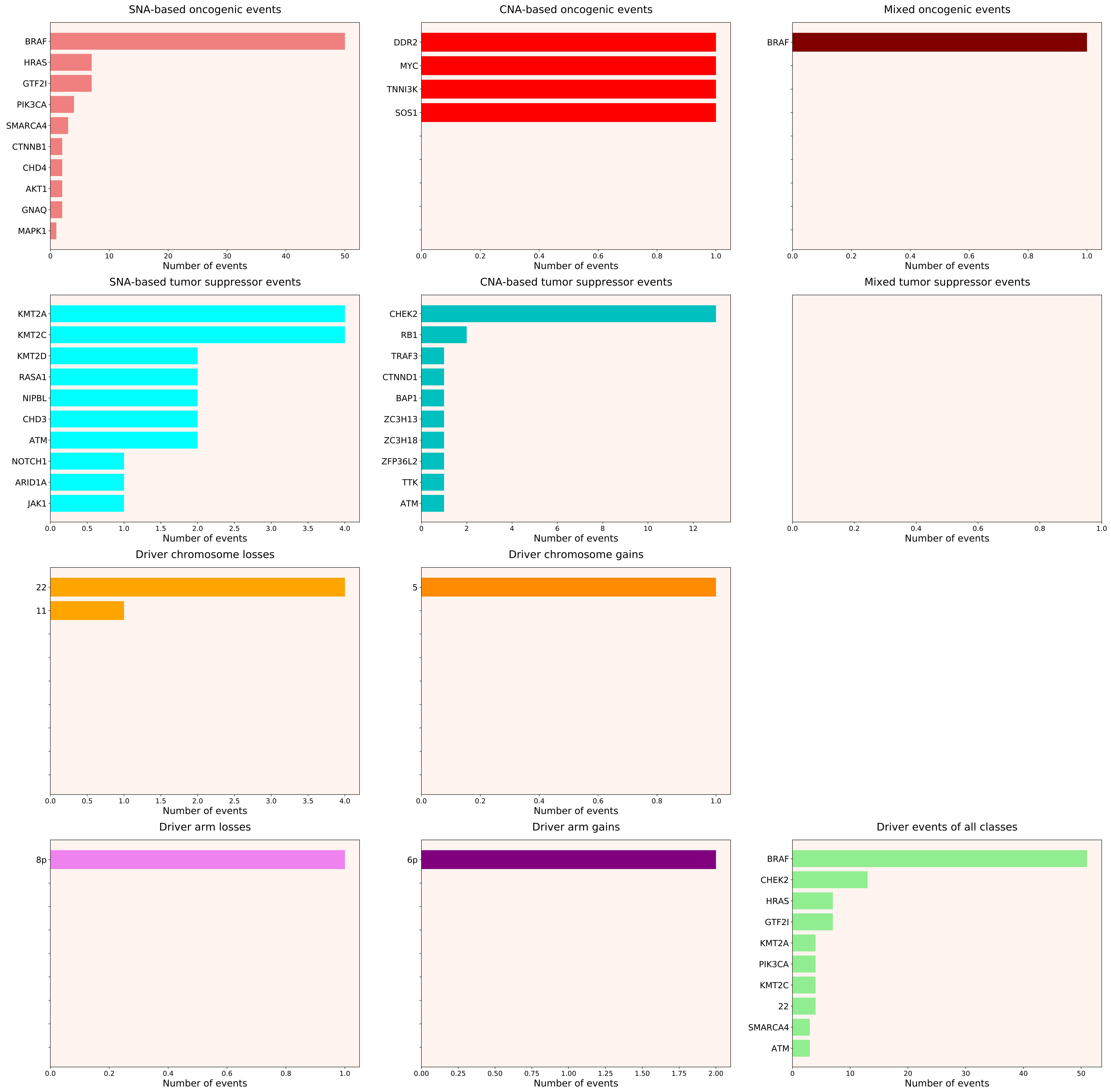

### 2021_8_16_20_32_distribution_events_detailed_4.pdf

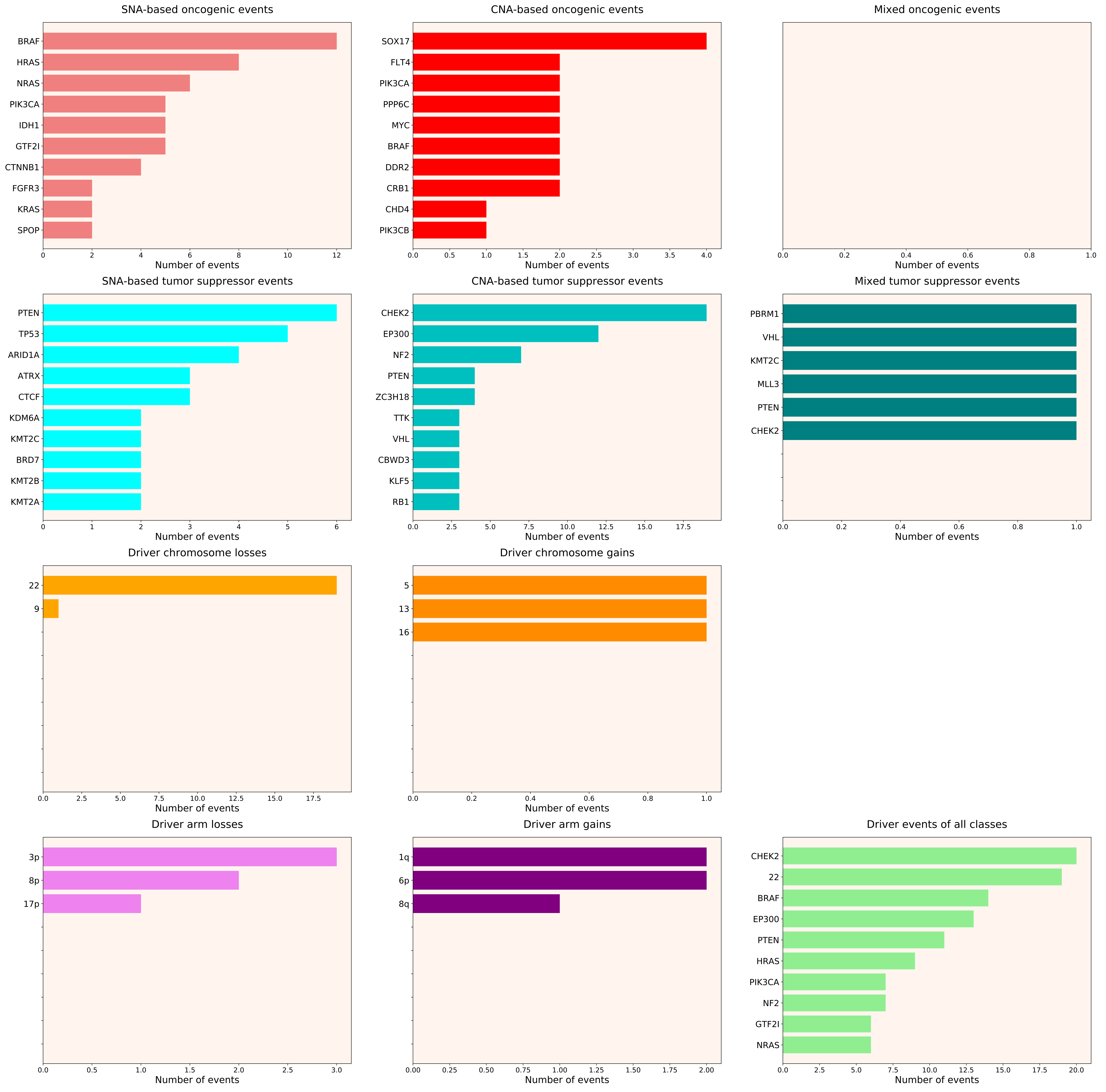

### 2021_8_16_20_32_distribution_events_detailed_5.pdf

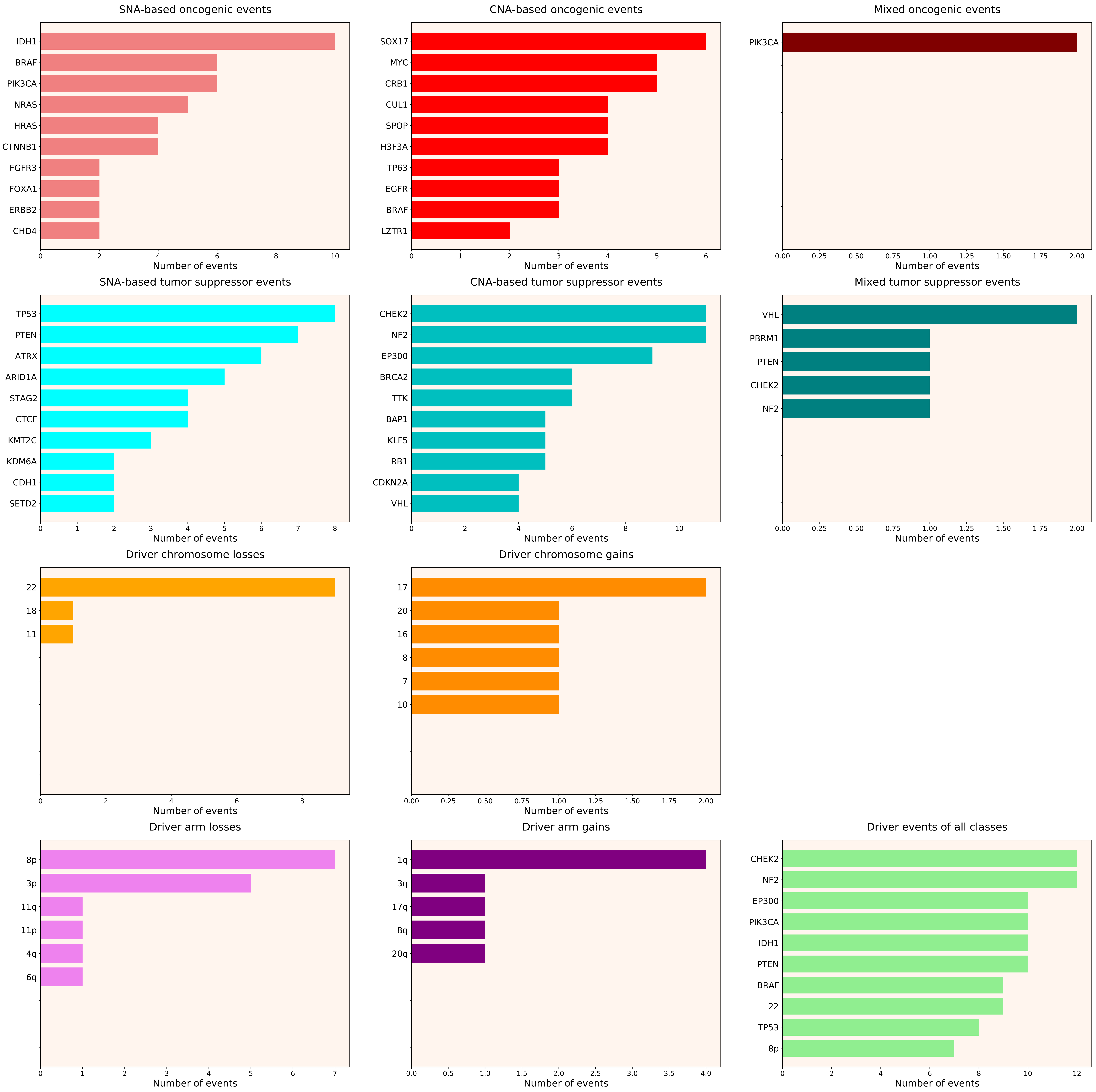

### 2021_8_16_20_32_distribution_events_detailed_6.pdf

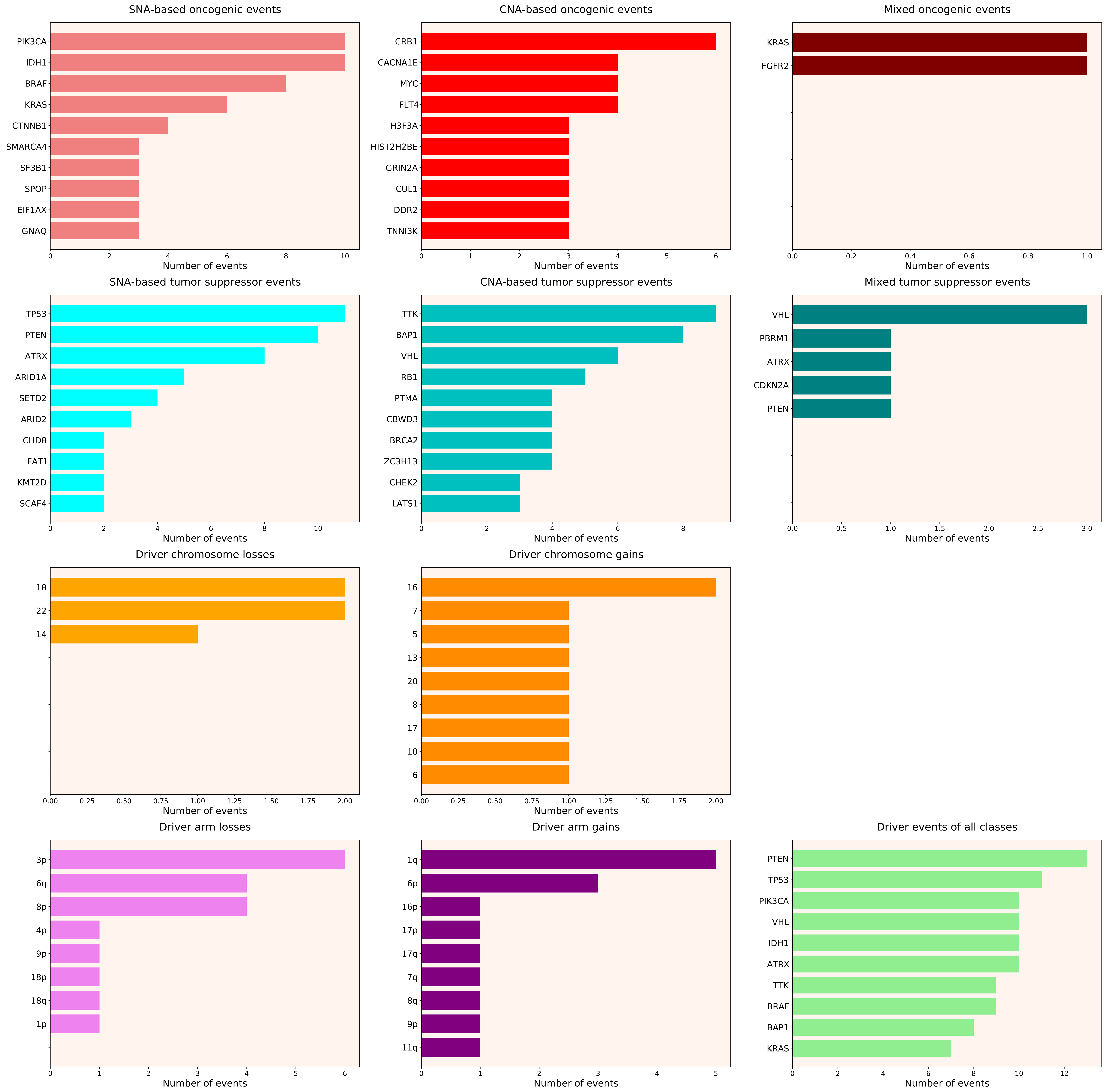

### 2021_8_16_20_32_distribution_events_detailed_7.pdf

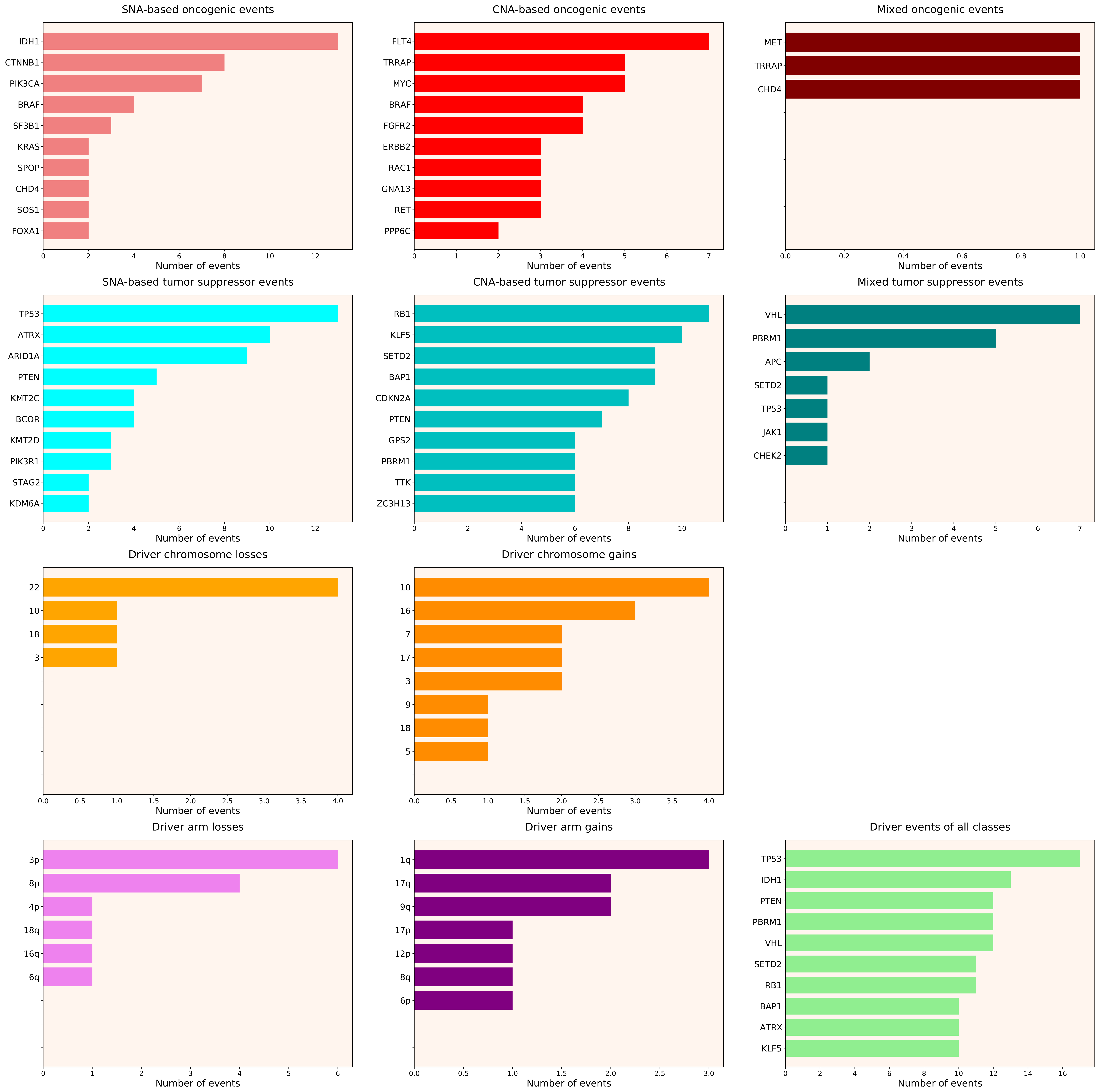

### 2021_8_16_20_32_distribution_events_detailed_10.pdf

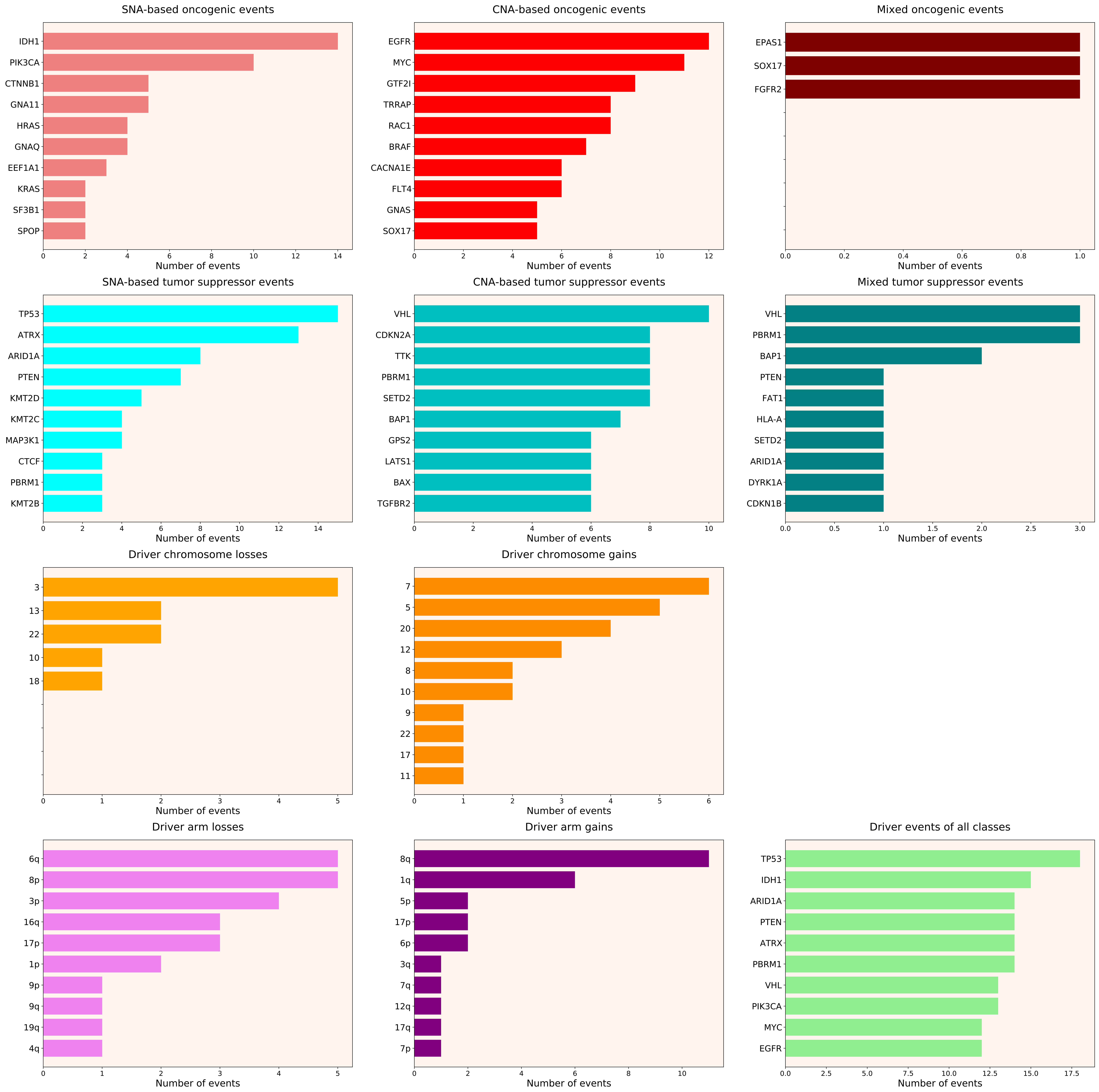

### 2021_8_16_20_32_distribution_events_detailed_11.pdf

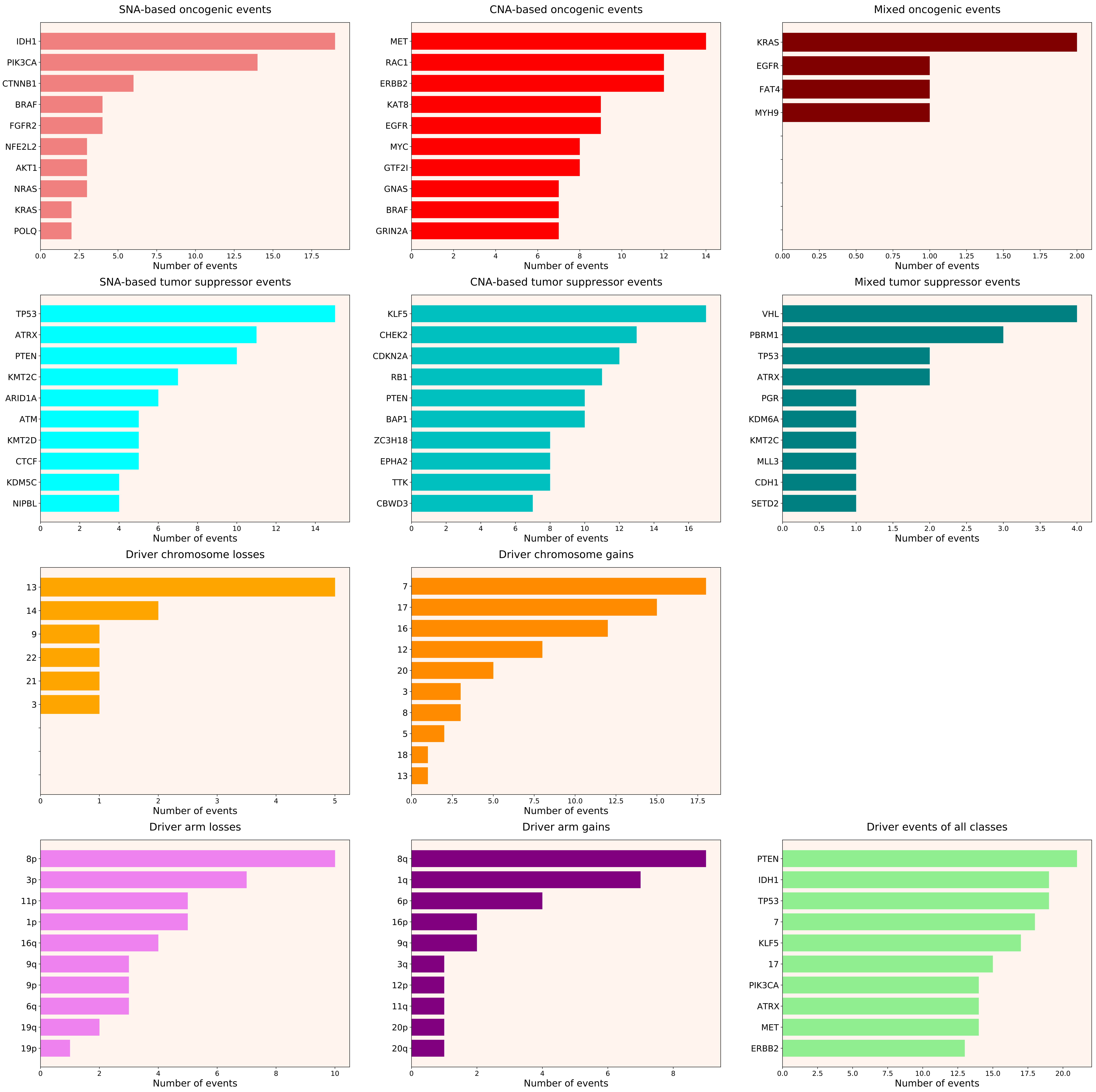

### 2021_8_16_20_32_distribution_events_detailed_14.pdf

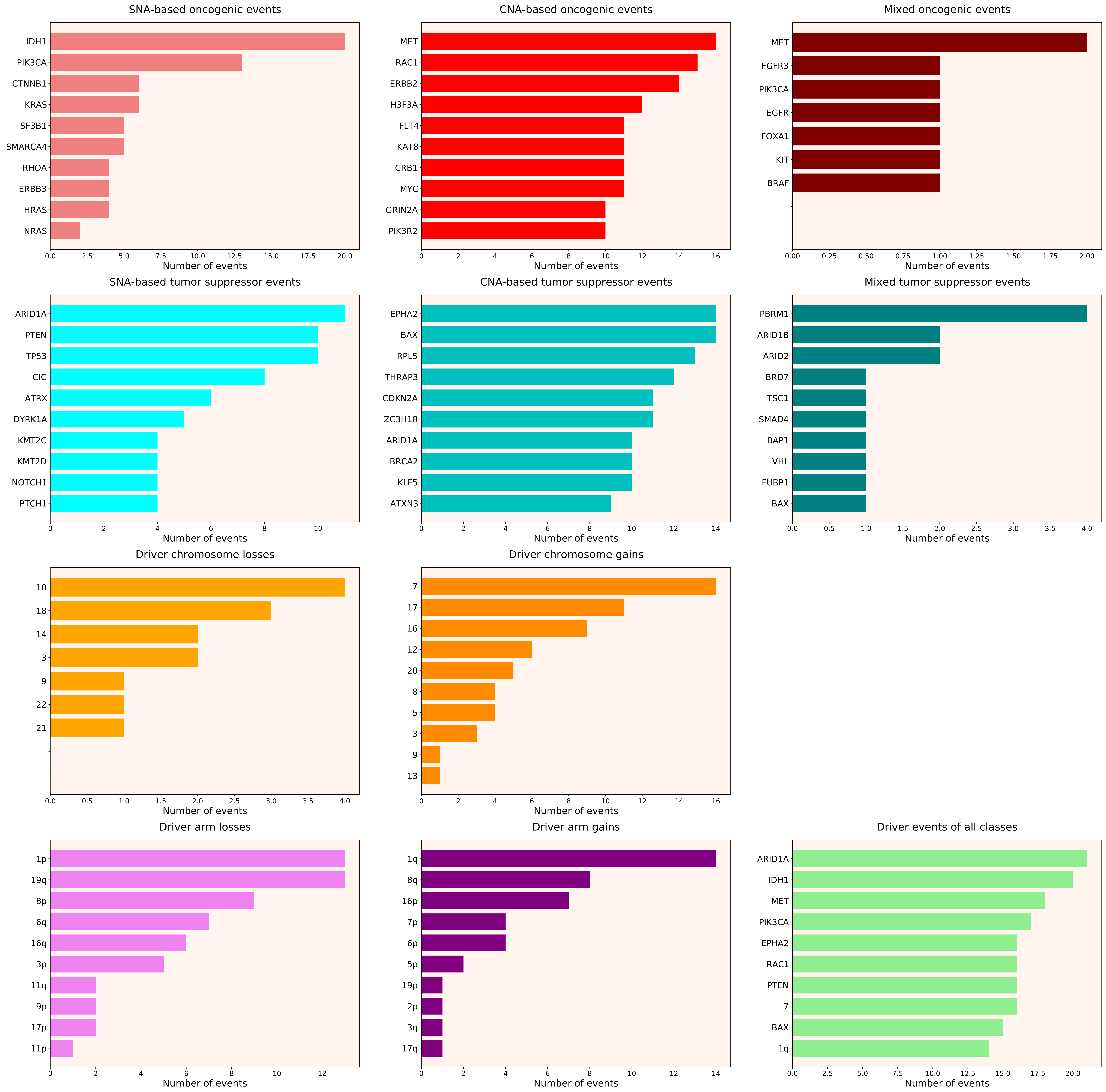

### 2021_8_16_20_32_distribution_events_detailed_15.pdf

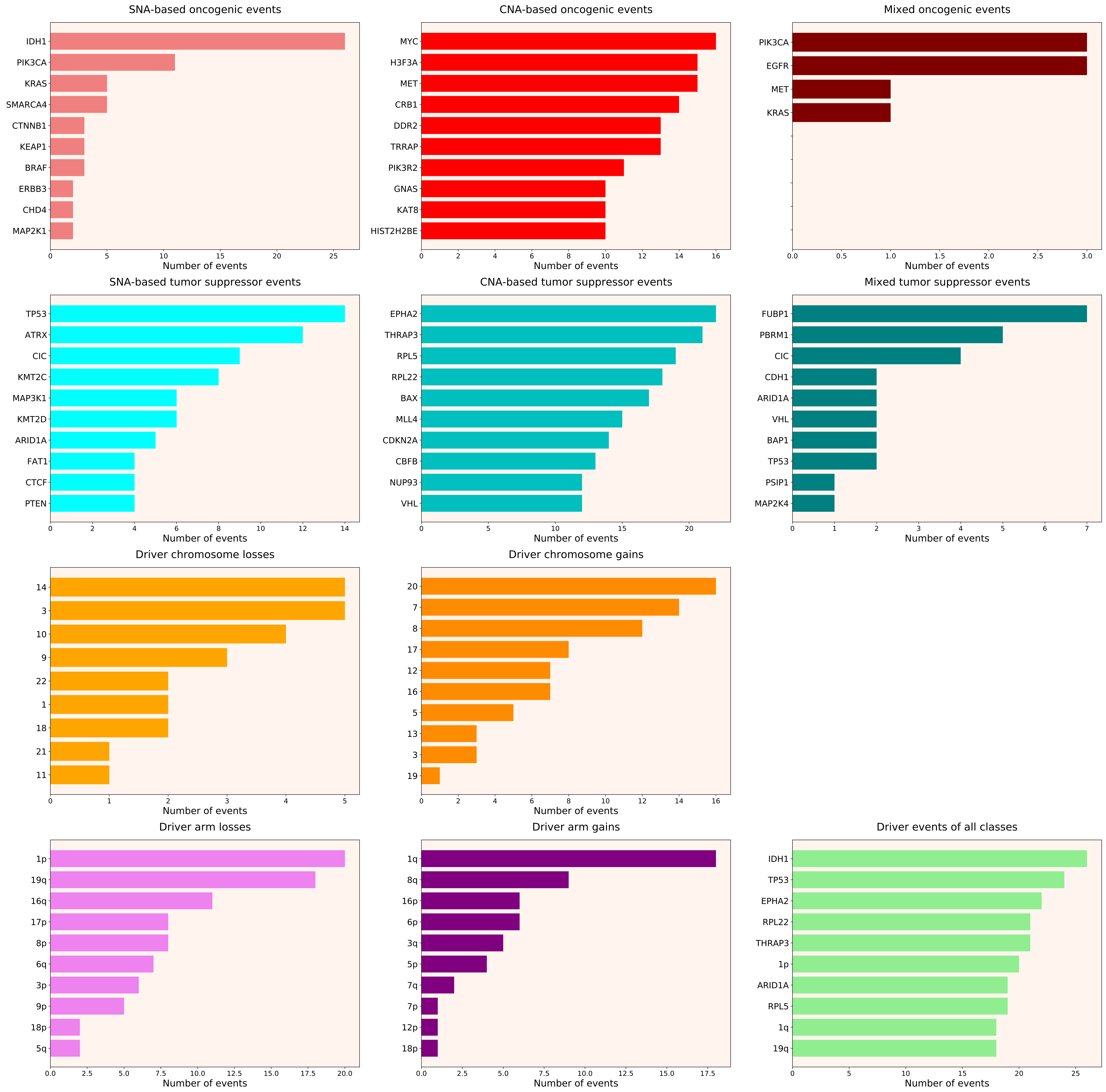

### 2021_8_16_20_32_distribution_events_detailed_16.pdf

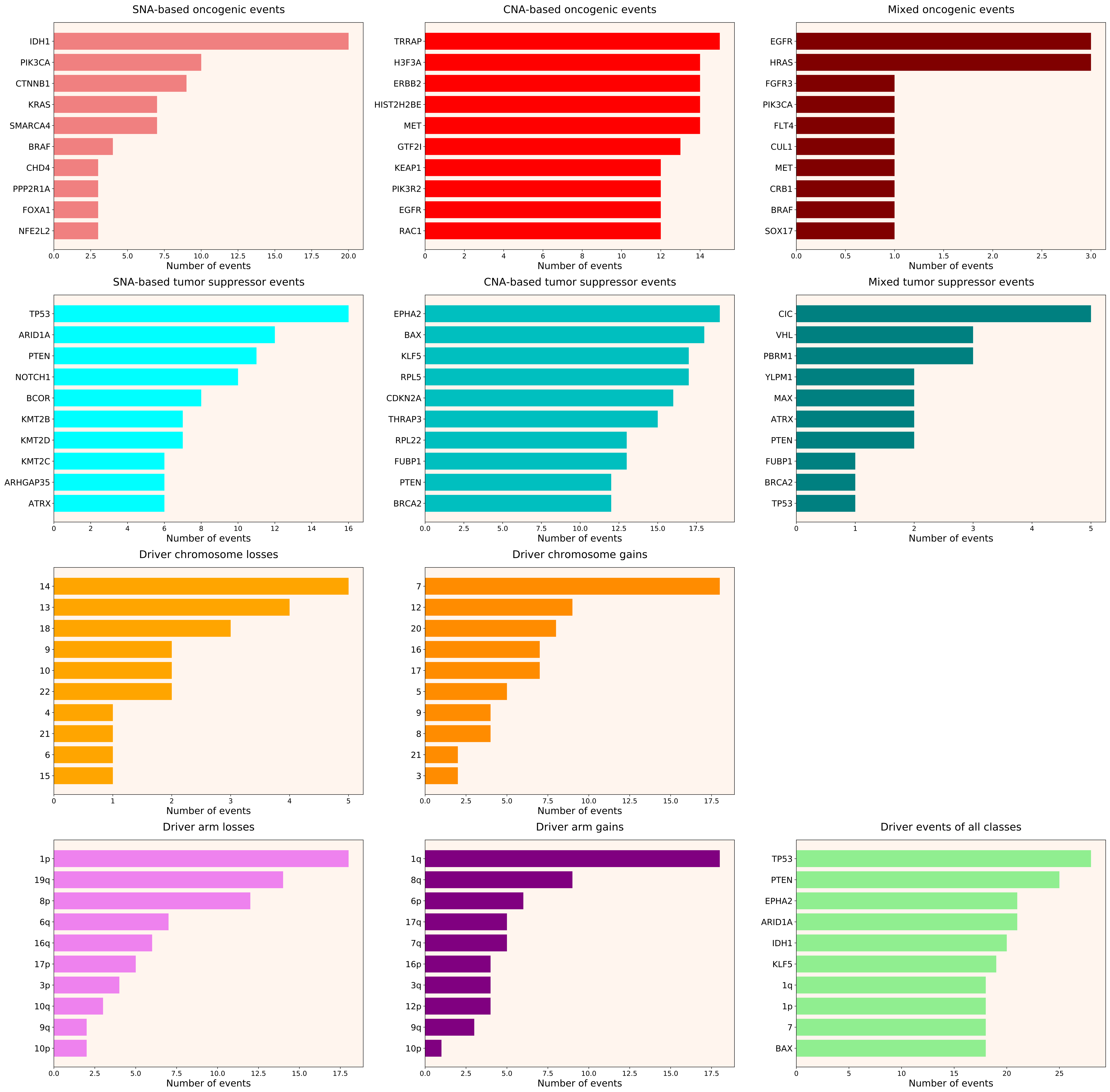

### 2021_8_16_20_32_distribution_events_detailed_17.pdf

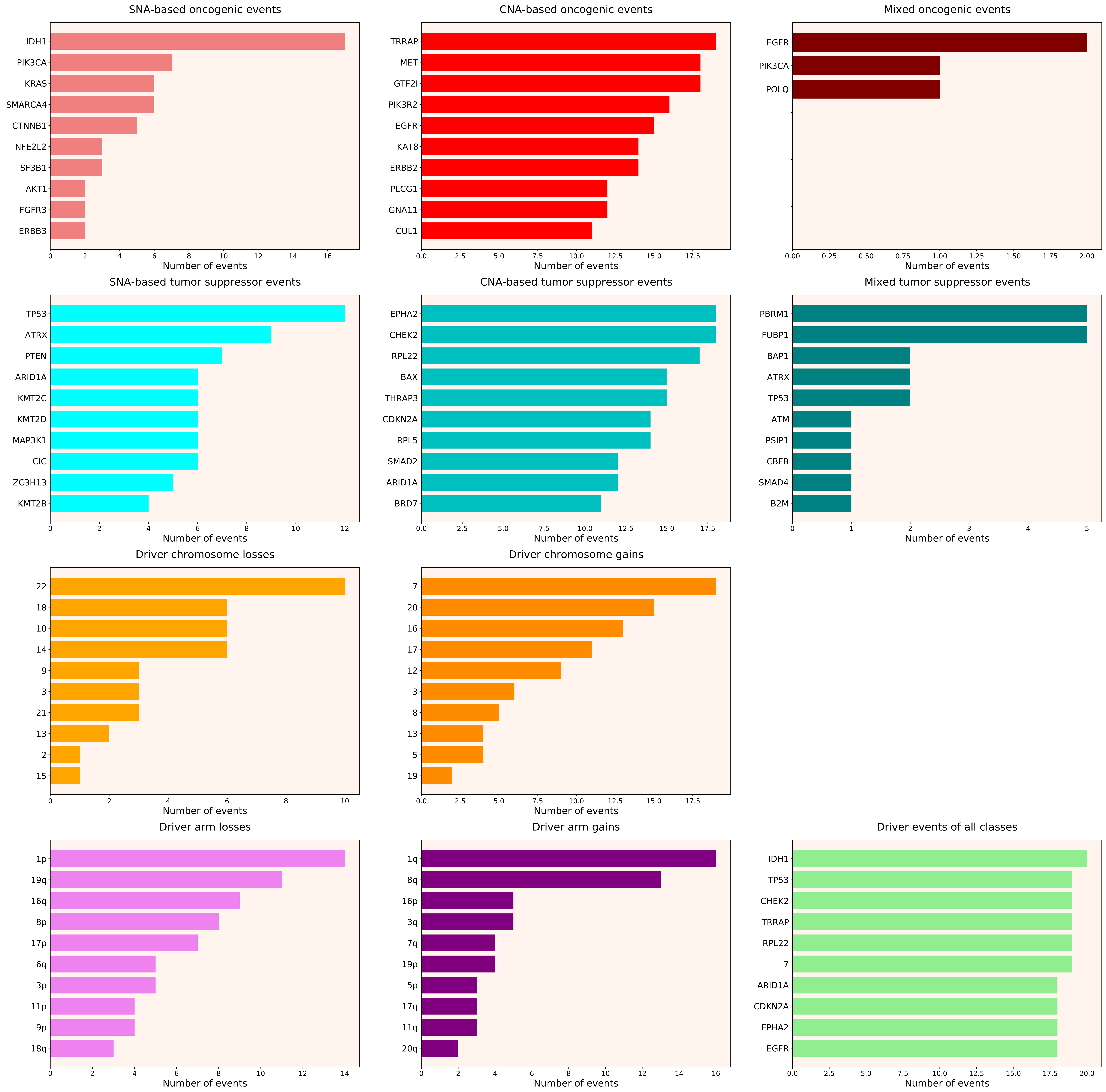

### 2021_8_16_20_32_distribution_events_detailed_28.pdf

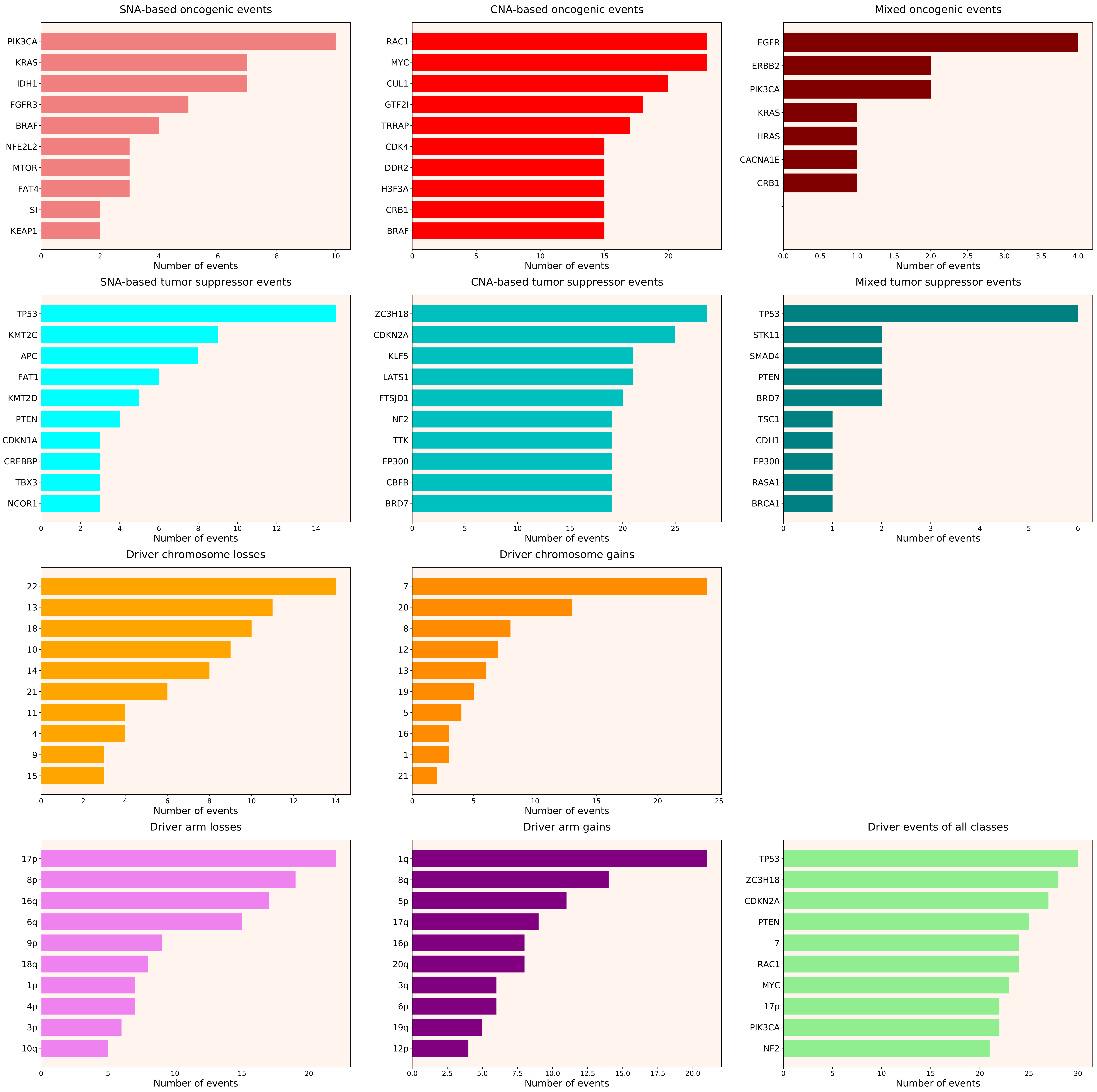

### 2021_8_16_20_32_distribution_events_detailed_29.pdf

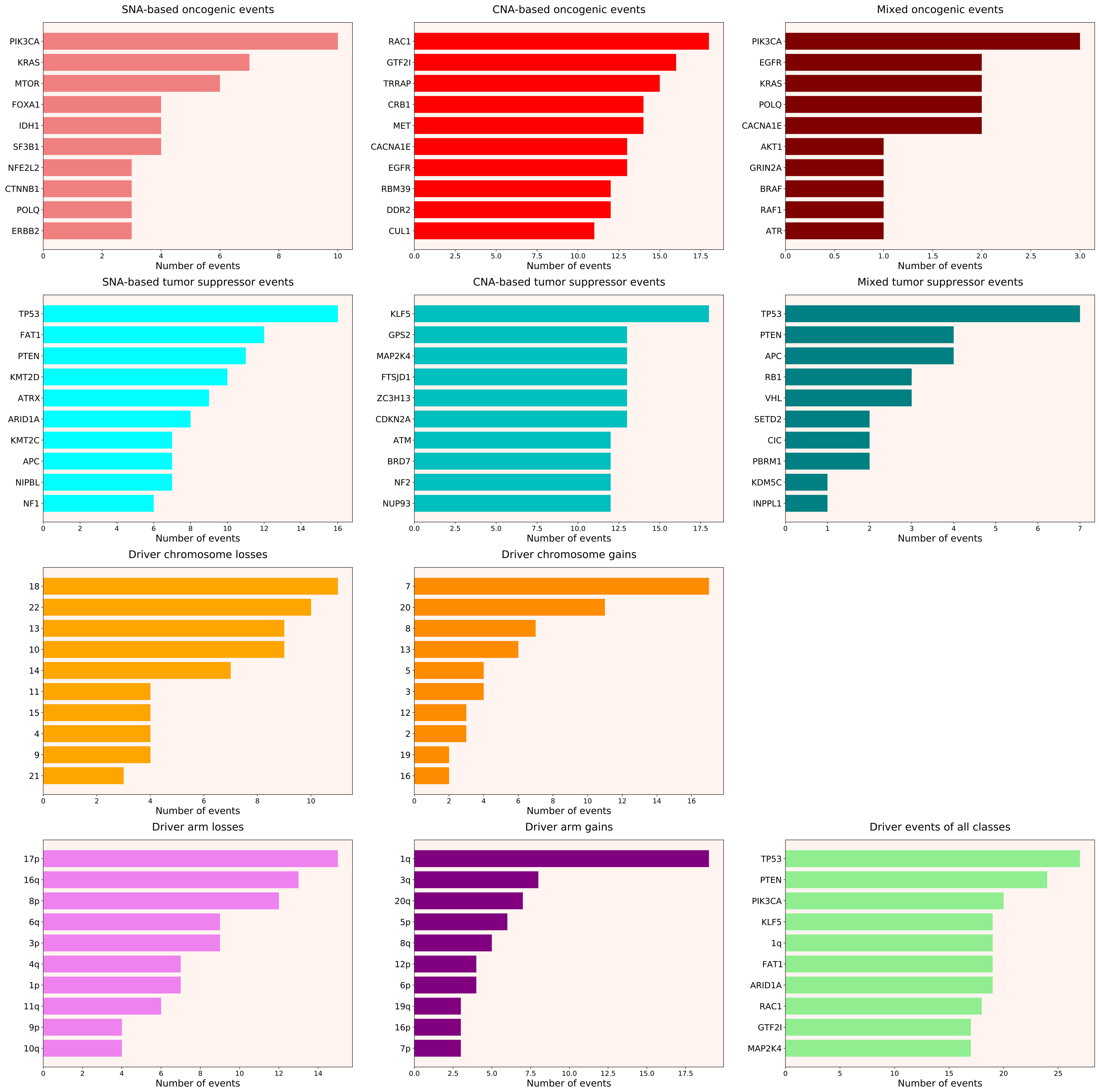

### 2021_8_16_20_32_distribution_events_detailed_38.pdf

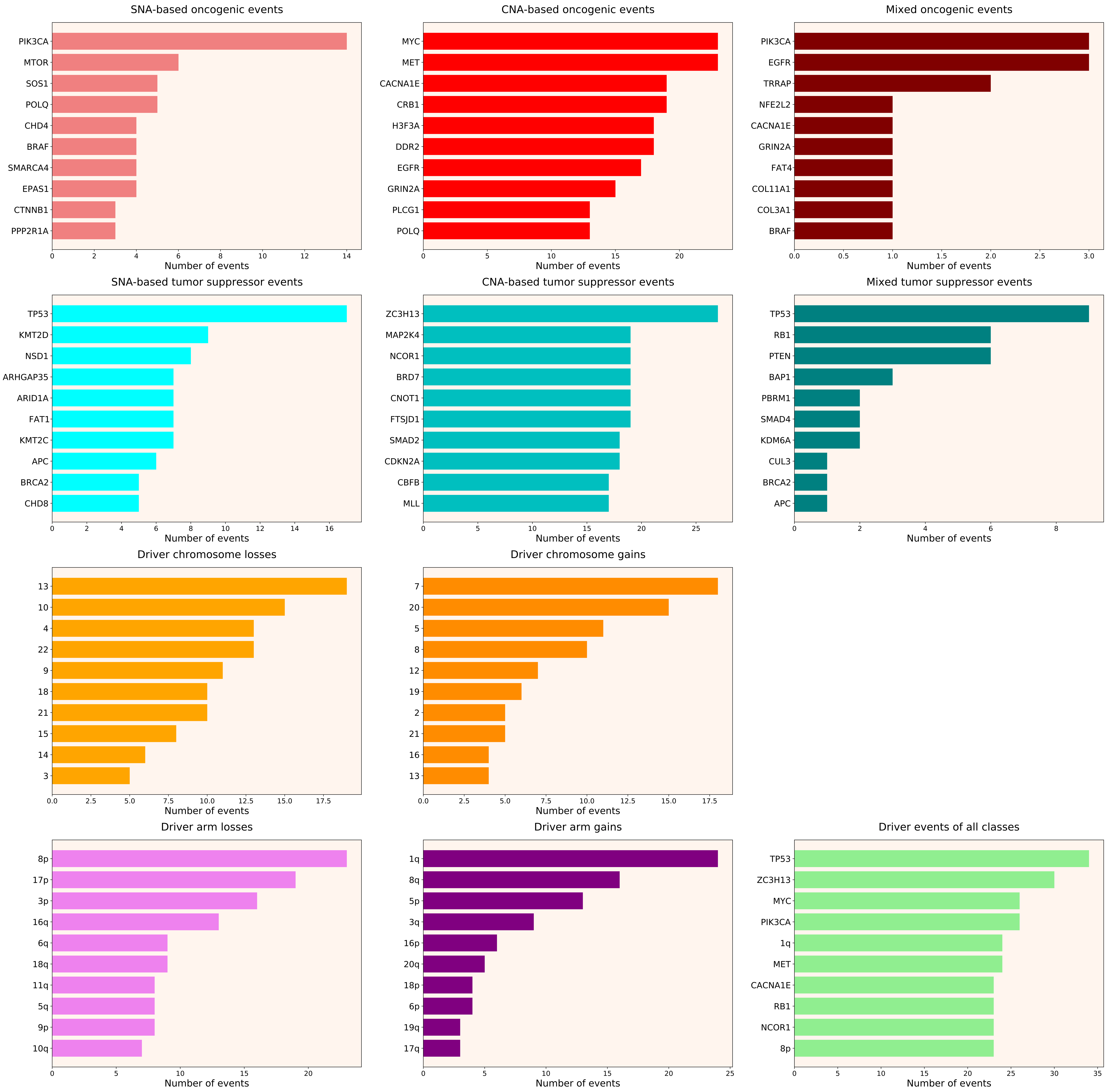

### 2021_8_16_20_32_distribution_events_detailed_39.pdf

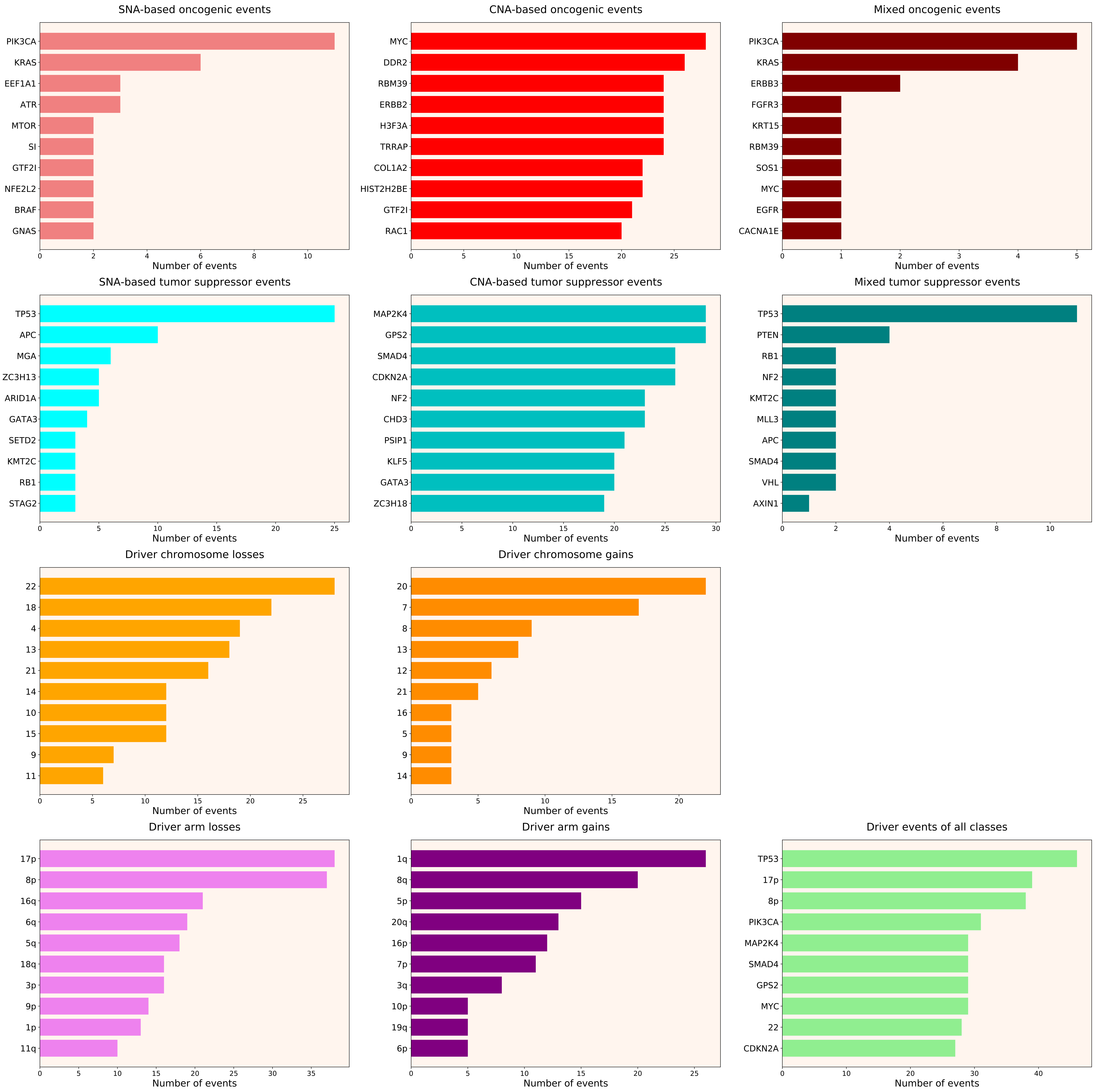

### 2021_8_16_20_32_distribution_events_detailed_DSI.pdf

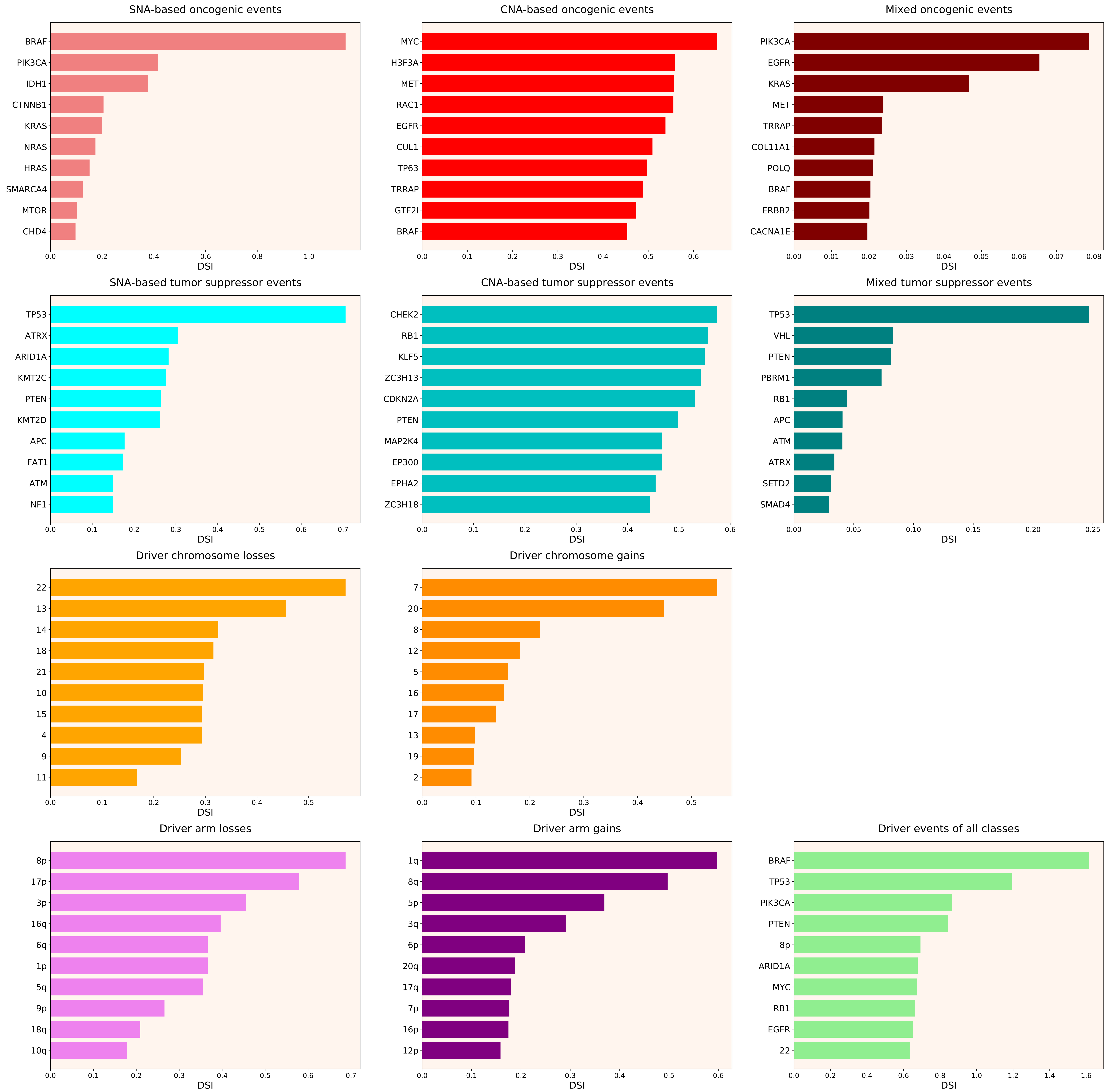

### 2021_8_16_20_32_distribution_events_detailed_DSI_females.pdf

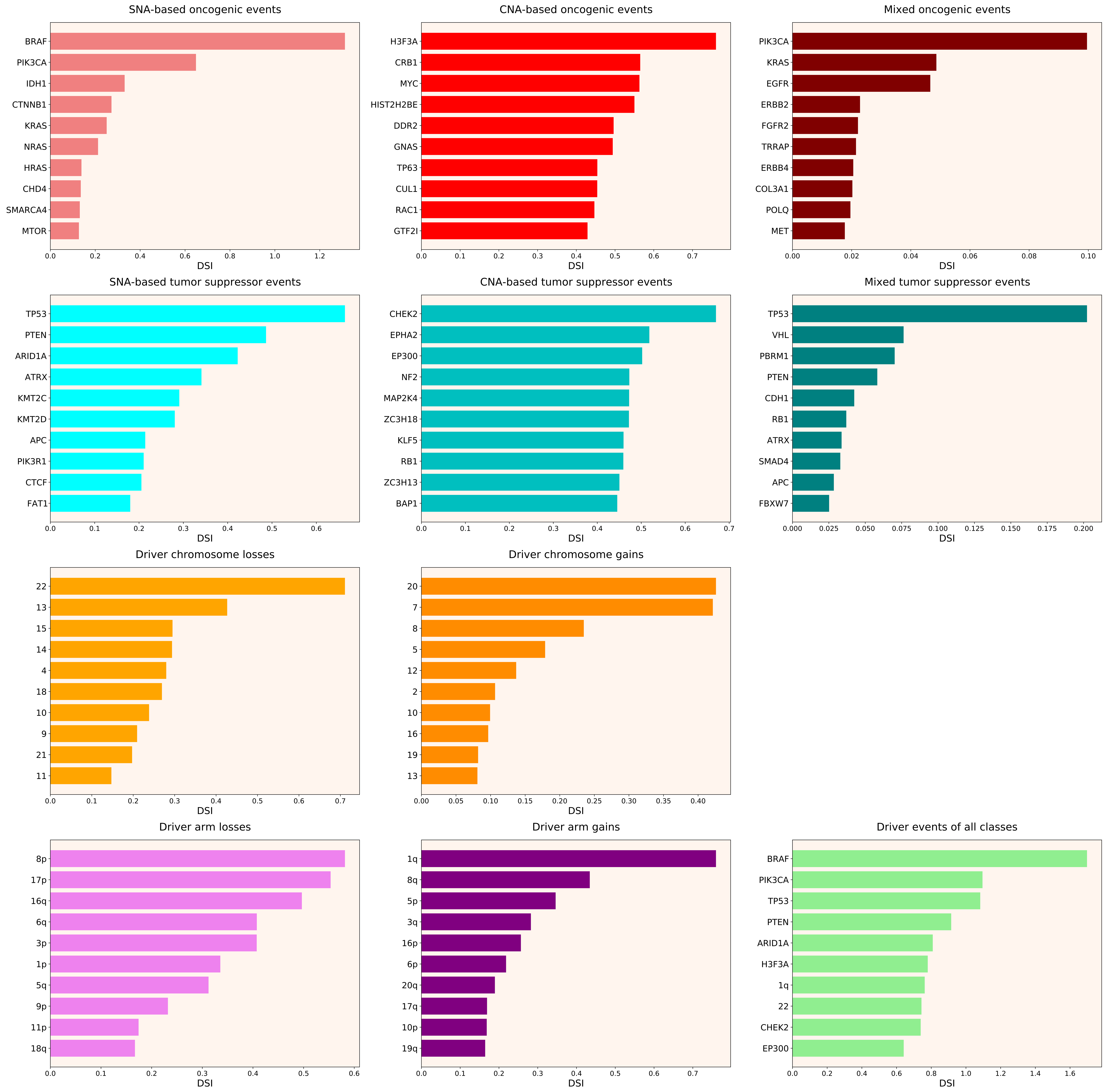

### 2021_8_16_20_32_distribution_events_detailed_DSI_males.pdf

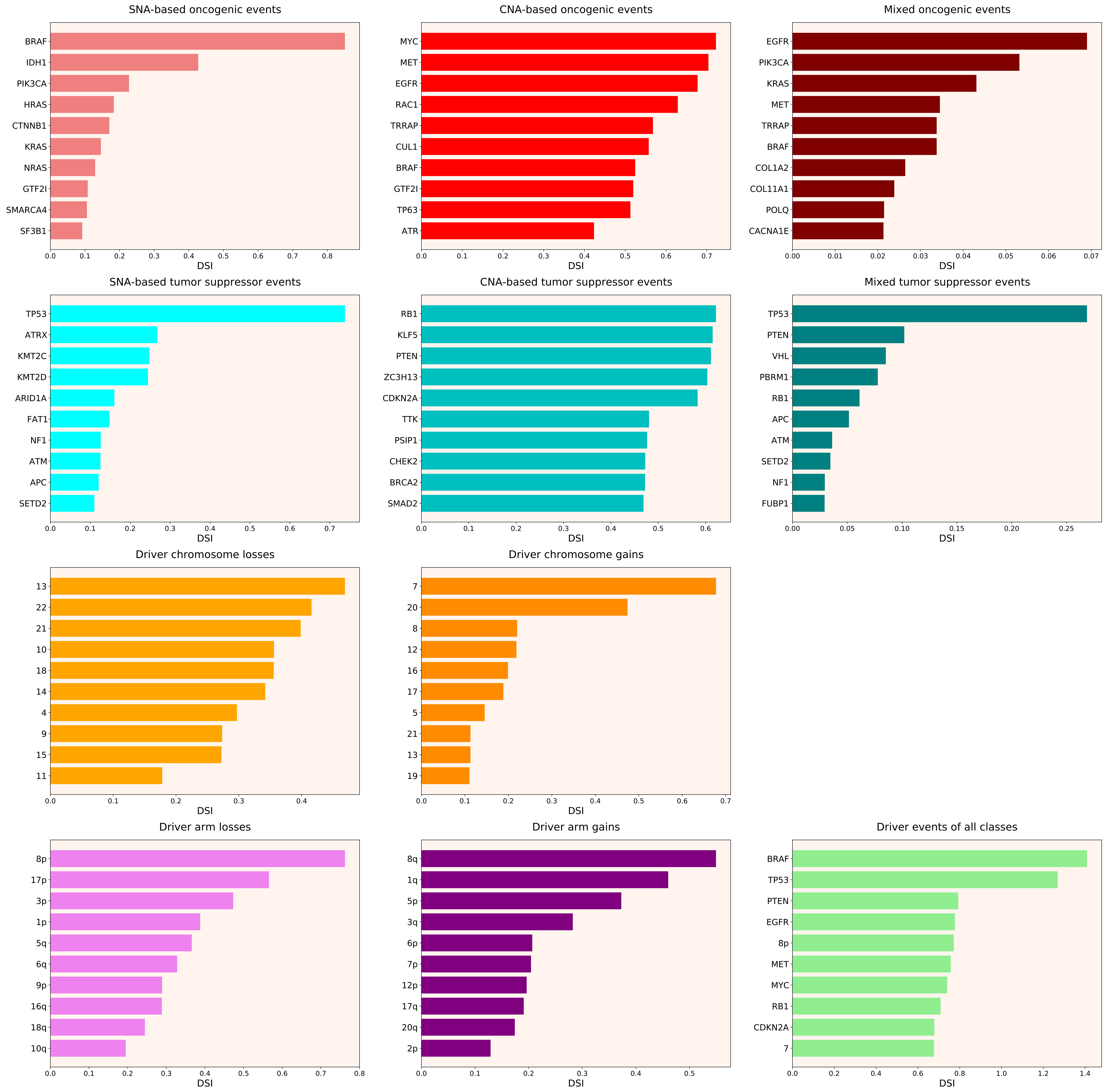

### 2021_8_16_20_32_distribution_events_detailed_females_11.pdf

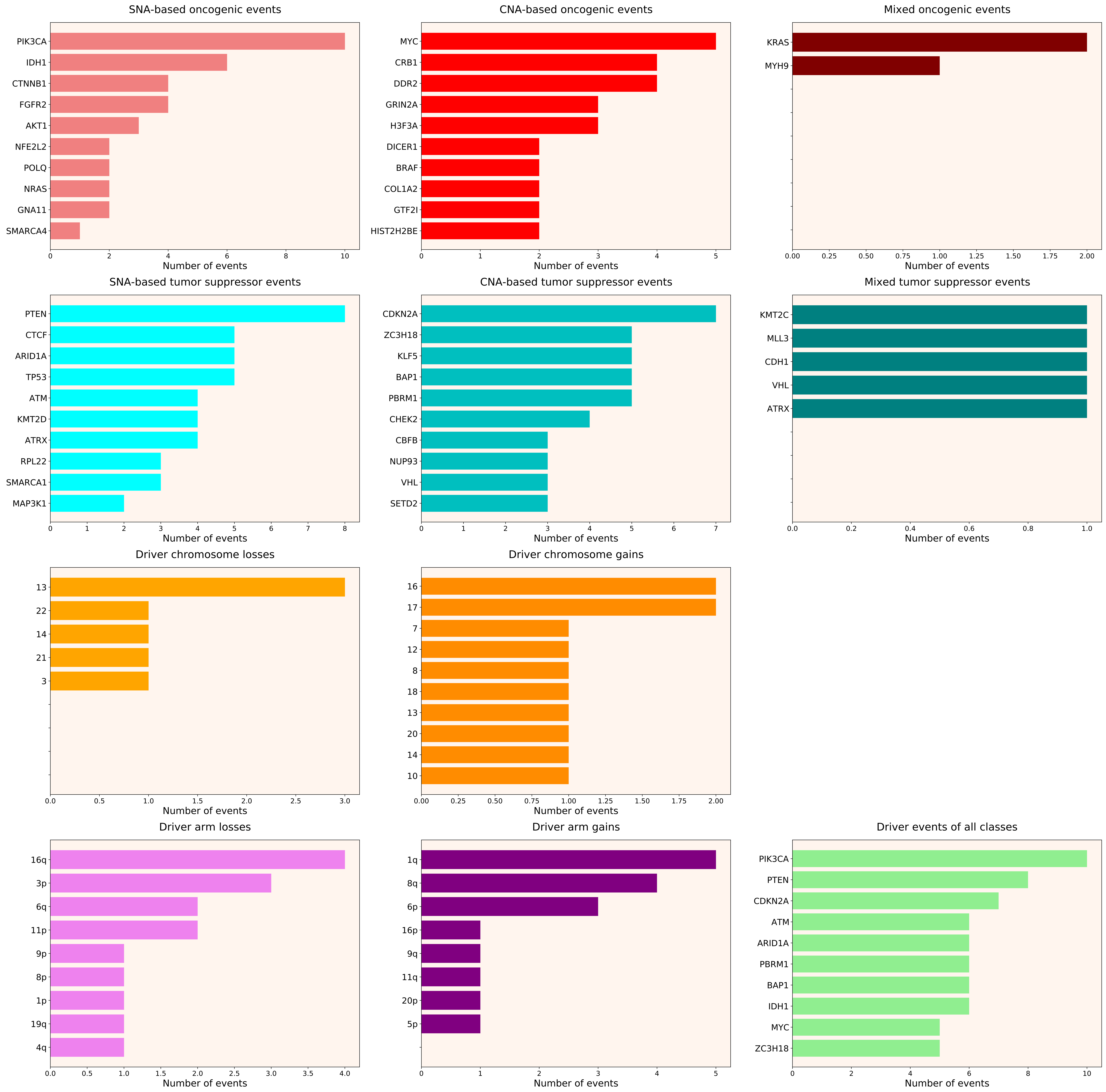

### 2021_8_16_20_32_distribution_events_detailed_females_12.pdf

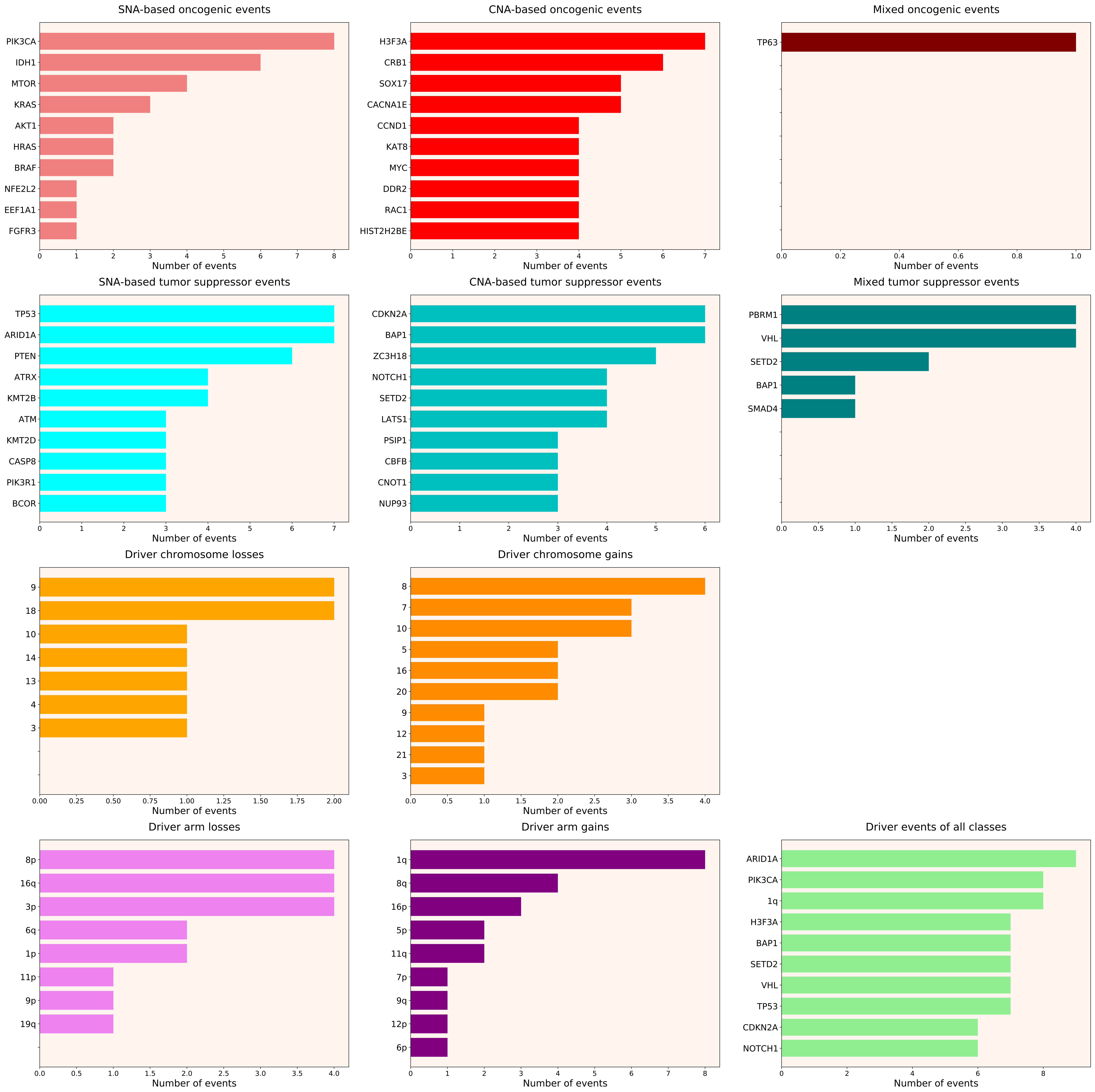

### 2021_8_16_20_32_distribution_events_detailed_females_13.pdf

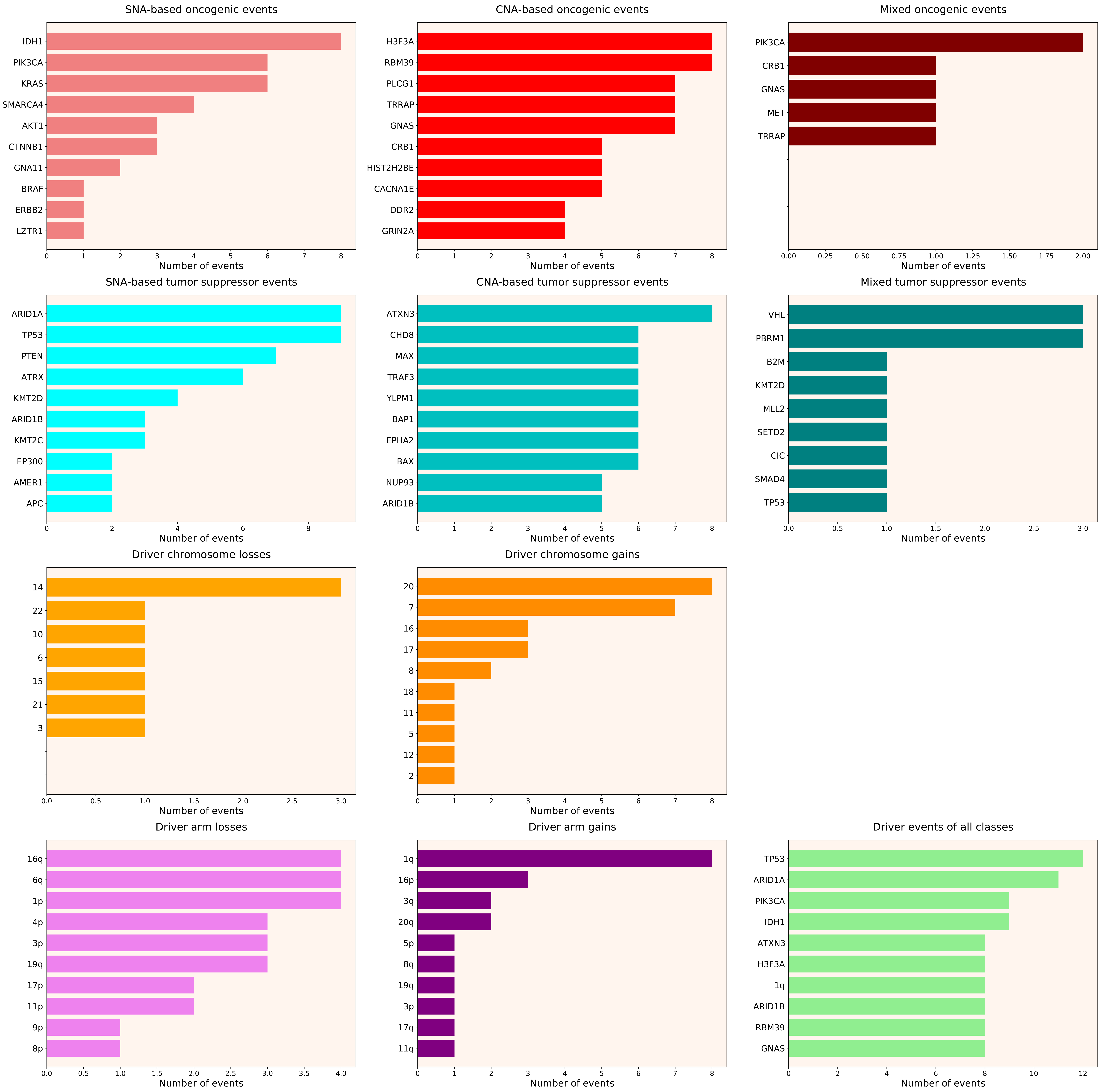

### 2021_8_16_20_32_distribution_events_detailed_females_14.pdf

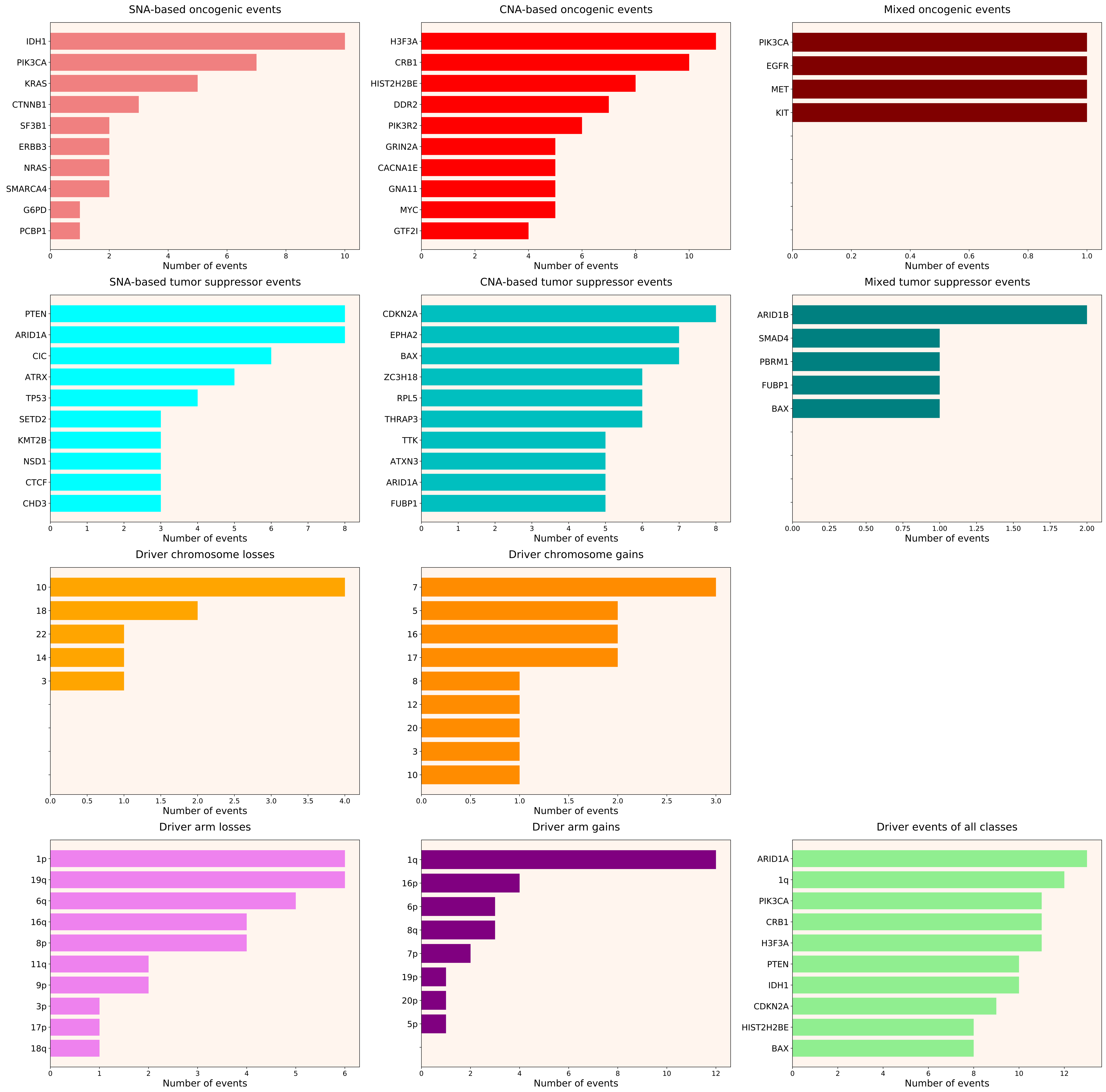

### 2021_8_16_20_32_distribution_events_detailed_females_15.pdf

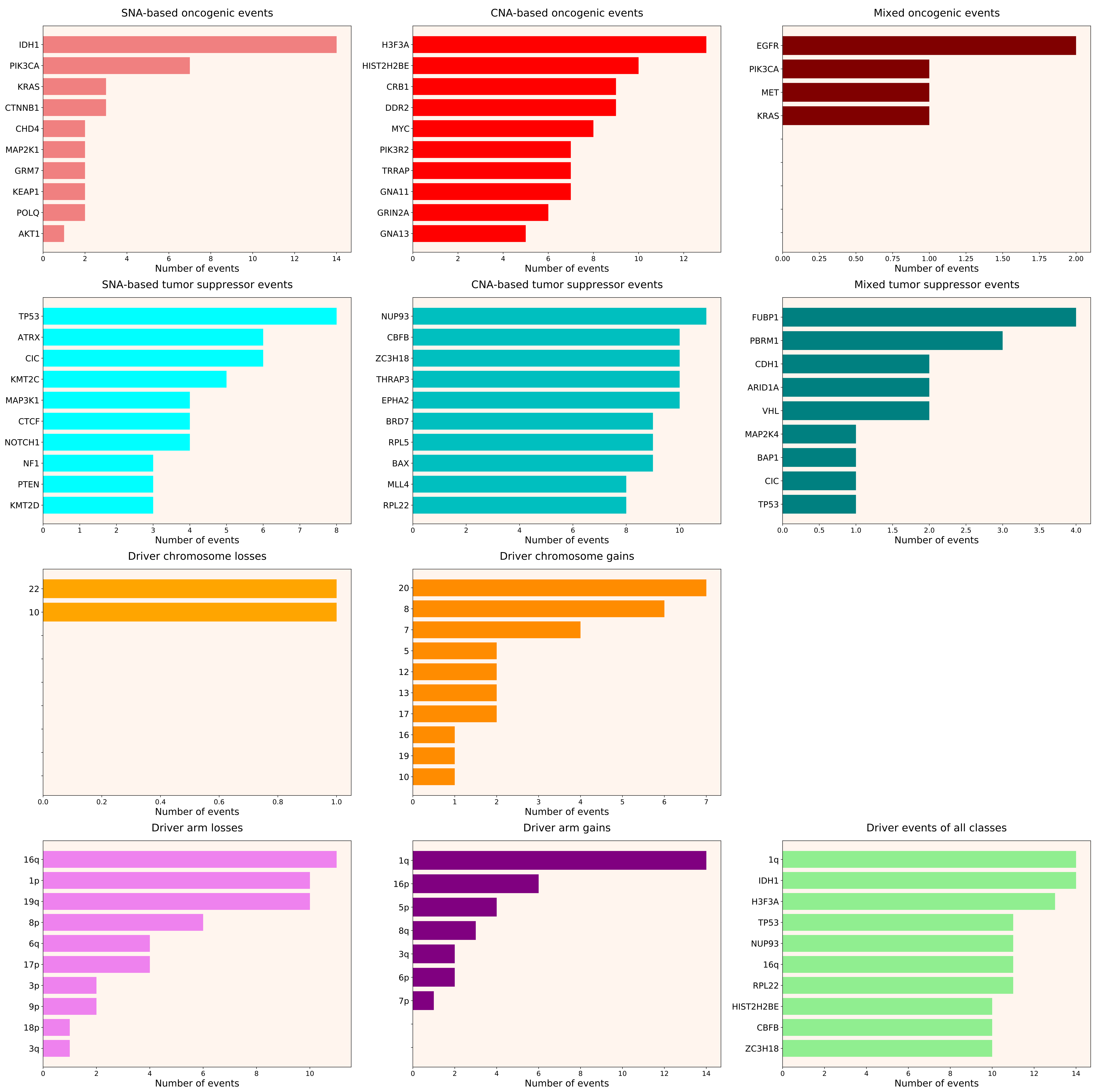

### 2021_8_16_20_32_distribution_events_detailed_females_16.pdf

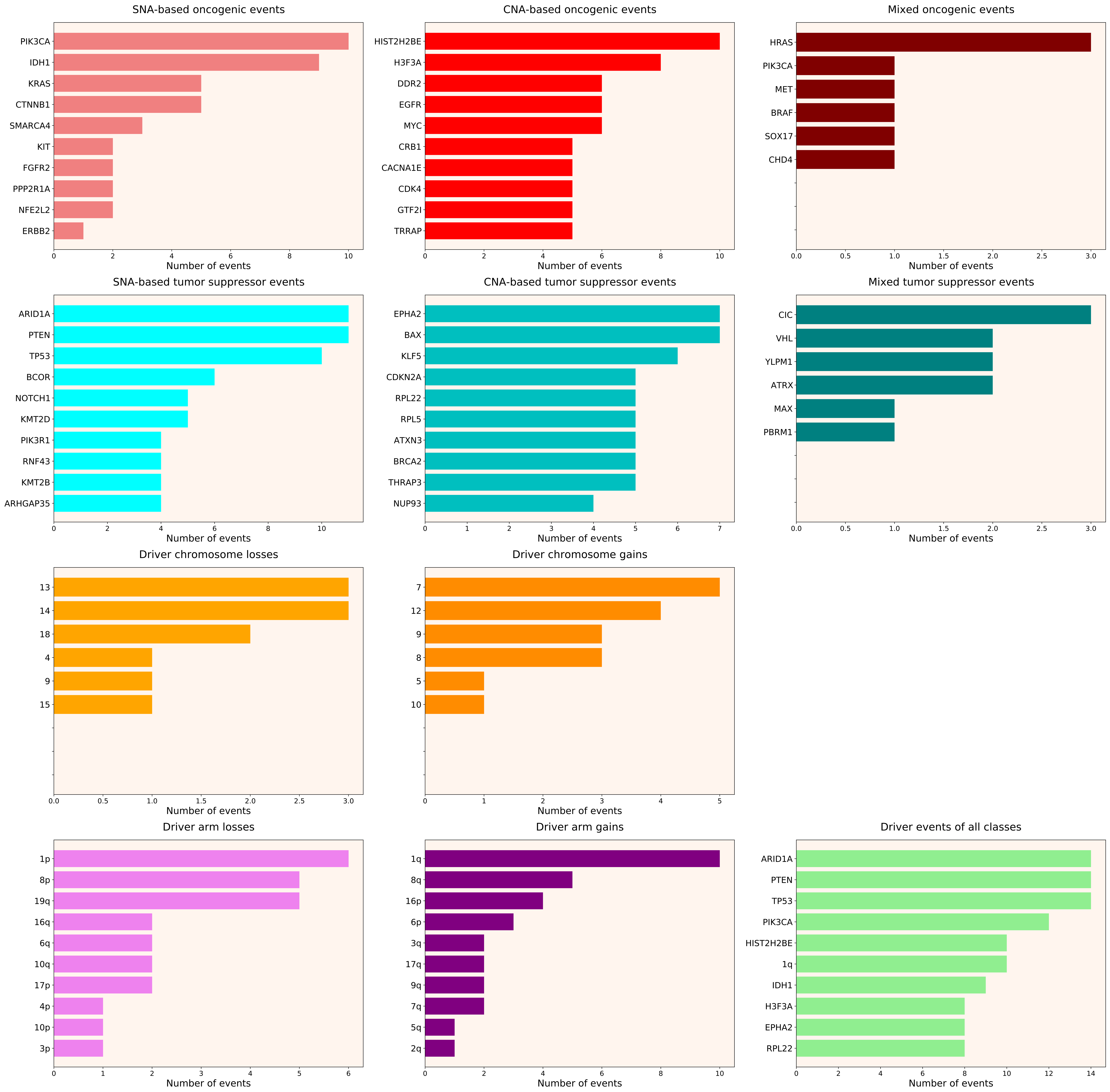

### 2021_8_16_20_32_distribution_events_detailed_females_17.pdf

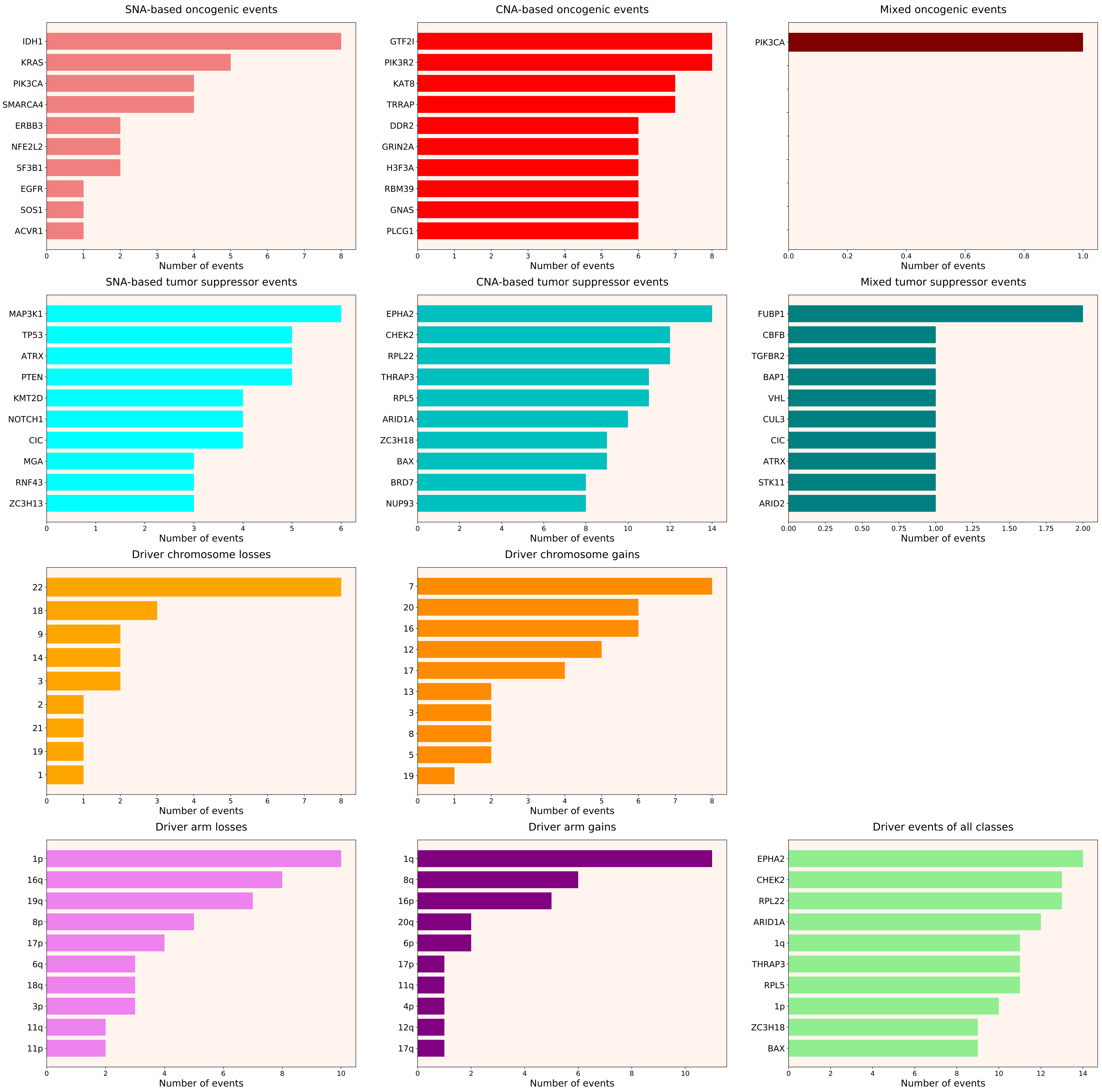

### 2021_8_16_20_32_distribution_events_detailed_females_28.pdf

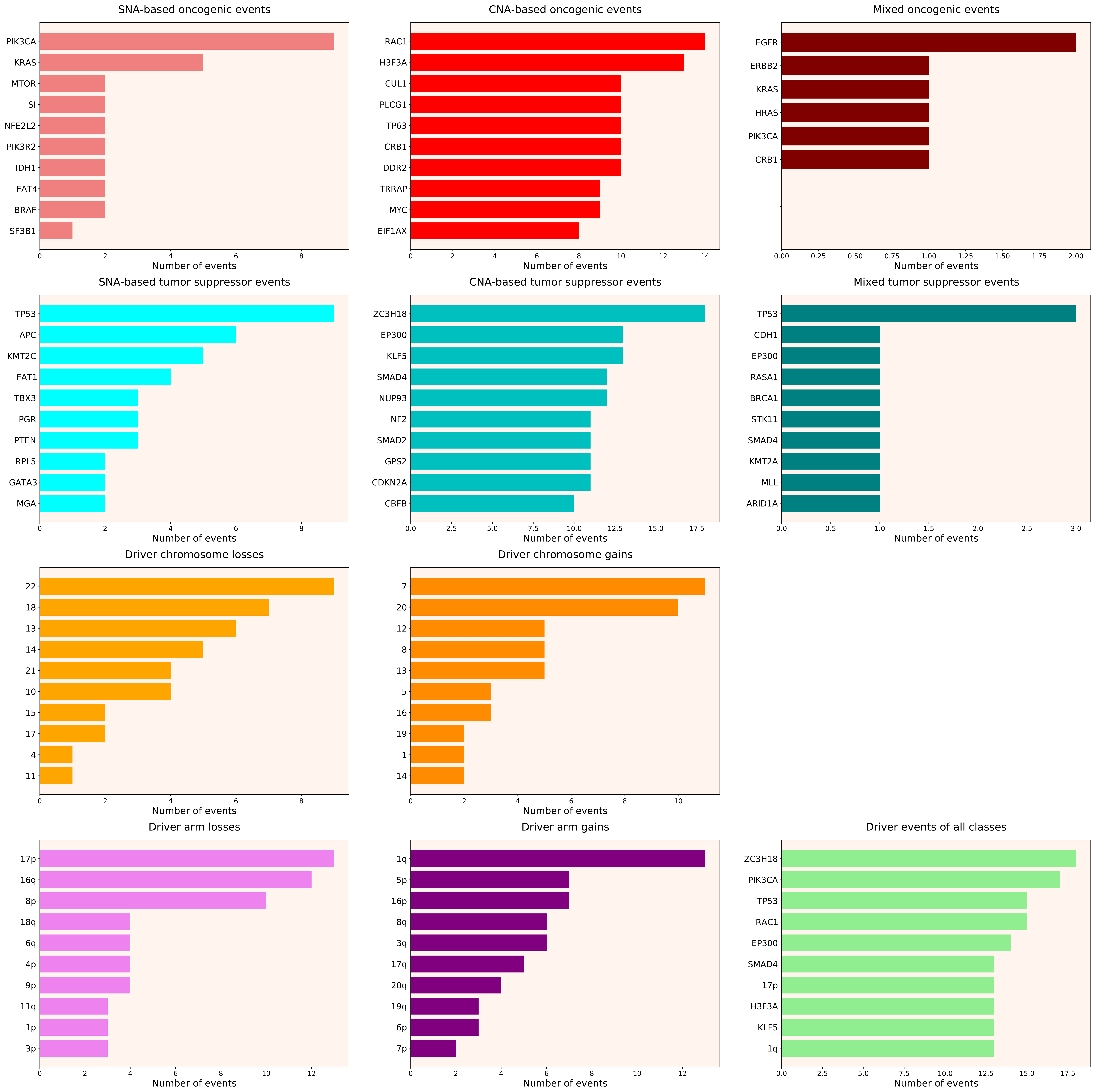

### 2021_8_16_20_32_distribution_events_detailed_females_29.pdf

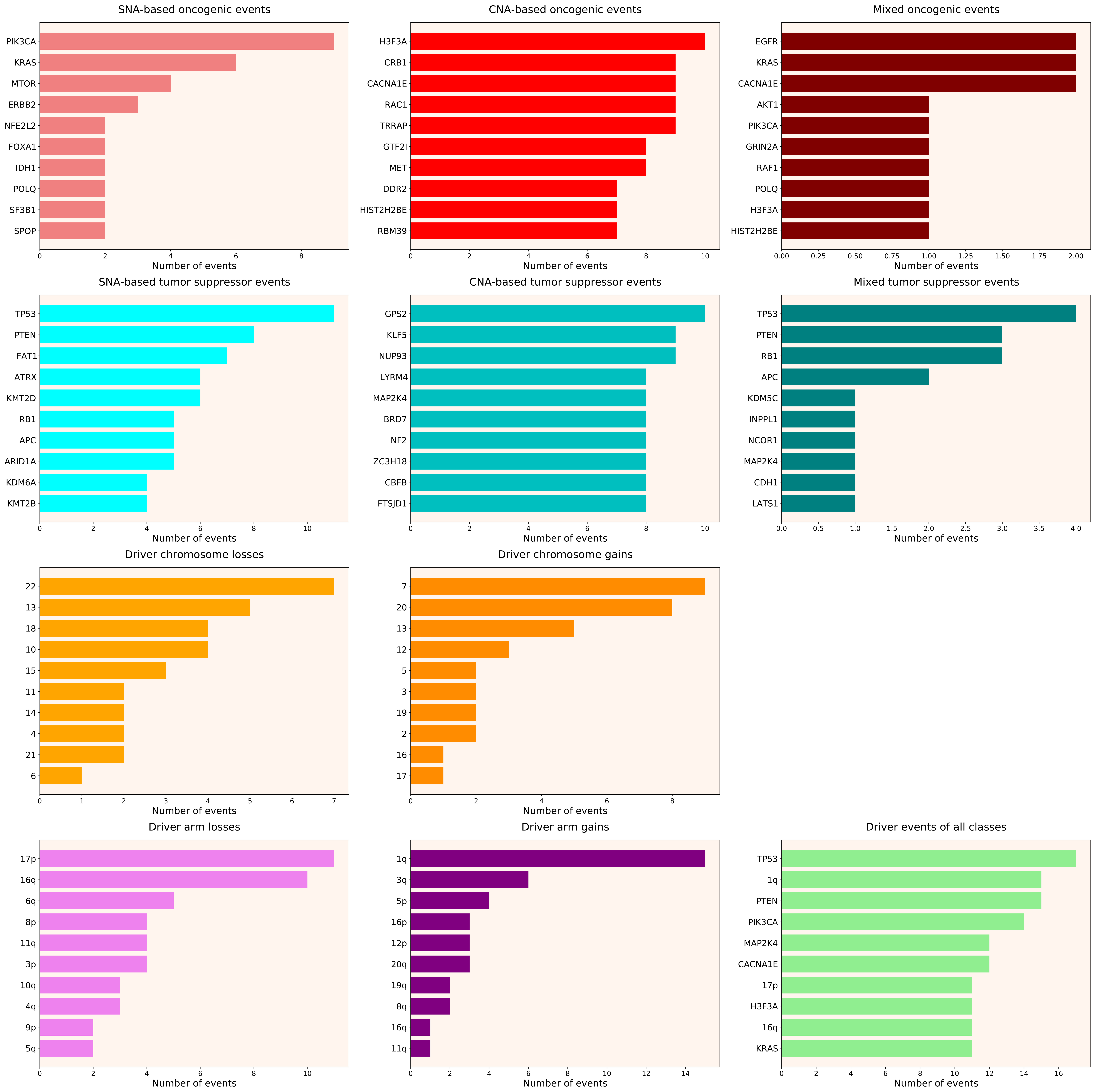

### 2021_8_16_20_32_distribution_events_detailed_females_39.pdf

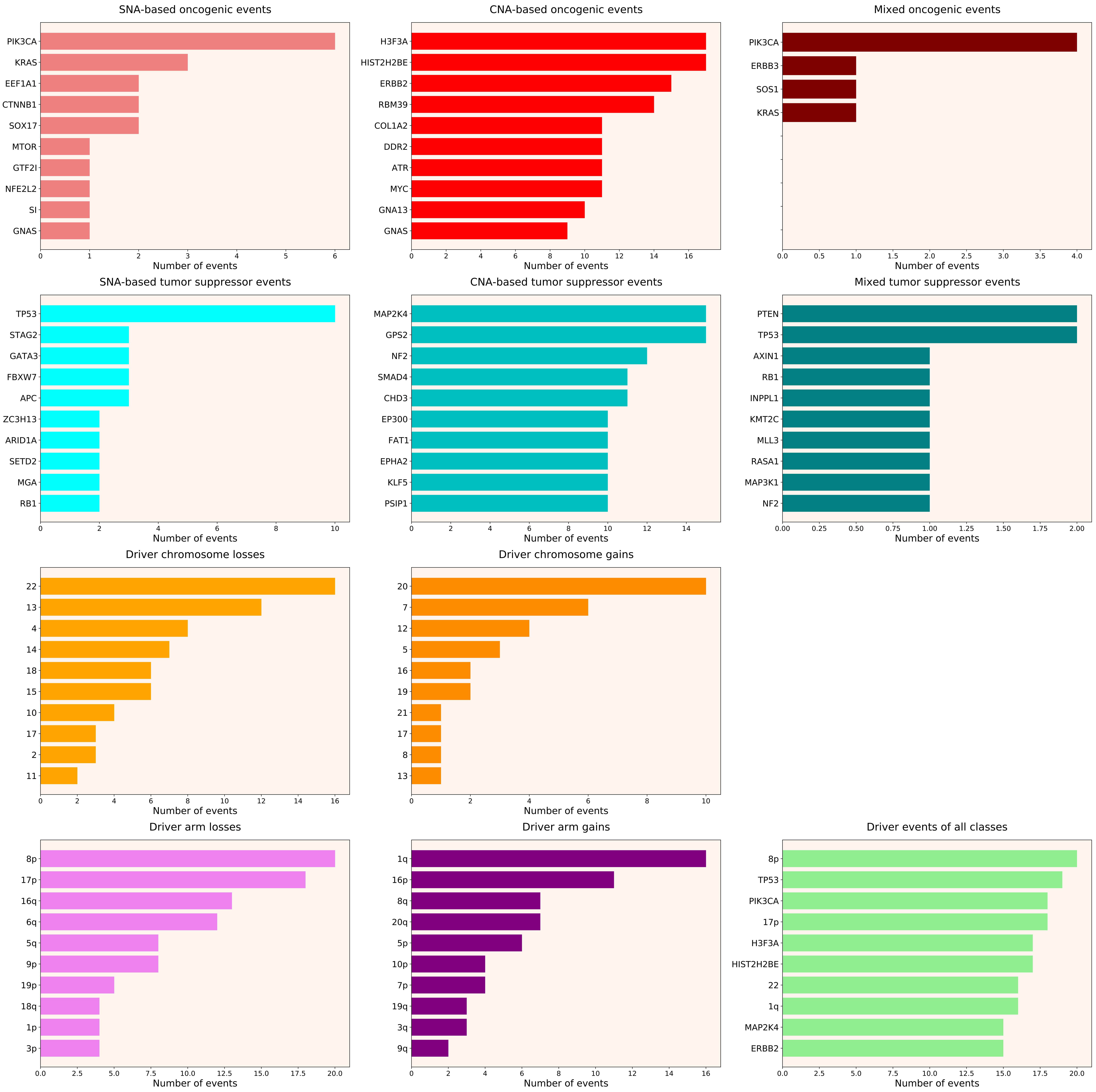

### 2021_8_16_20_32_distribution_events_detailed_males_1.pdf

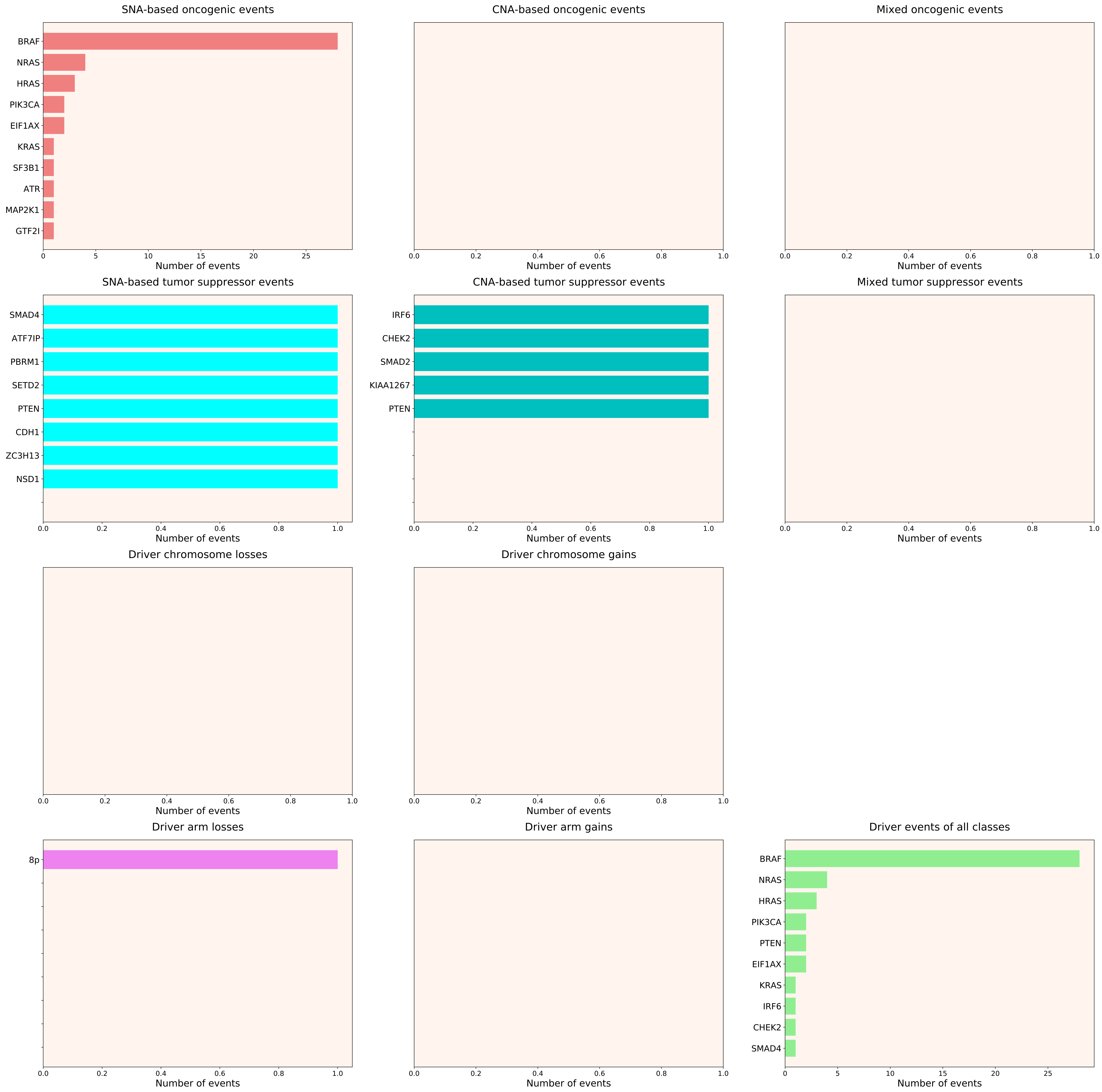

### 2021_8_16_20_32_distribution_events_detailed_males_4.pdf

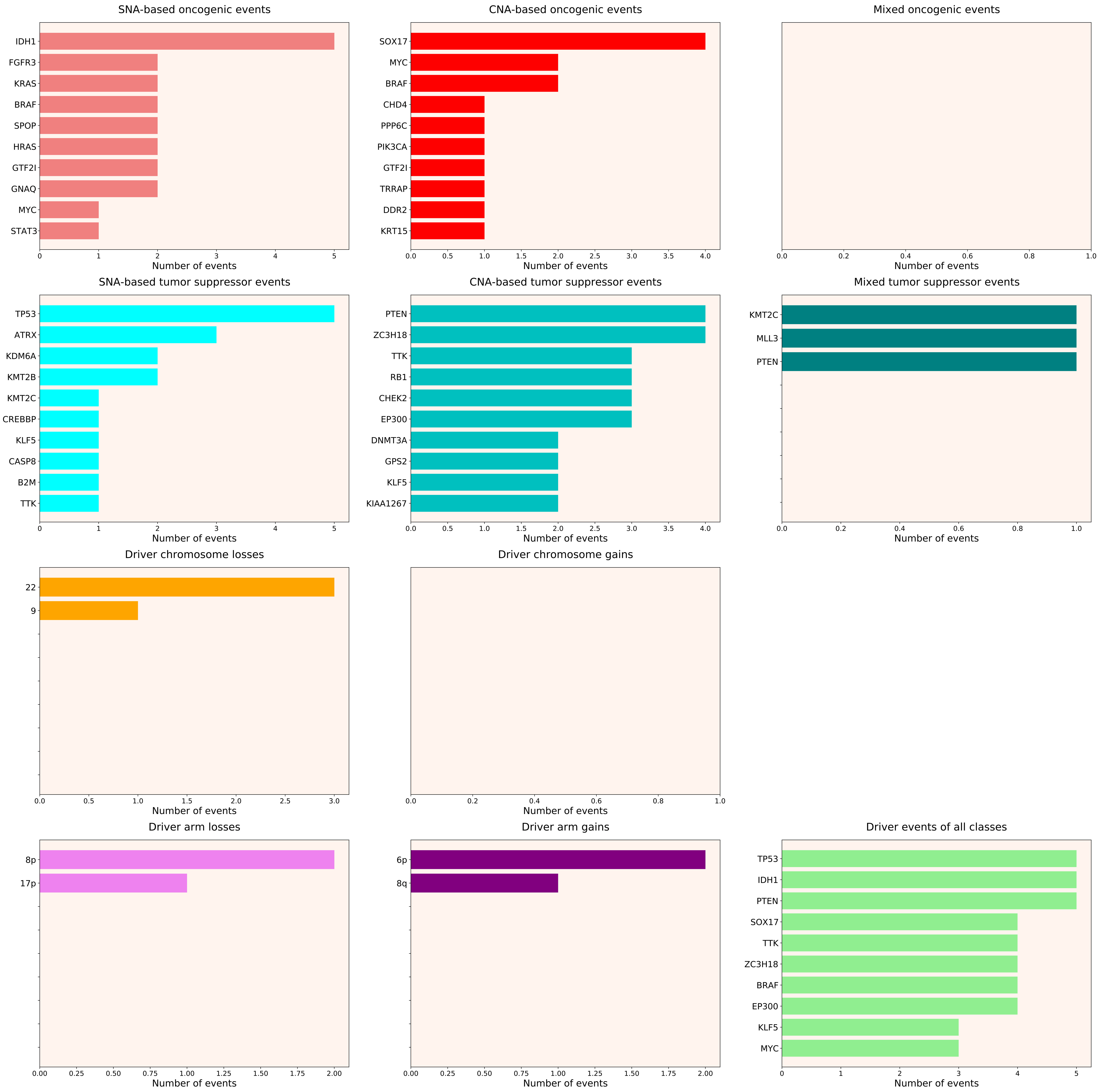

### 2021_8_16_20_32_distribution_events_detailed_males_5.pdf

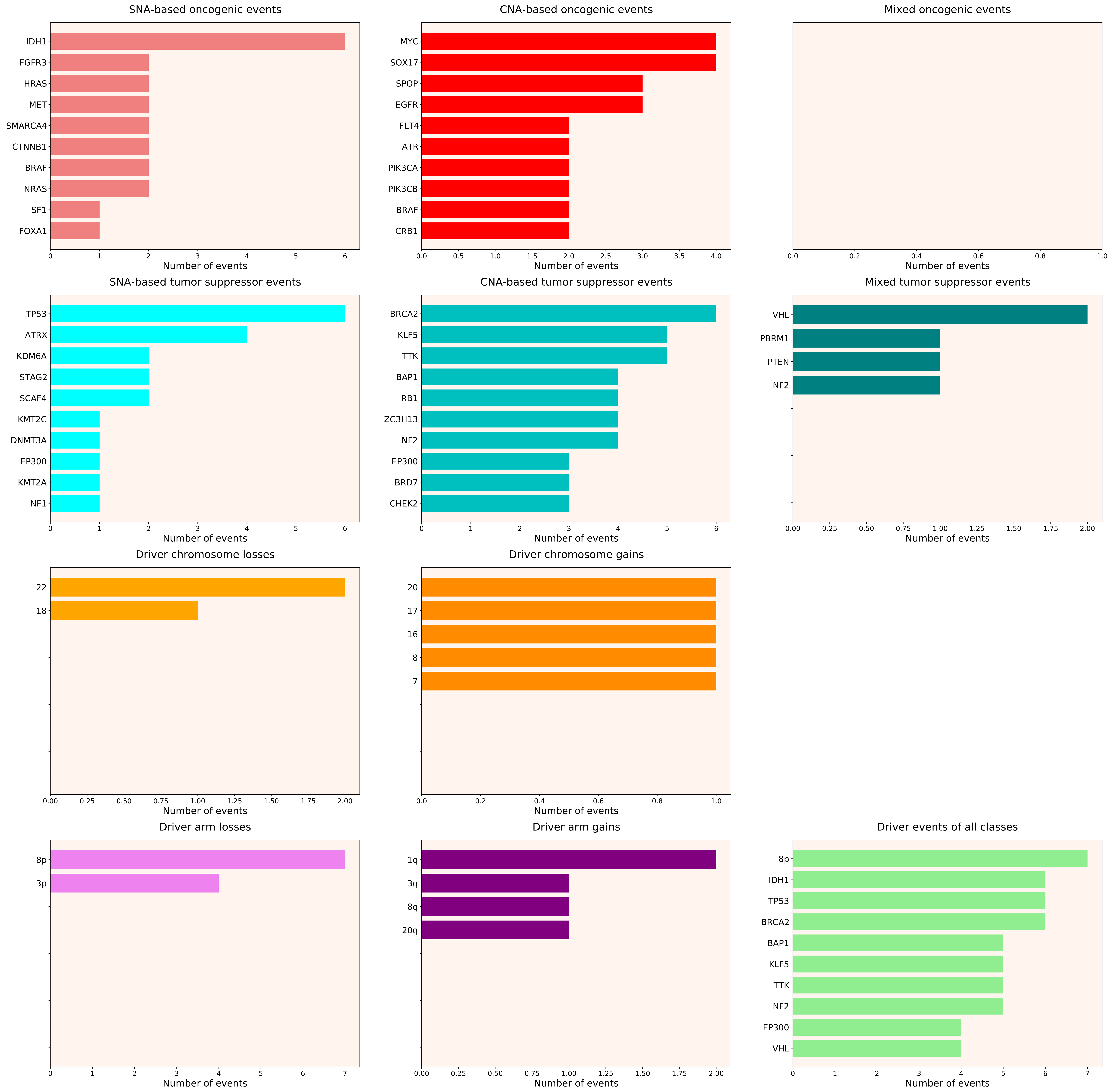

### 2021_8_16_20_32_distribution_events_detailed_males_6.pdf

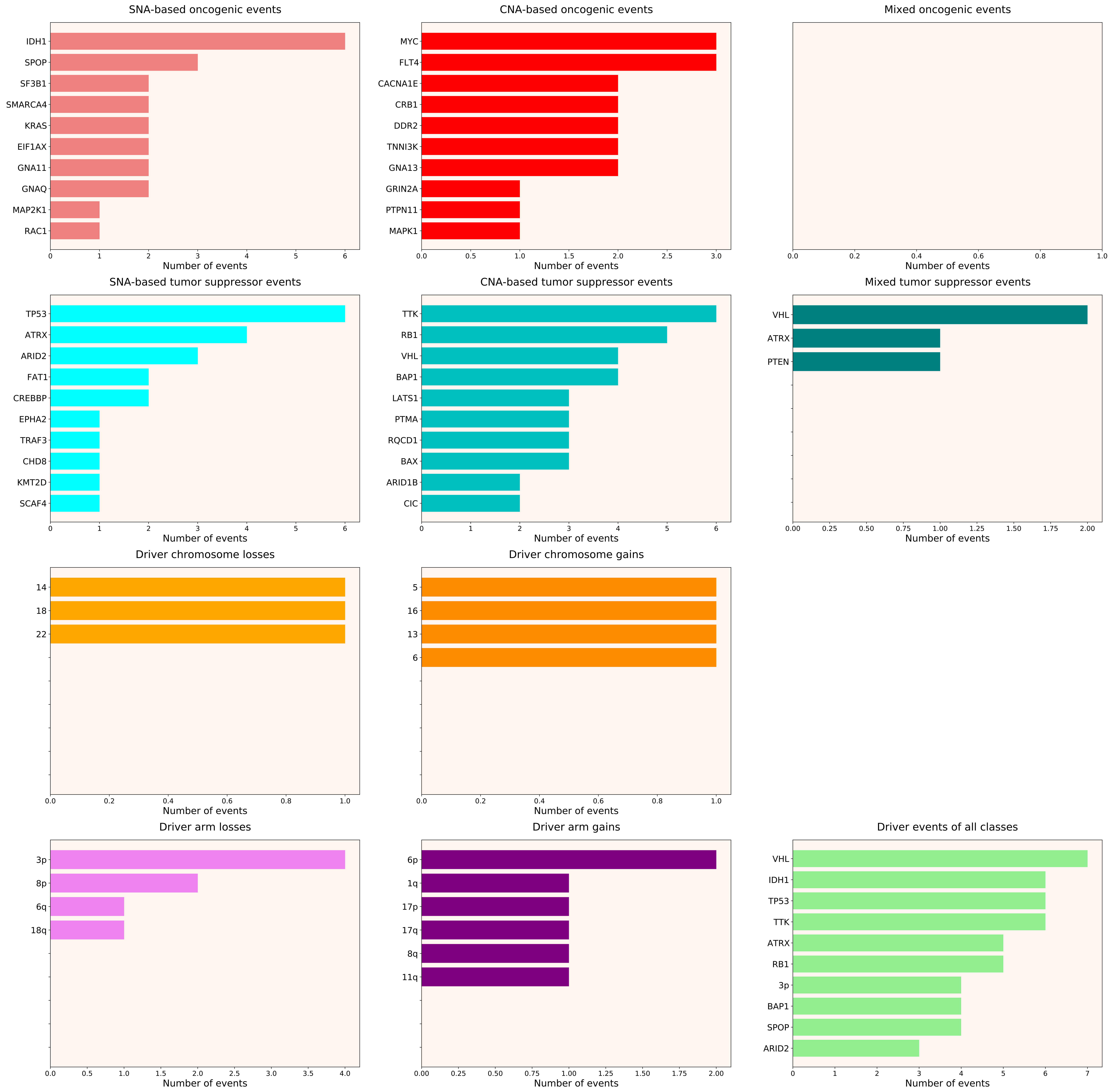

### 2021_8_16_20_32_distribution_events_detailed_males_7.pdf

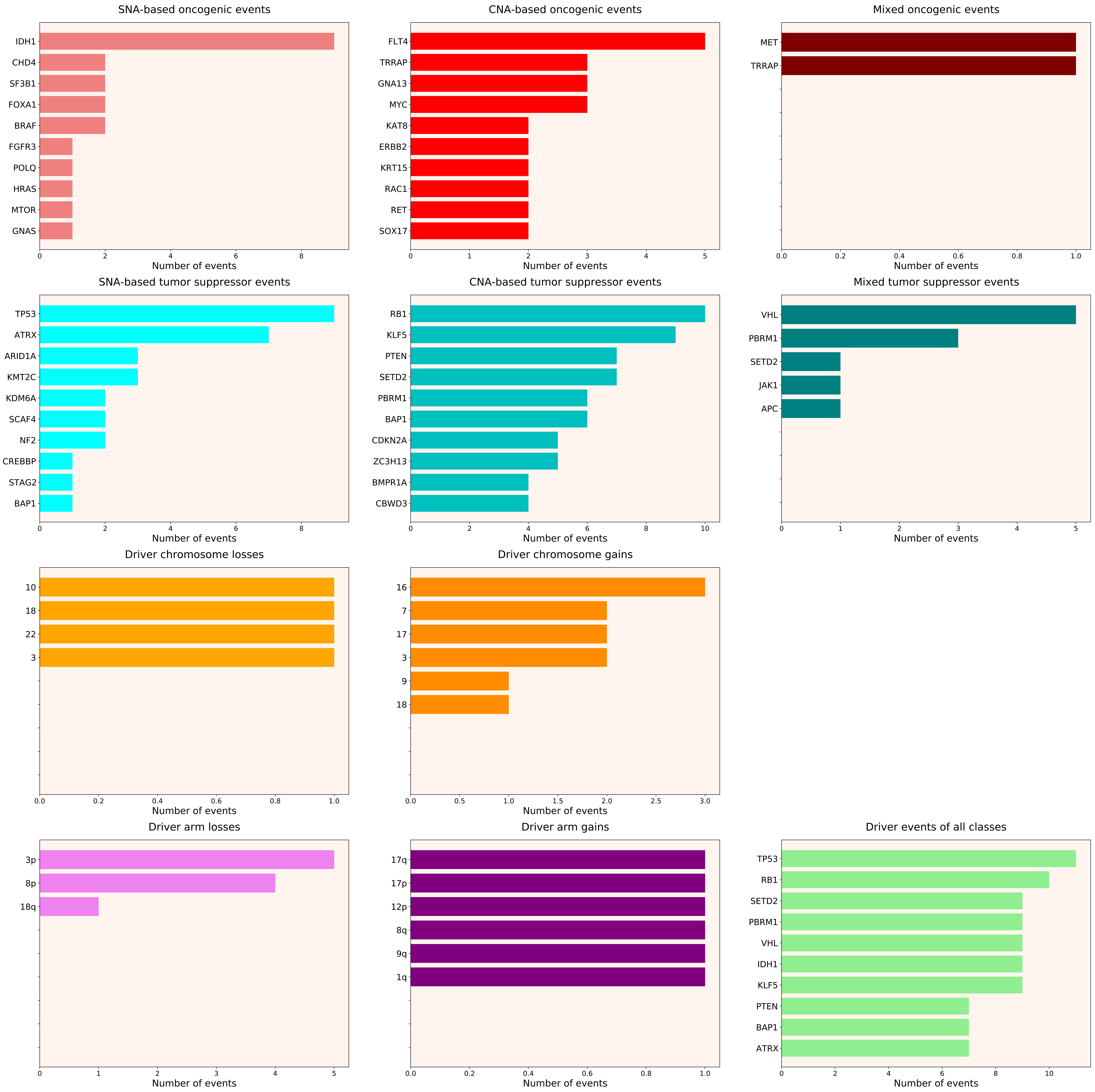

### 2021_8_16_20_32_distribution_events_detailed_males_10.pdf

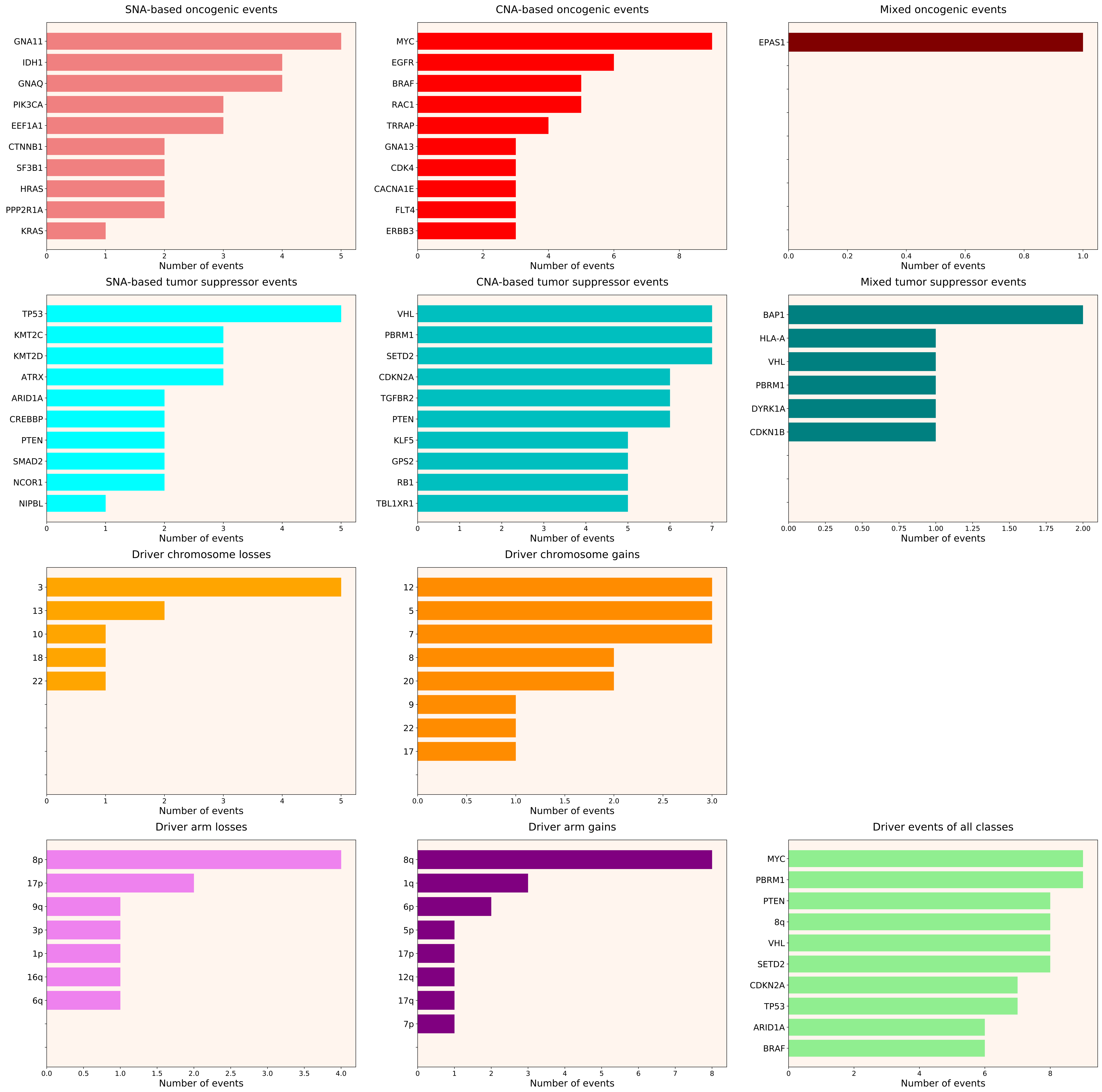

### 2021_8_16_20_32_distribution_events_detailed_males_11.pdf

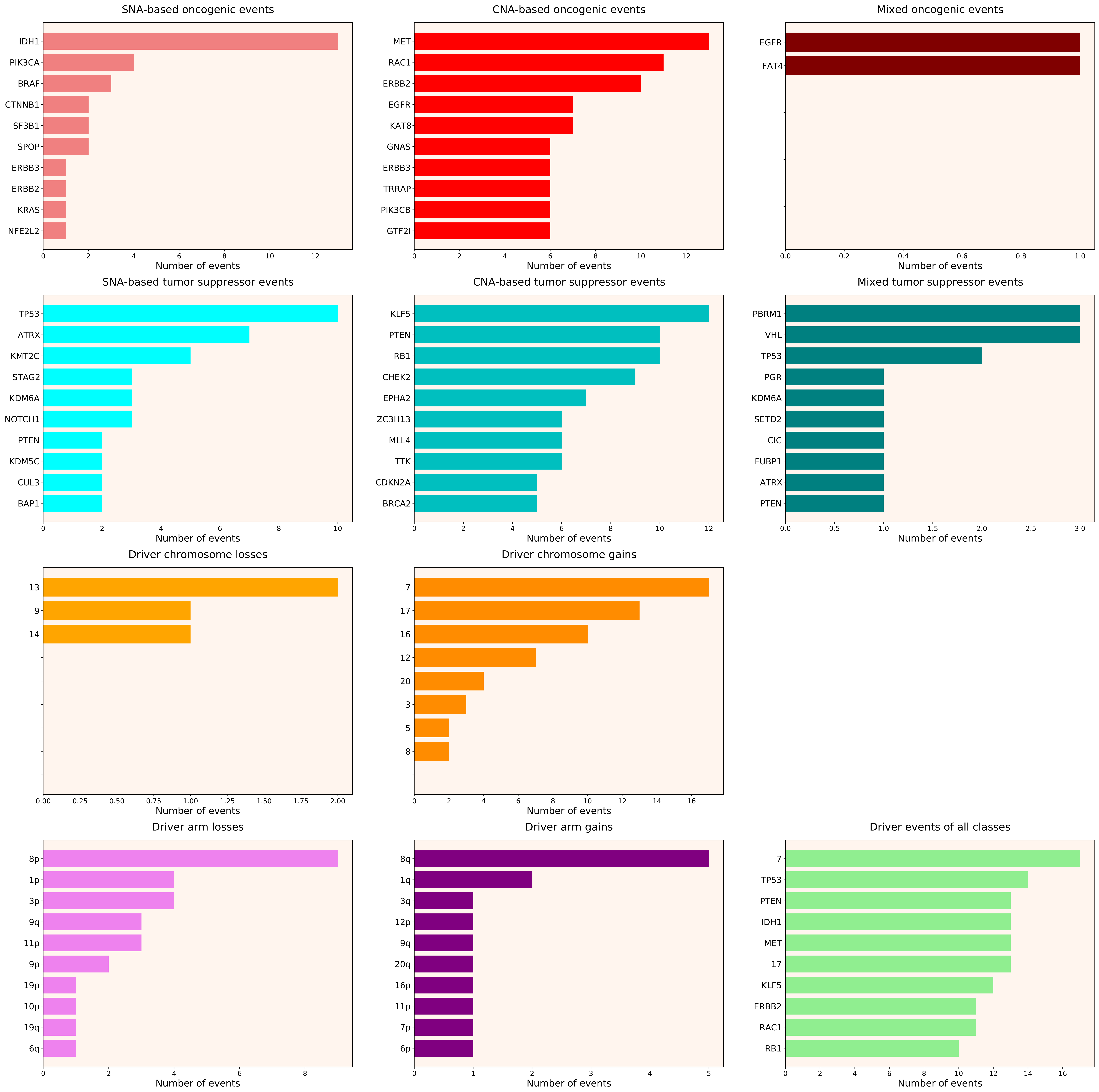

### 2021_8_16_20_32_distribution_events_detailed_males_12.pdf

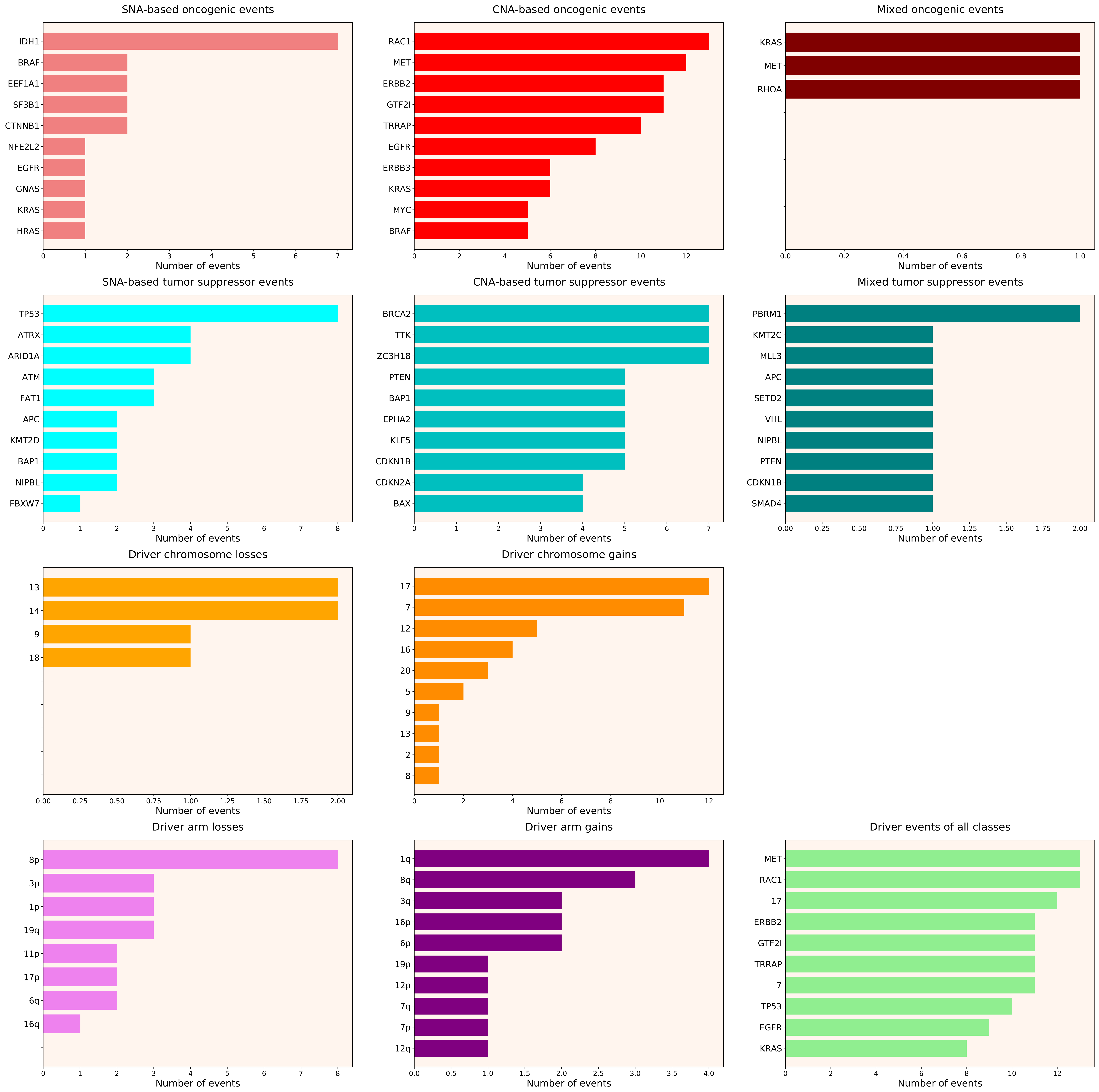

### 2021_8_16_20_32_distribution_events_detailed_males_13.pdf

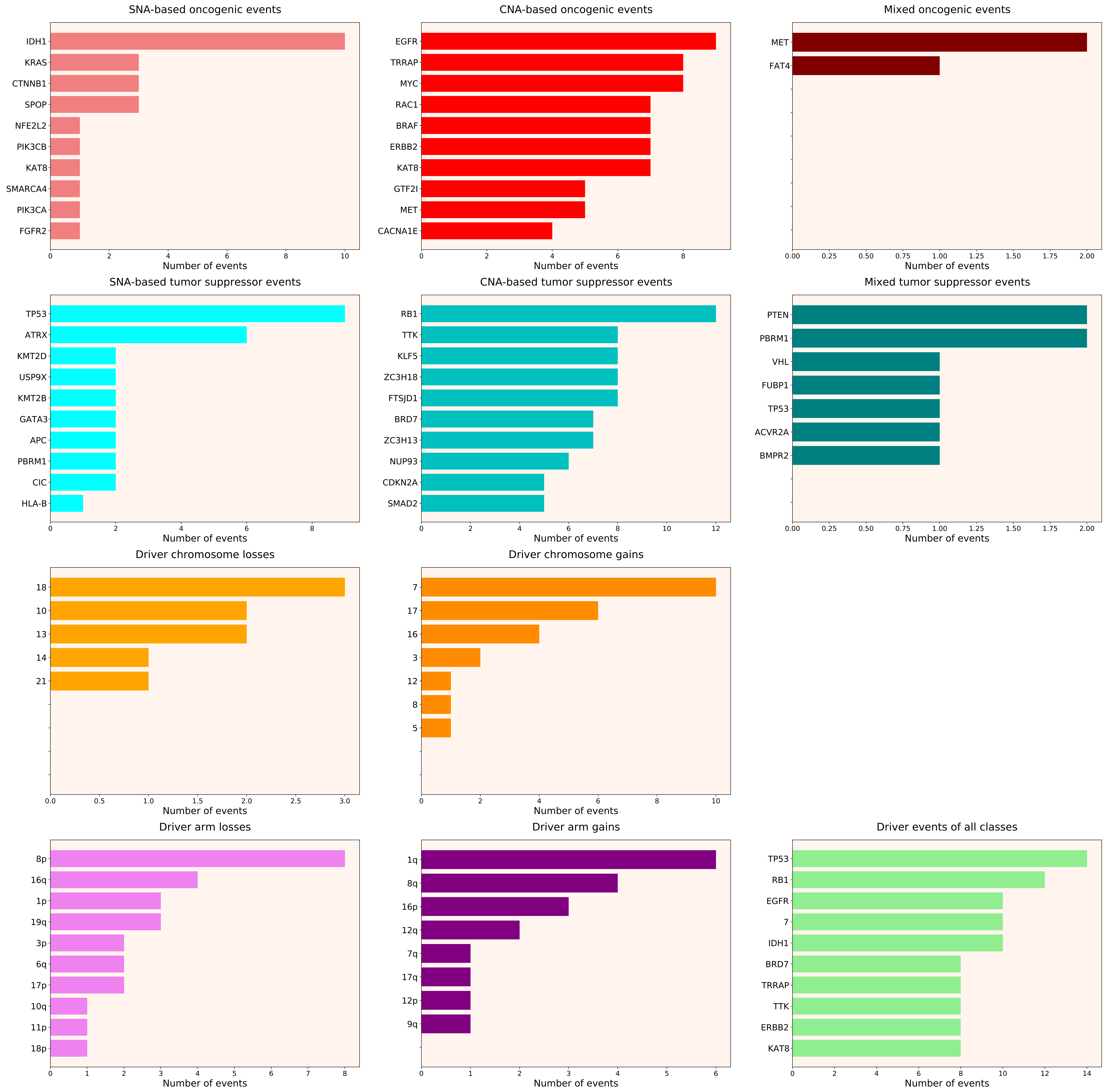

### 2021_8_16_20_32_distribution_events_detailed_males_14.pdf

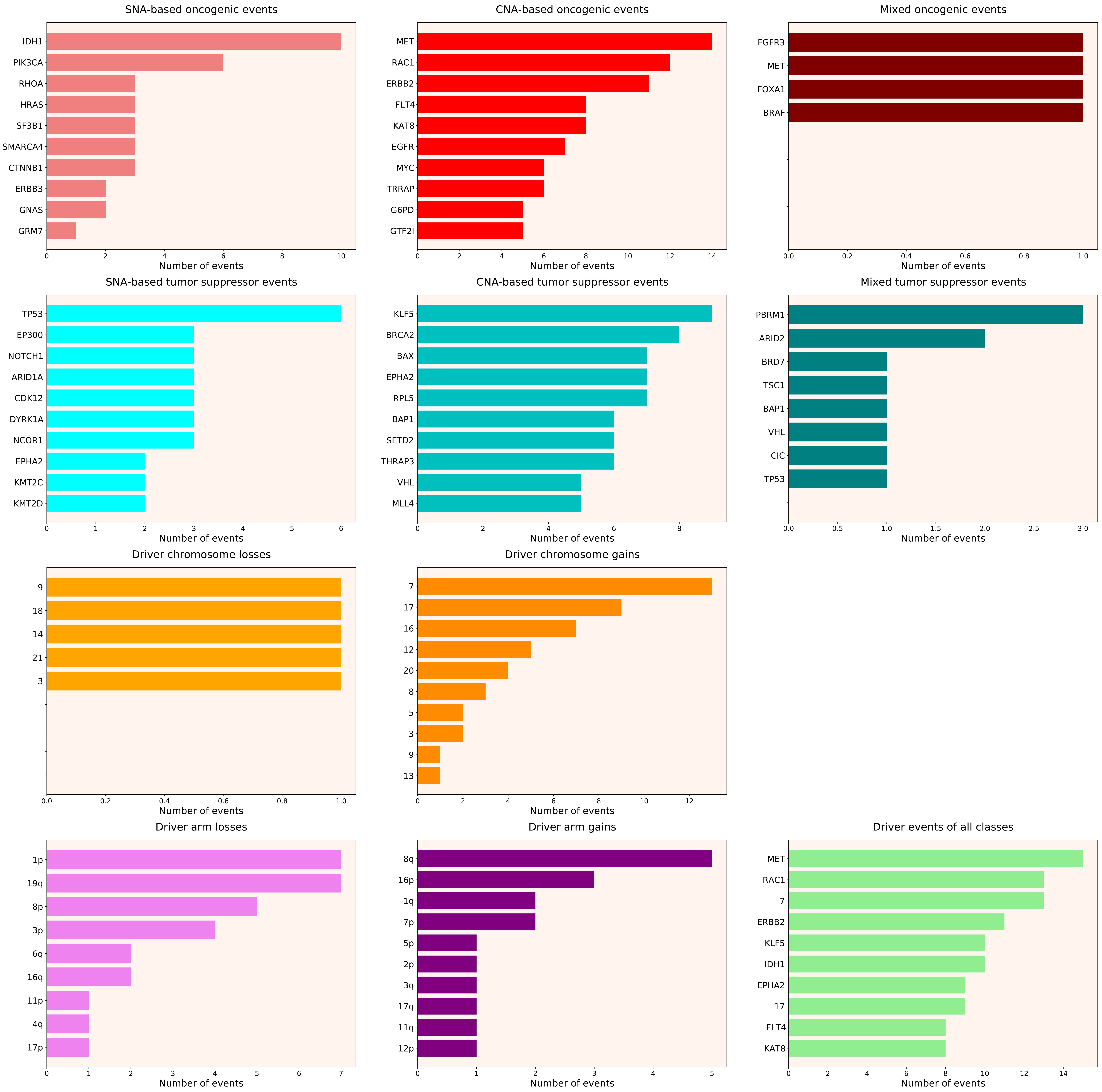

### 2021_8_16_20_32_distribution_events_detailed_males_15.pdf

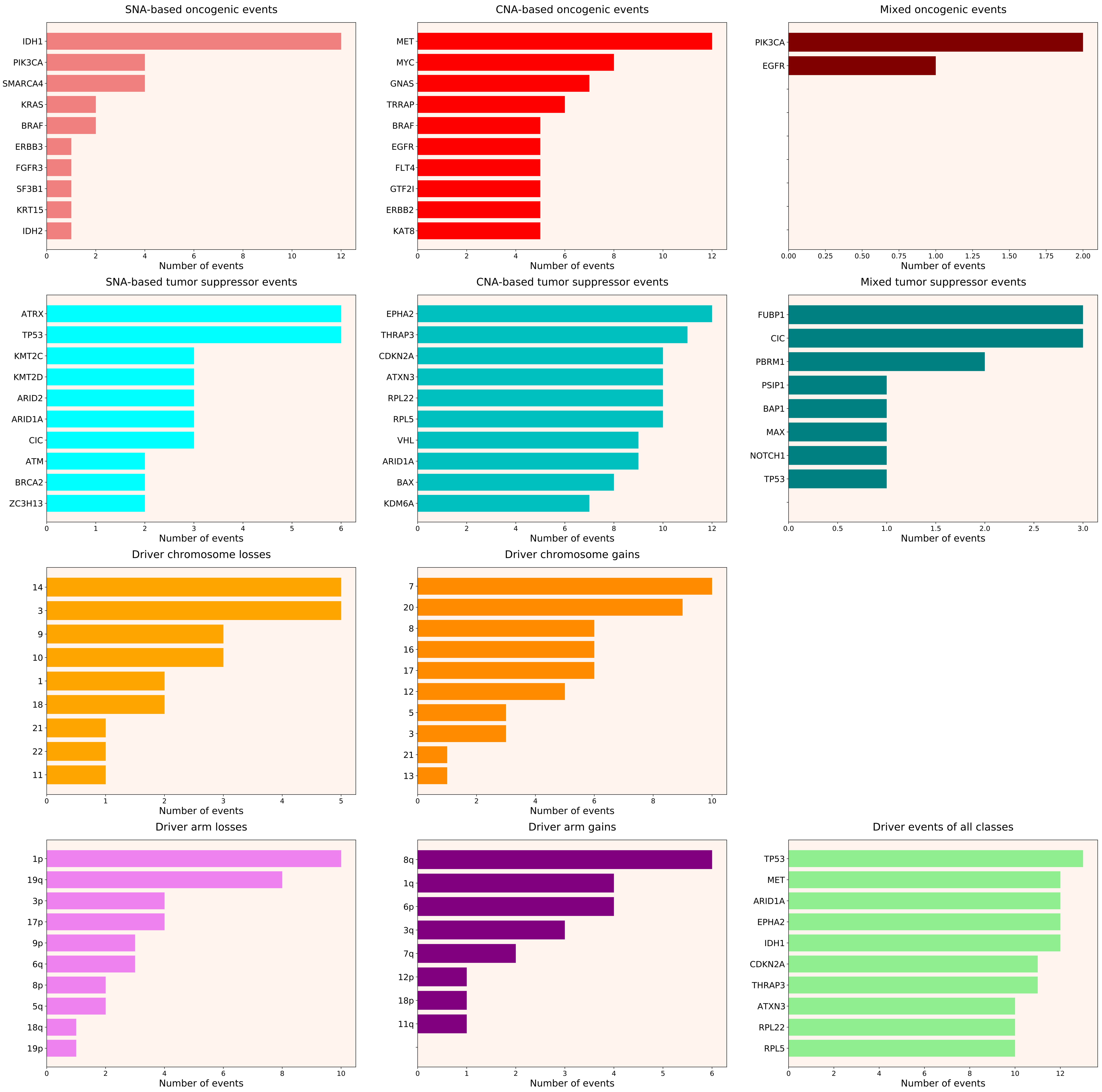

### 2021_8_16_20_32_distribution_events_detailed_males_28.pdf

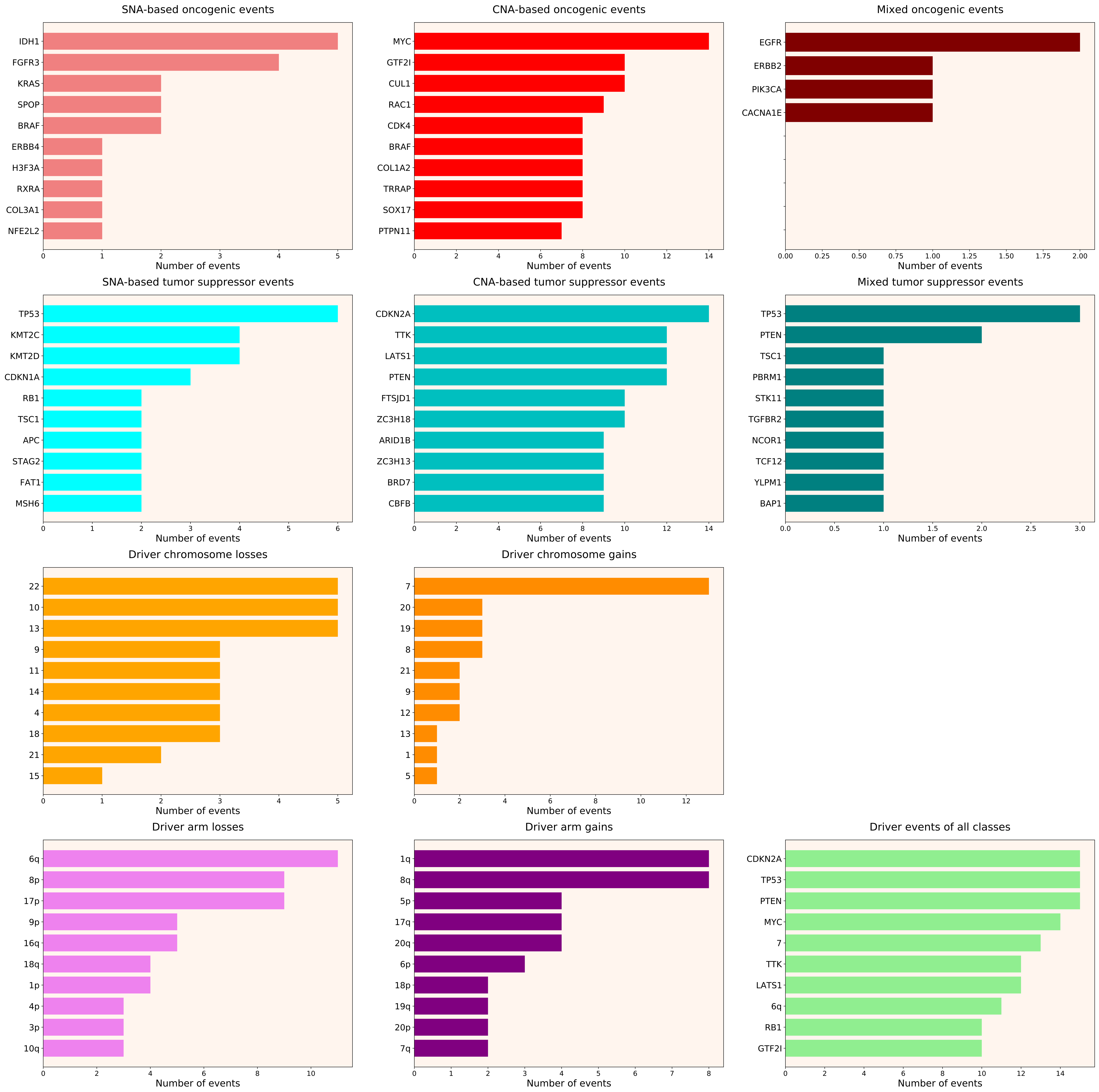

### 2021_8_16_20_32_distribution_events_detailed_males_29.pdf

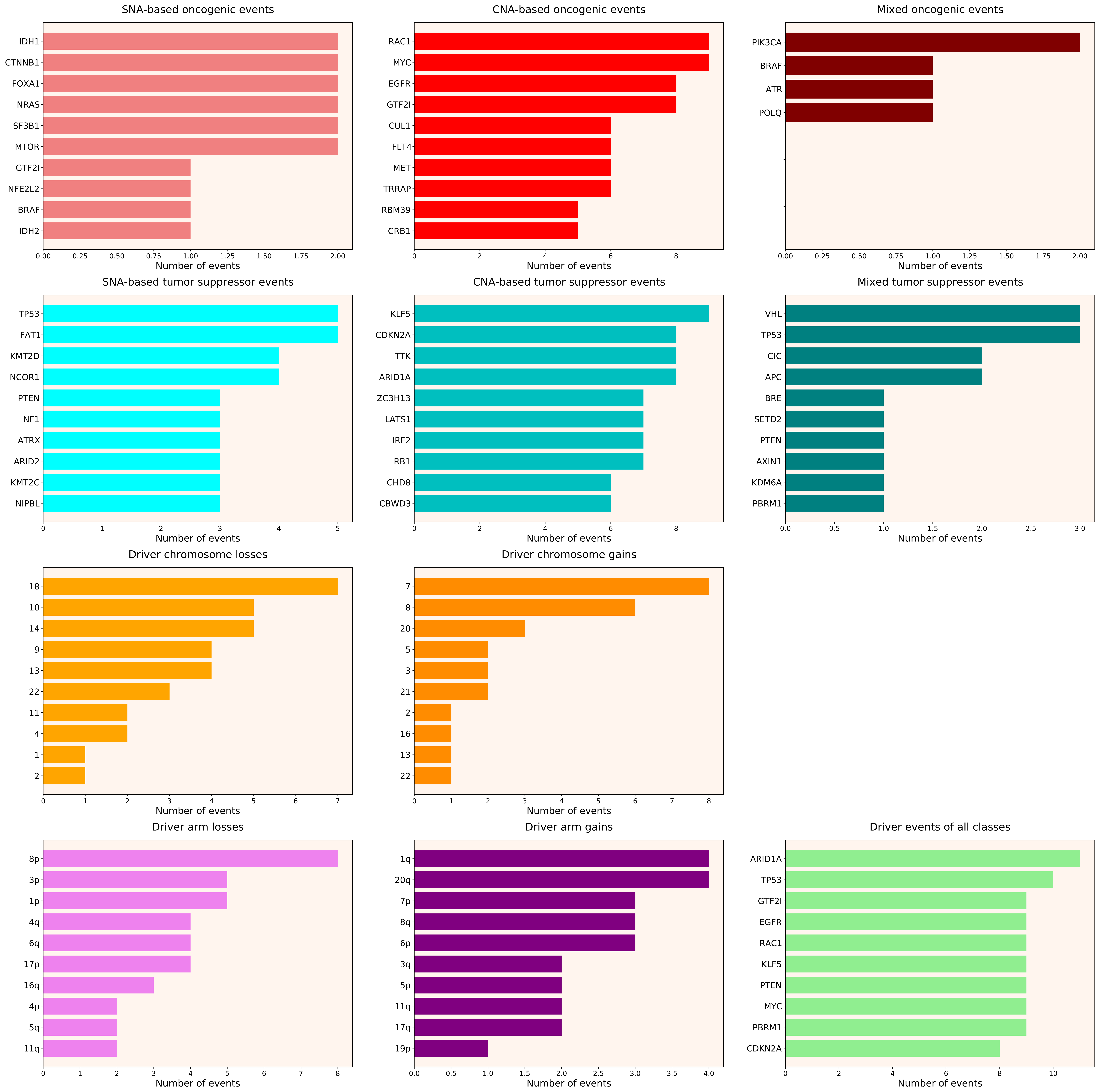

### 2021_8_16_20_32_distribution_events_detailed_males_38.pdf

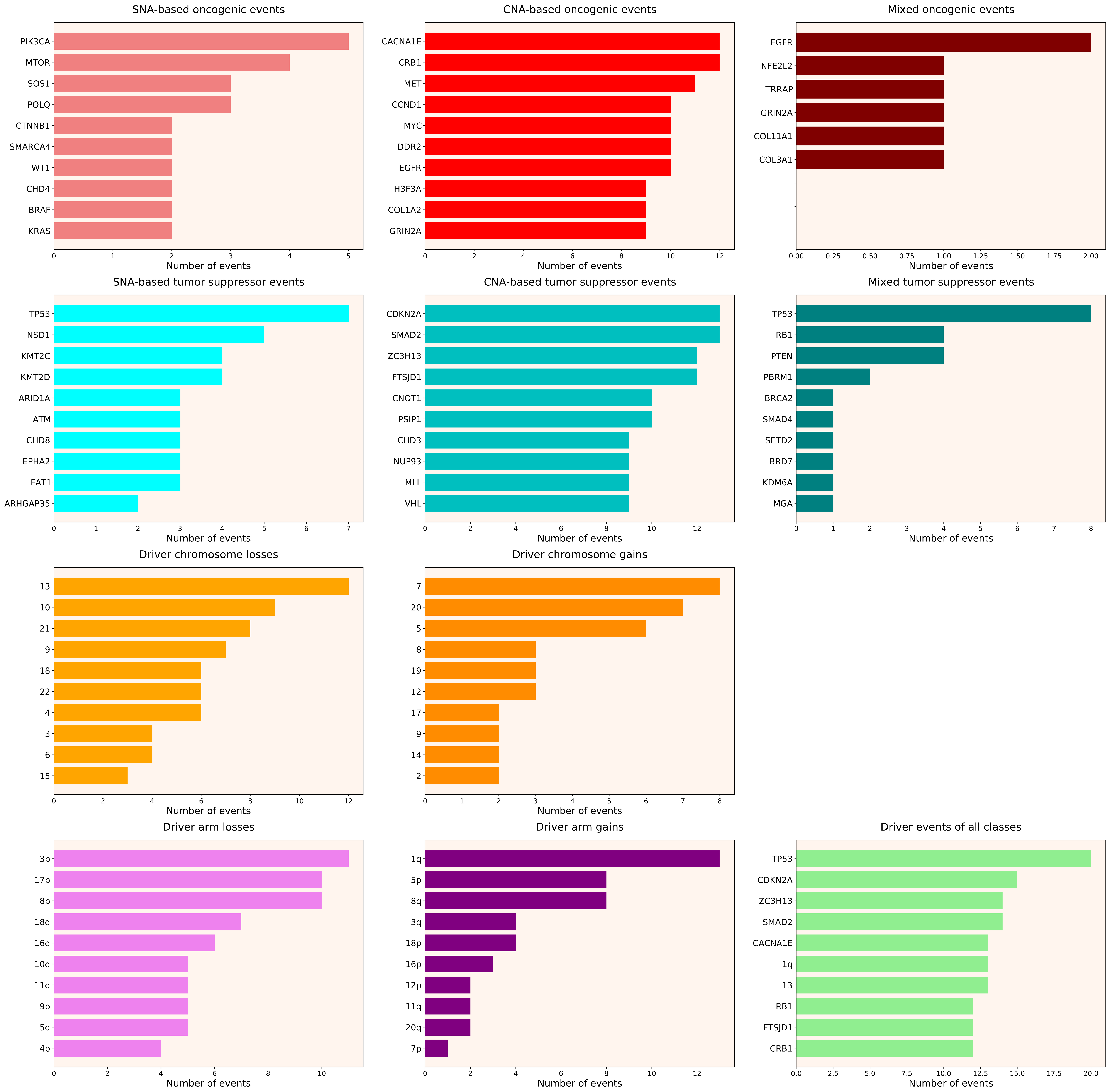

### 2021_8_16_20_32_distribution_events_detailed_males_39.pdf

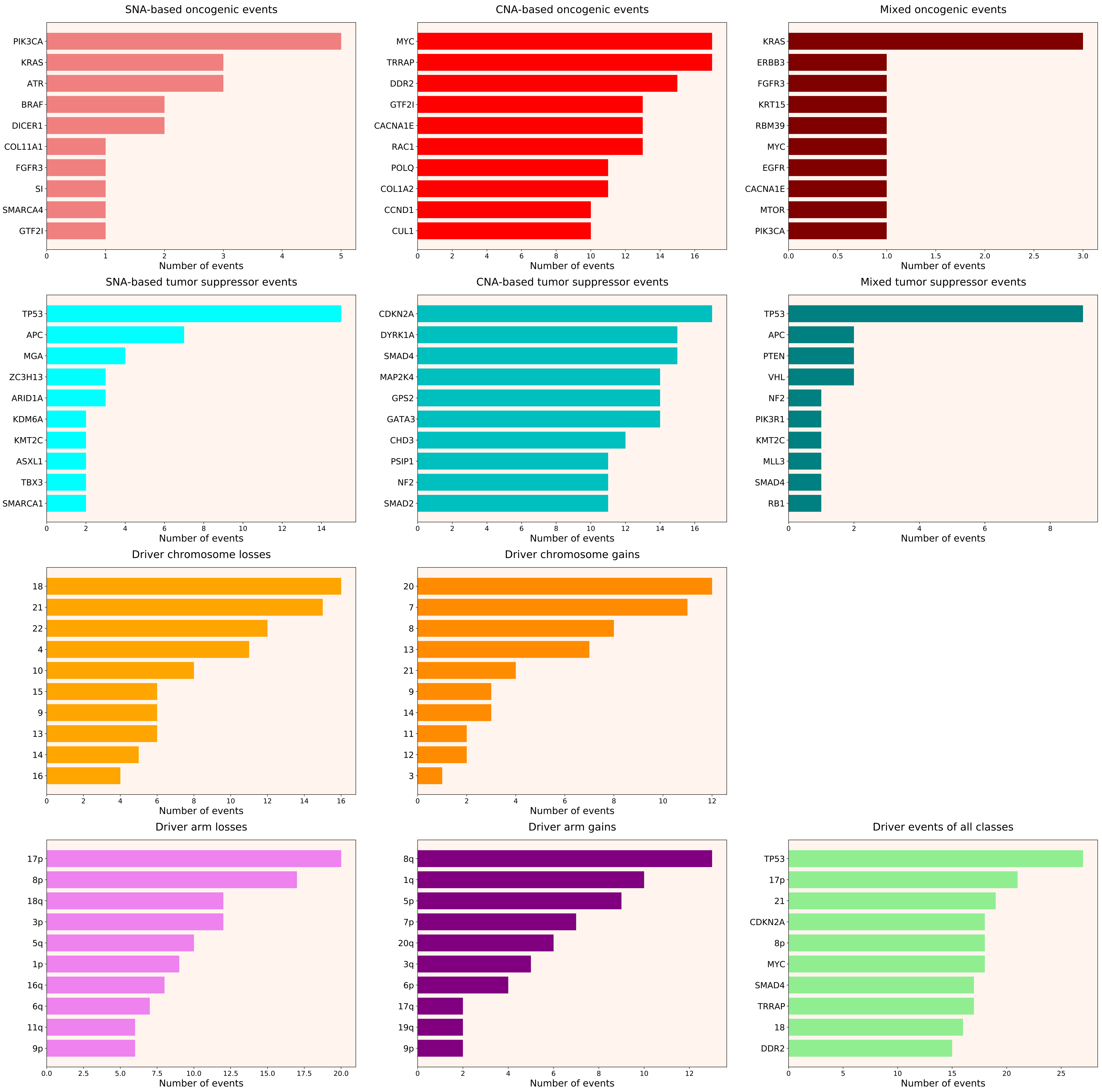

### 2021_8_16_20_32_distribution_events_detailed_NDSI.pdf

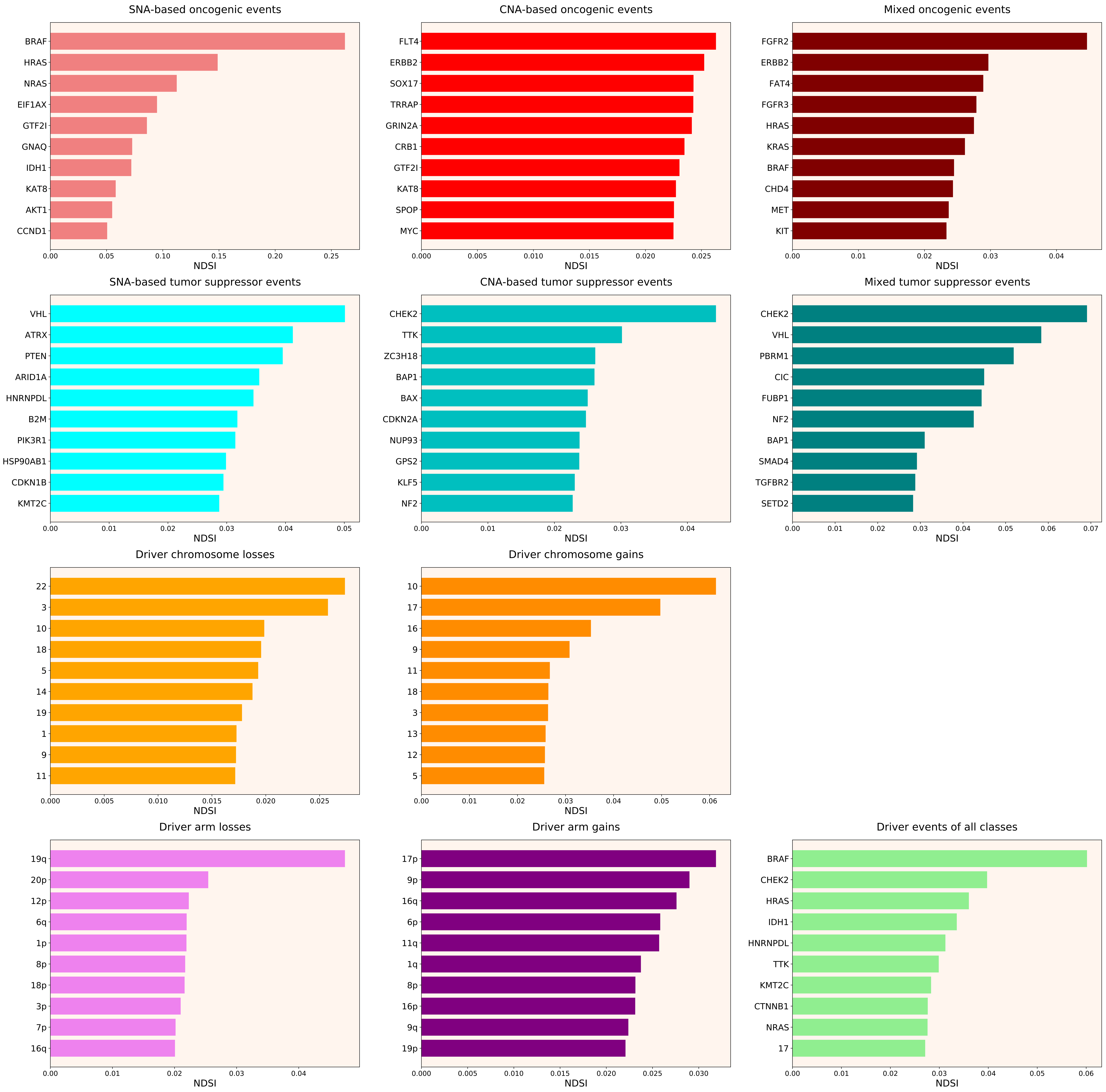

### 2021_8_16_20_32_distribution_events_detailed_NDSI_females.pdf

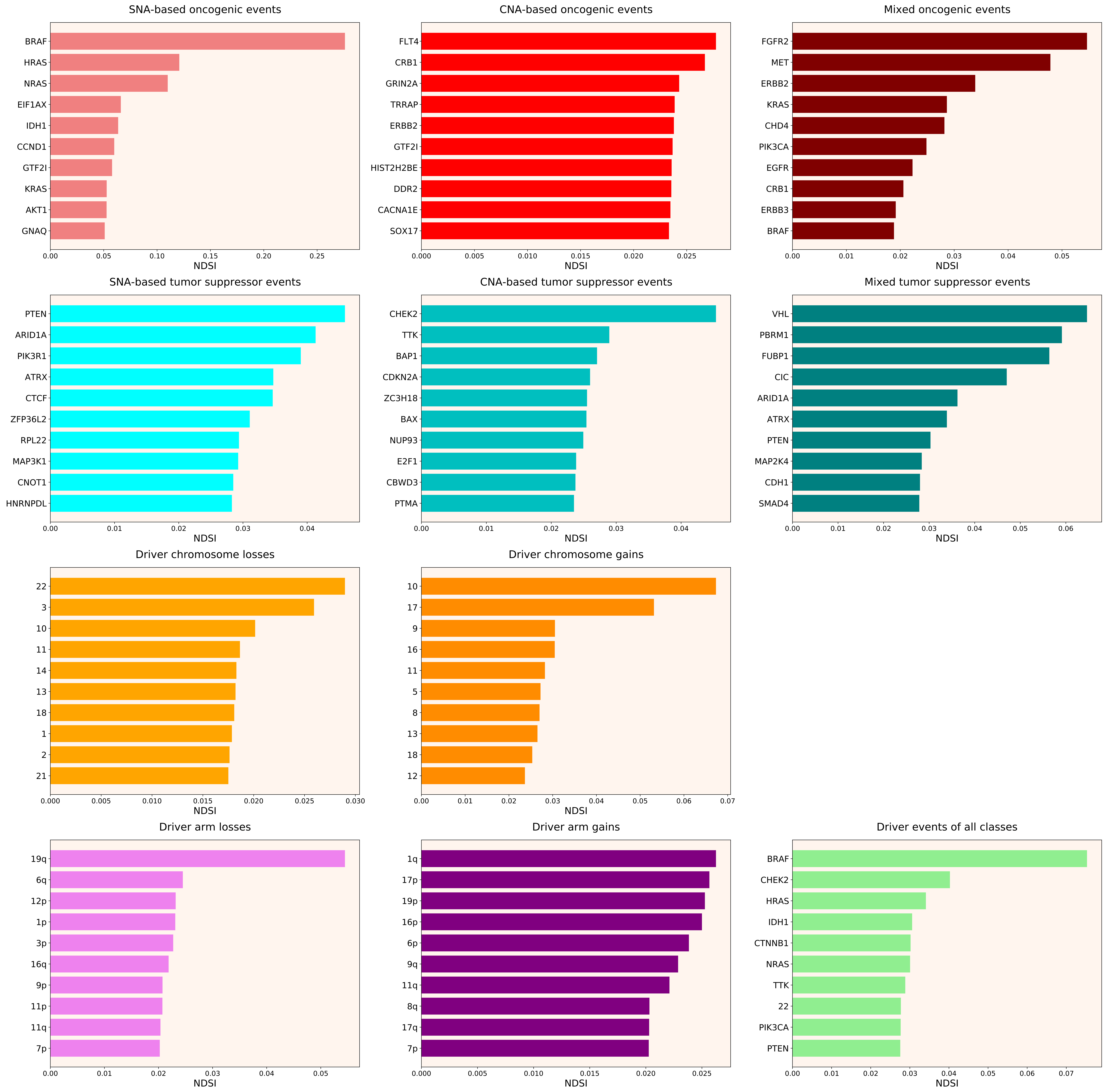

### 2021_8_16_20_32_distribution_events_detailed_NDSI_males.pdf

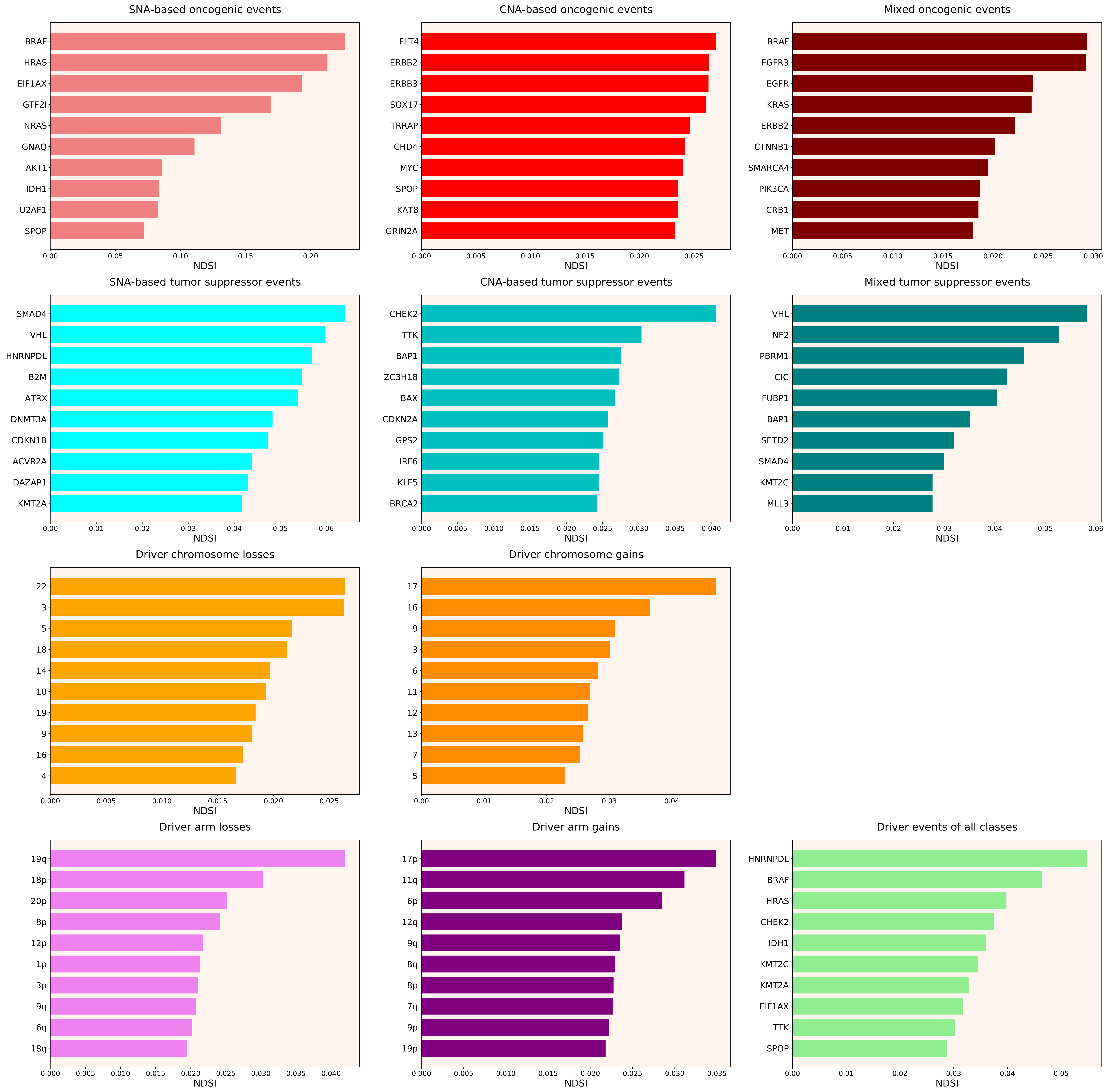

### 2021_8_16_20_32_LUSC.pdf

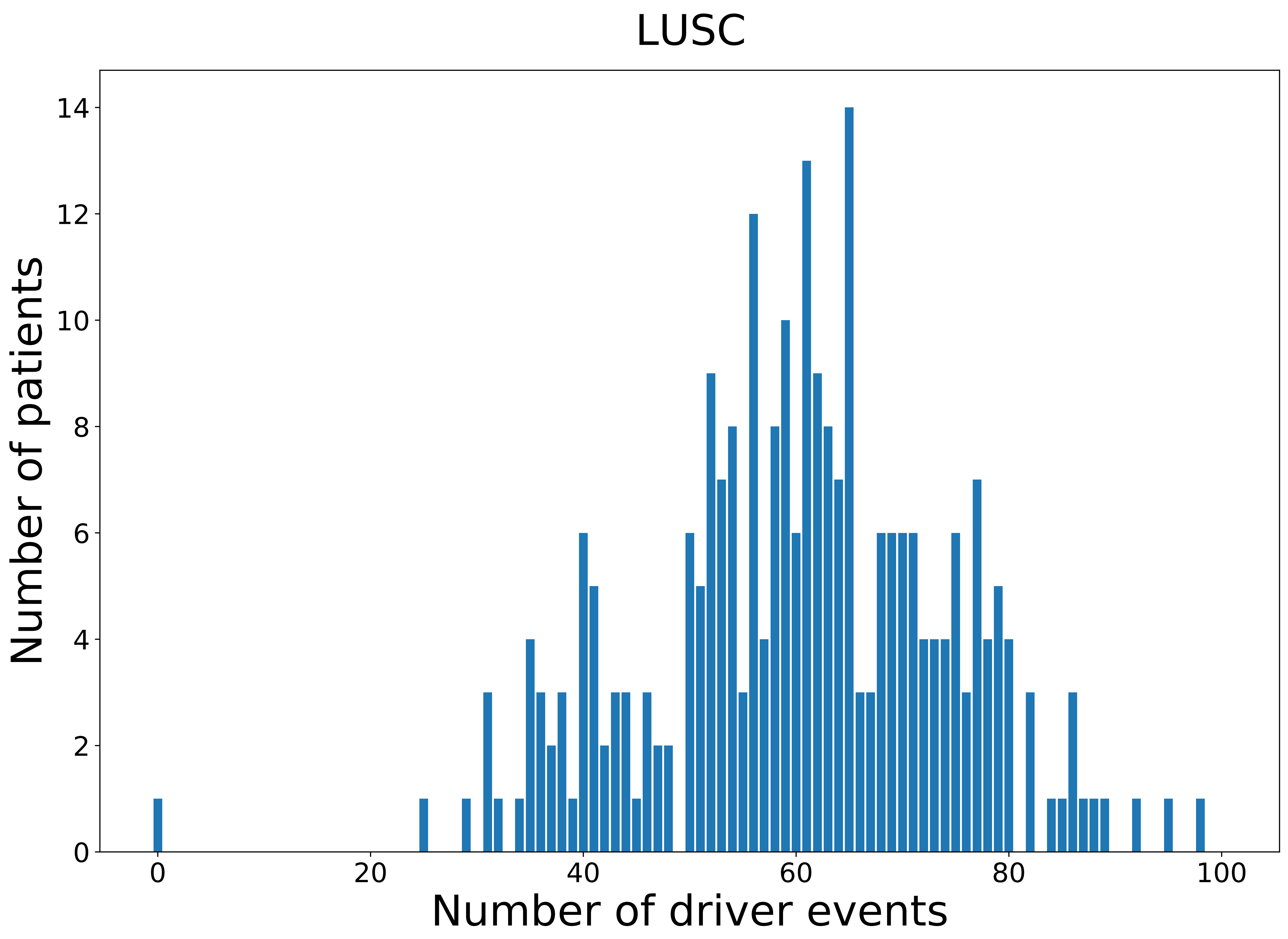

### 2021_8_16_20_32_LUSC_FEMALE.pdf

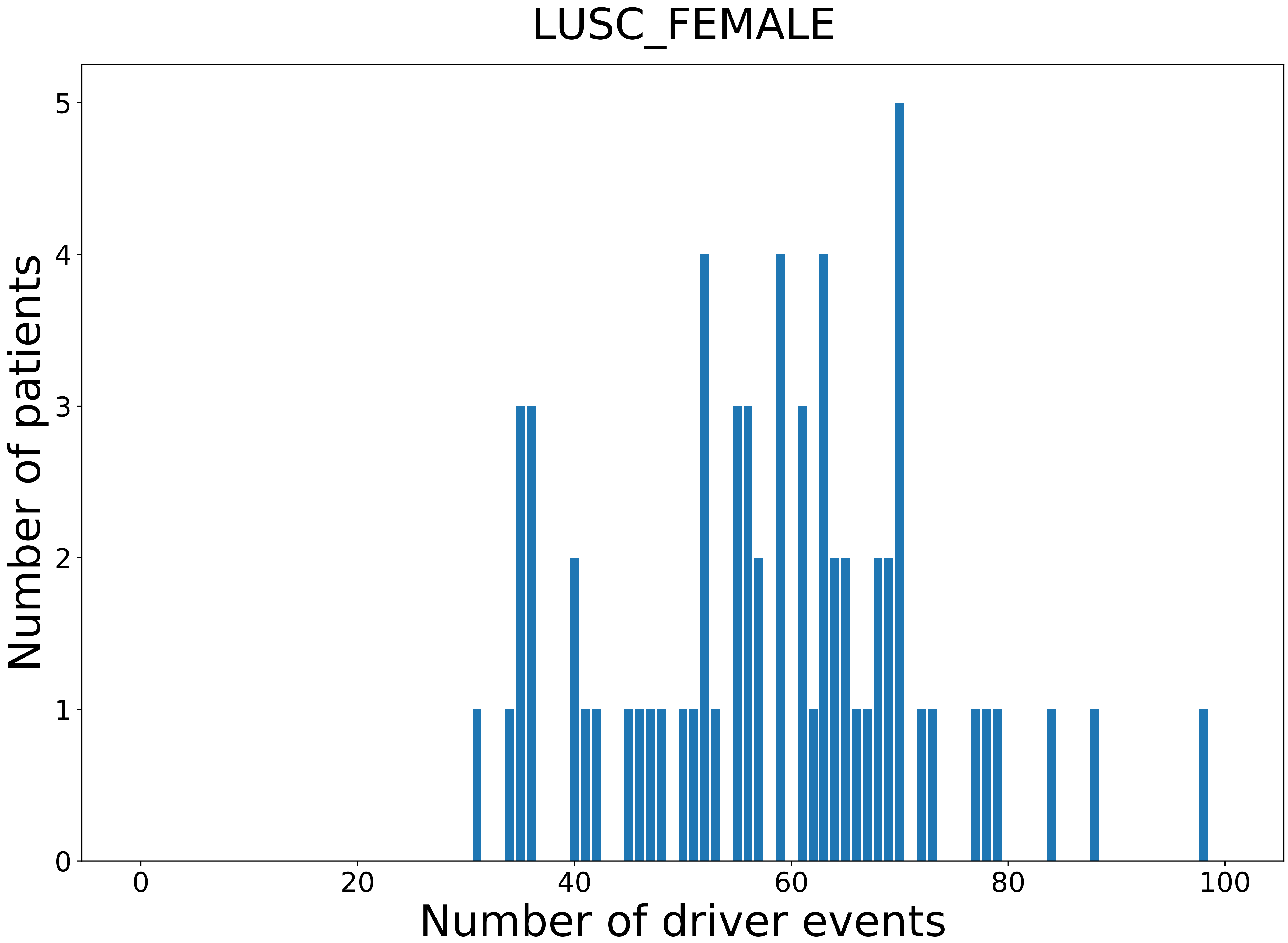

### 2021_8_16_20_32_MESO_MALE.pdf

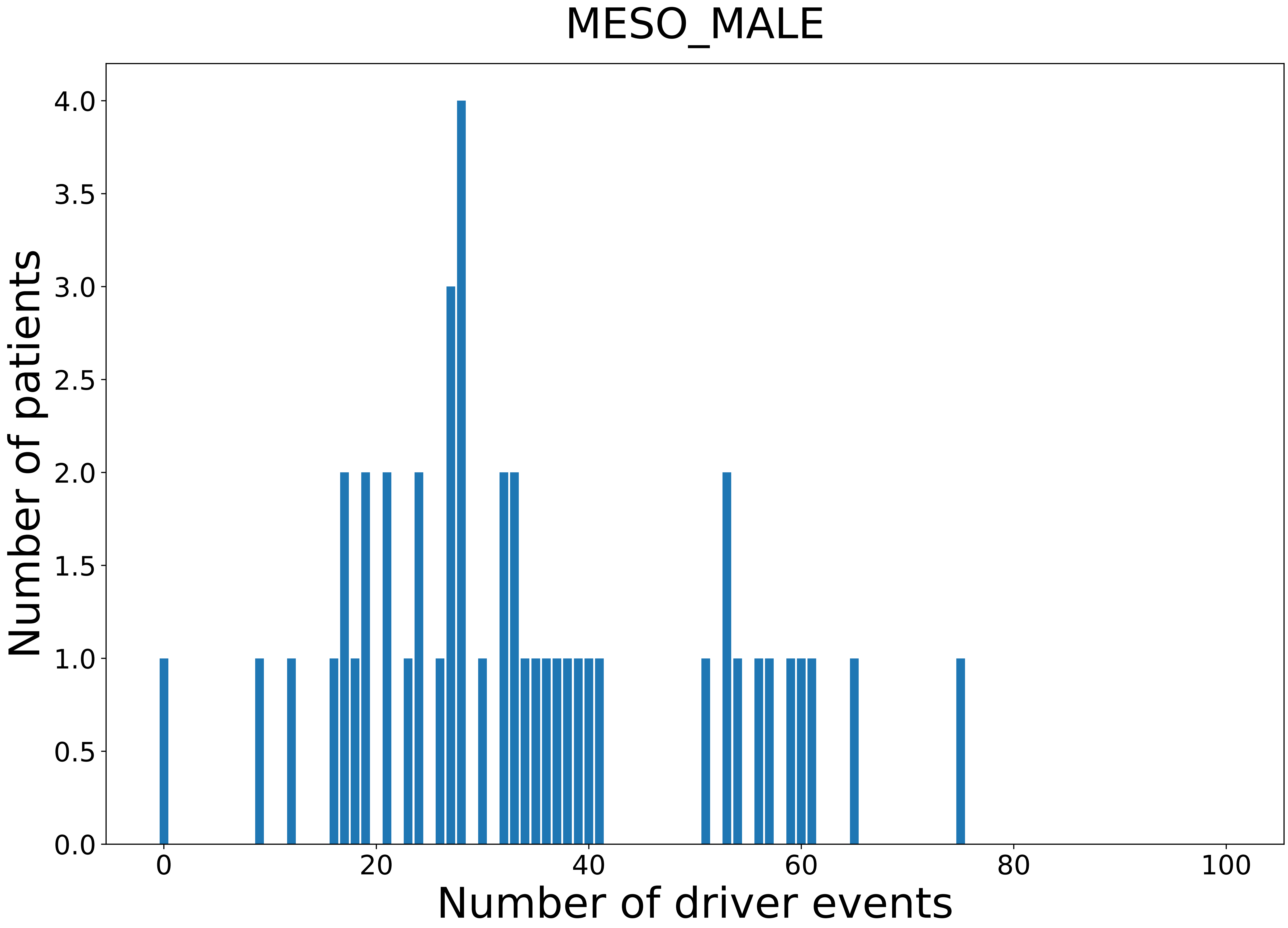

### 2021_8_16_20_32_PANCAN.pdf

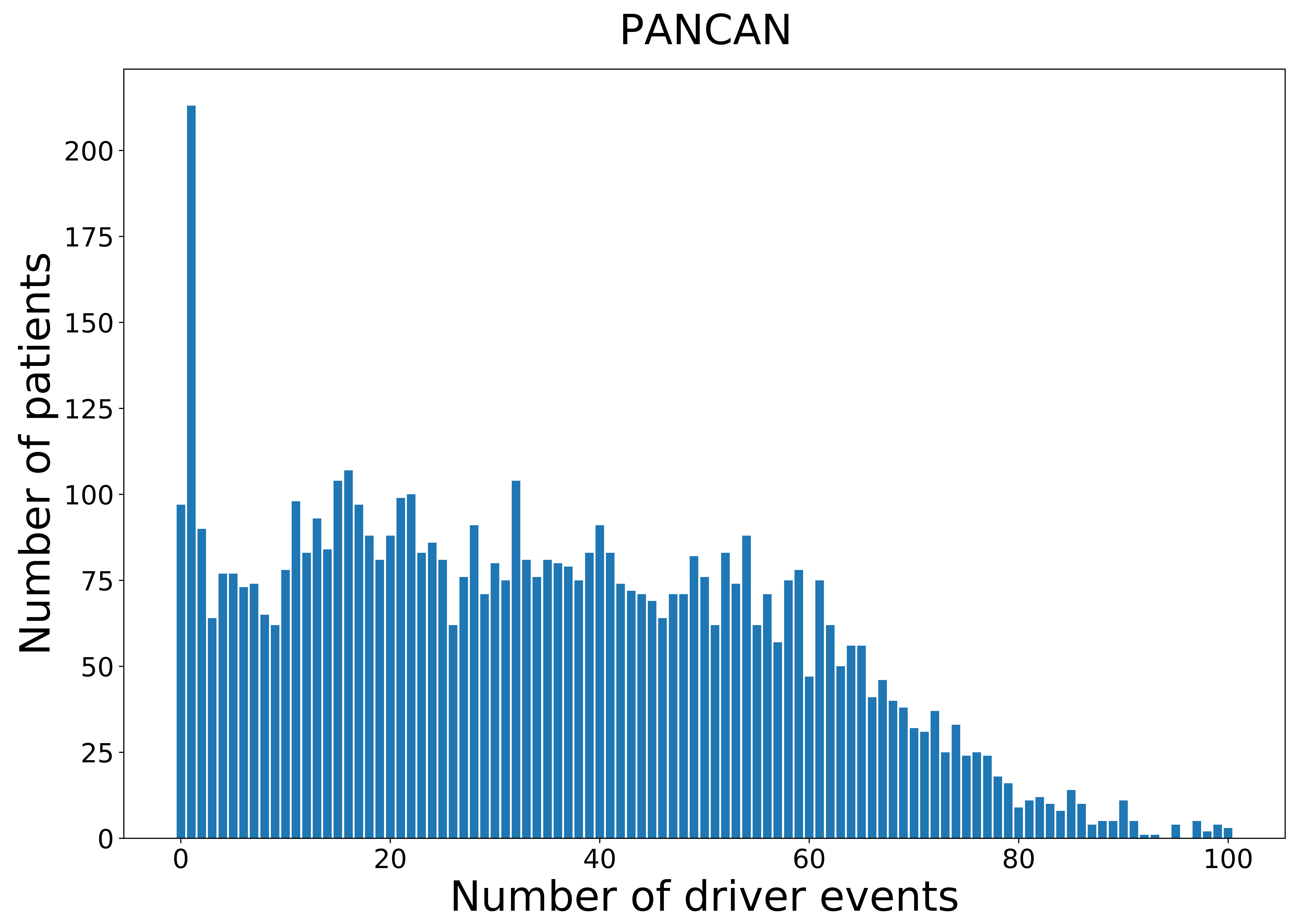

### 2021_8_16_20_32_PCPG.pdf

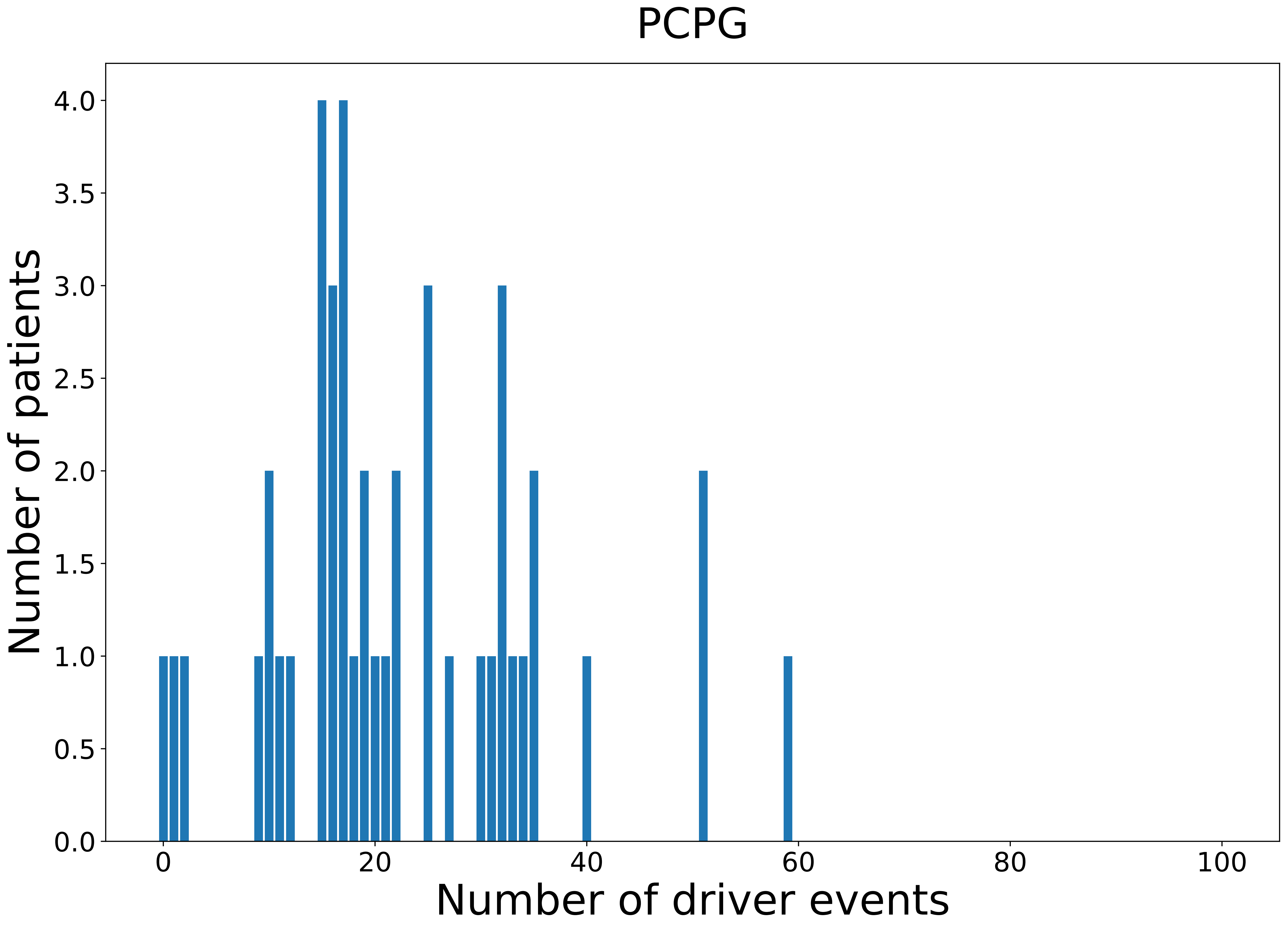

### 2021_8_16_20_32_TGCT_MALE.pdf

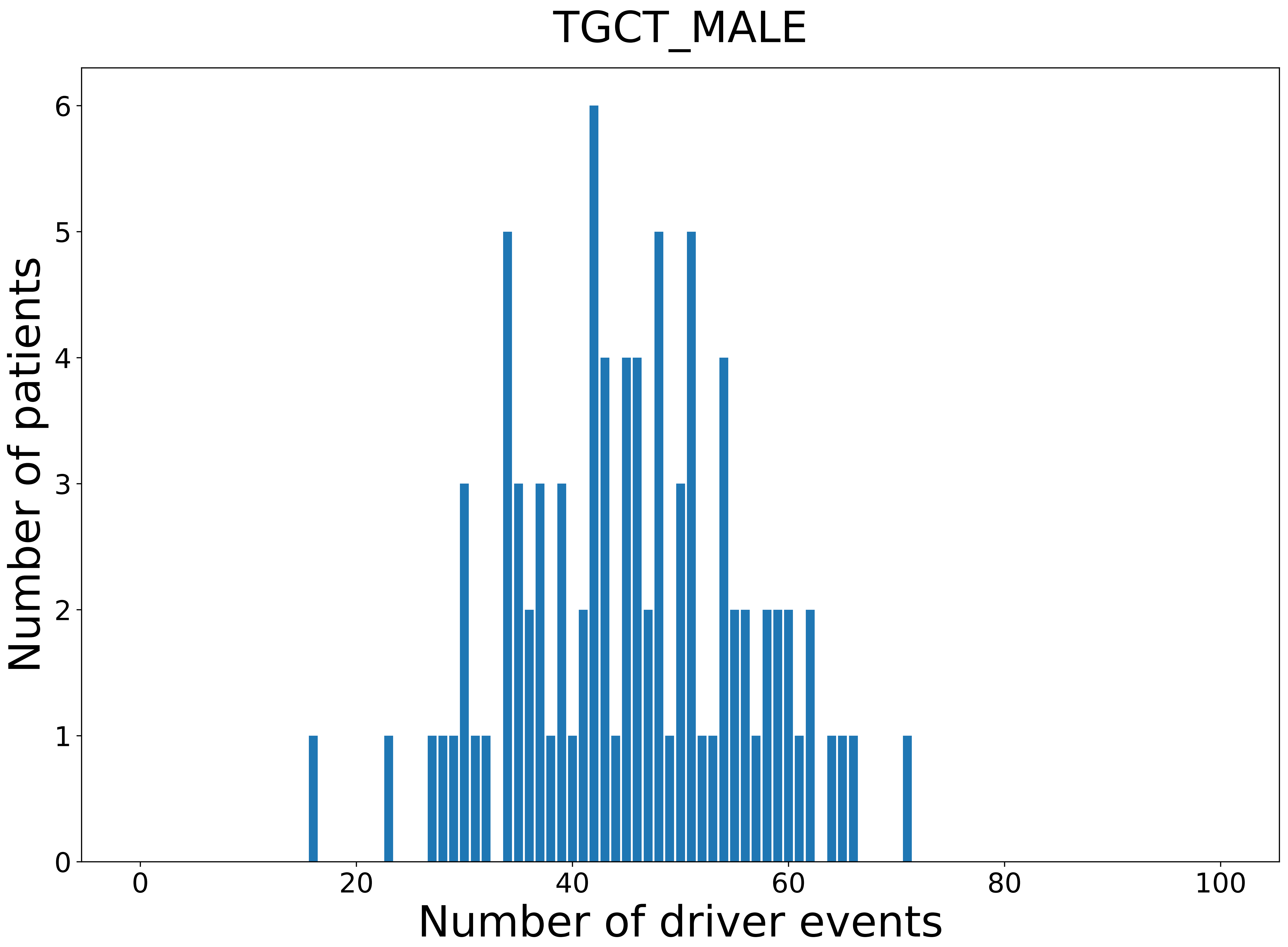
