## Supplementary figures and images for "Novel Driver Strength Index highlights important cancer genes in TCGA PanCanAtlas patients"

### 2021_8_16_20_32_ACC.pdf

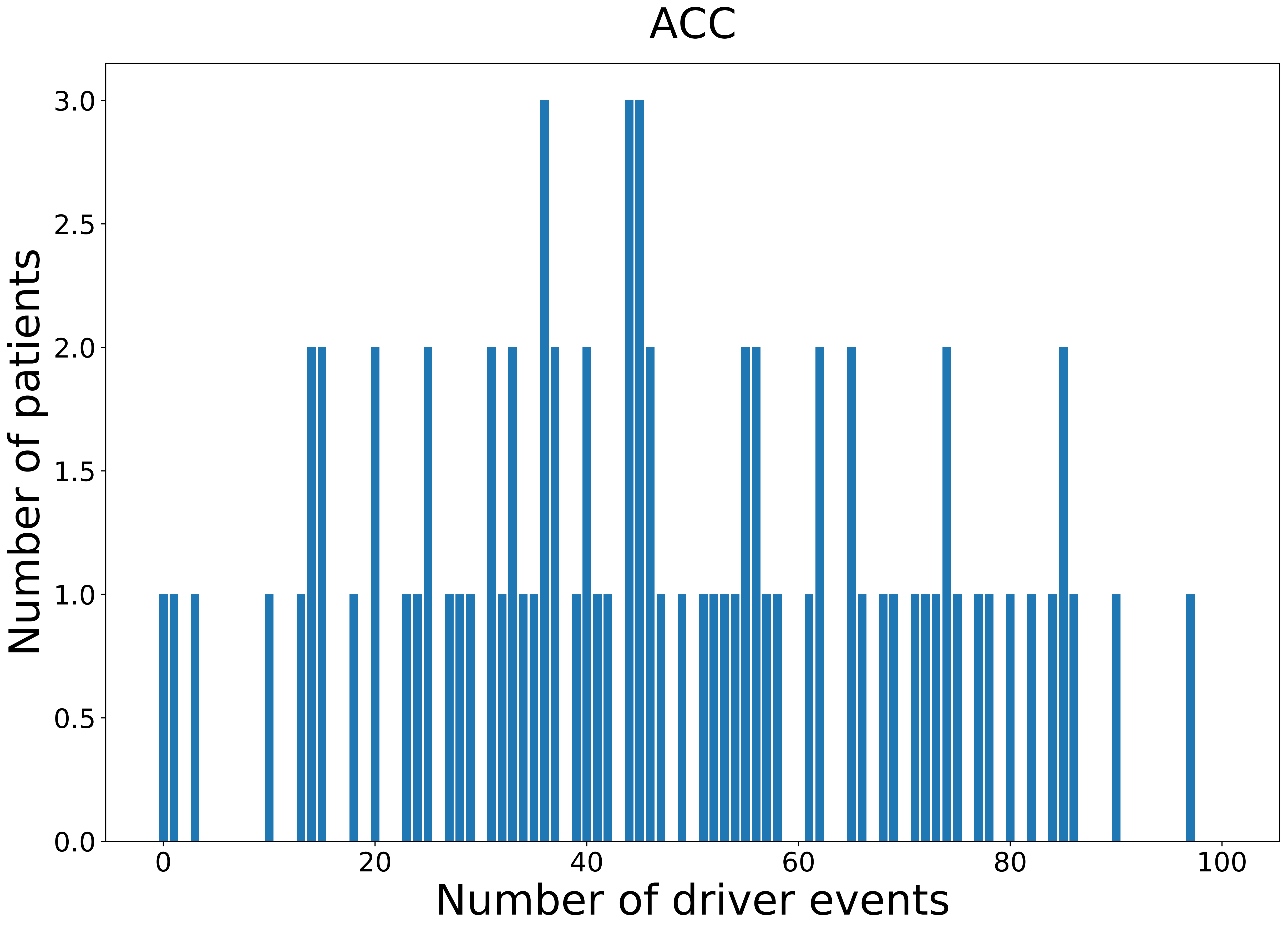

### 2021_8_16_20_32_ACC_FEMALE.pdf

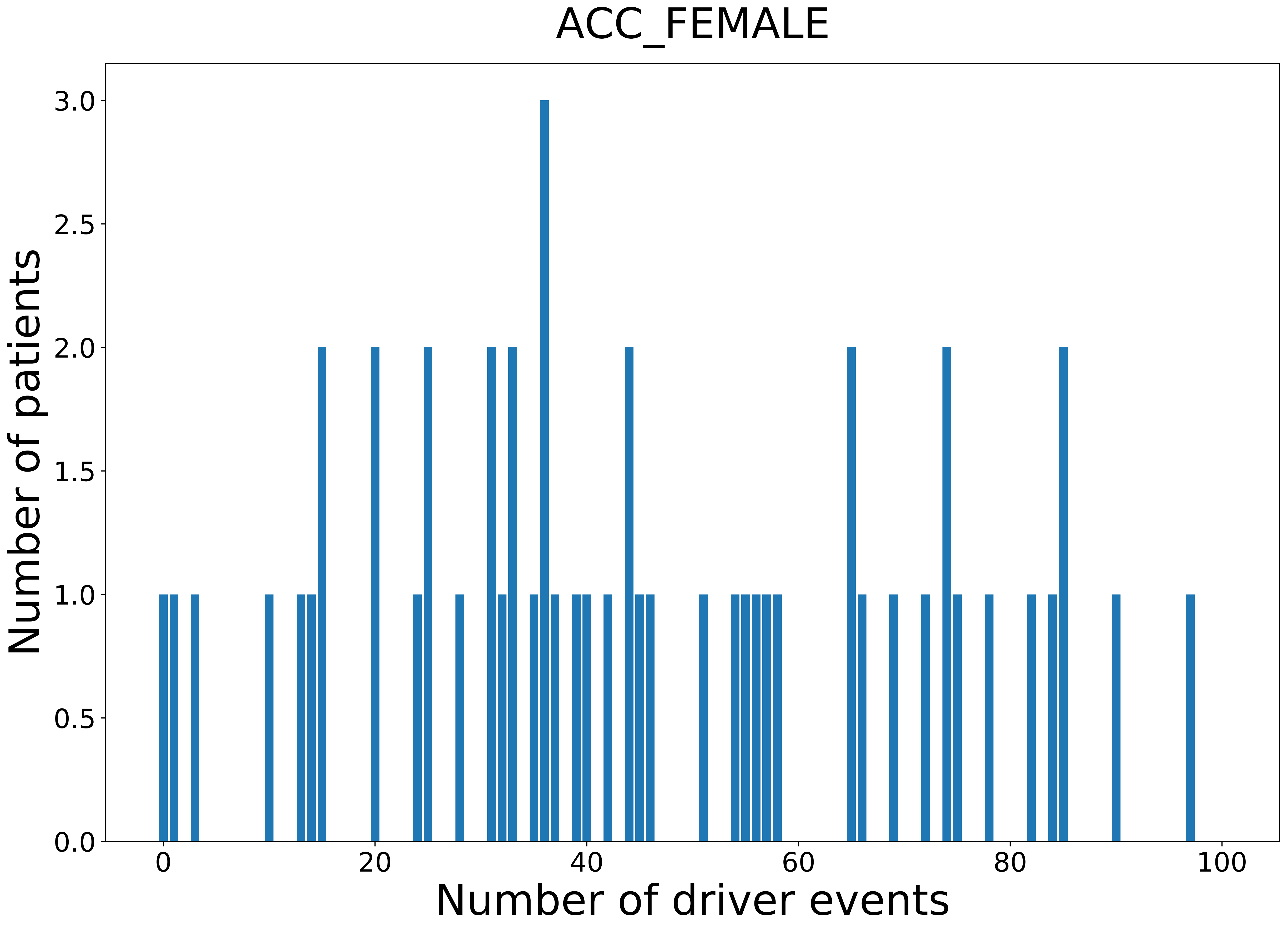

### 2021_8_16_20_32_ACC_MALE.pdf

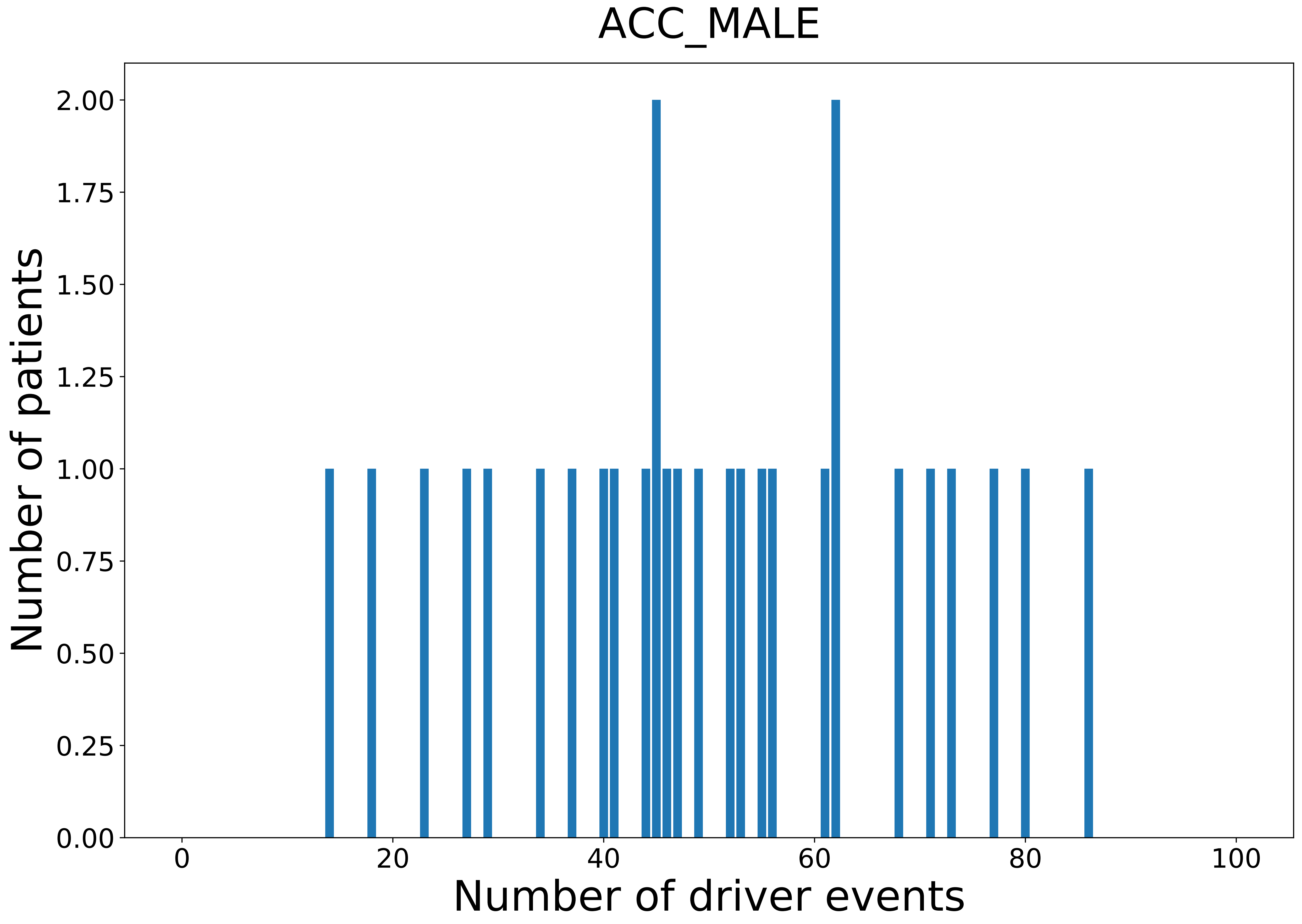

### 2021_8_16_20_32_BLCA.pdf

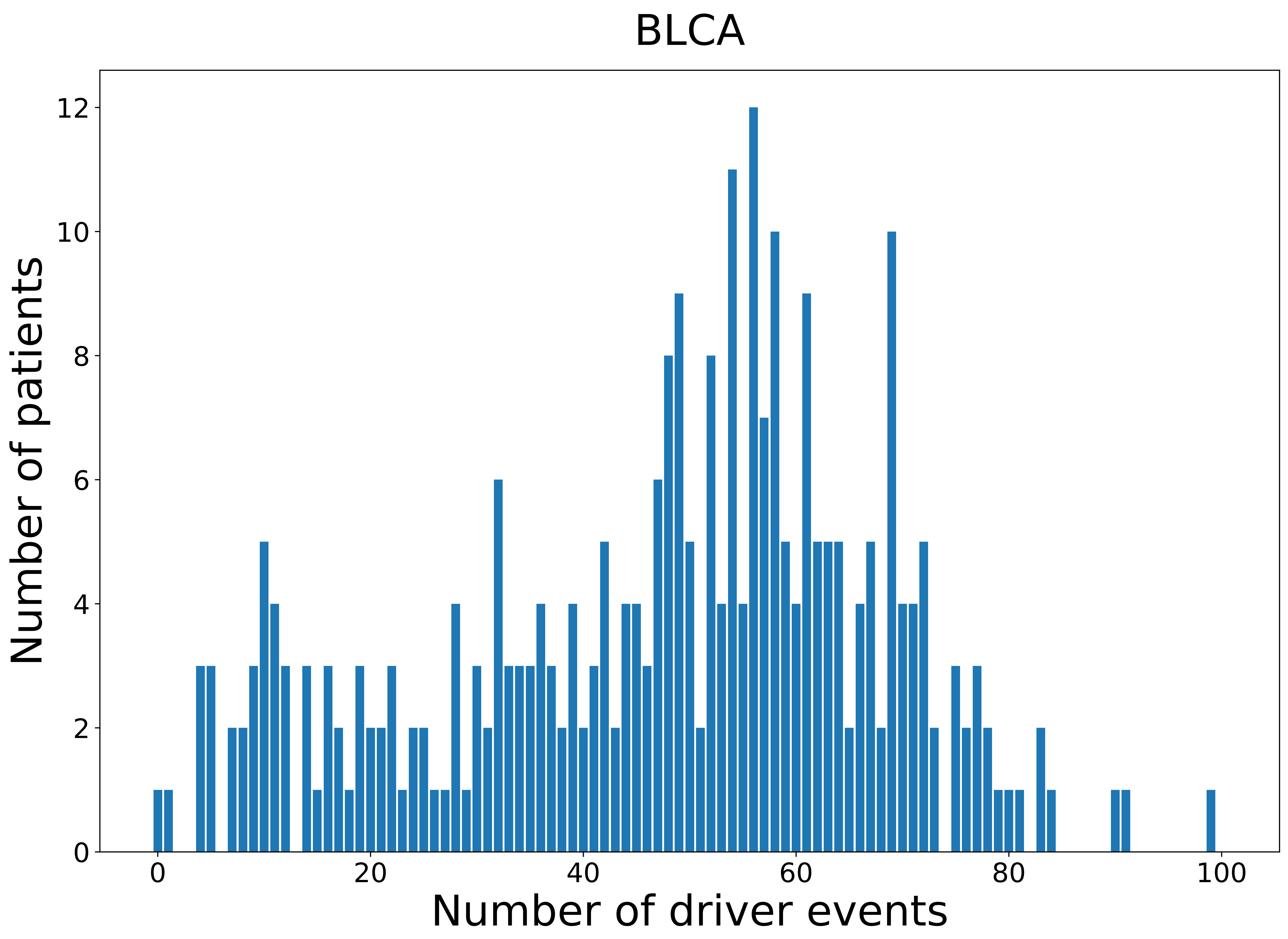

### 2021_8_16_20_32_BLCA_FEMALE.pdf

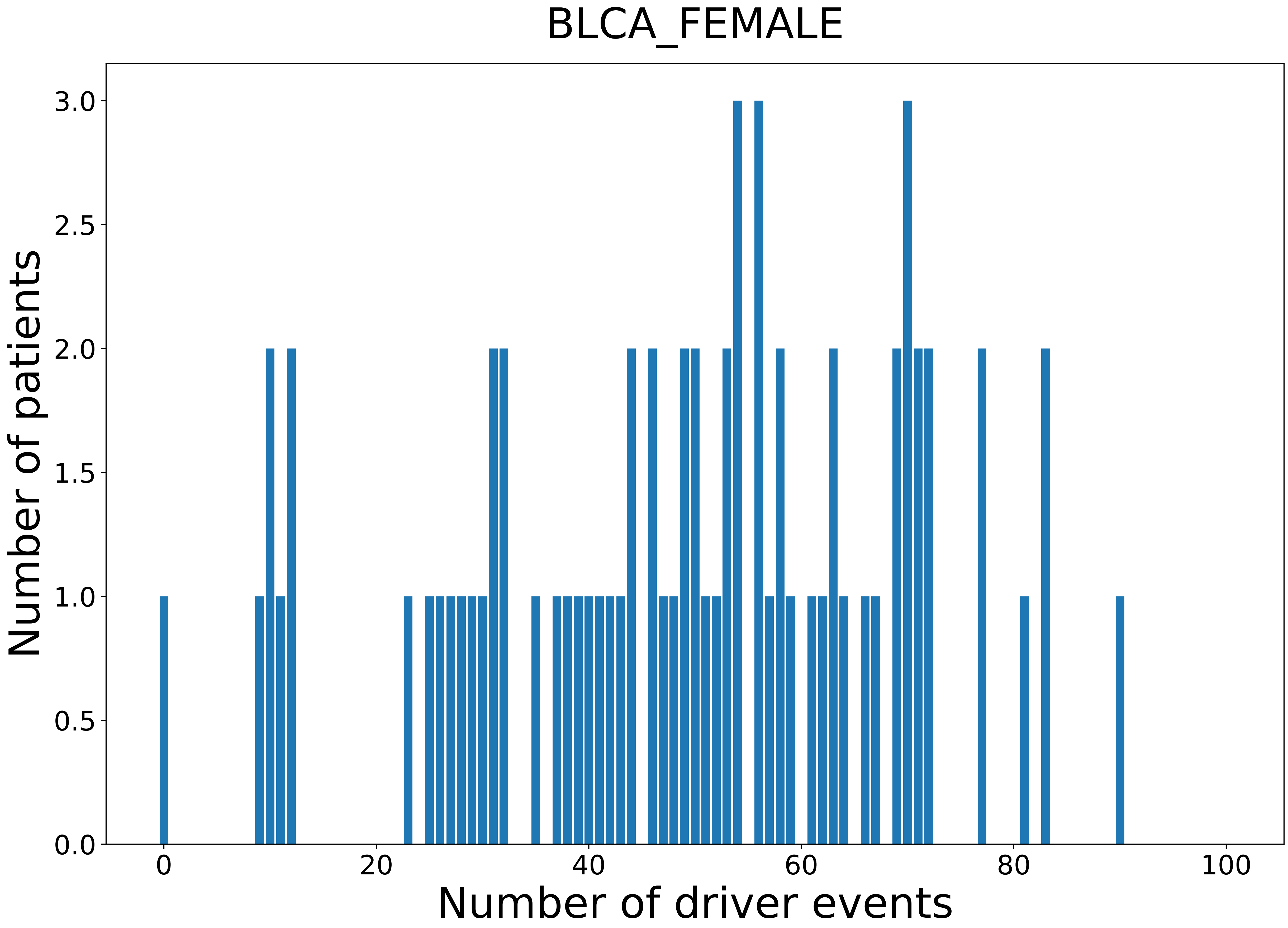

### 2021_8_16_20_32_BLCA_MALE.pdf

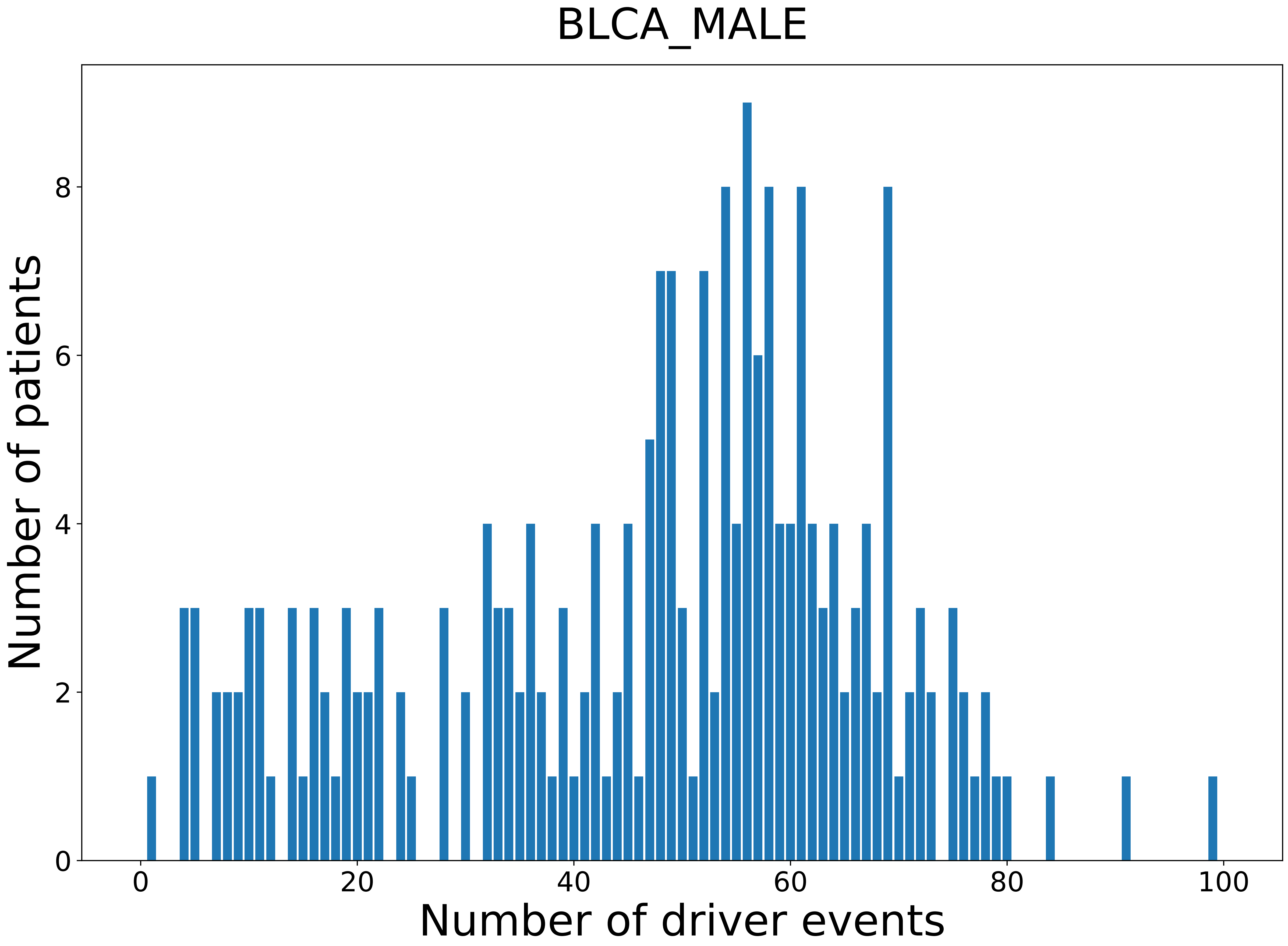

### 2021_8_16_20_32_BRCA.pdf

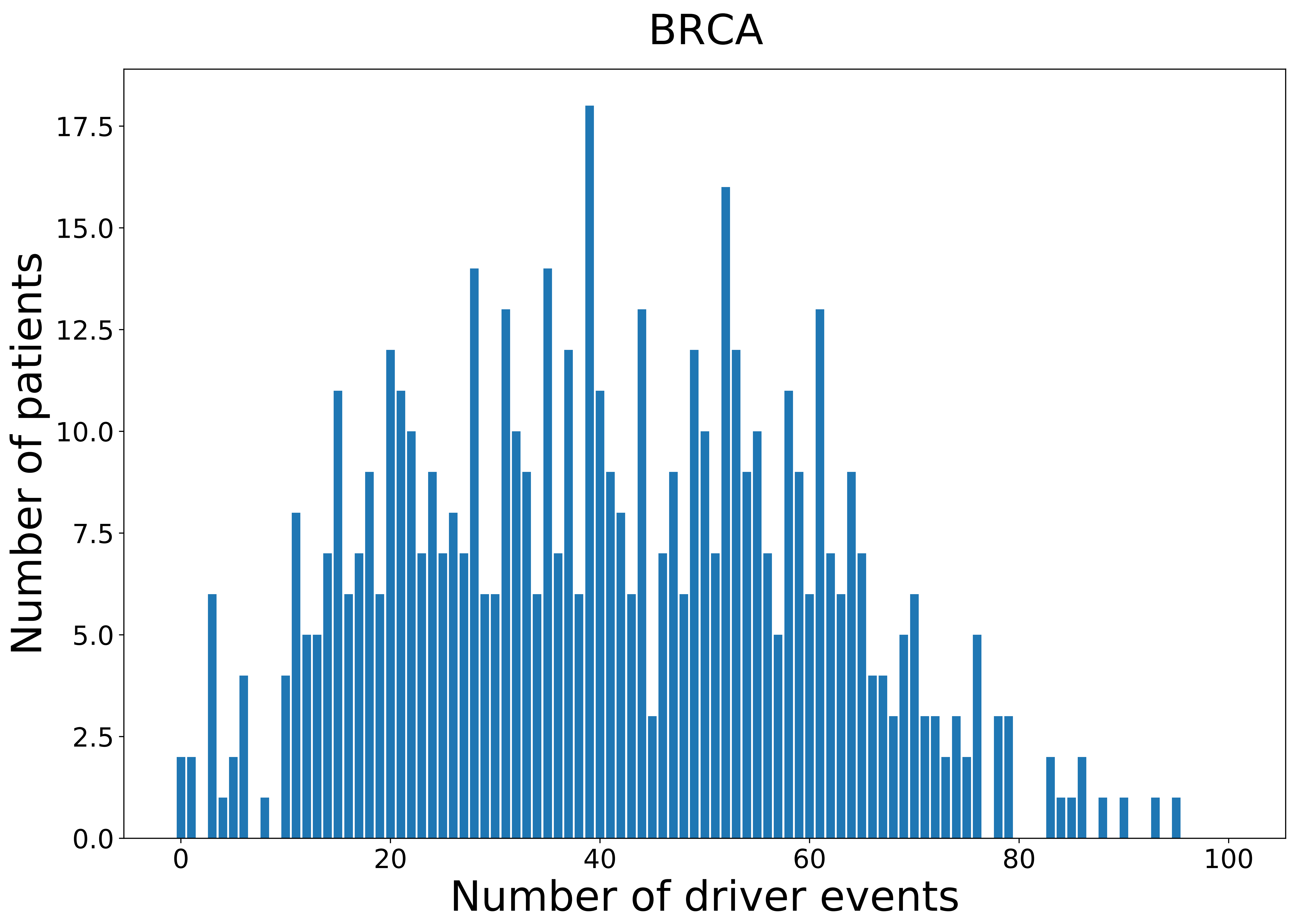

### 2021_8_16_20_32_BRCA_FEMALE.pdf

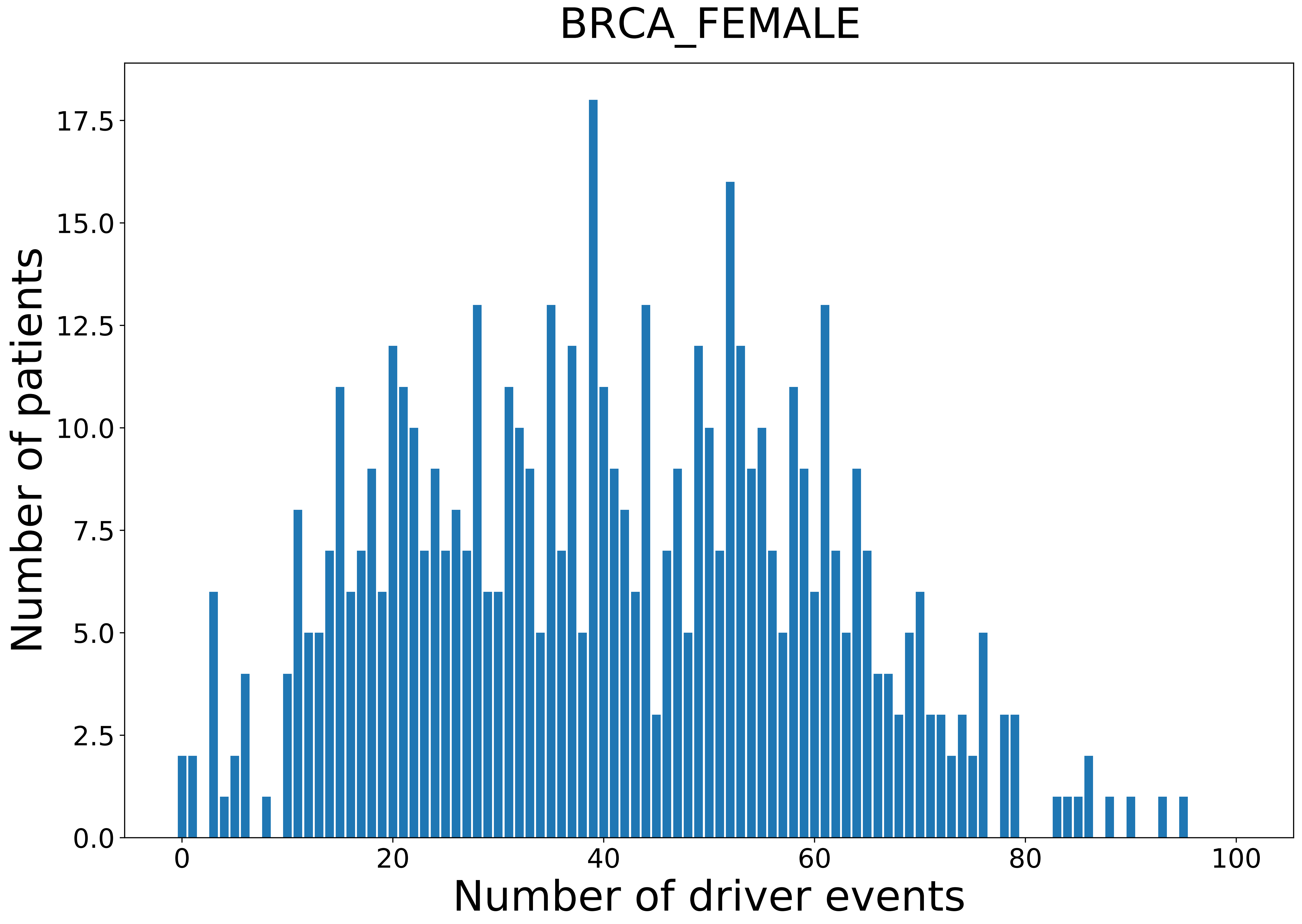

### 2021_8_16_20_32_CESC.pdf

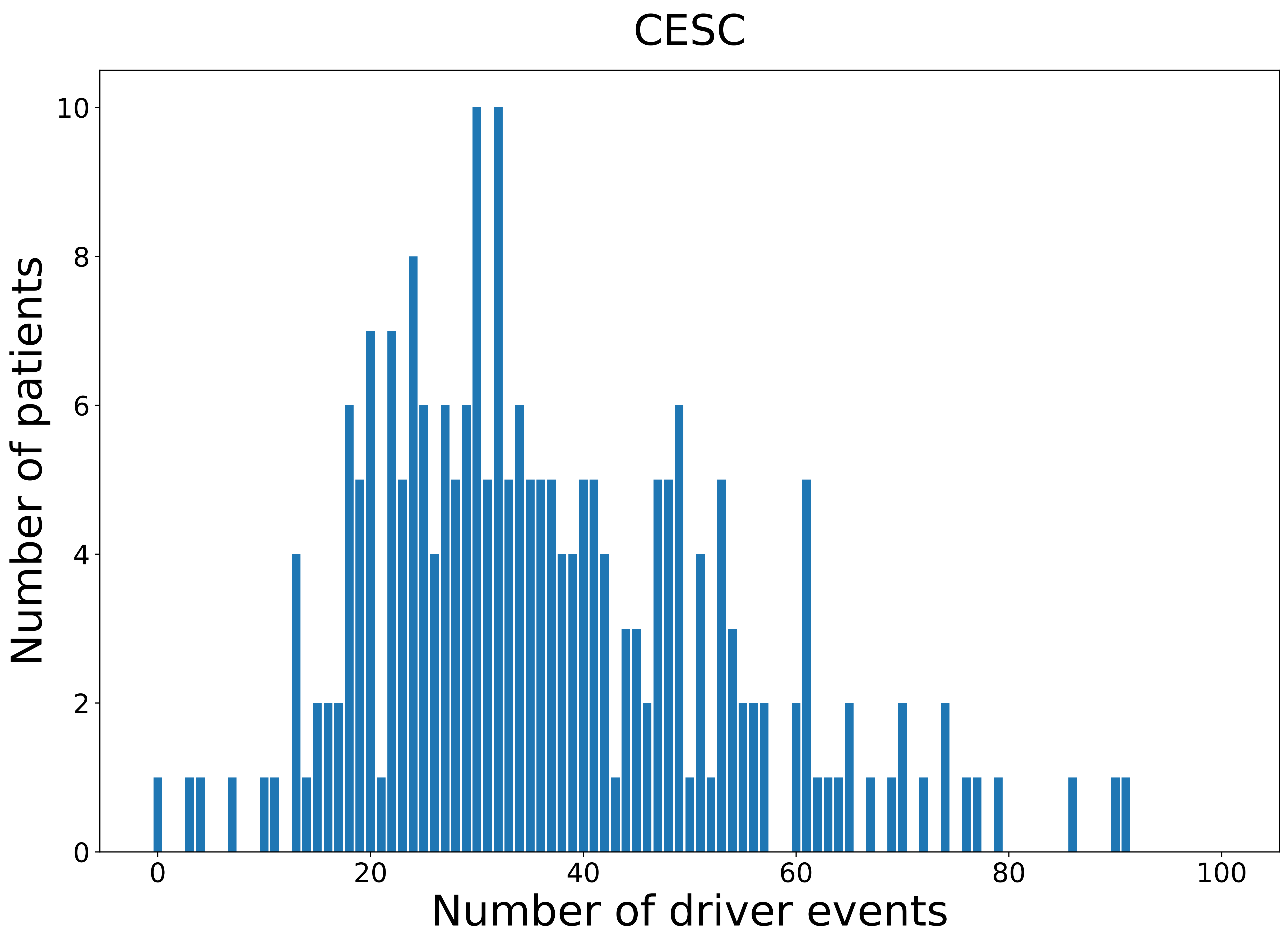

### 2021_8_16_20_32_CESC_FEMALE.pdf

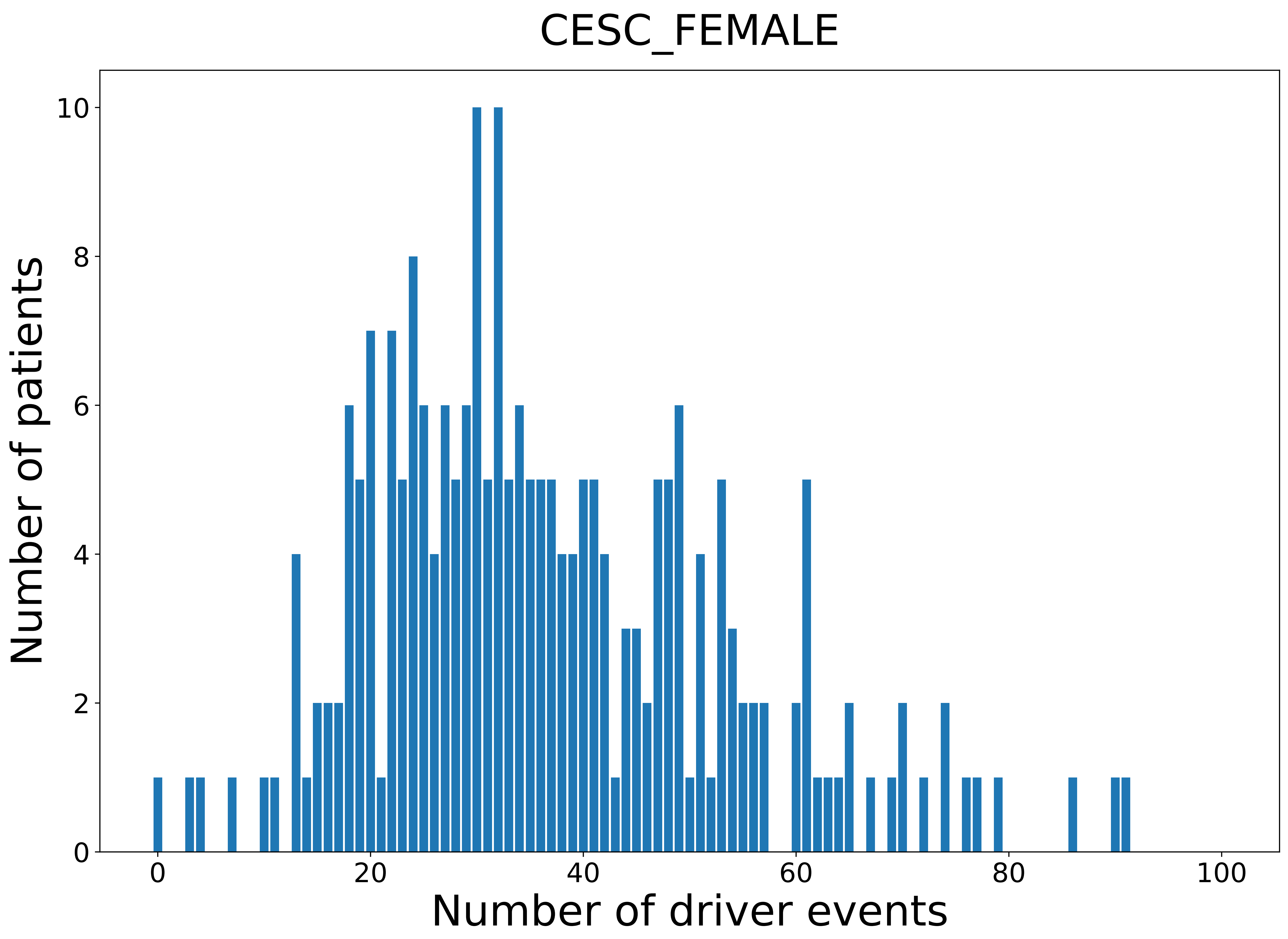

### 2021_8_16_20_32_CHOL.pdf

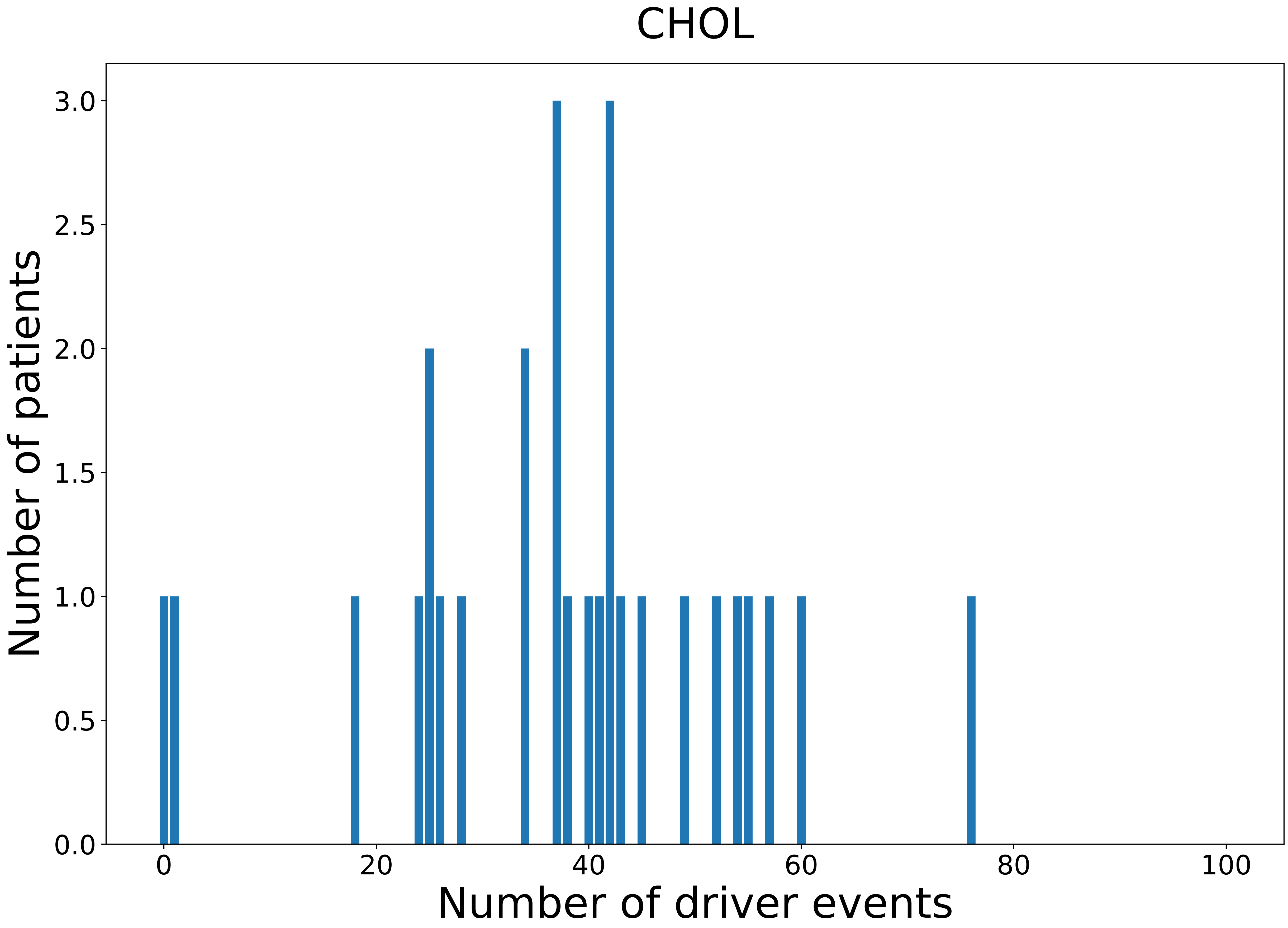

### 2021_8_16_20_32_CHOL_FEMALE.pdf

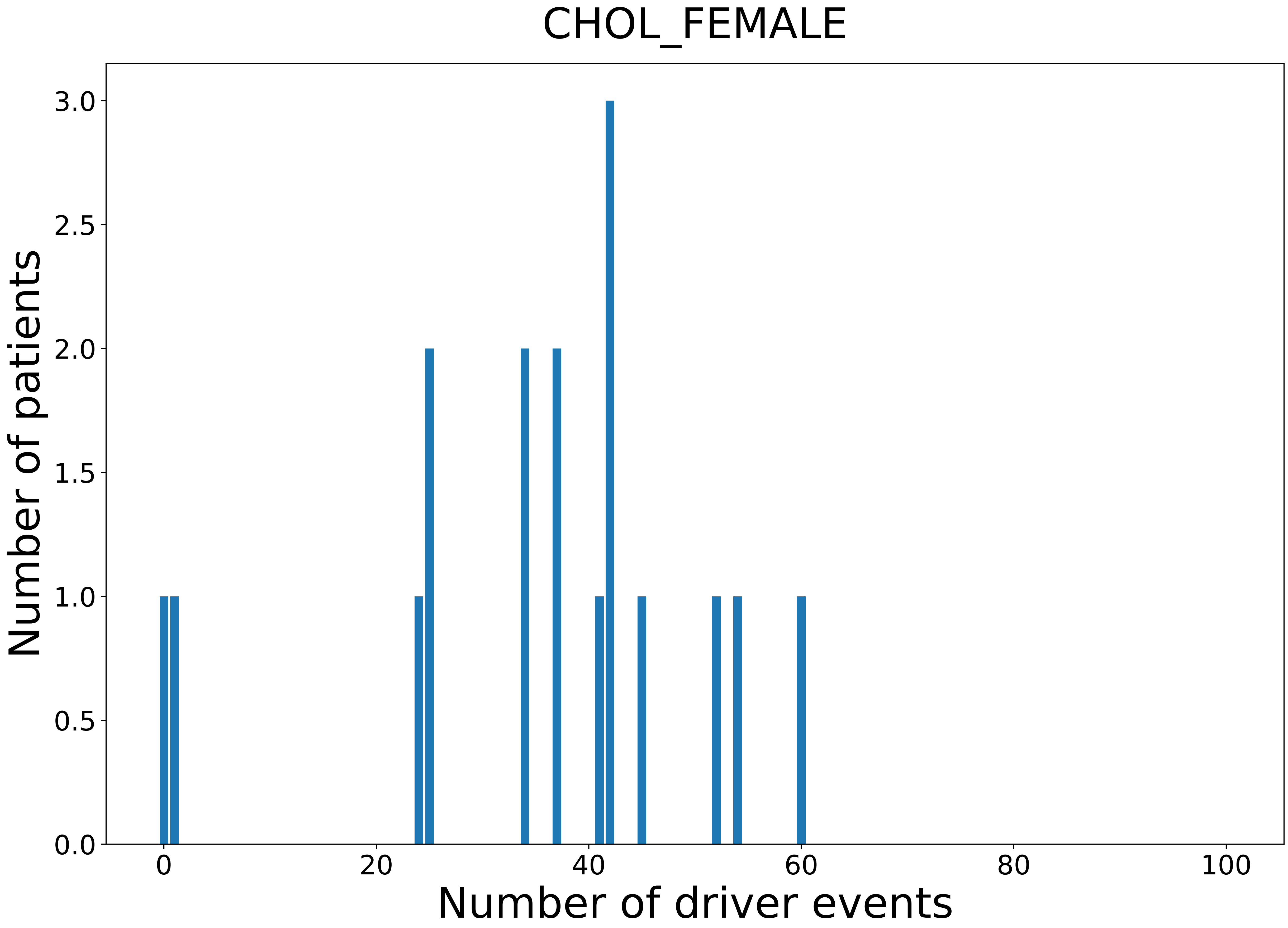

### 2021_8_16_20_32_CHOL_MALE.pdf

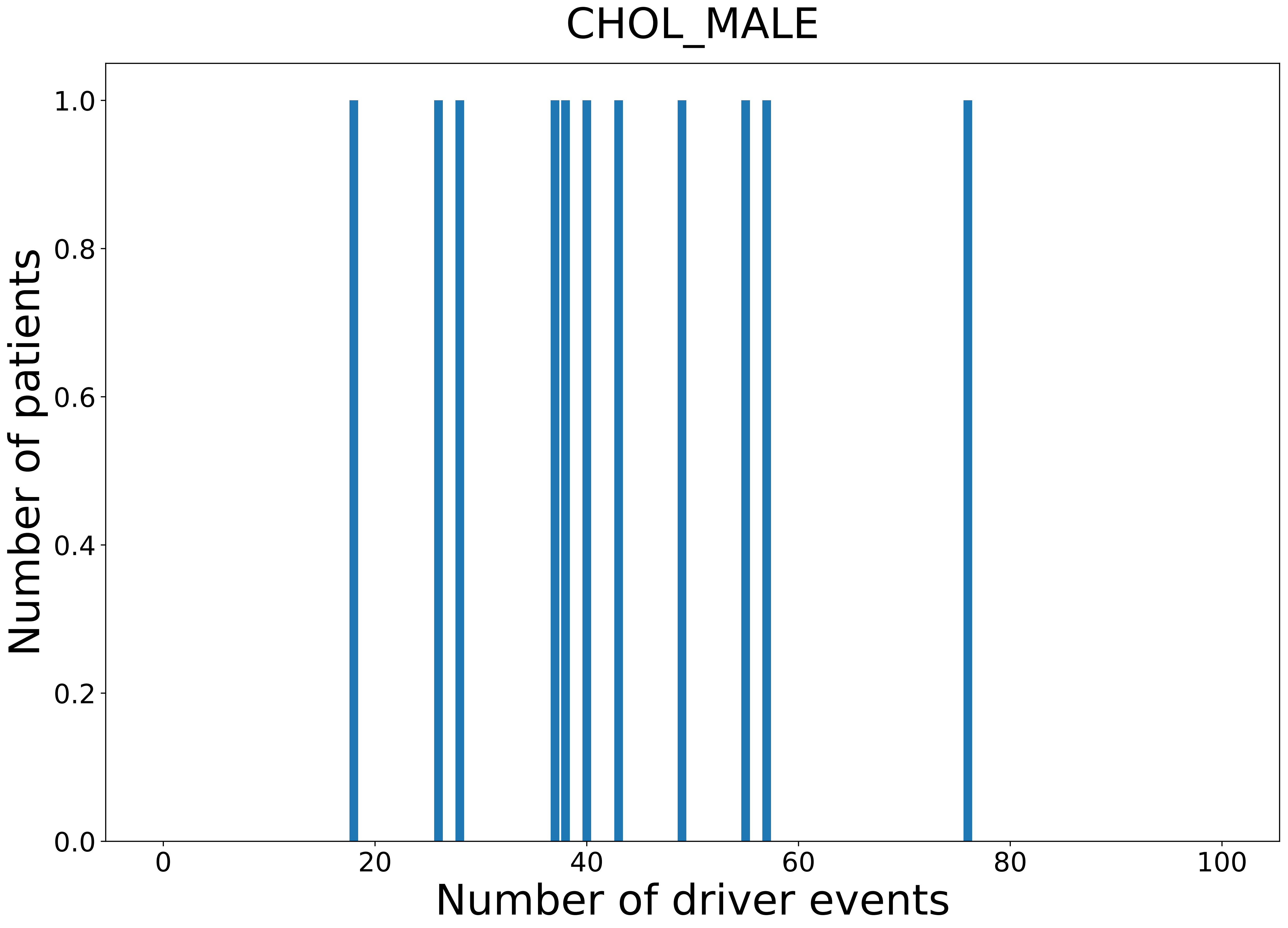

### 2021_8_16_20_32_COAD.pdf

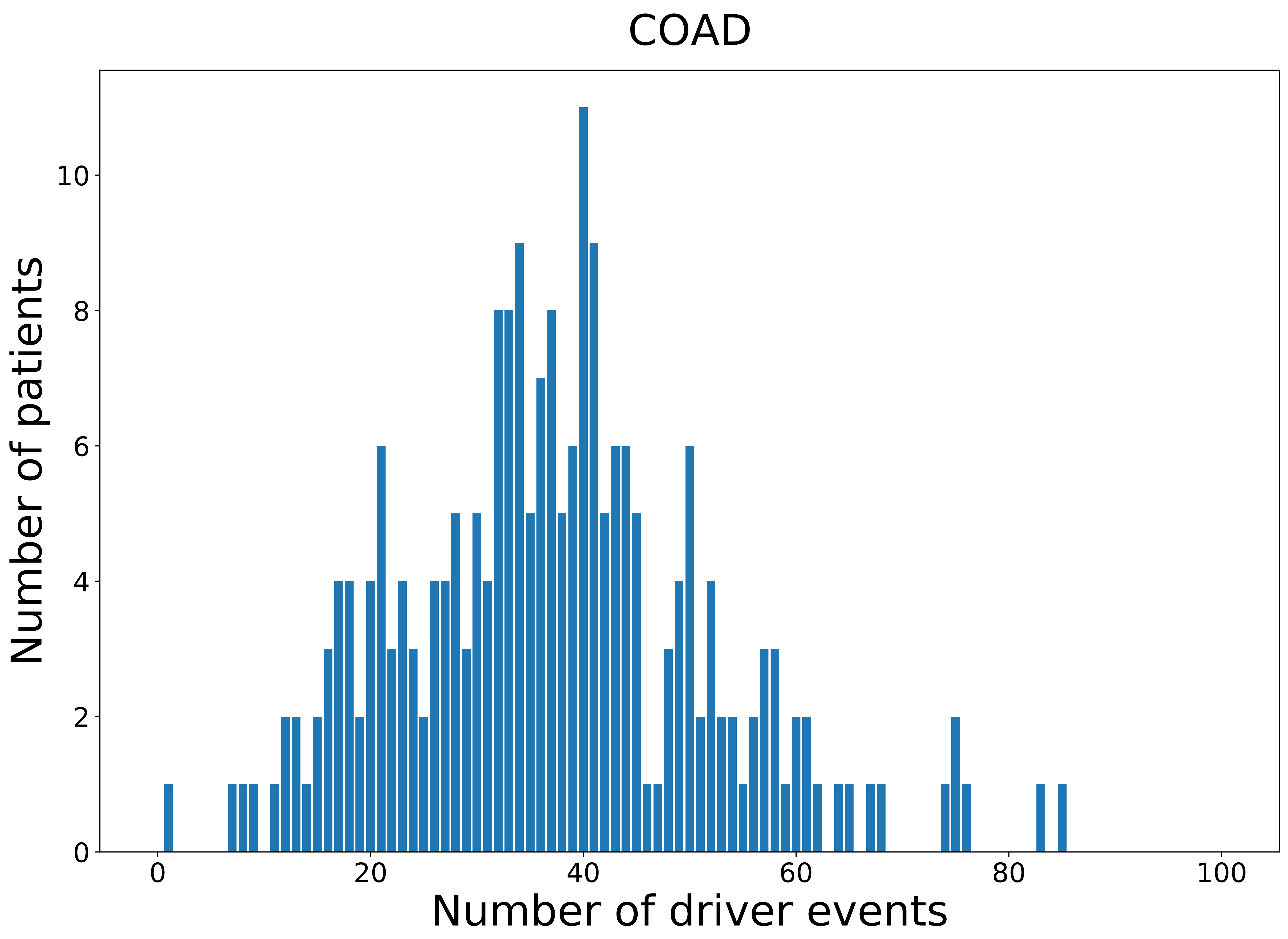

### 2021_8_16_20_32_COAD_FEMALE.pdf

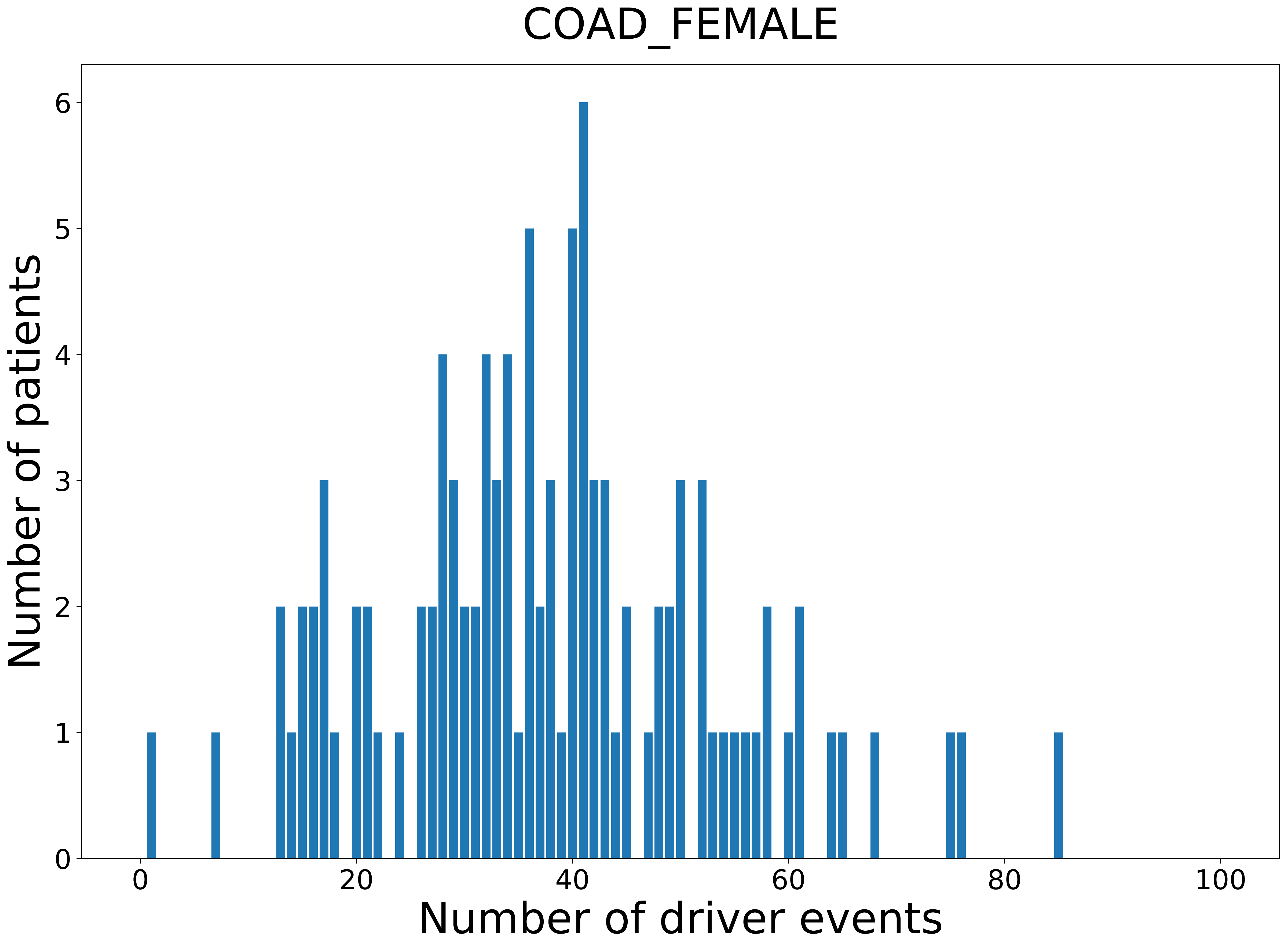

### 2021_8_16_20_32_COAD_MALE.pdf

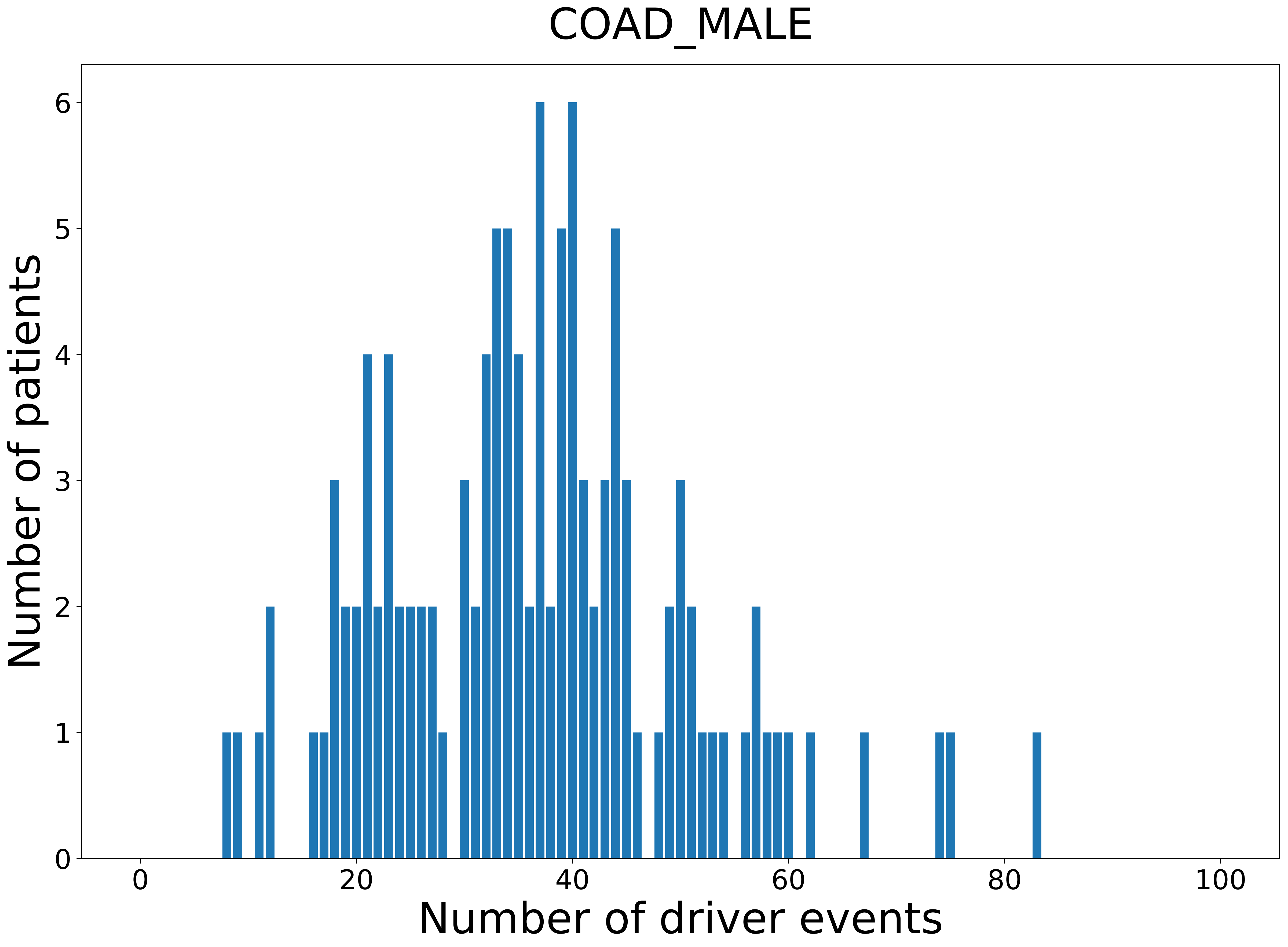

### 2021_8_16_20_32_distribution_age.pdf

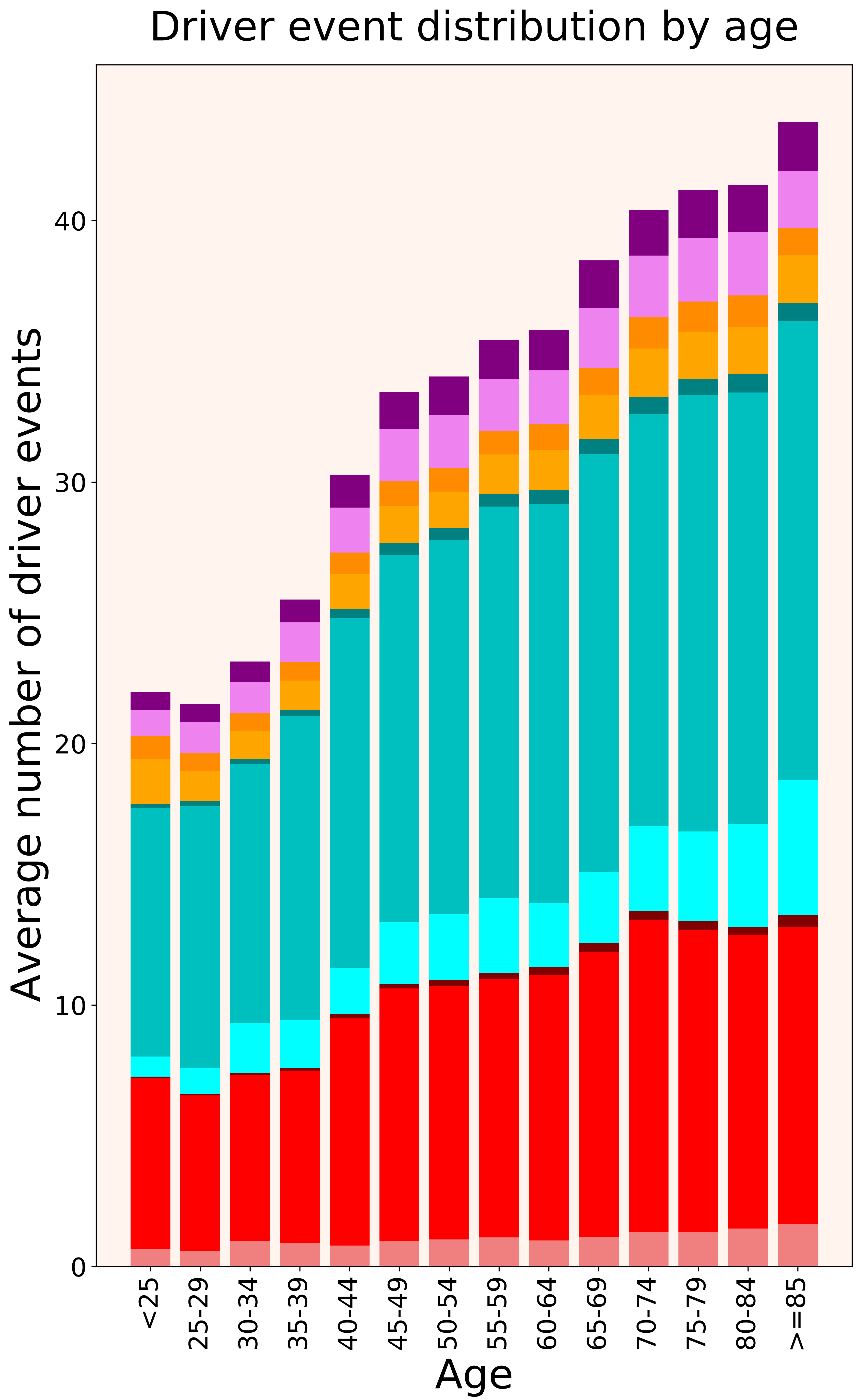

### 2021_8_16_20_32_distribution_age_females.pdf

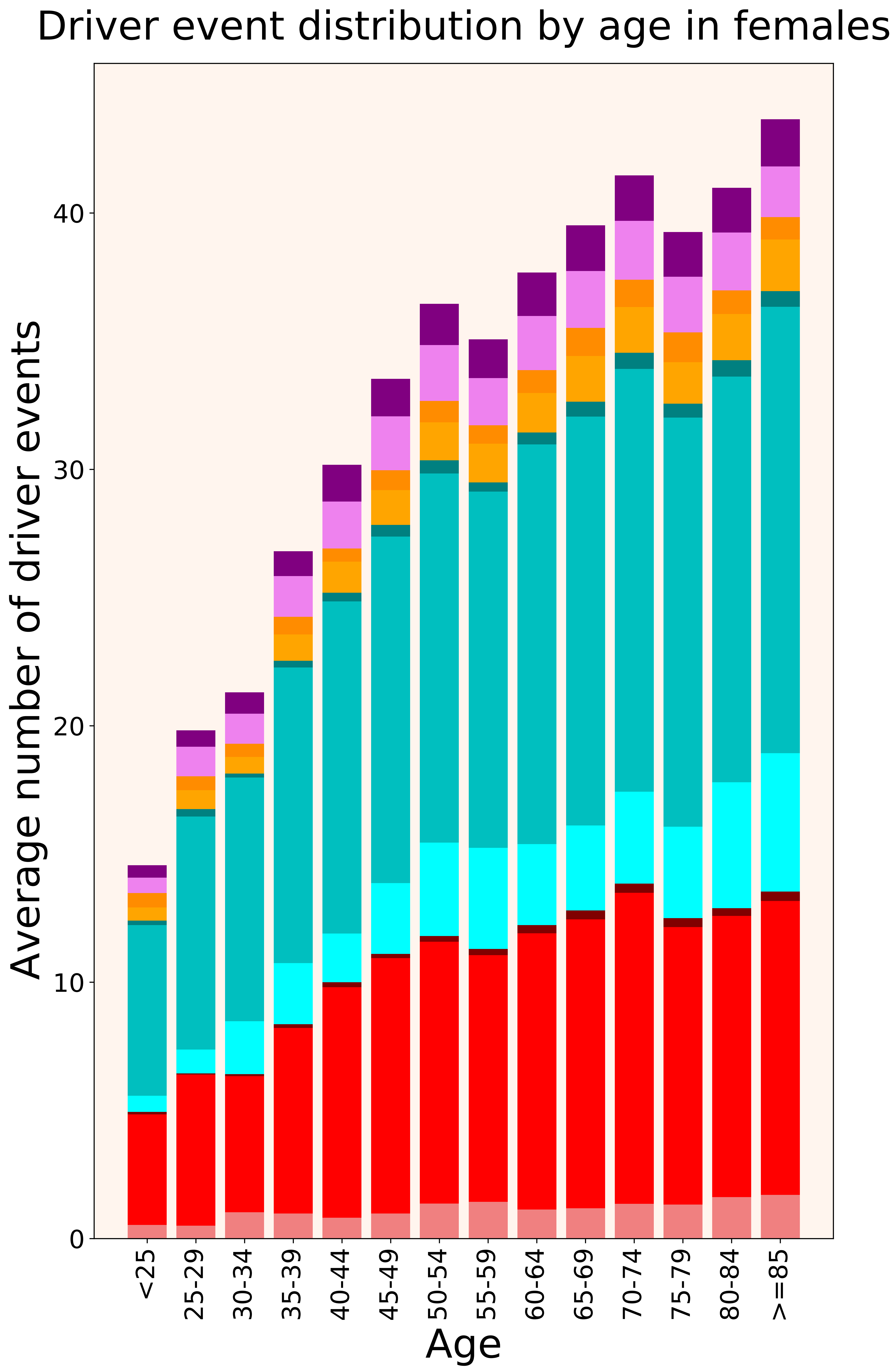

### 2021_8_16_20_32_distribution_age_males.pdf

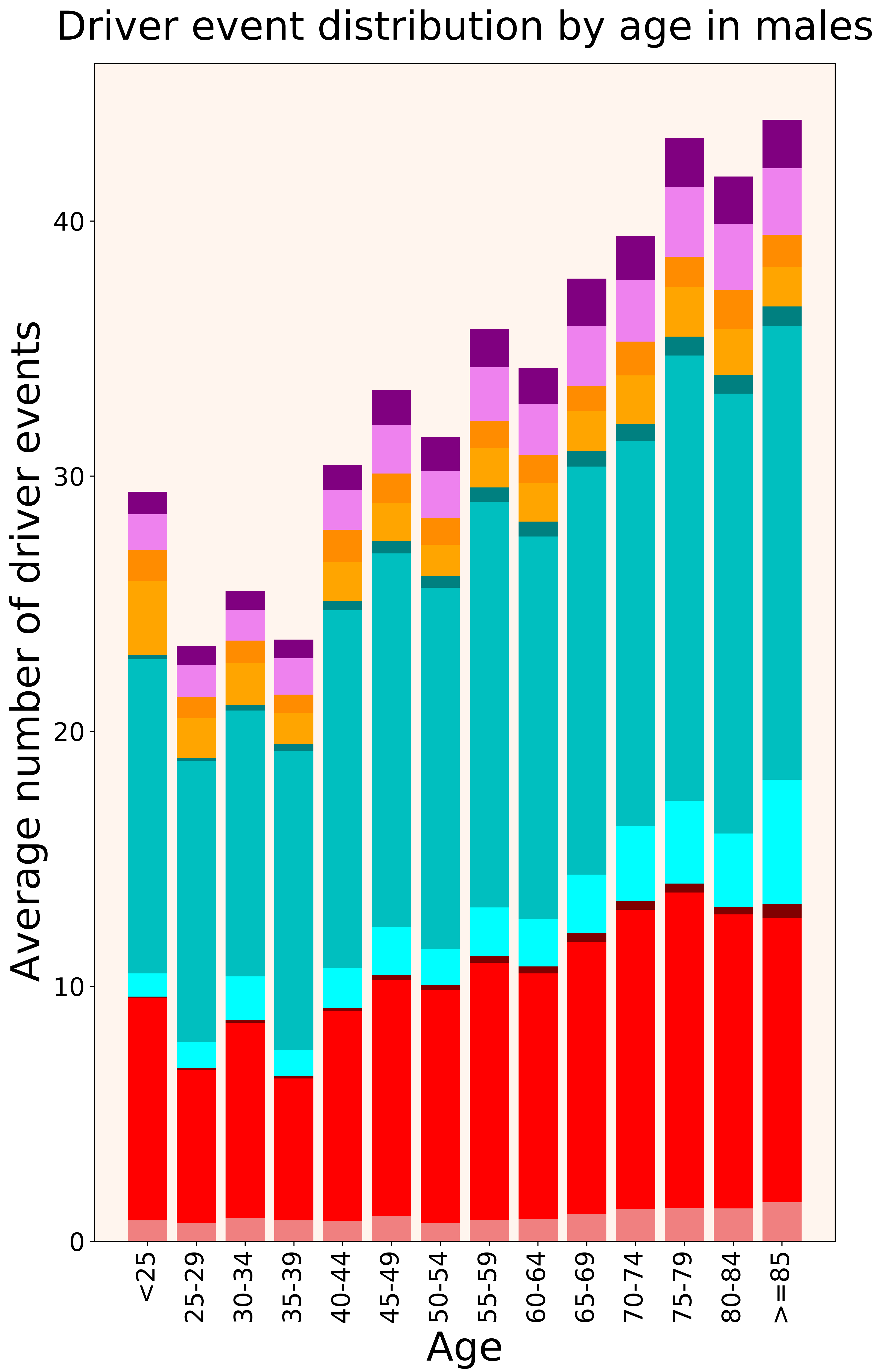

### 2021_8_16_20_32_distribution_cohorts.pdf

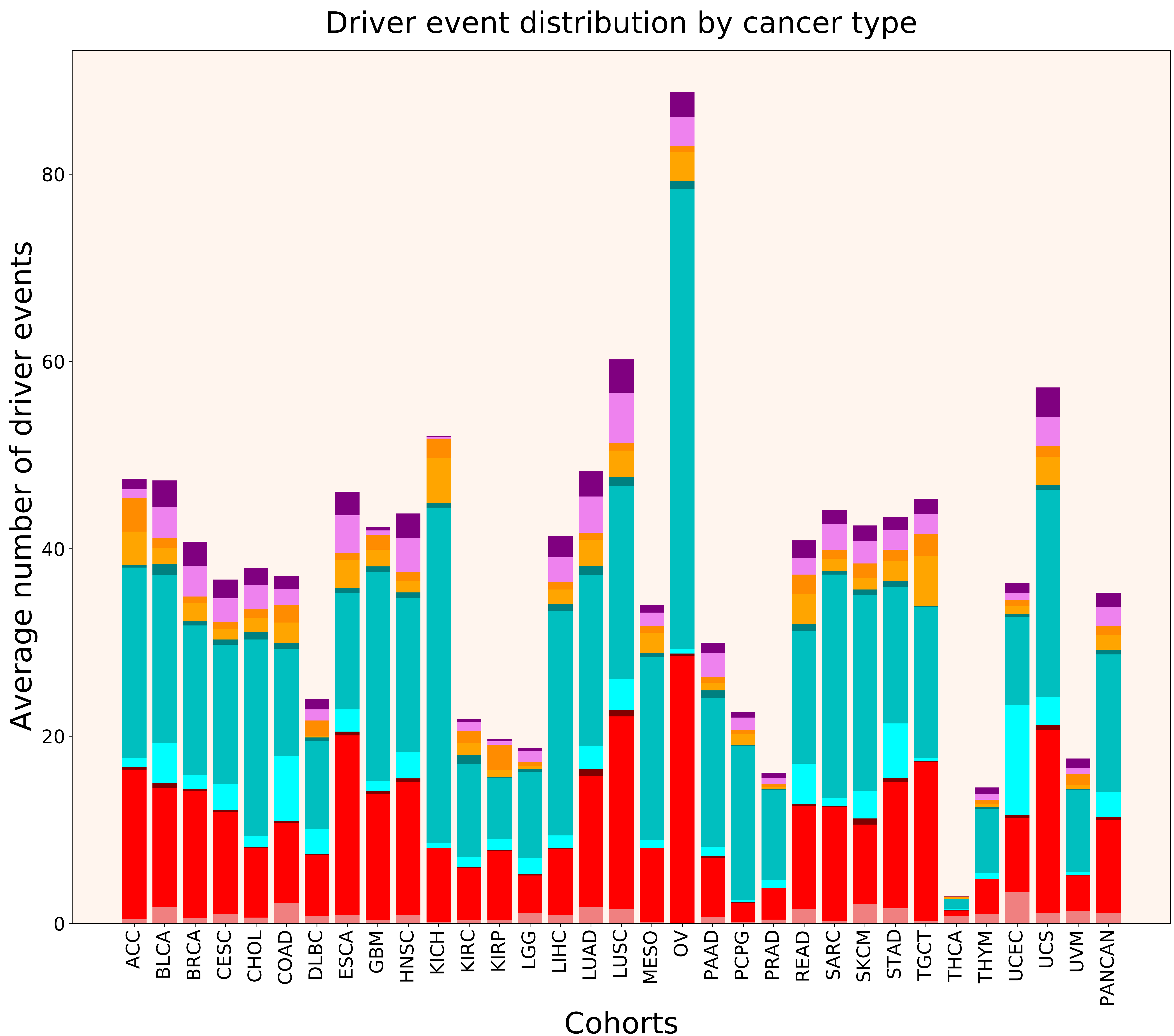

### 2021_8_16_20_32_distribution_cohorts_females.pdf

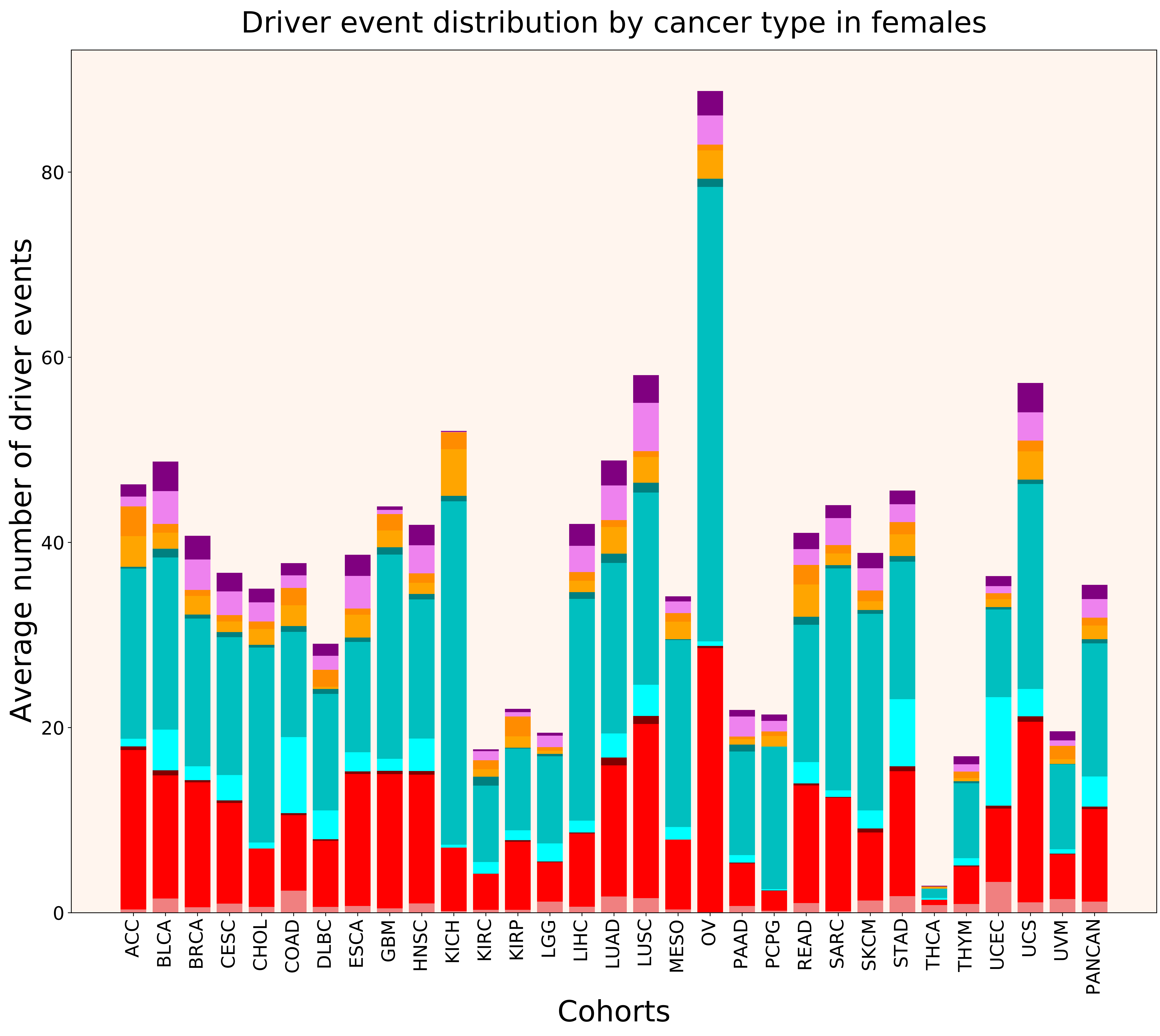

### 2021_8_16_20_32_distribution_cohorts_males.pdf

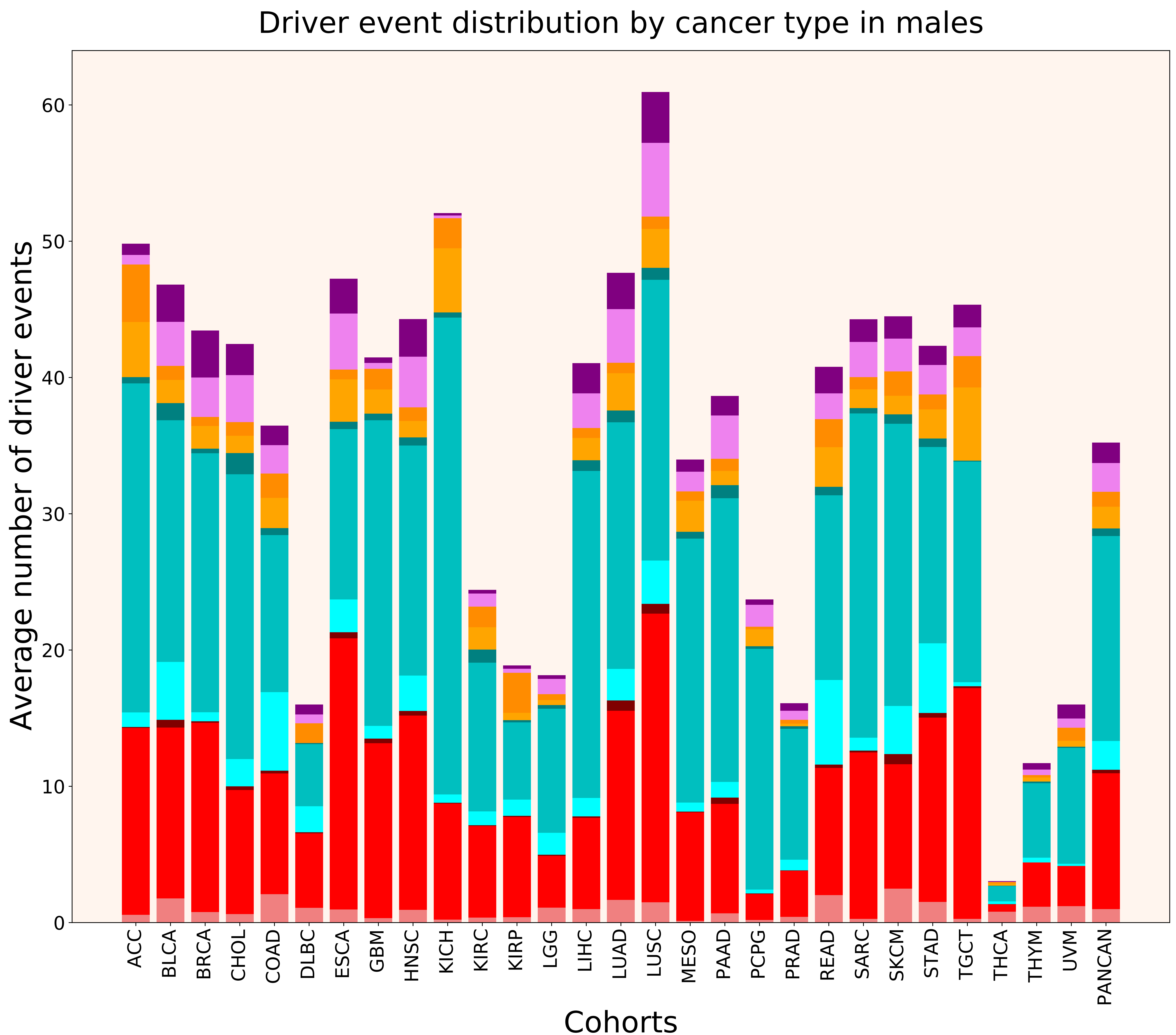

### 2021_8_16_20_32_distribution_events_detailed.pdf

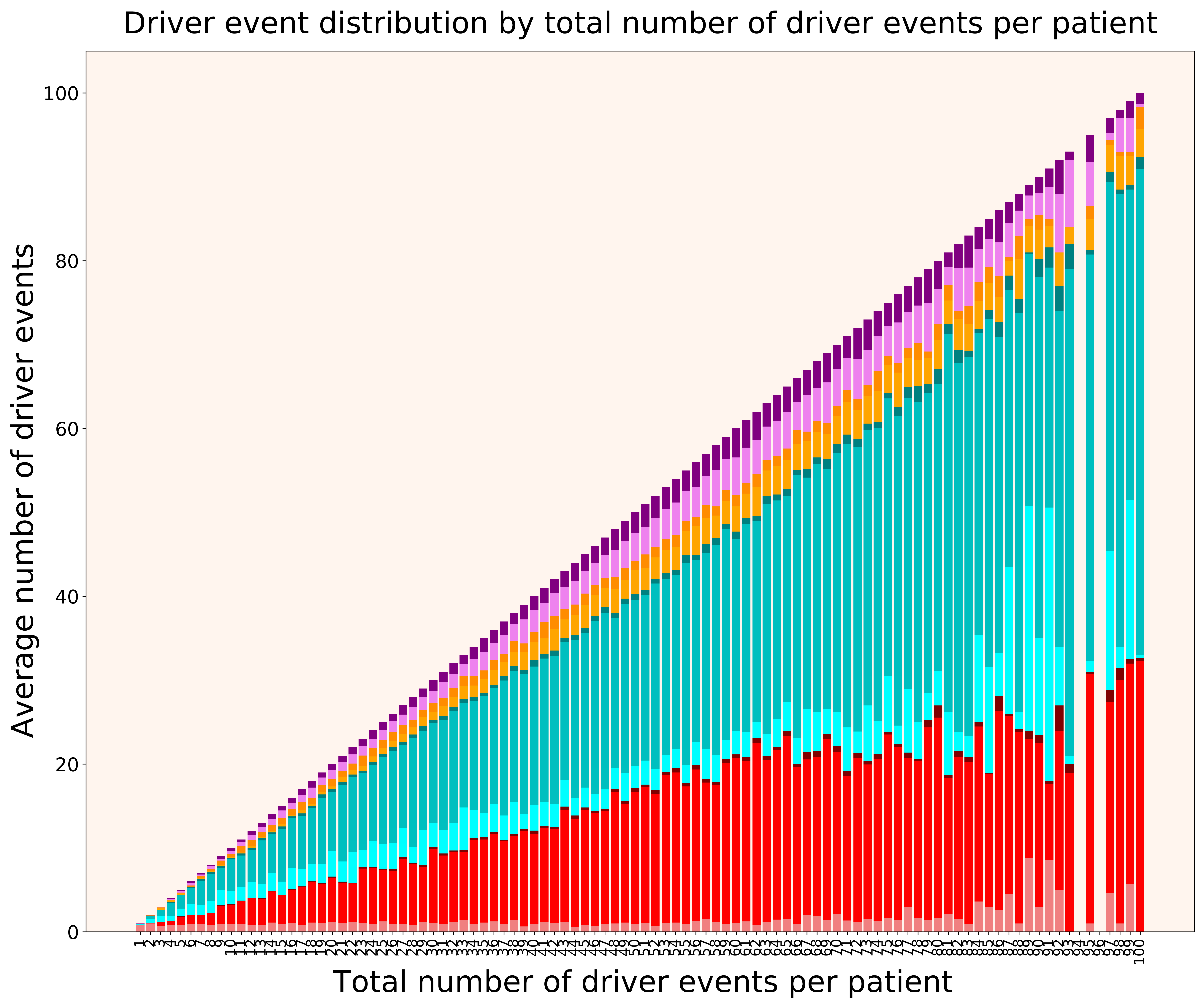

### 2021_8_16_20_32_distribution_events_detailed_females.pdf

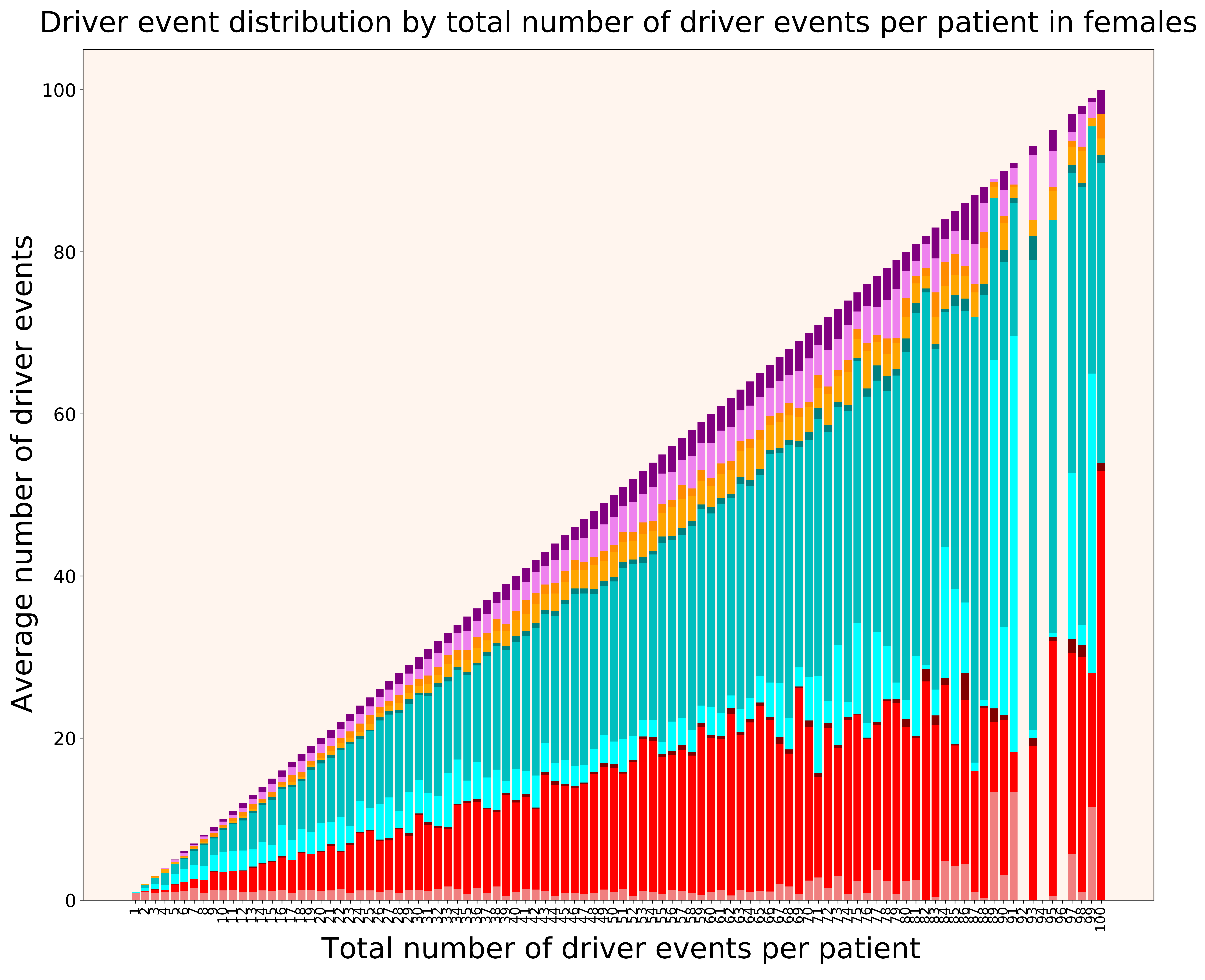

### 2021_8_16_20_32_distribution_events_detailed_males.pdf

#### Driver event distribution by total number of driver events per patient in males

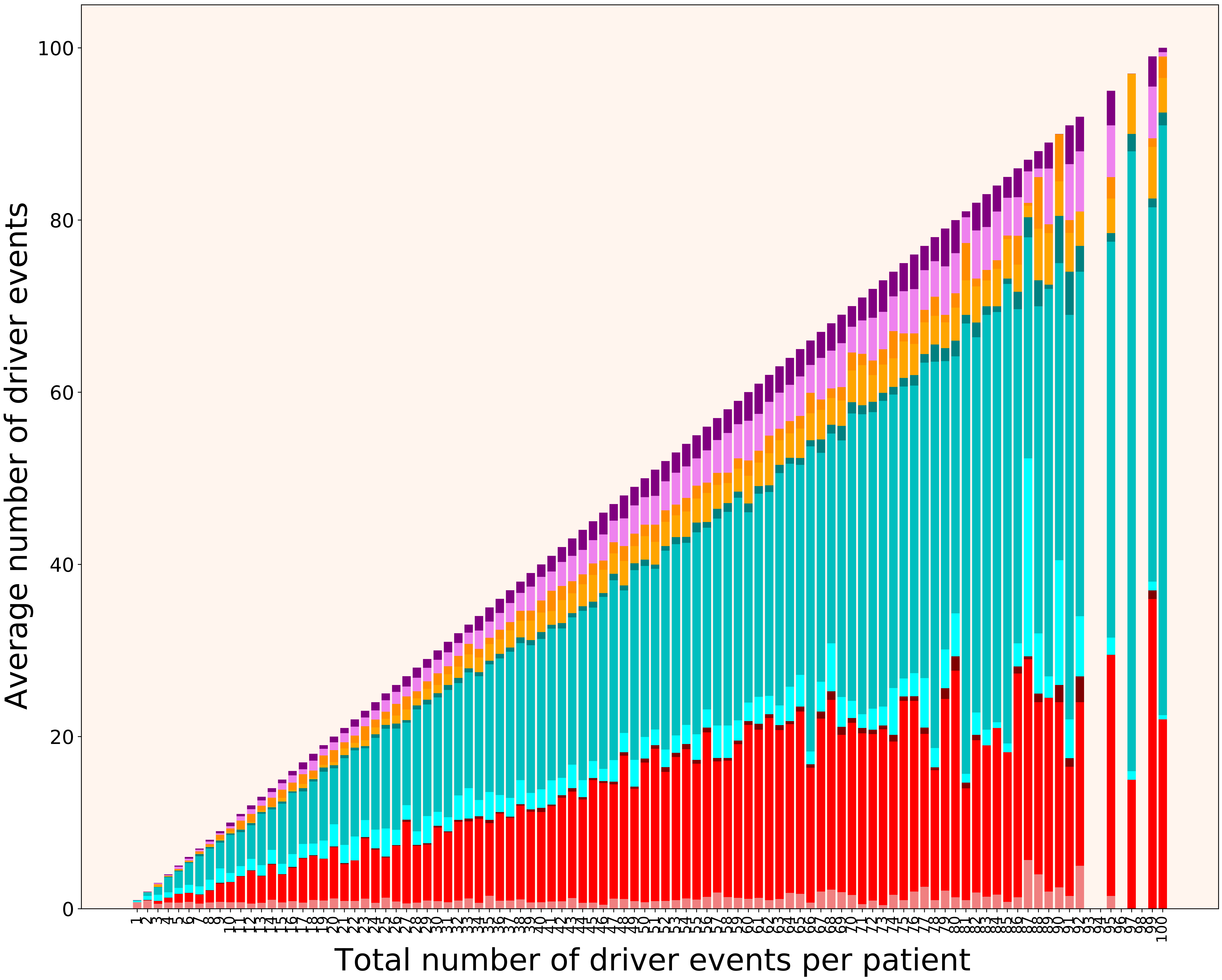

### 2021_8_16_20_32_distribution_gender.pdf

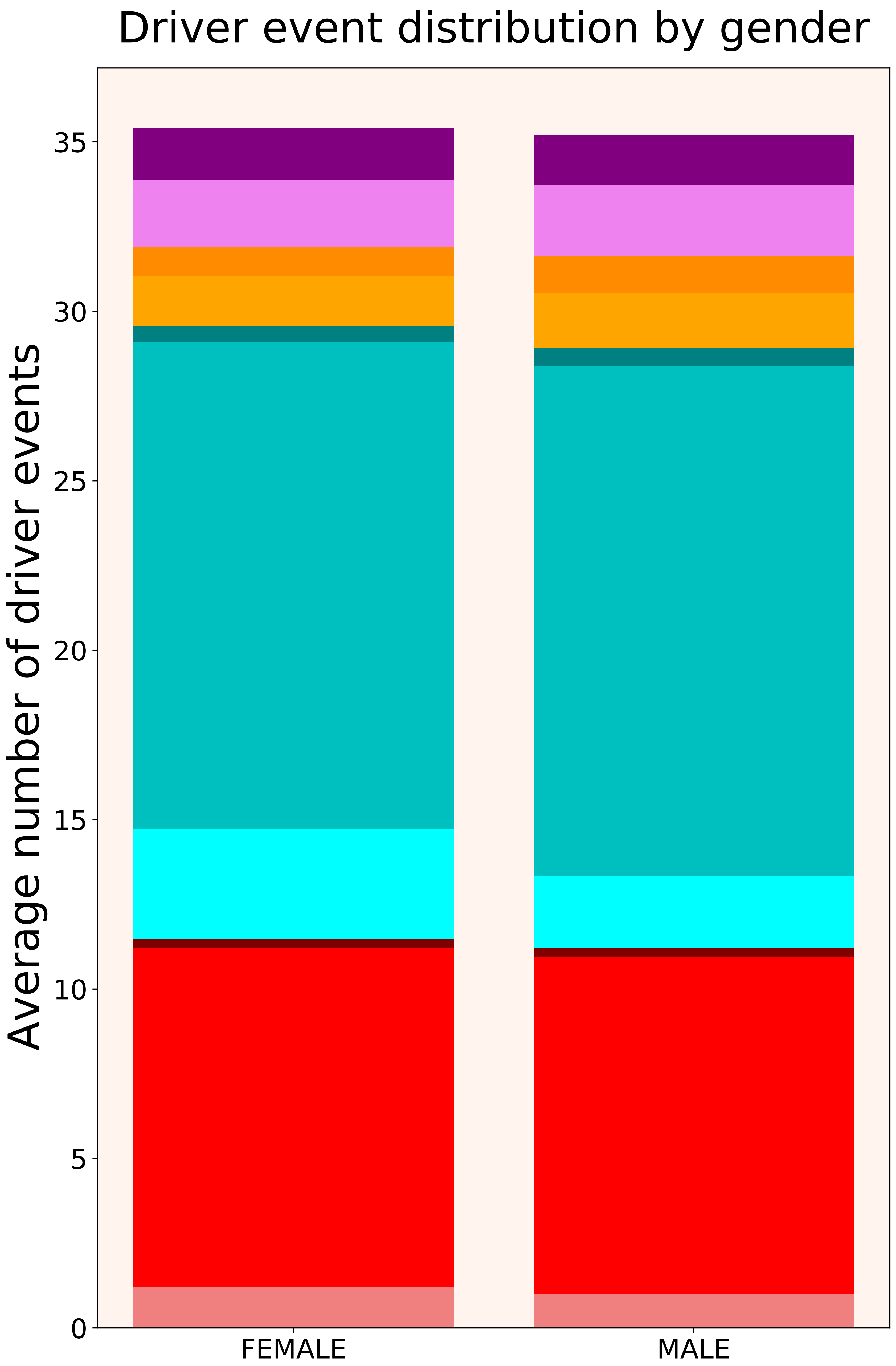

### 2021_8_16_20_32_distribution_stages.pdf

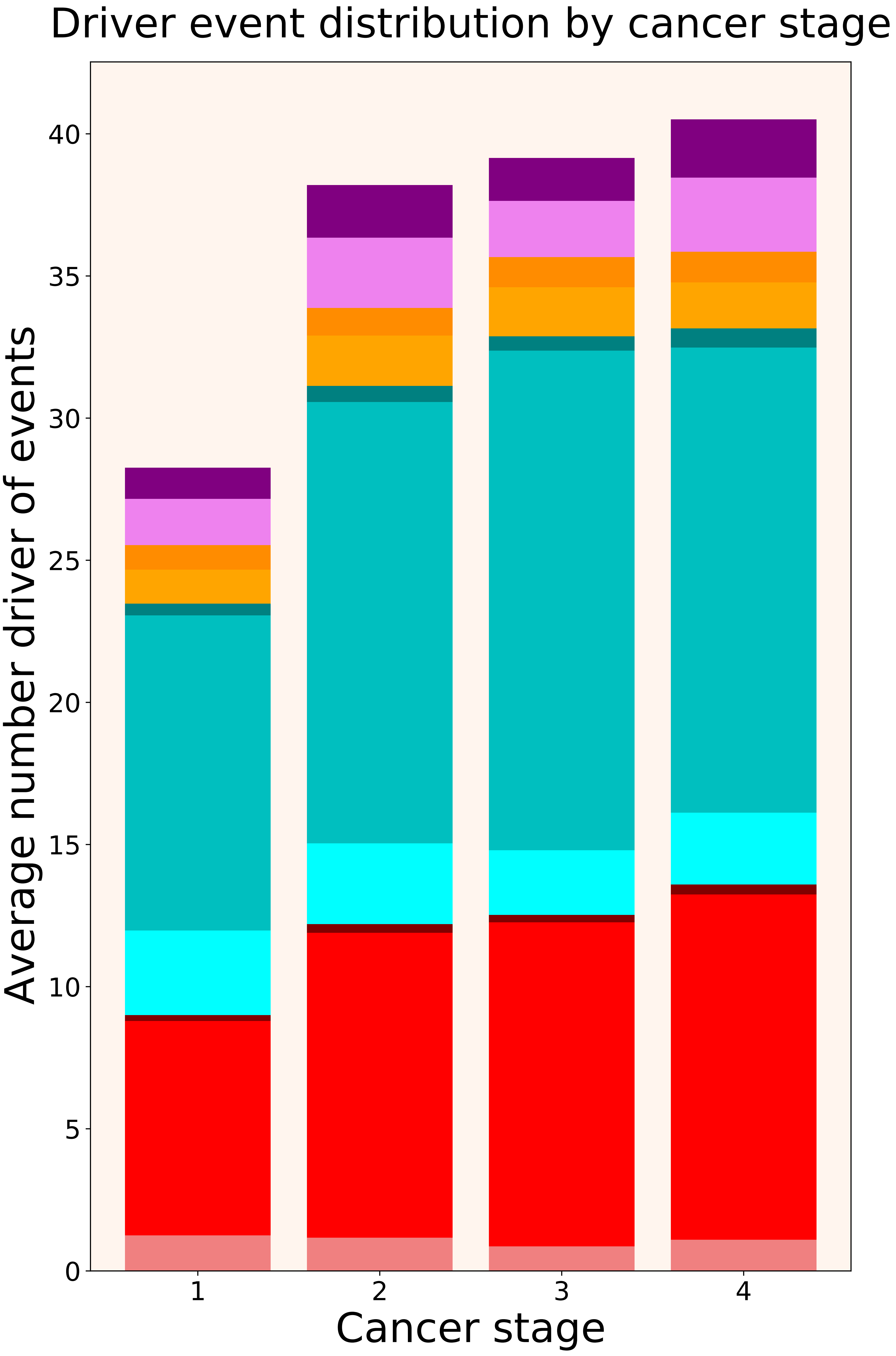

### 2021_8_16_20_32_distribution_stages_females.pdf

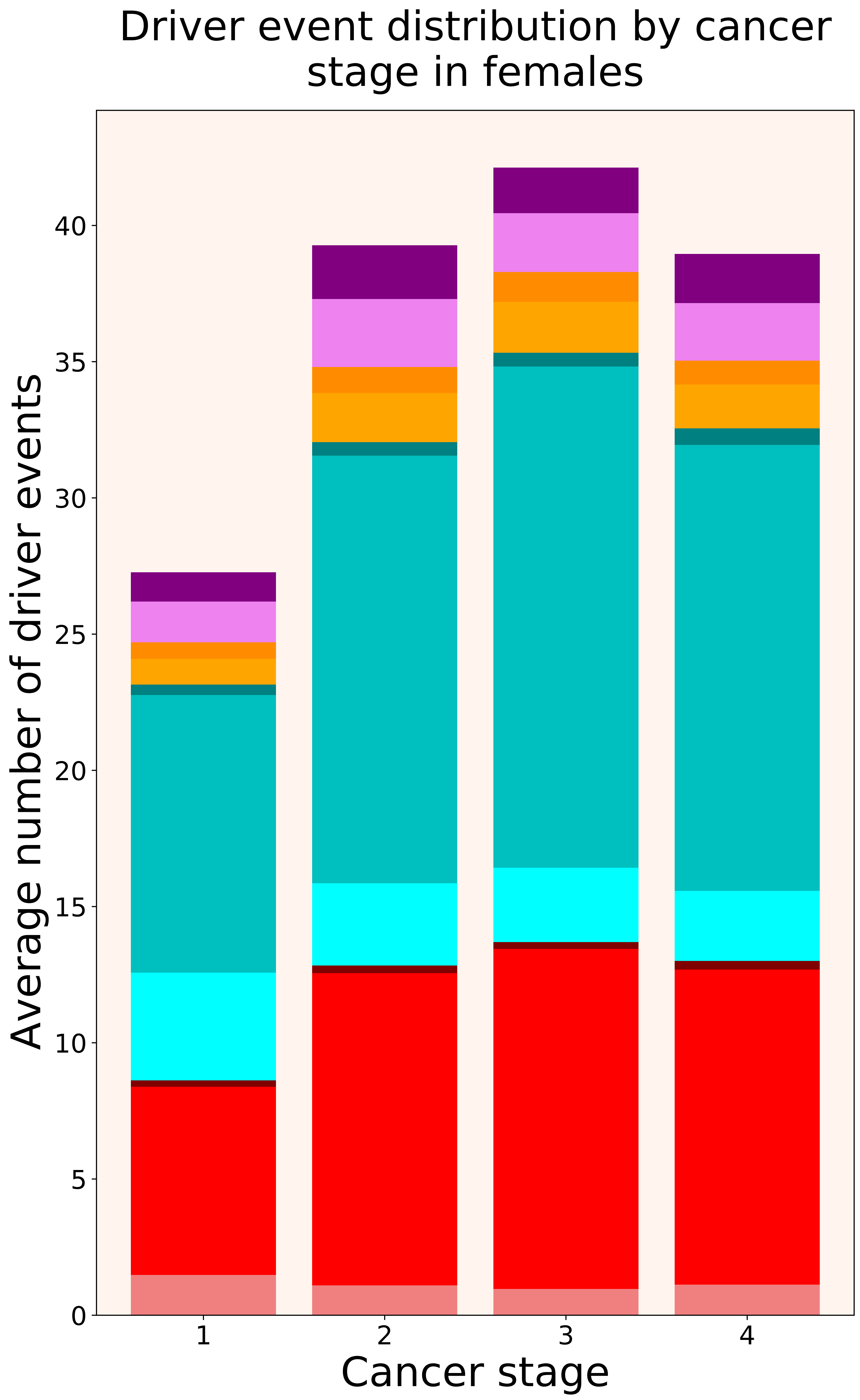

### 2021_8_16_20_32_distribution_stages_males.pdf

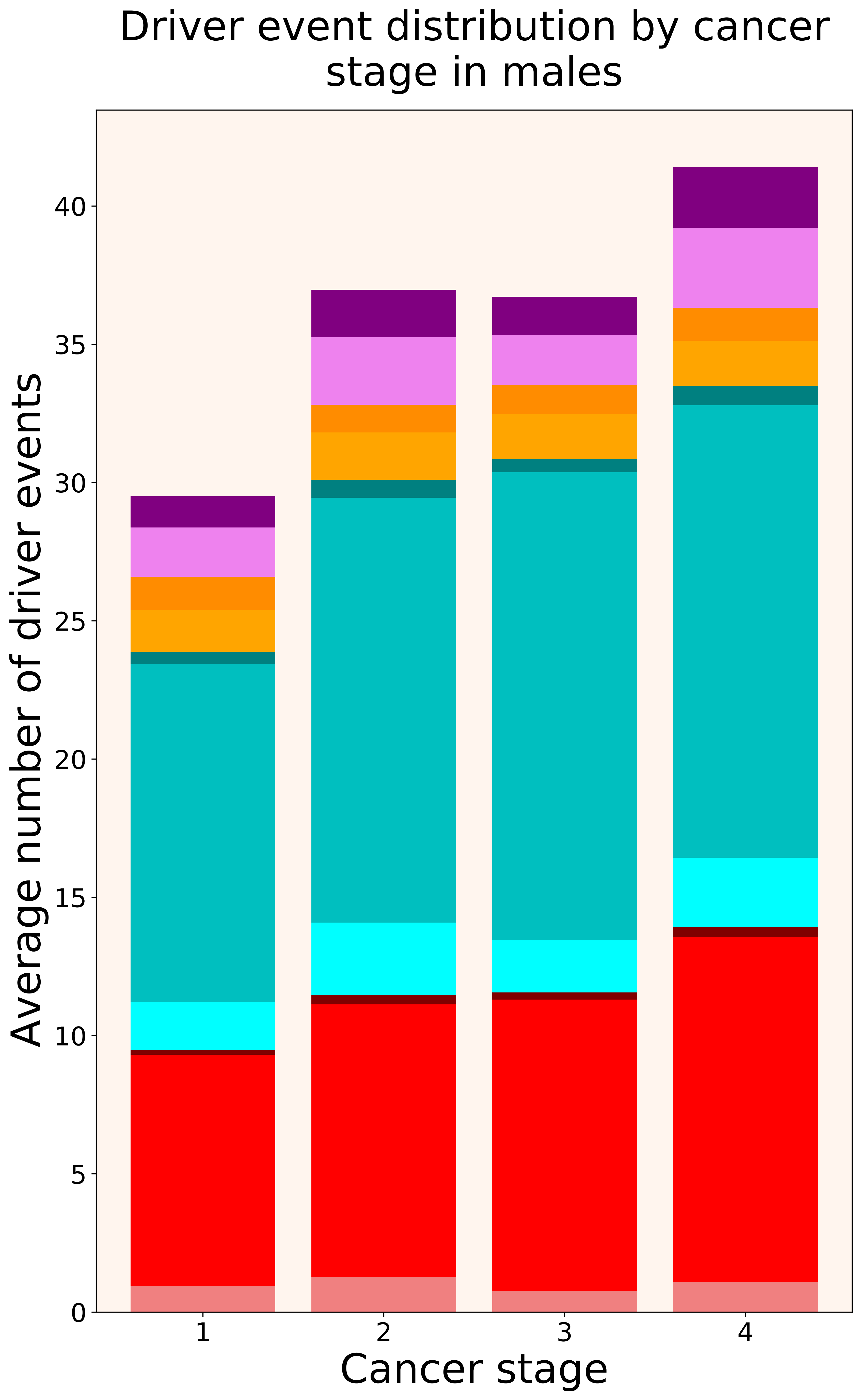

### 2021_8_16_20_32_DLBC.pdf

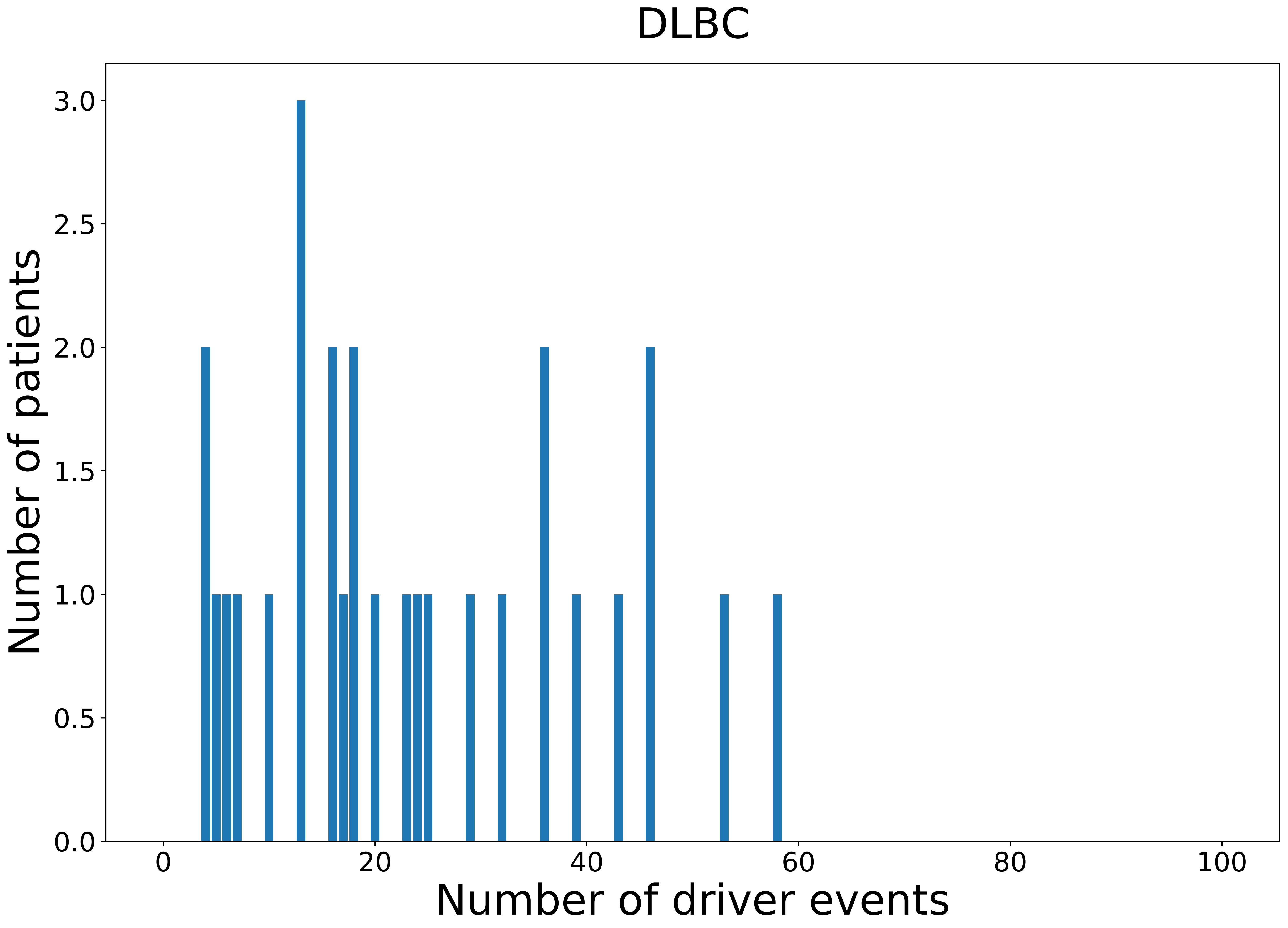

### 2021_8_16_20_32_DLBC_FEMALE.pdf

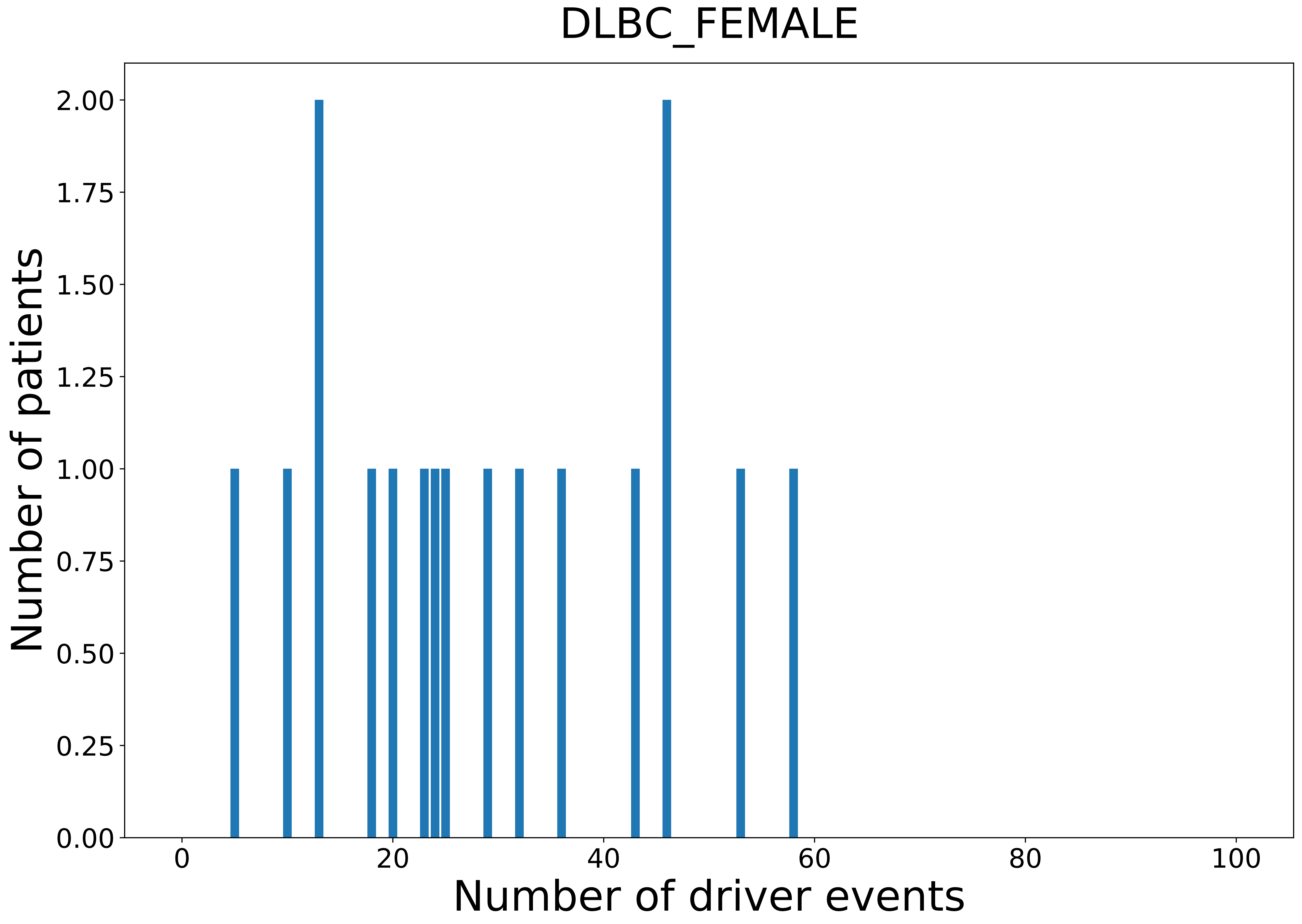

### 2021_8_16_20_32_DLBC_MALE.pdf

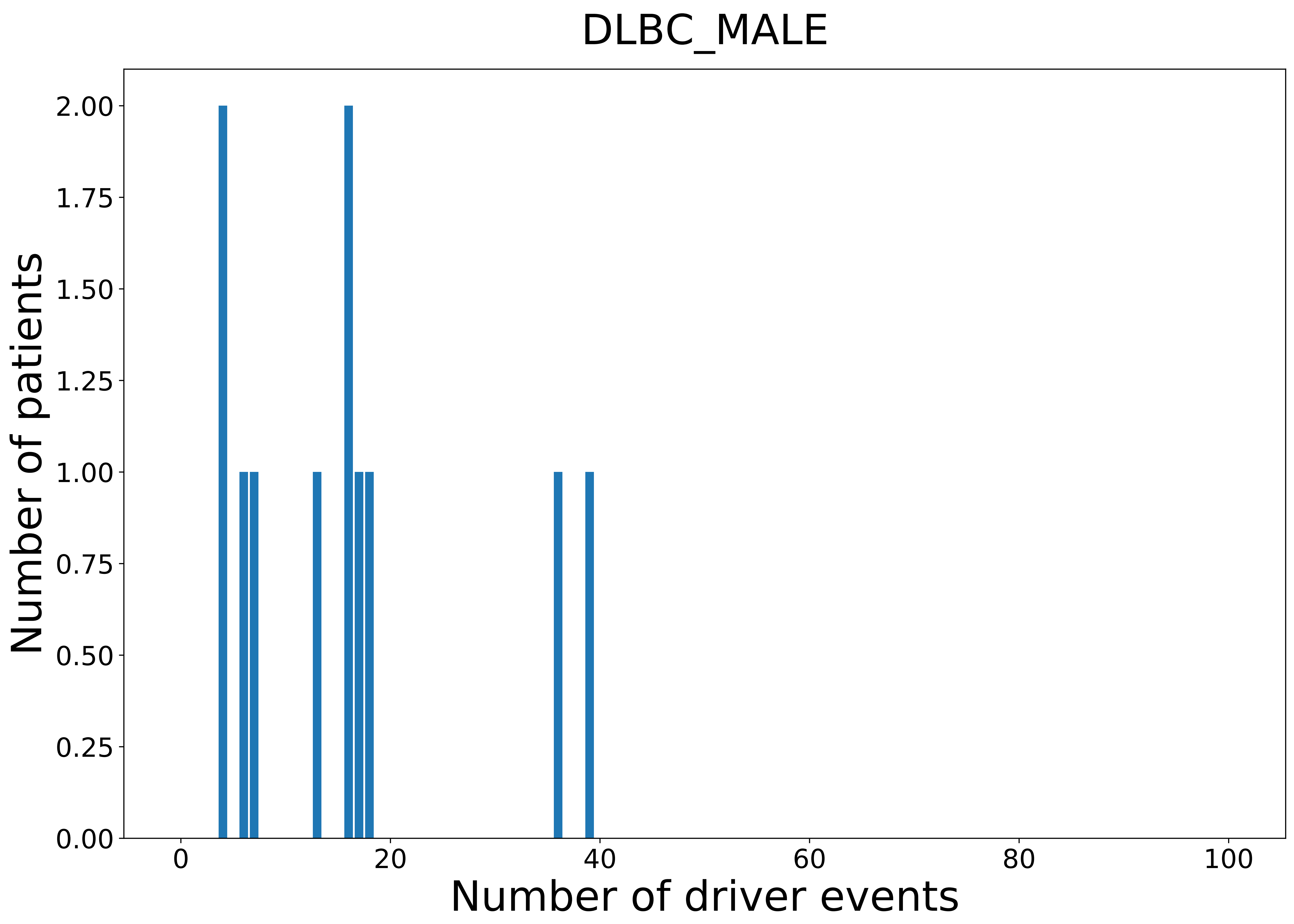

### 2021_8_16_20_32_ESCA.pdf

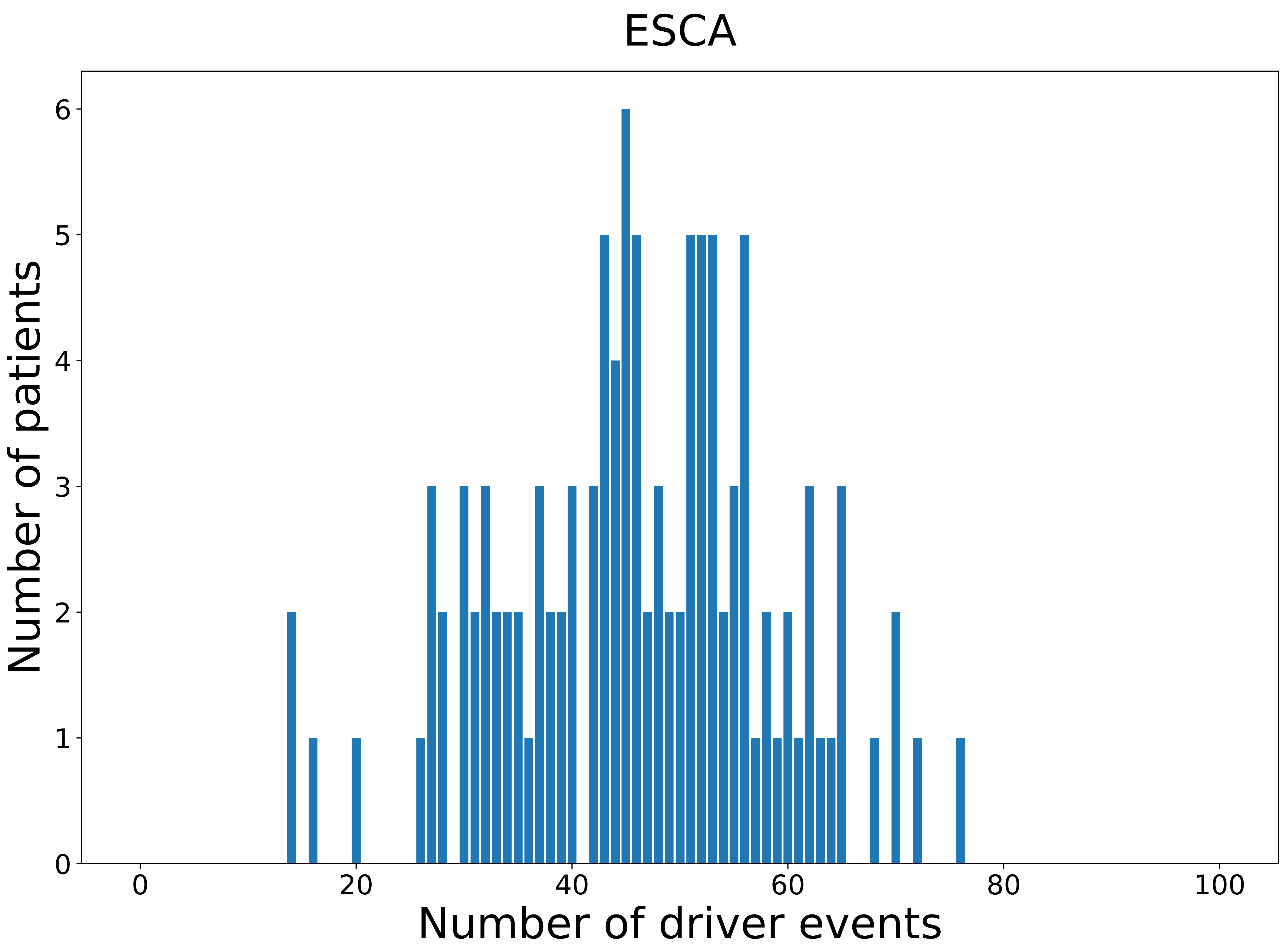

### 2021_8_16_20_32_ESCA_FEMALE.pdf

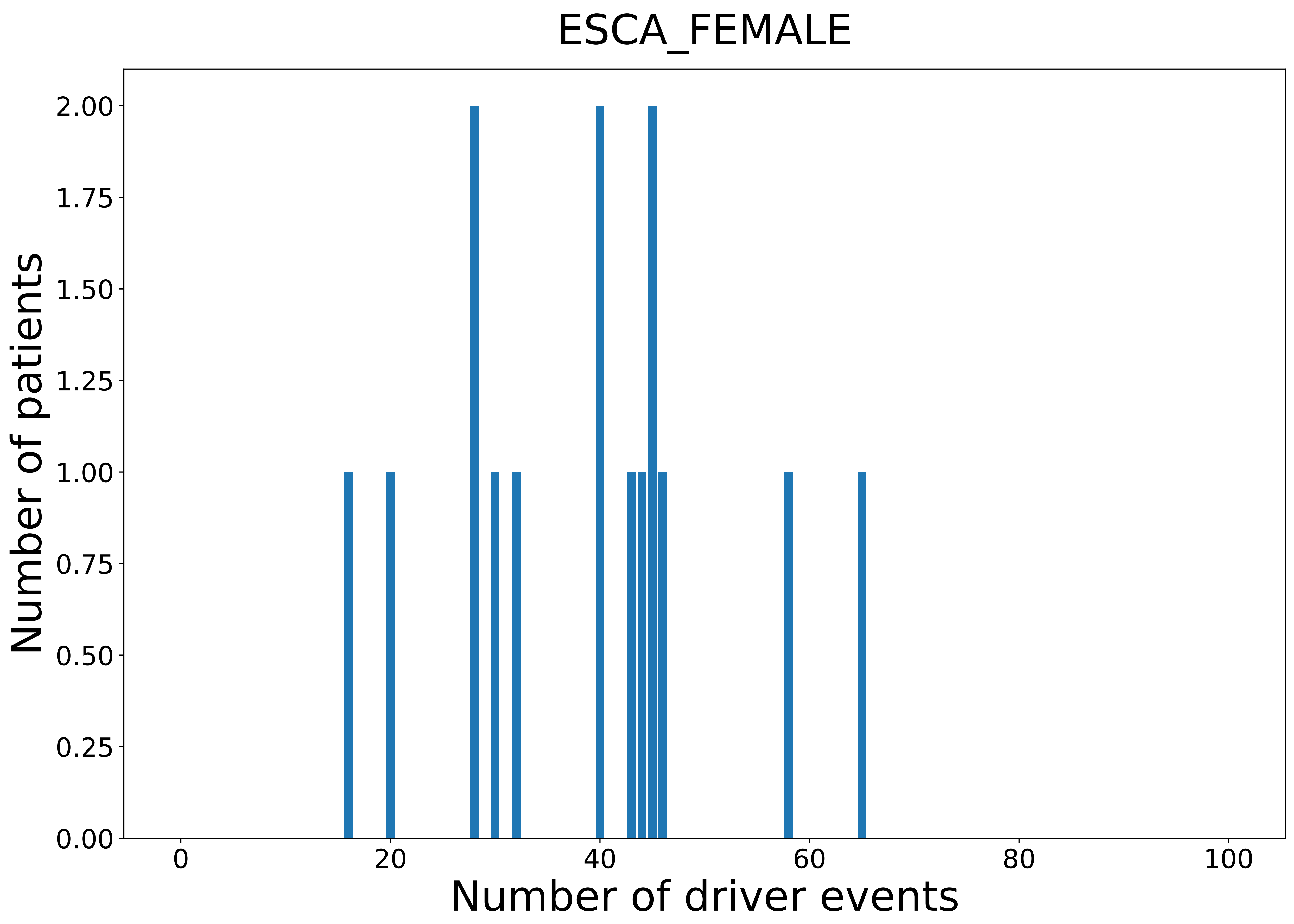

### 2021_8_16_20_32_ESCA_MALE.pdf

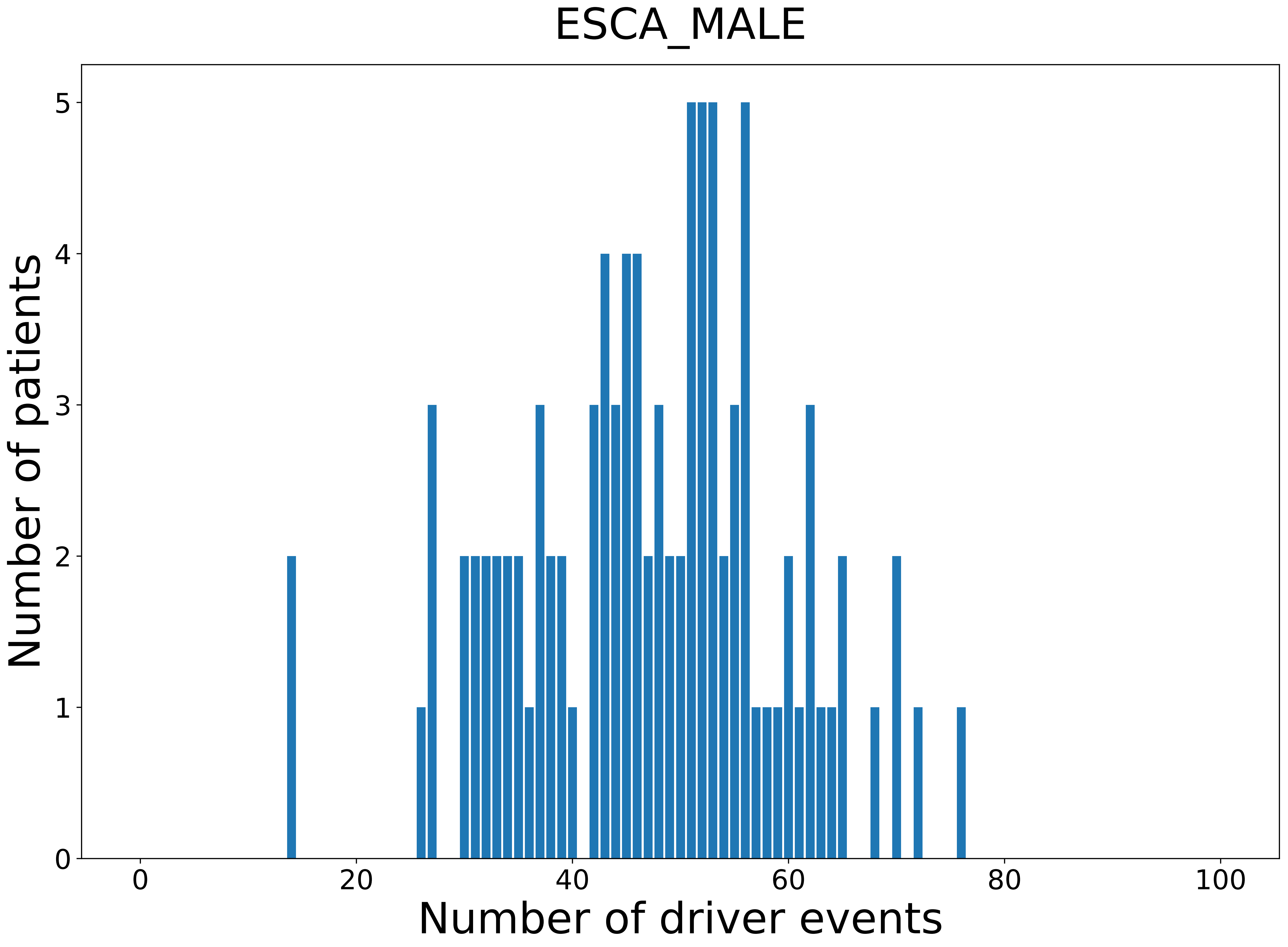

### 2021_8_16_20_32_GBM.pdf

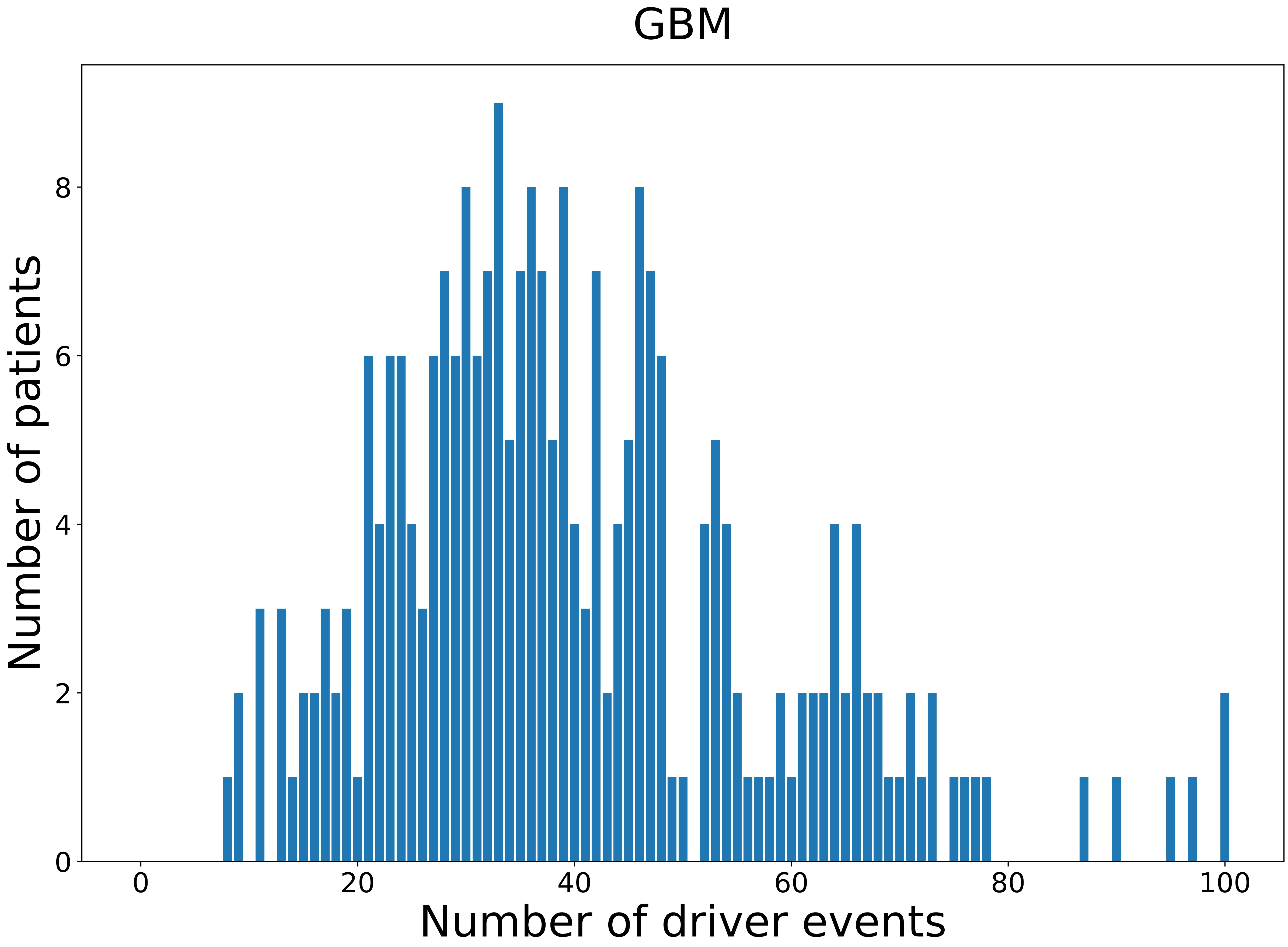

### 2021_8_16_20_32_GBM_FEMALE.pdf

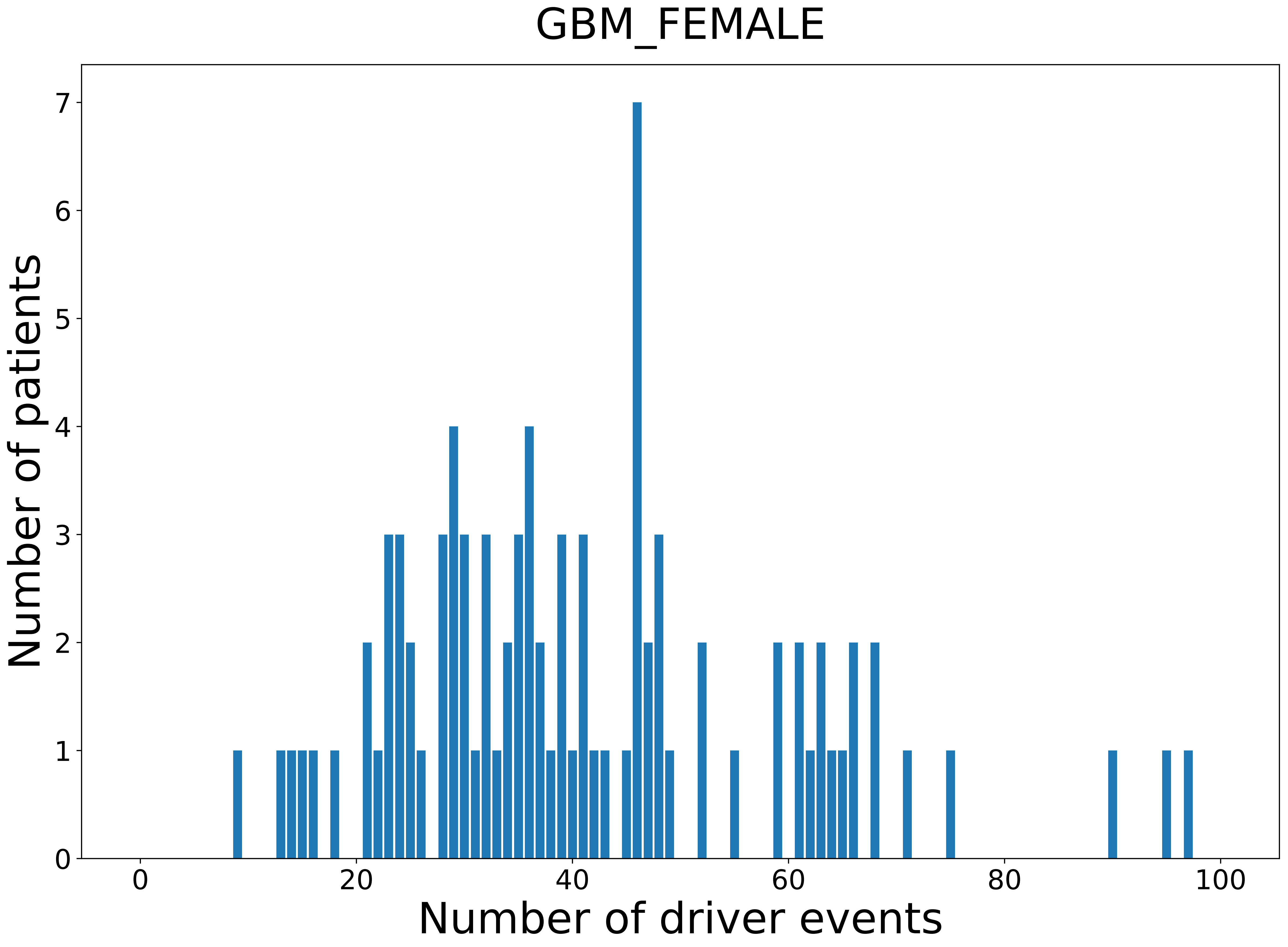

### 2021_8_16_20_32_GBM_MALE.pdf

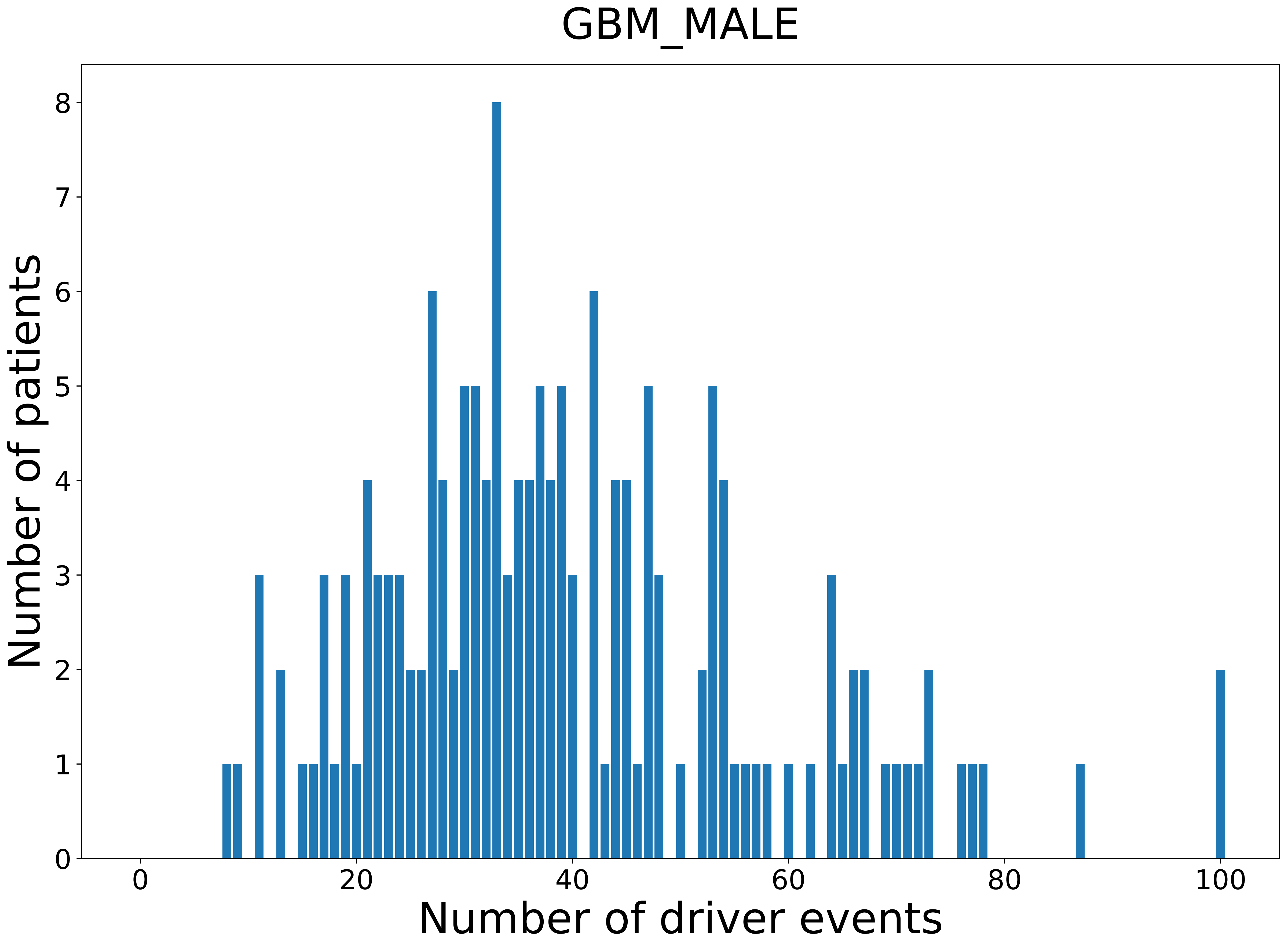

### 2021_8_16_20_32_HNSC.pdf

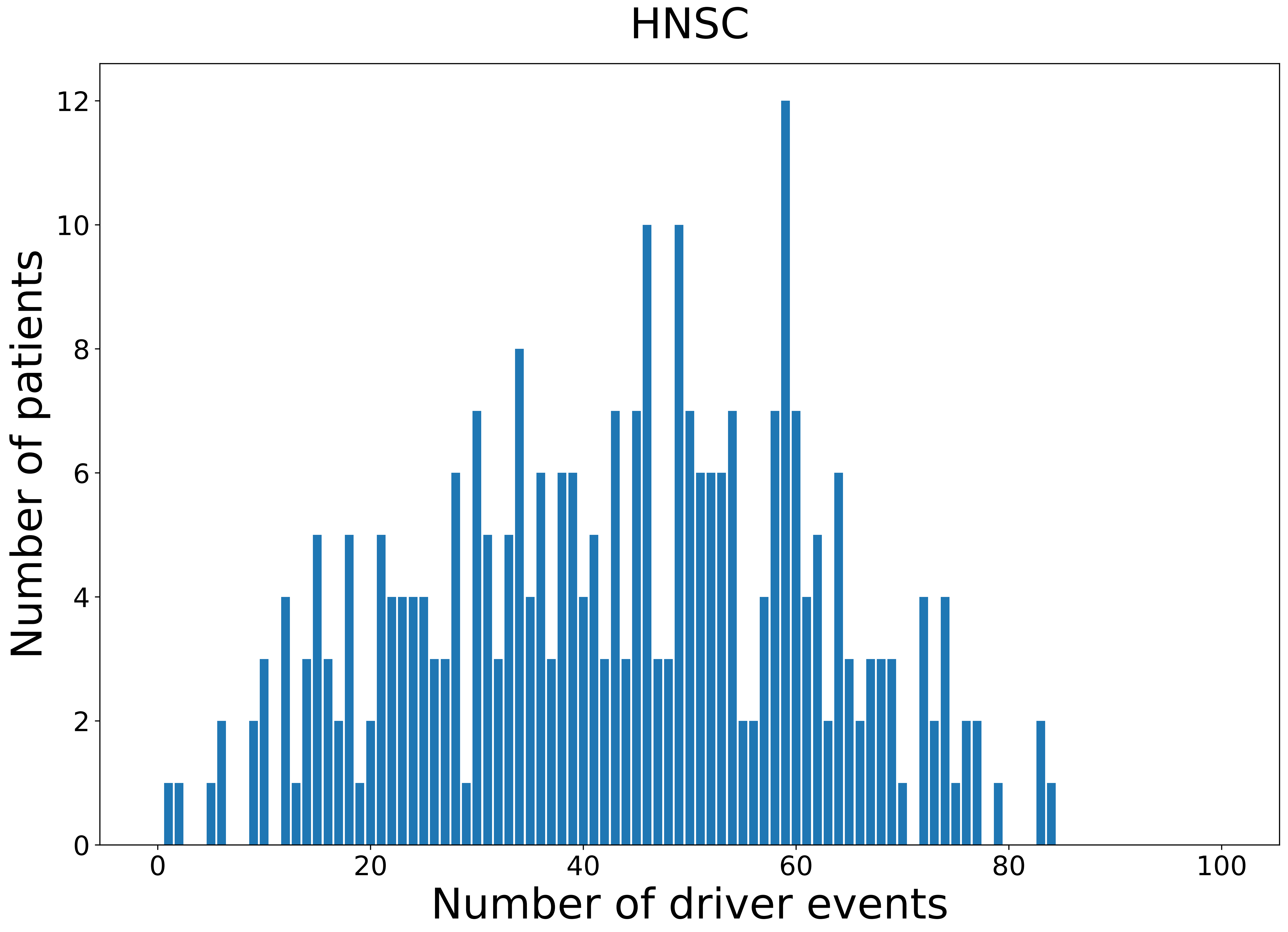

### 2021_8_16_20_32_HNSC_FEMALE.pdf

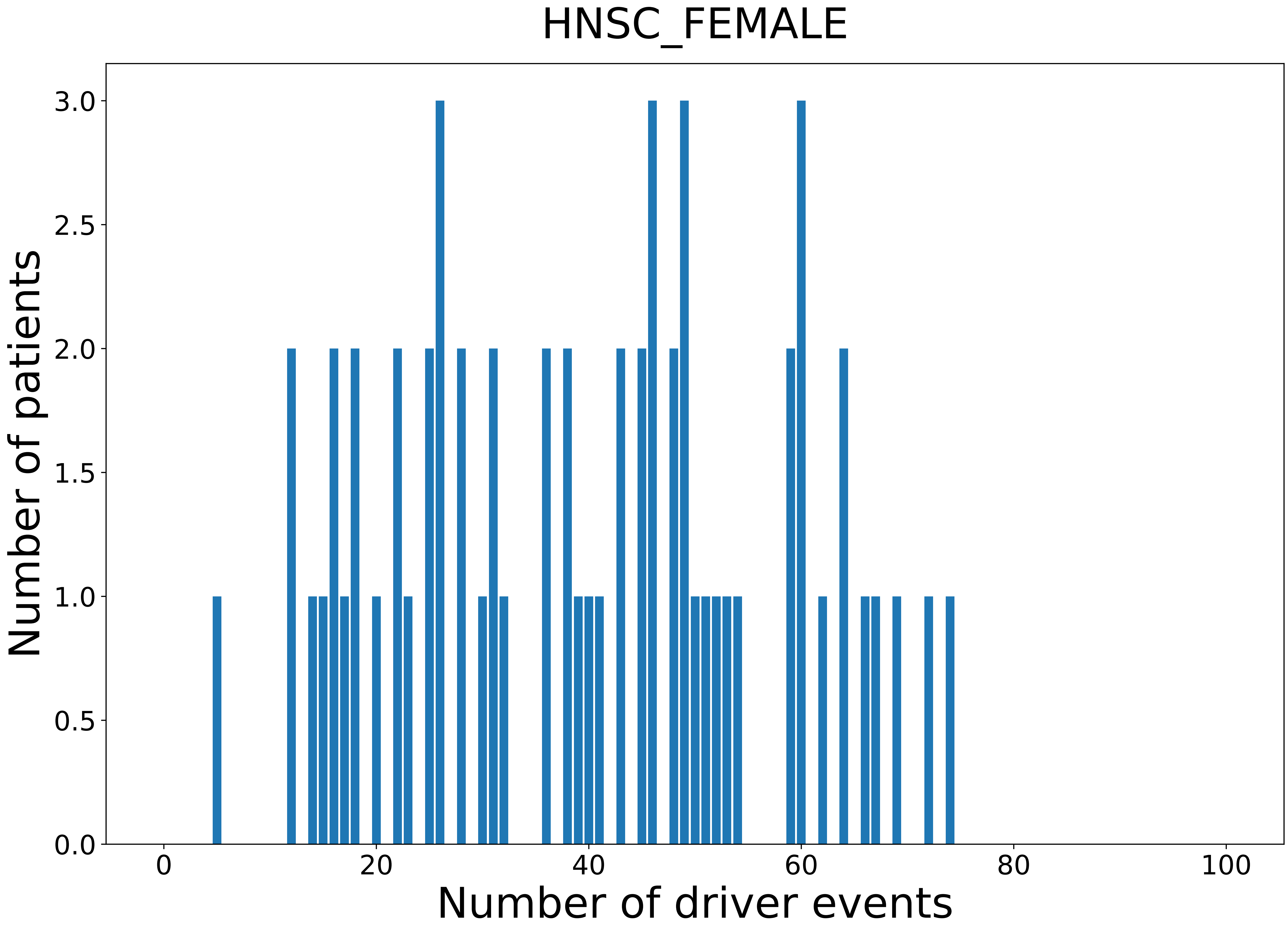

### 2021_8_16_20_32_HNSC_MALE.pdf

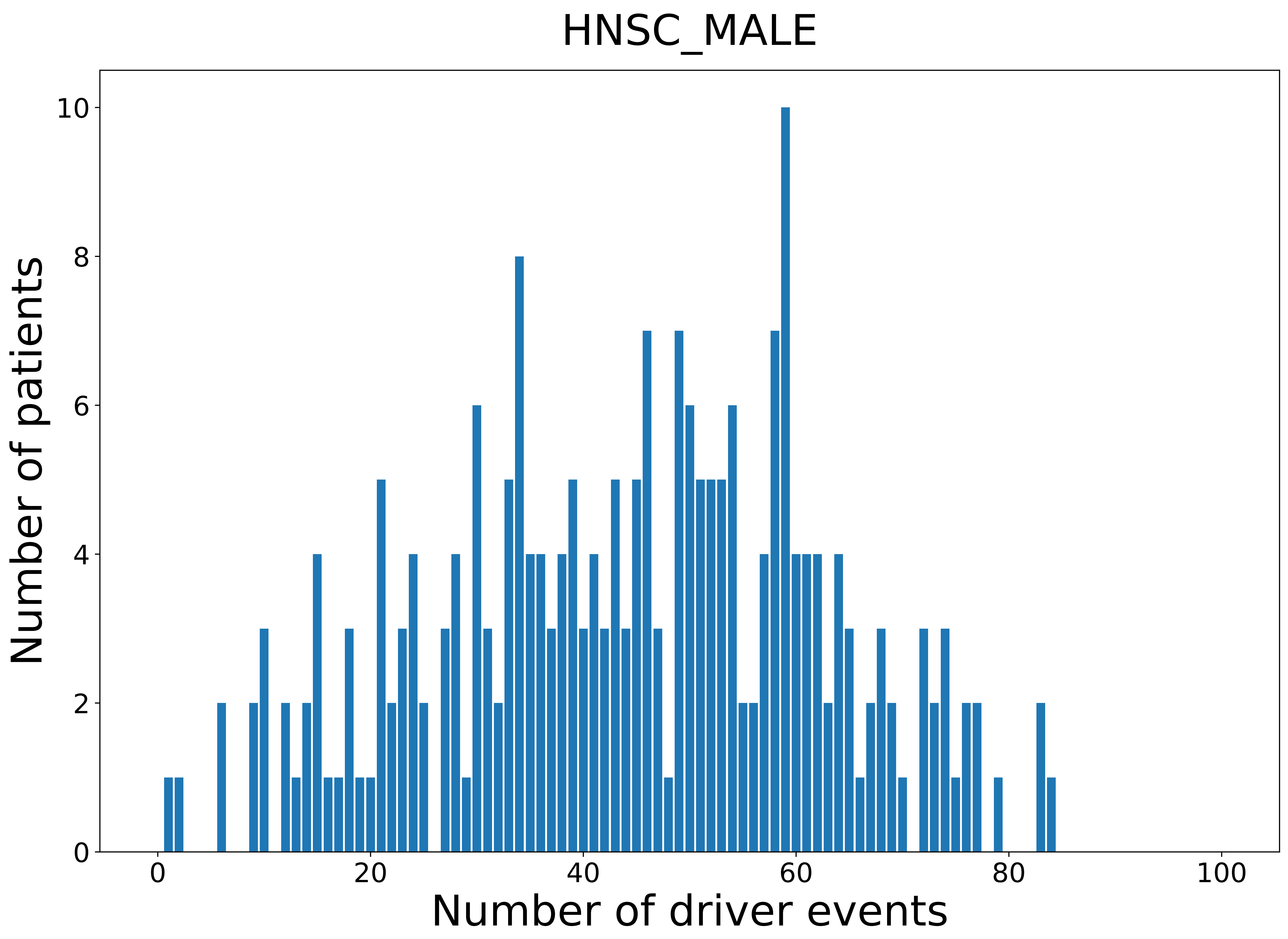

### 2021_8_16_20_32_KICH.pdf

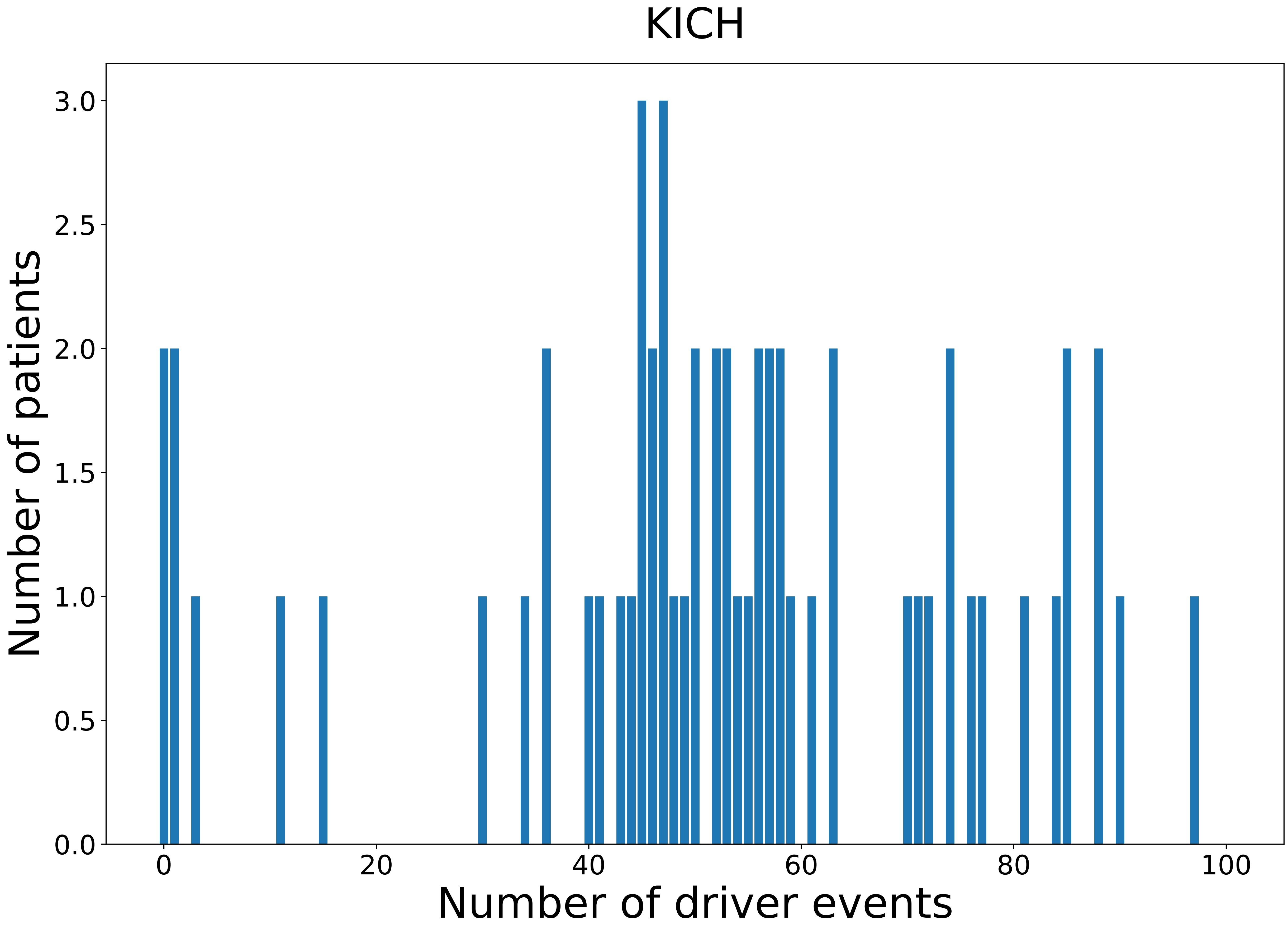

### 2021_8_16_20_32_KICH_FEMALE.pdf

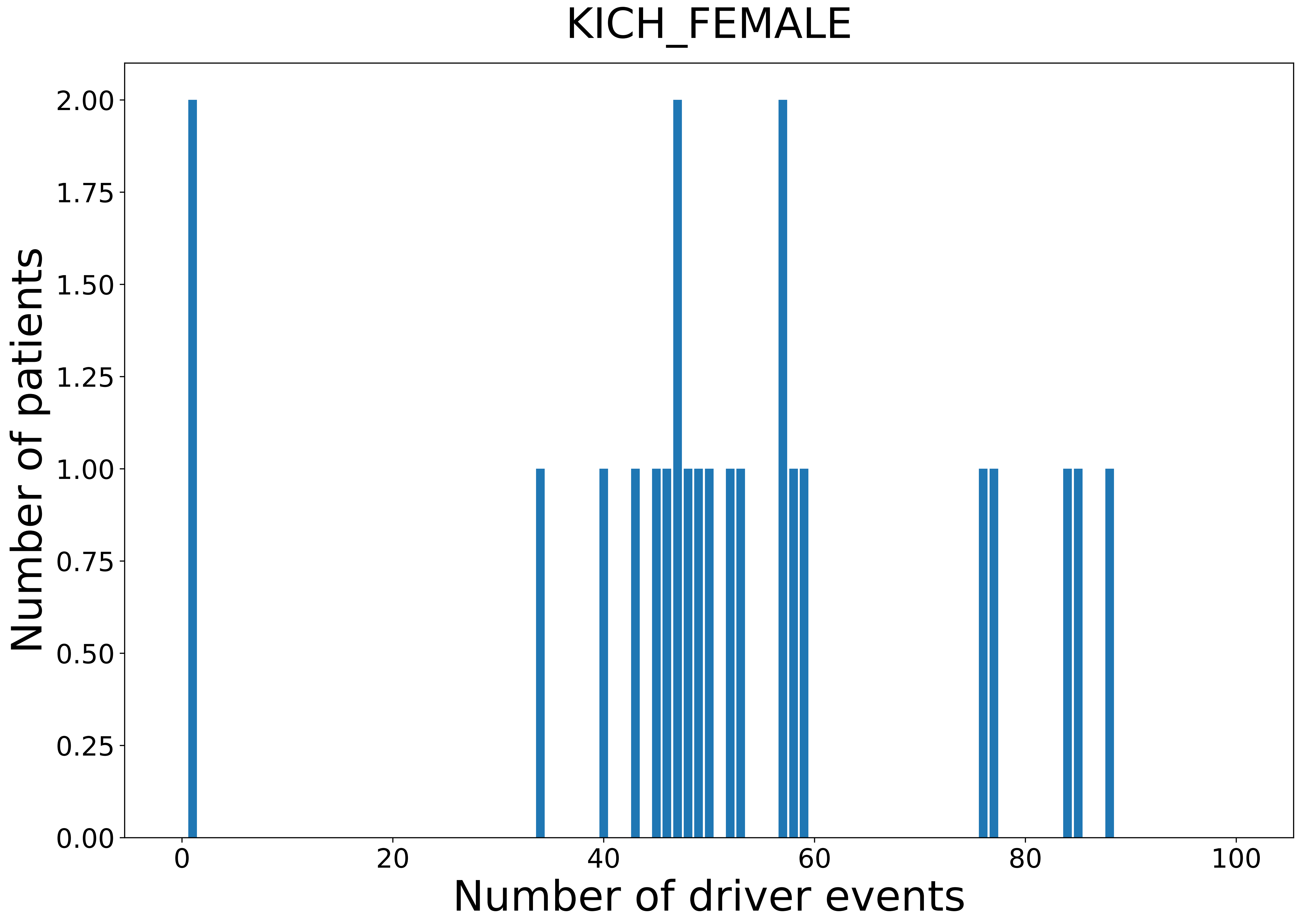

### 2021_8_16_20_32_KICH_MALE.pdf

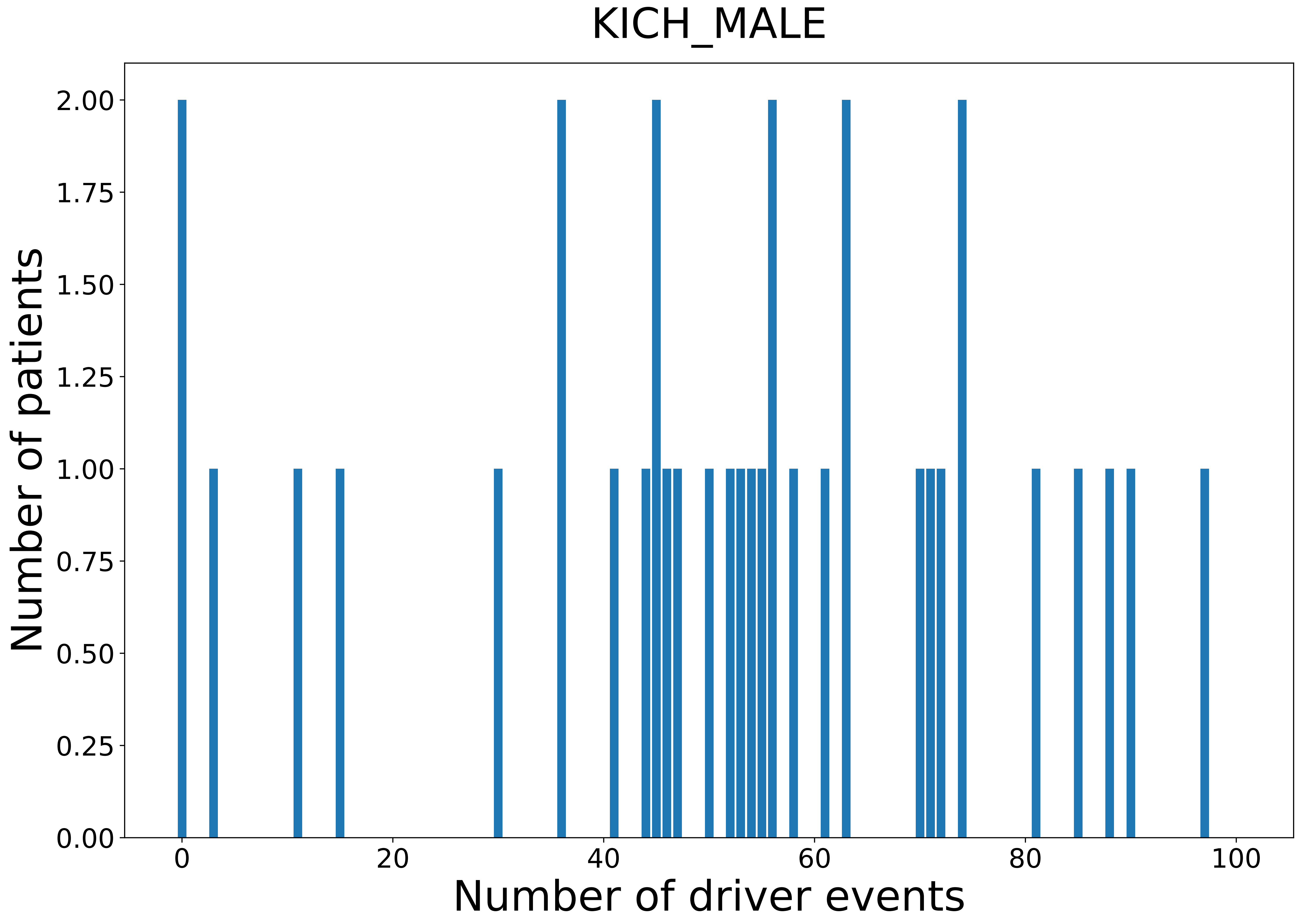

### 2021_8_16_20_32_KIRC.pdf

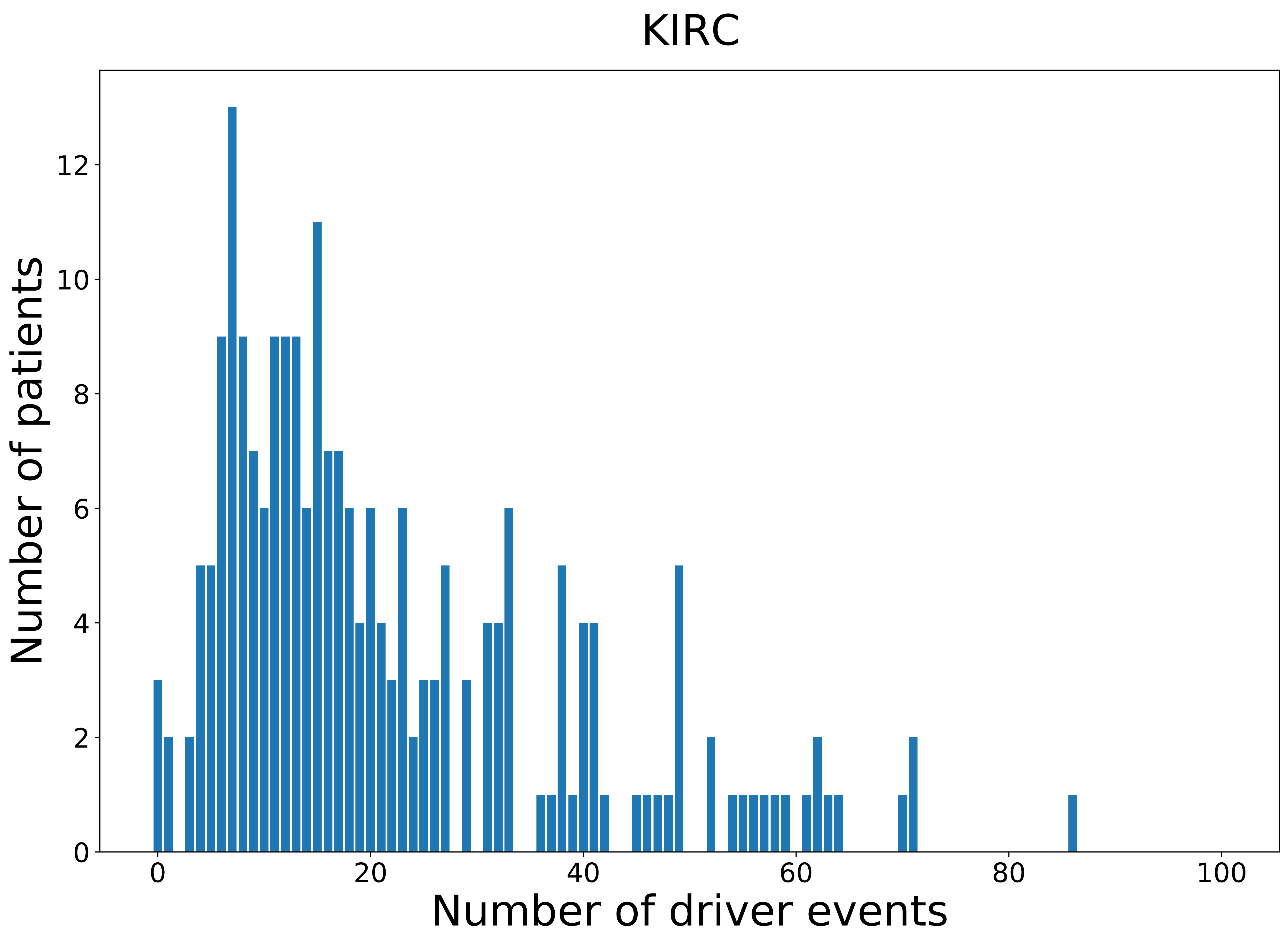

### 2021_8_16_20_32_KIRC_FEMALE.pdf

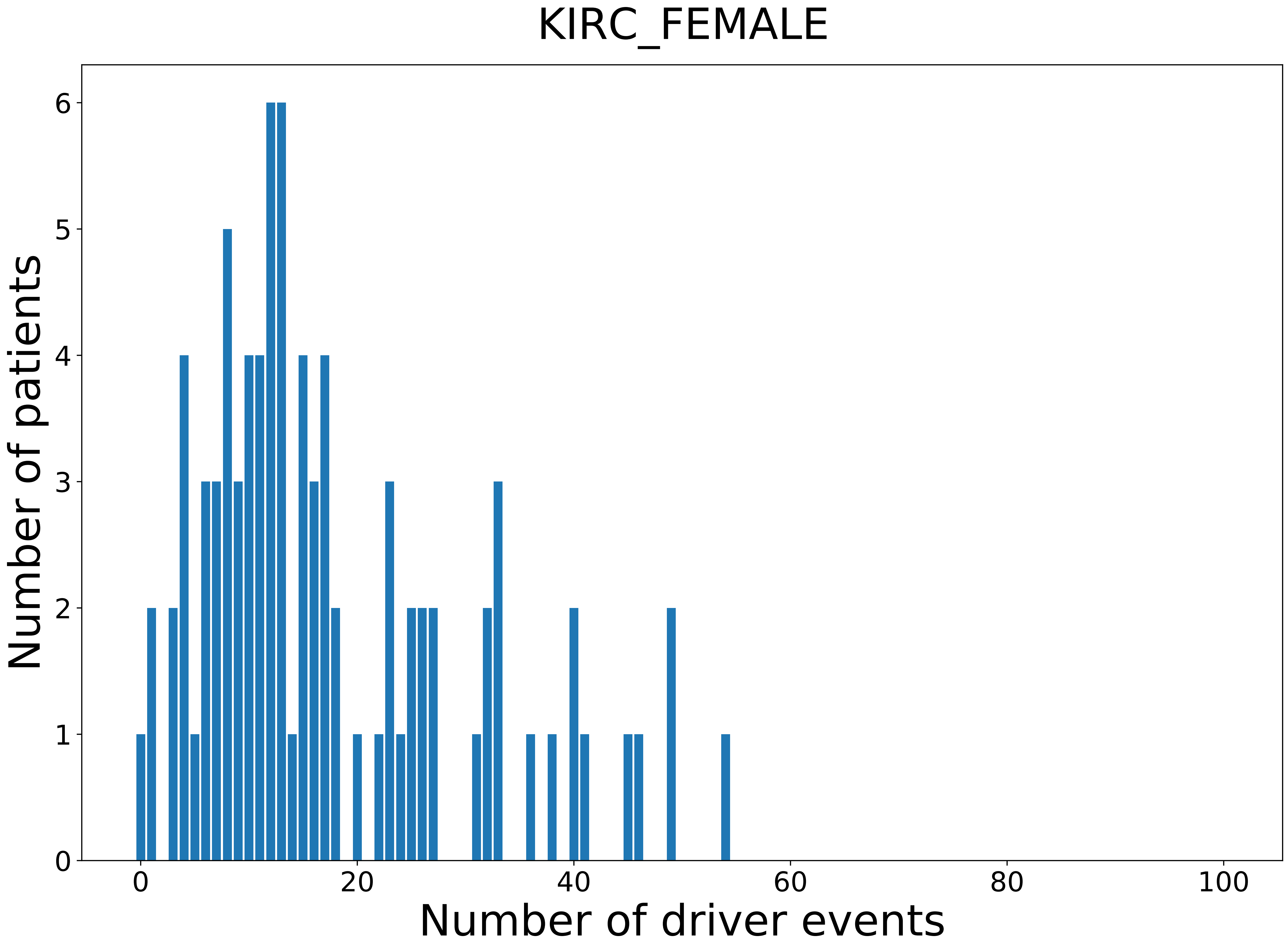

### 2021_8_16_20_32_KIRC_MALE.pdf

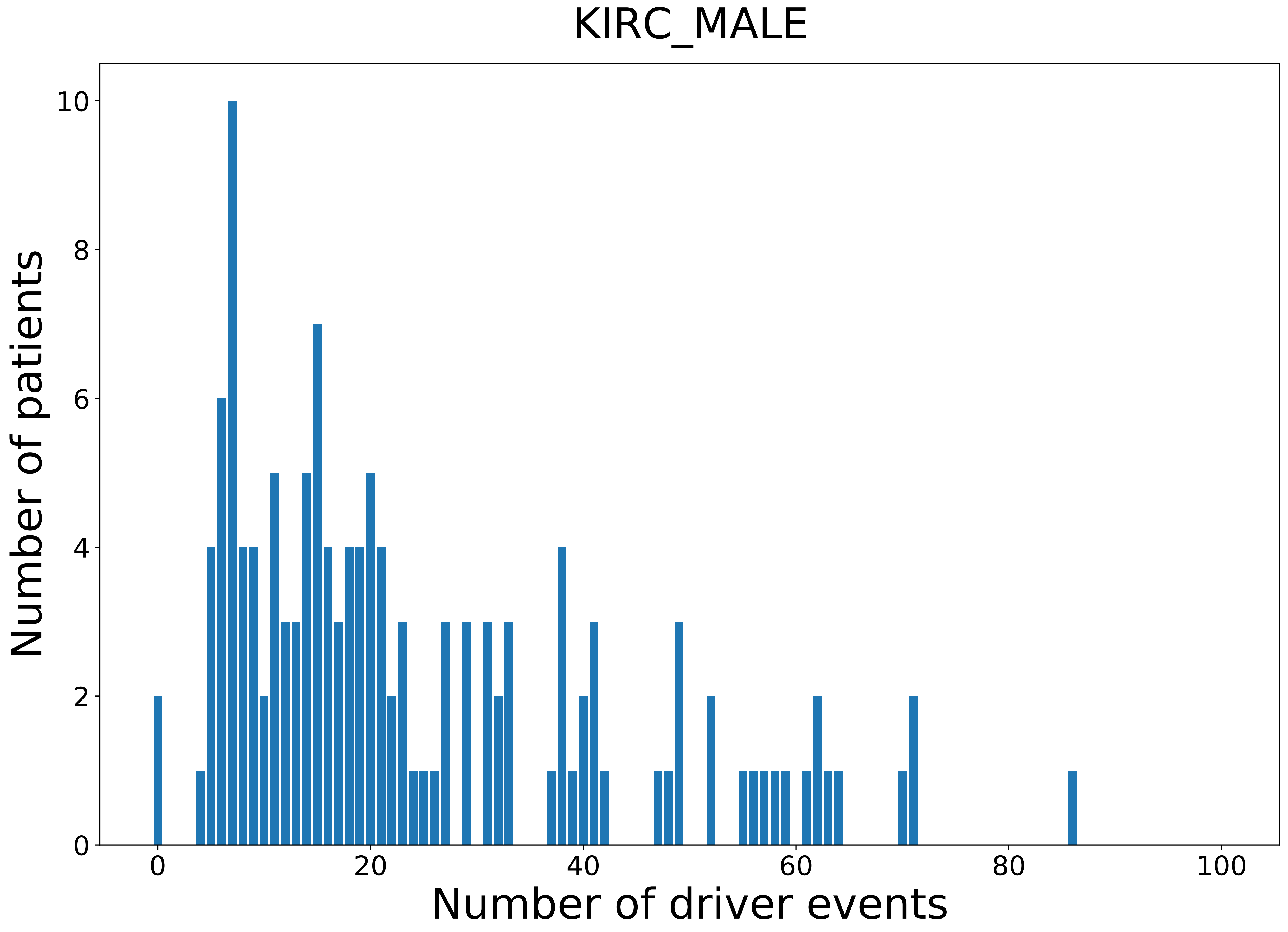

### 2021_8_16_20_32_KIRP.pdf

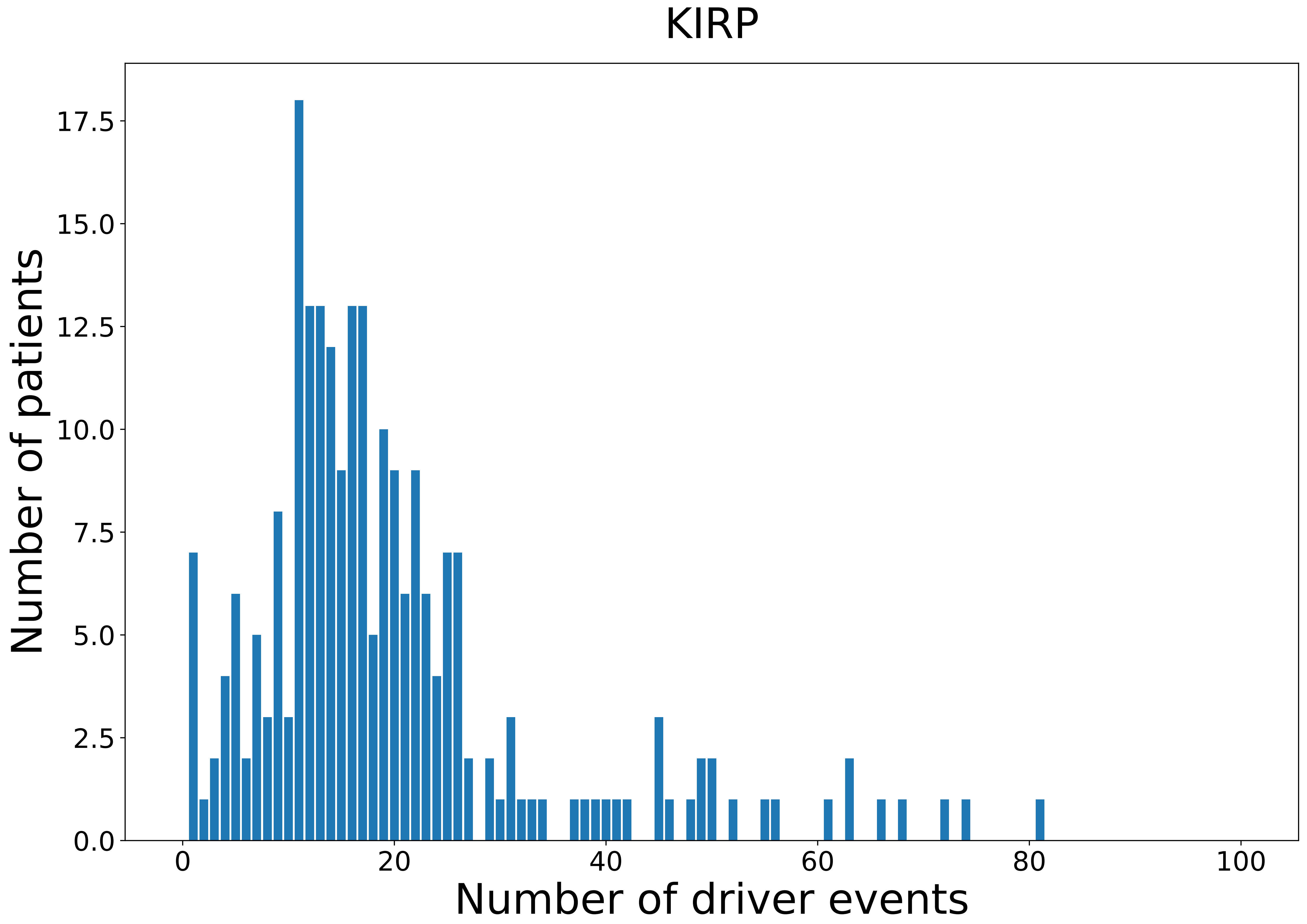

### 2021_8_16_20_32_KIRP_FEMALE.pdf

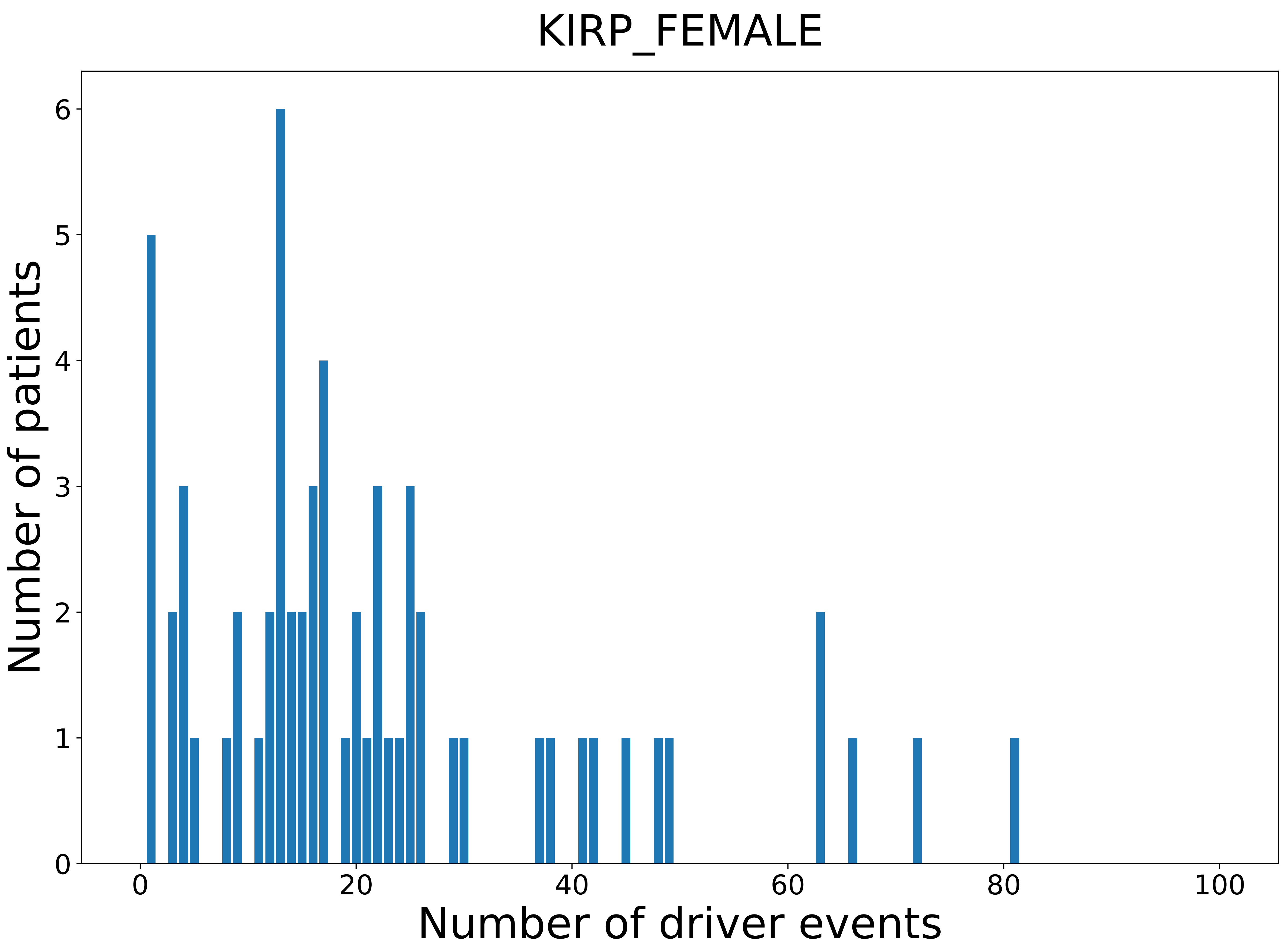

### 2021_8_16_20_32_KIRP_MALE.pdf

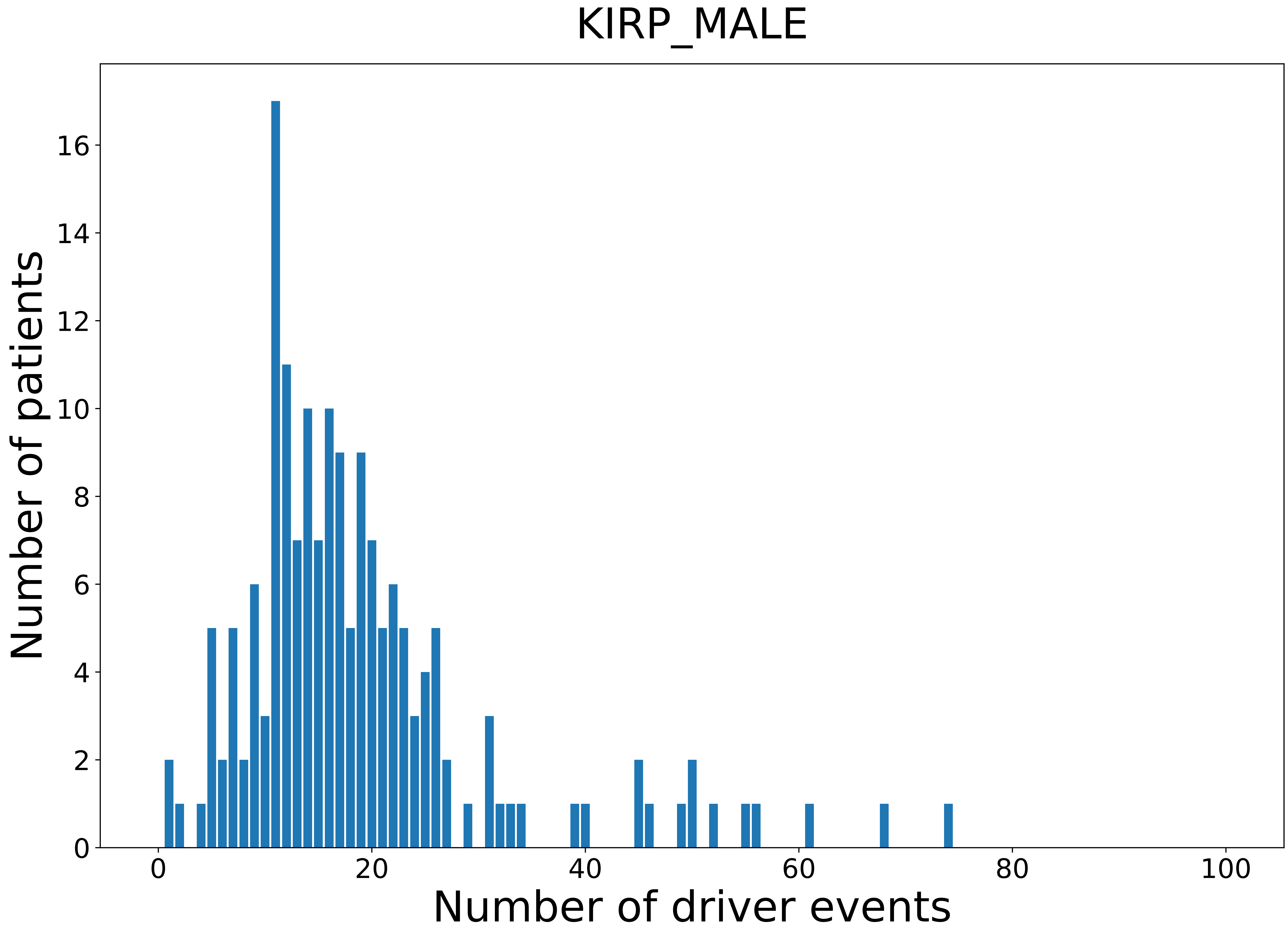

### 2021_8_16_20_32_LGG.pdf

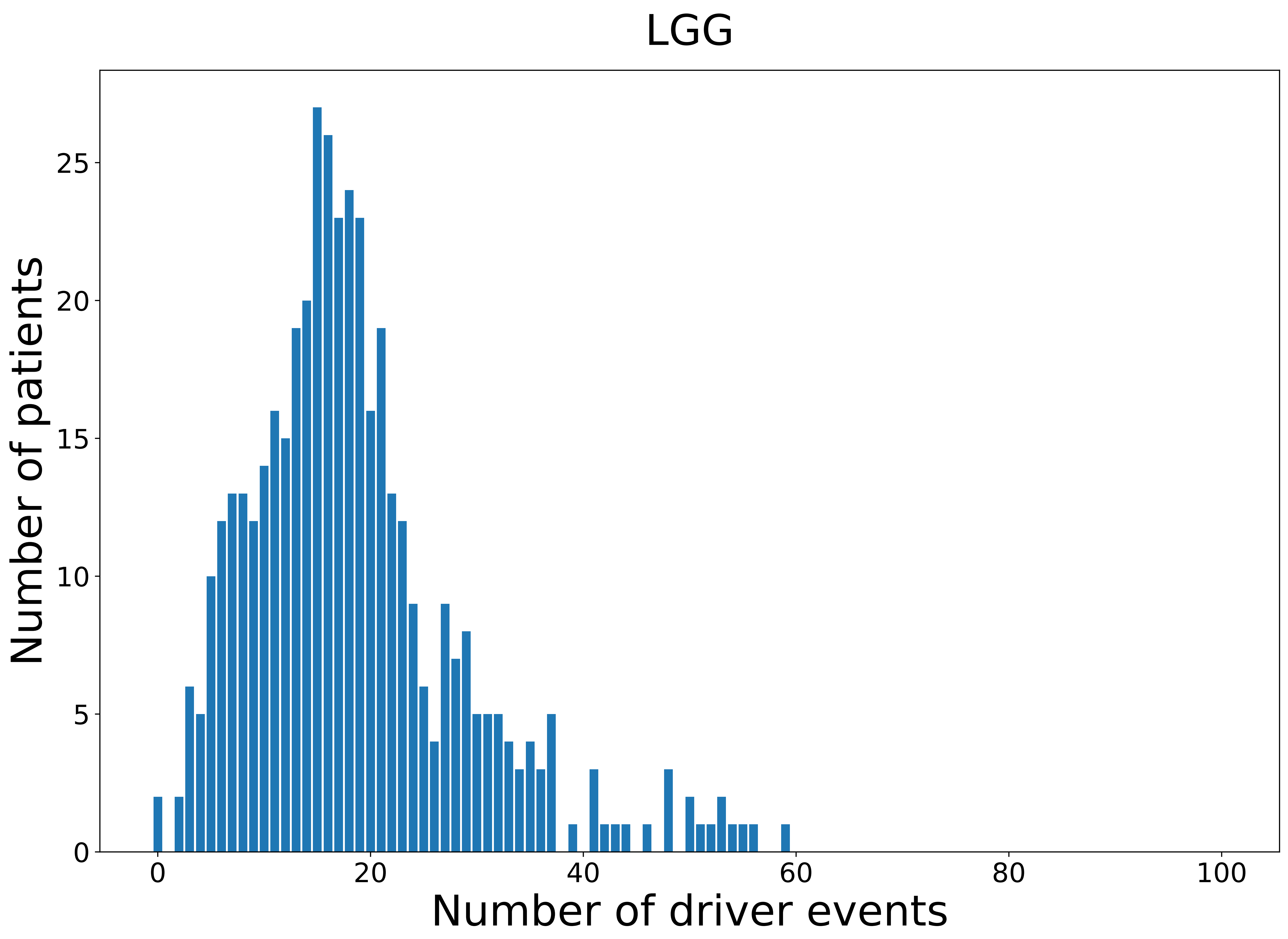

### 2021_8_16_20_32_LGG_FEMALE.pdf

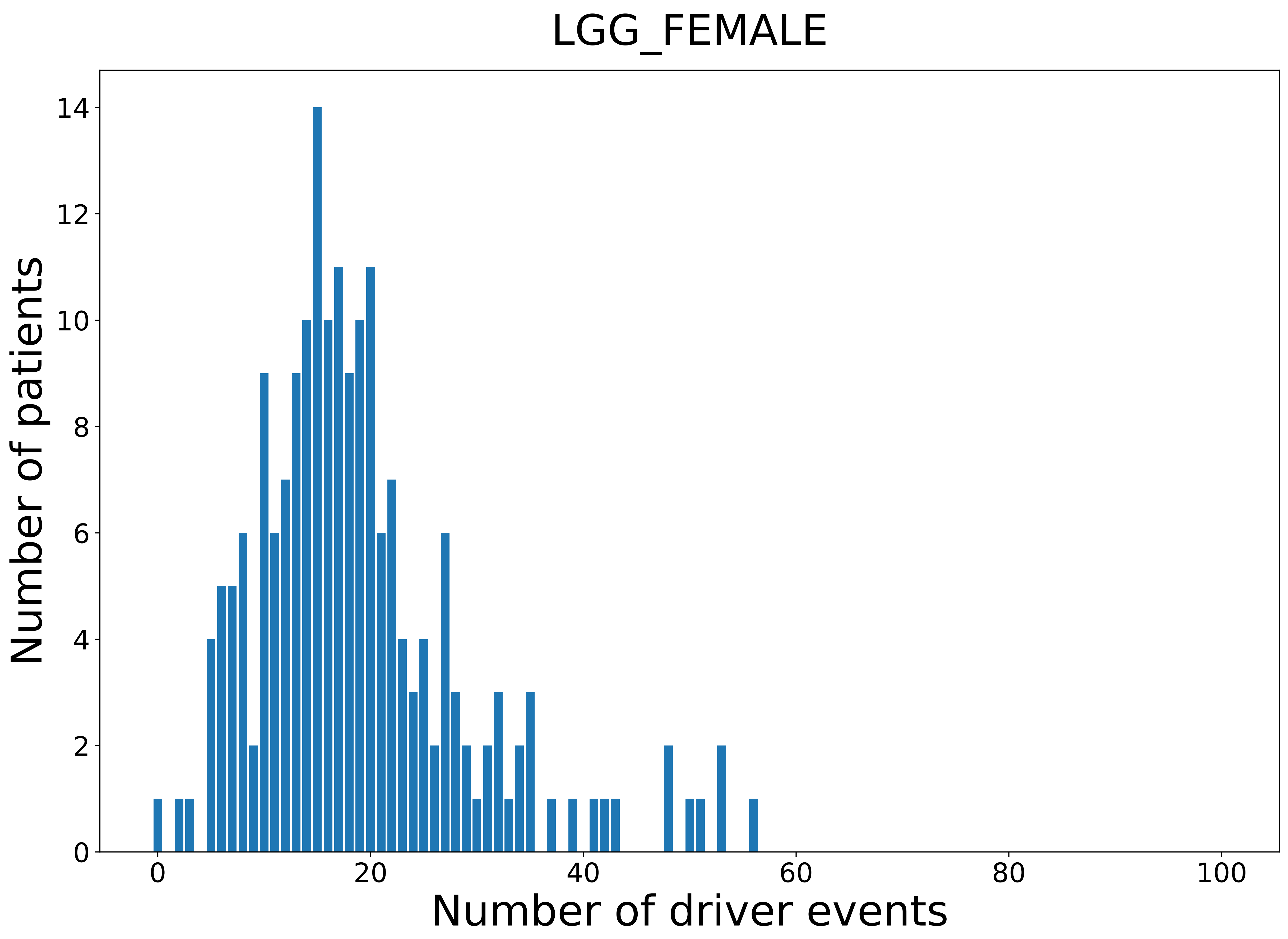

### 2021_8_16_20_32_LGG_MALE.pdf

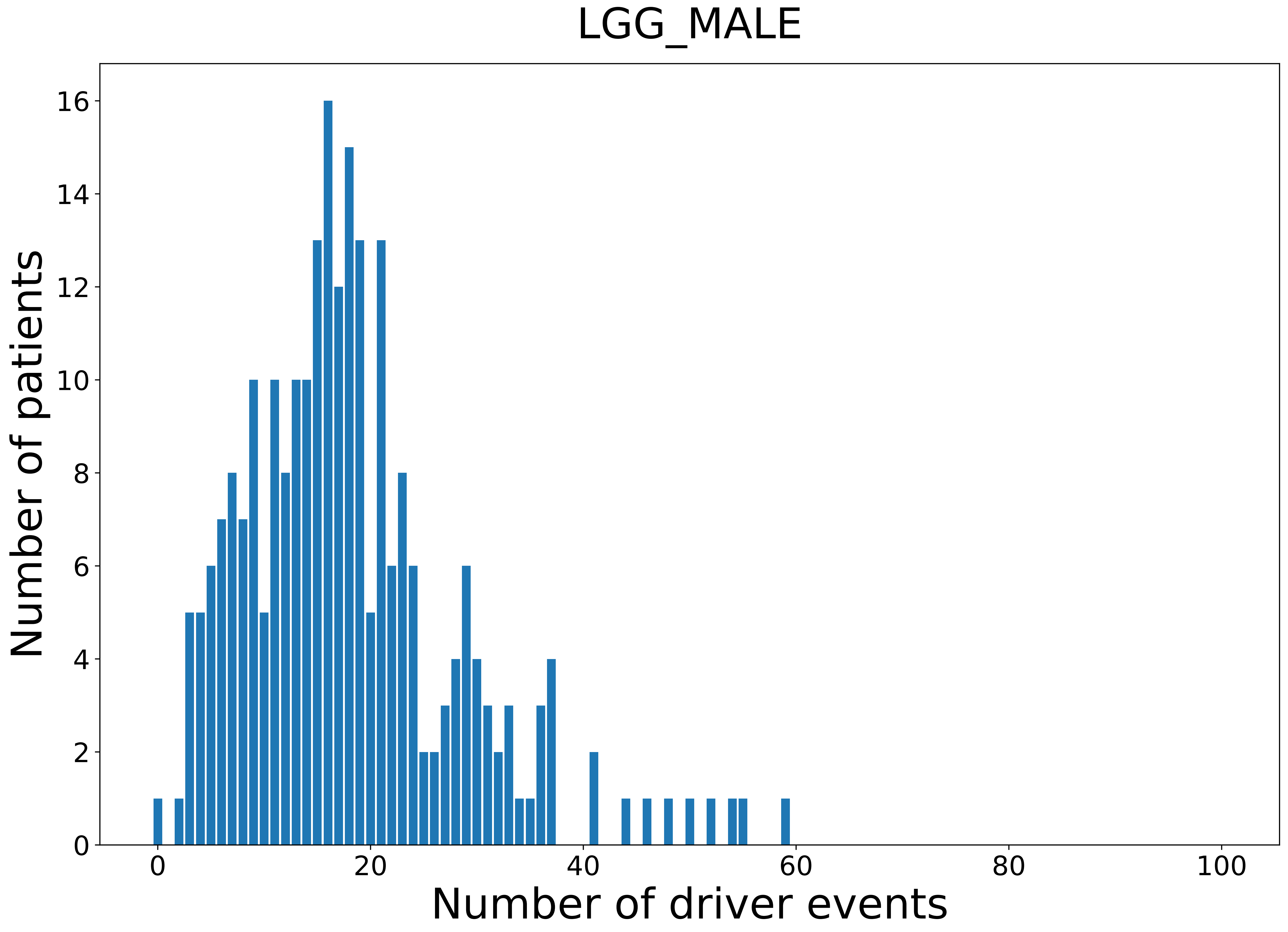

### 2021_8_16_20_32_LIHC.pdf

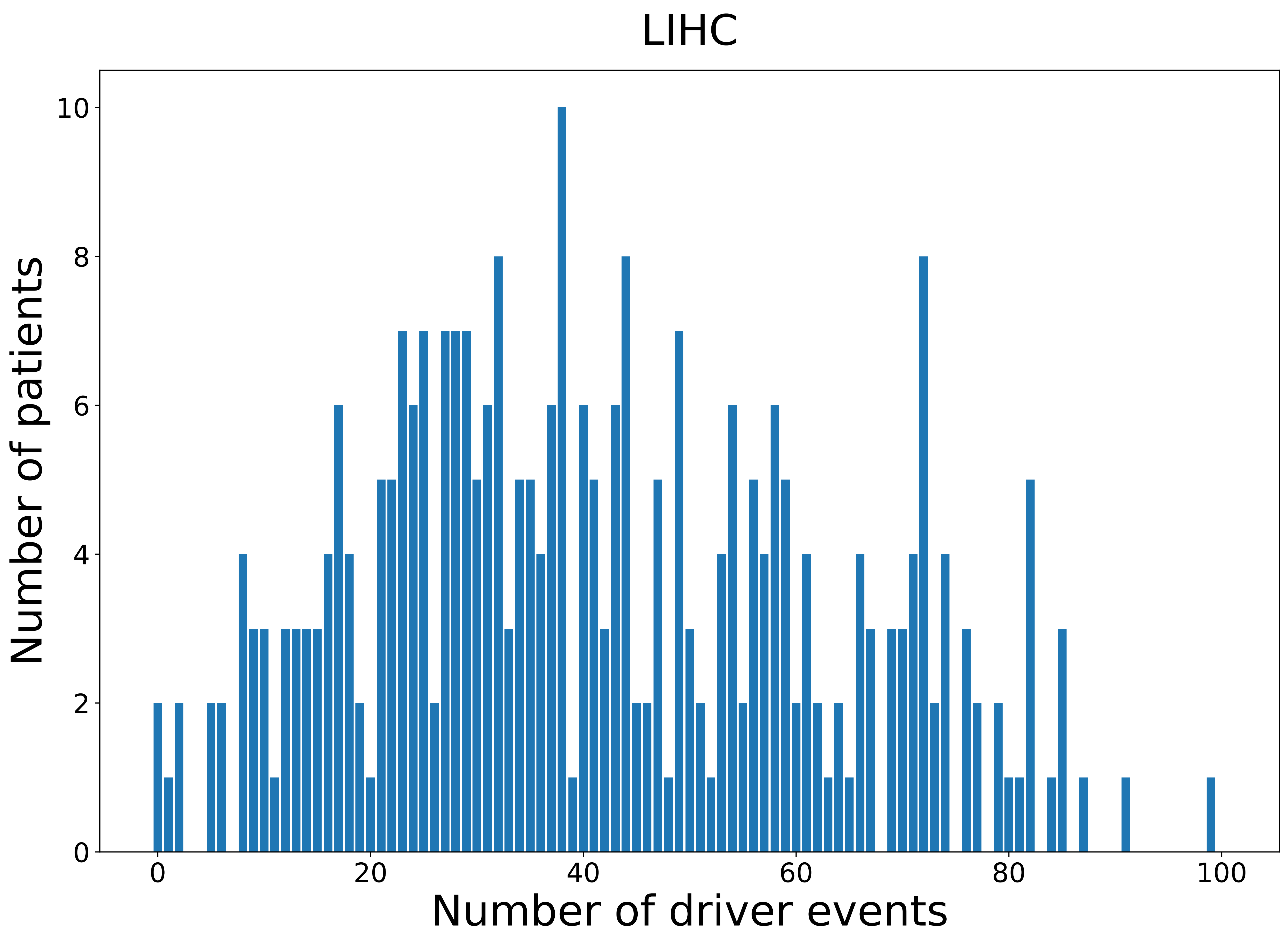

### 2021_8_16_20_32_LIHC_FEMALE.pdf

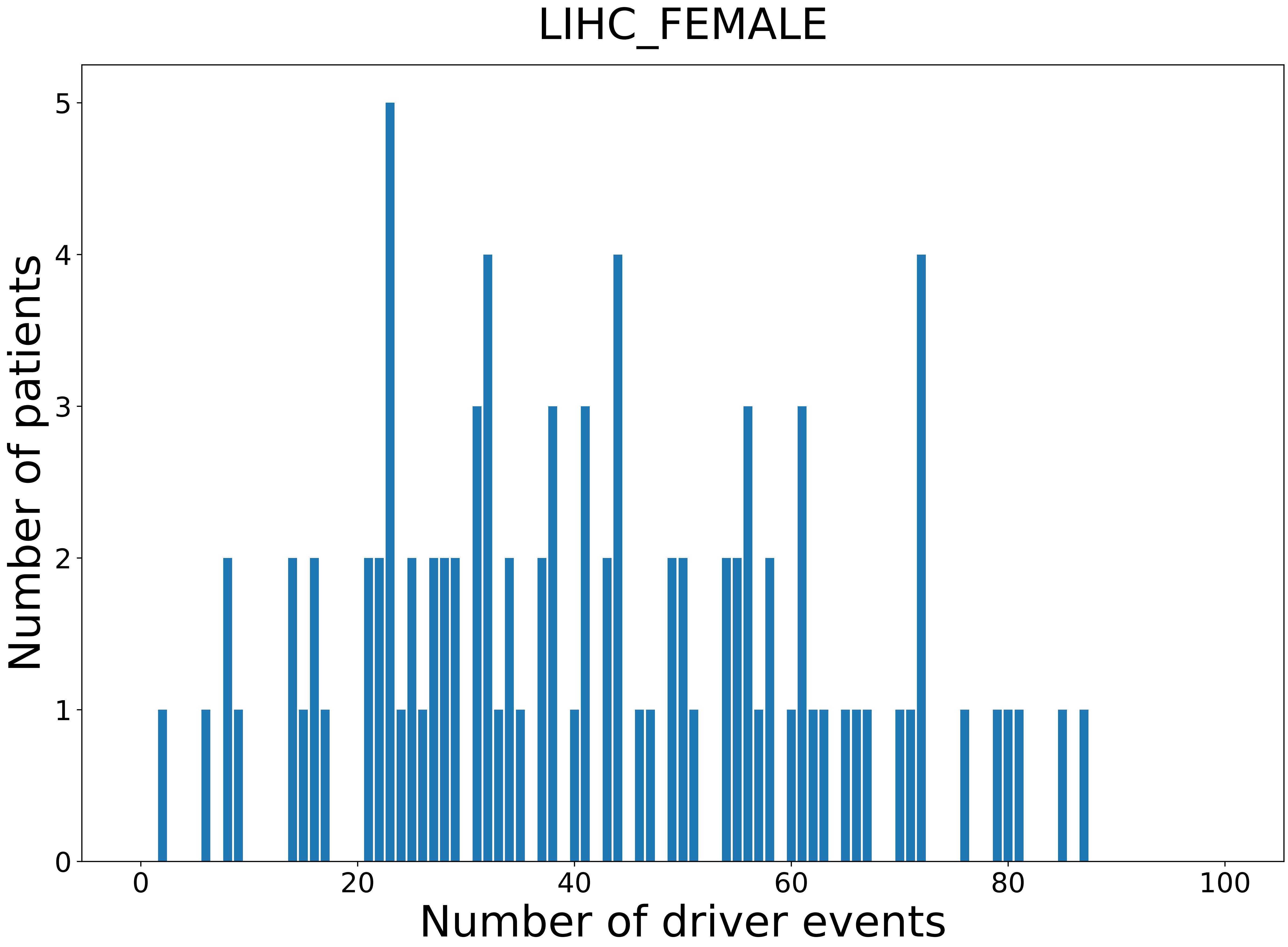

### 2021_8_16_20_32_LIHC_MALE.pdf

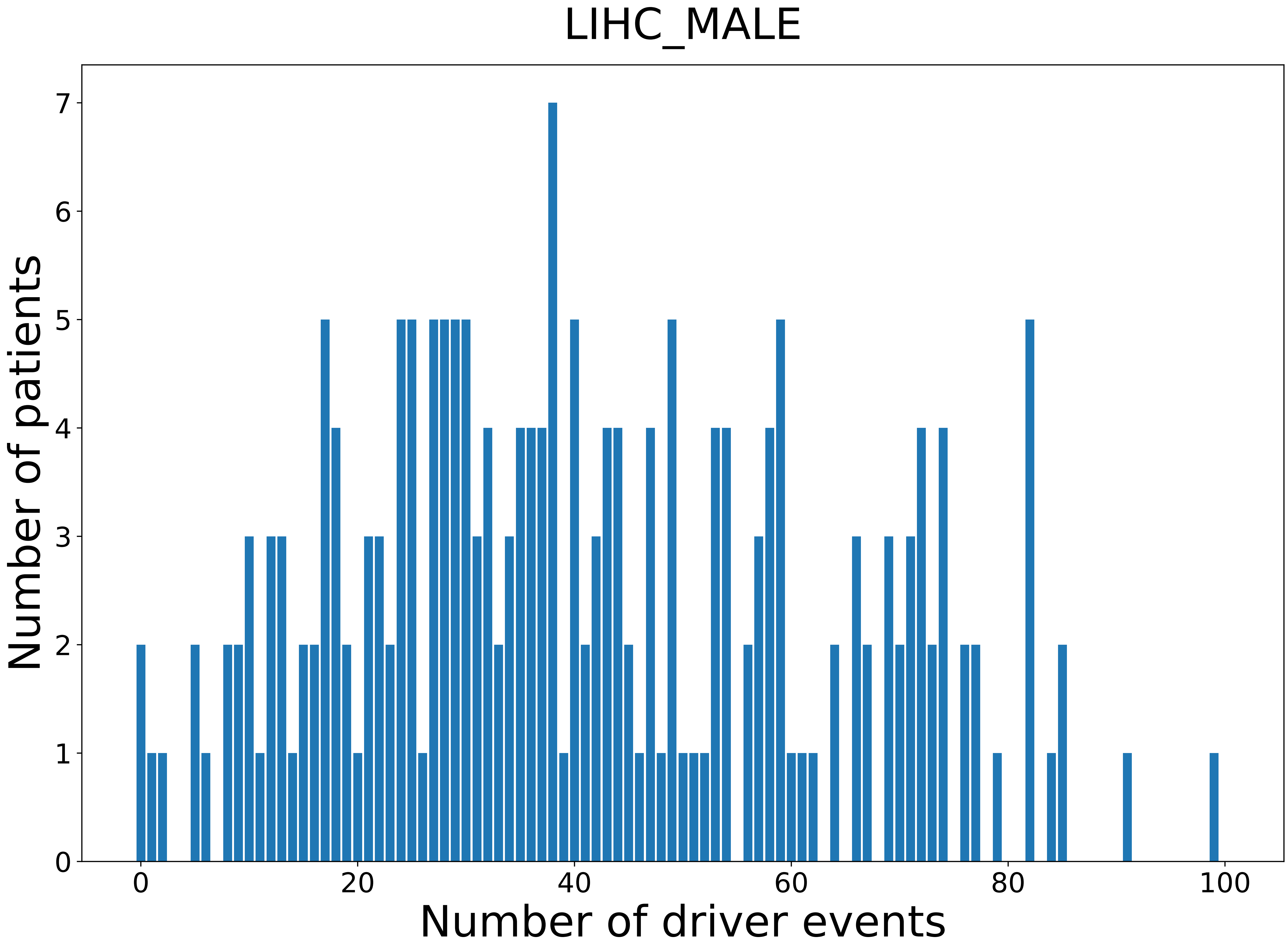

### 2021_8_16_20_32_LUAD.pdf

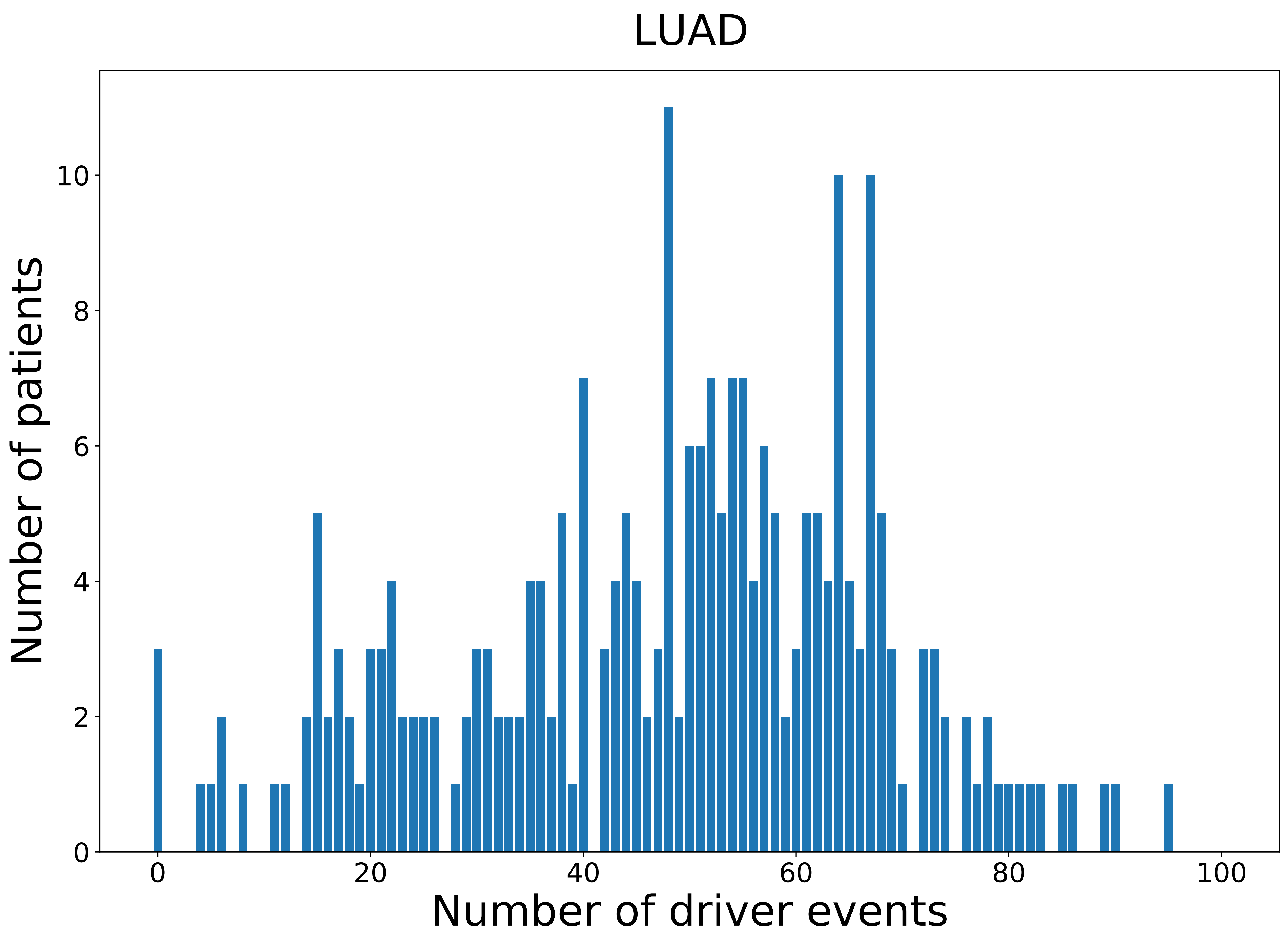

### 2021_8_16_20_32_LUAD_FEMALE.pdf

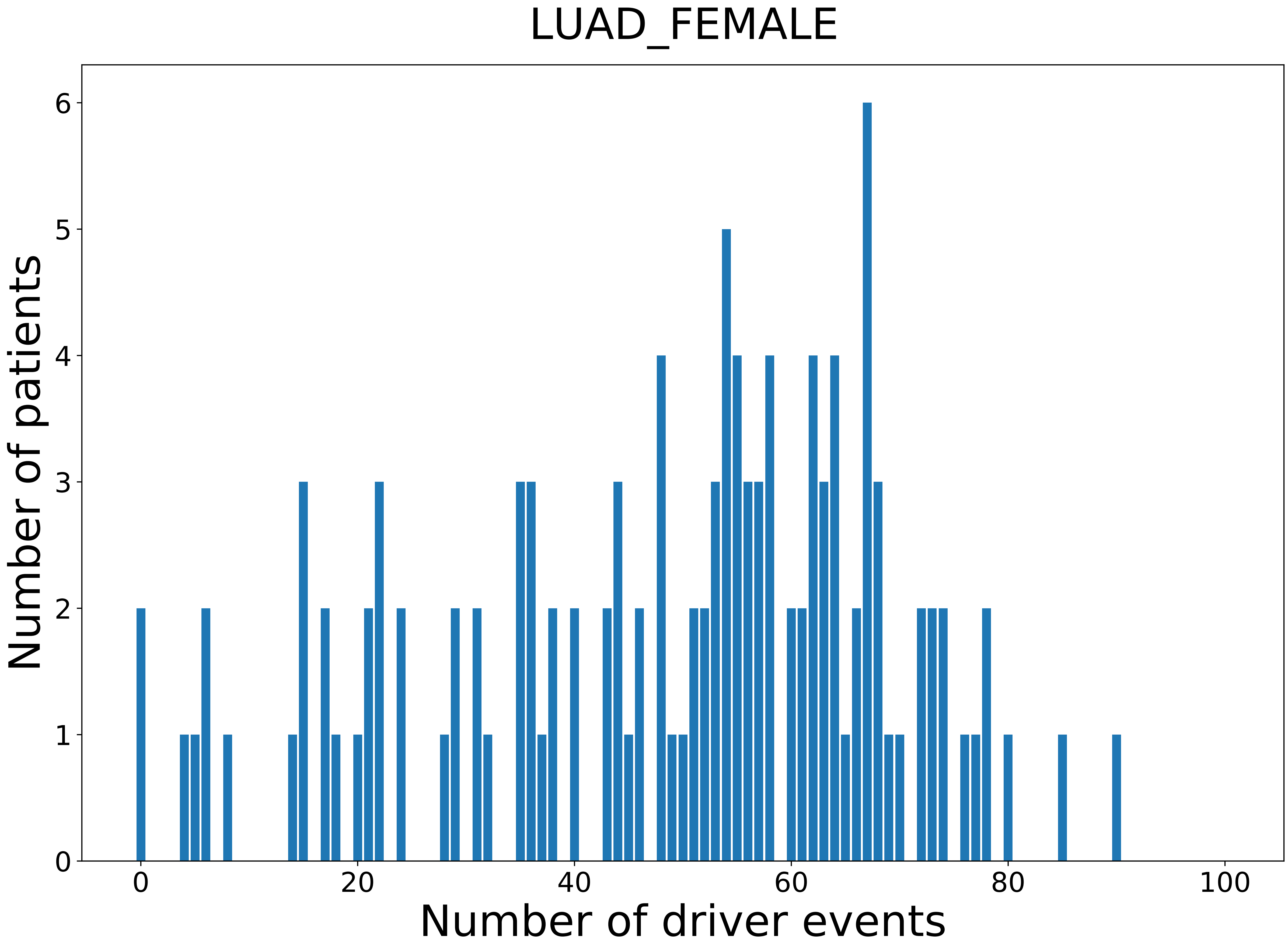

### 2021_8_16_20_32_LUAD_MALE.pdf

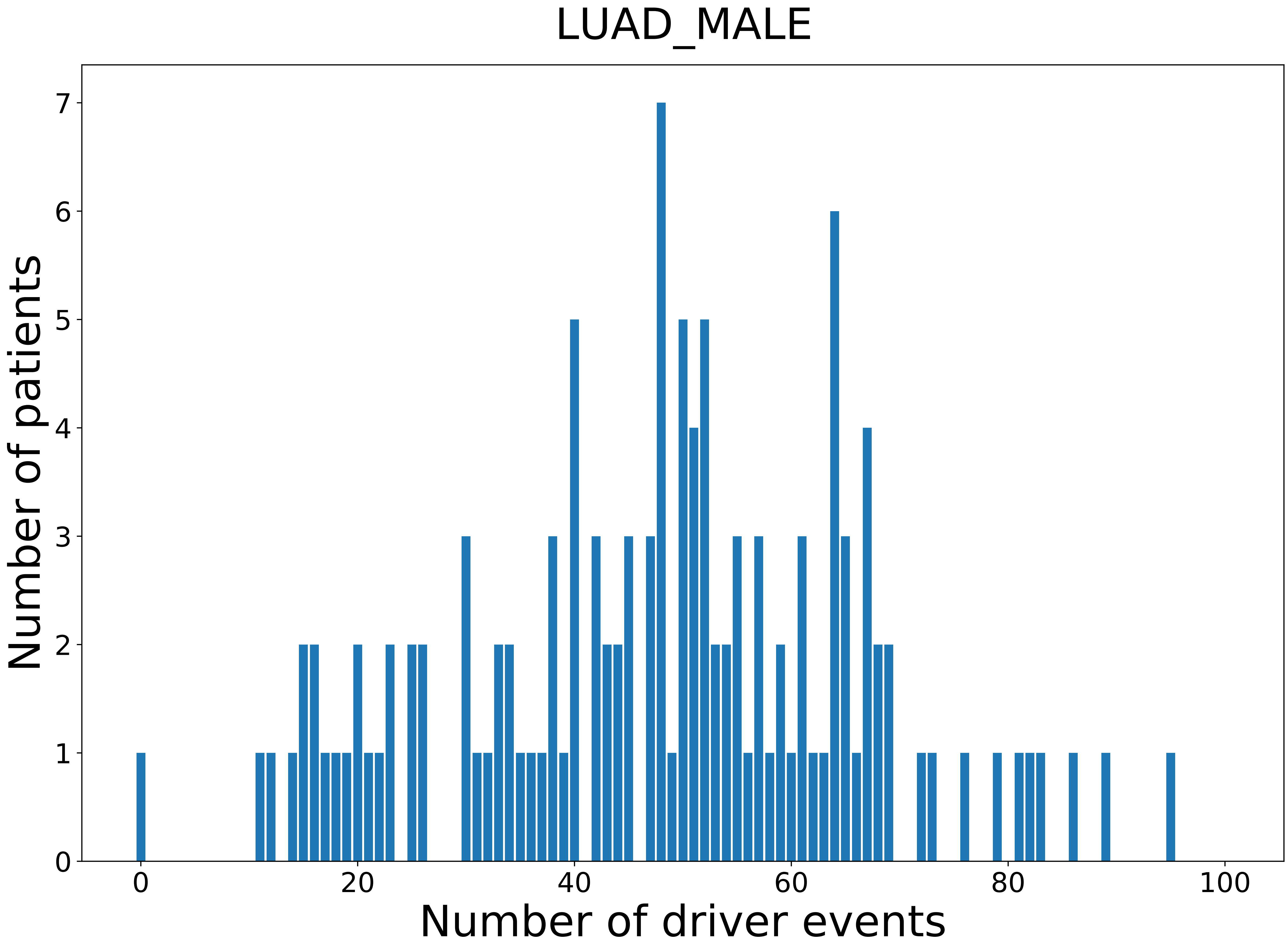

### 2021_8_16_20_32_LUSC_MALE.pdf

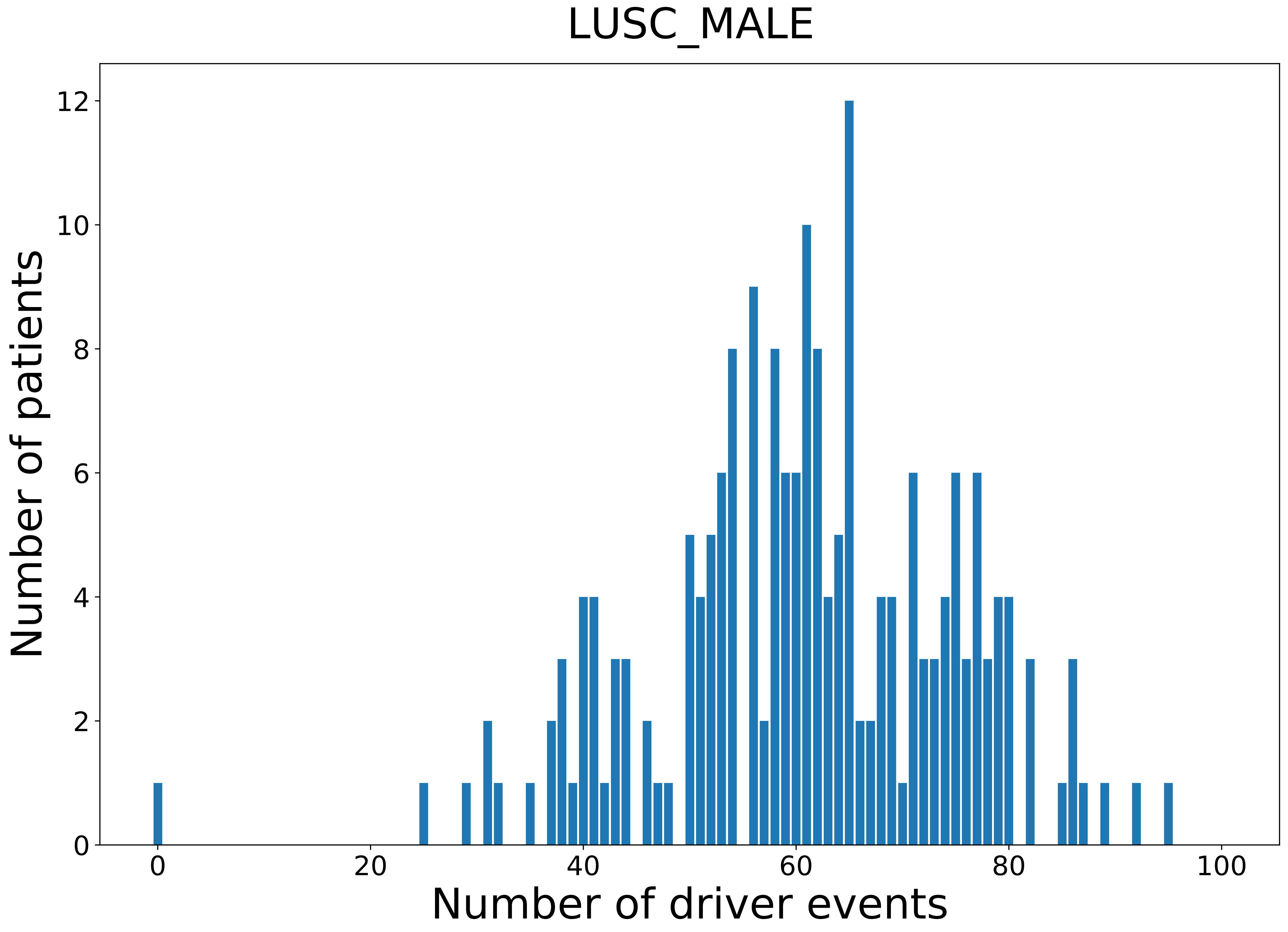

### 2021_8_16_20_32_MESO.pdf

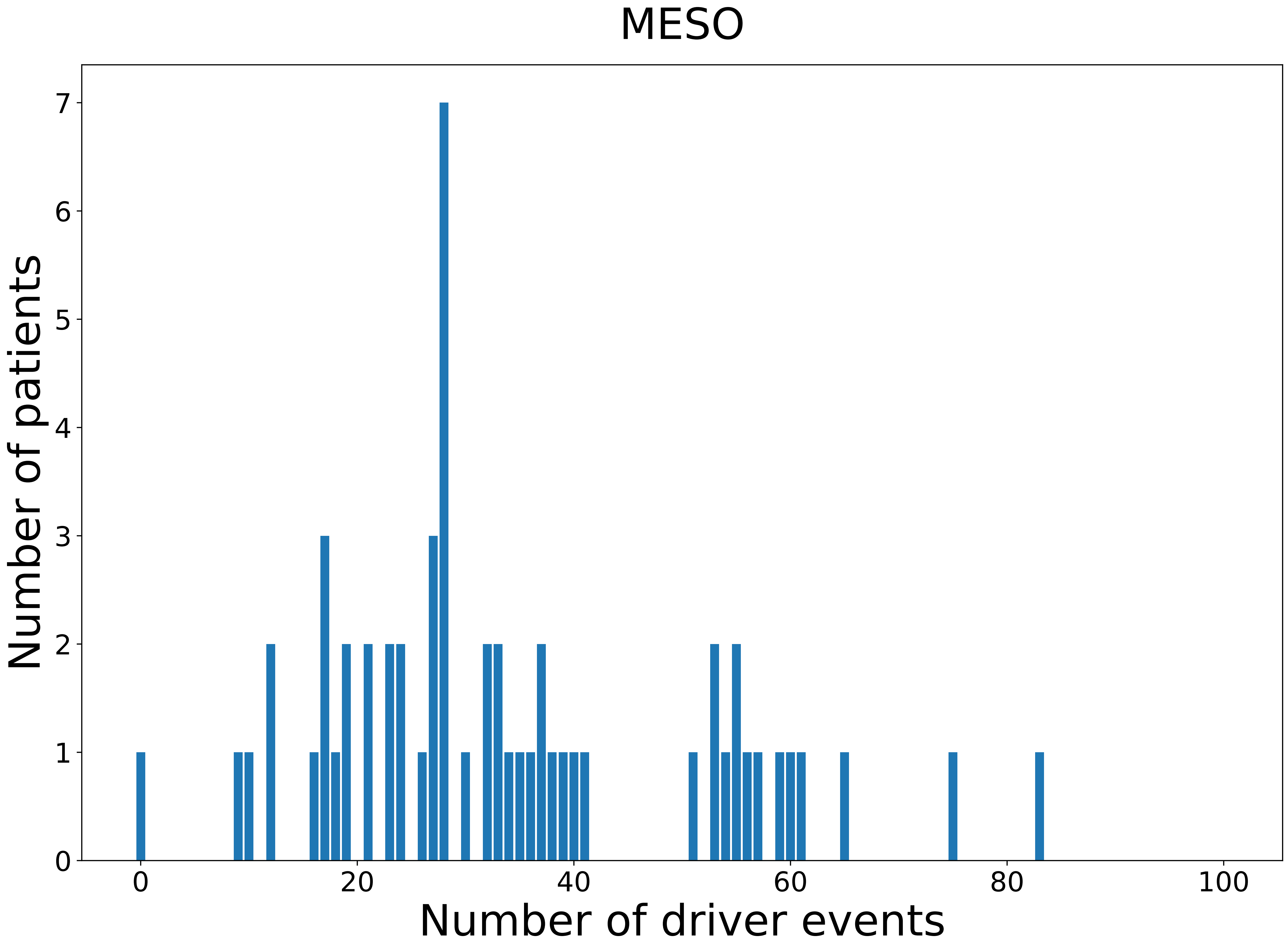

### 2021_8_16_20_32_MESO_FEMALE.pdf

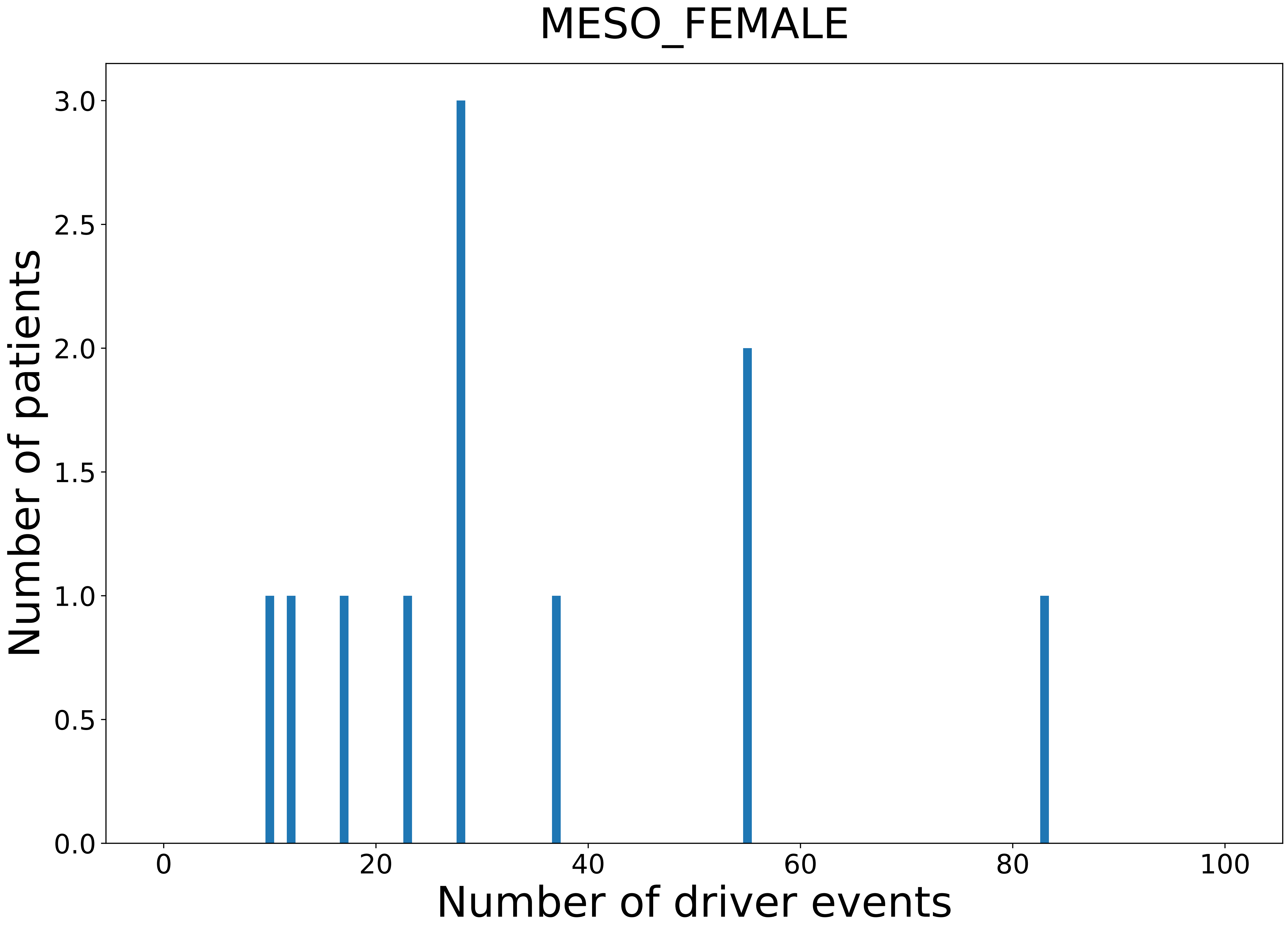

### 2021_8_16_20_32_OV.pdf

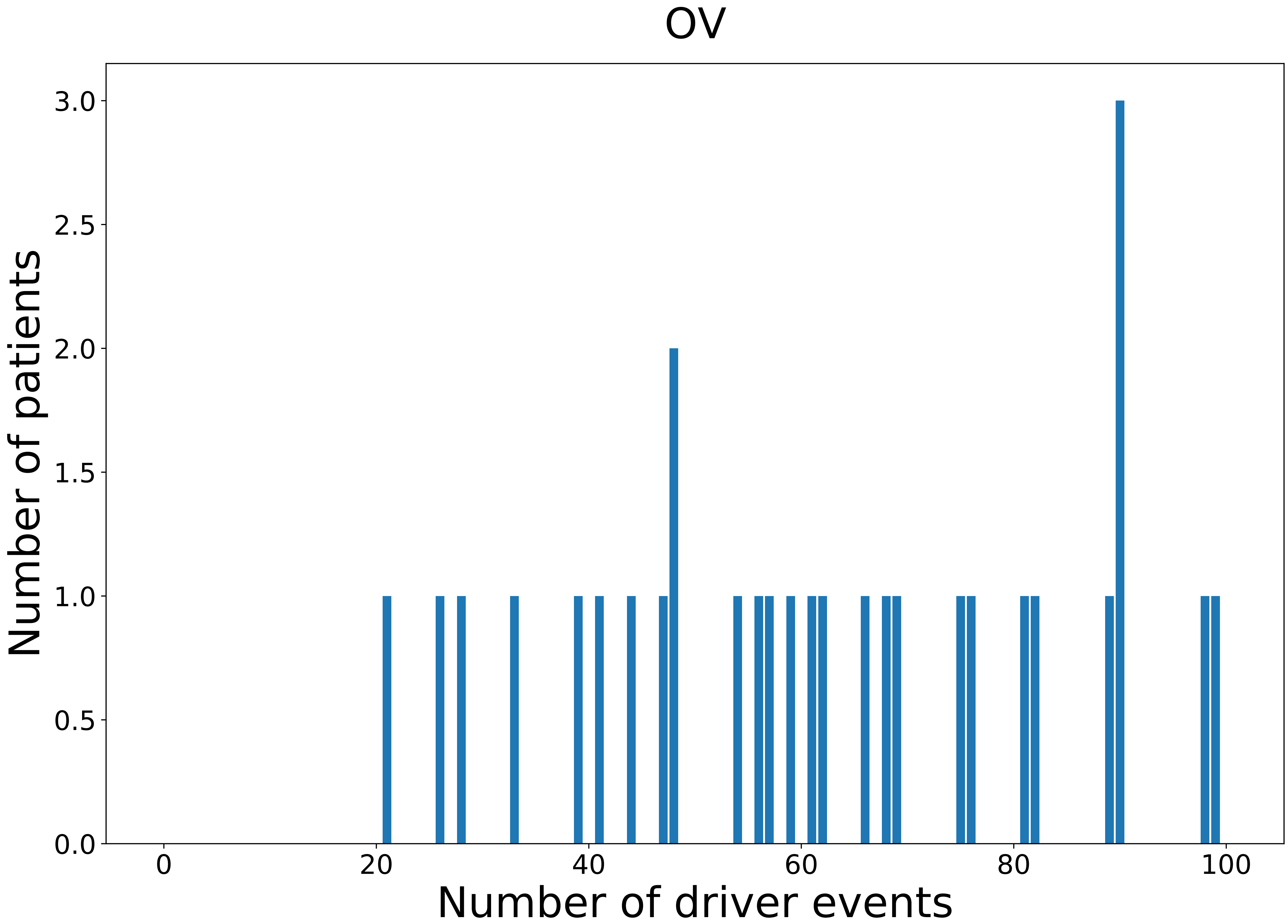

### 2021_8_16_20_32_OV_FEMALE.pdf

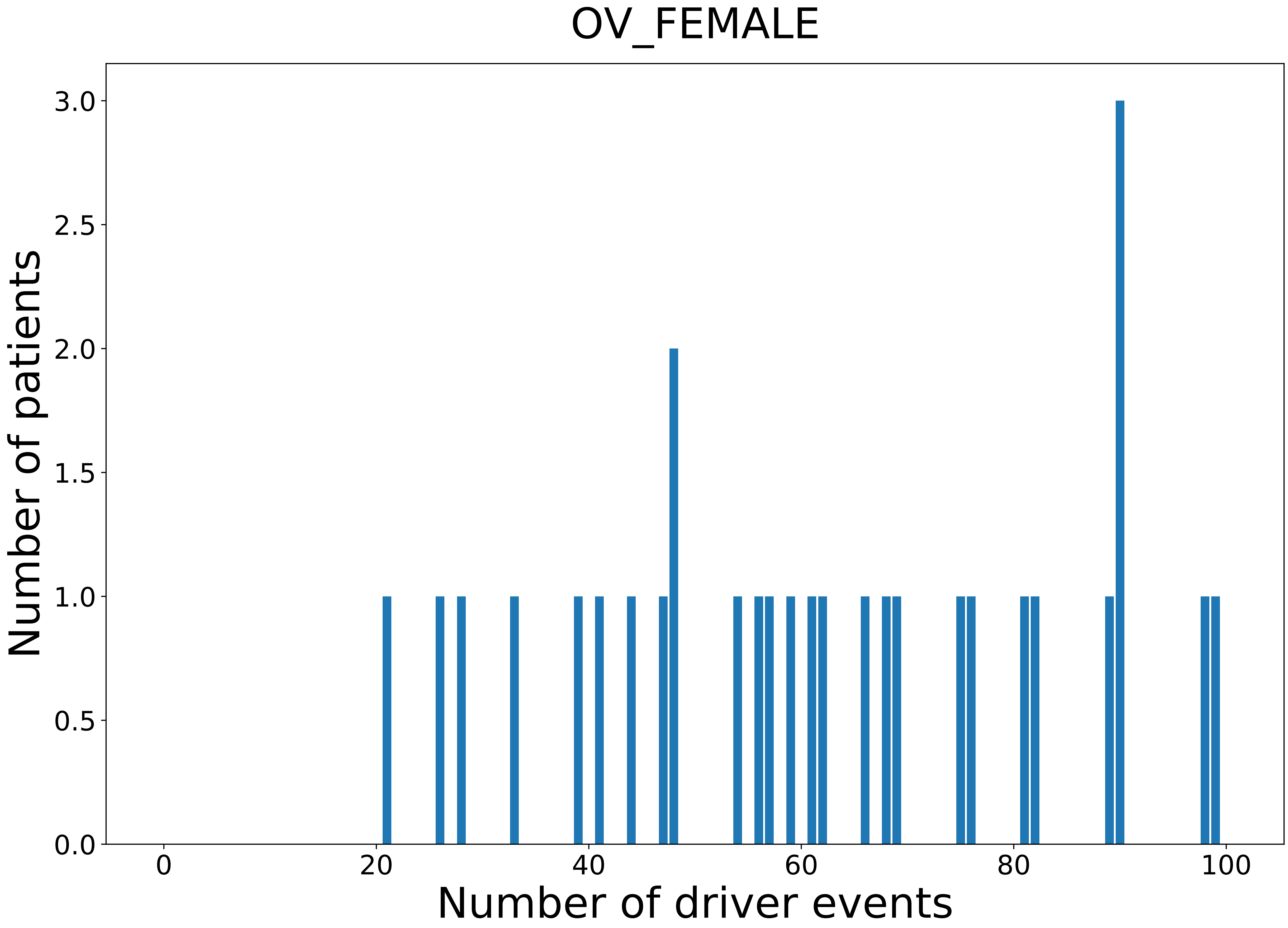

### 2021_8_16_20_32_PAAD.pdf

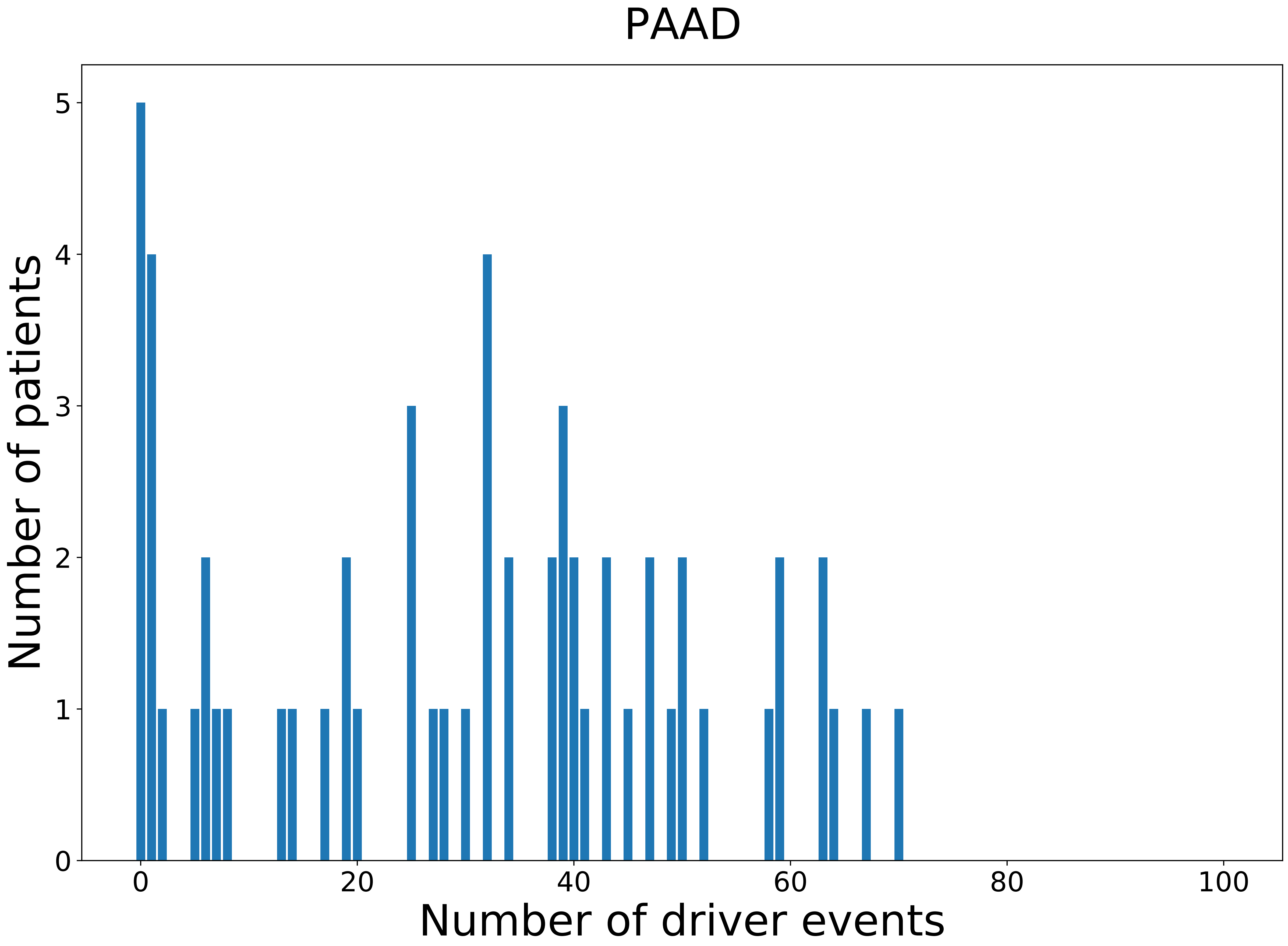

### 2021_8_16_20_32_PAAD_FEMALE.pdf

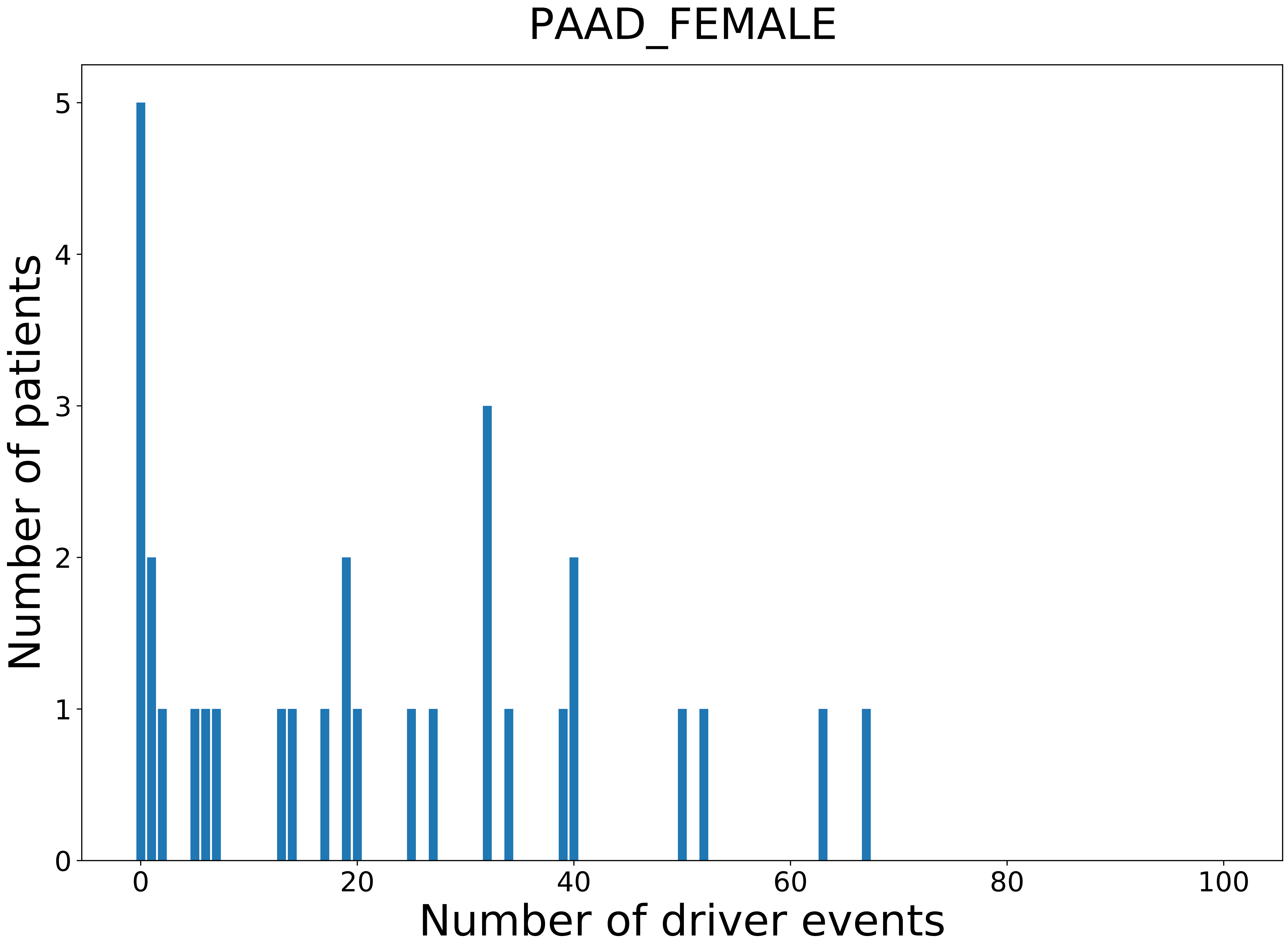

### 2021_8_16_20_32_PAAD_MALE.pdf

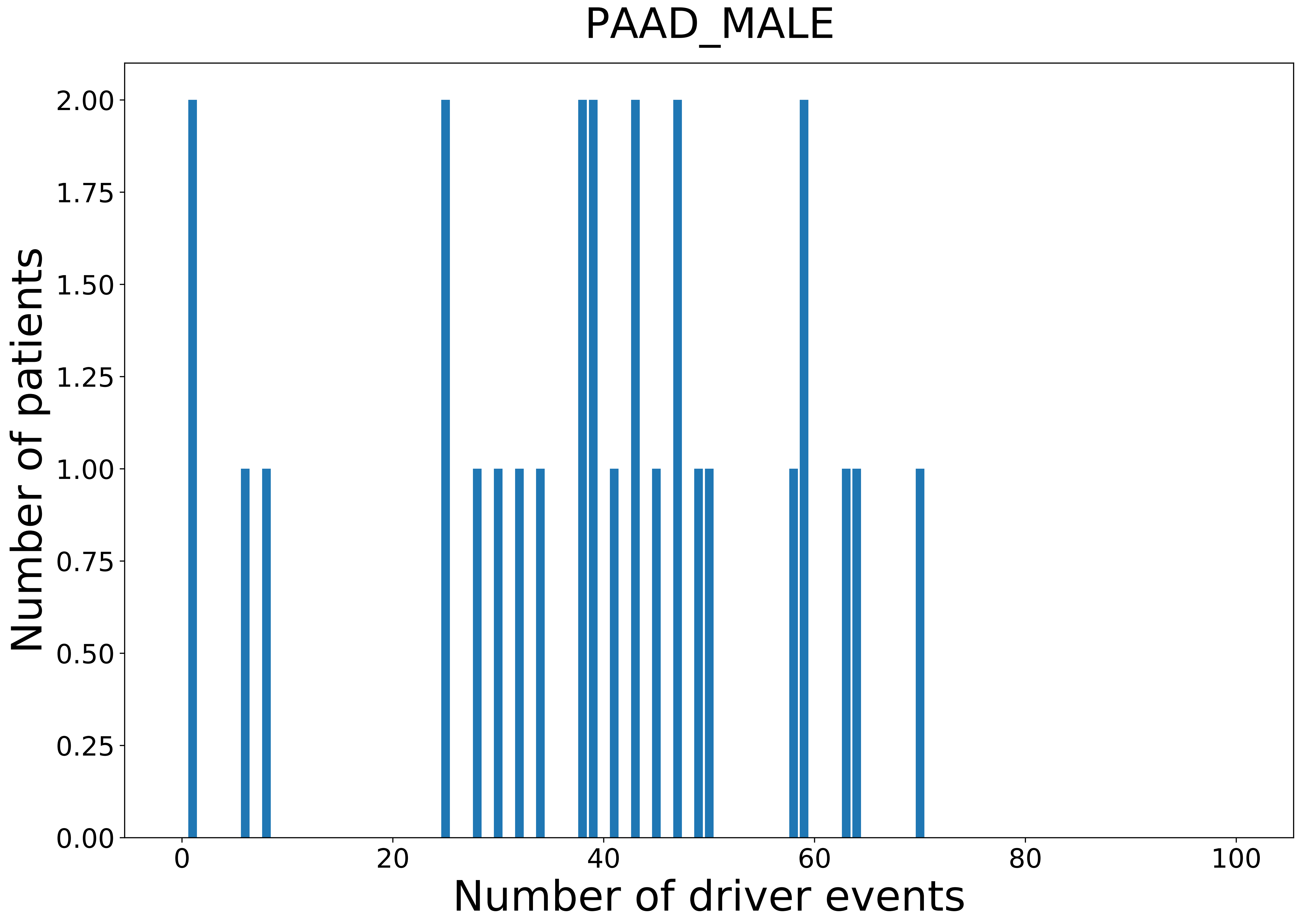

### 2021_8_16_20_32_PANCAN_FEMALE.pdf

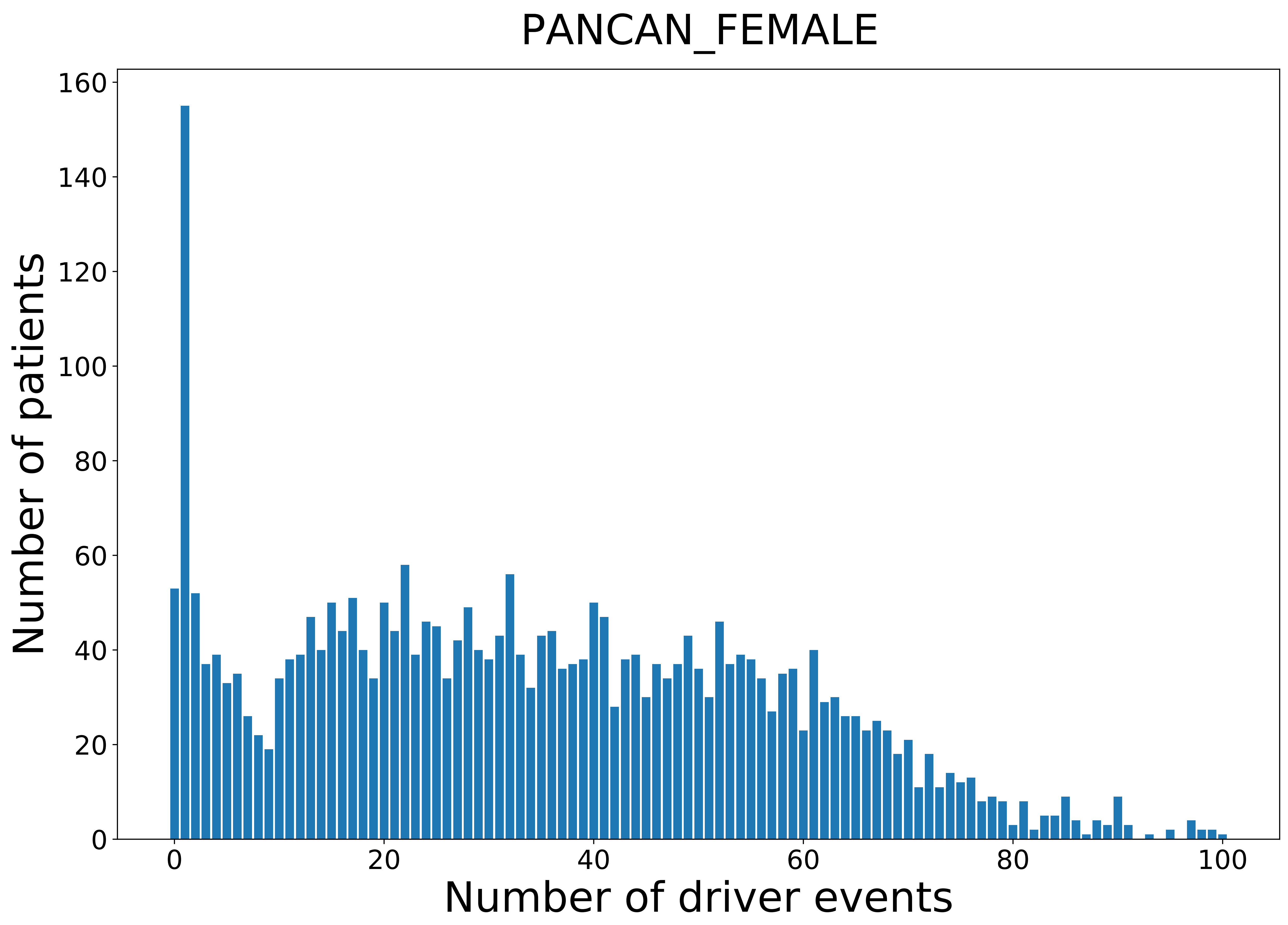

### 2021_8_16_20_32_PANCAN_MALE.pdf

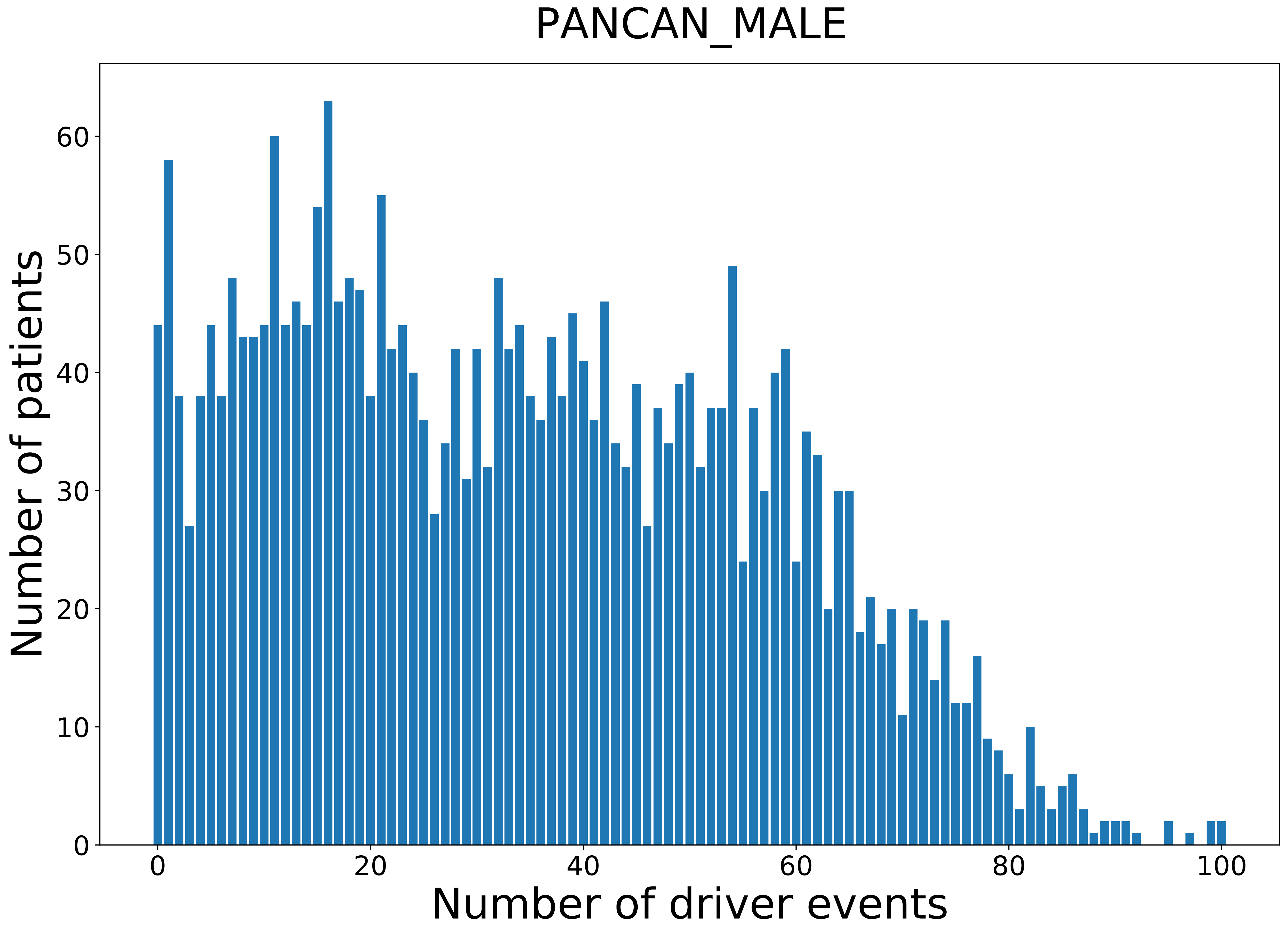

### 2021_8_16_20_32_PCPG_FEMALE.pdf

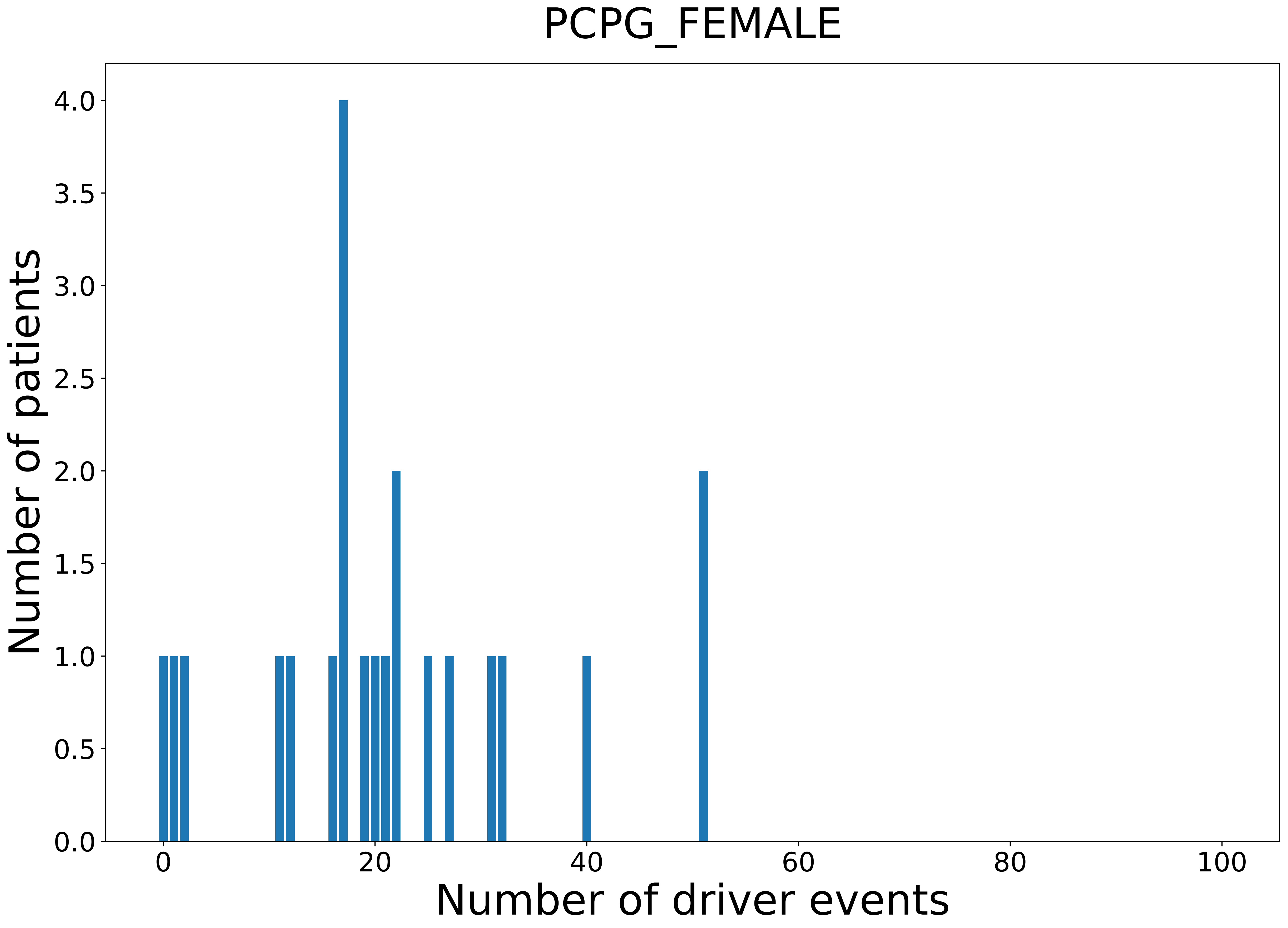

### 2021_8_16_20_32_PCPG_MALE.pdf

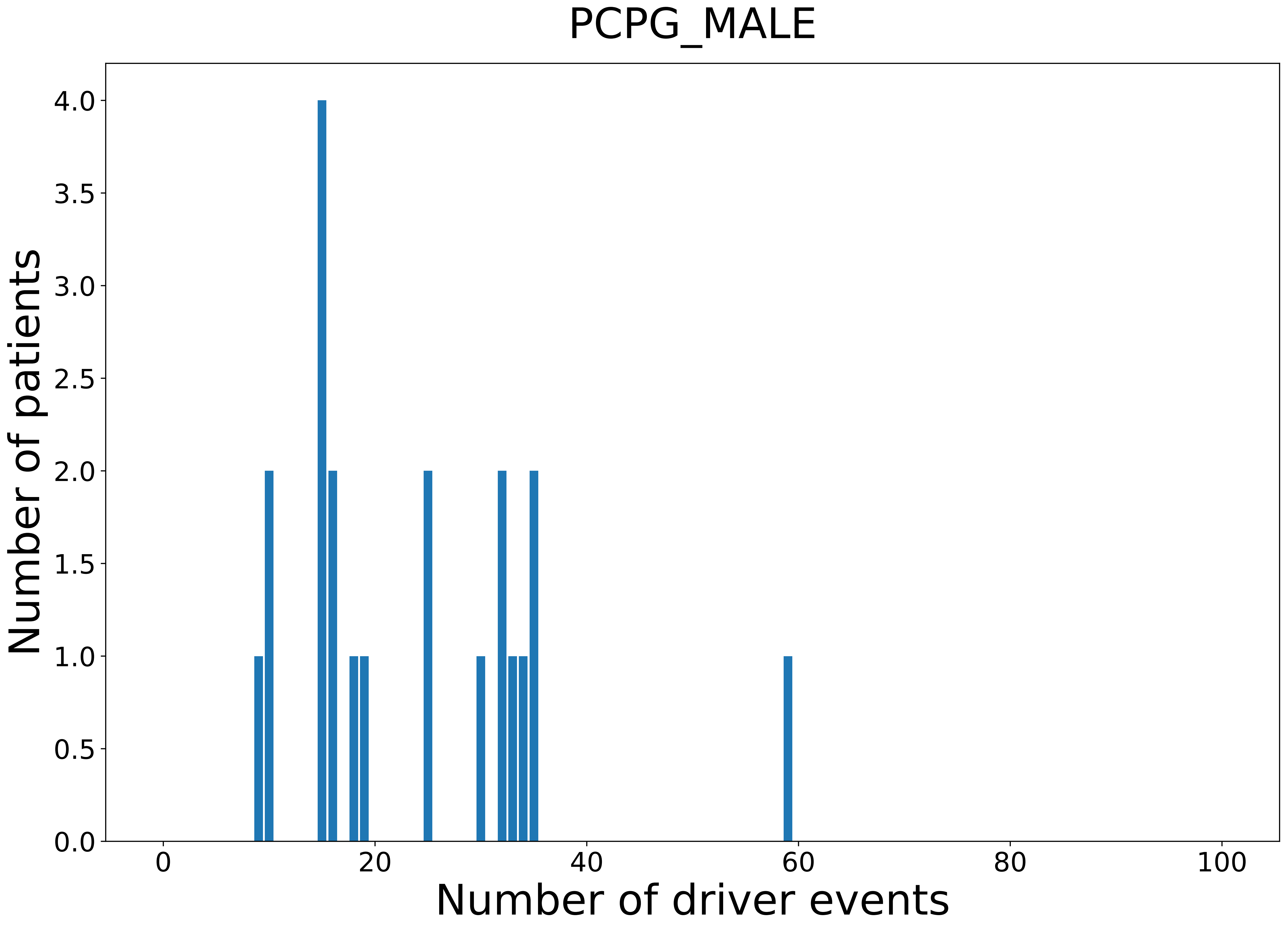

### 2021_8_16_20_32_PRAD.pdf

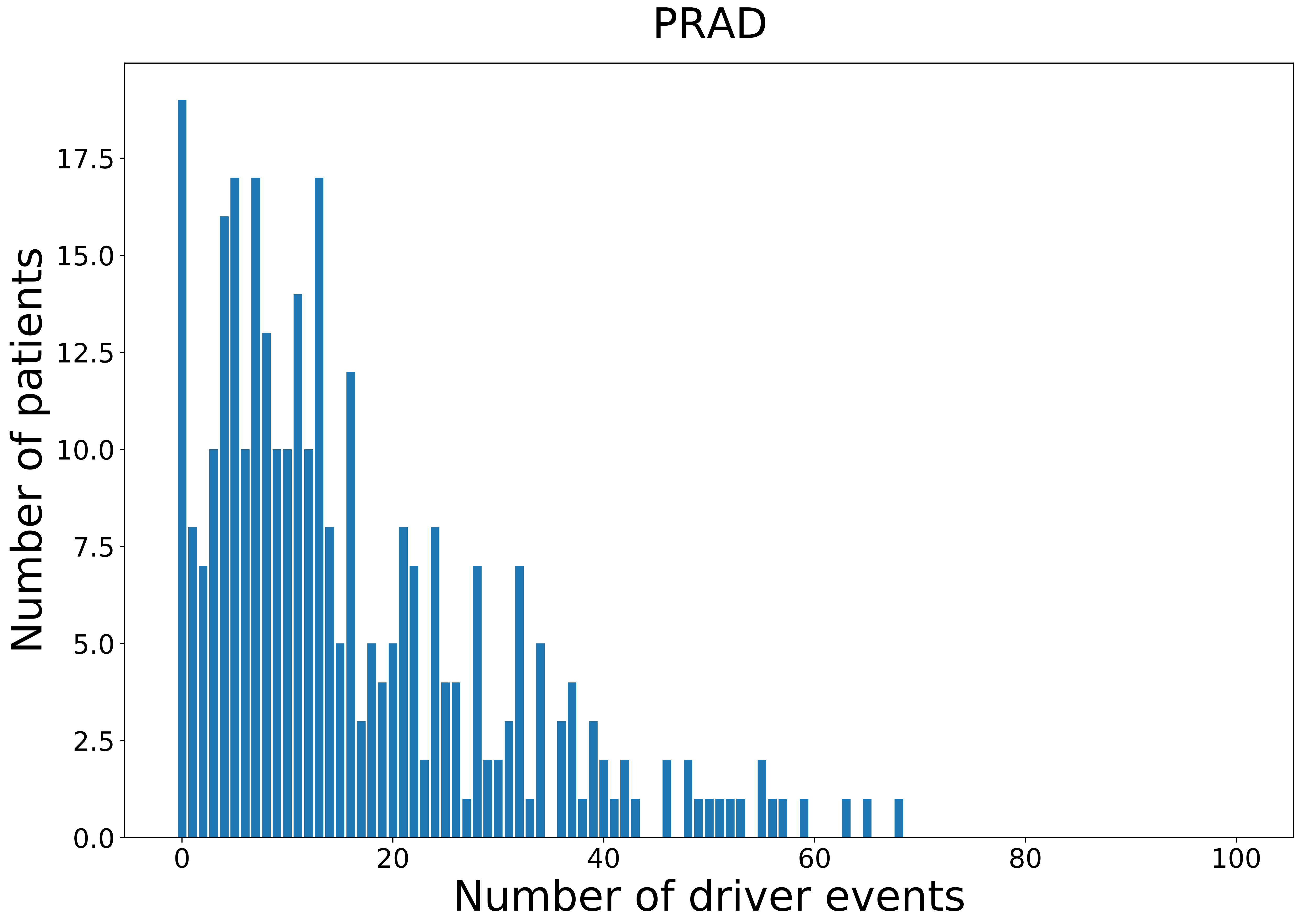

### 2021_8_16_20_32_PRAD_MALE.pdf

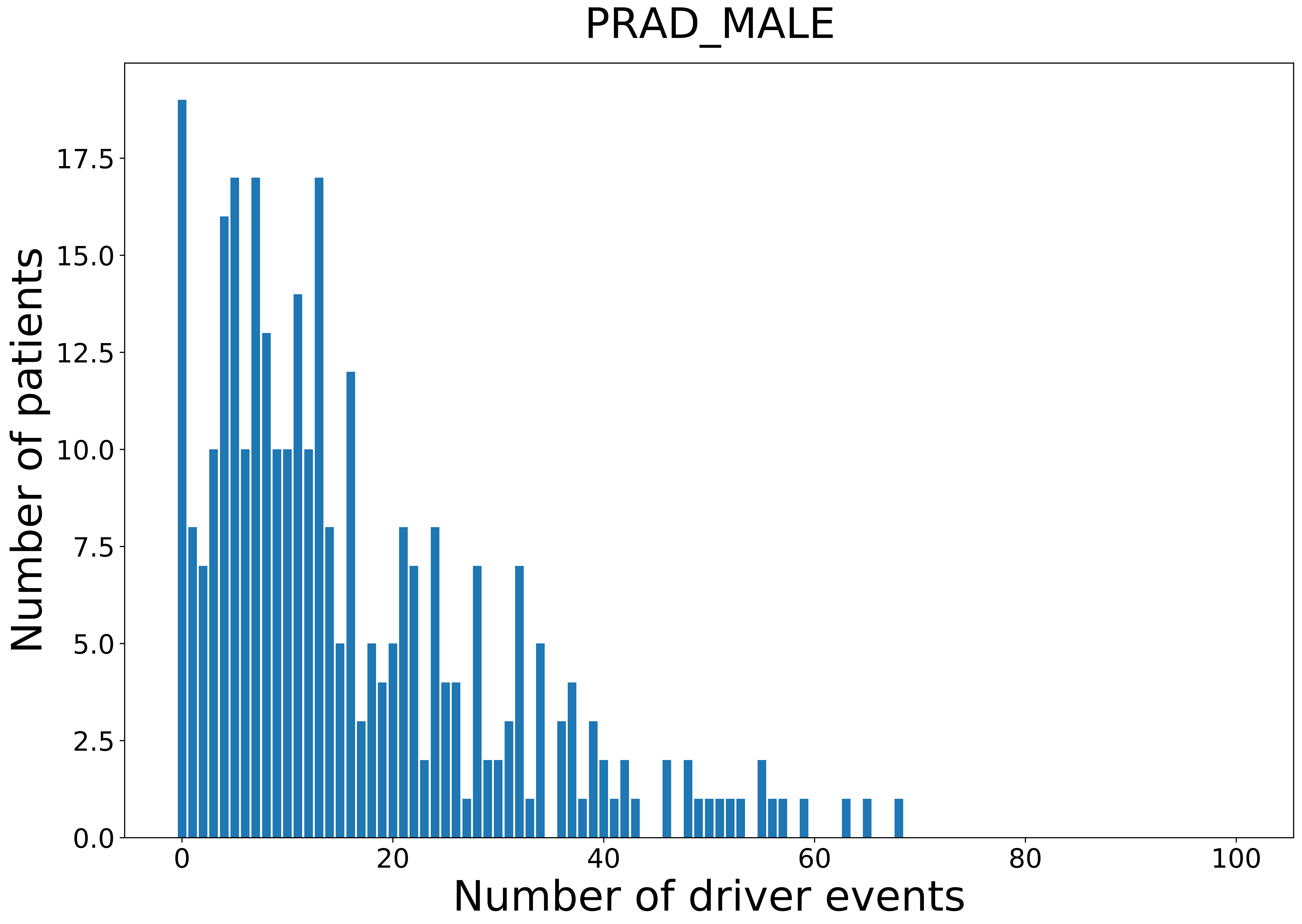

### 2021_8_16_20_32_READ.pdf

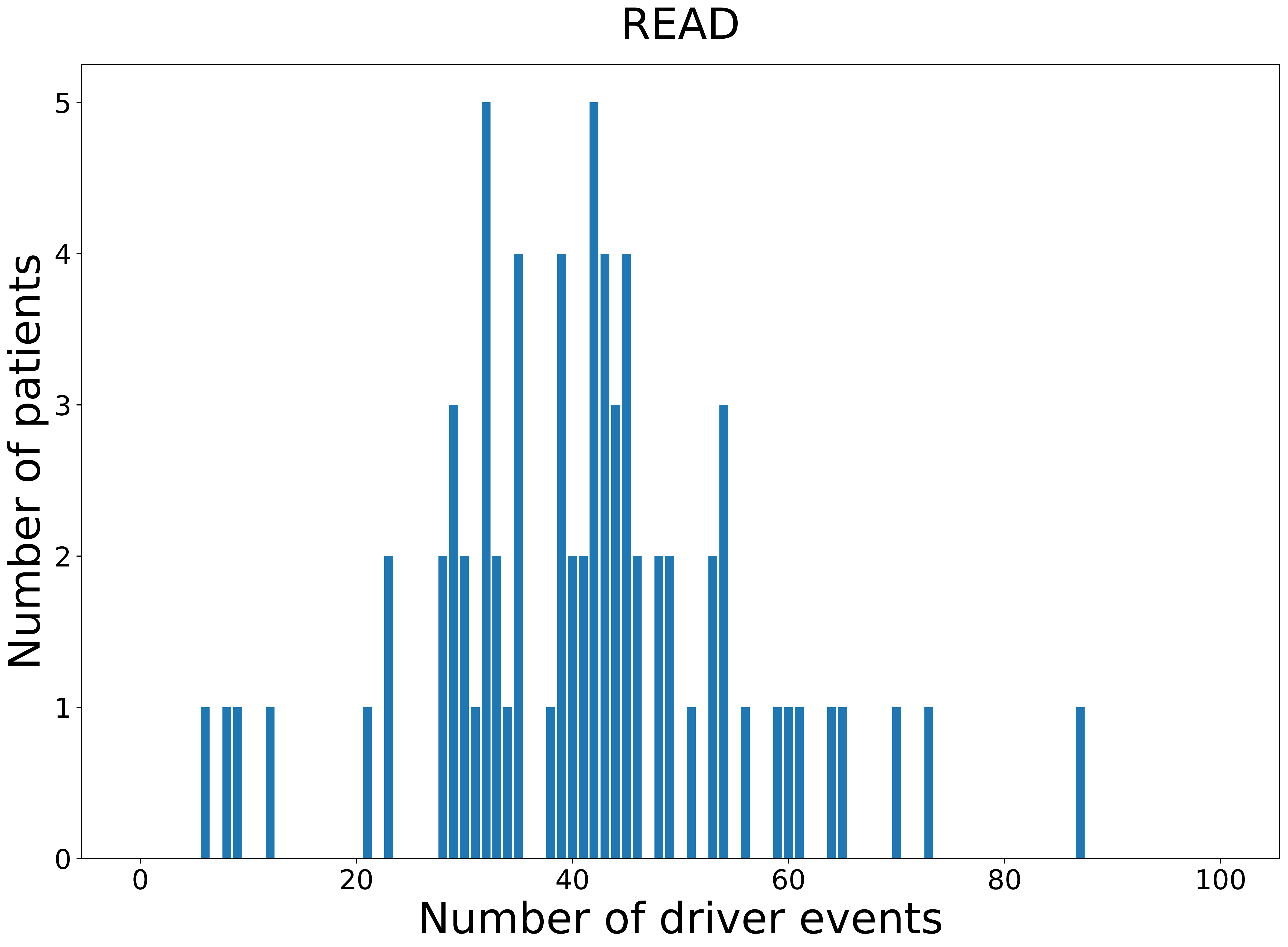

### 2021_8_16_20_32_READ_FEMALE.pdf

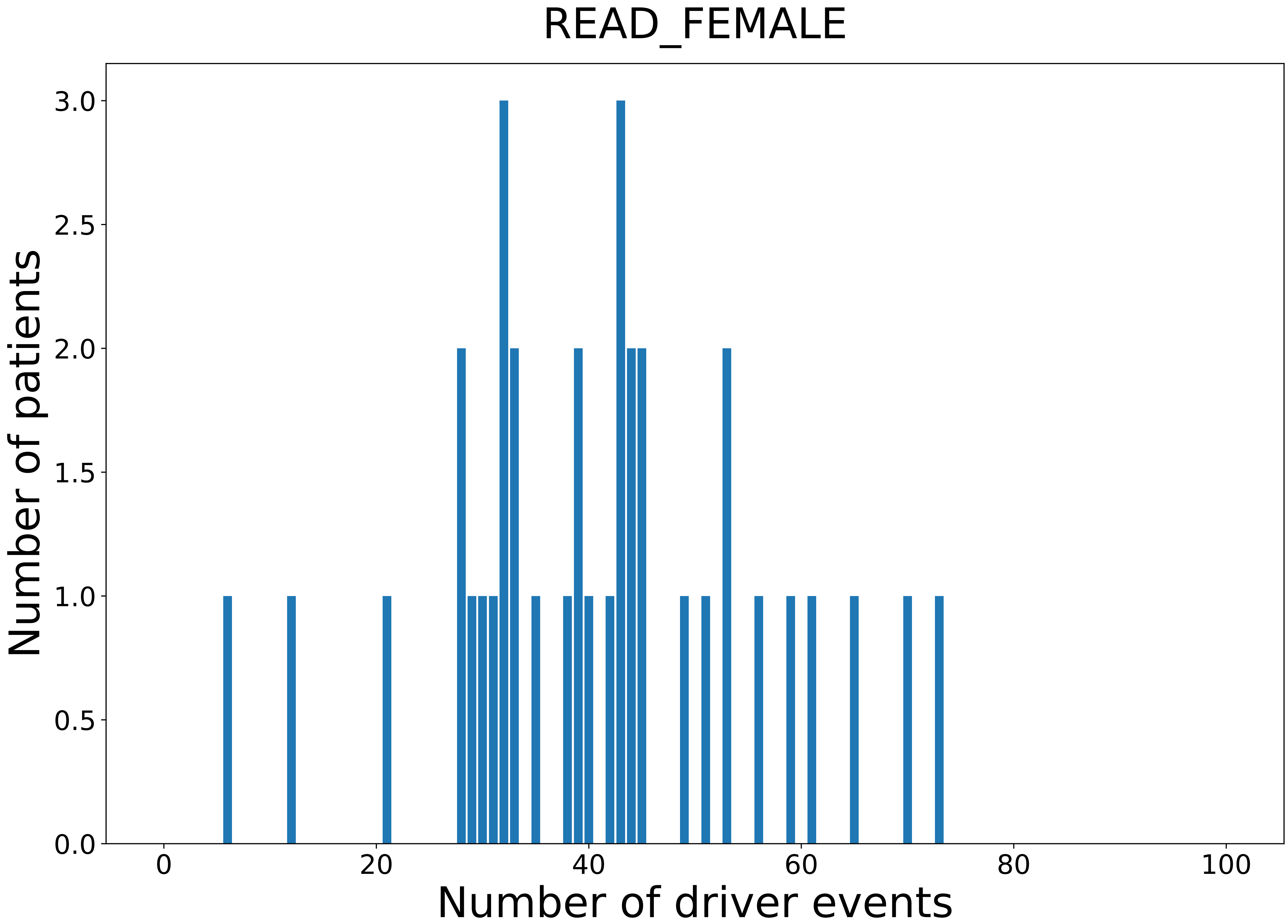

### 2021_8_16_20_32_READ_MALE.pdf

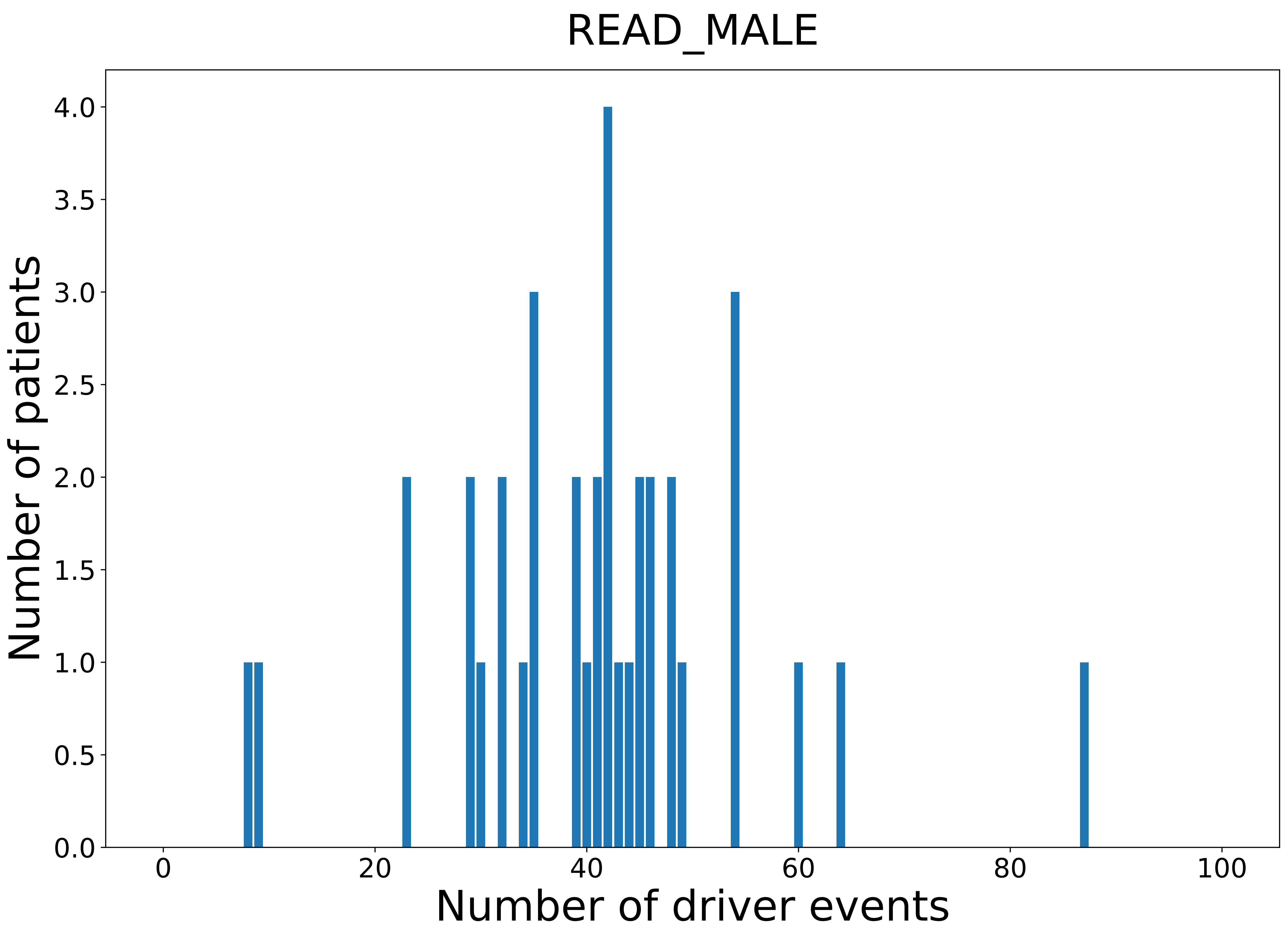

### 2021_8_16_20_32_SARC.pdf

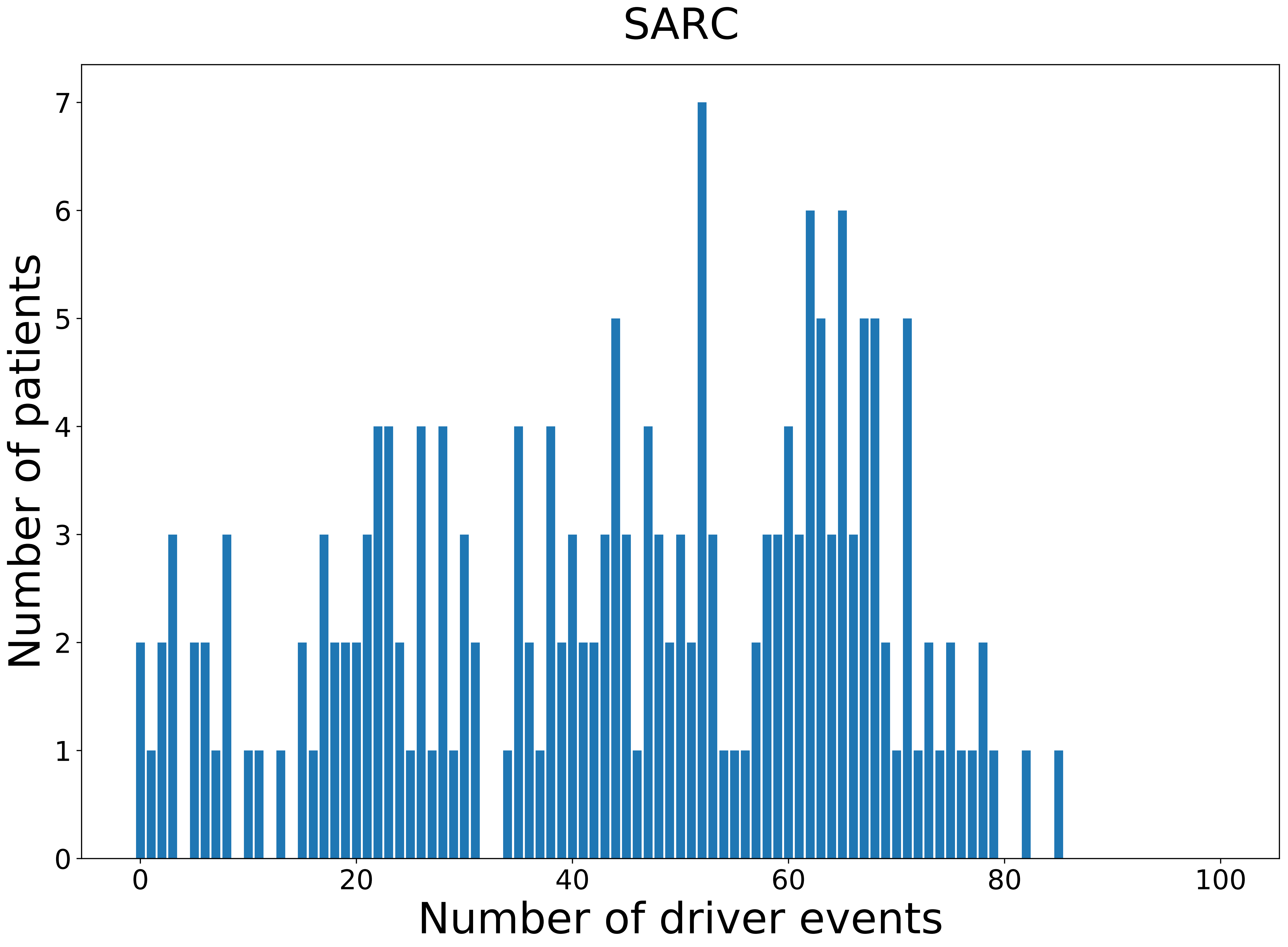

### 2021_8_16_20_32_SARC_FEMALE.pdf

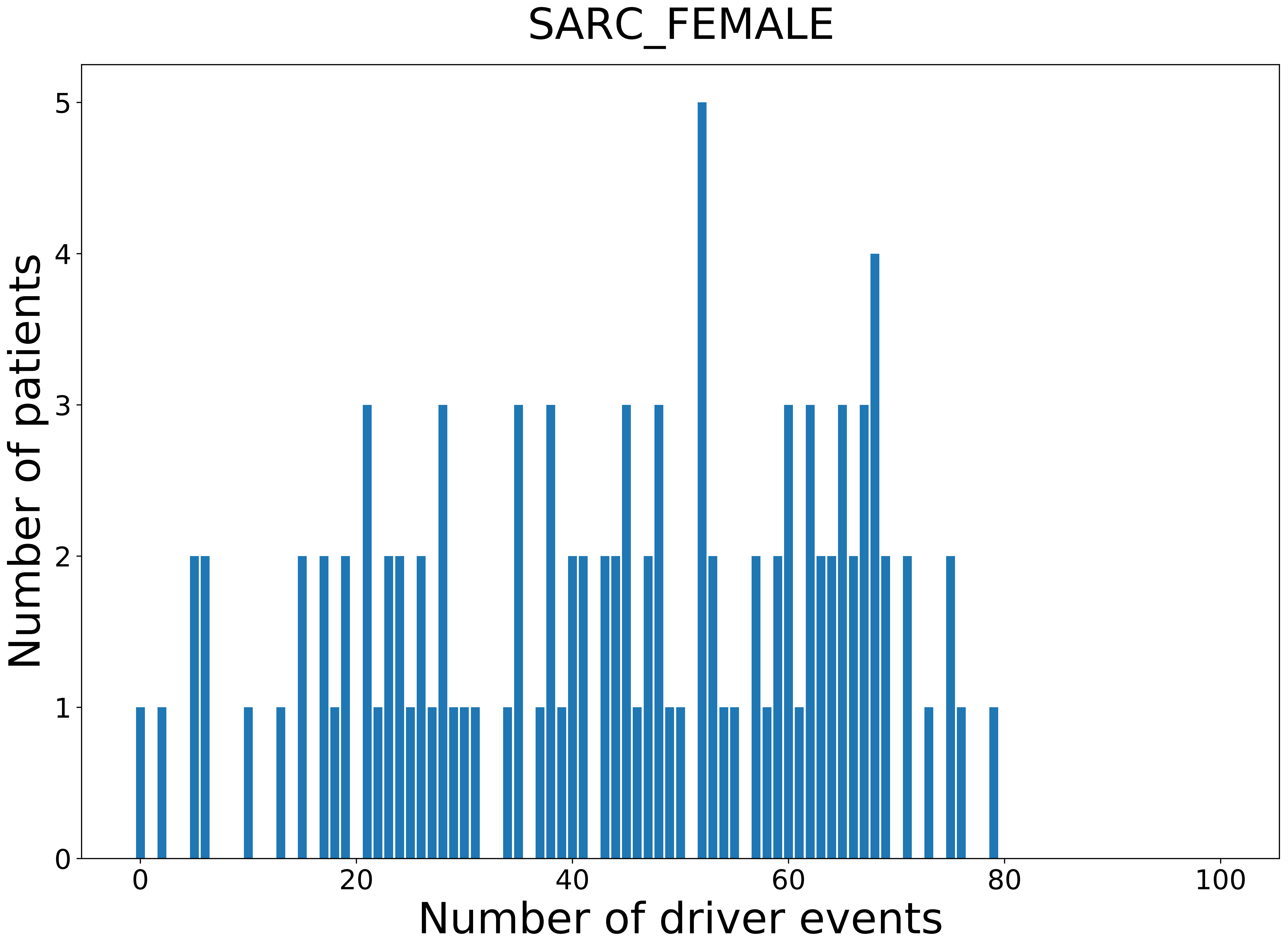

### 2021_8_16_20_32_SARC_MALE.pdf

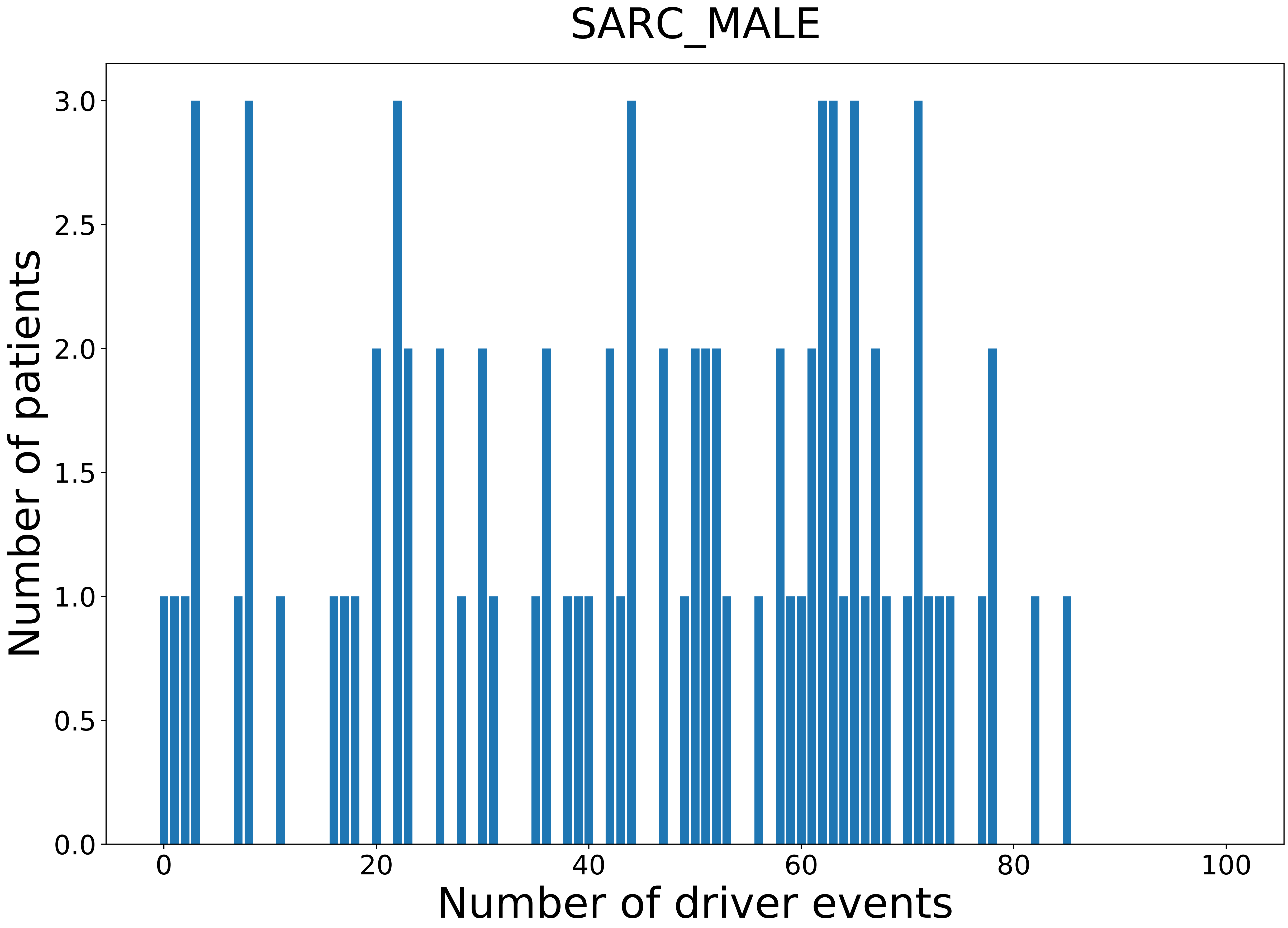

### 2021_8_16_20_32_SKCM.pdf

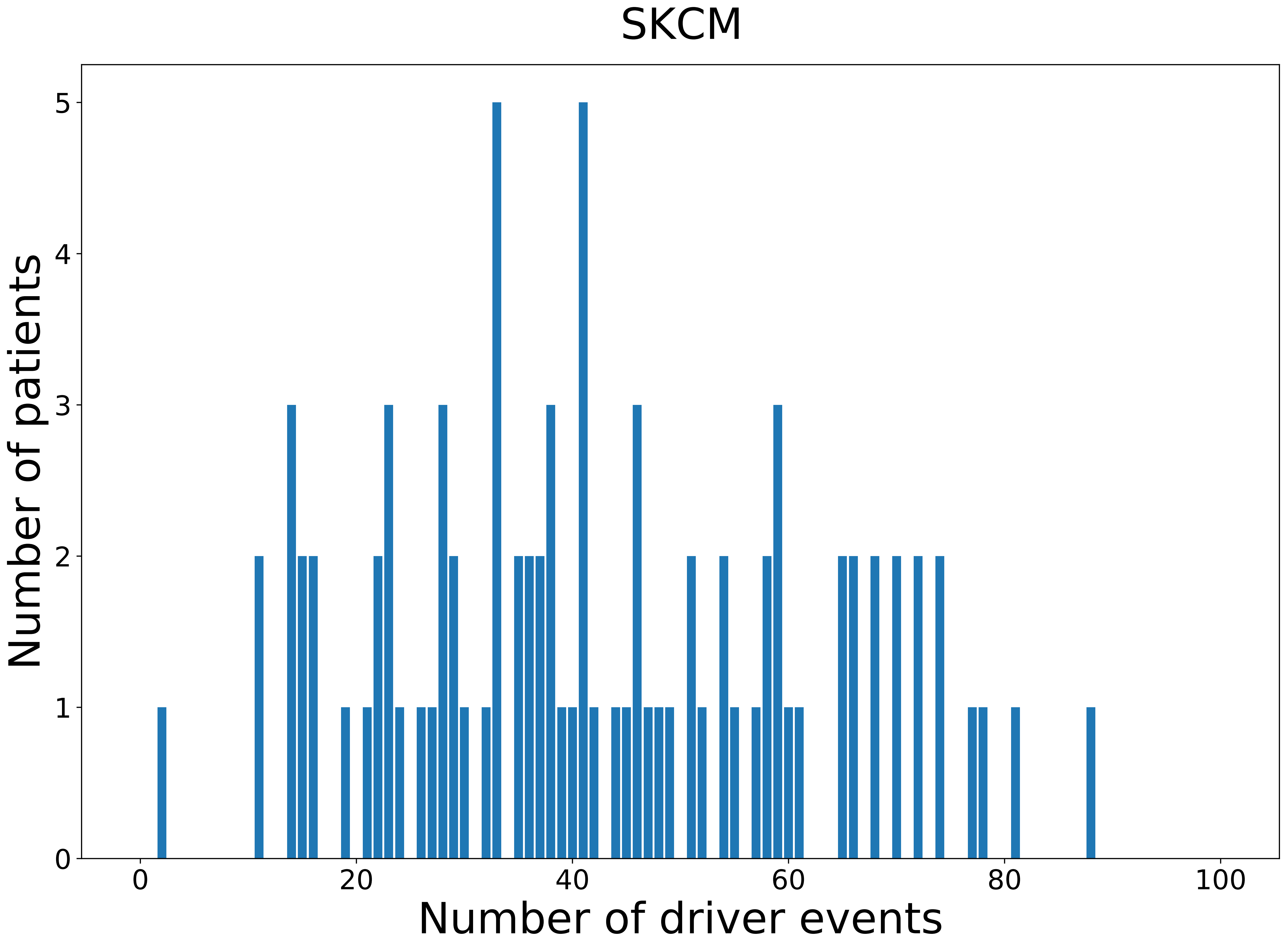

### 2021_8_16_20_32_SKCM_FEMALE.pdf

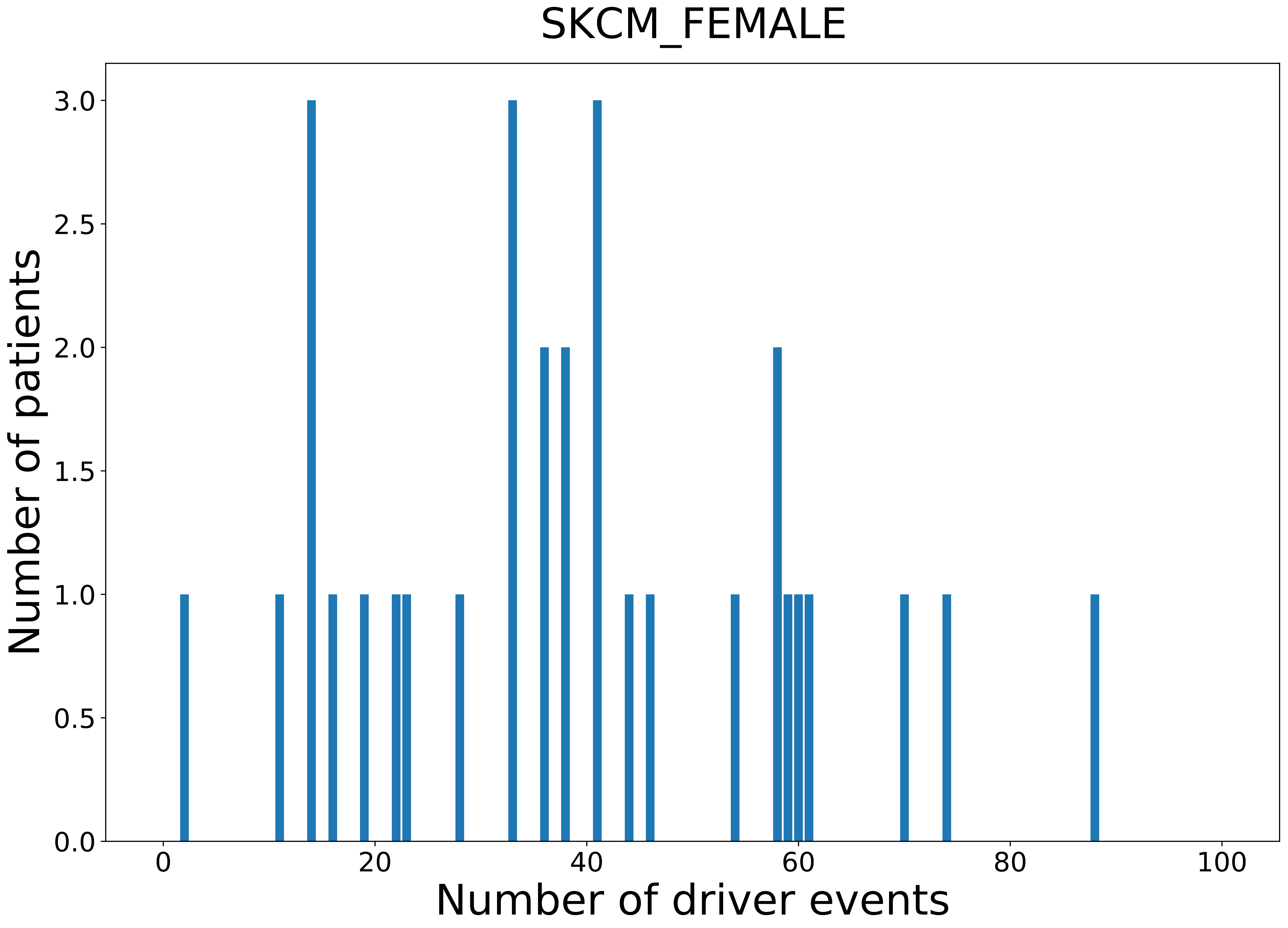

### 2021_8_16_20_32_SKCM_MALE.pdf

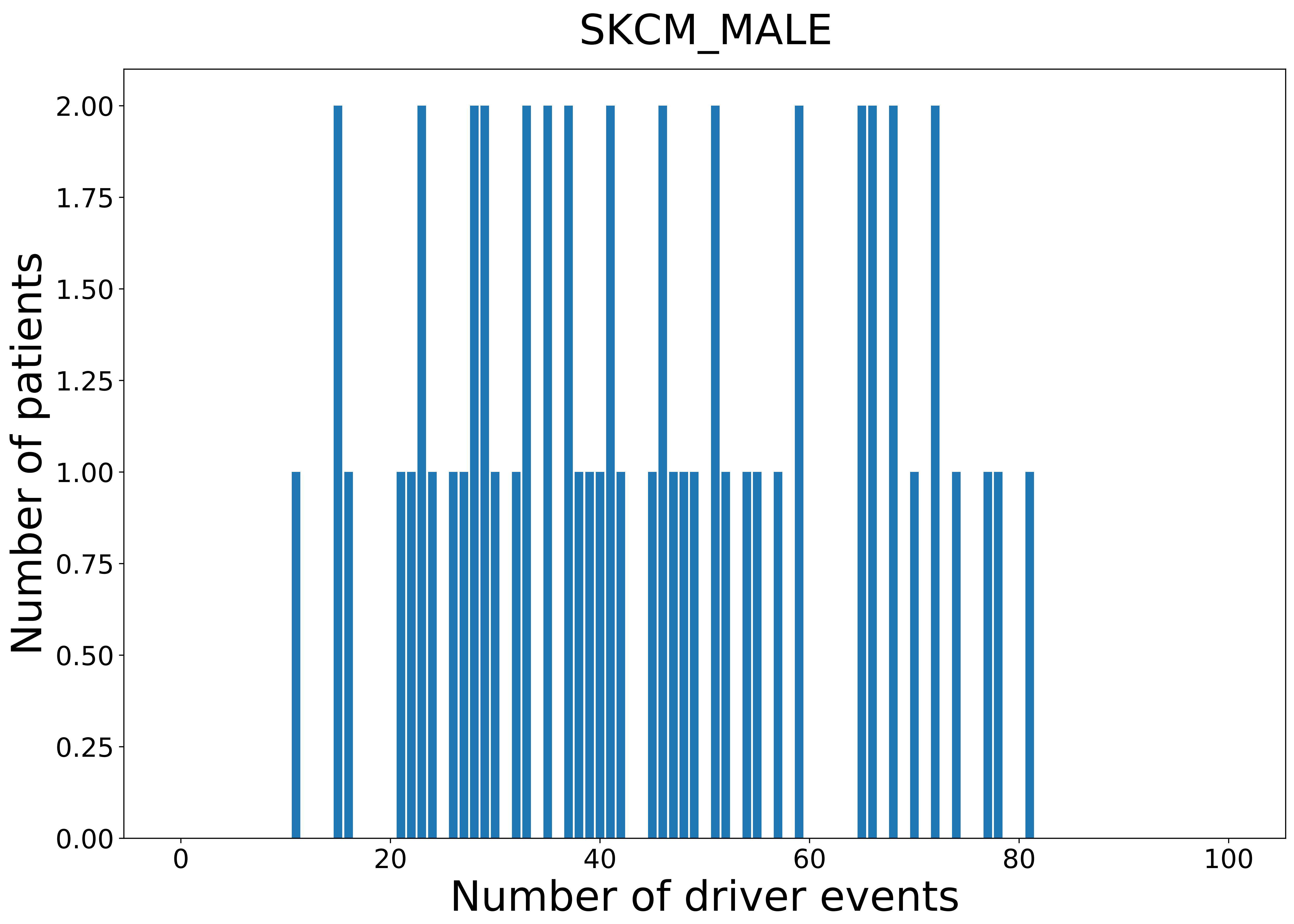

### 2021_8_16_20_32_STAD.pdf

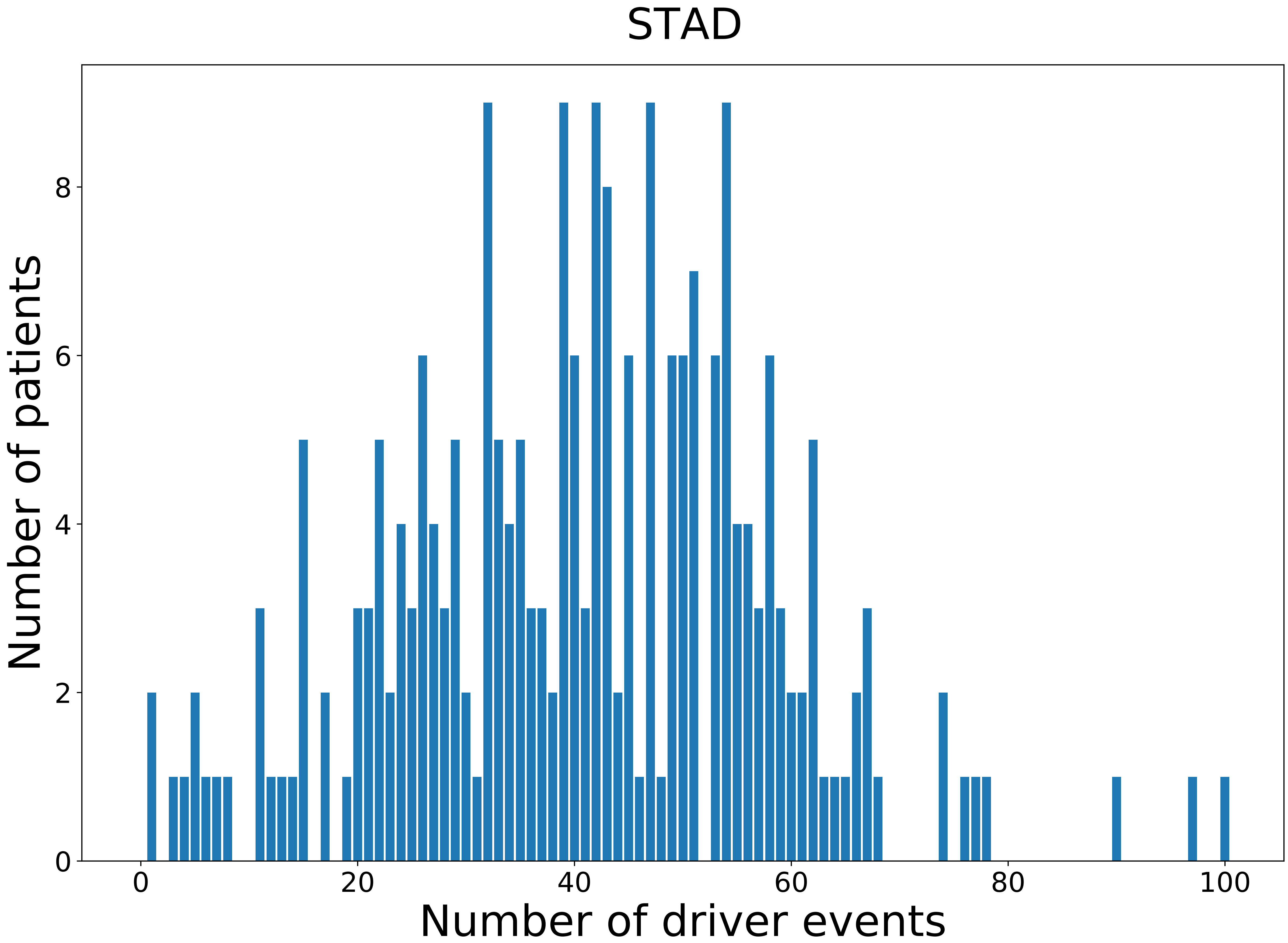

### 2021_8_16_20_32_STAD_FEMALE.pdf

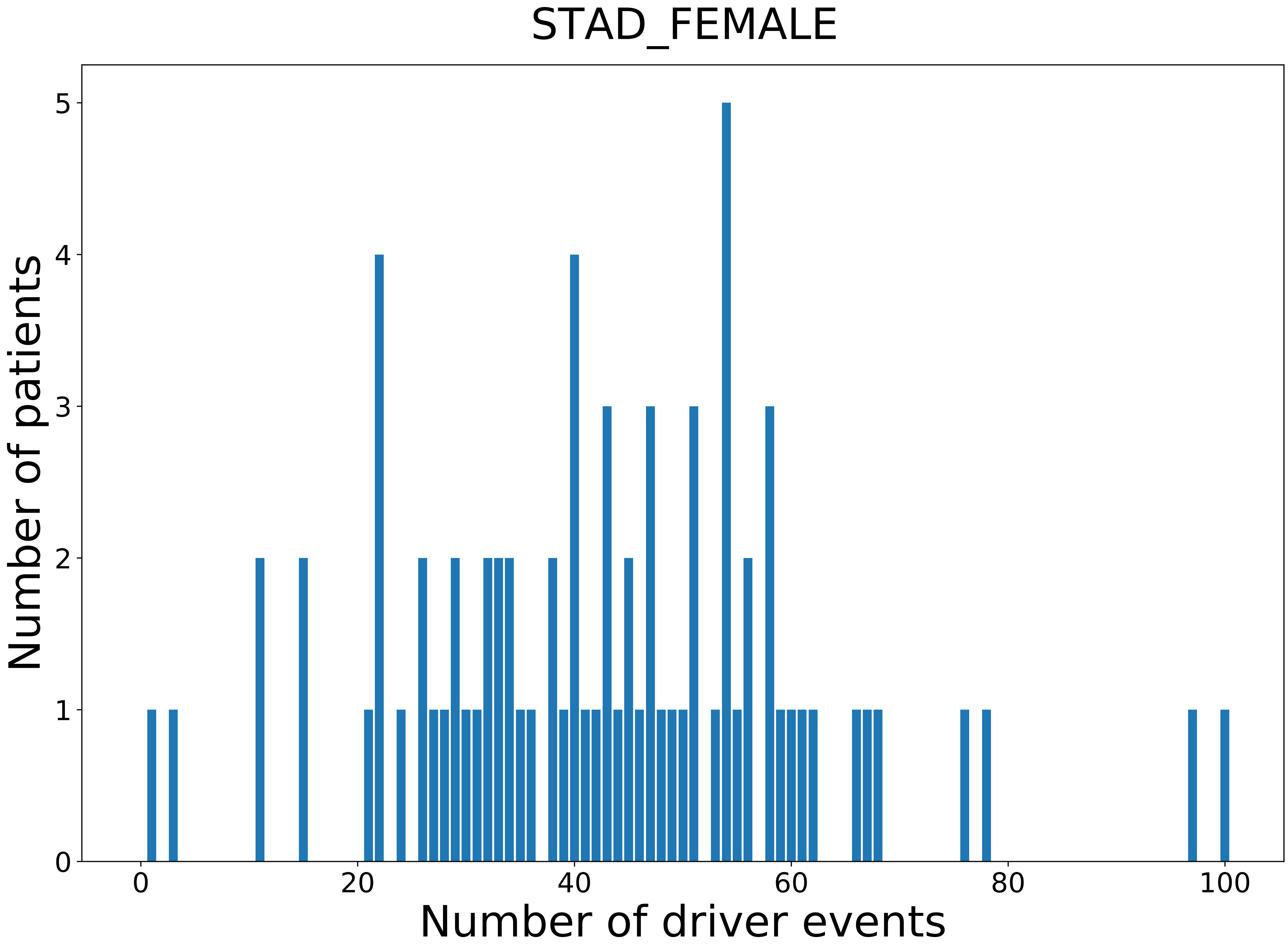

### 2021_8_16_20_32_STAD_MALE.pdf

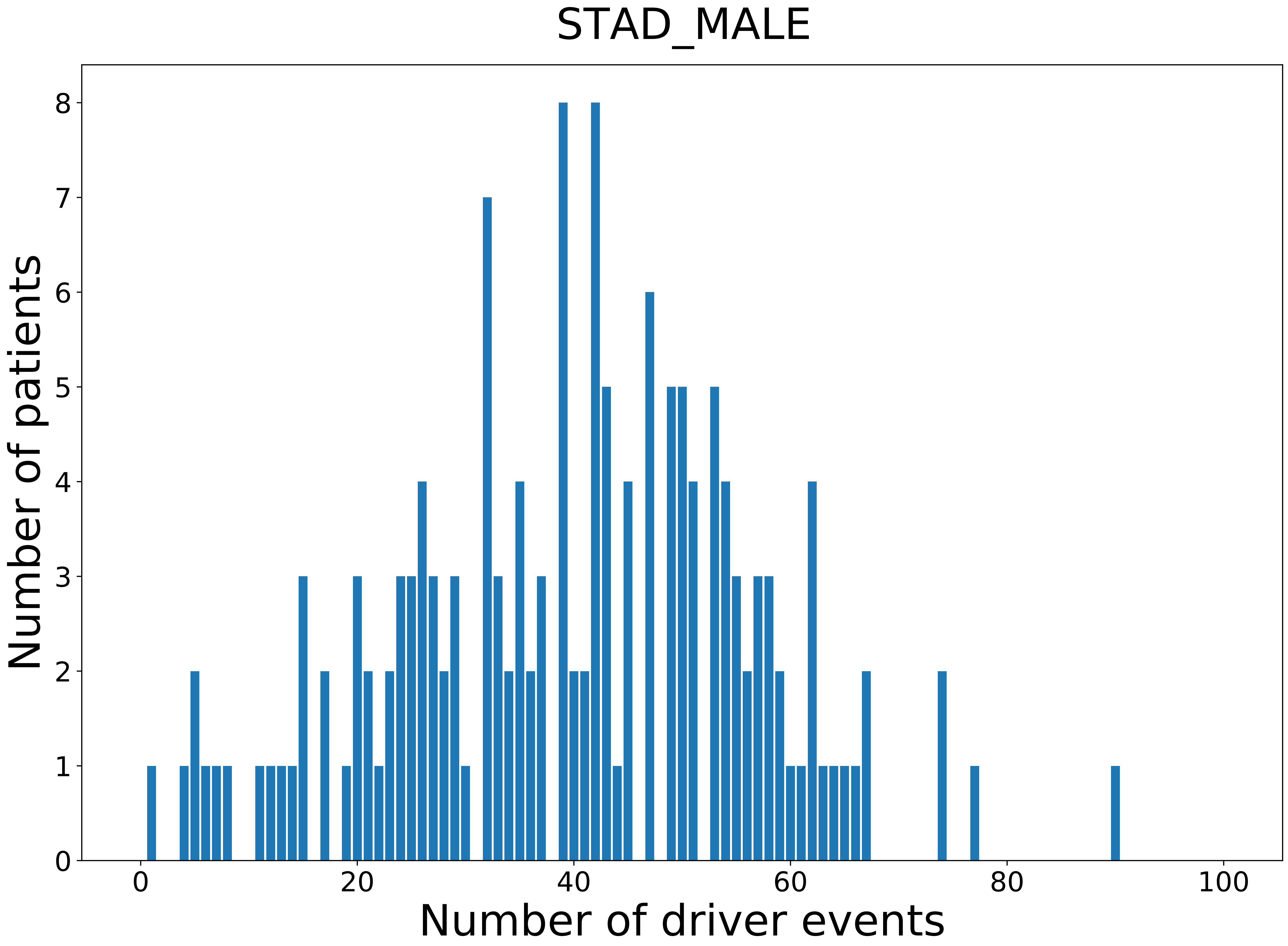

### 2021_8_16_20_32_TGCT.pdf

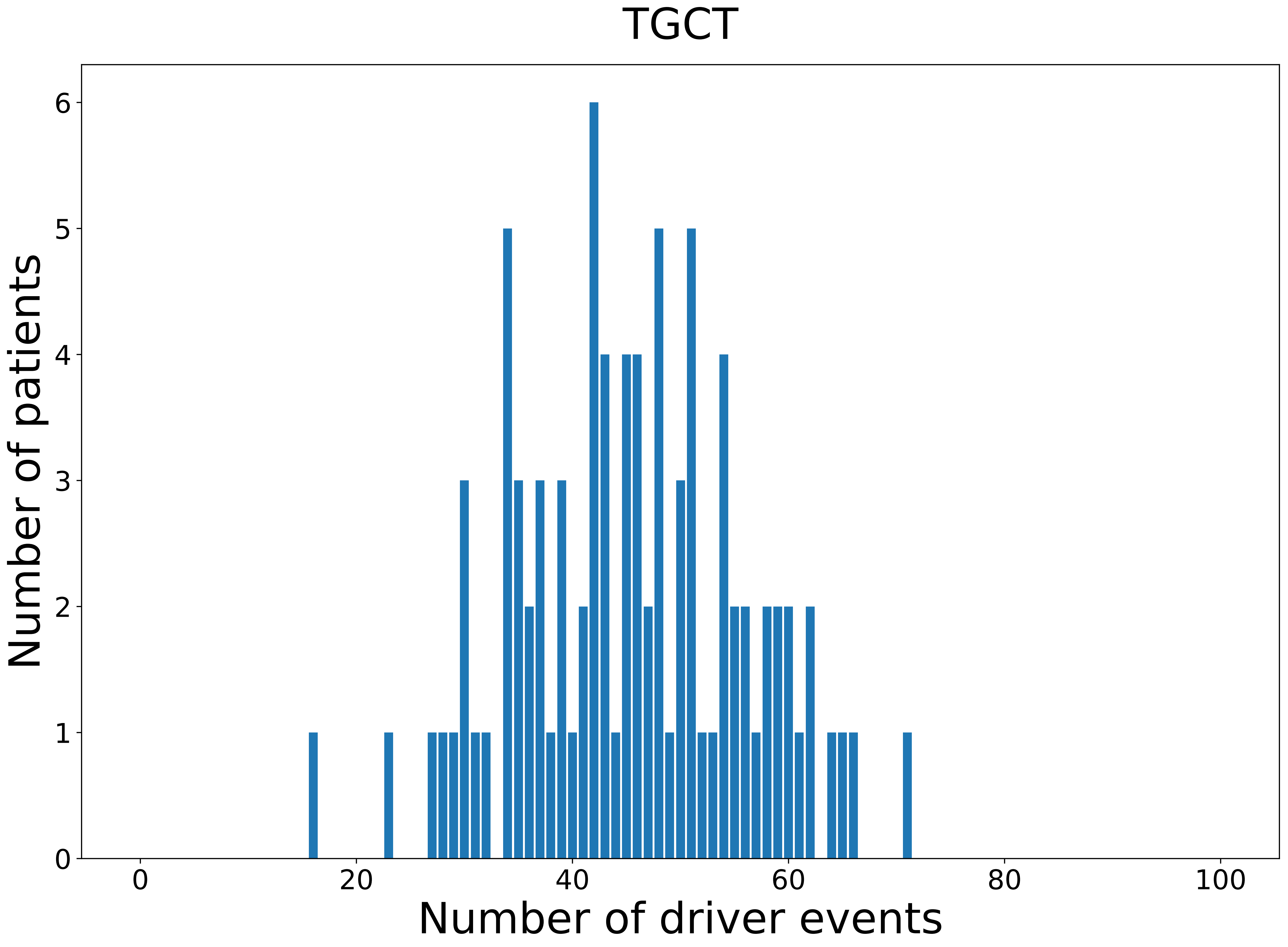

### 2021_8_16_20_32_THCA.pdf

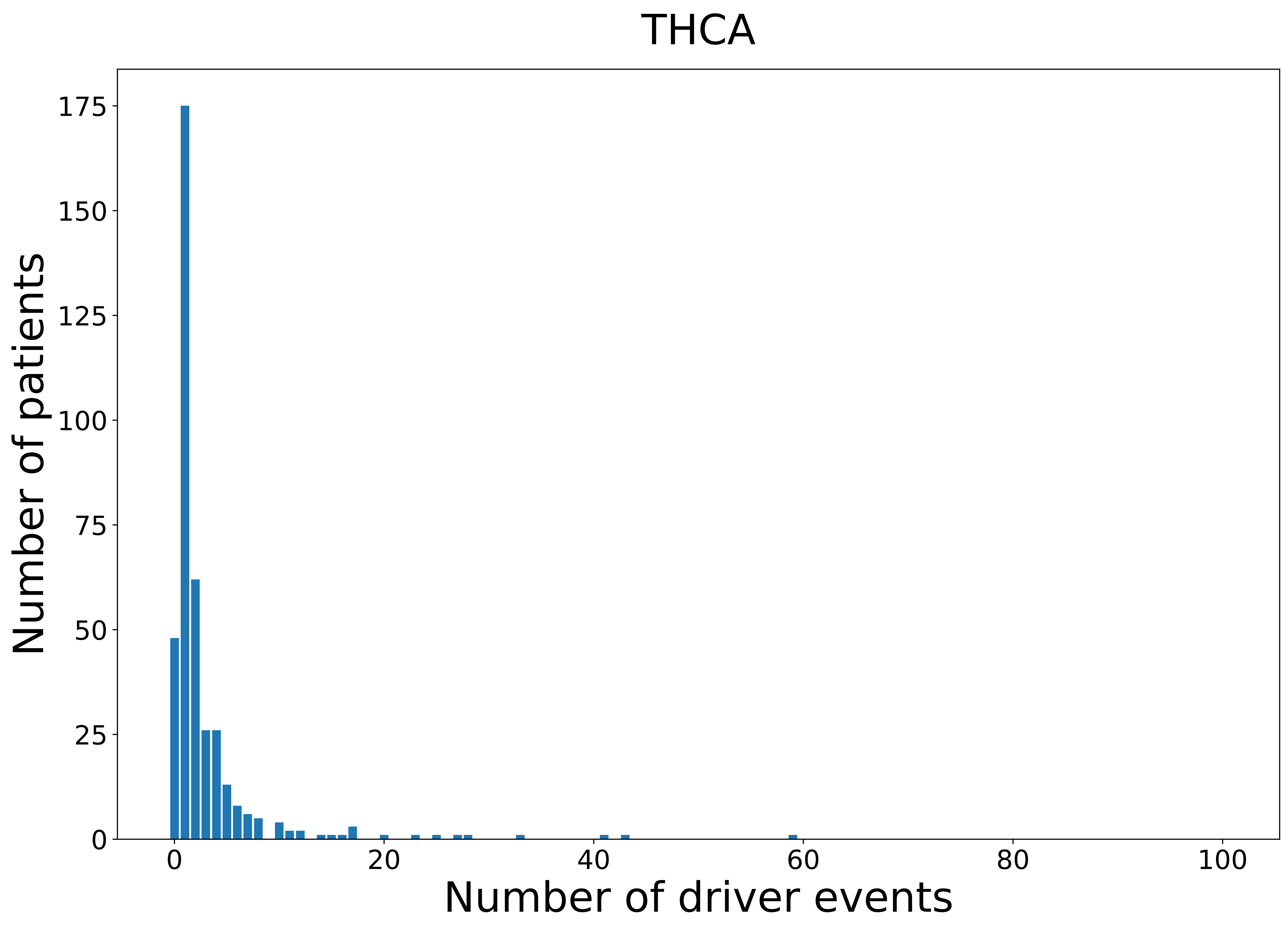

### 2021_8_16_20_32_THCA_FEMALE.pdf

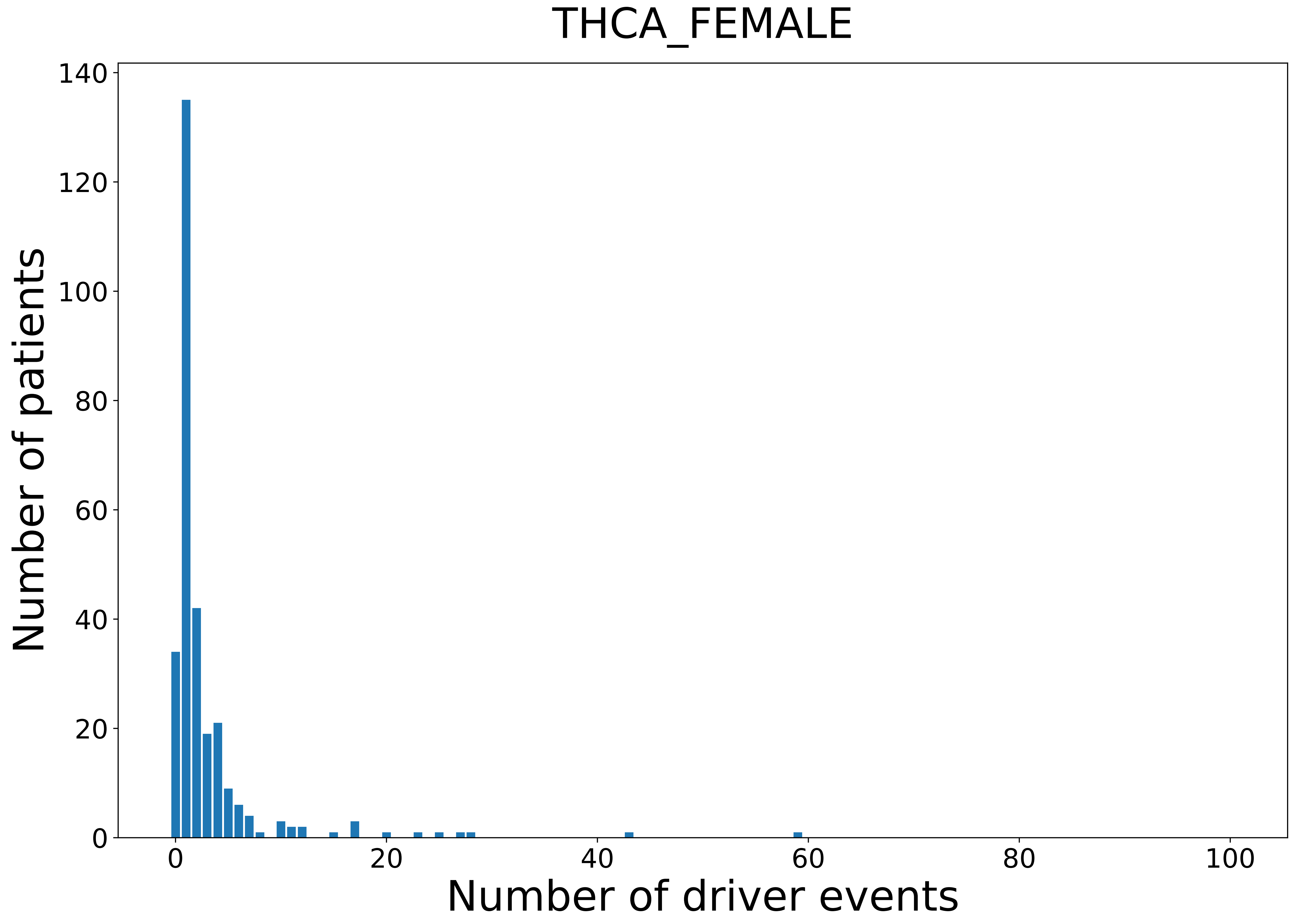

### 2021_8_16_20_32_THCA_MALE.pdf

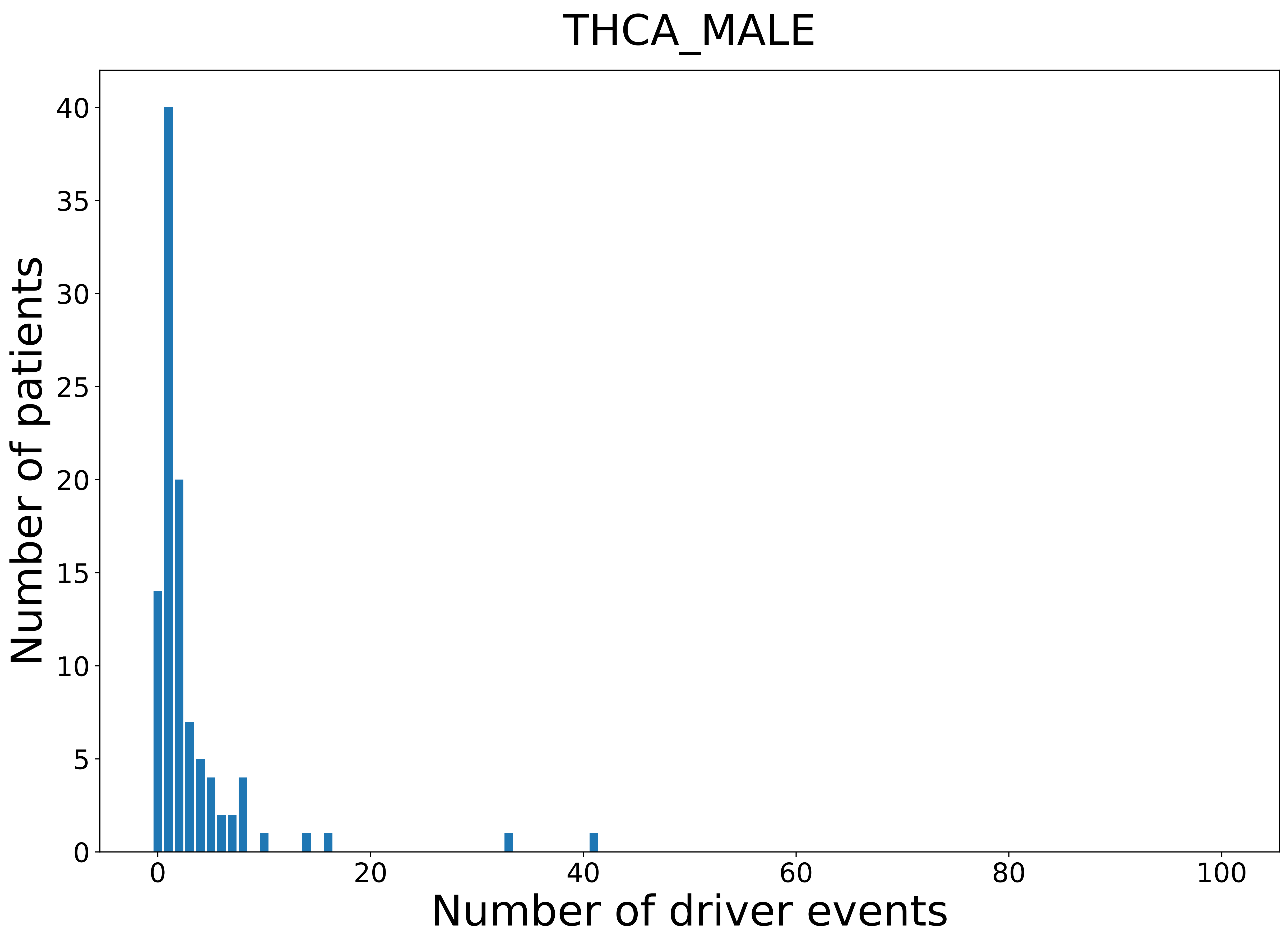

### 2021_8_16_20_32_THYM.pdf

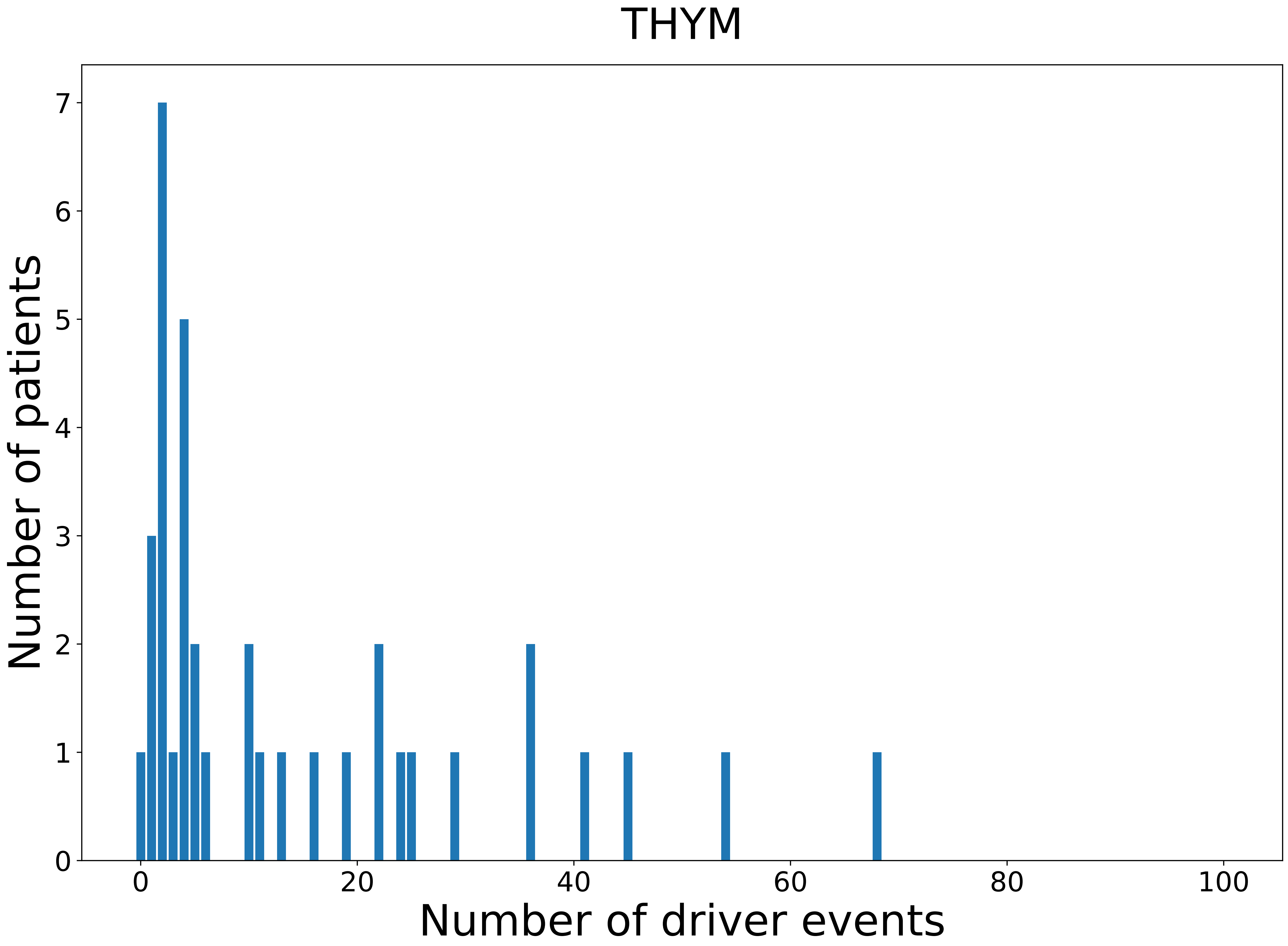

### 2021_8_16_20_32_THYM_FEMALE.pdf

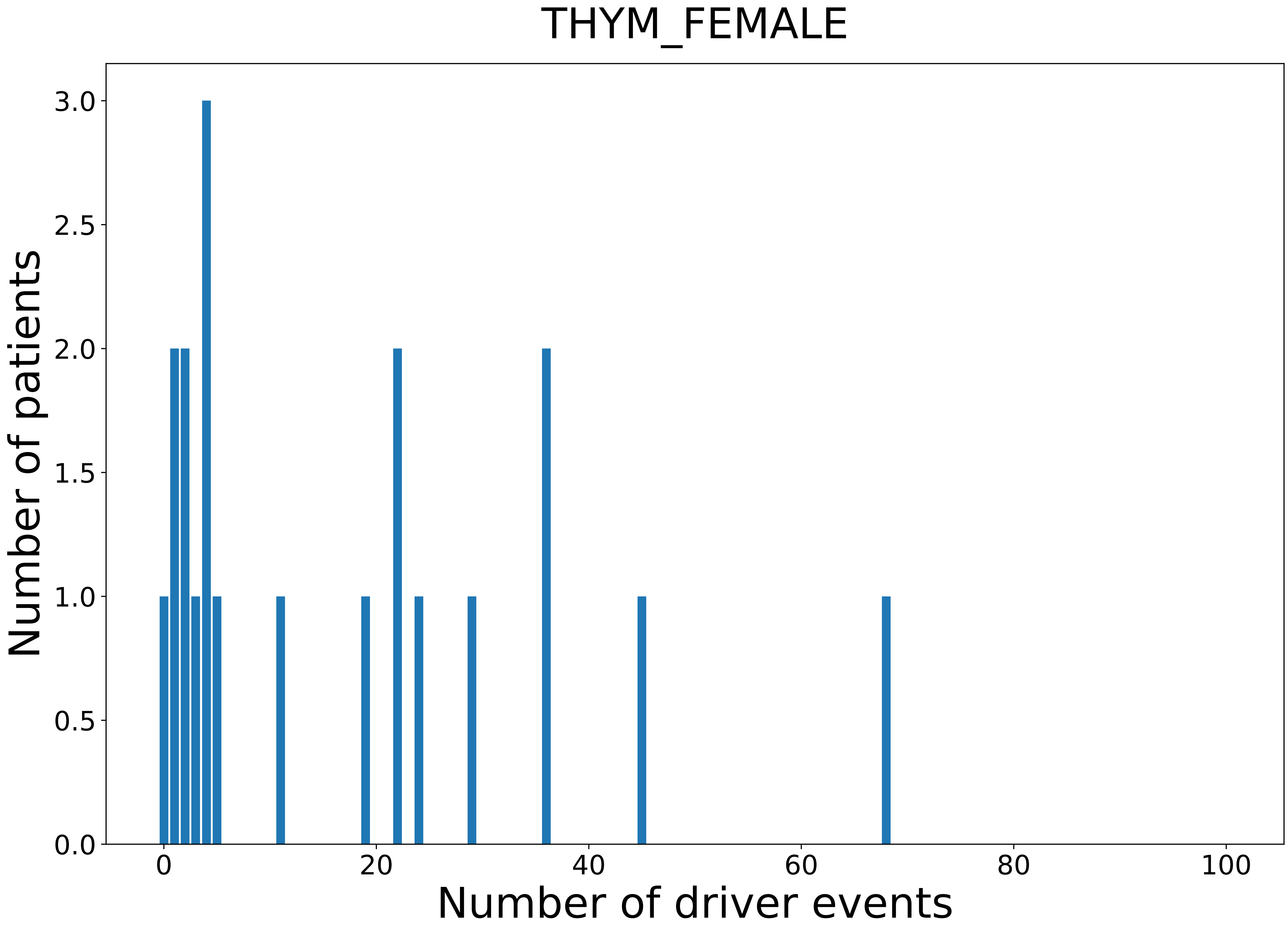

### 2021_8_16_20_32_THYM_MALE.pdf

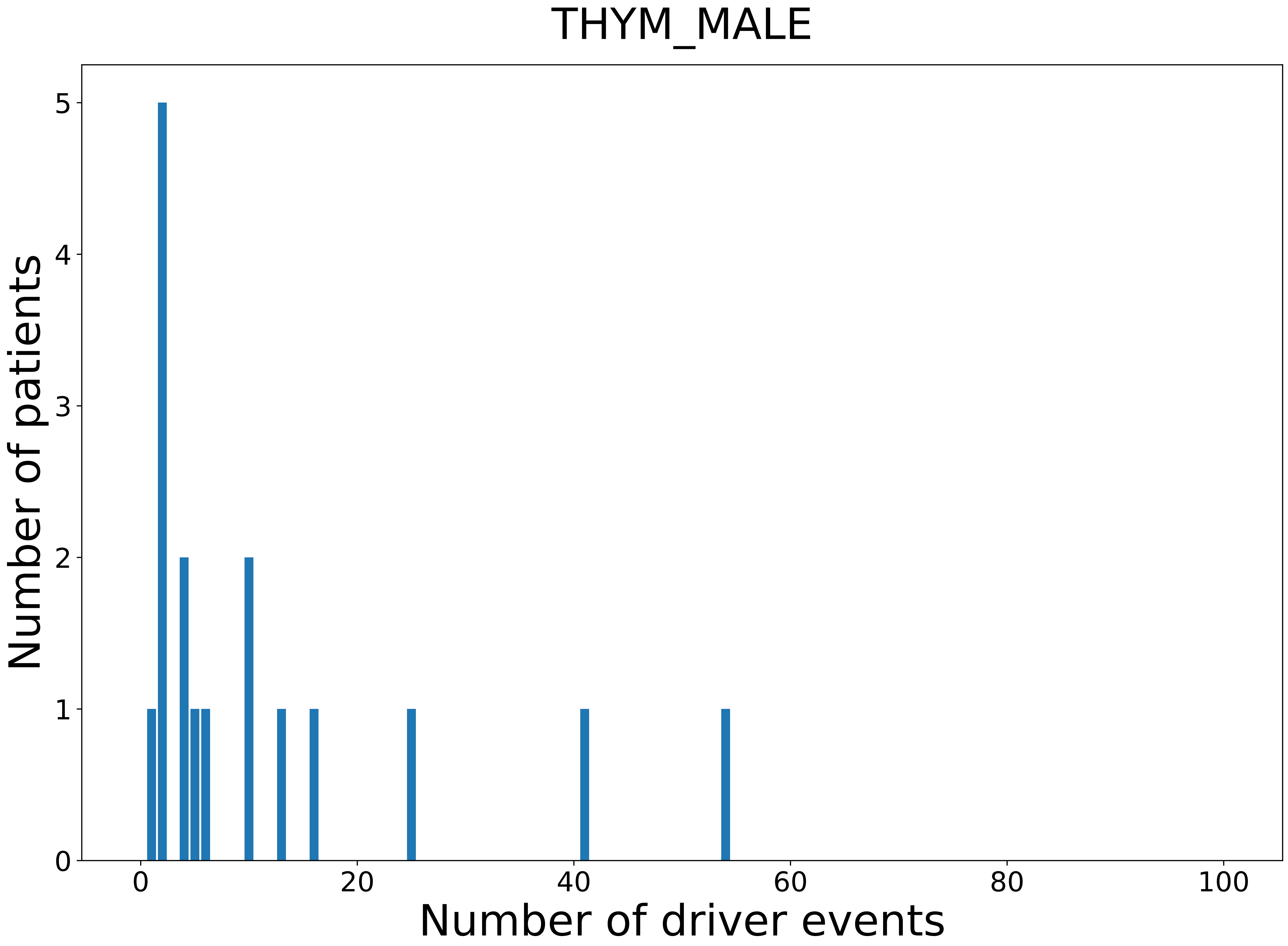

### 2021_8_16_20_32_UCEC.pdf

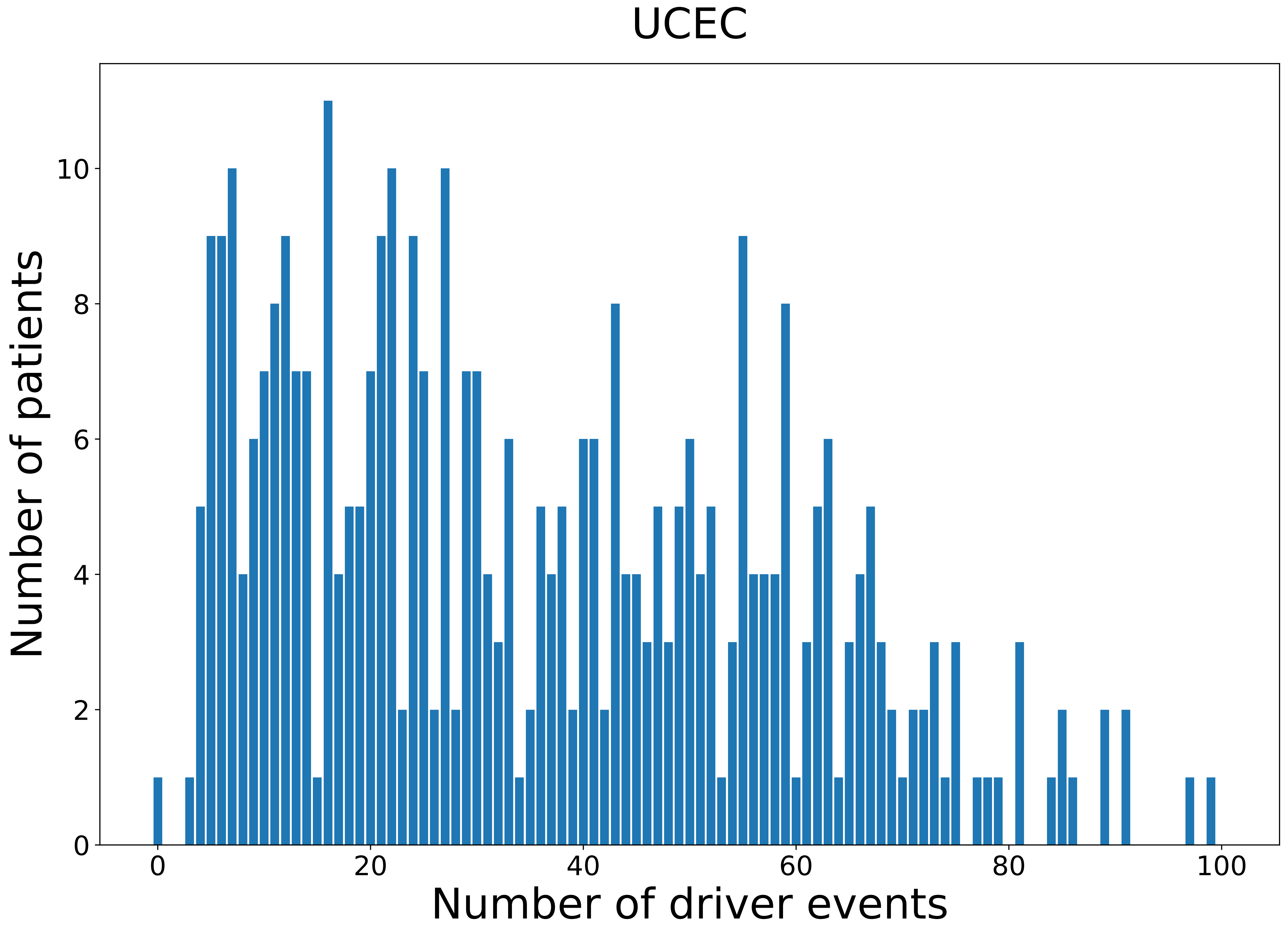

### 2021_8_16_20_32_UCEC_FEMALE.pdf

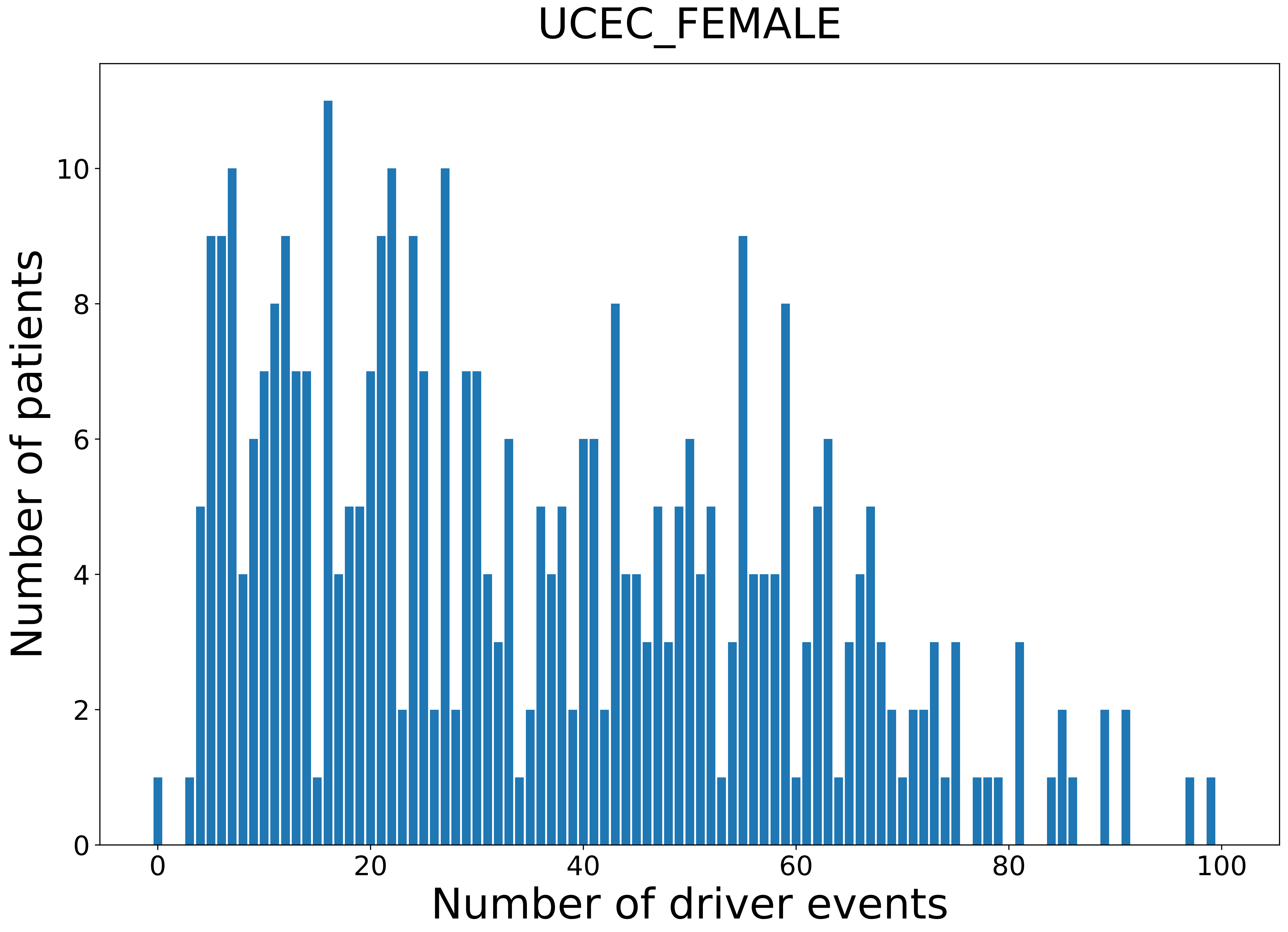

### 2021_8_16_20_32_UCS.pdf

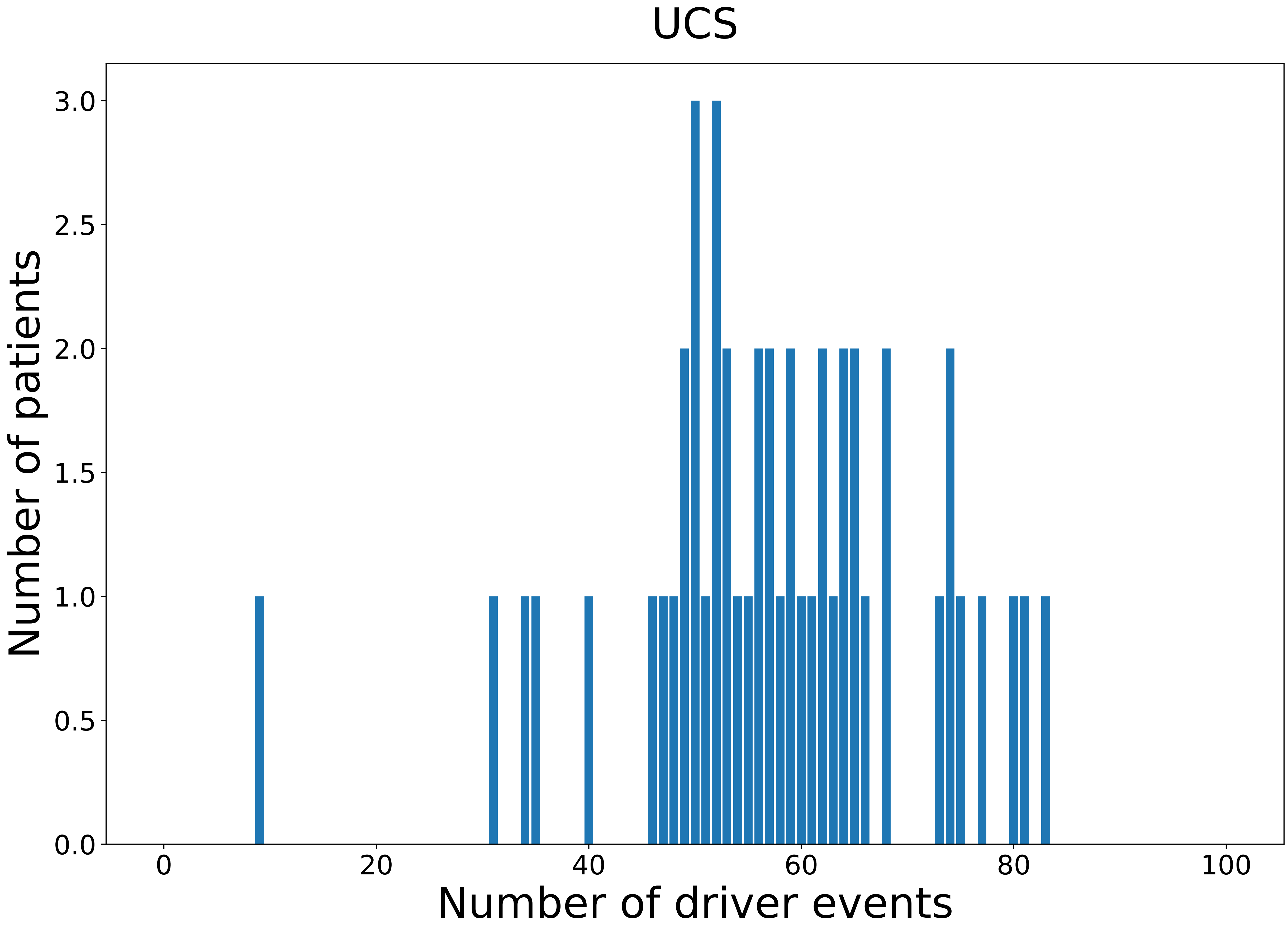

### 2021_8_16_20_32_UCS_FEMALE.pdf

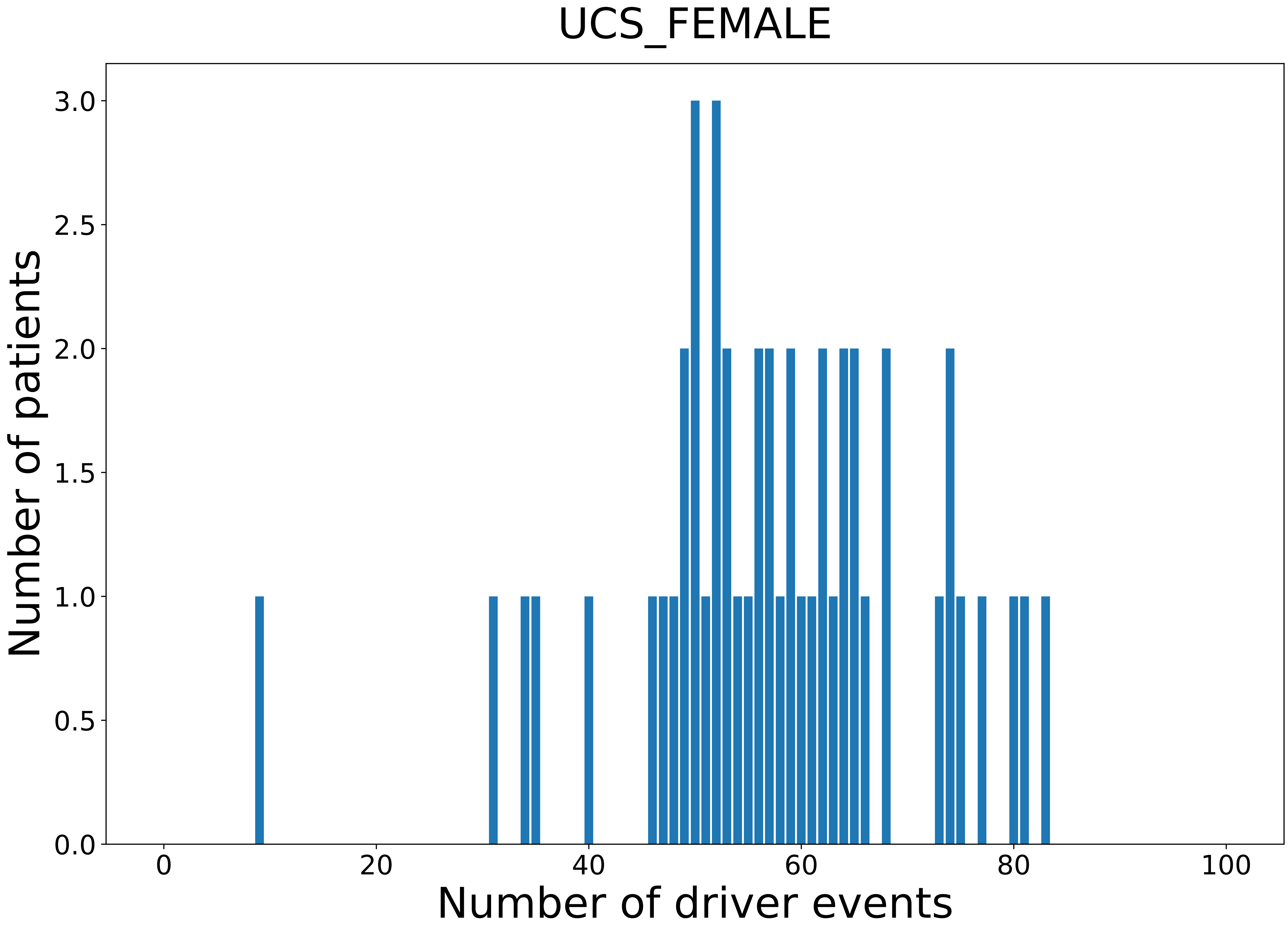

### 2021_8_16_20_32_UVM.pdf

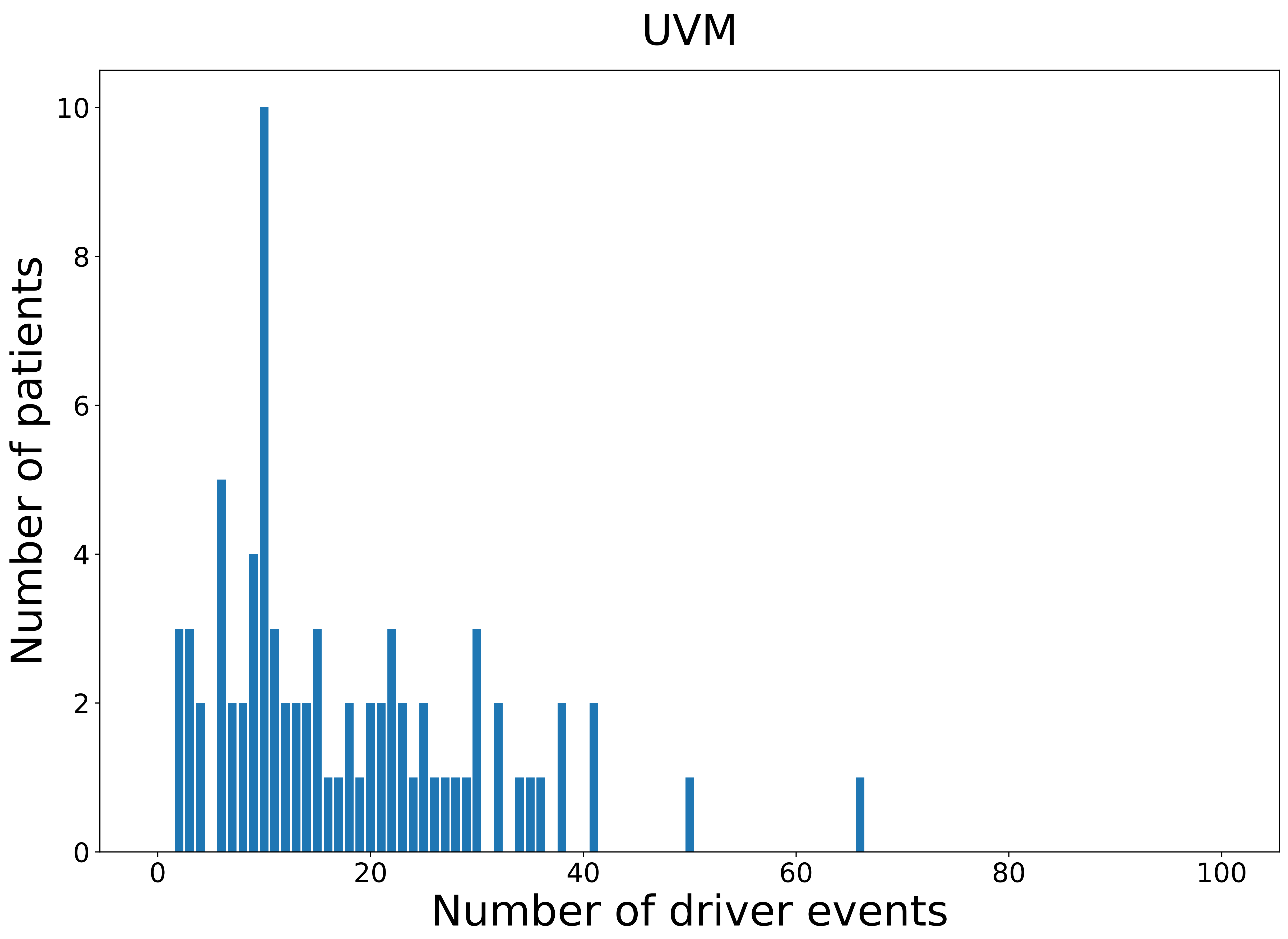

### 2021_8_16_20_32_UVM_FEMALE.pdf

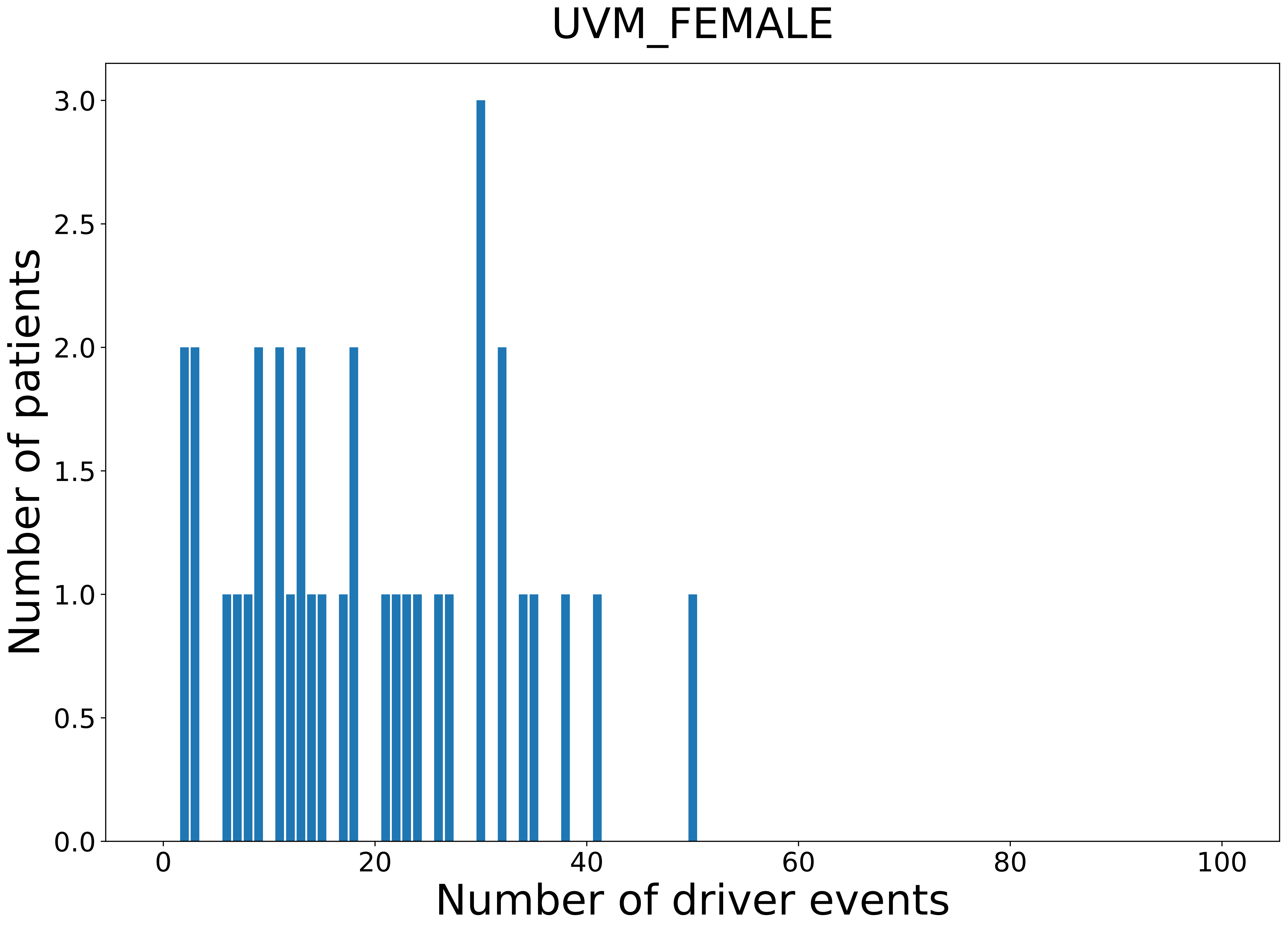

### 2021_8_16_20_32_UVM_MALE.pdf

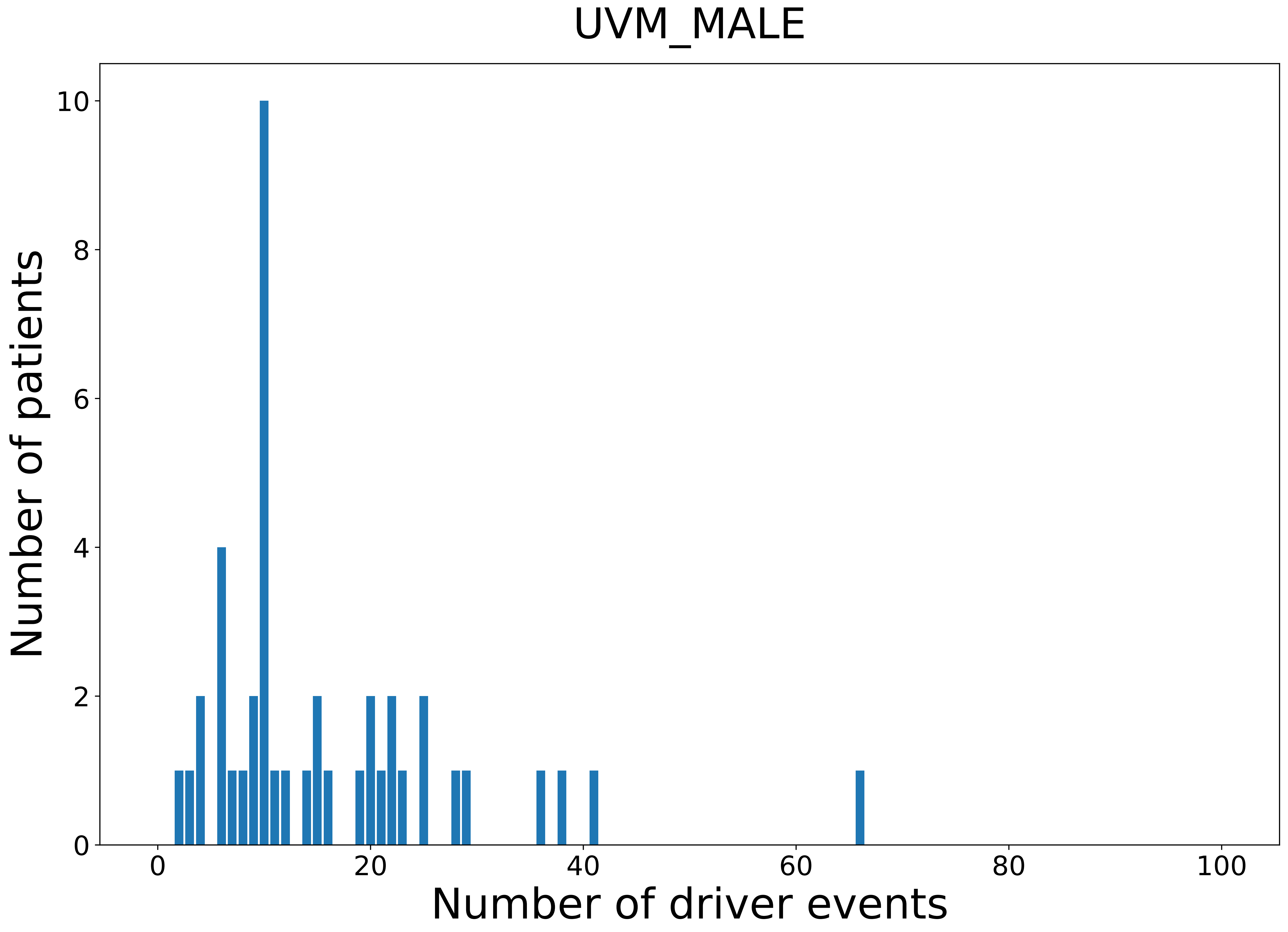

### legeng.png

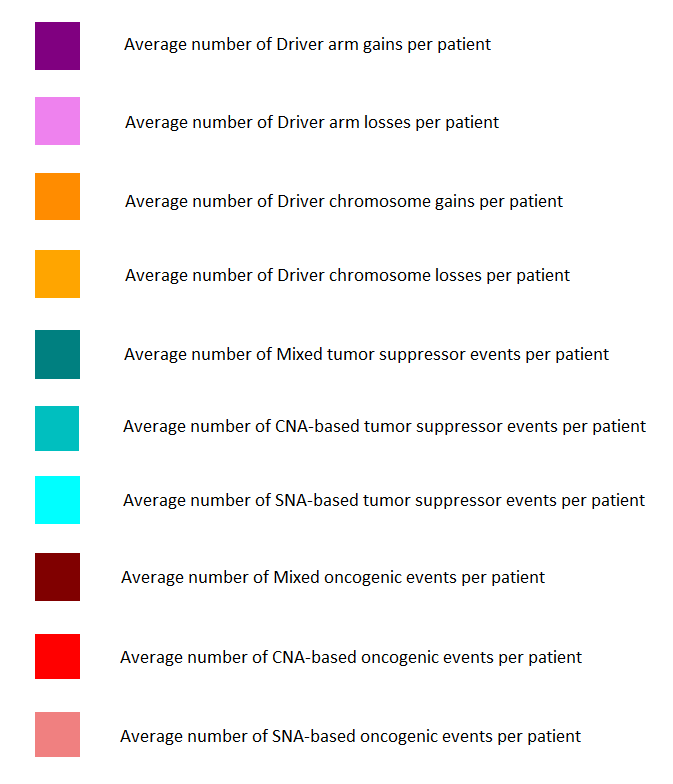
