## Supplementary figures and images for "Novel Driver Strength Index highlights important cancer genes in TCGA PanCanAtlas patients"

### 2021_8_16_14_9_distribution_events_detailed_28.pdf

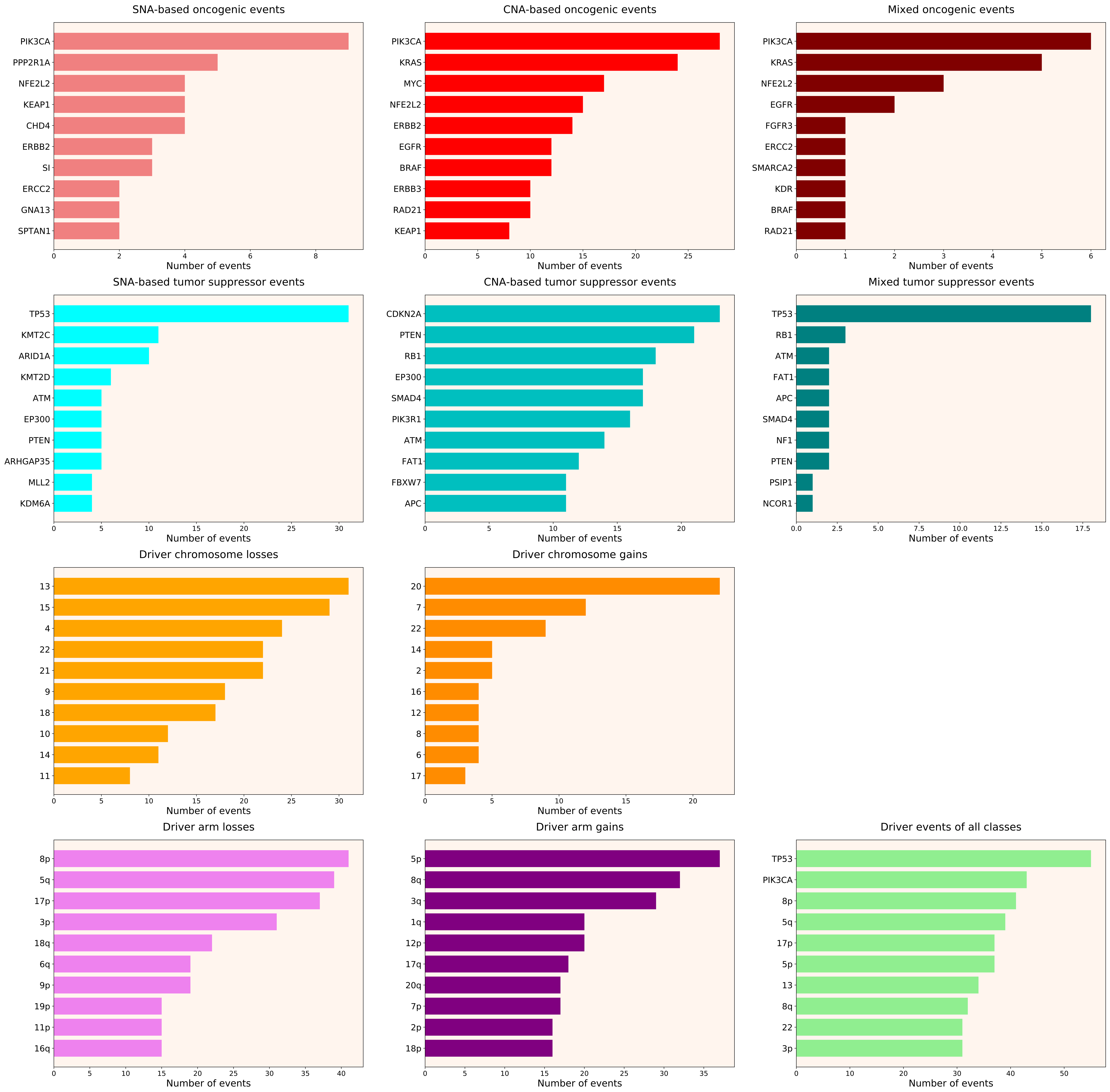

### 2021_8_16_14_9_distribution_events_detailed_females_8.pdf

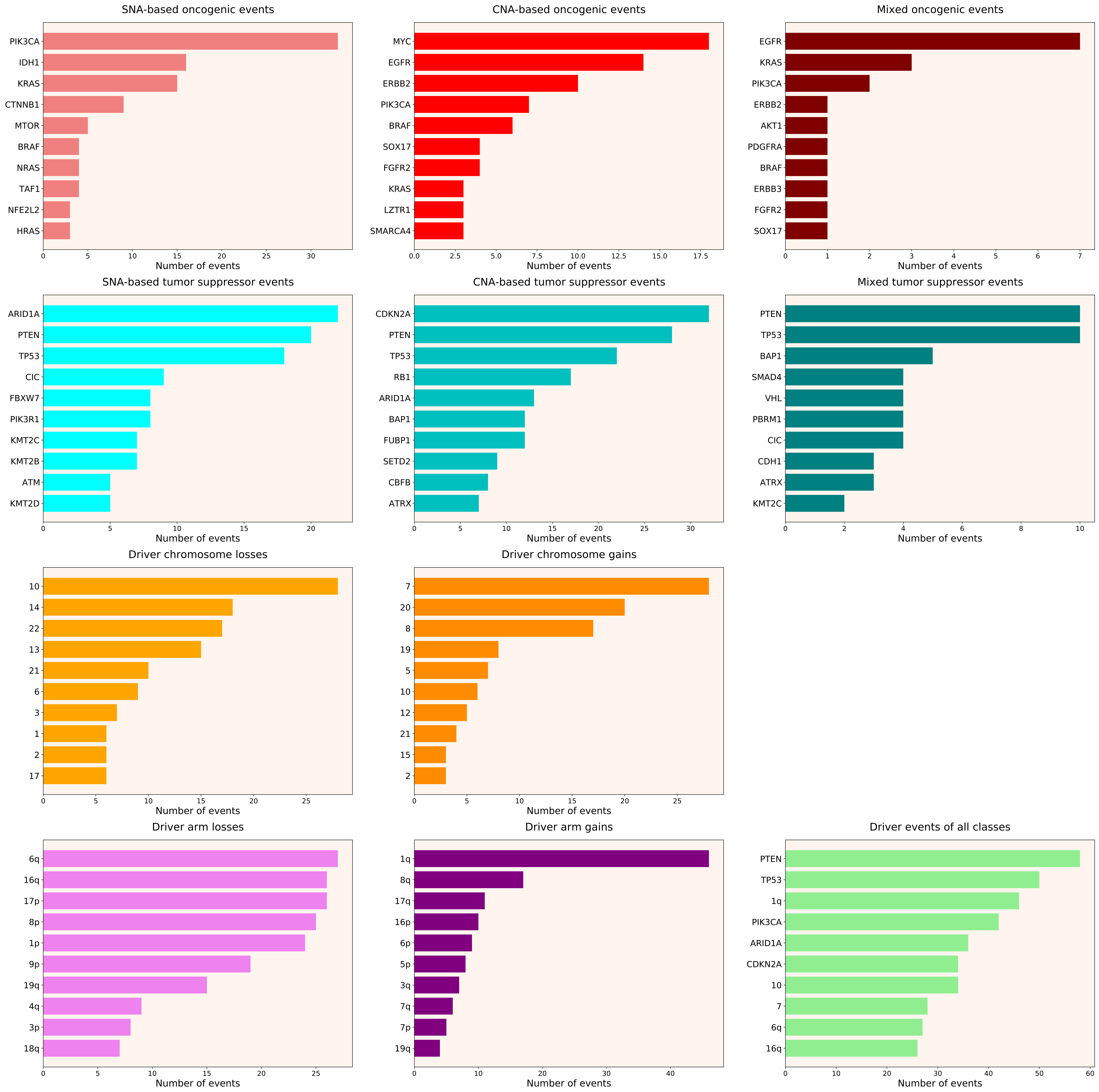

### 2021_8_16_14_9_distribution_events_detailed_females_25.pdf

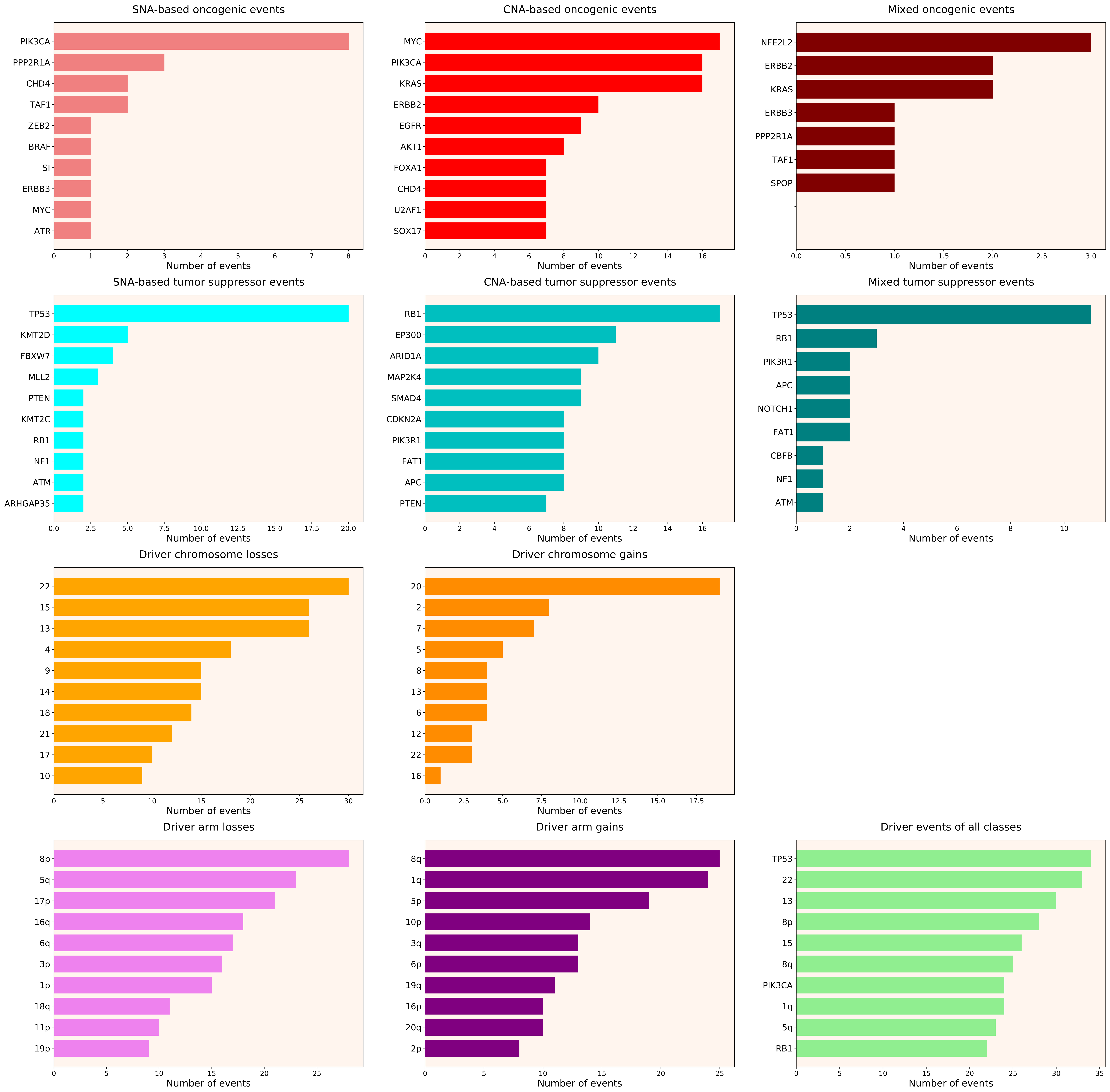

### 2021_8_16_14_9_distribution_events_detailed_males_21.pdf

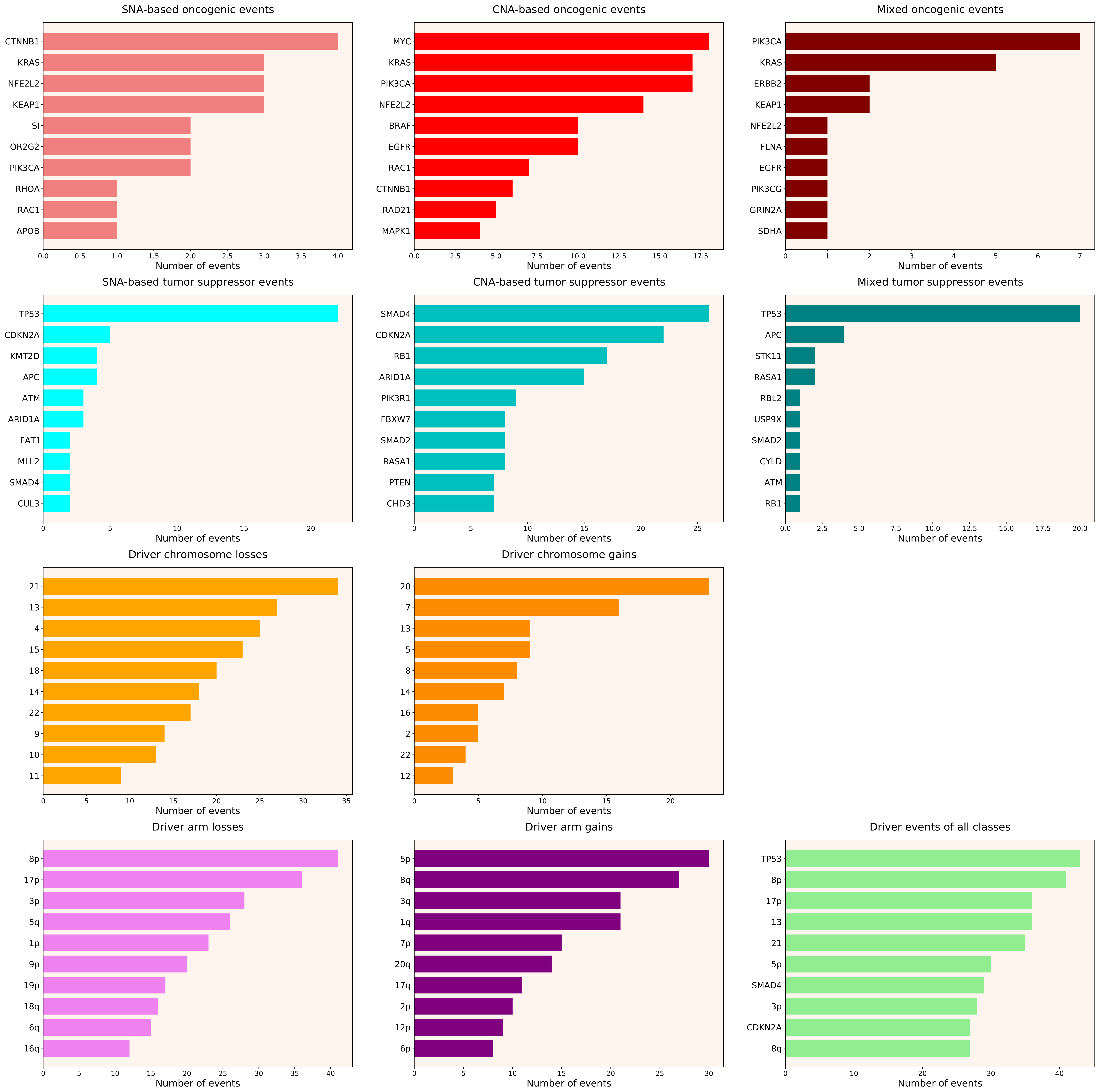

### 2021_8_16_14_9_distribution_events_detailed_males_35.pdf

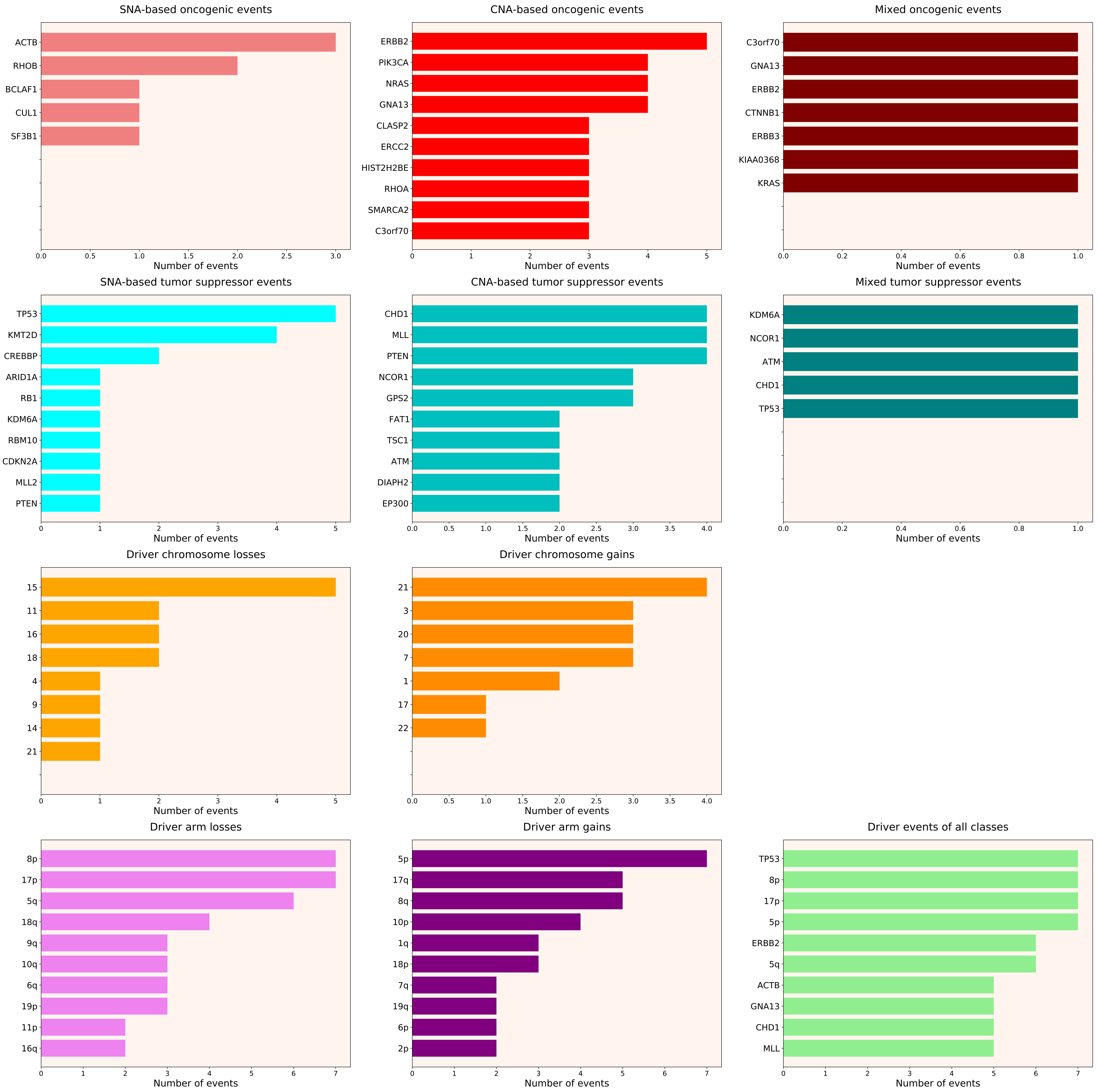
