## Supplementary figures and images for "Novel Driver Strength Index highlights important cancer genes in TCGA PanCanAtlas patients"

### 2021_8_16_14_9_ACC.pdf

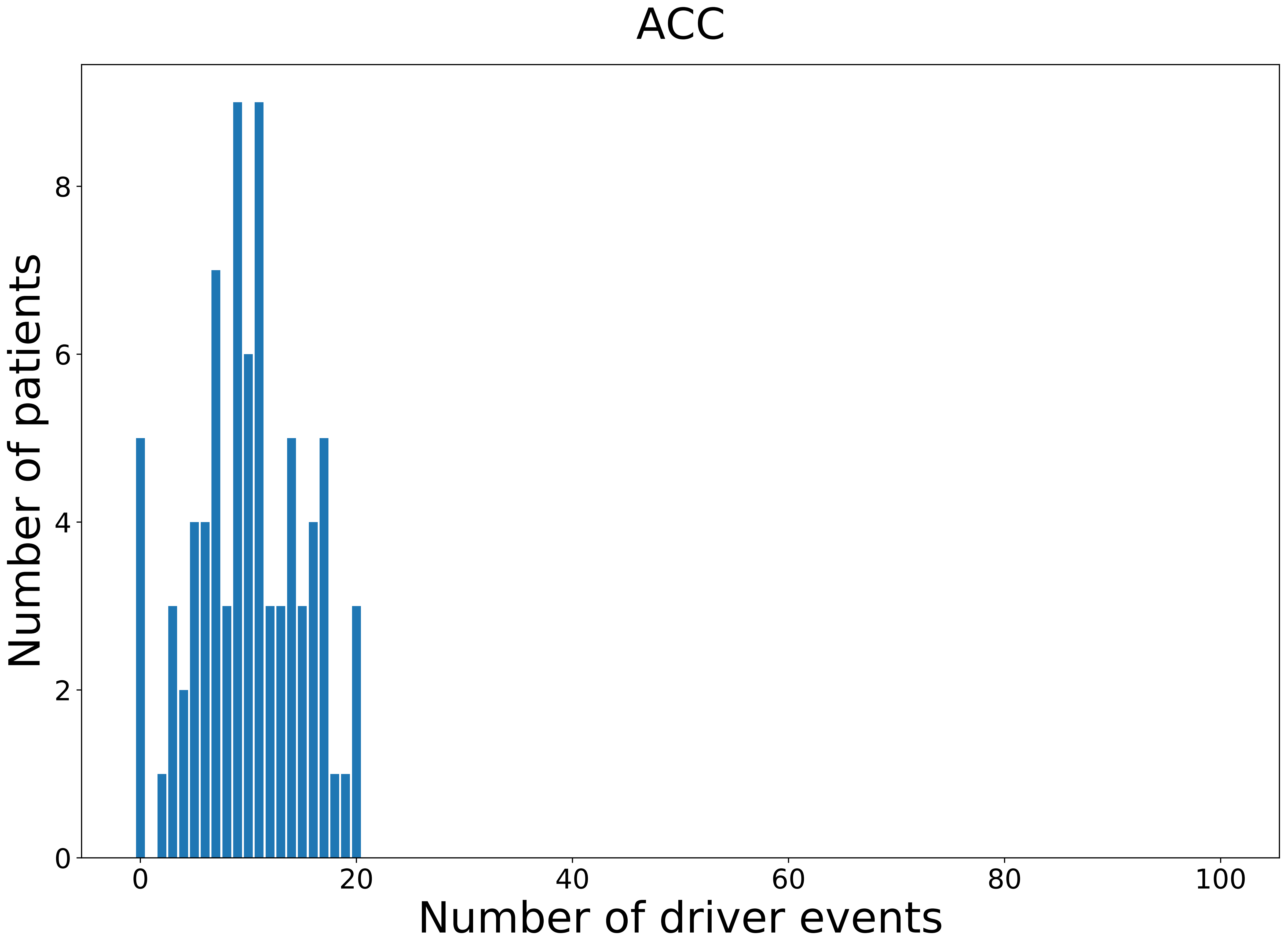

### 2021_8_16_14_9_ACC_FEMALE.pdf

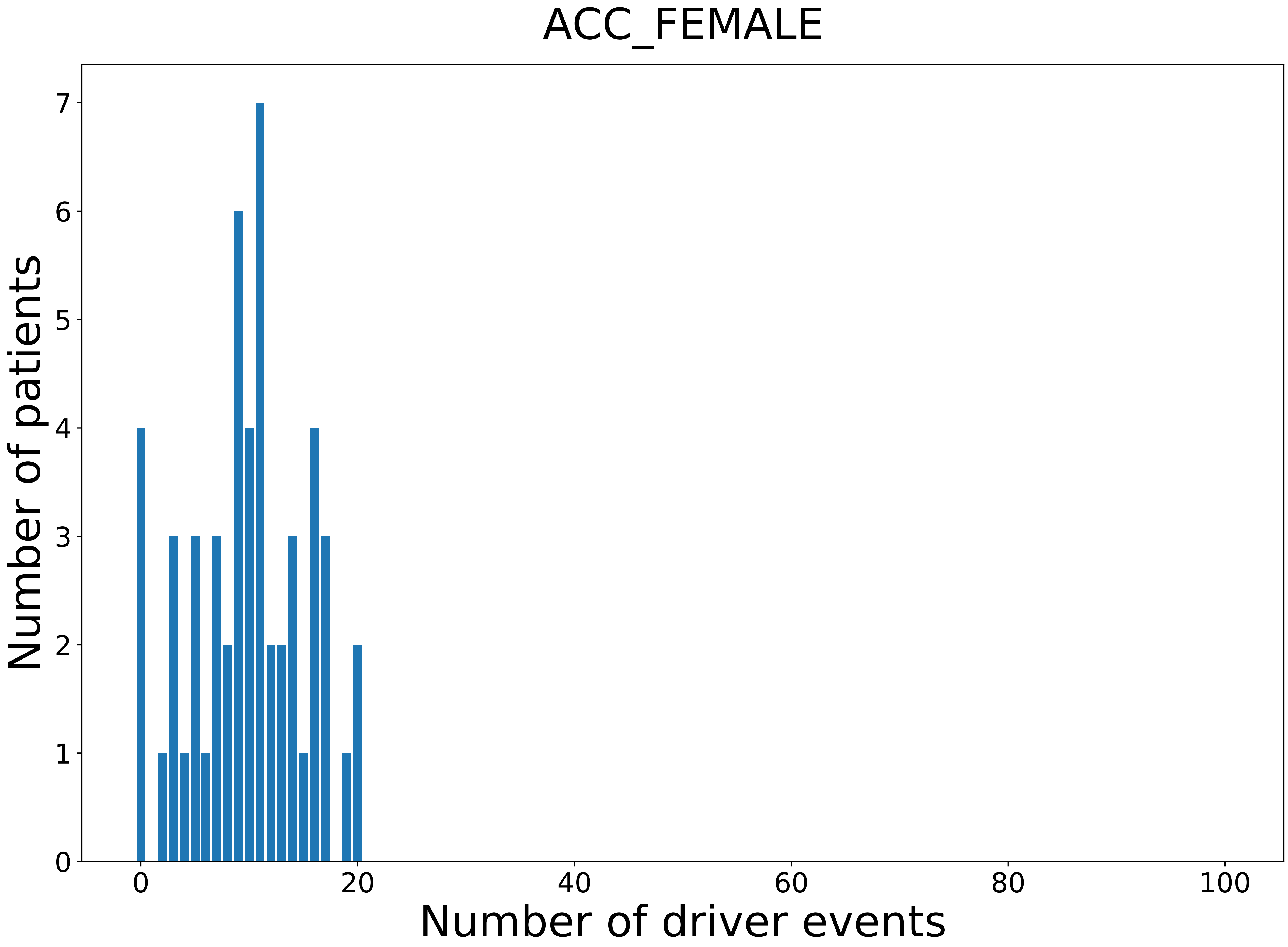

### 2021_8_16_14_9_ACC_MALE.pdf

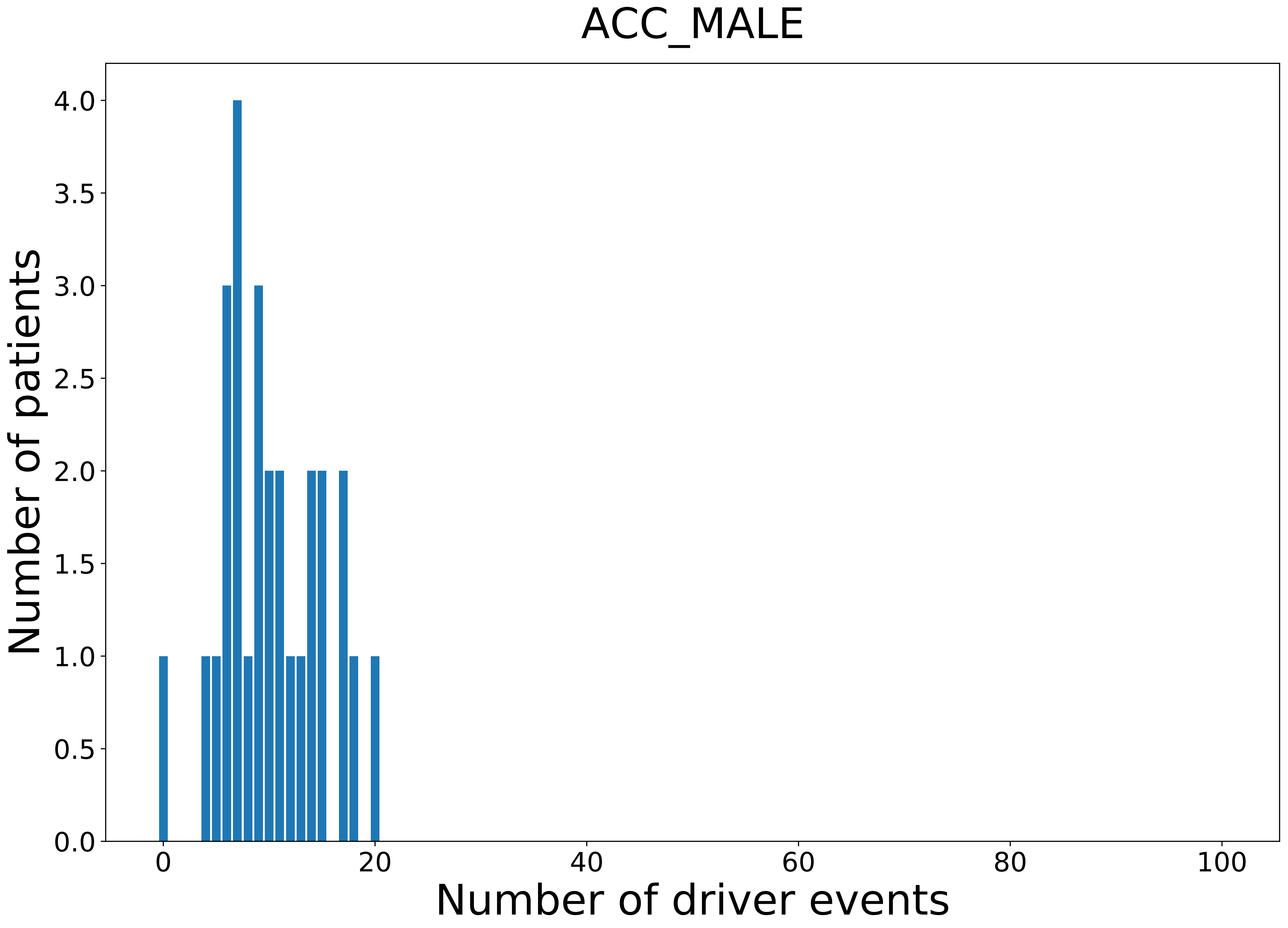

### 2021_8_16_14_9_BLCA.pdf

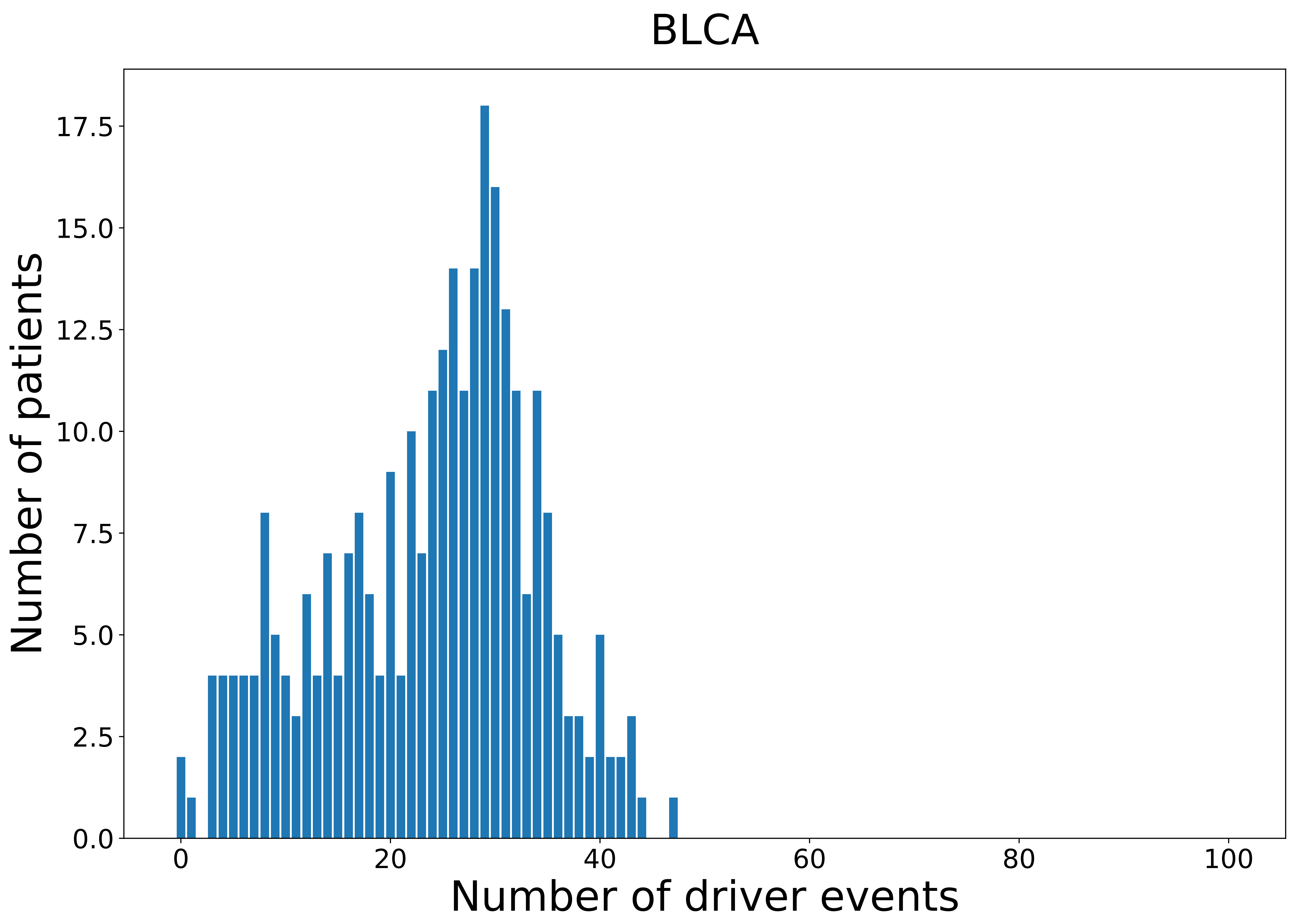

### 2021_8_16_14_9_BLCA_MALE.pdf

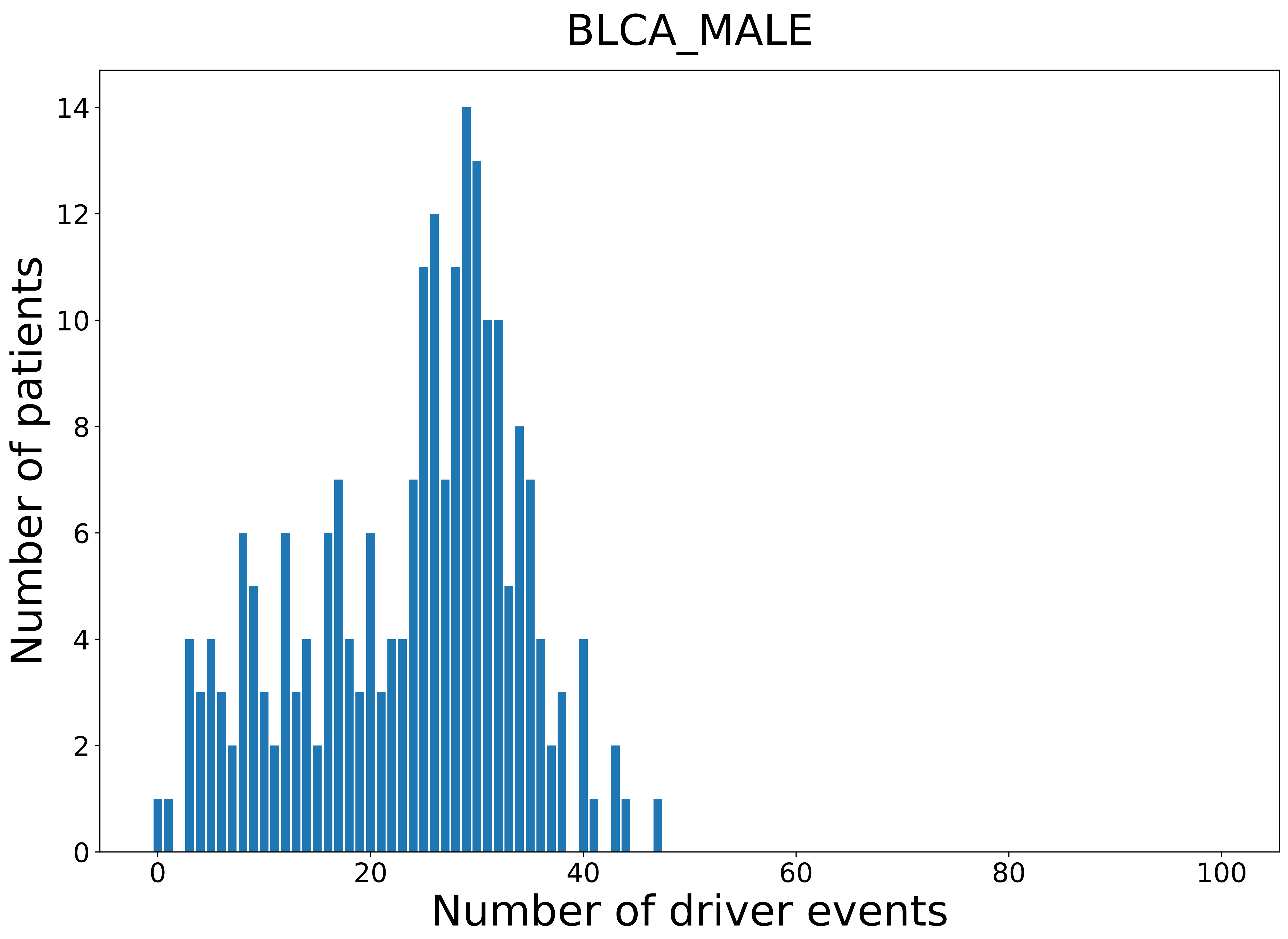

### 2021_8_16_14_9_CESC.pdf

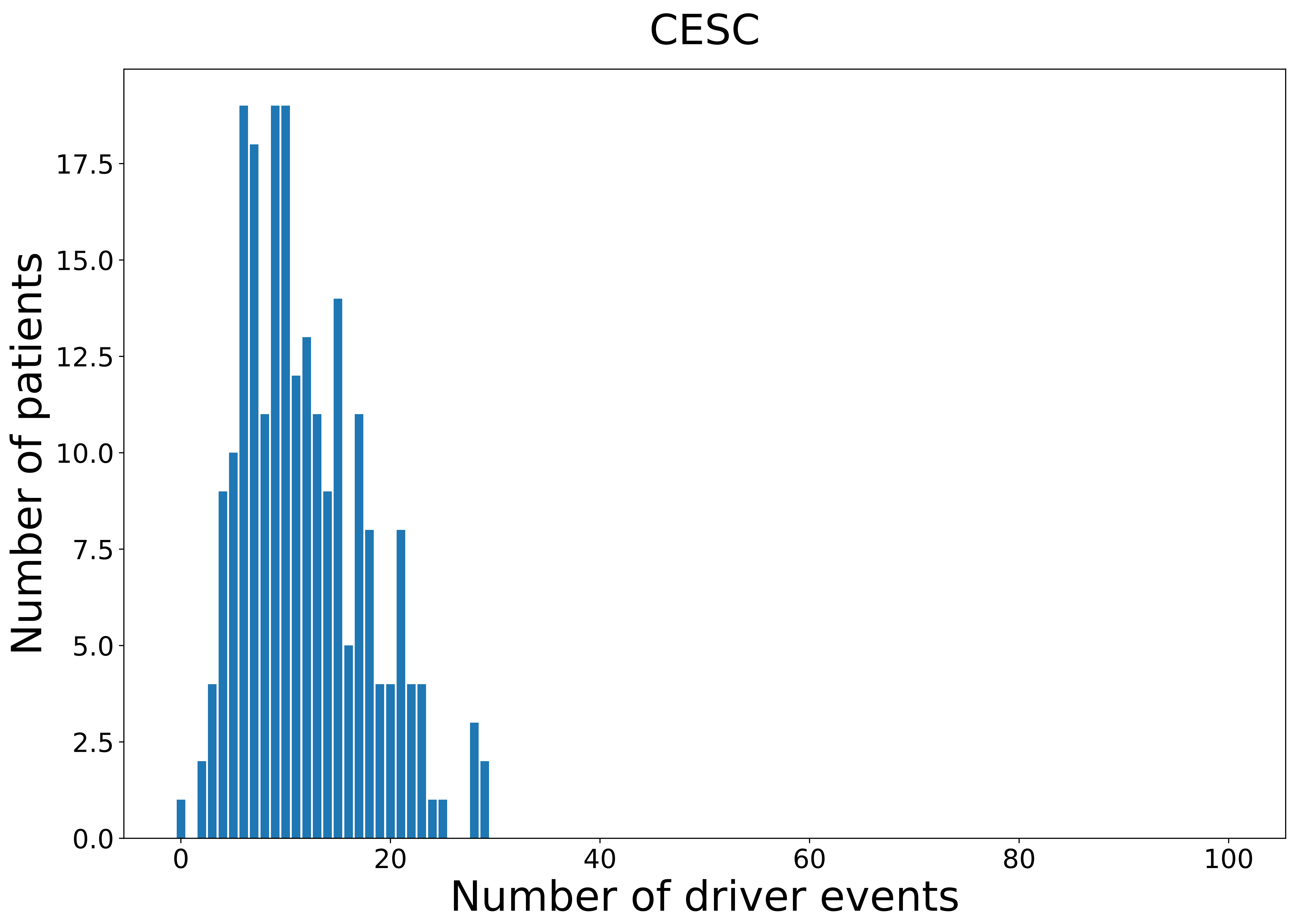

### 2021_8_16_14_9_CESC_FEMALE.pdf

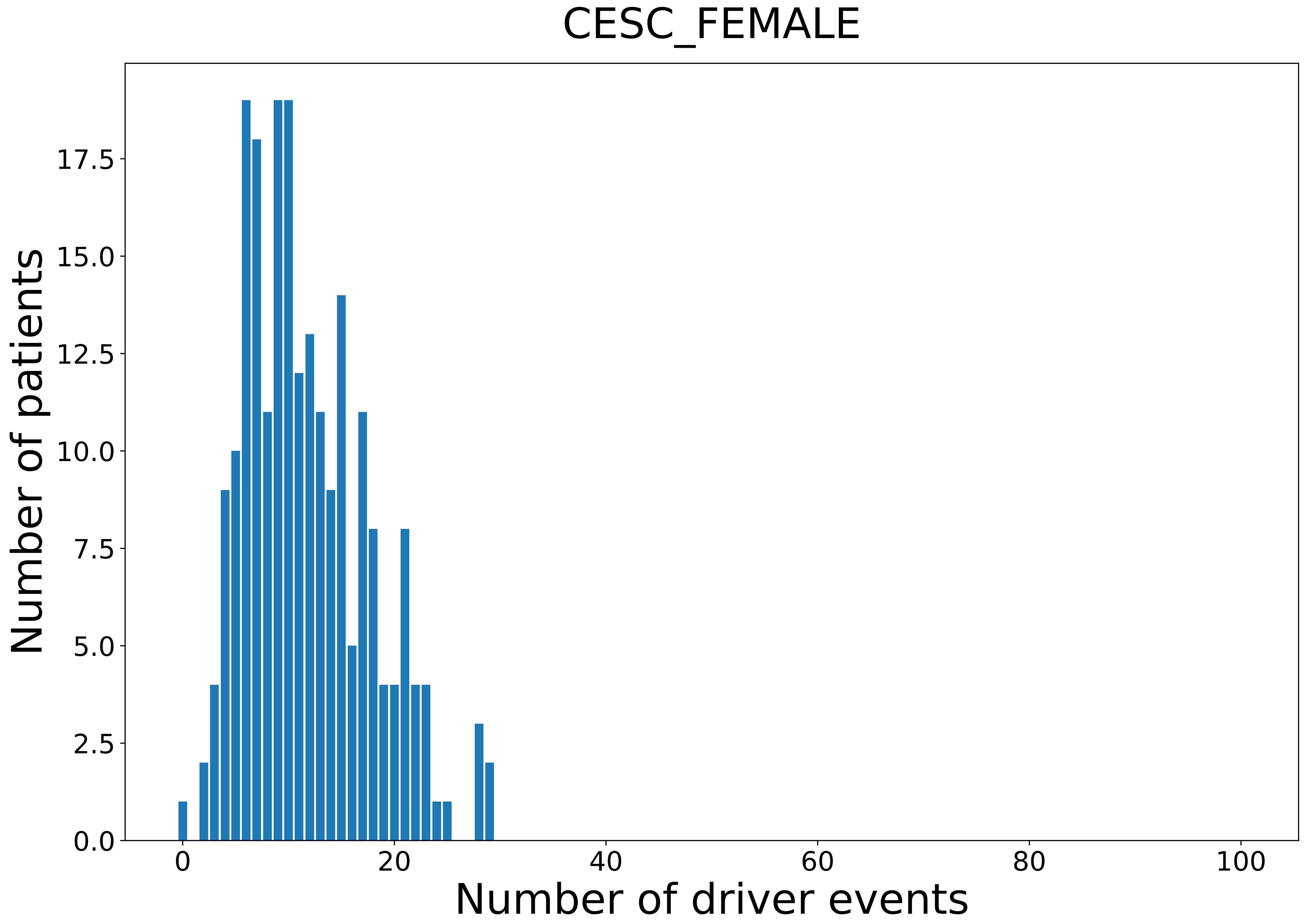

### 2021_8_16_14_9_CHOL_MALE.pdf

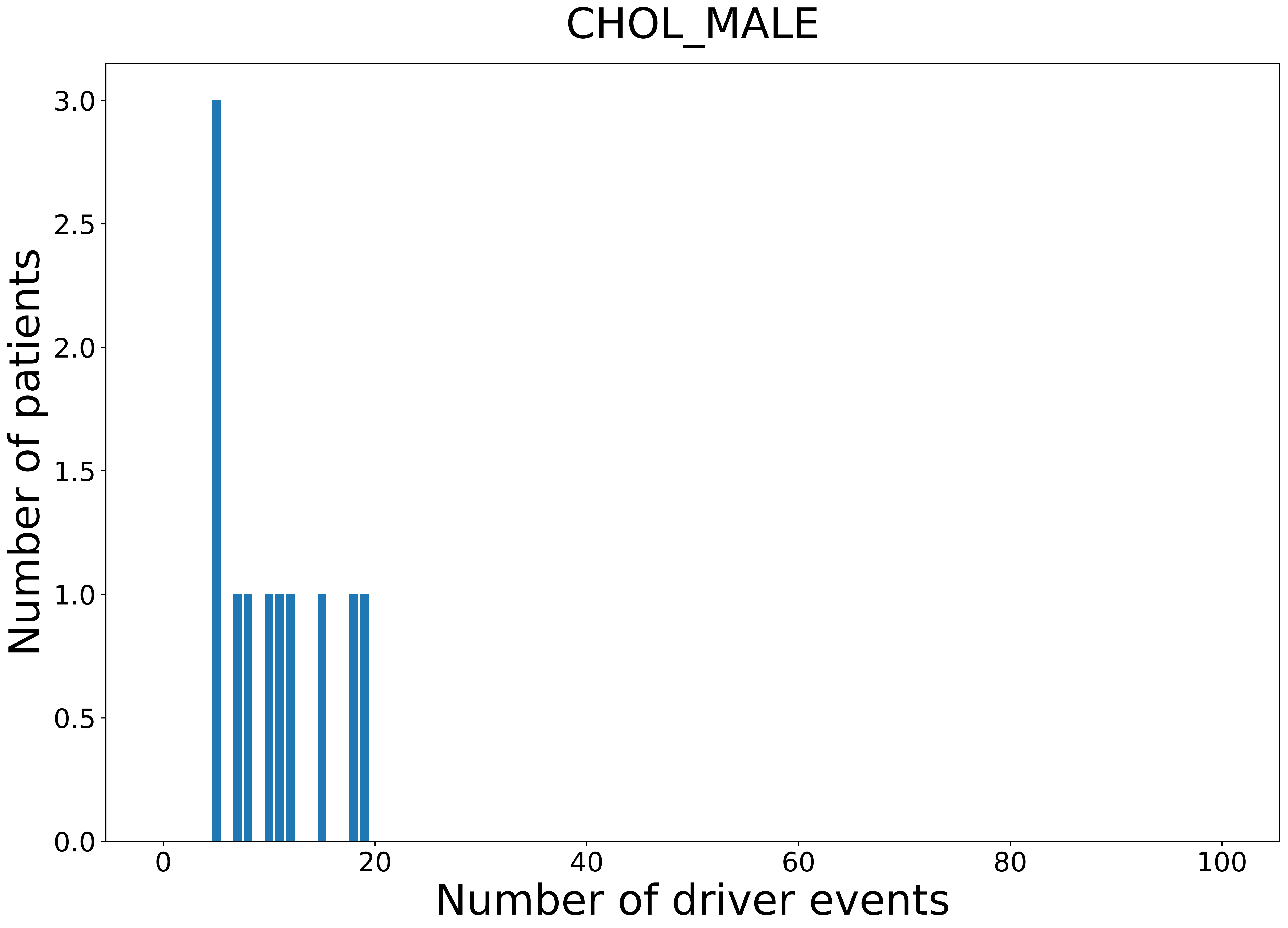

### 2021_8_16_14_9_COAD_FEMALE.pdf

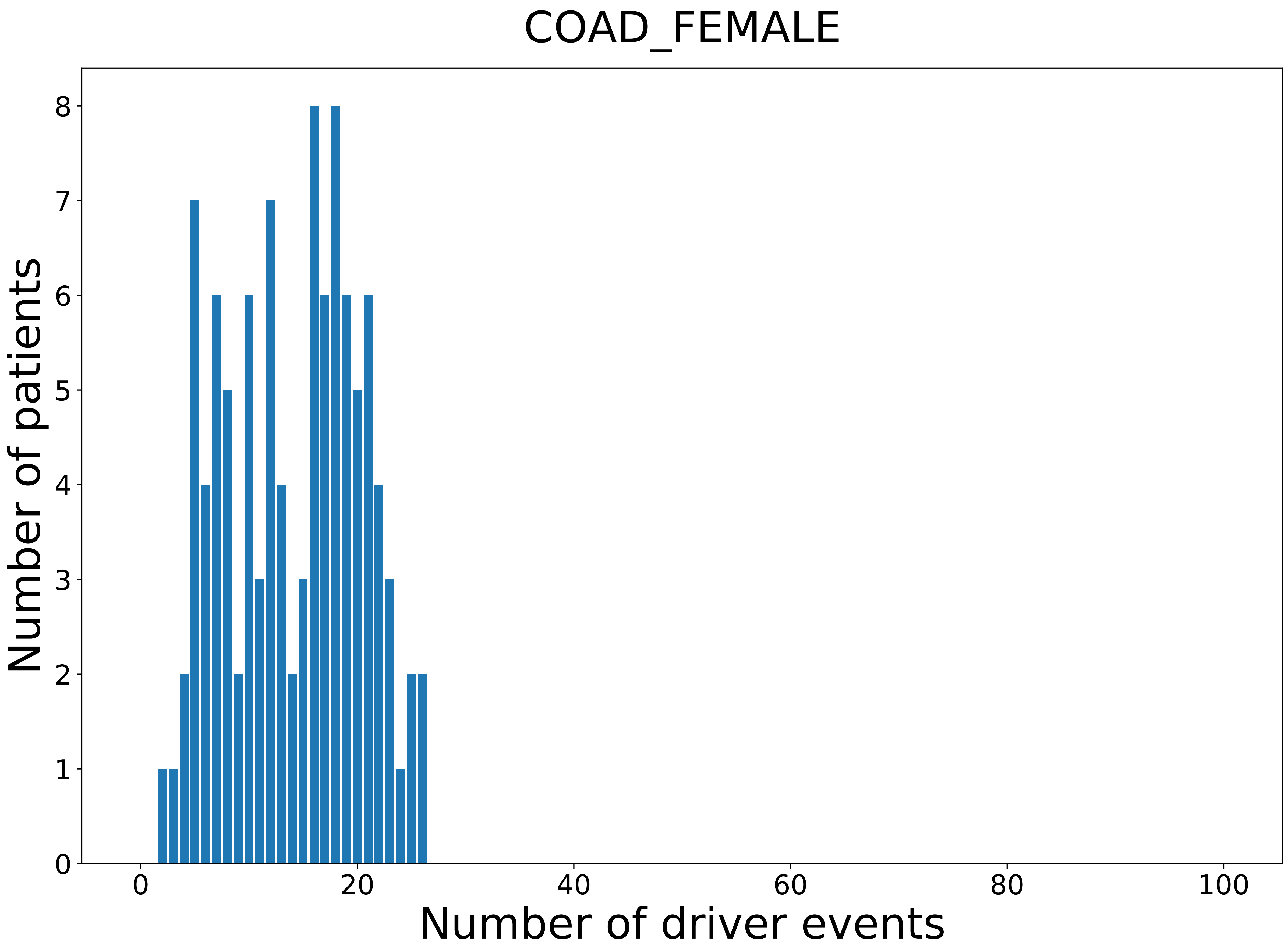

### 2021_8_16_14_9_COAD_MALE.pdf

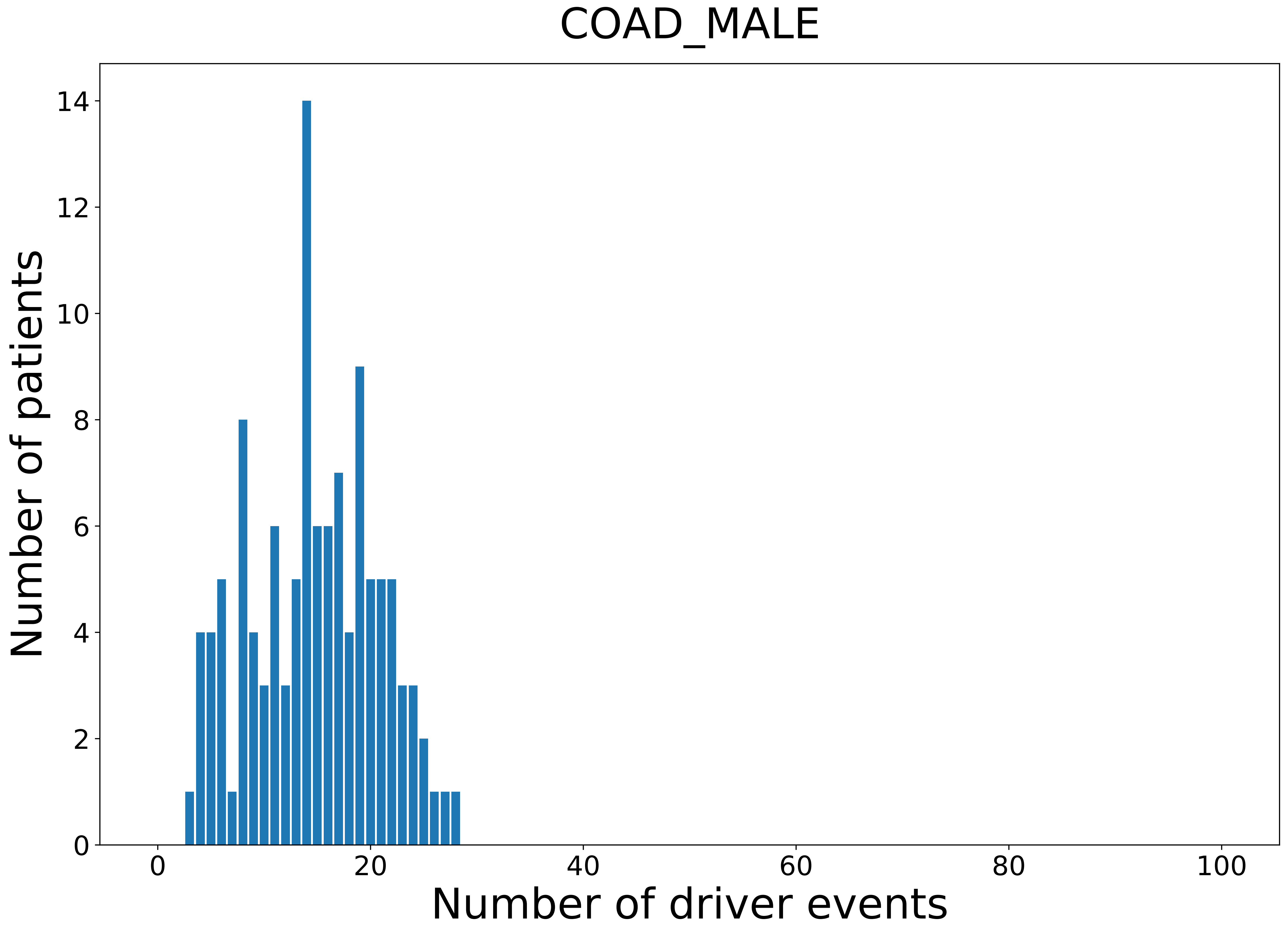

### 2021_8_16_14_9_distribution_events_detailed_10.pdf

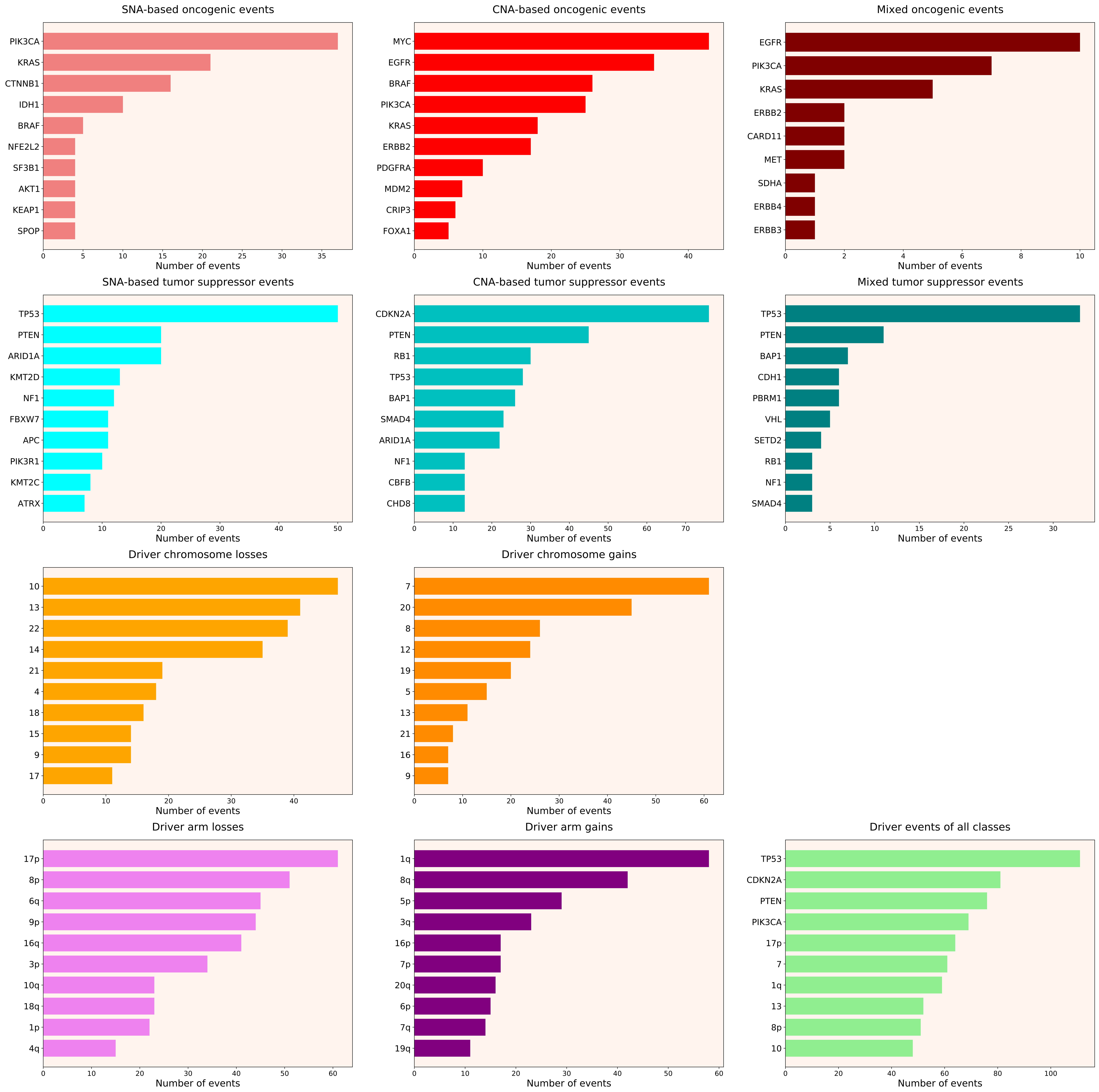

### 2021_8_16_14_9_distribution_events_detailed_11.pdf

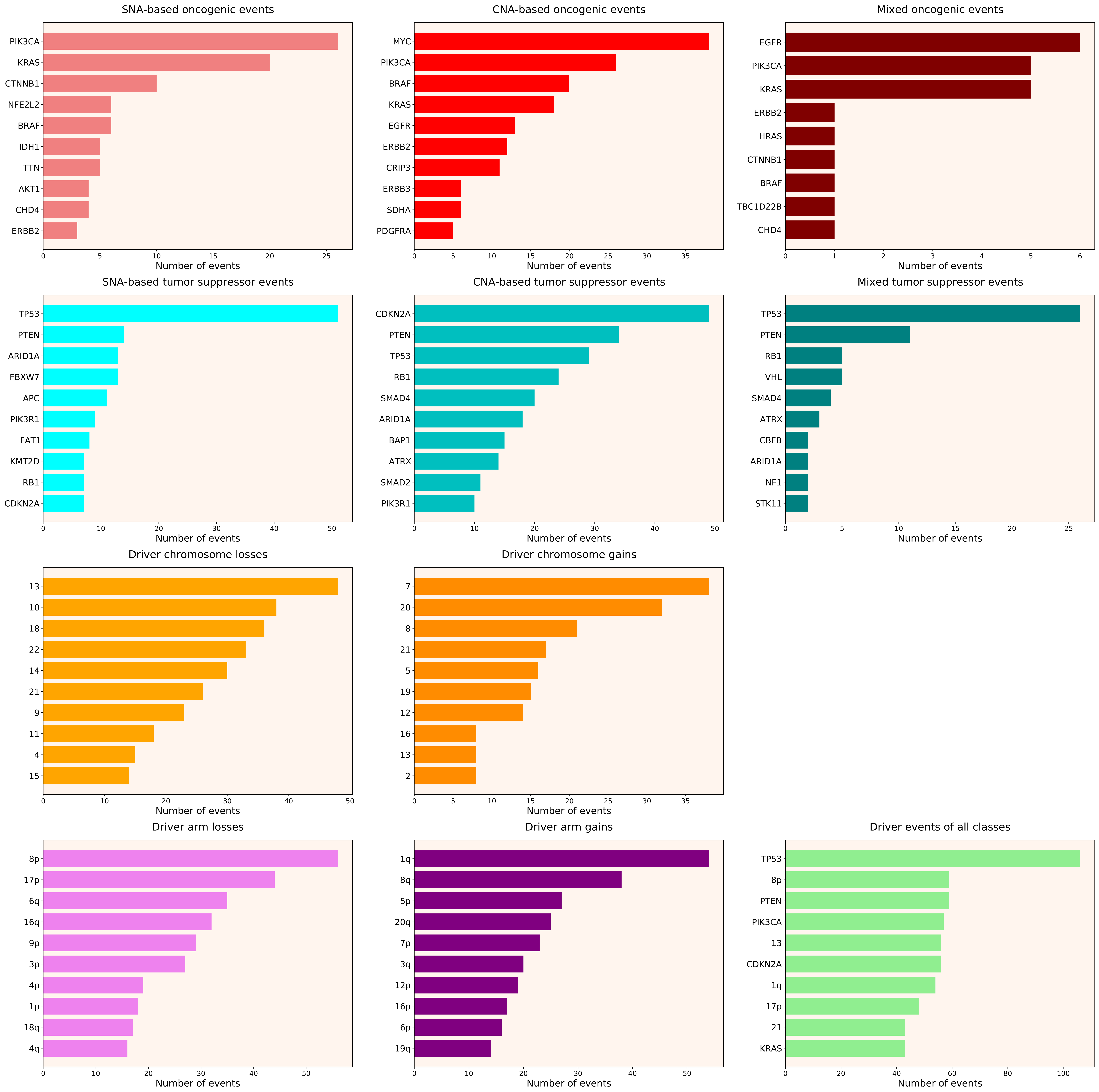

### 2021_8_16_14_9_distribution_events_detailed_12.pdf

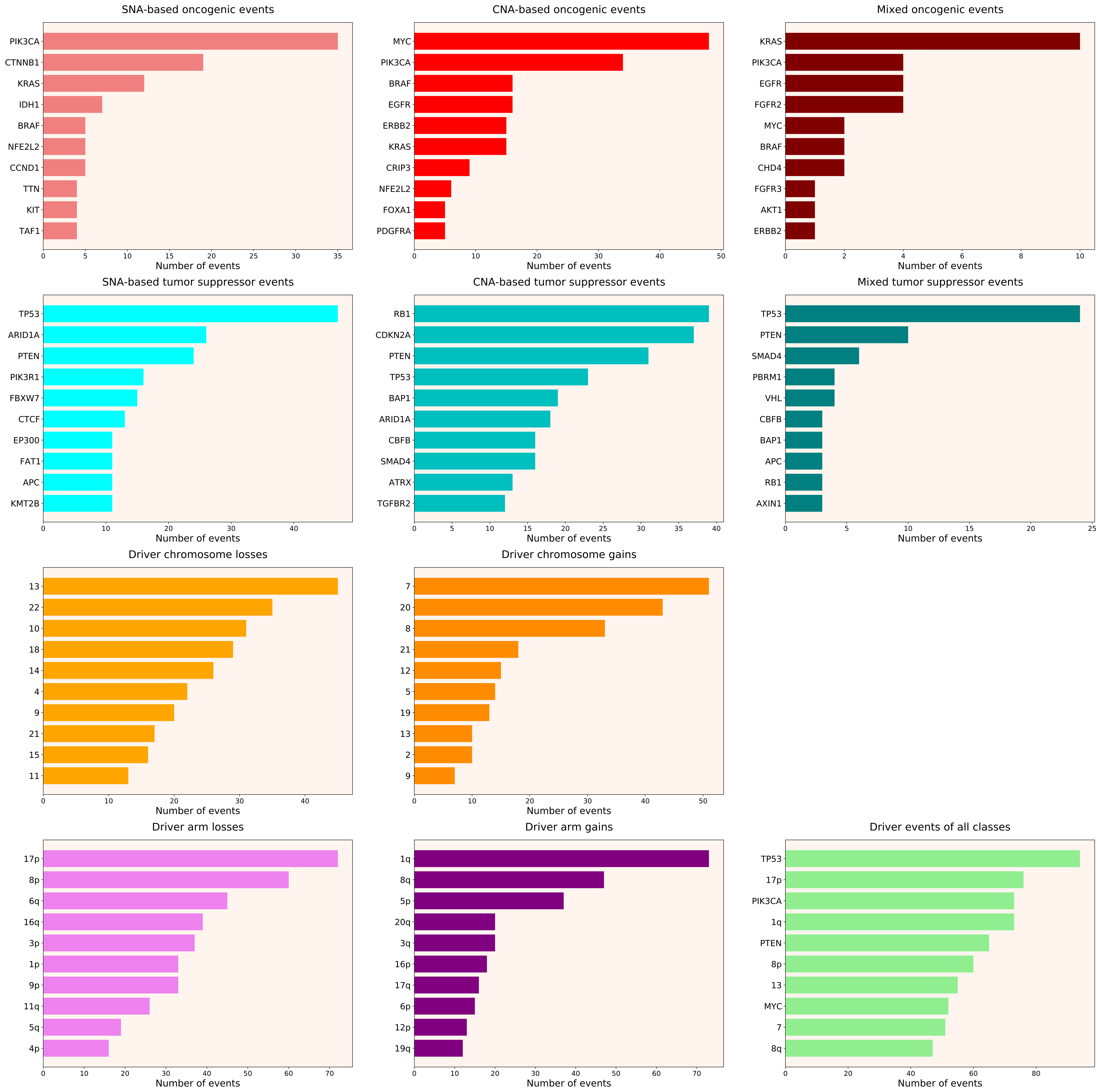

### 2021_8_16_14_9_distribution_events_detailed_13.pdf

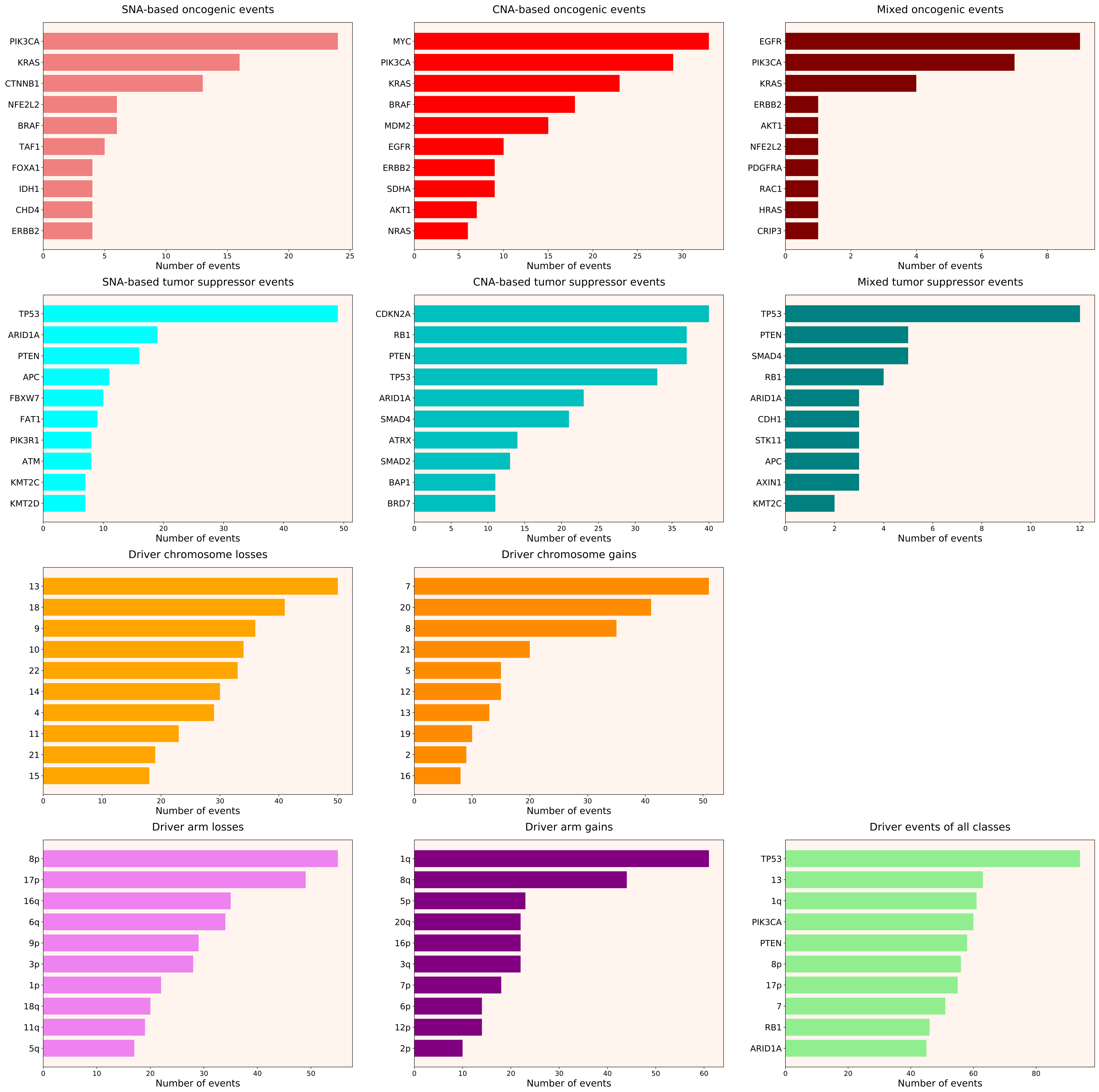

### 2021_8_16_14_9_distribution_events_detailed_15.pdf

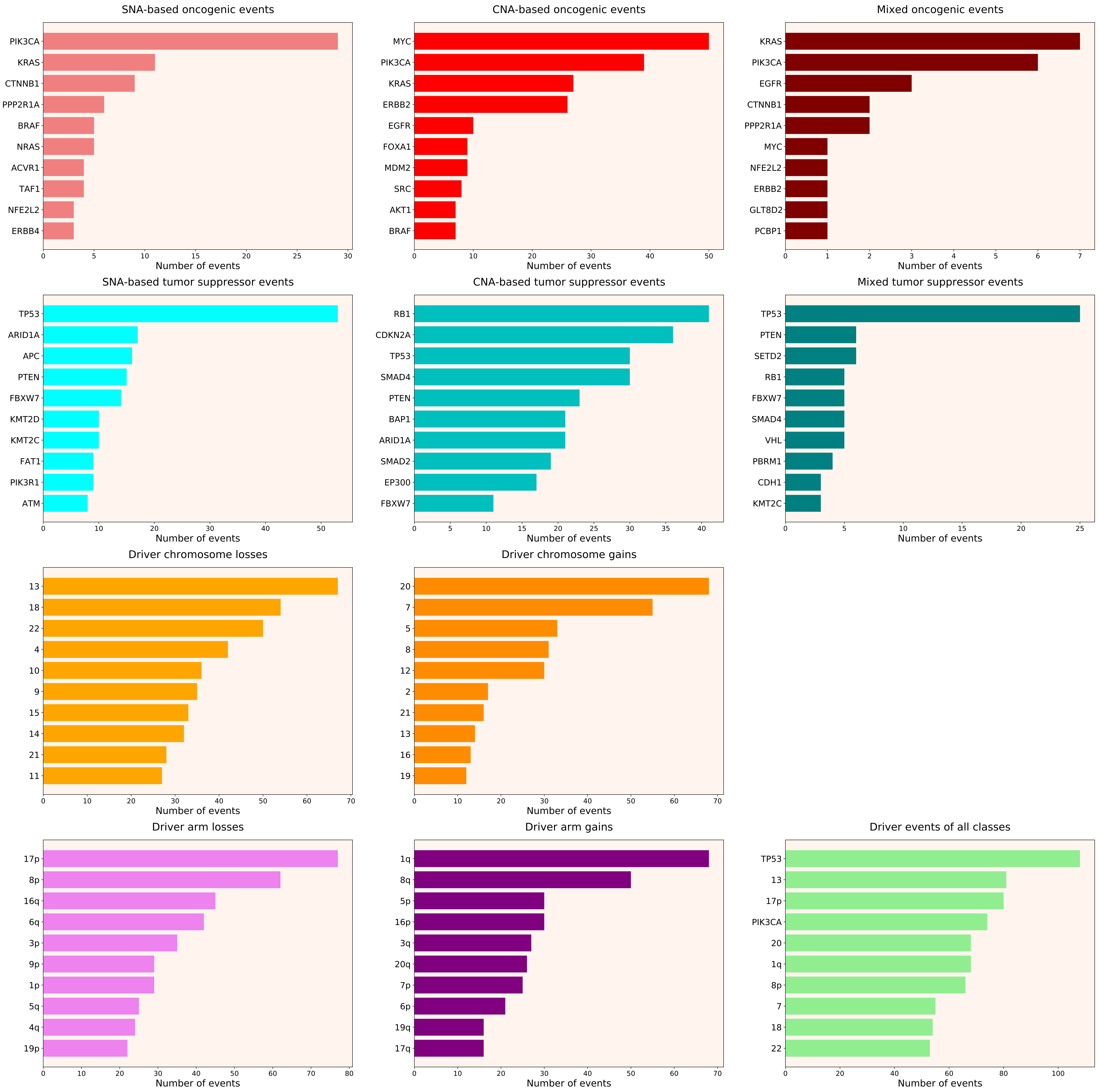

### 2021_8_16_14_9_distribution_events_detailed_29.pdf

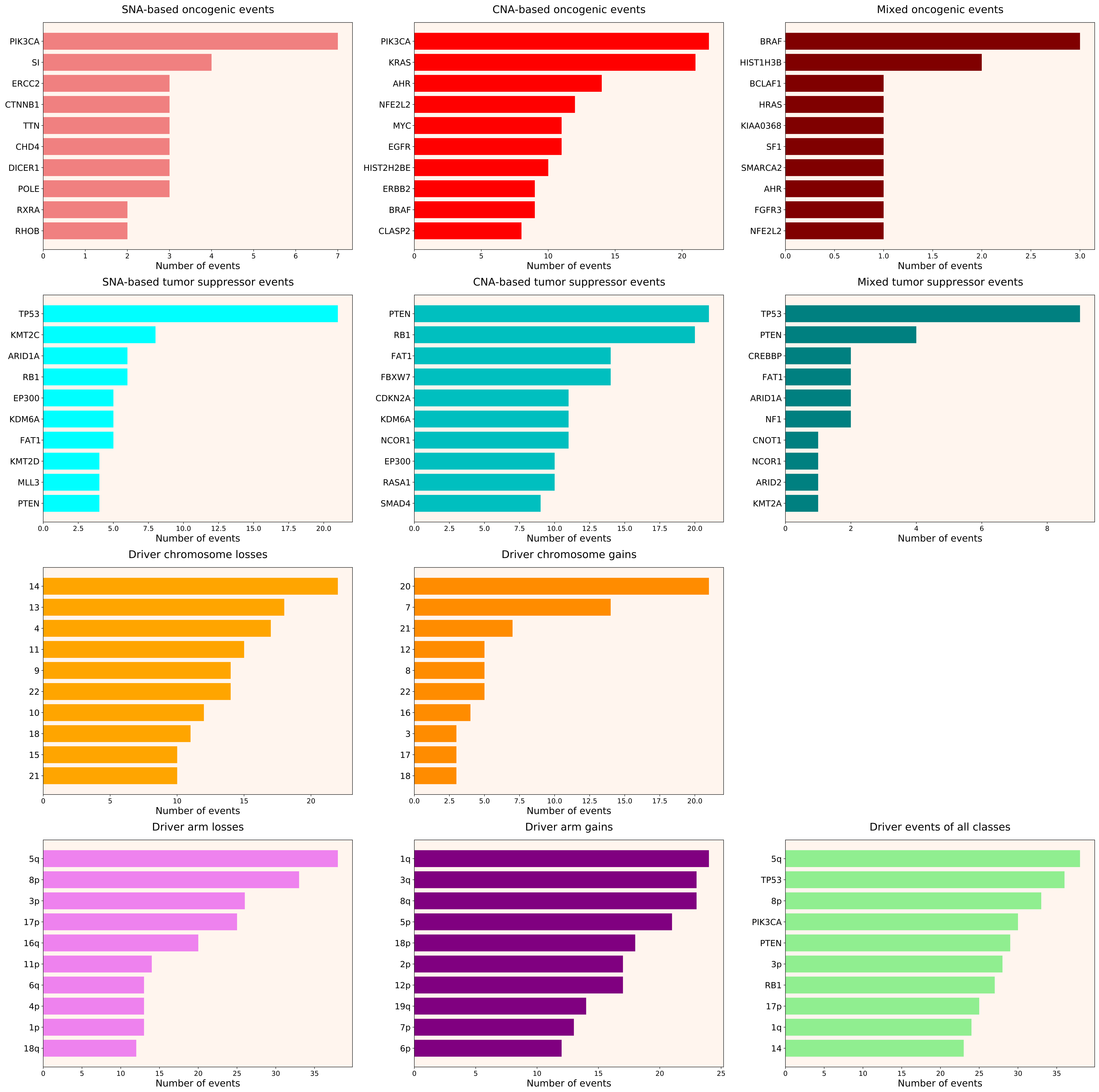

### 2021_8_16_14_9_distribution_events_detailed_38.pdf

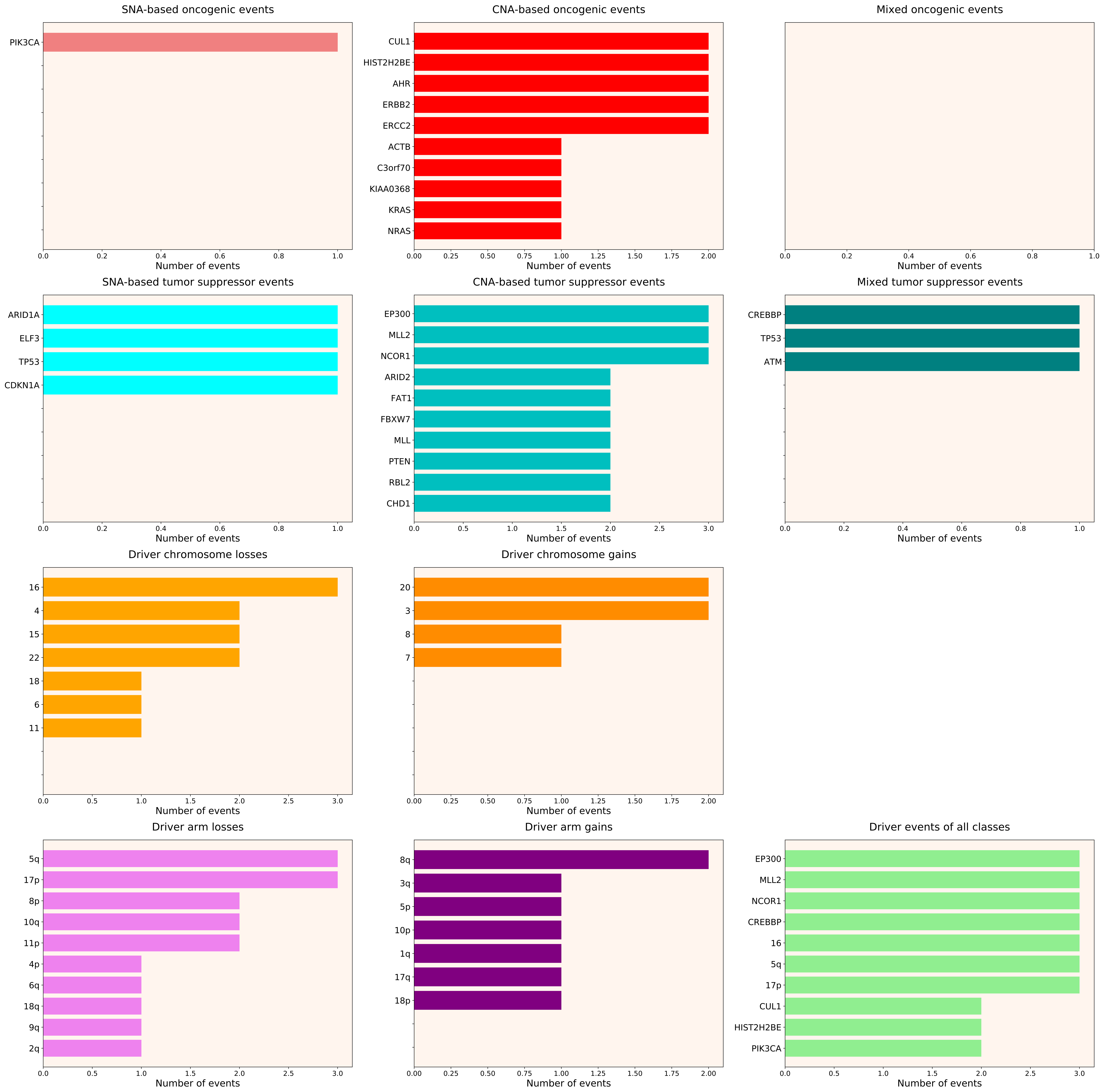

### 2021_8_16_14_9_distribution_events_detailed_39.pdf

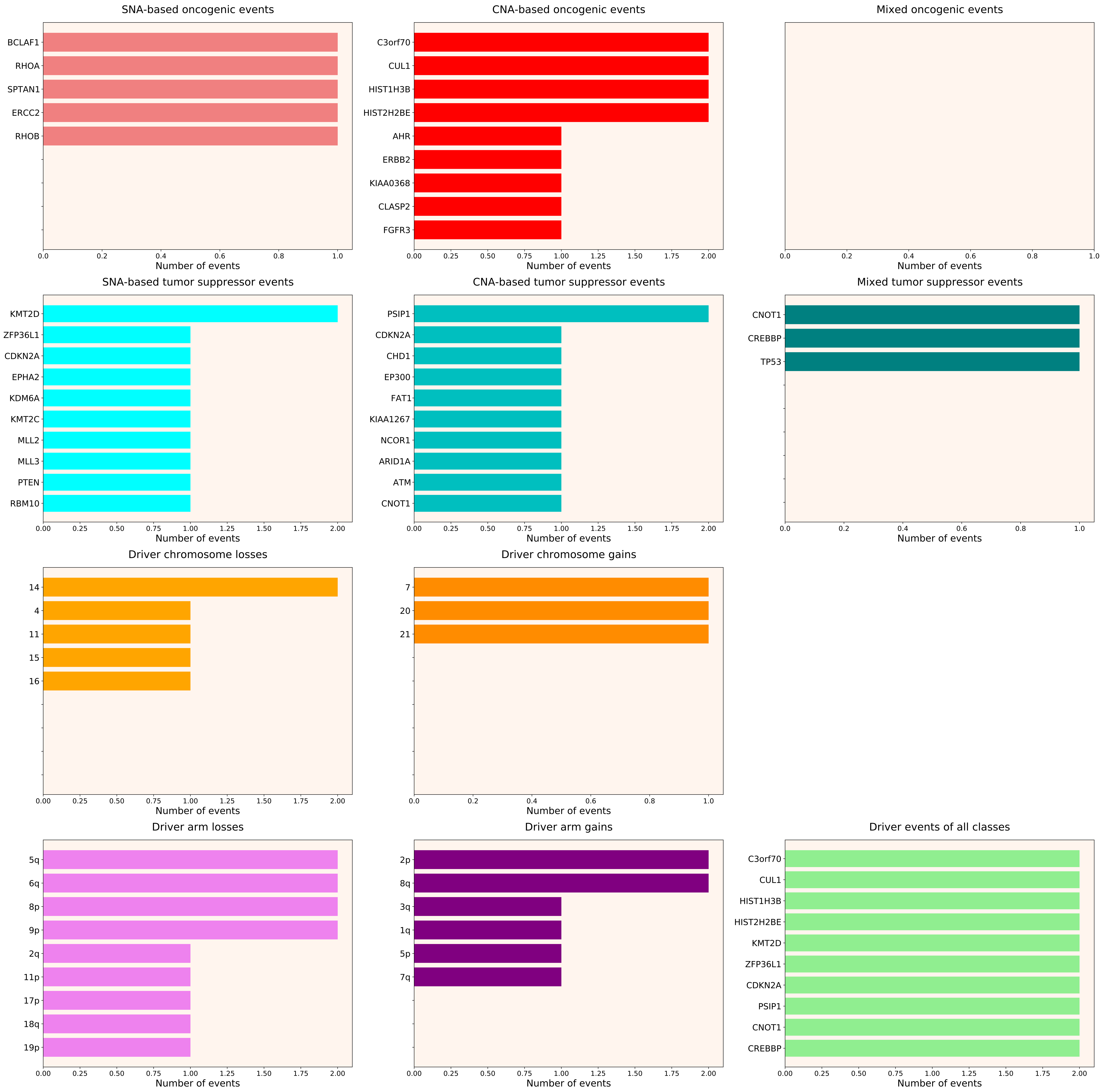

### 2021_8_16_14_9_distribution_events_detailed_DSI.pdf

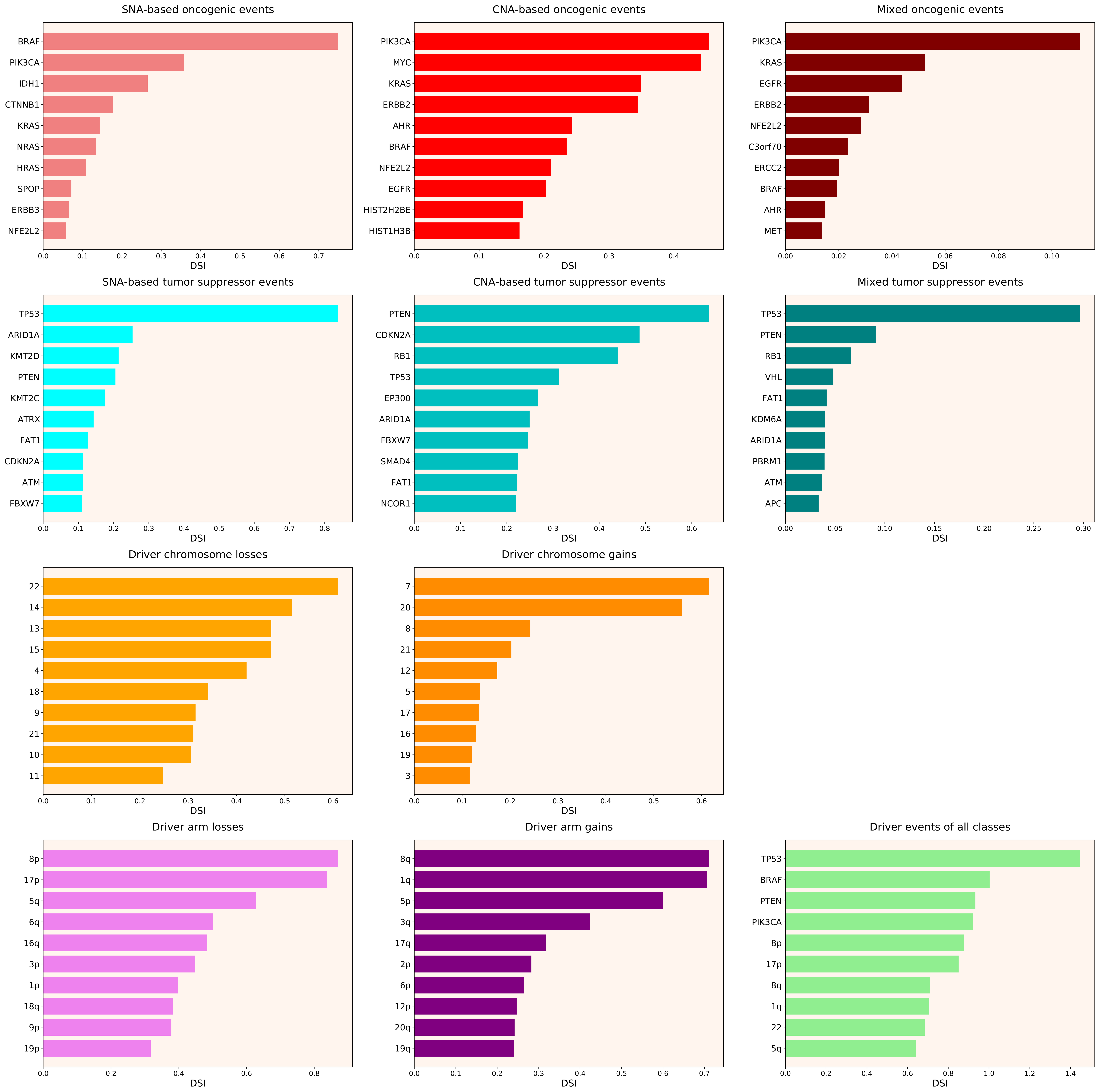

### 2021_8_16_14_9_distribution_events_detailed_DSI_females.pdf

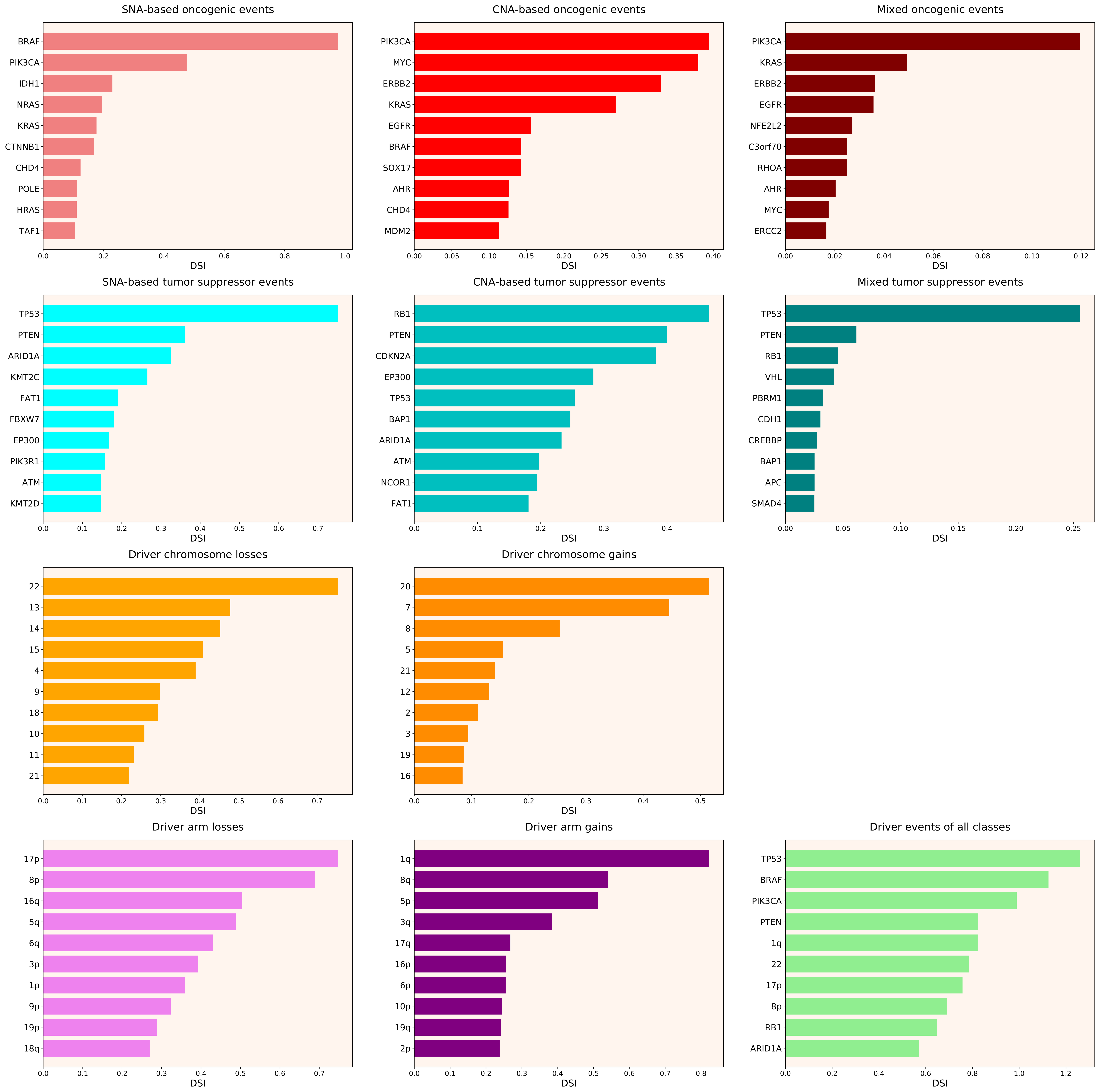

### 2021_8_16_14_9_distribution_events_detailed_DSI_males.pdf

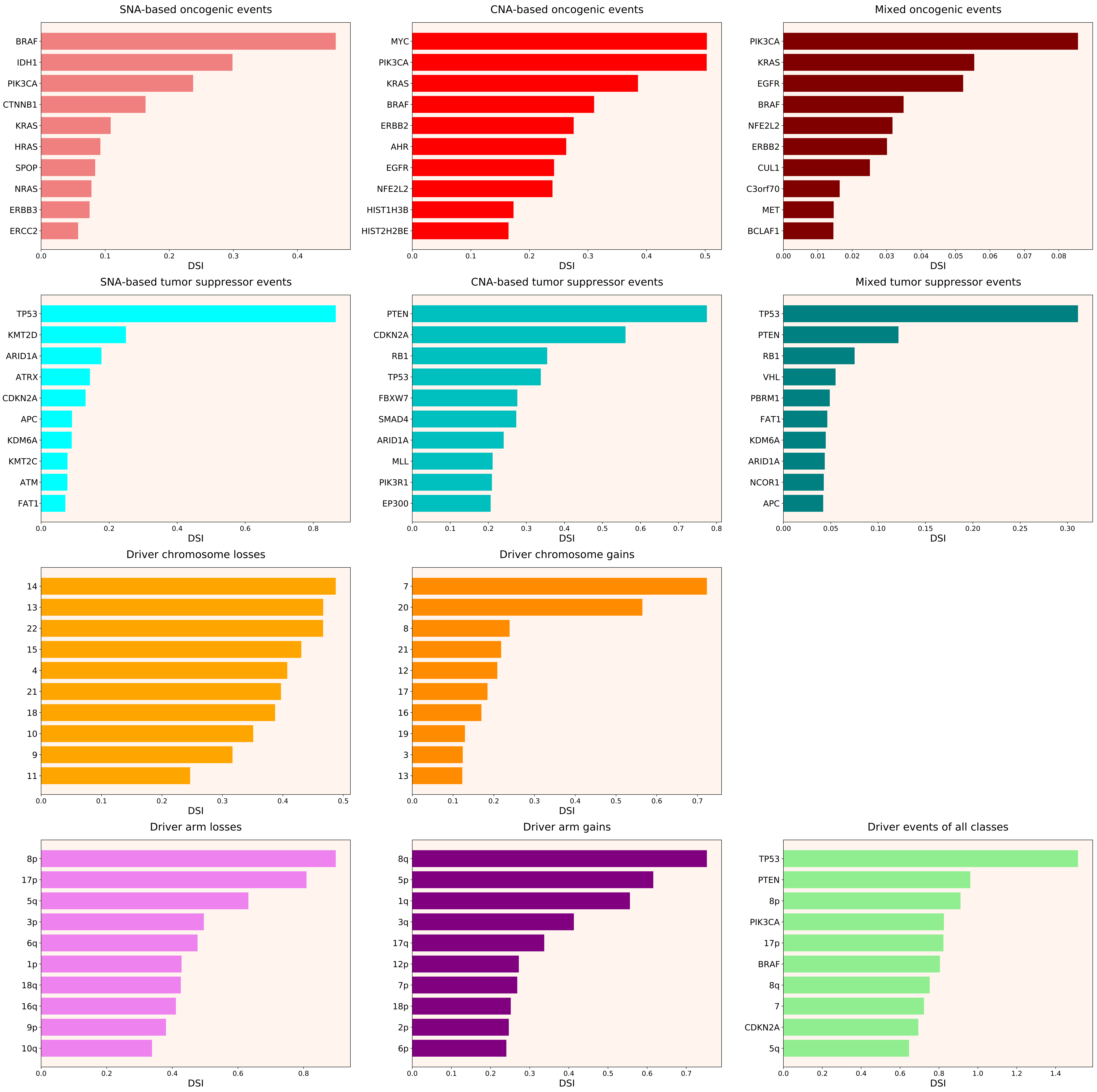

### 2021_8_16_14_9_distribution_events_detailed_females_9.pdf

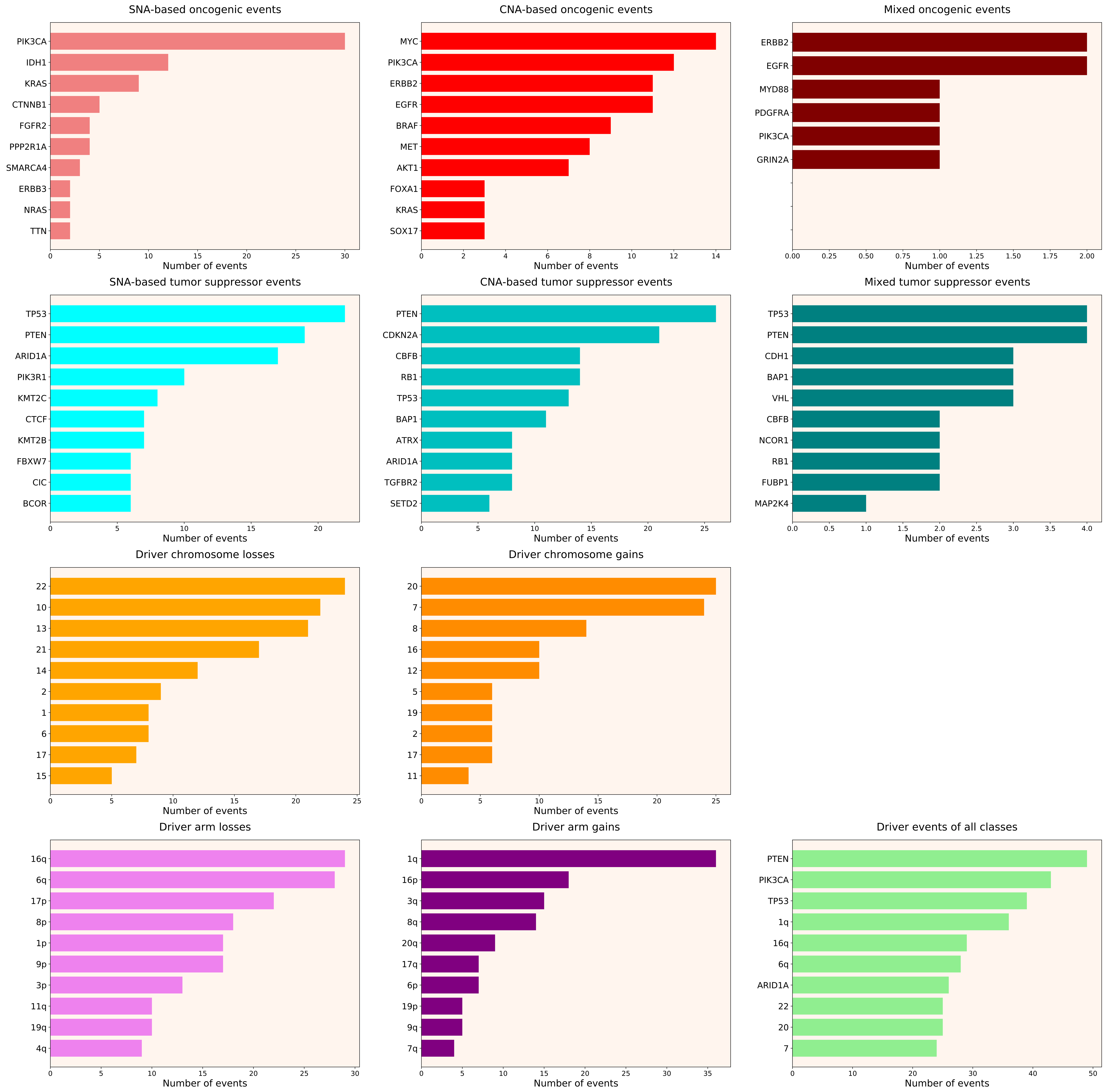

### 2021_8_16_14_9_distribution_events_detailed_females_18.pdf

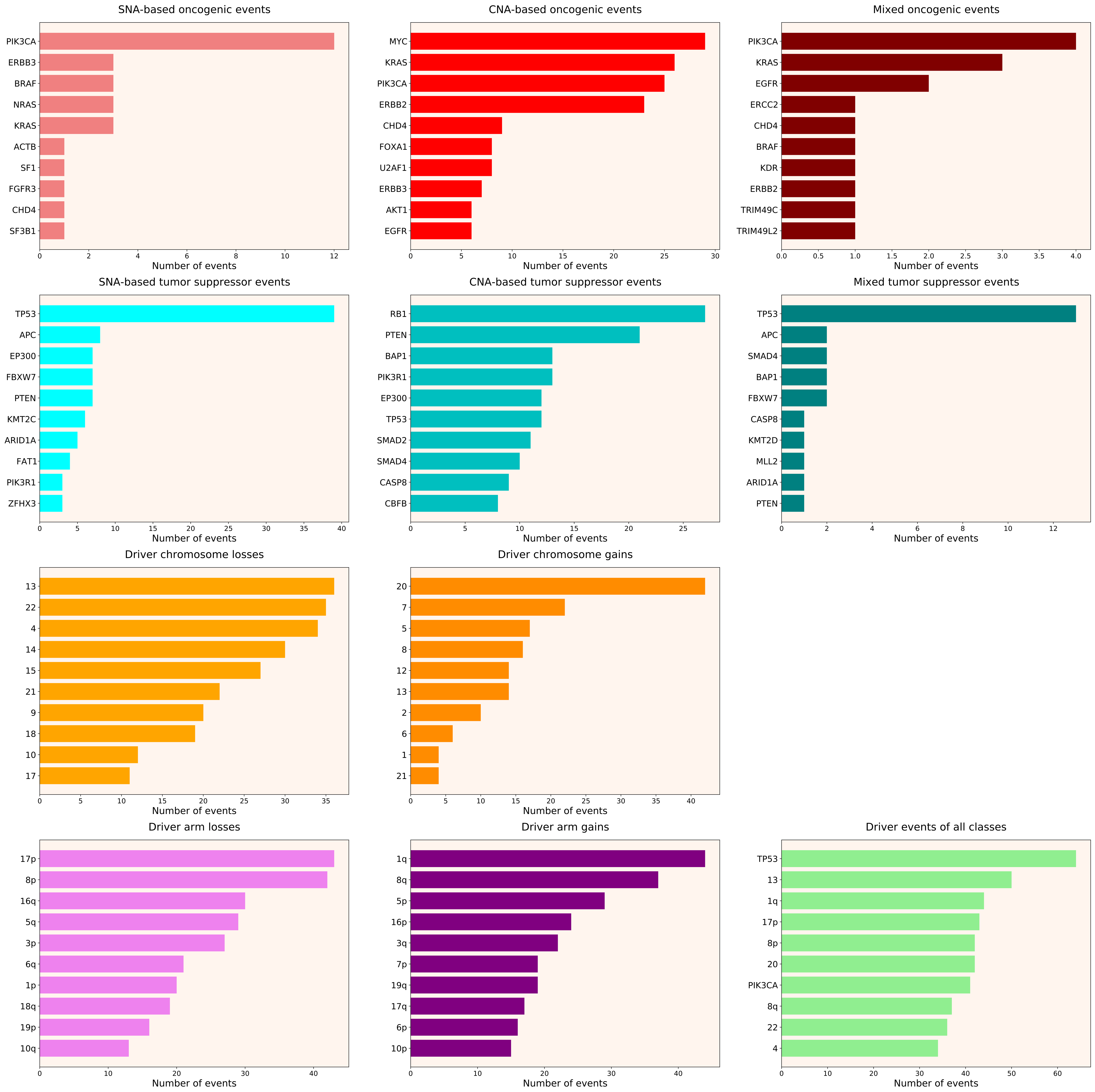

### 2021_8_16_14_9_distribution_events_detailed_females_19.pdf

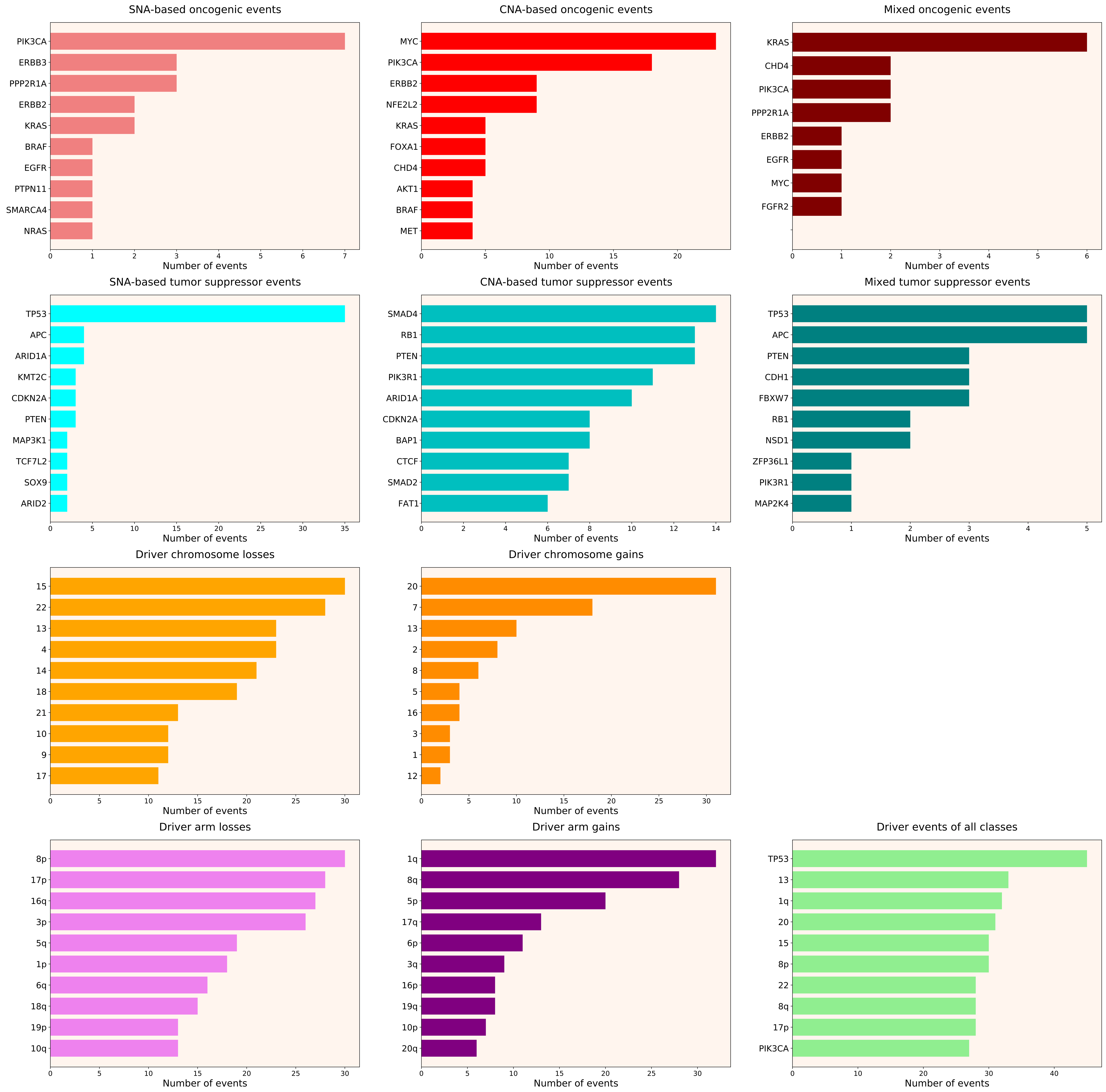

### 2021_8_16_14_9_distribution_events_detailed_females_20.pdf

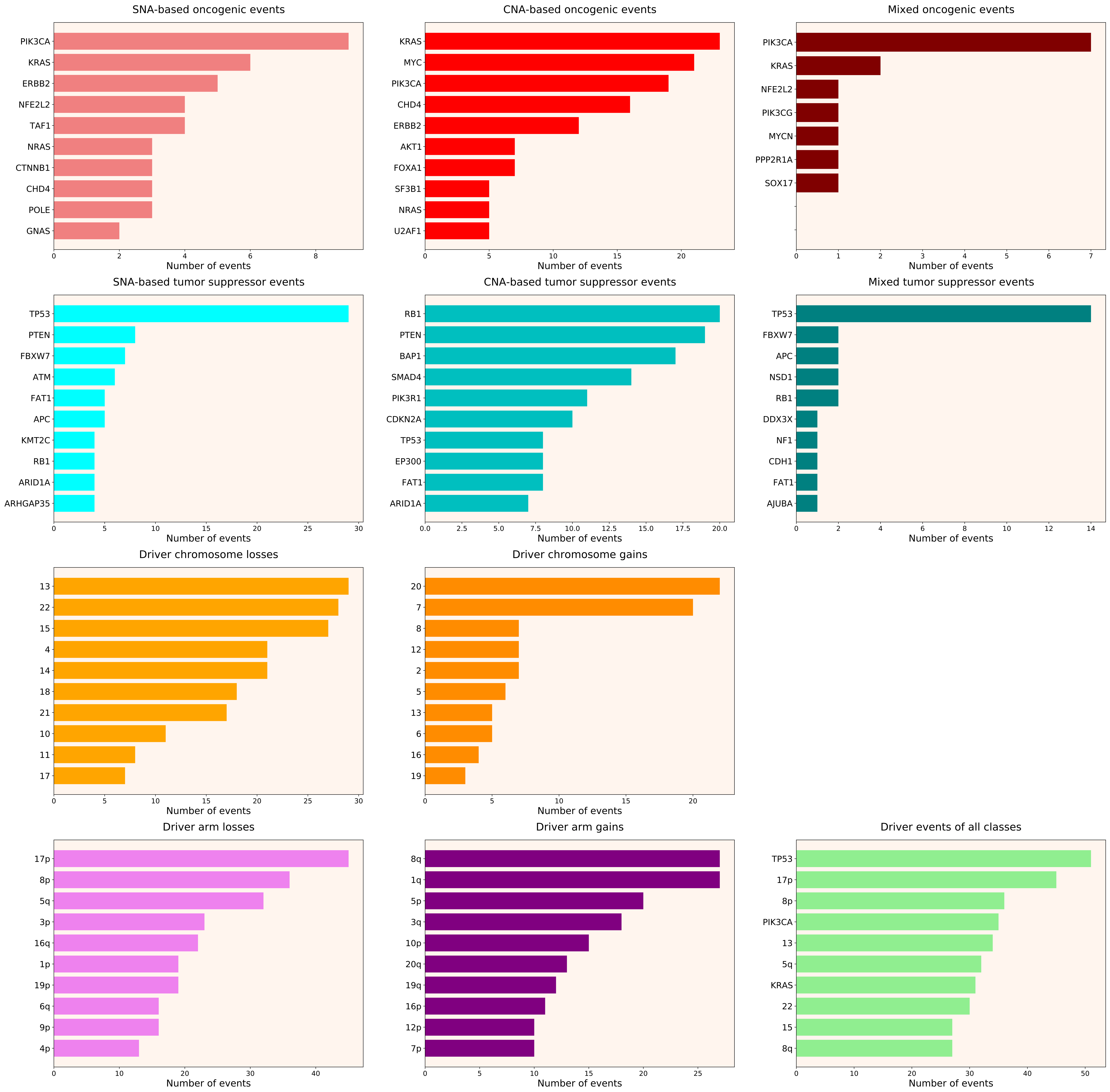

### 2021_8_16_14_9_distribution_events_detailed_females_21.pdf

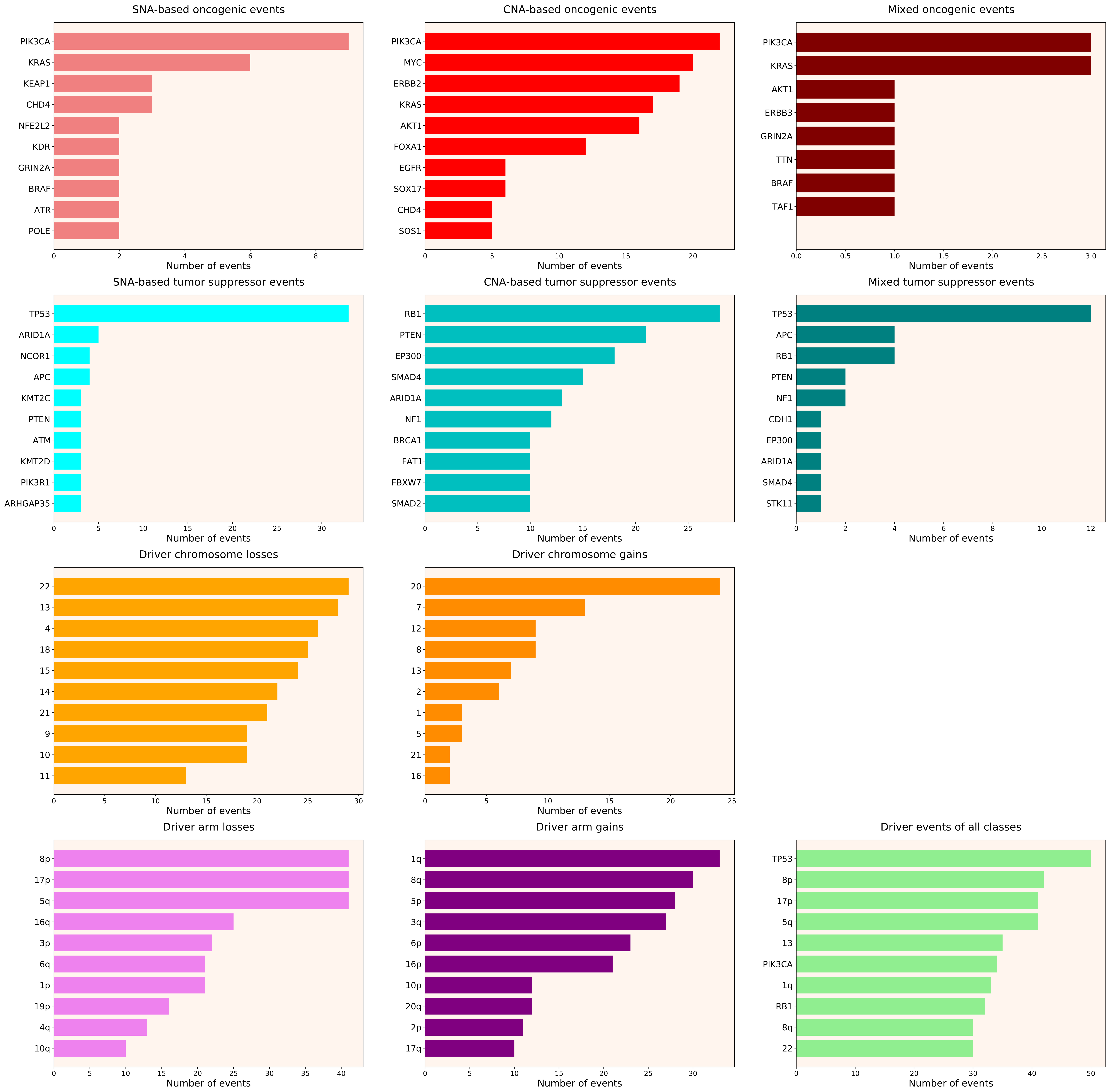

### 2021_8_16_14_9_distribution_events_detailed_females_22.pdf

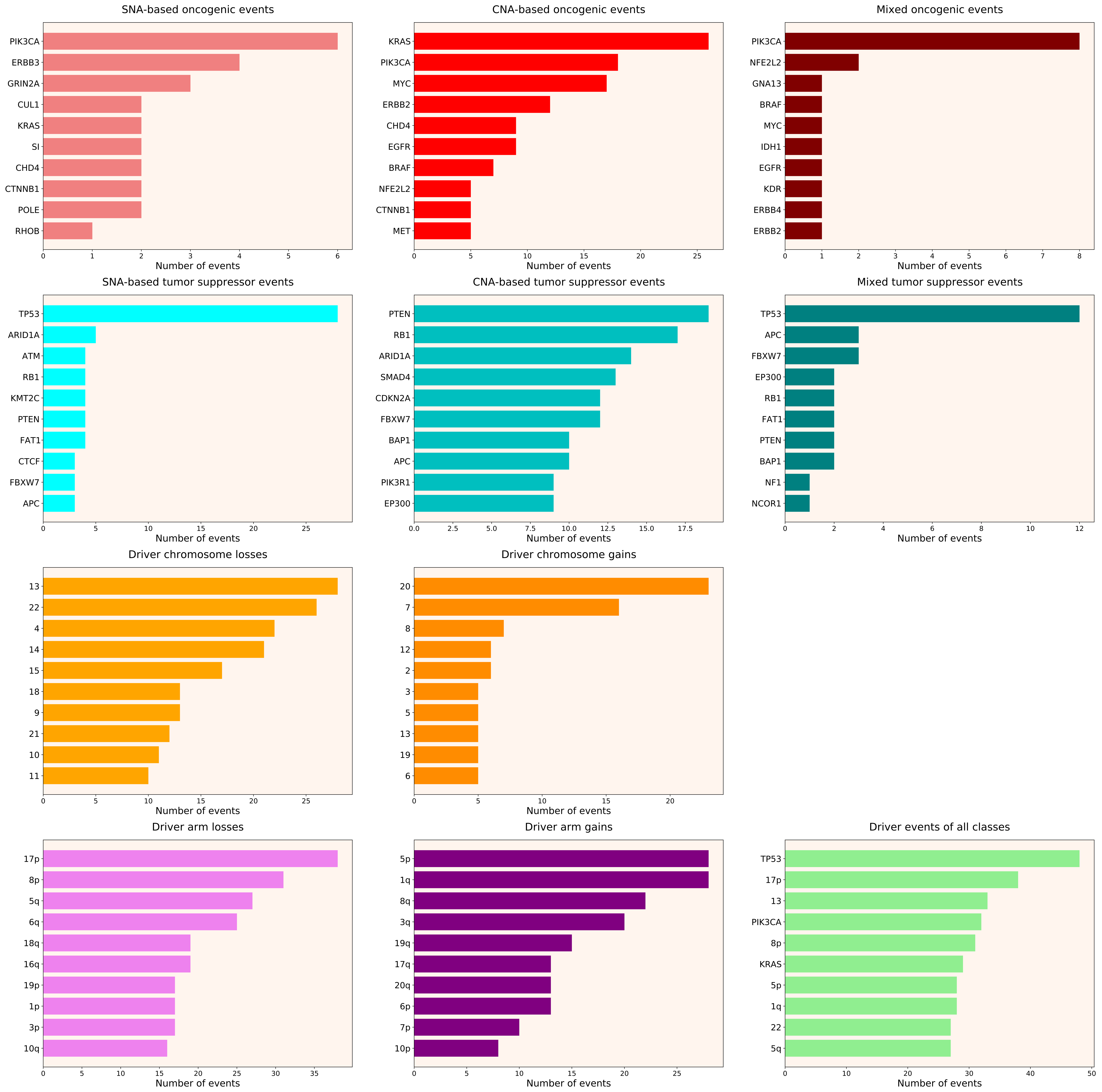

### 2021_8_16_14_9_distribution_events_detailed_females_23.pdf

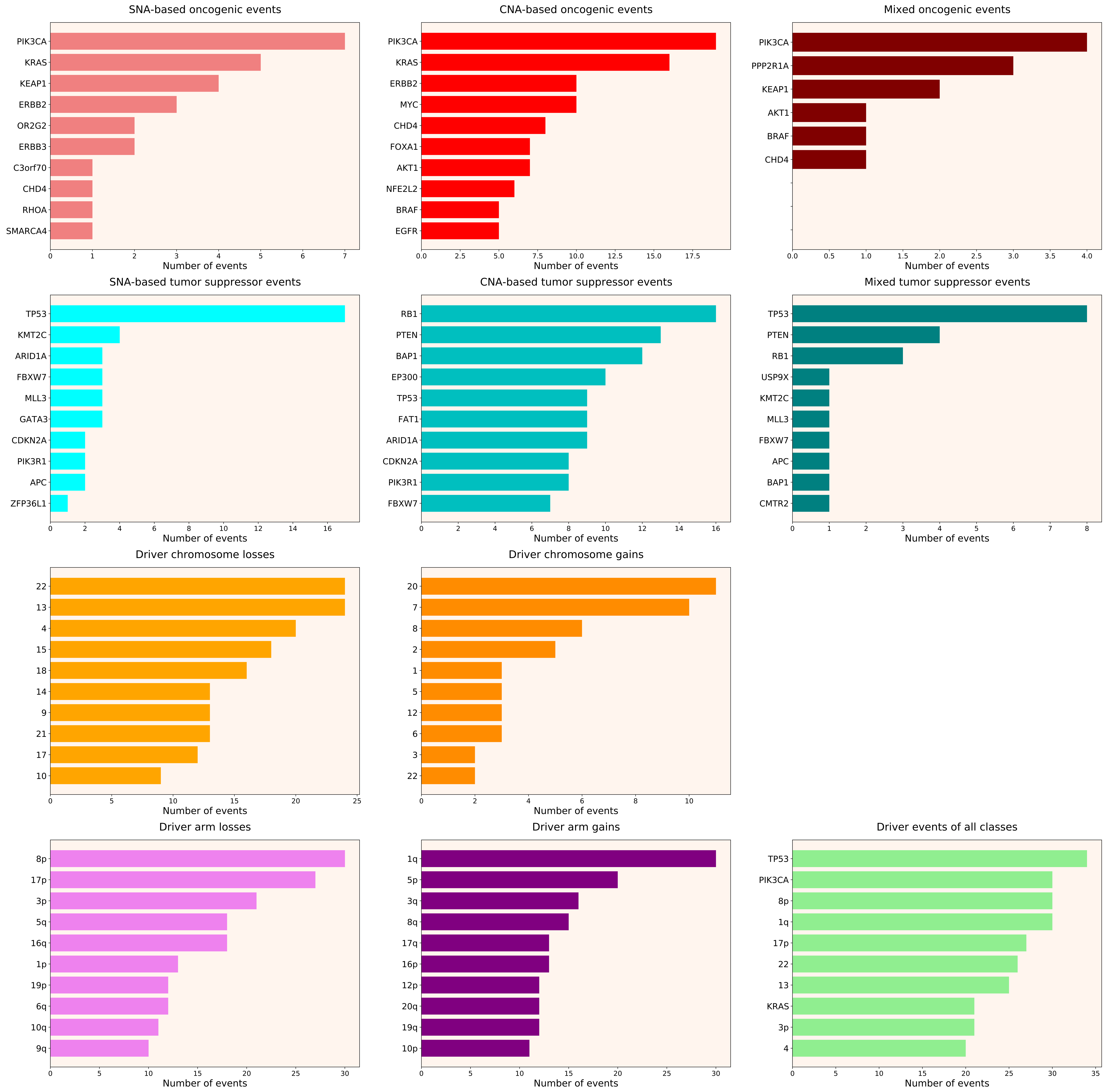

### 2021_8_16_14_9_distribution_events_detailed_females_24.pdf

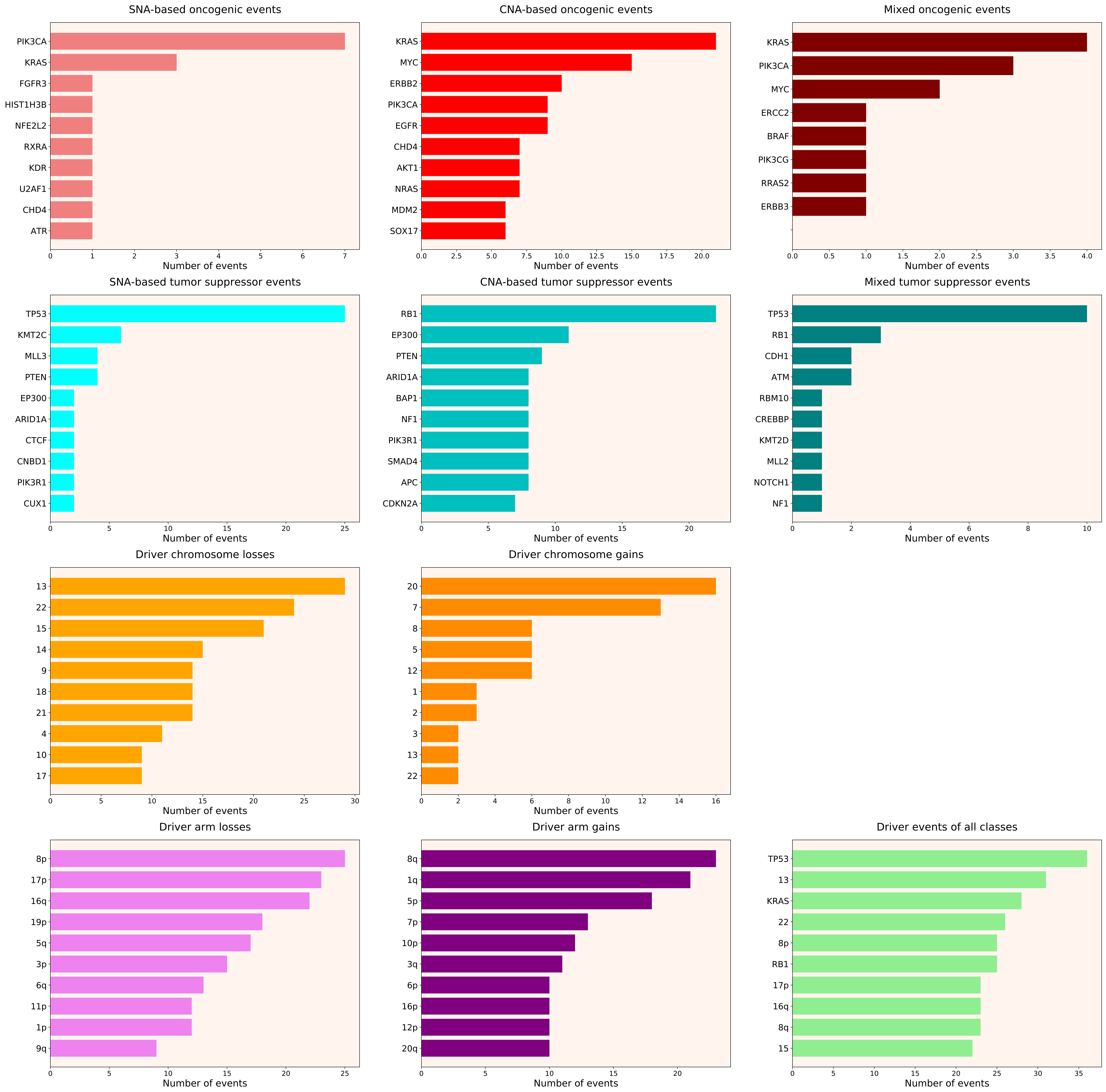

### 2021_8_16_14_9_distribution_events_detailed_females_30.pdf

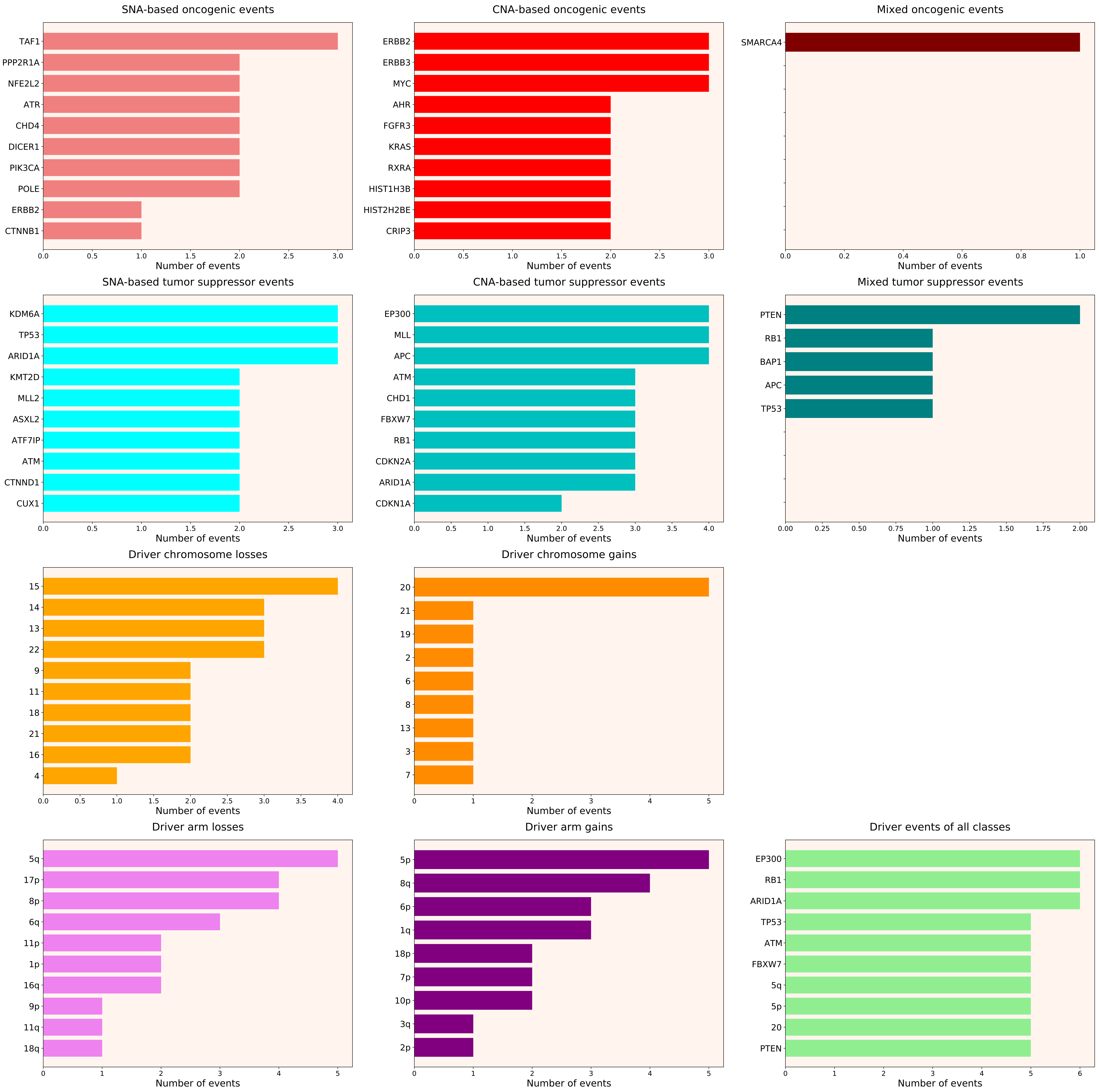

### 2021_8_16_14_9_distribution_events_detailed_females_31.pdf

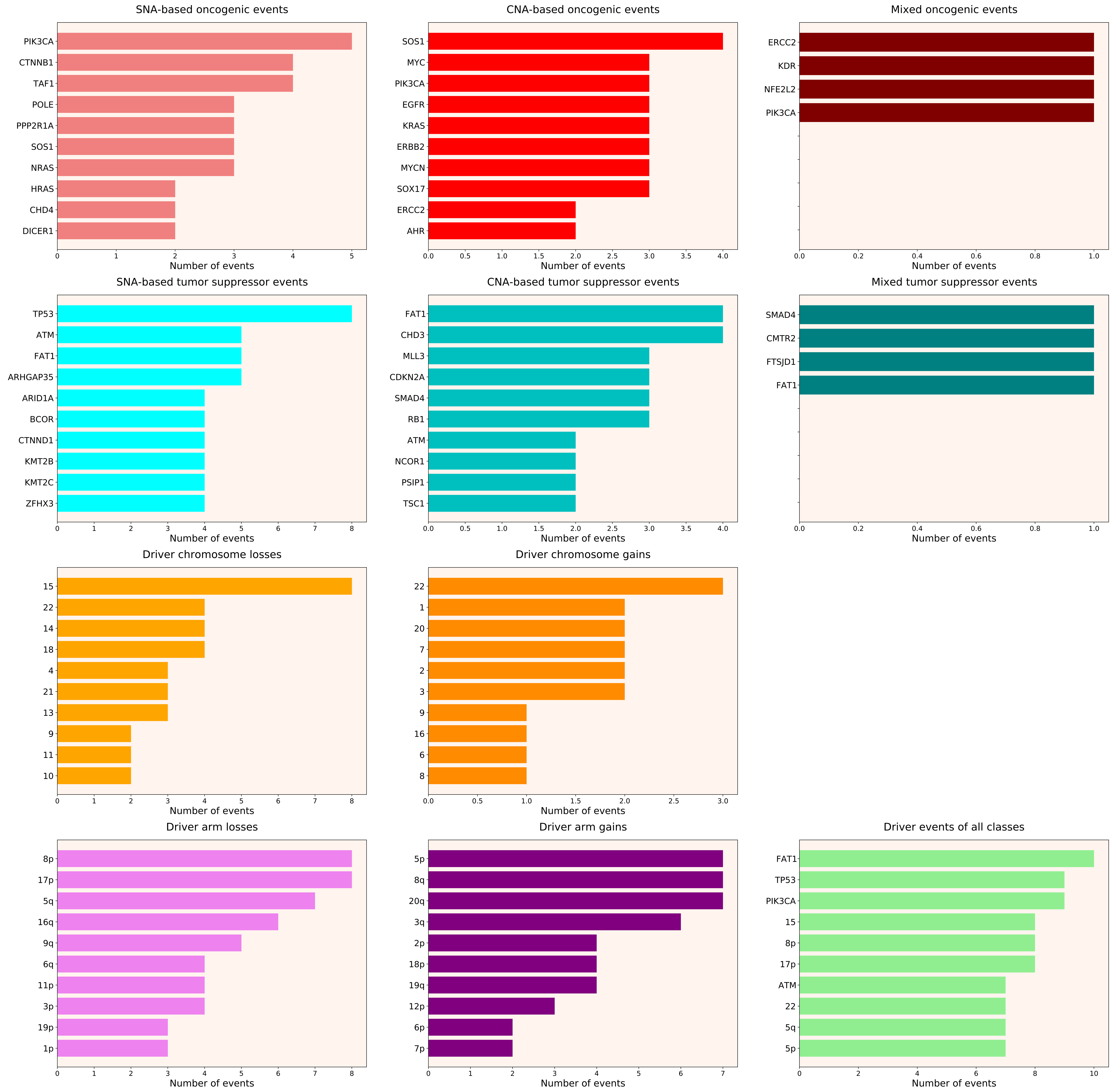

### 2021_8_16_14_9_distribution_events_detailed_females_34.pdf

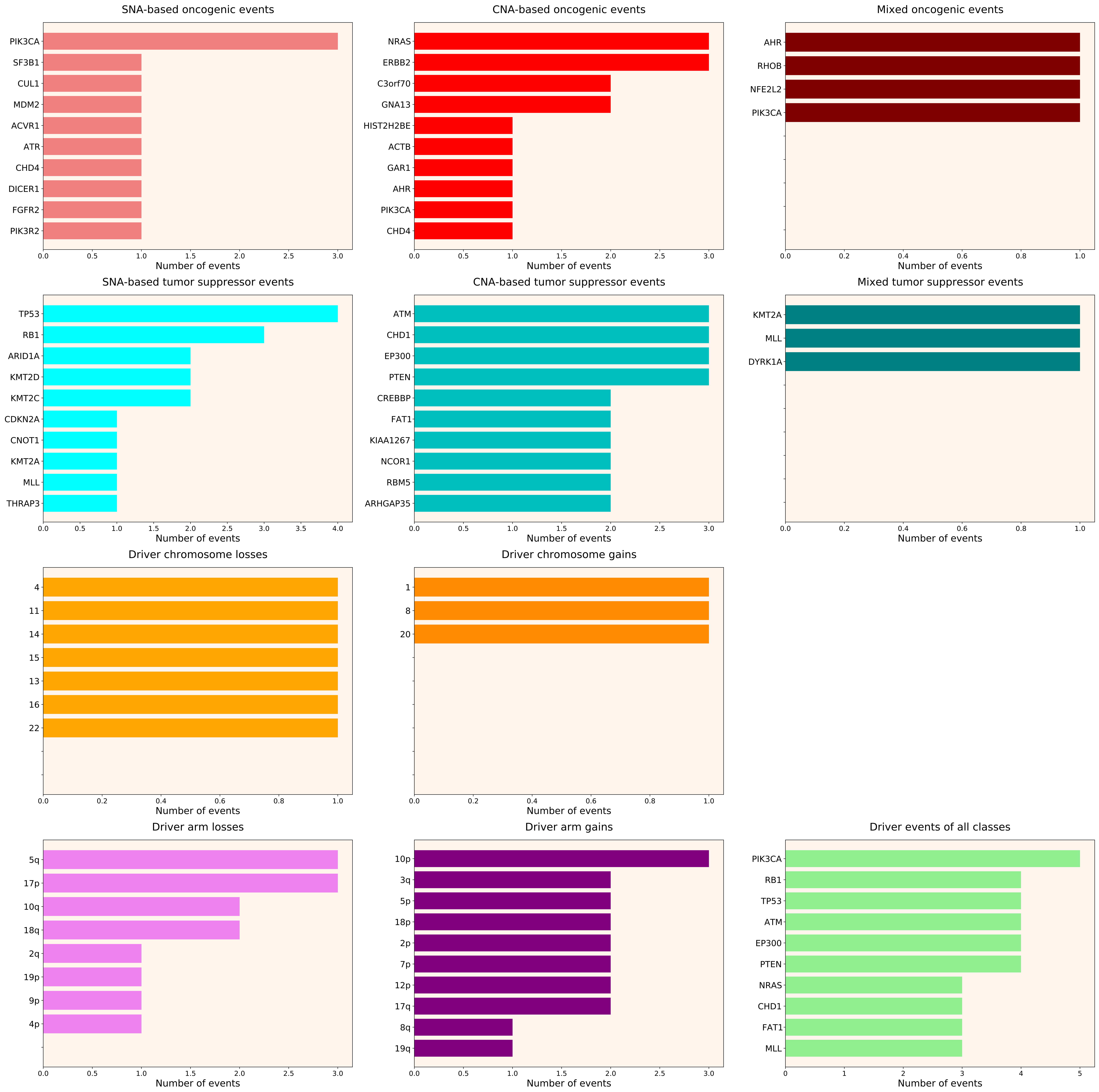

### 2021_8_16_14_9_distribution_events_detailed_females_35.pdf

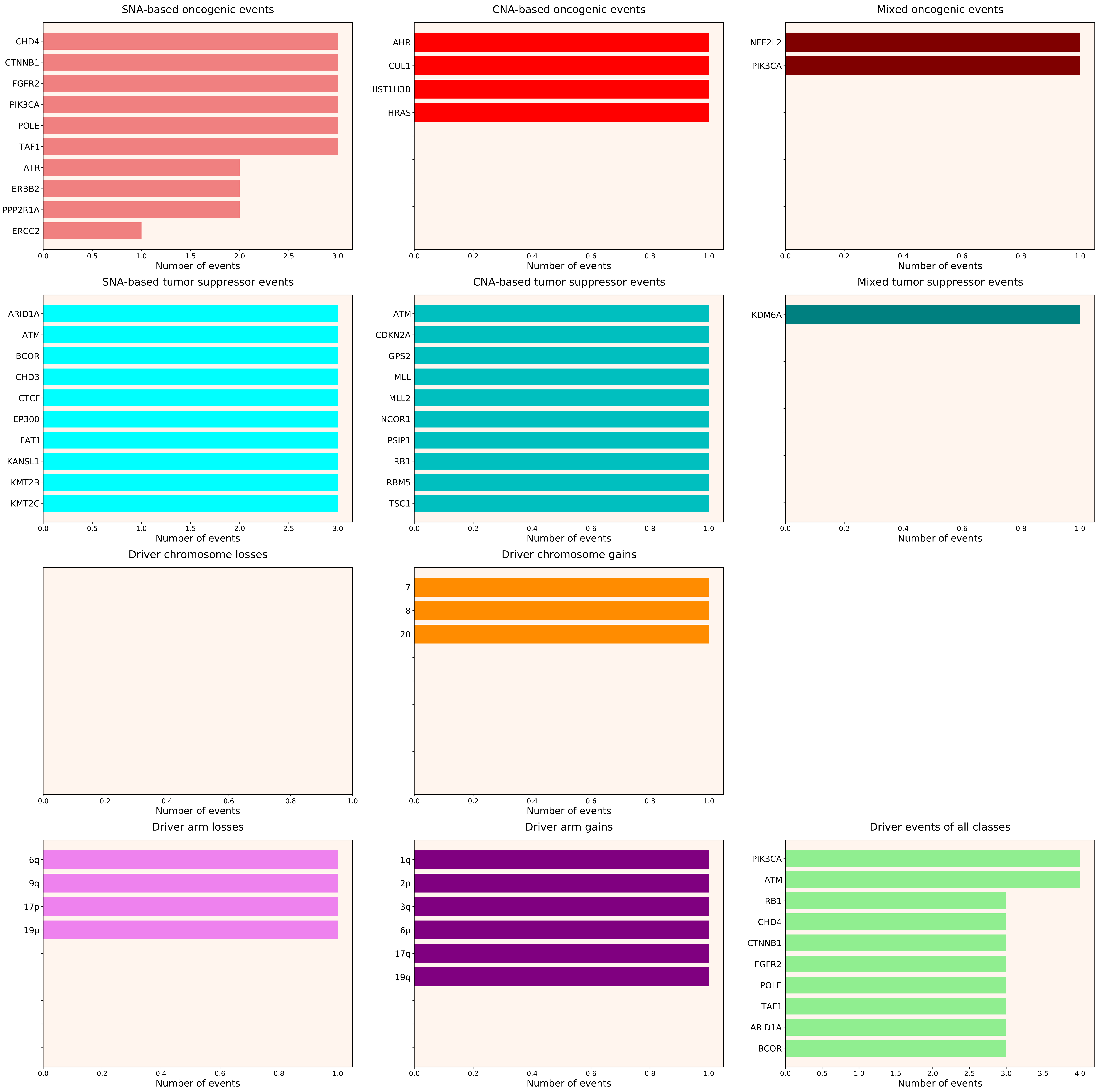

### 2021_8_16_14_9_distribution_events_detailed_females_36.pdf

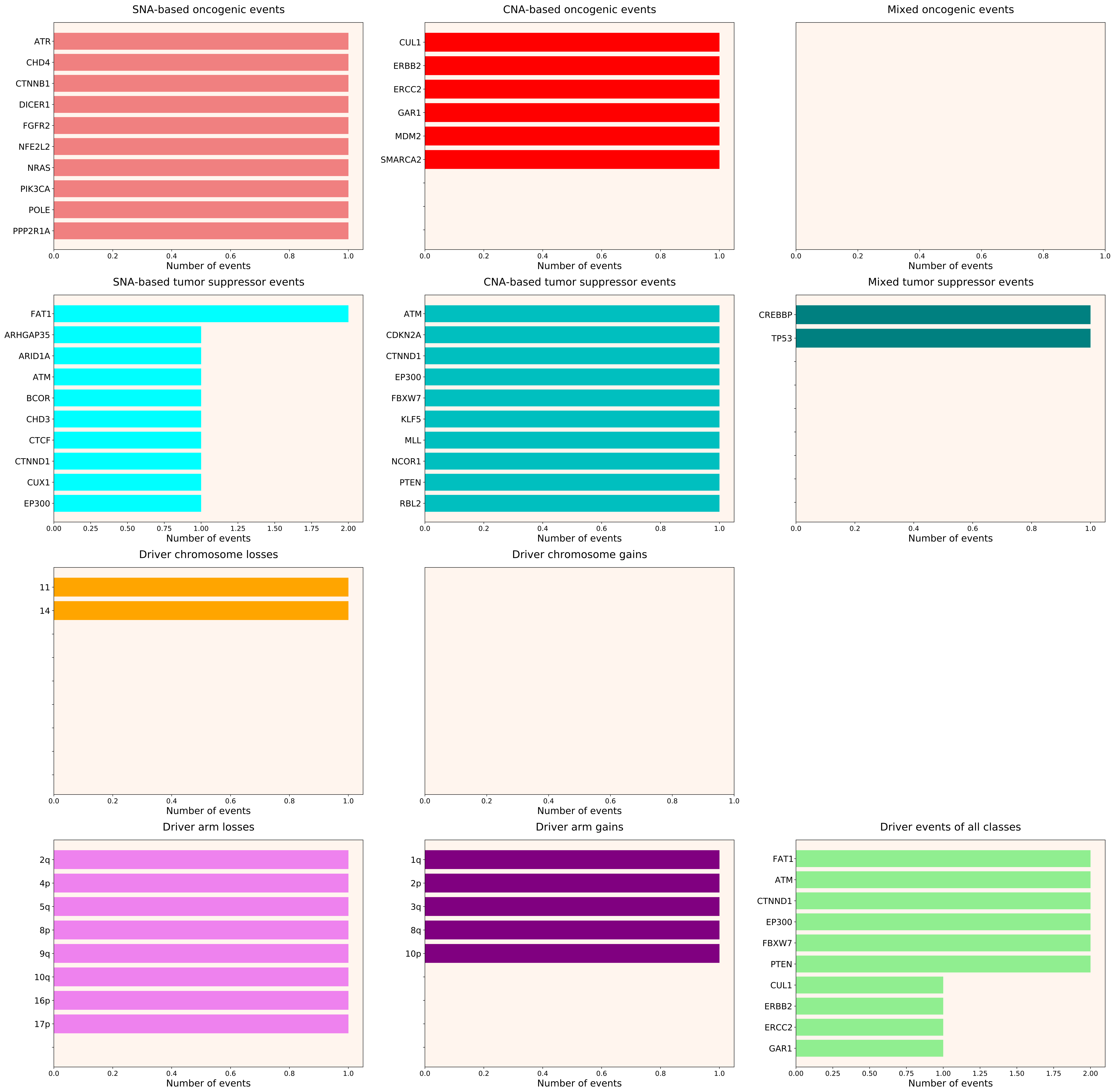

### 2021_8_16_14_9_distribution_events_detailed_females_37.pdf

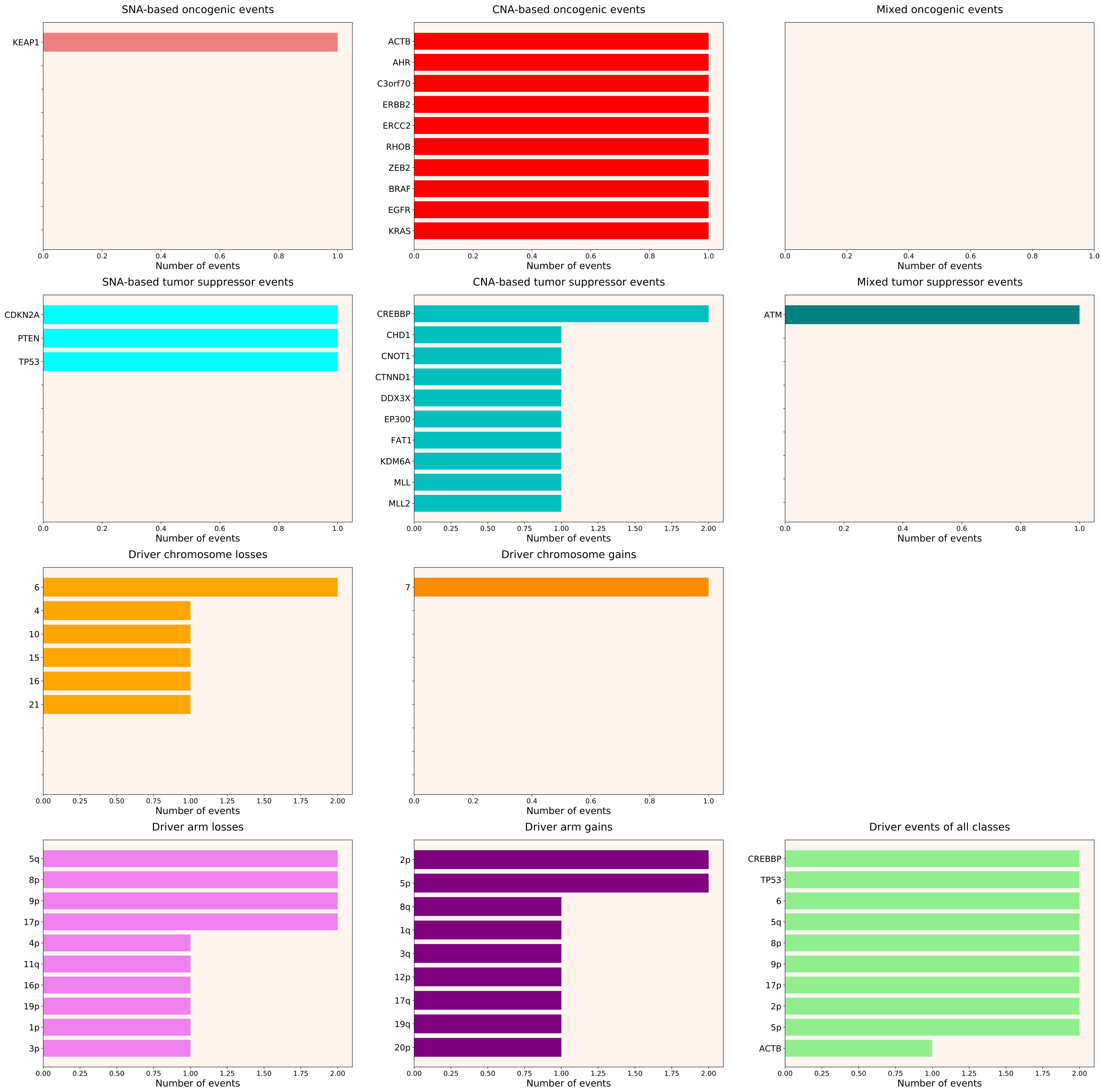

### 2021_8_16_14_9_distribution_events_detailed_males_8.pdf

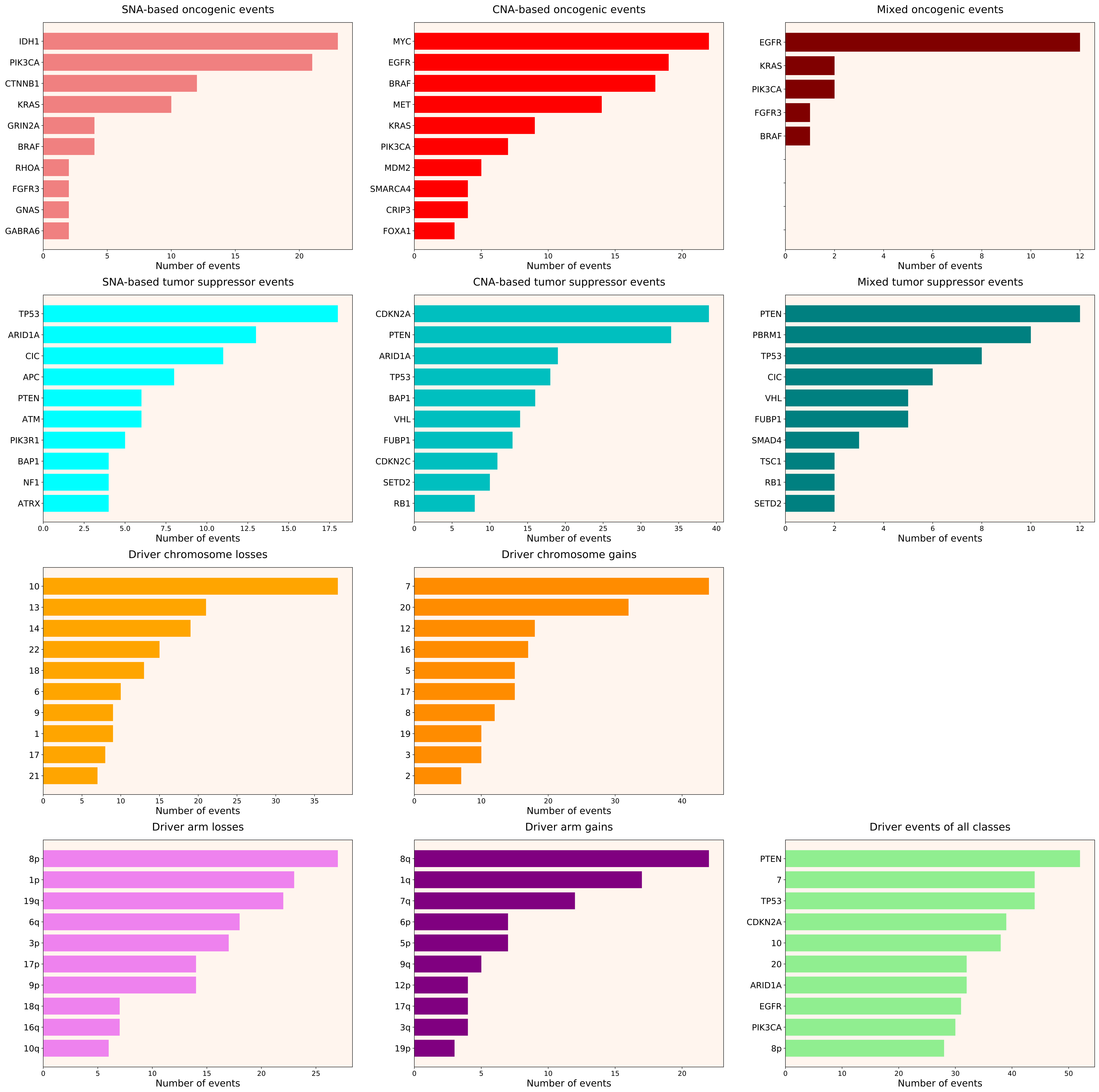

### 2021_8_16_14_9_distribution_events_detailed_males_9.pdf

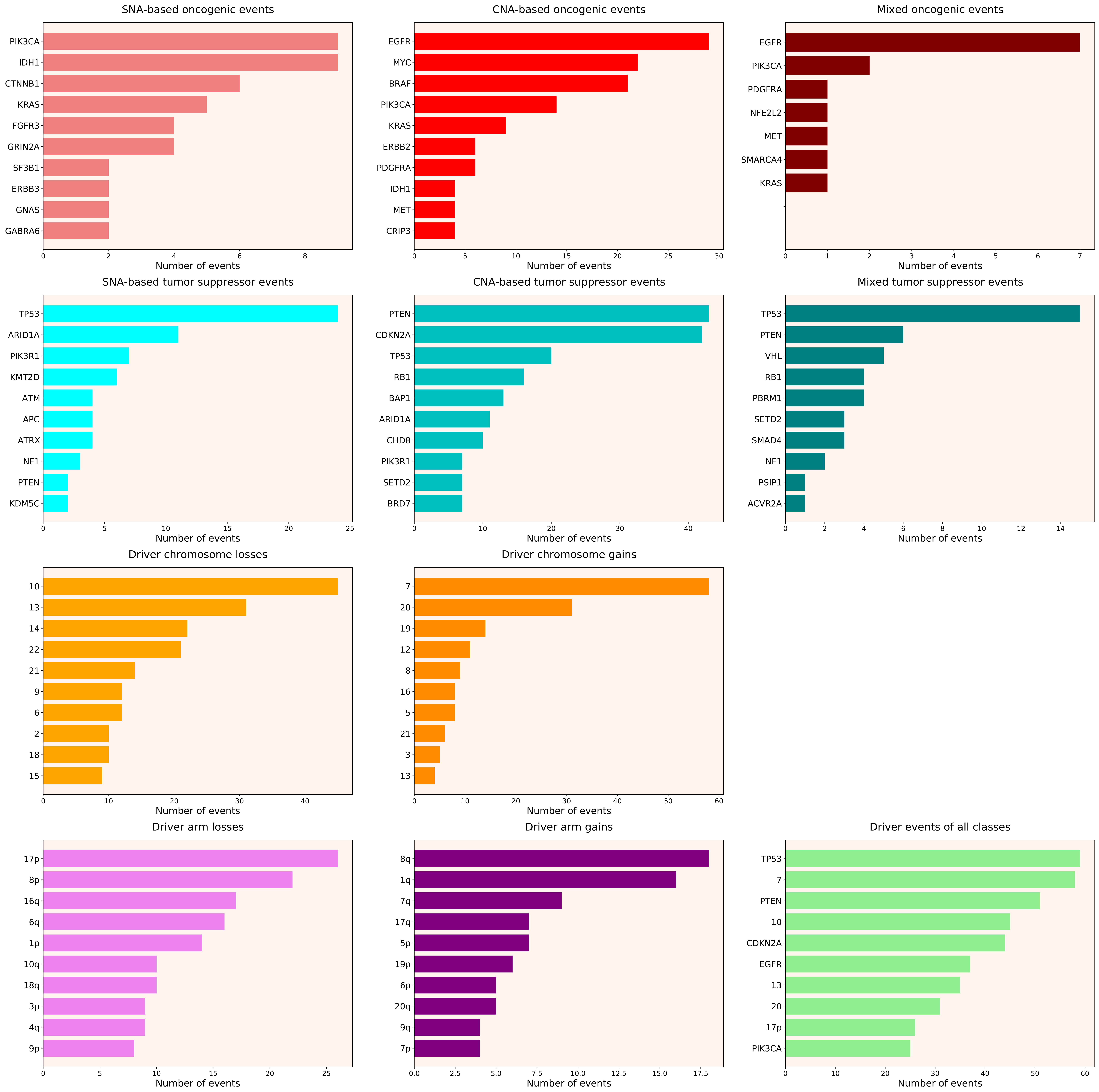

### 2021_8_16_14_9_distribution_events_detailed_males_18.pdf

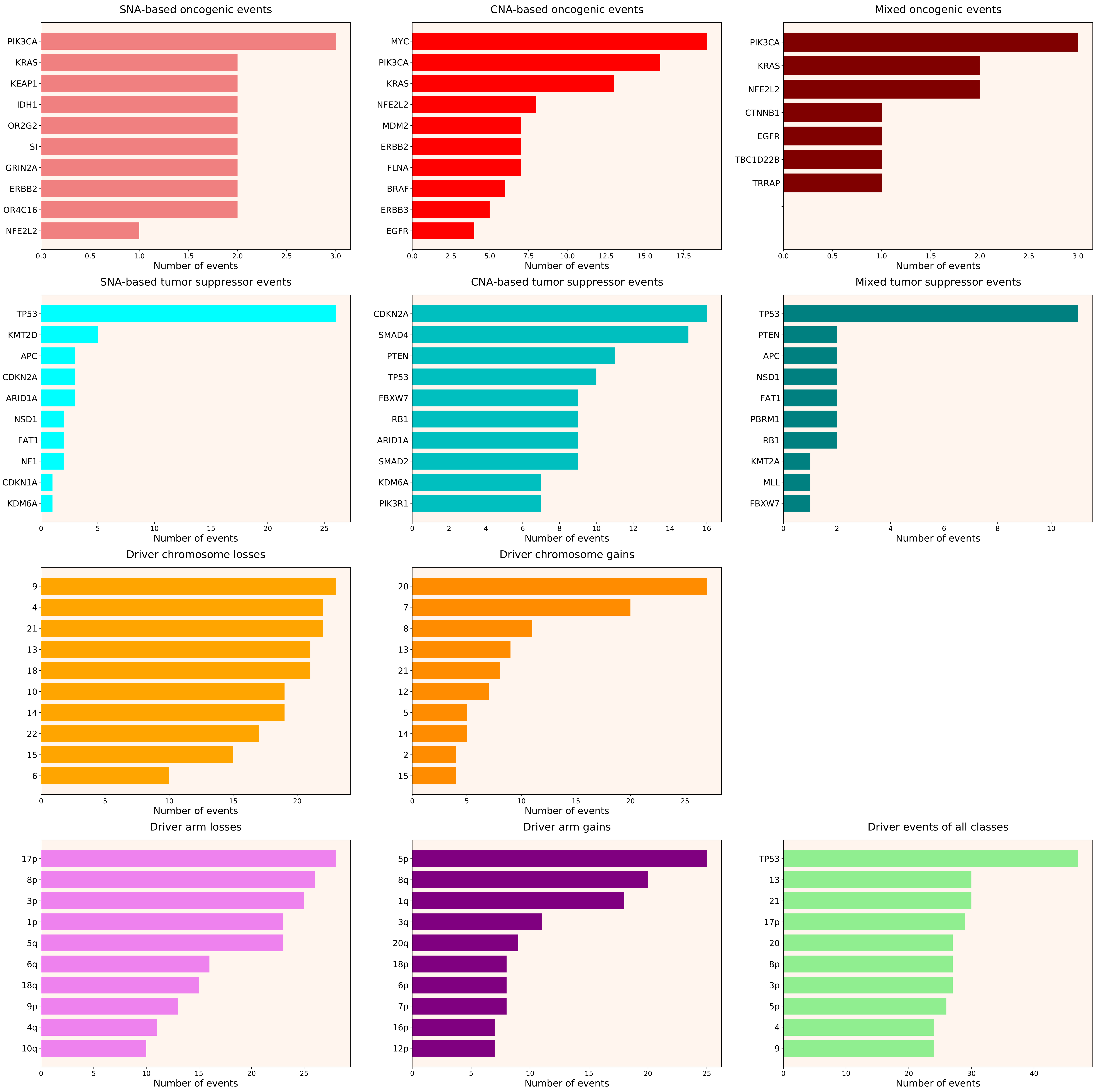

### 2021_8_16_14_9_distribution_events_detailed_males_19.pdf

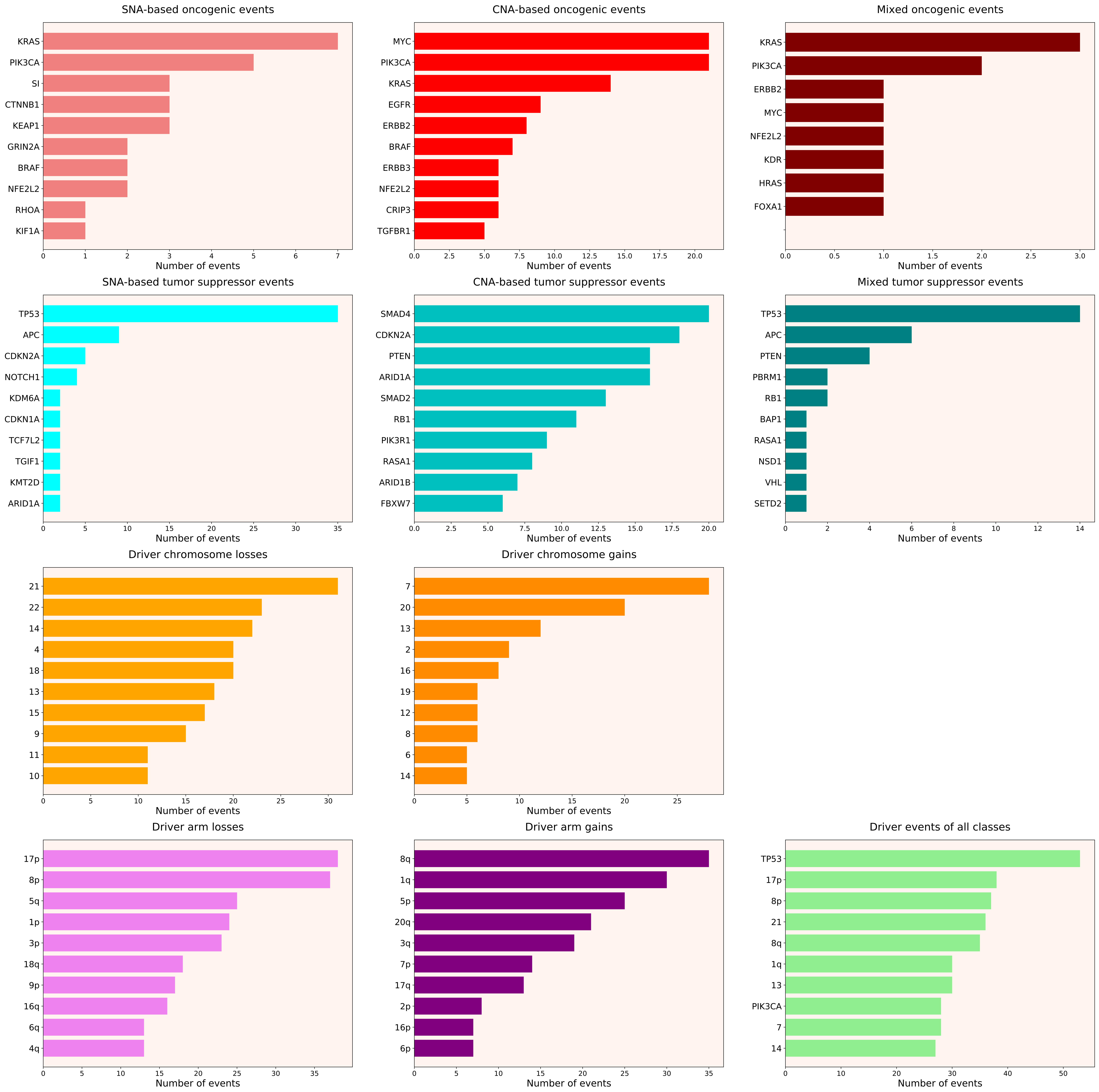

### 2021_8_16_14_9_distribution_events_detailed_males_20.pdf

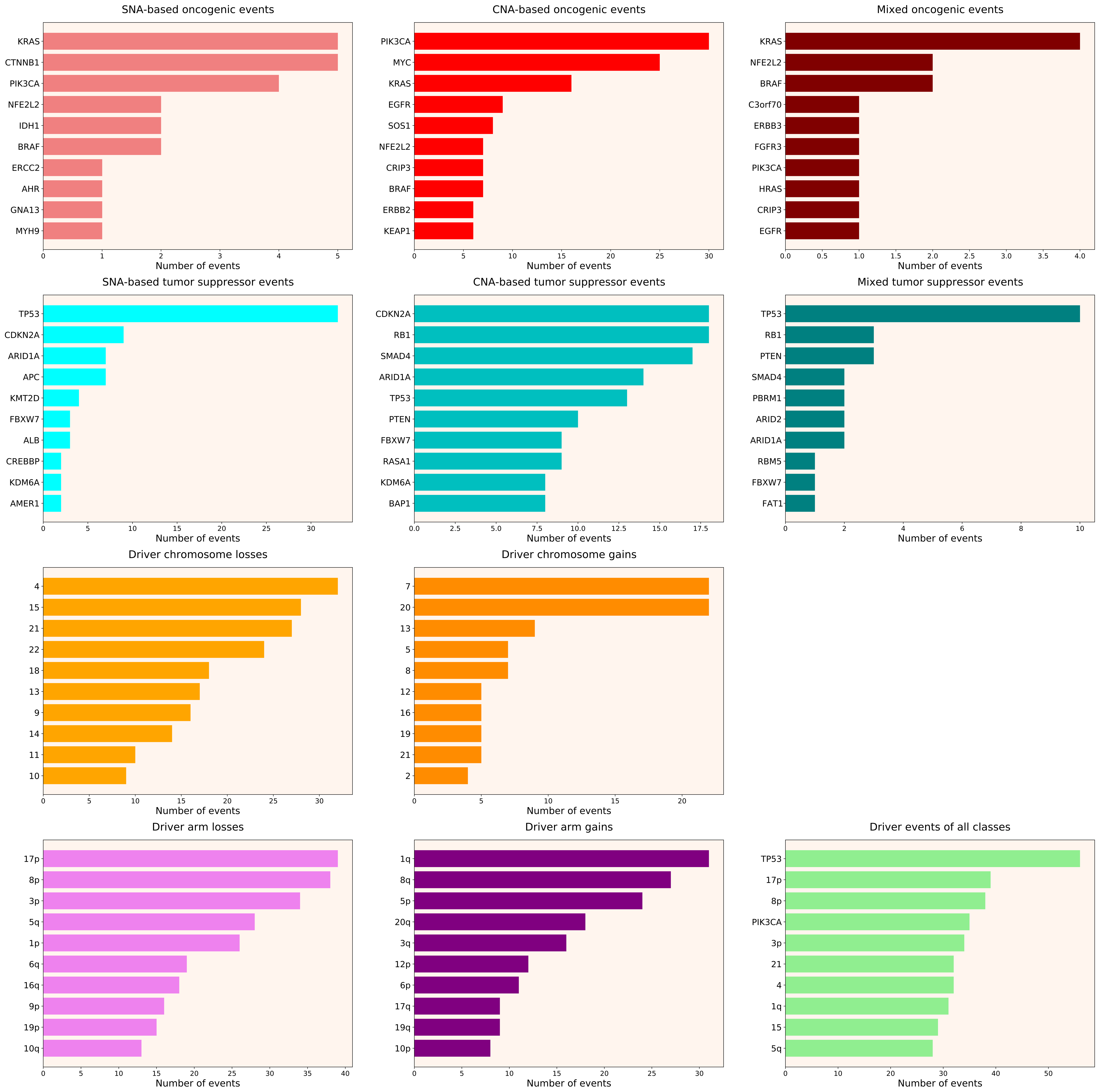

### 2021_8_16_14_9_distribution_events_detailed_males_24.pdf

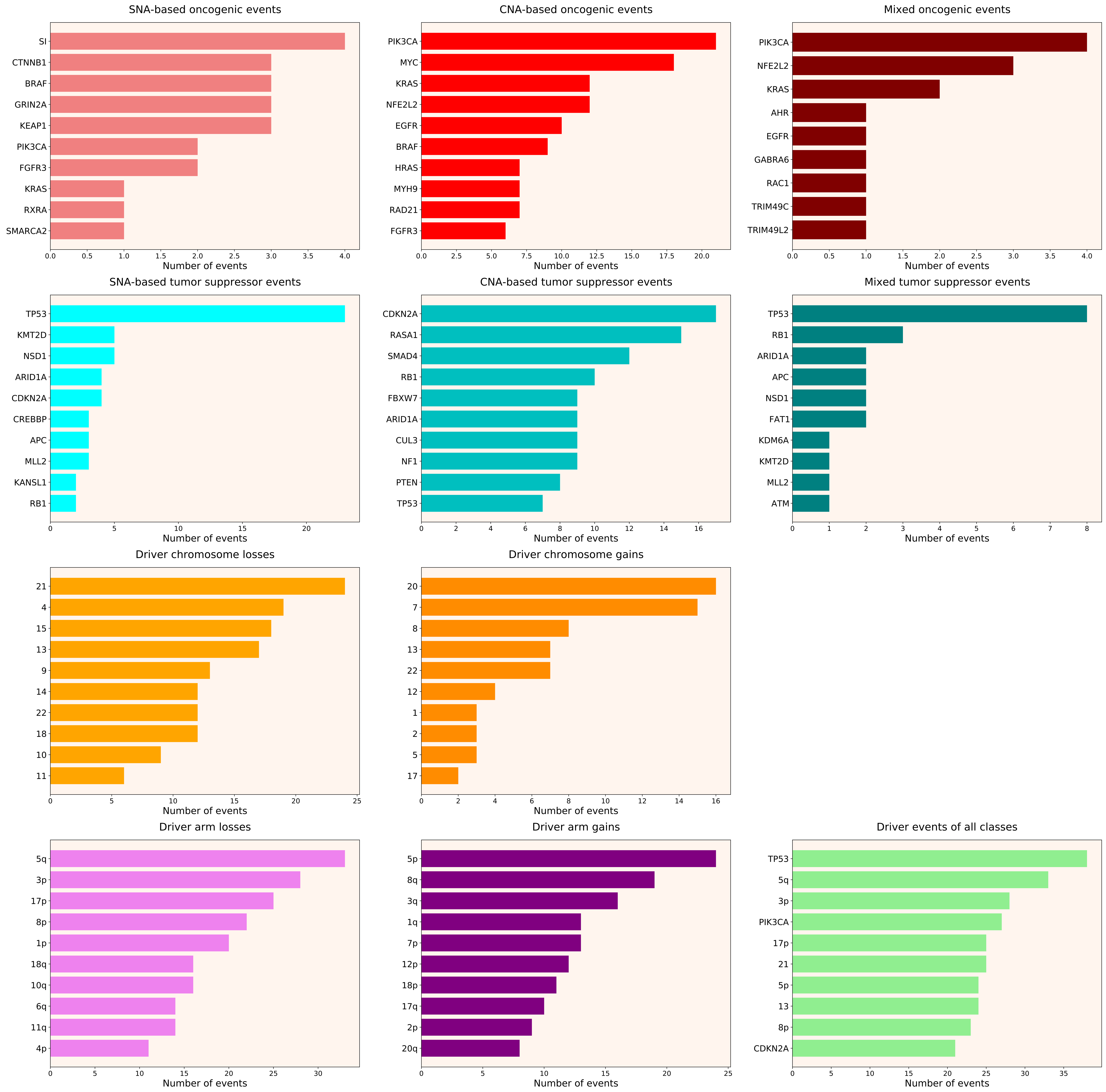

### 2021_8_16_14_9_distribution_events_detailed_males_25.pdf

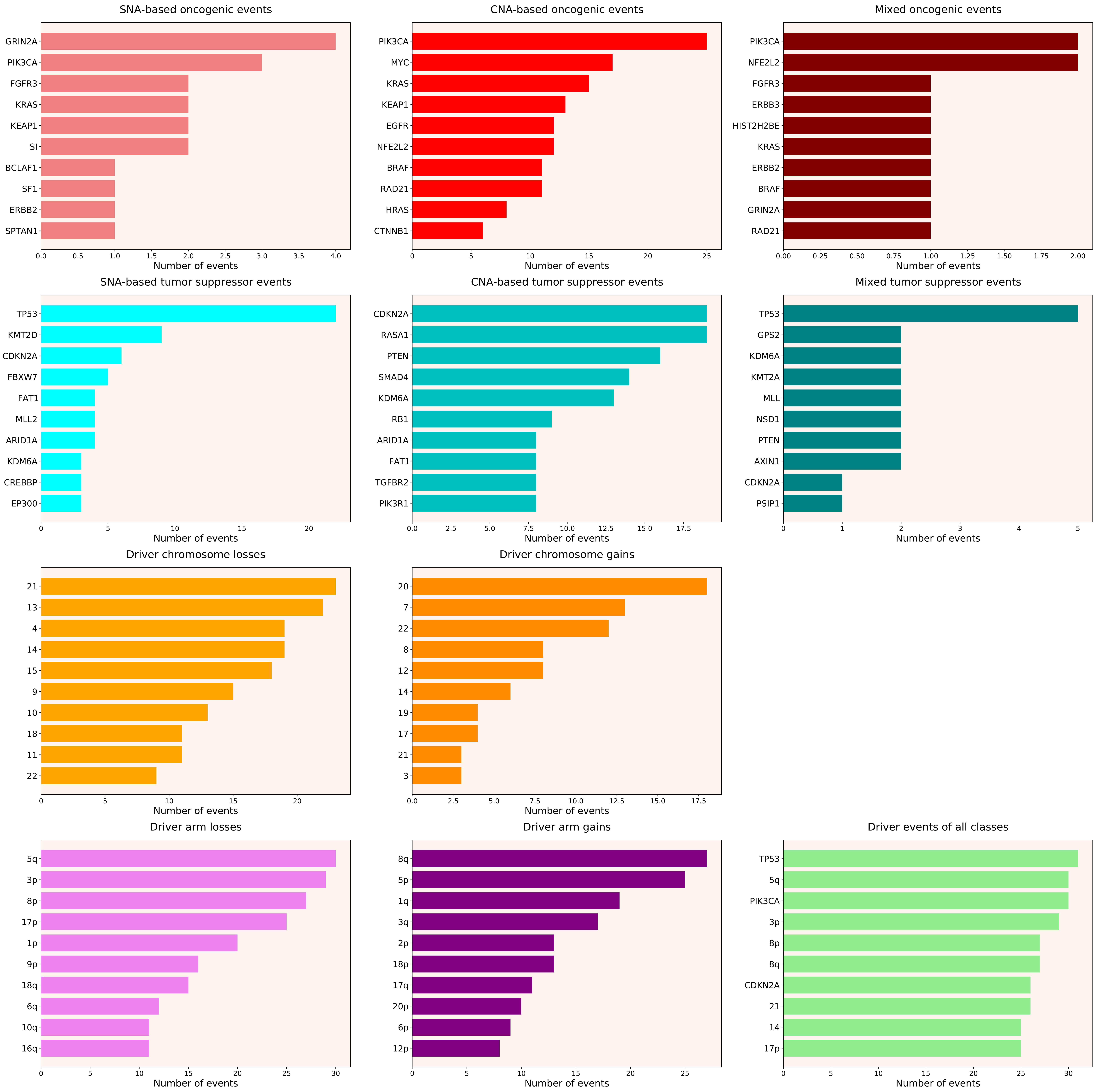

### 2021_8_16_14_9_distribution_events_detailed_males_26.pdf

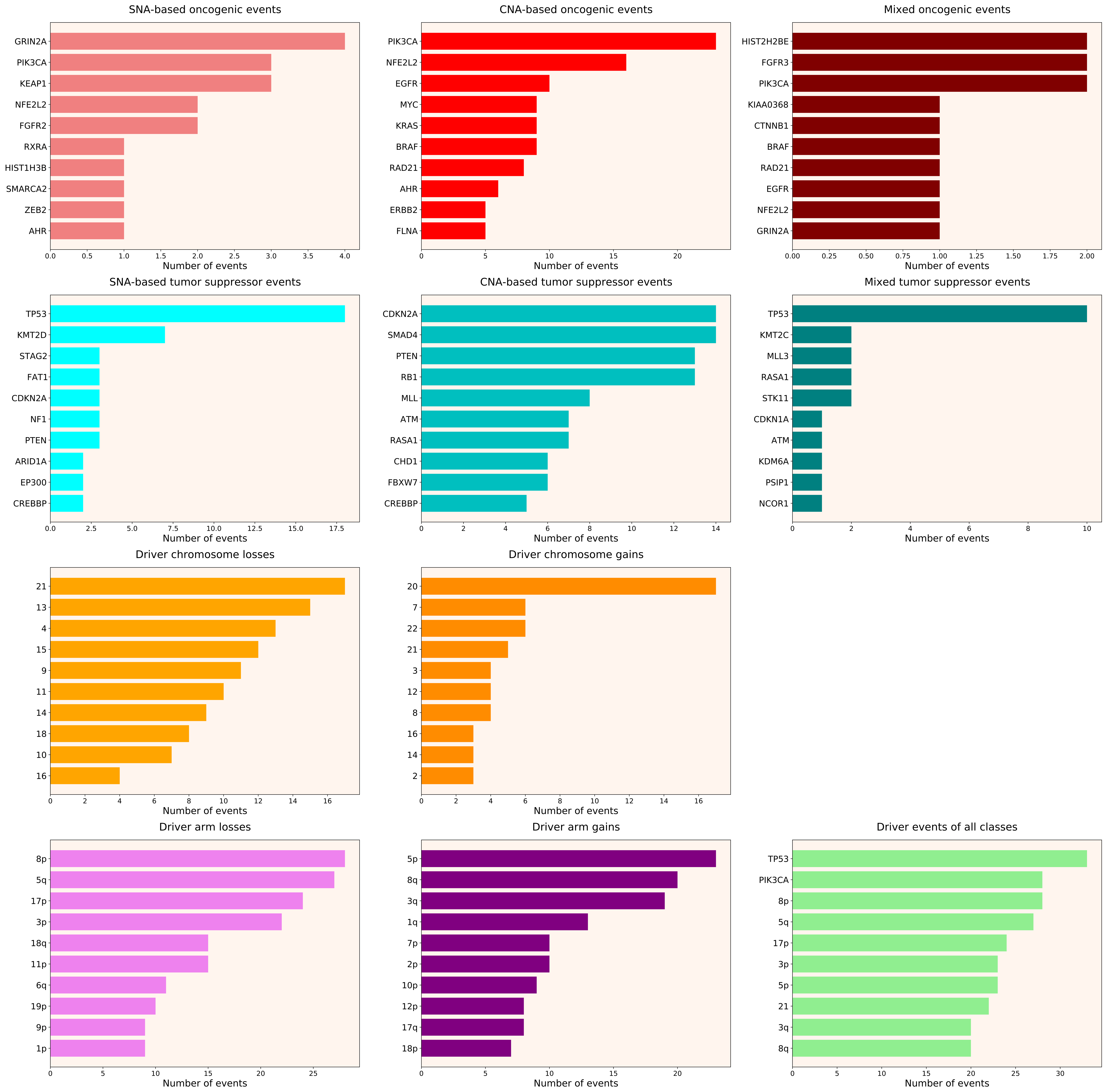

### 2021_8_16_14_9_distribution_events_detailed_males_27.pdf

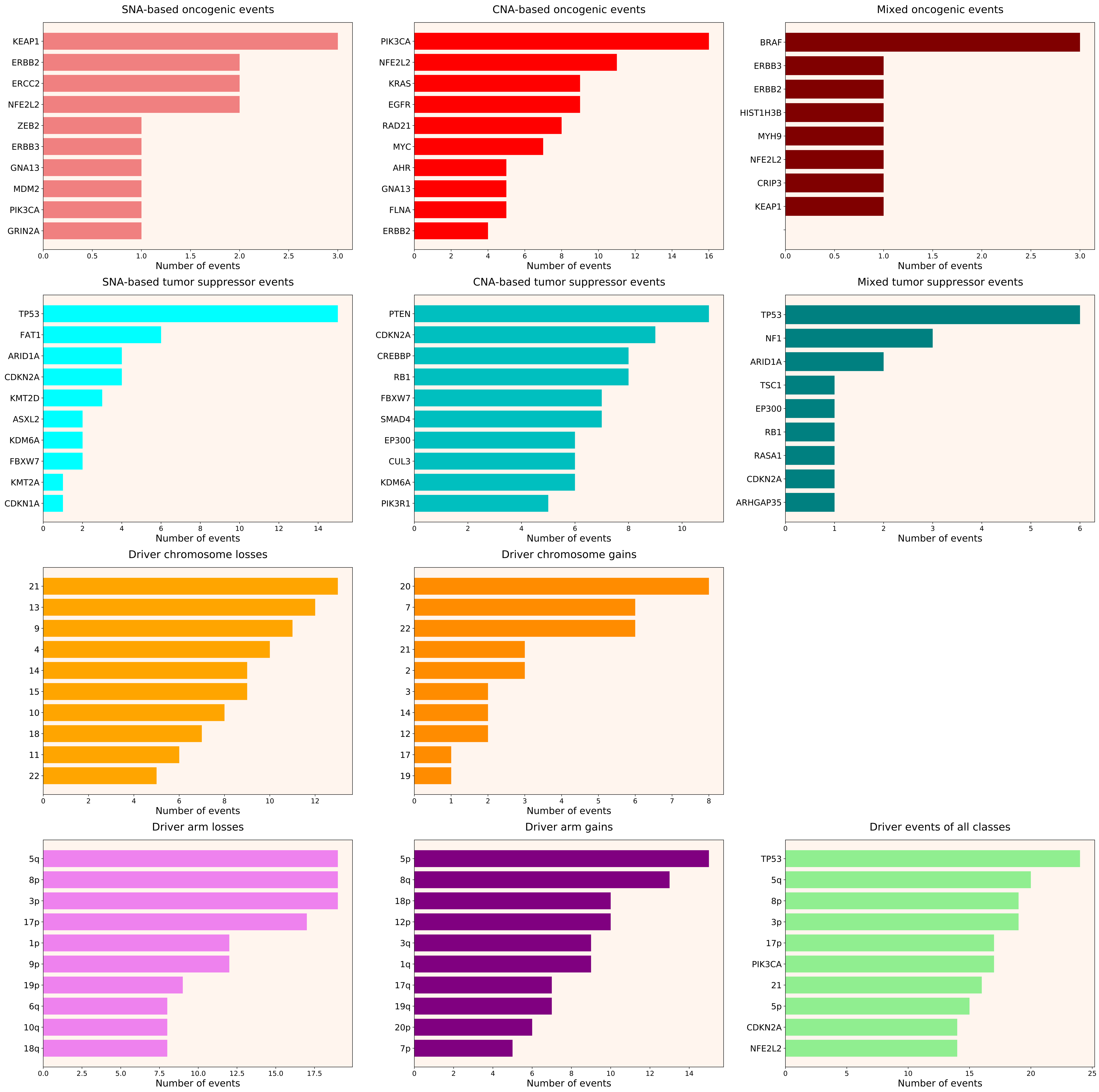

### 2021_8_16_14_9_distribution_events_detailed_males_30.pdf

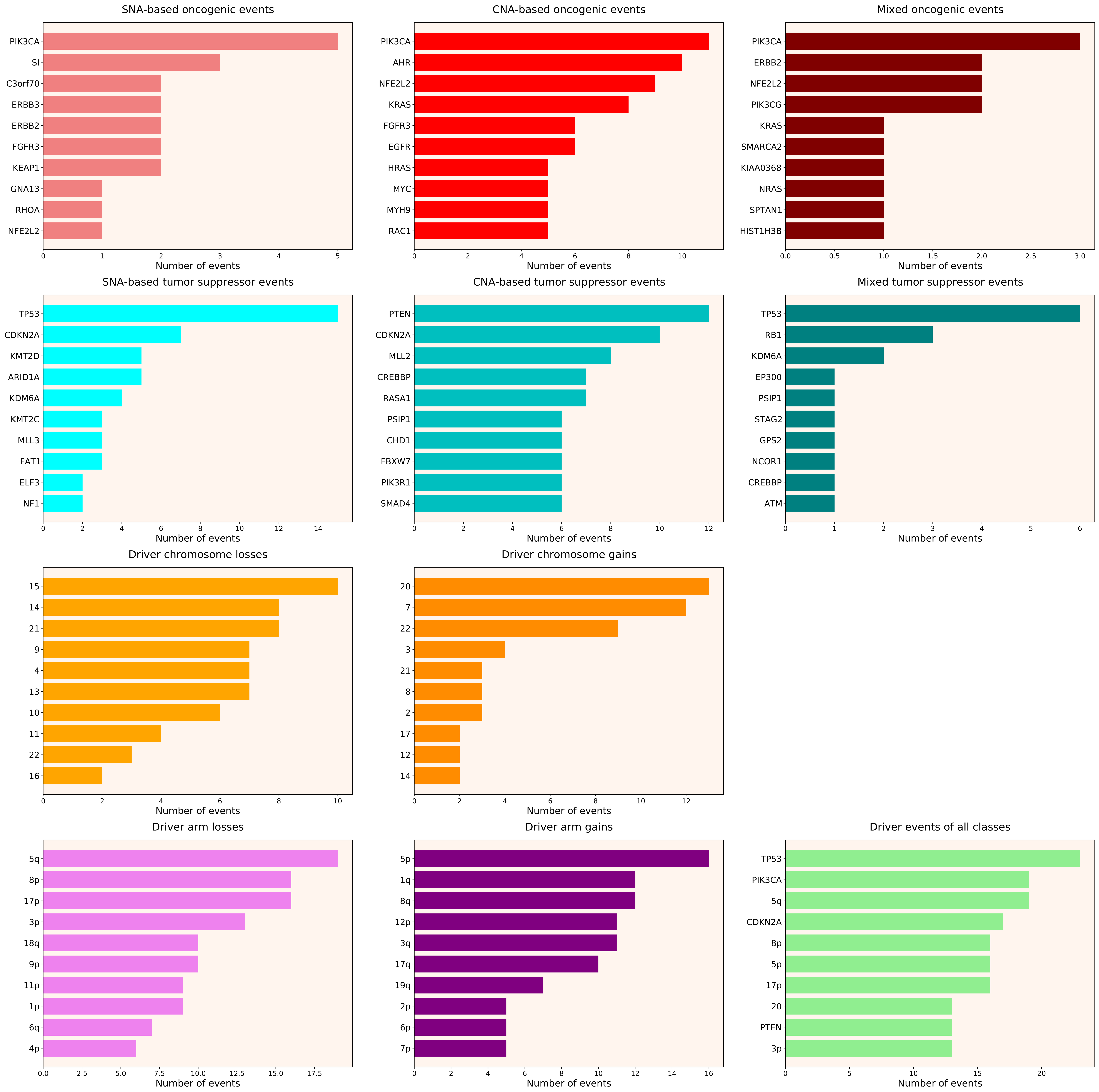

### 2021_8_16_14_9_distribution_events_detailed_males_31.pdf

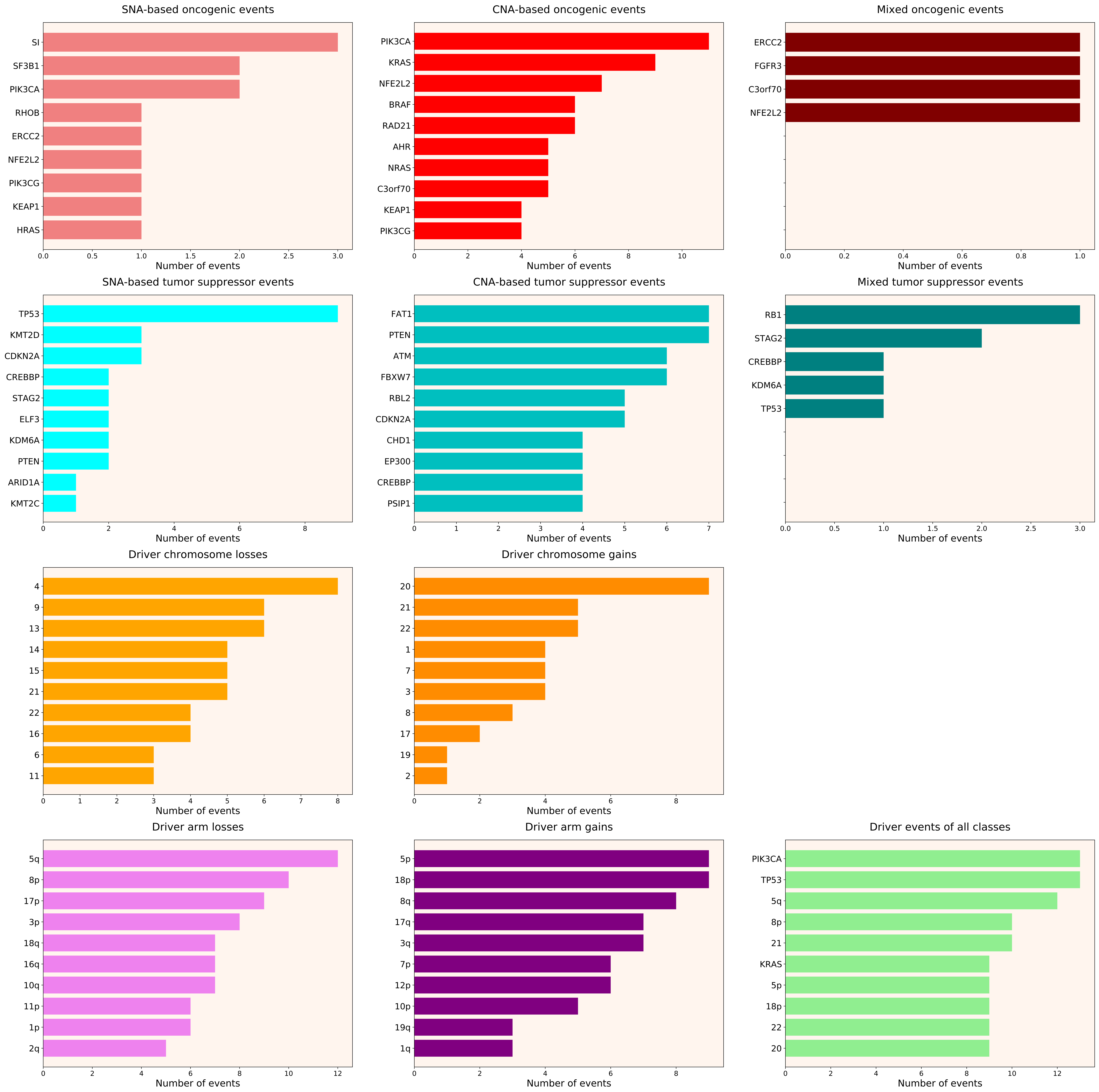

### 2021_8_16_14_9_distribution_events_detailed_males_32.pdf

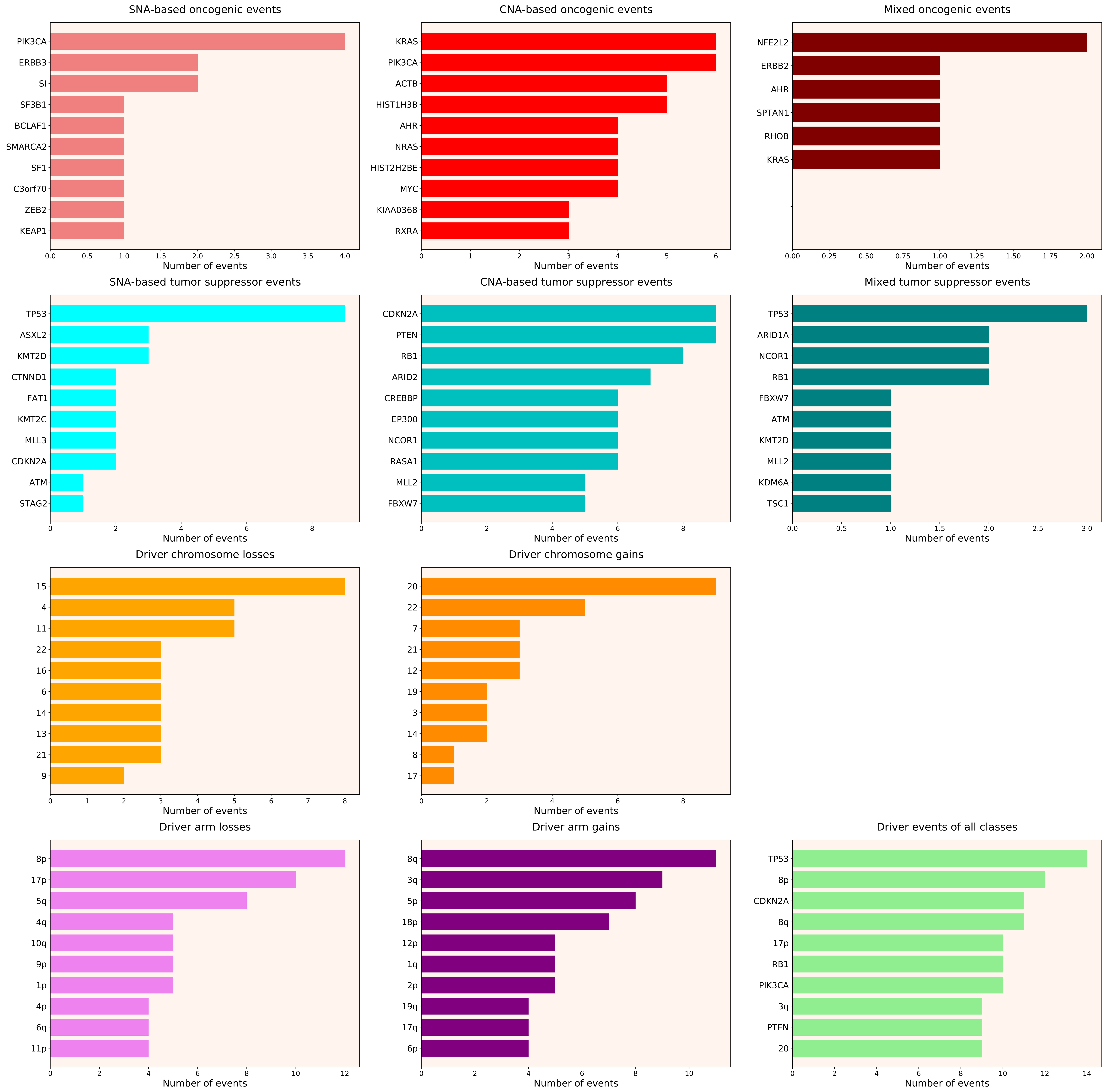

### 2021_8_16_14_9_distribution_events_detailed_males_33.pdf

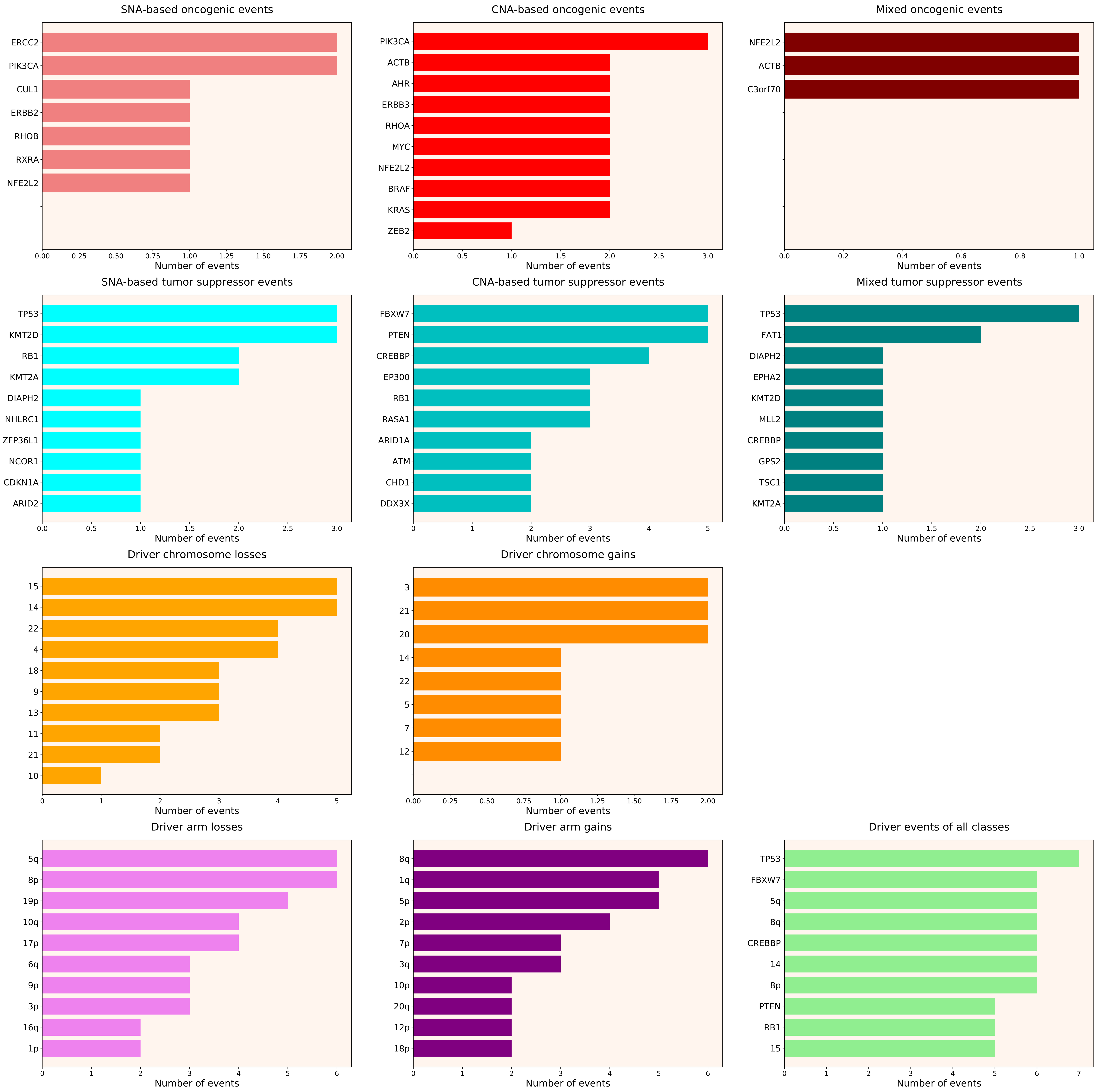

### 2021_8_16_14_9_distribution_events_detailed_males_34.pdf

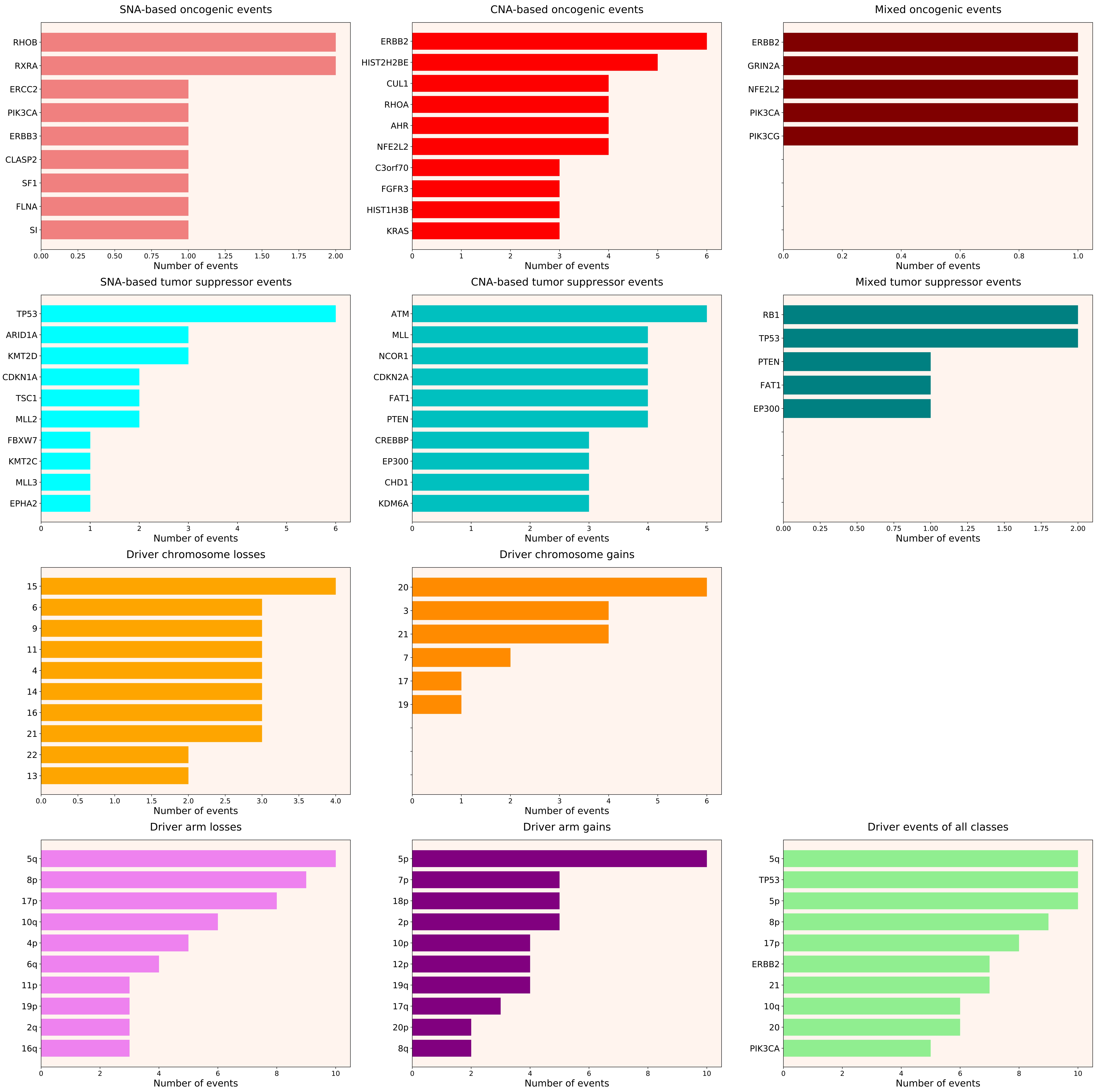

### 2021_8_16_14_9_distribution_events_detailed_NDSI.pdf

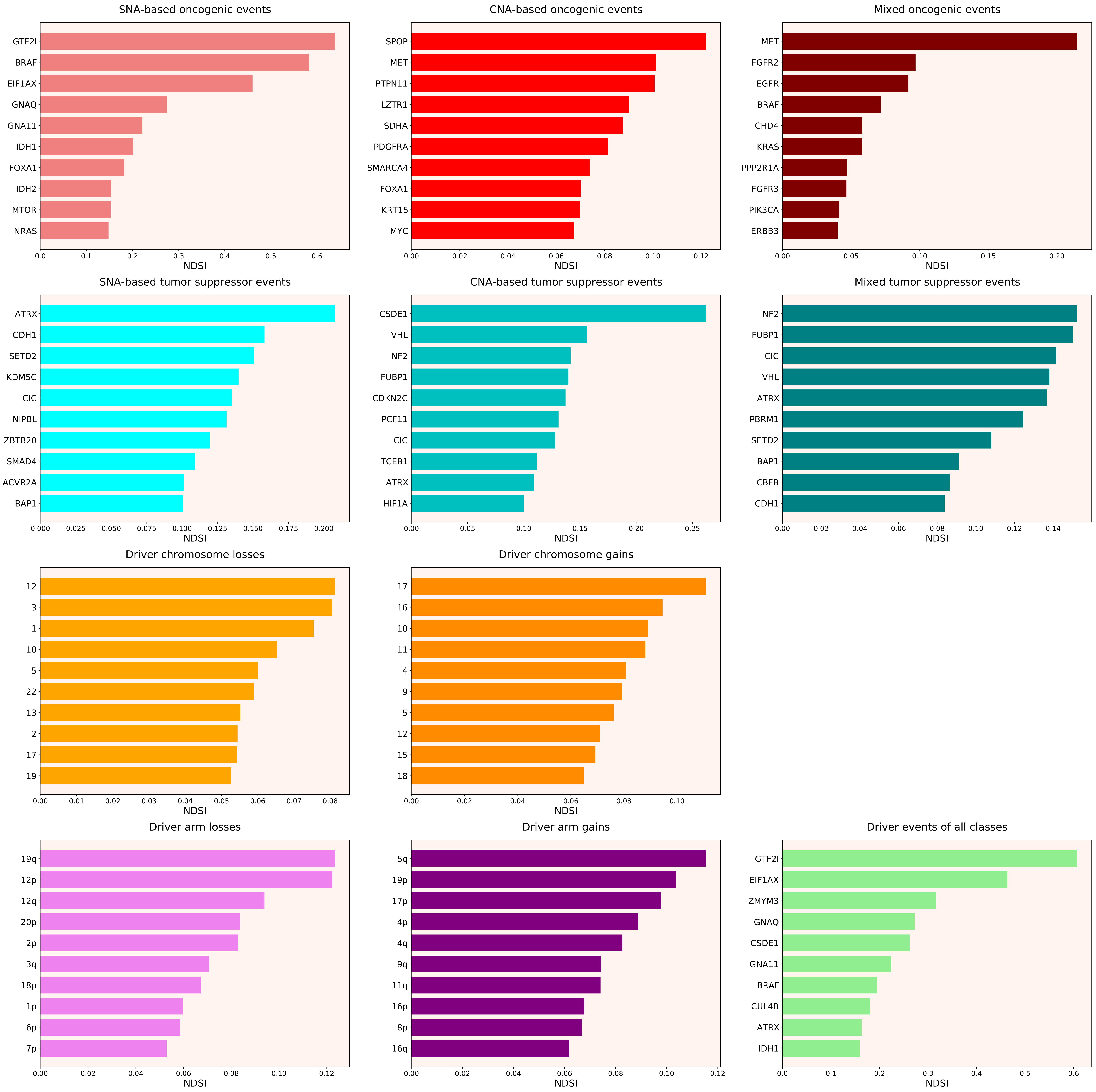

### 2021_8_16_14_9_distribution_events_detailed_NDSI_females.pdf

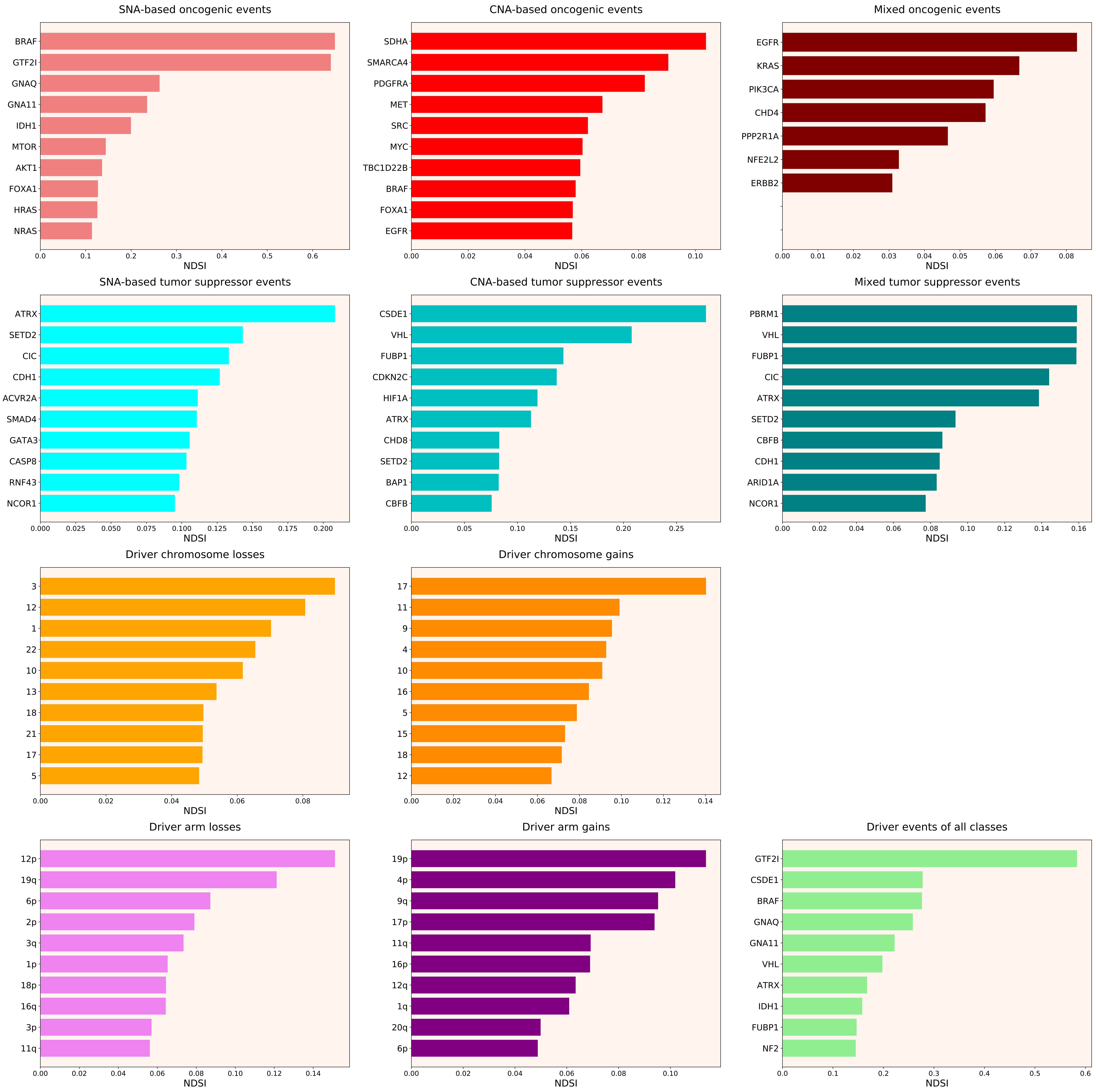

### 2021_8_16_14_9_distribution_events_detailed_NDSI_males.pdf

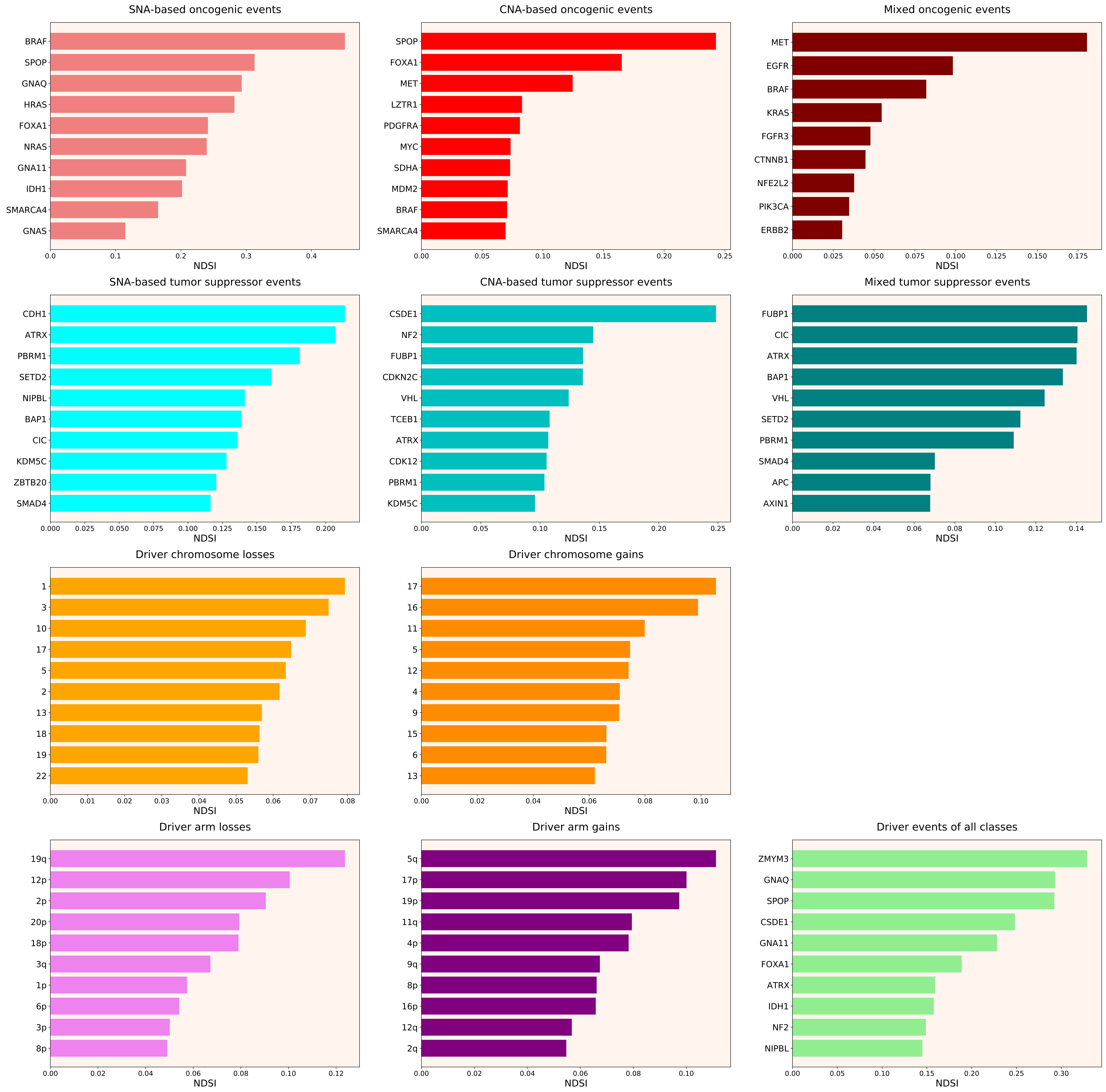

### 2021_8_16_14_9_ESCA.pdf

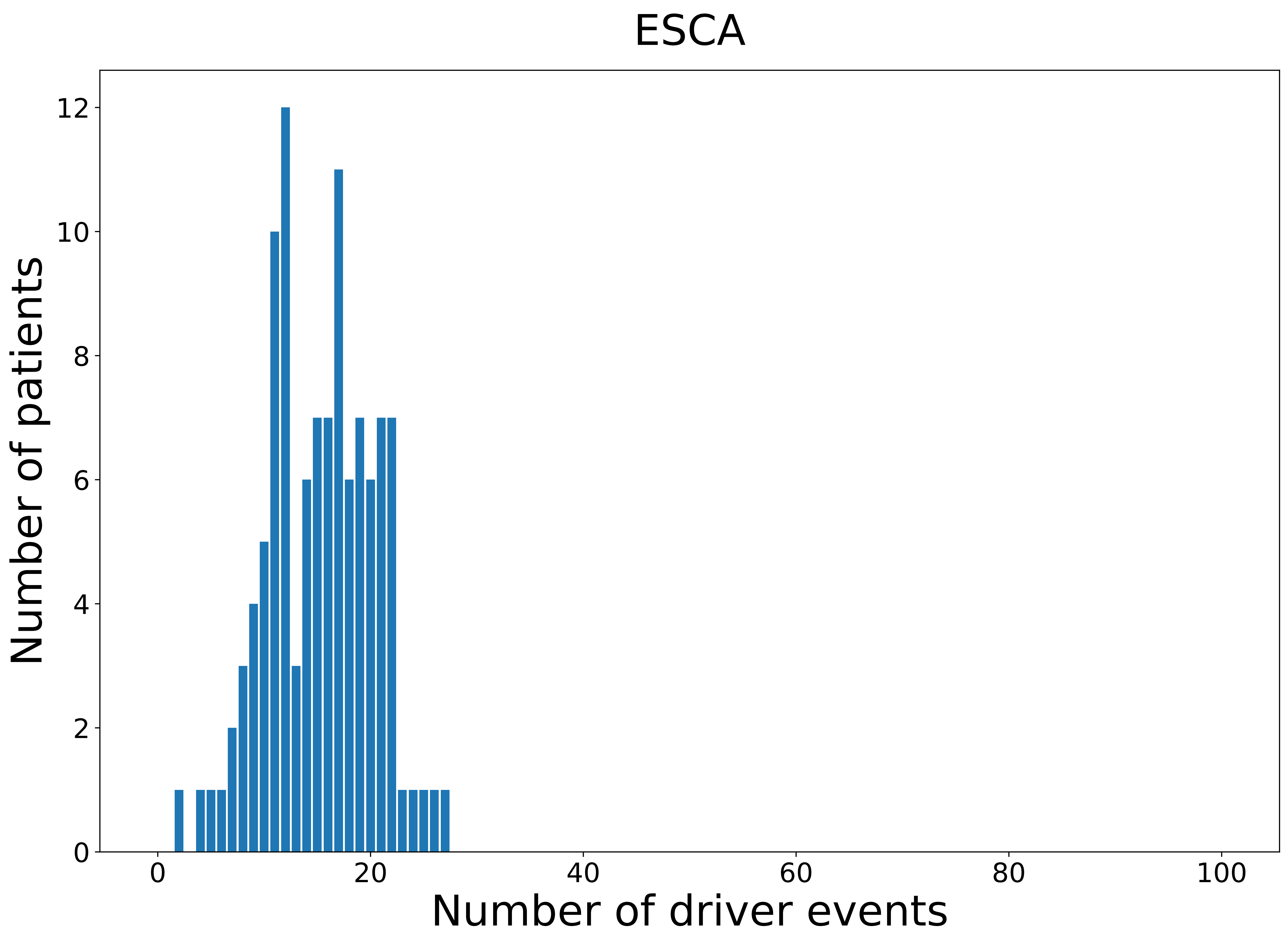

### 2021_8_16_14_9_ESCA_MALE.pdf

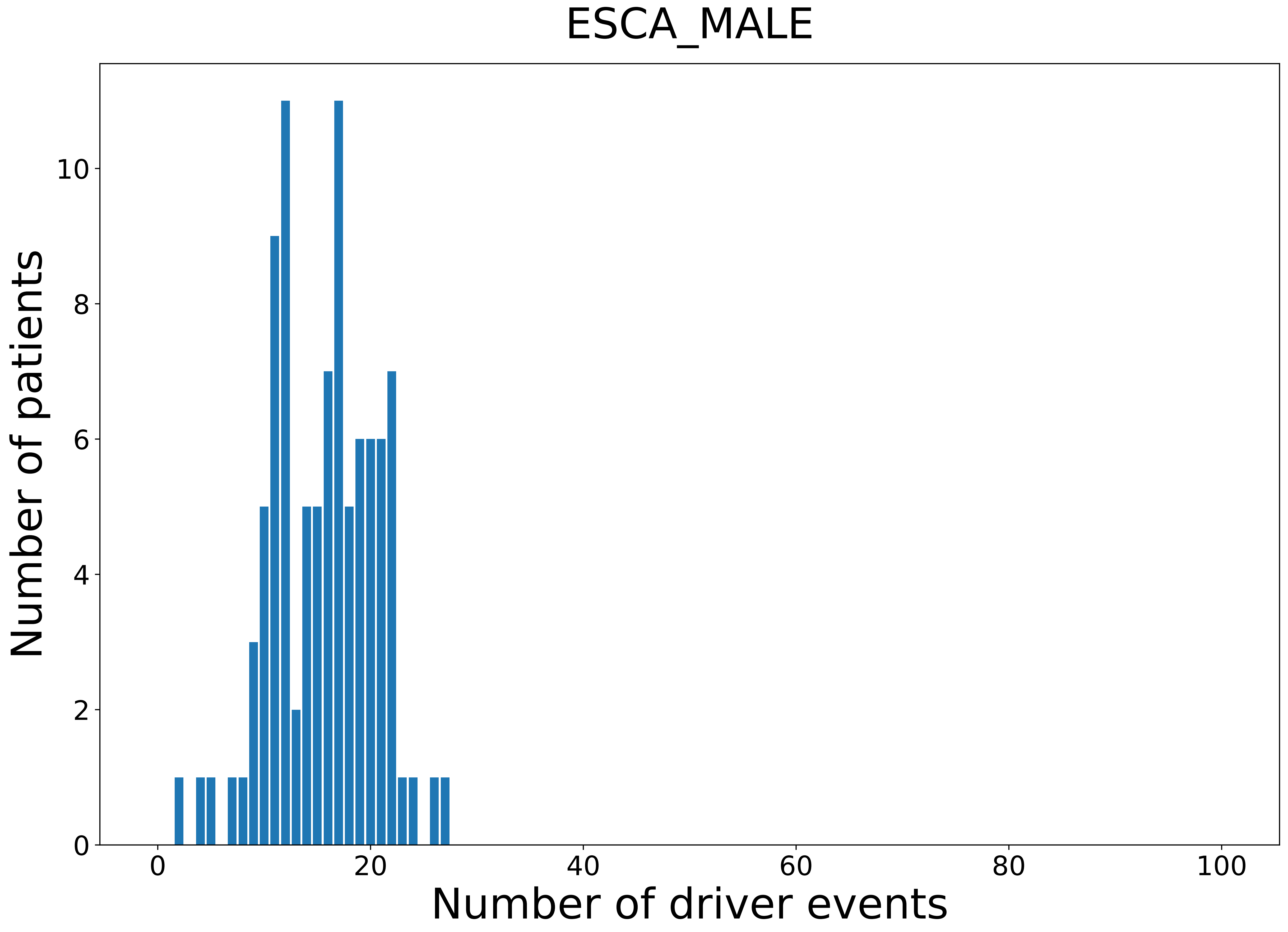

### 2021_8_16_14_9_GBM.pdf

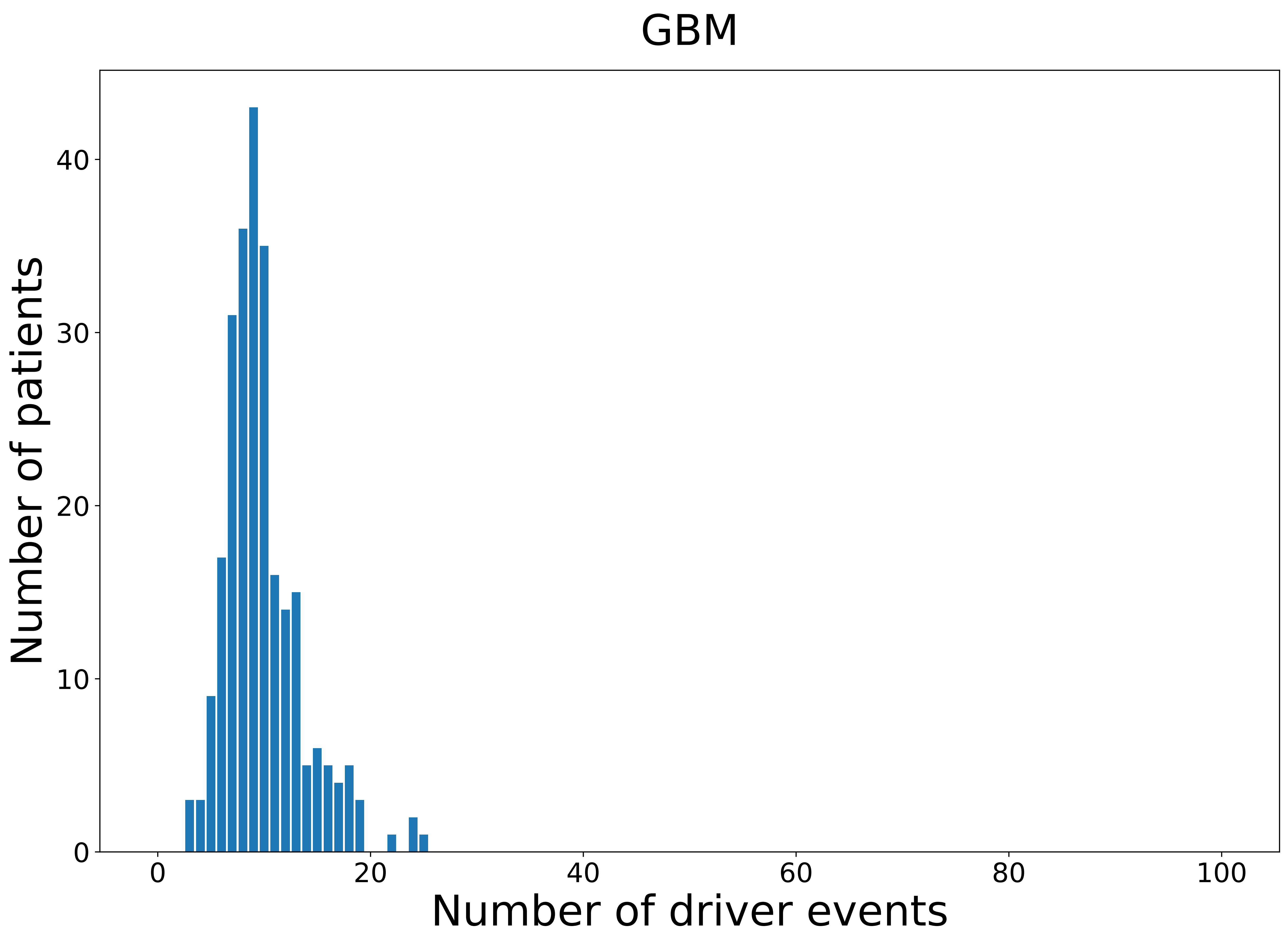

### 2021_8_16_14_9_GBM_FEMALE.pdf

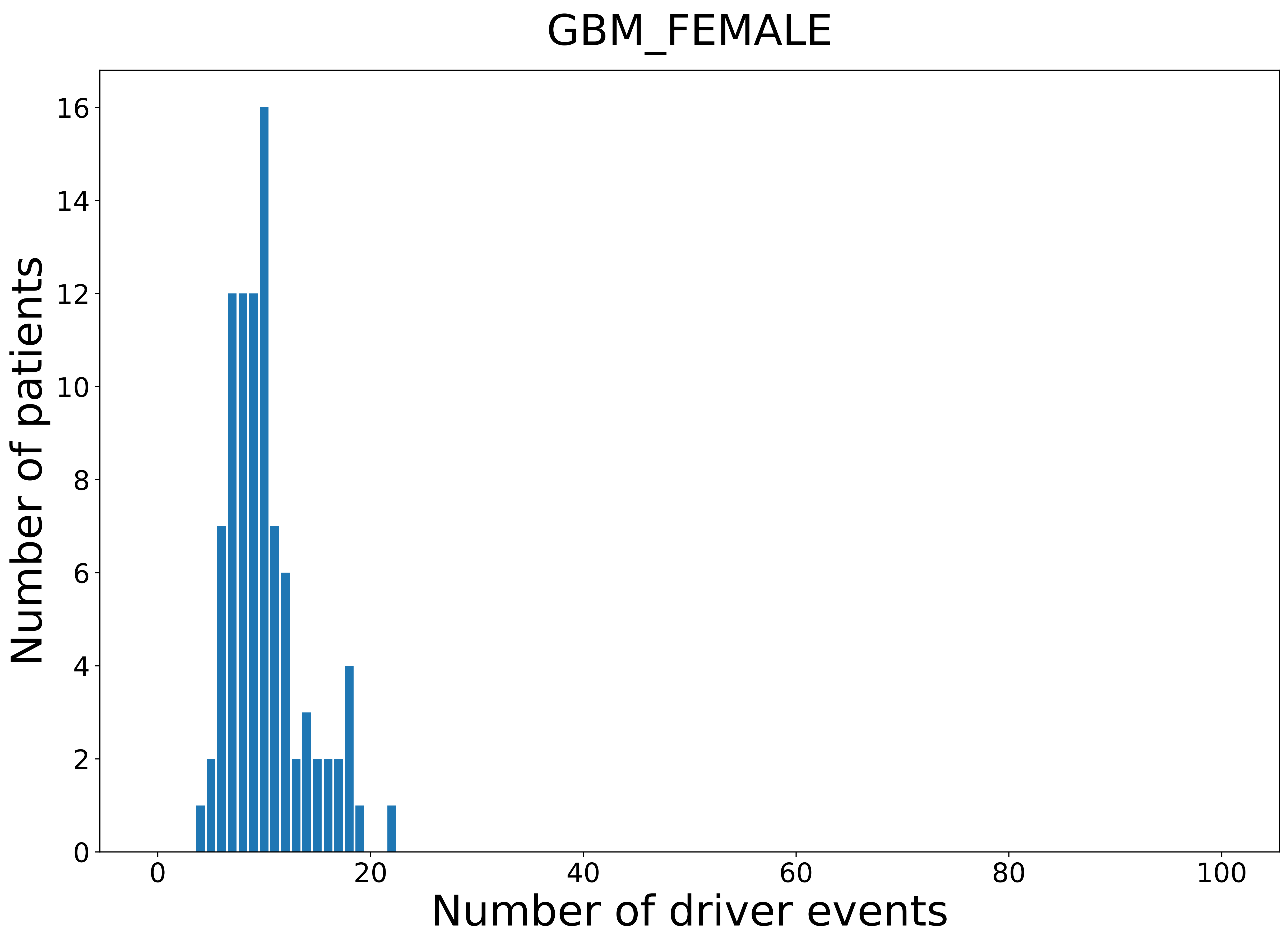

### 2021_8_16_14_9_HNSC.pdf

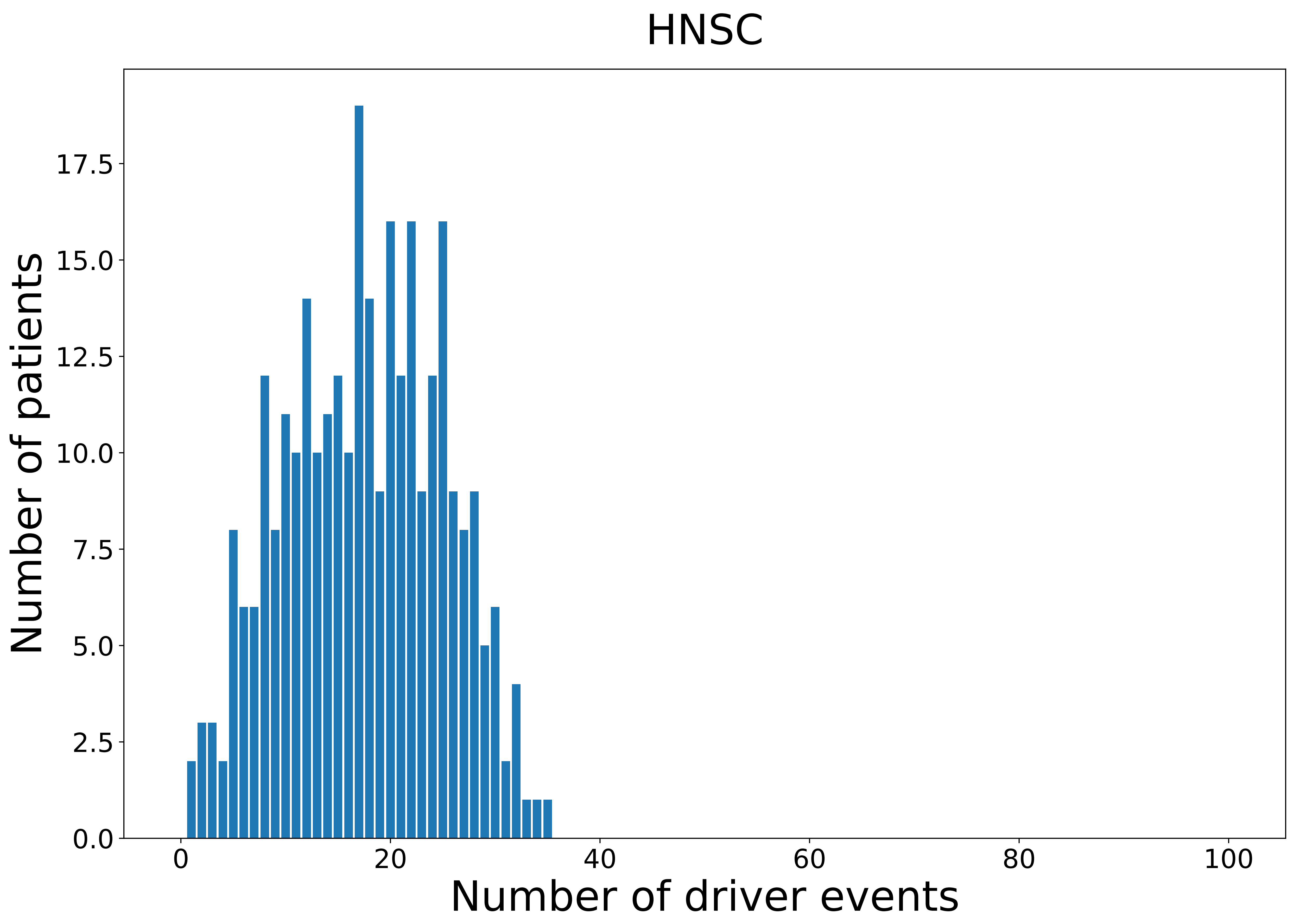

### 2021_8_16_14_9_HNSC_FEMALE.pdf

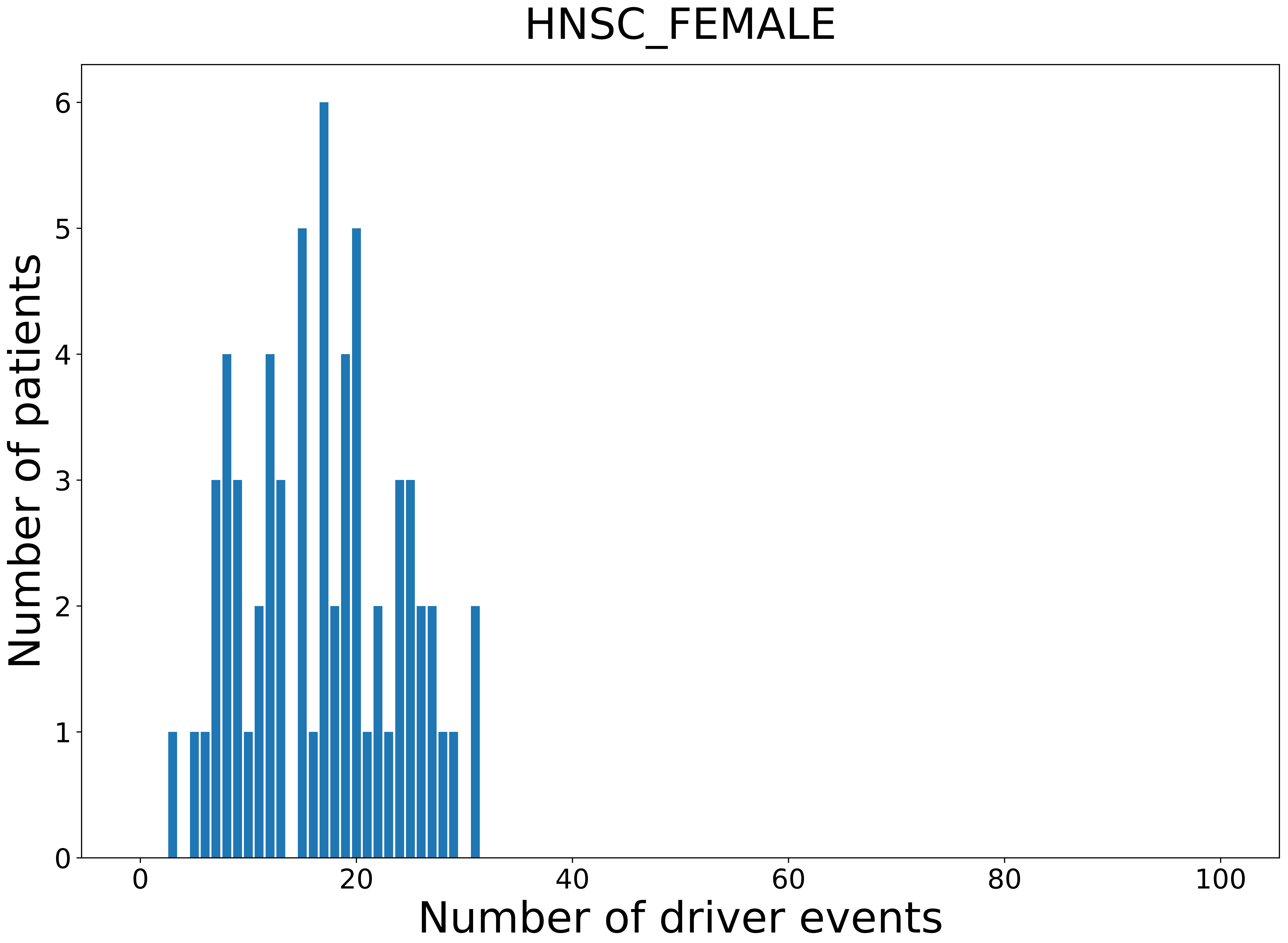

### 2021_8_16_14_9_HNSC_MALE.pdf

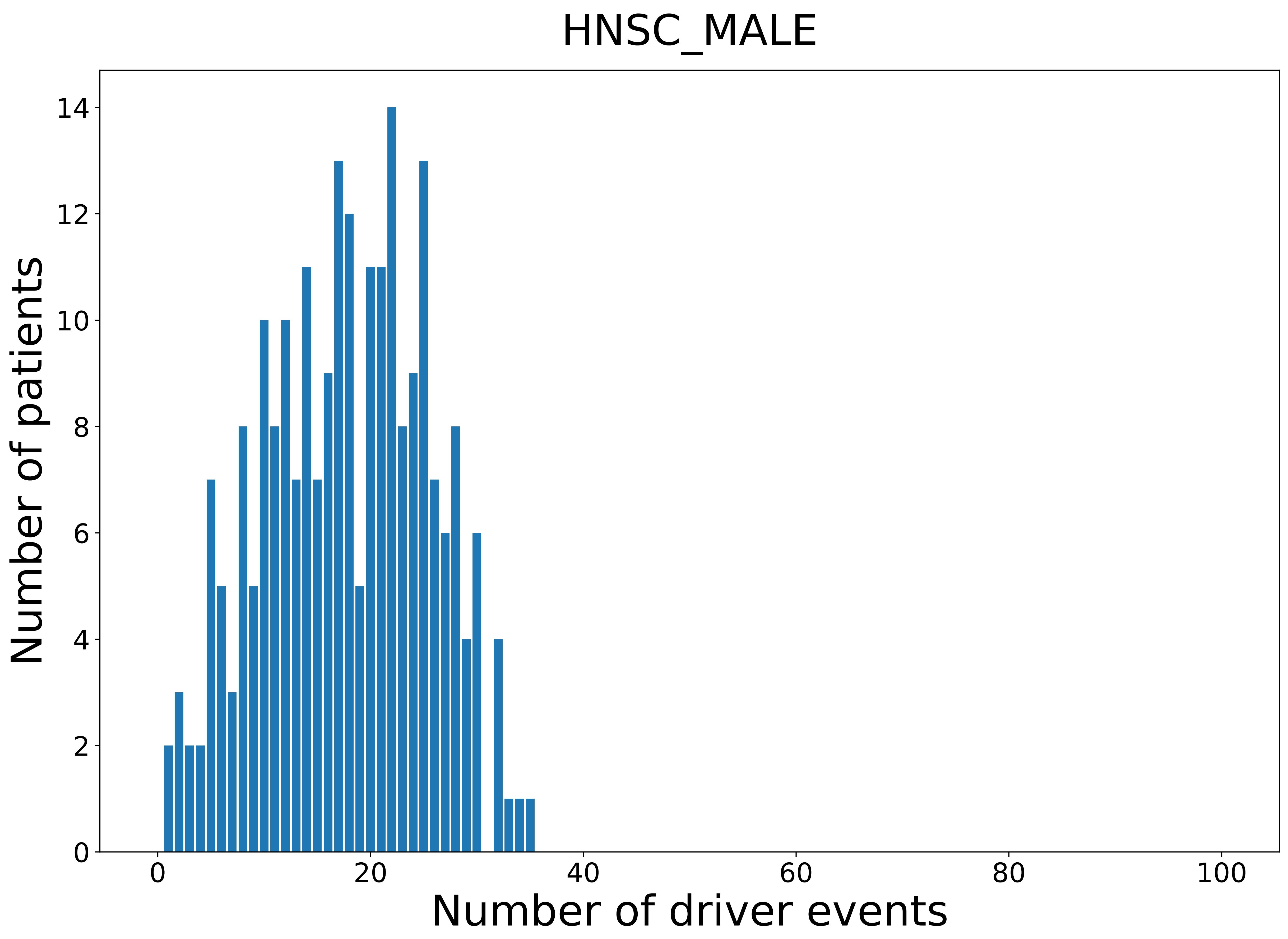

### 2021_8_16_14_9_KICH_MALE.pdf

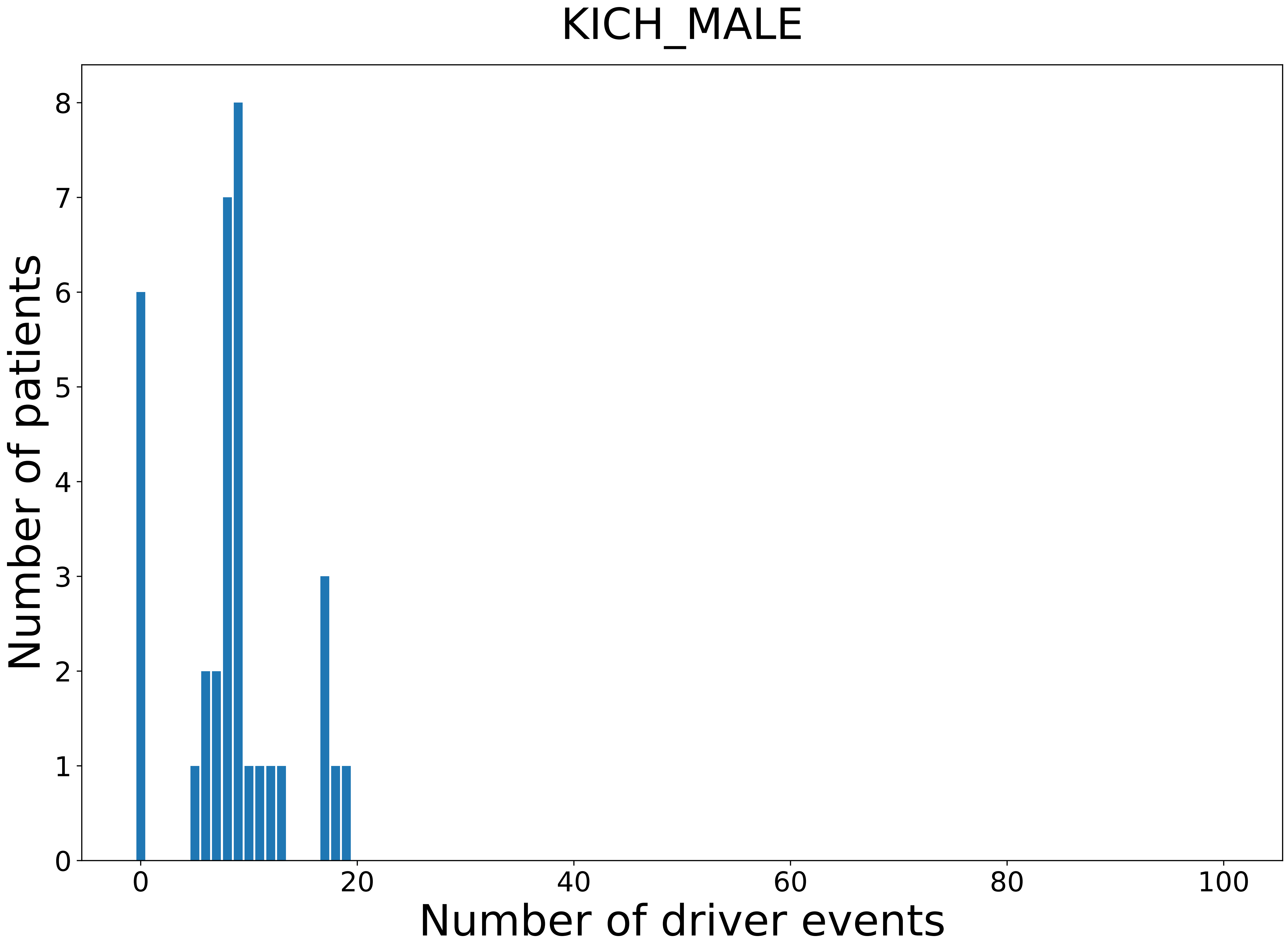

### 2021_8_16_14_9_KIRC.pdf

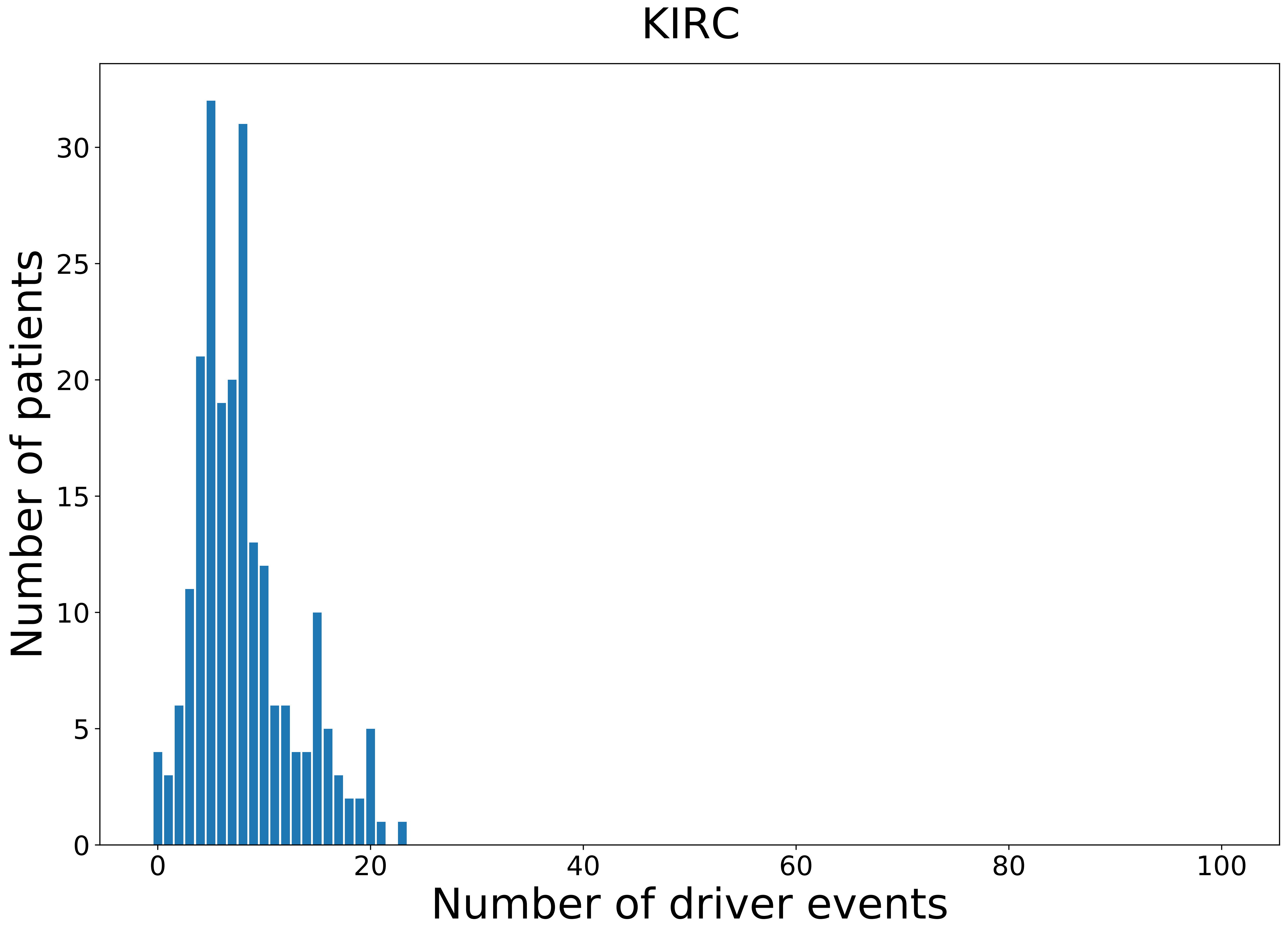

### 2021_8_16_14_9_KIRC_MALE.pdf

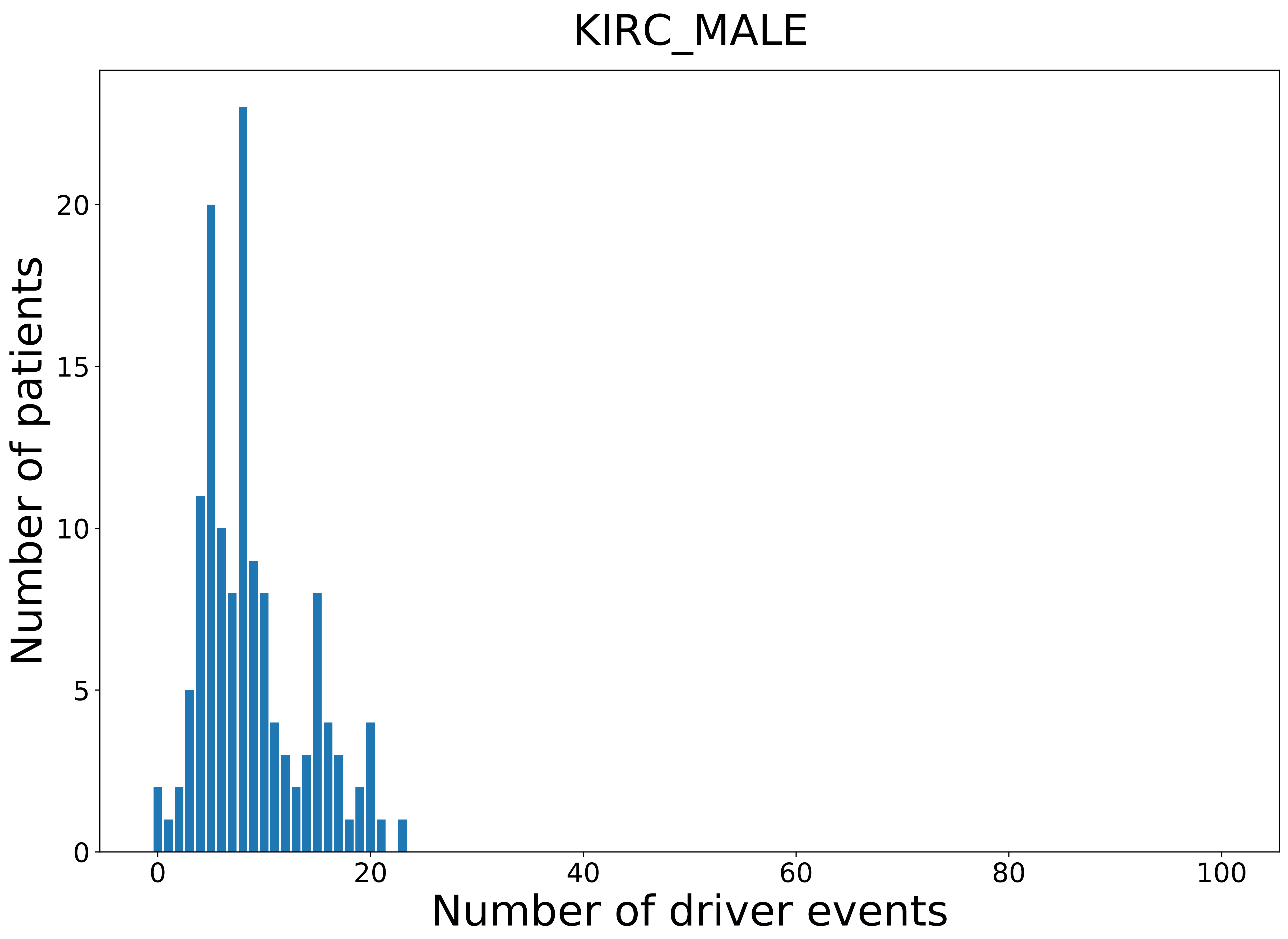

### 2021_8_16_14_9_KIRP_MALE.pdf

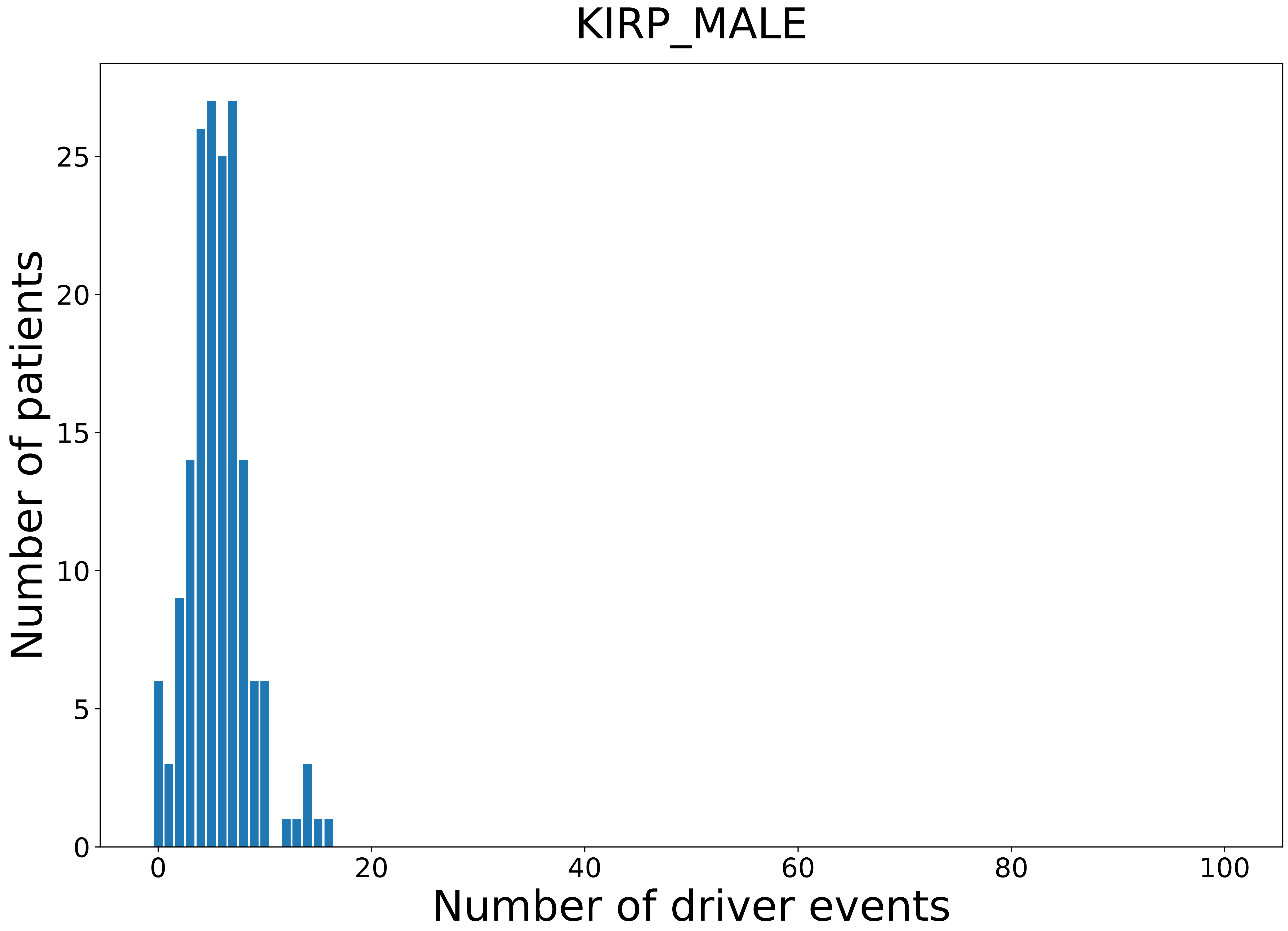

### 2021_8_16_14_9_LGG.pdf

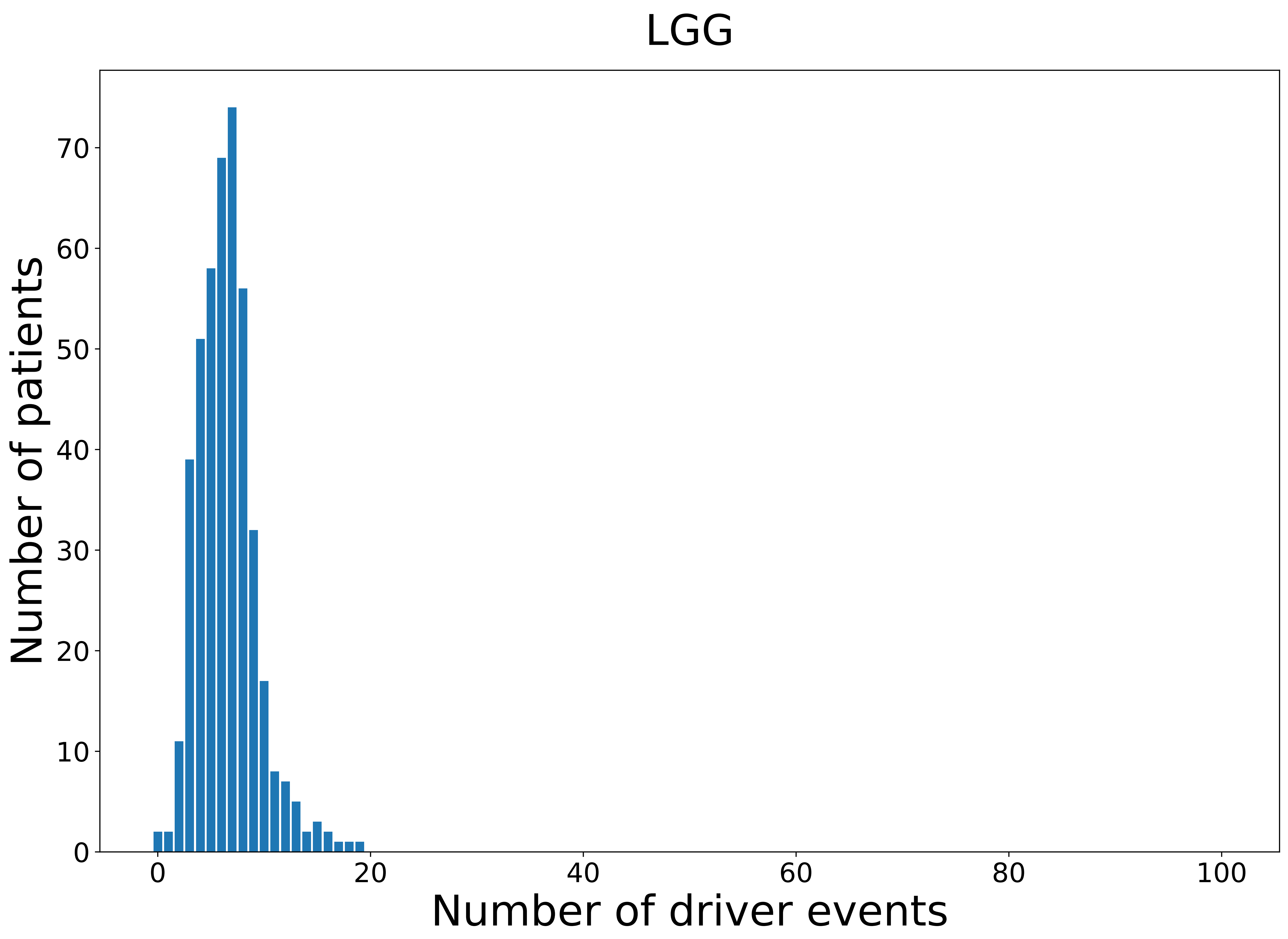

### 2021_8_16_14_9_LGG_FEMALE.pdf

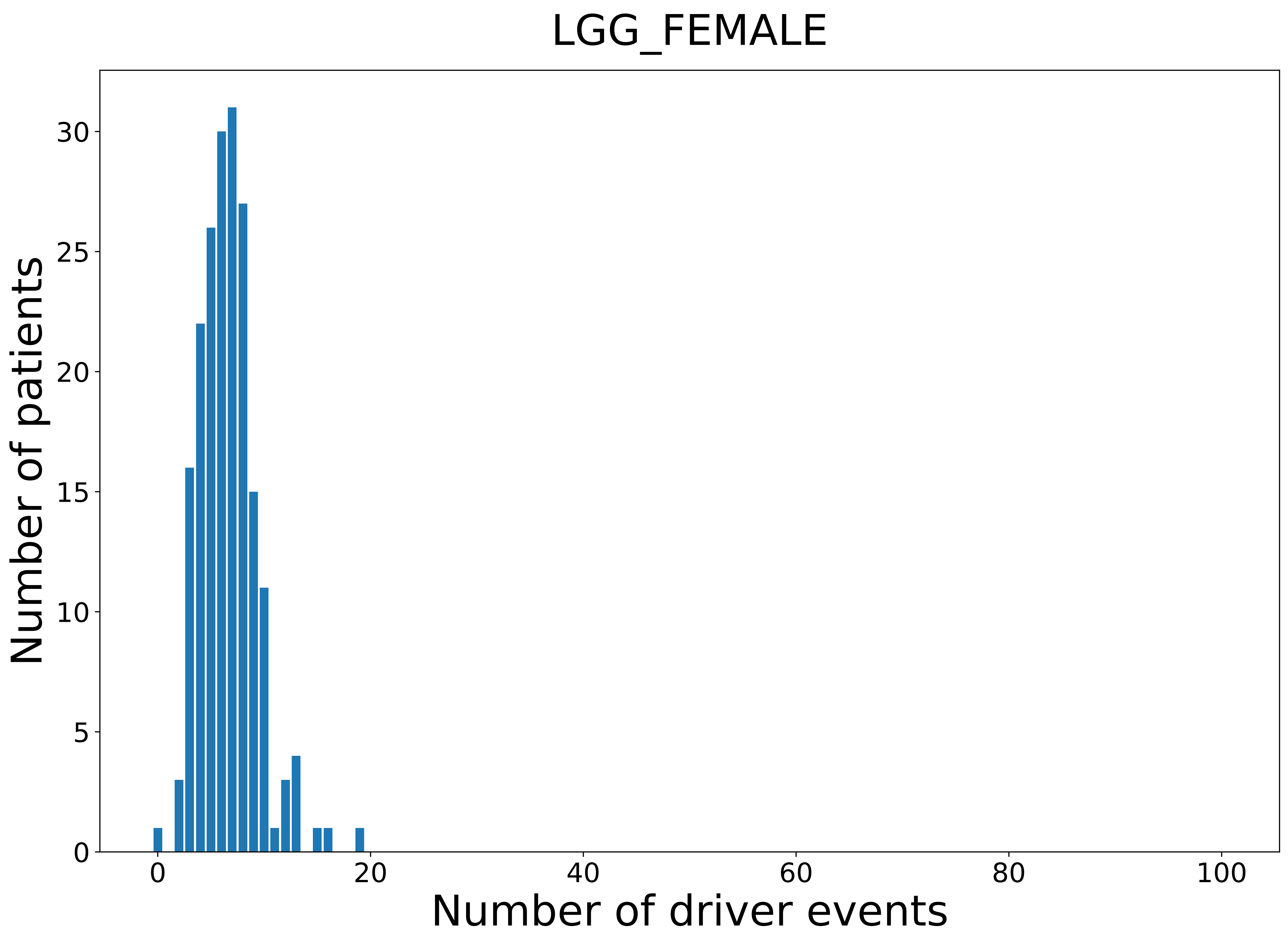

### 2021_8_16_14_9_LIHC.pdf

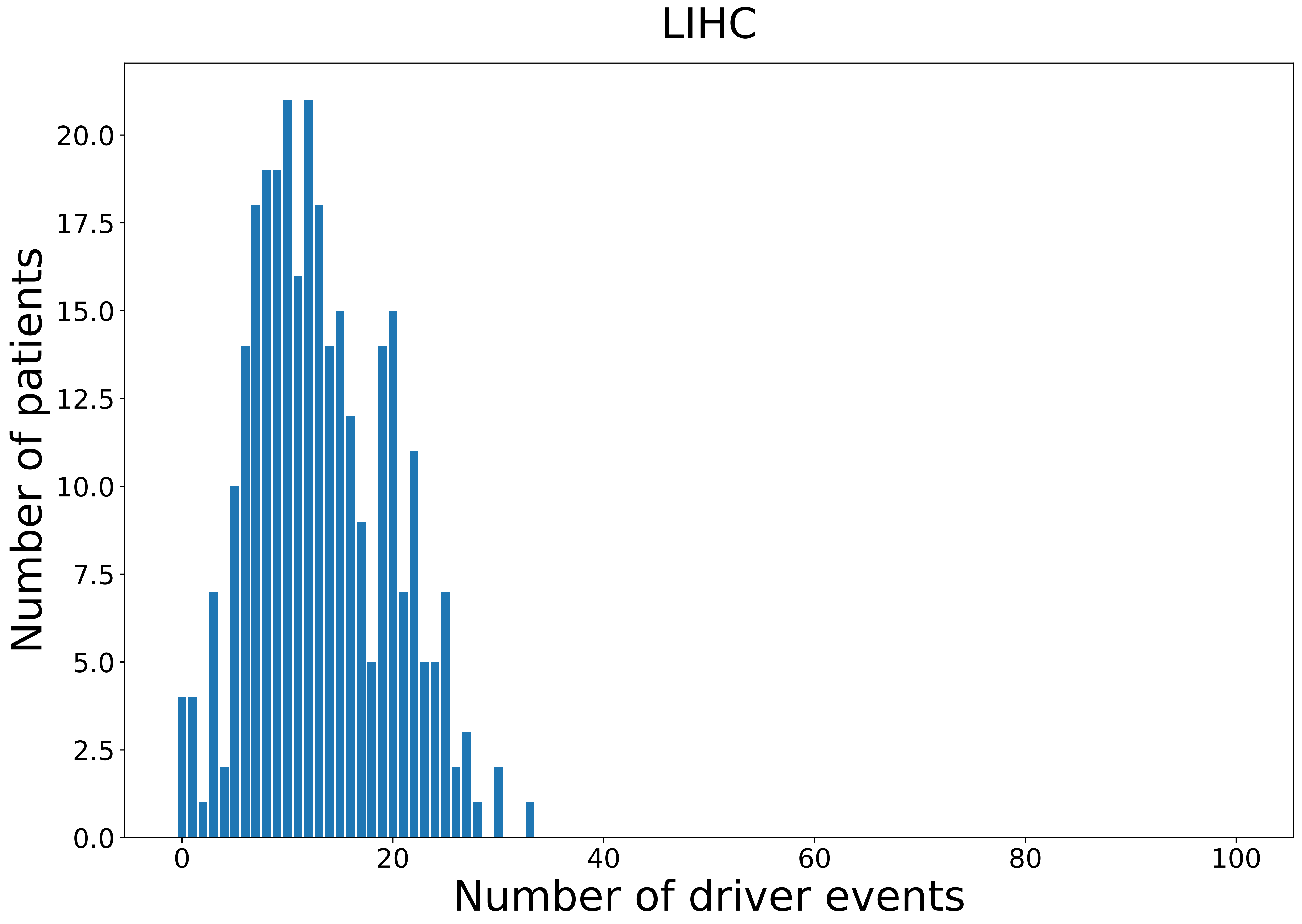

### 2021_8_16_14_9_LIHC_MALE.pdf

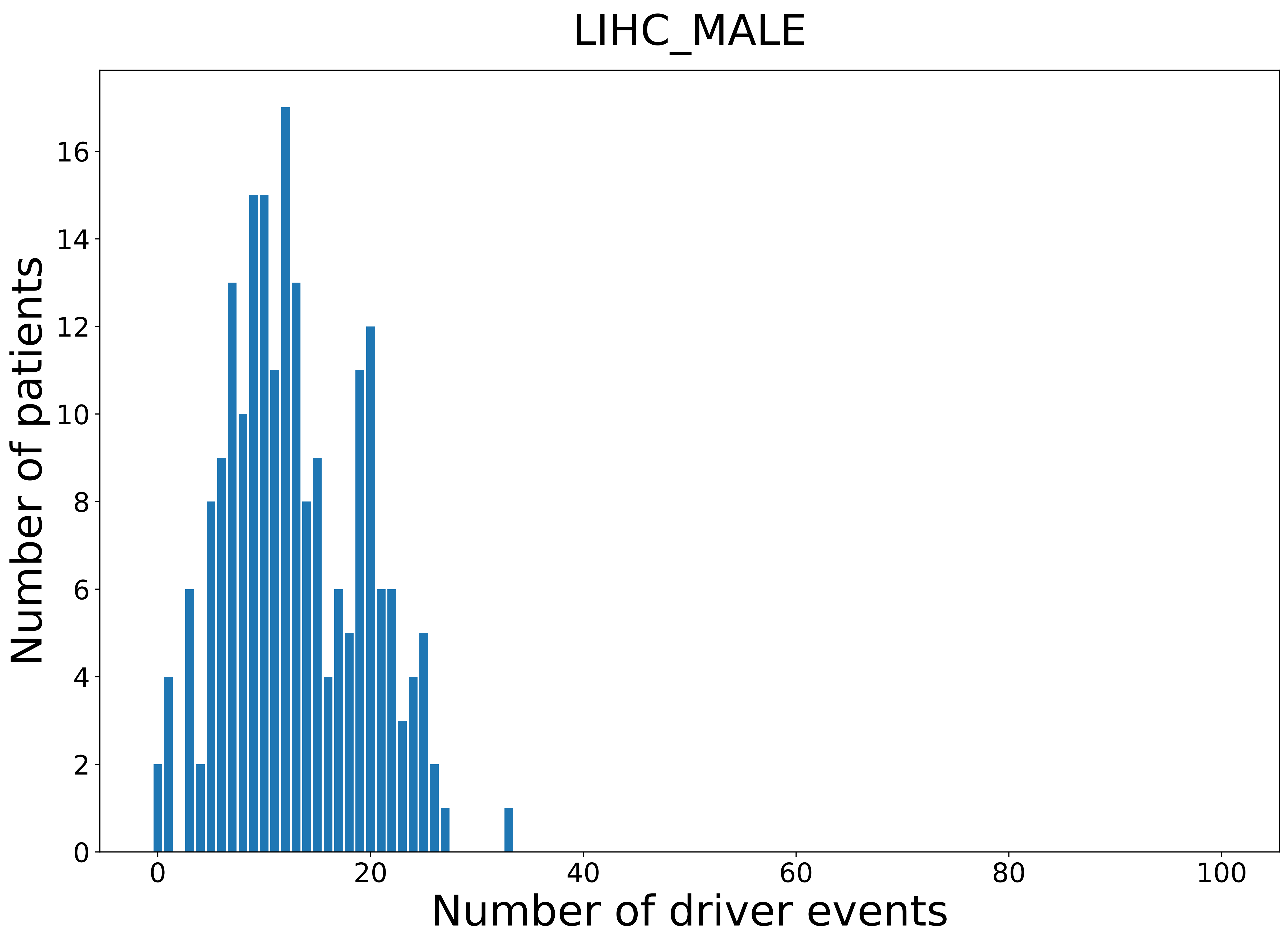

### 2021_8_16_14_9_LUAD.pdf

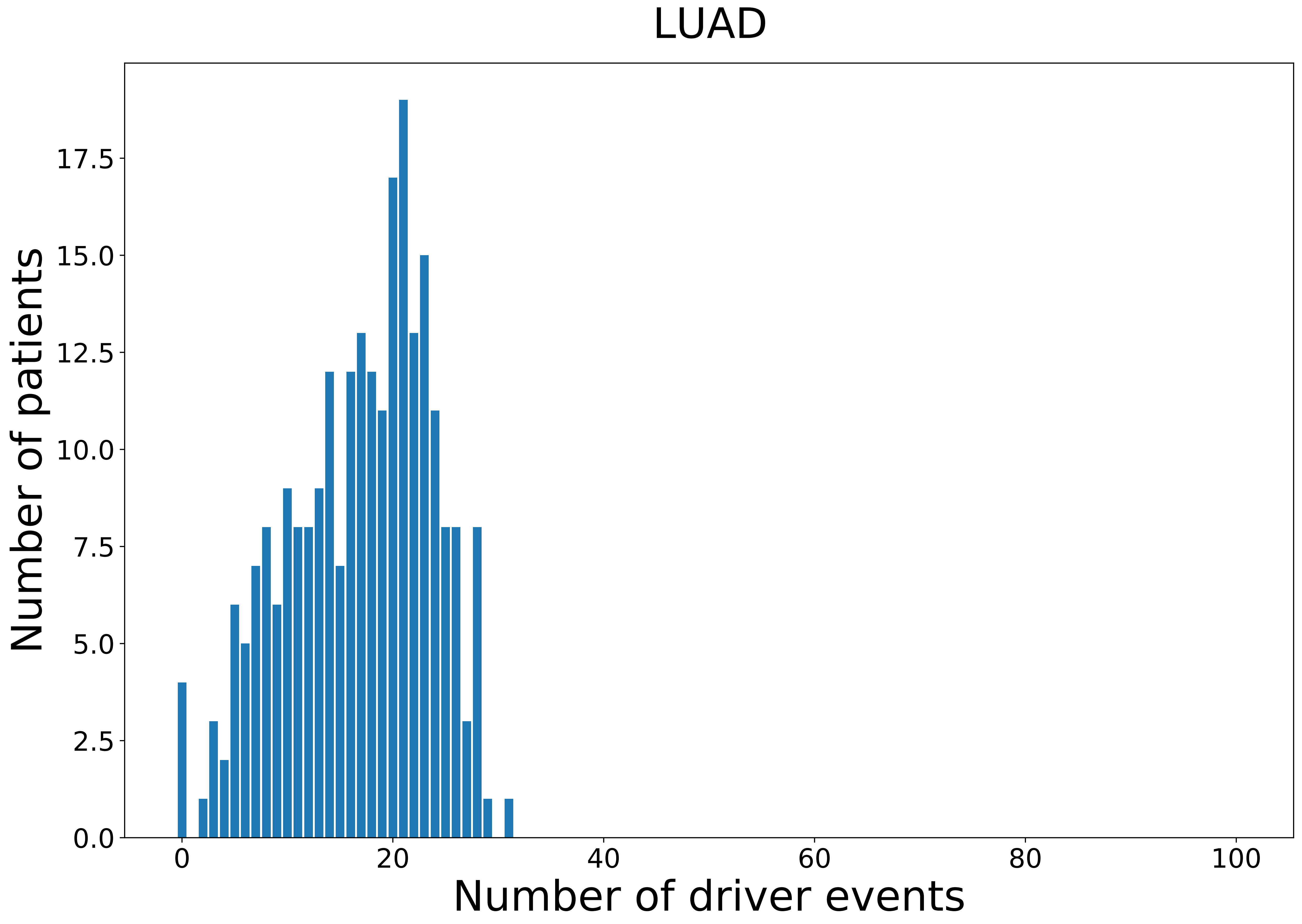

### 2021_8_16_14_9_LUAD_FEMALE.pdf

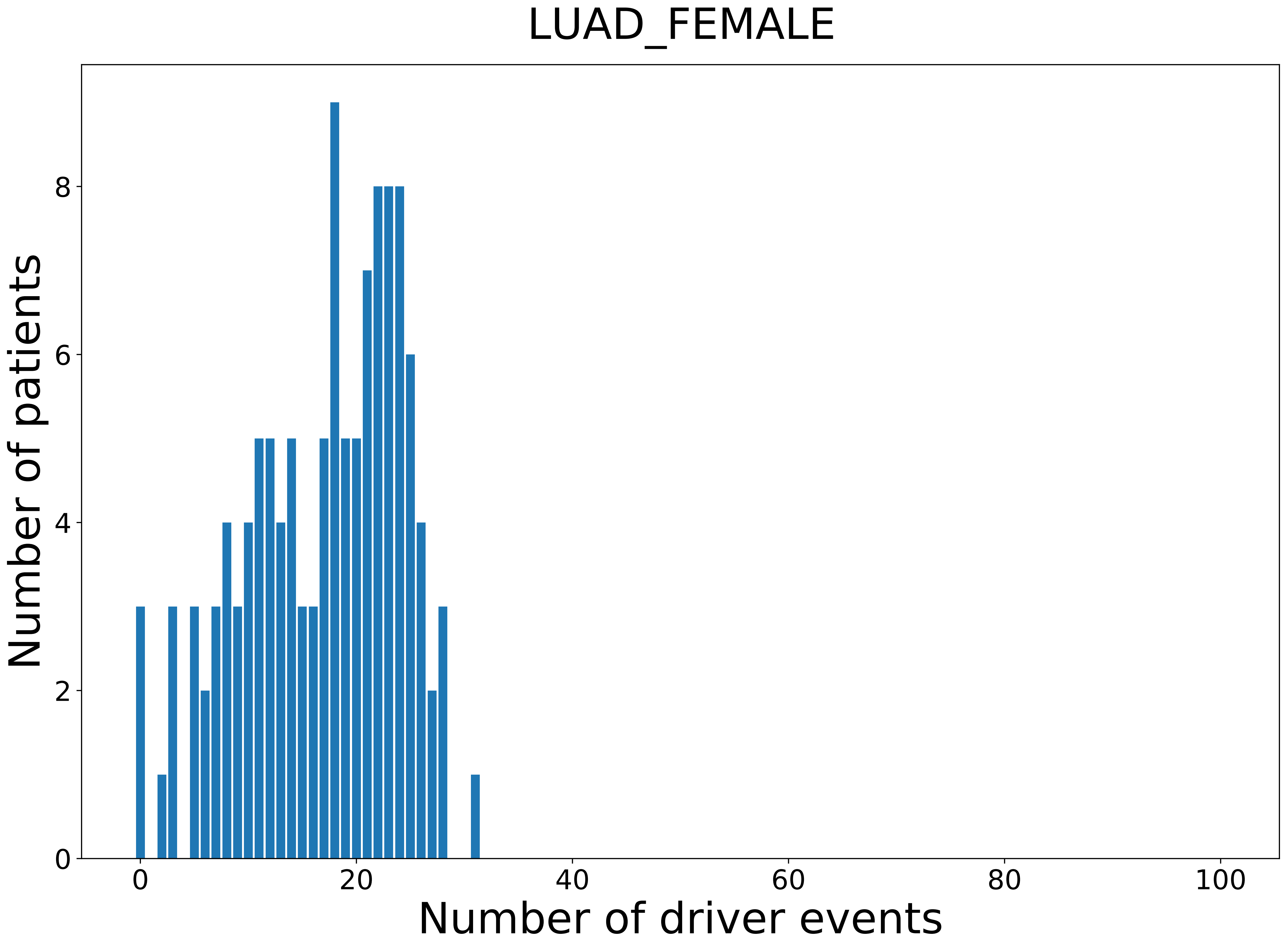

### 2021_8_16_14_9_LUSC.pdf

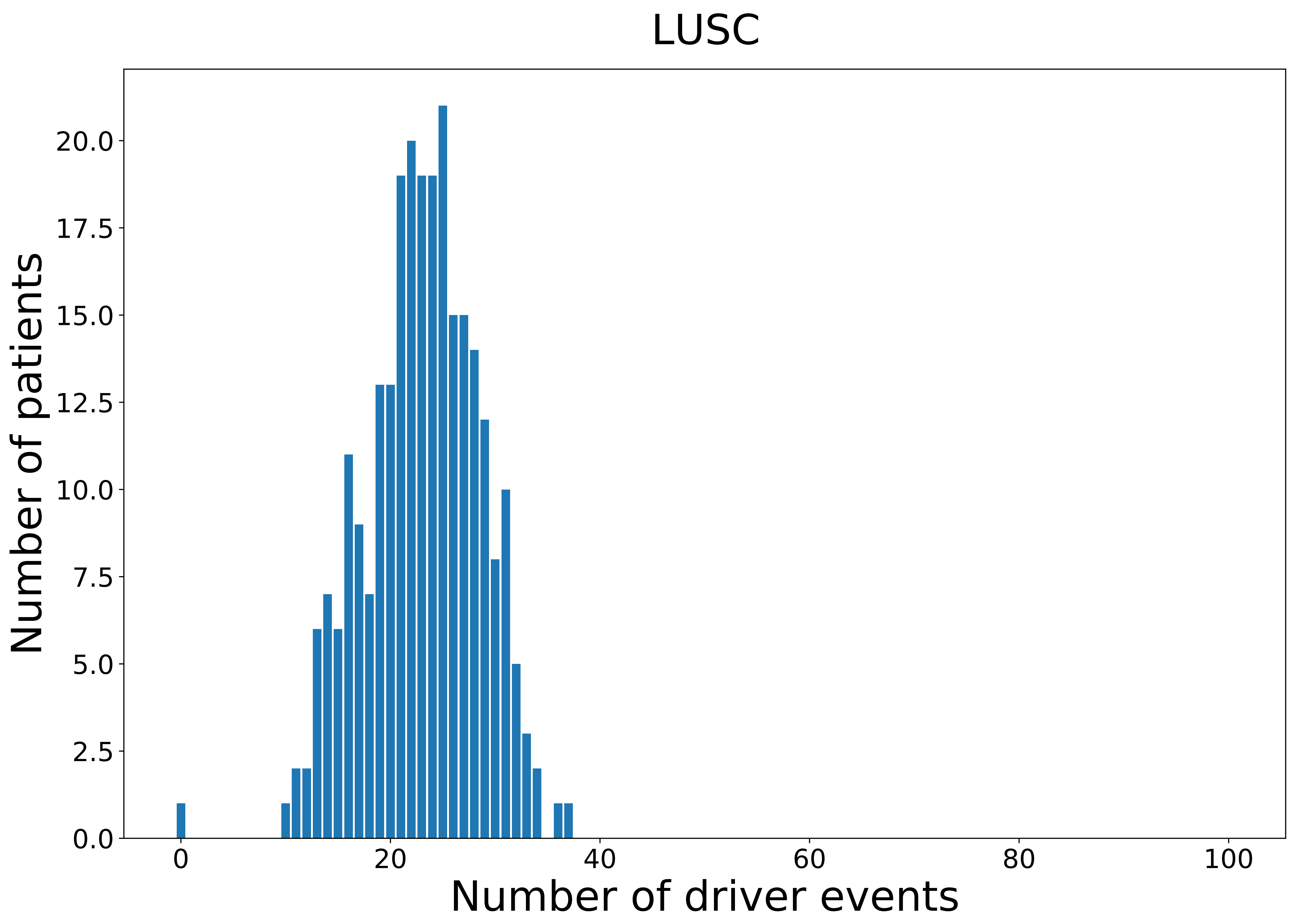

### 2021_8_16_14_9_MESO_FEMALE.pdf

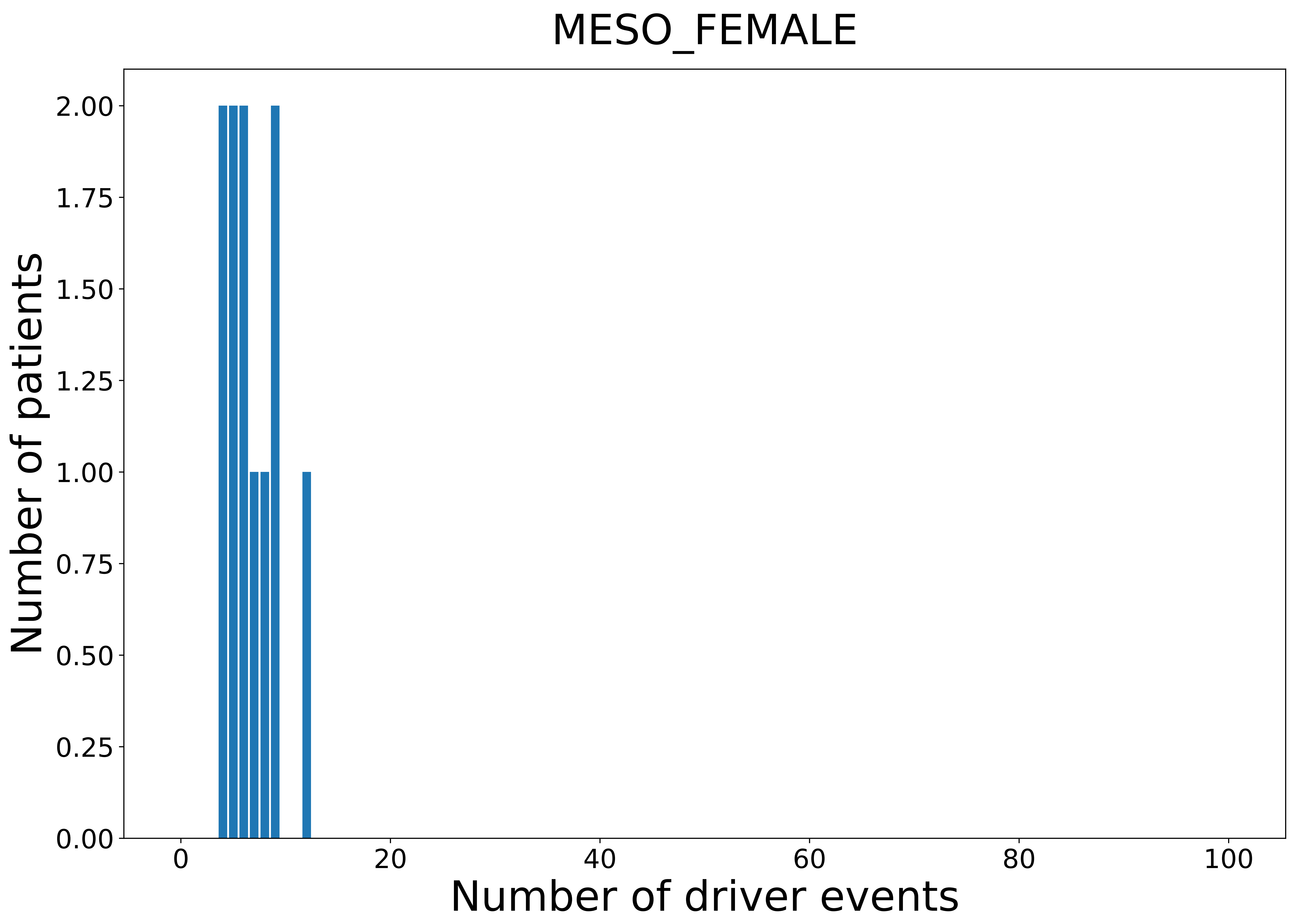

### 2021_8_16_14_9_OV_FEMALE.pdf

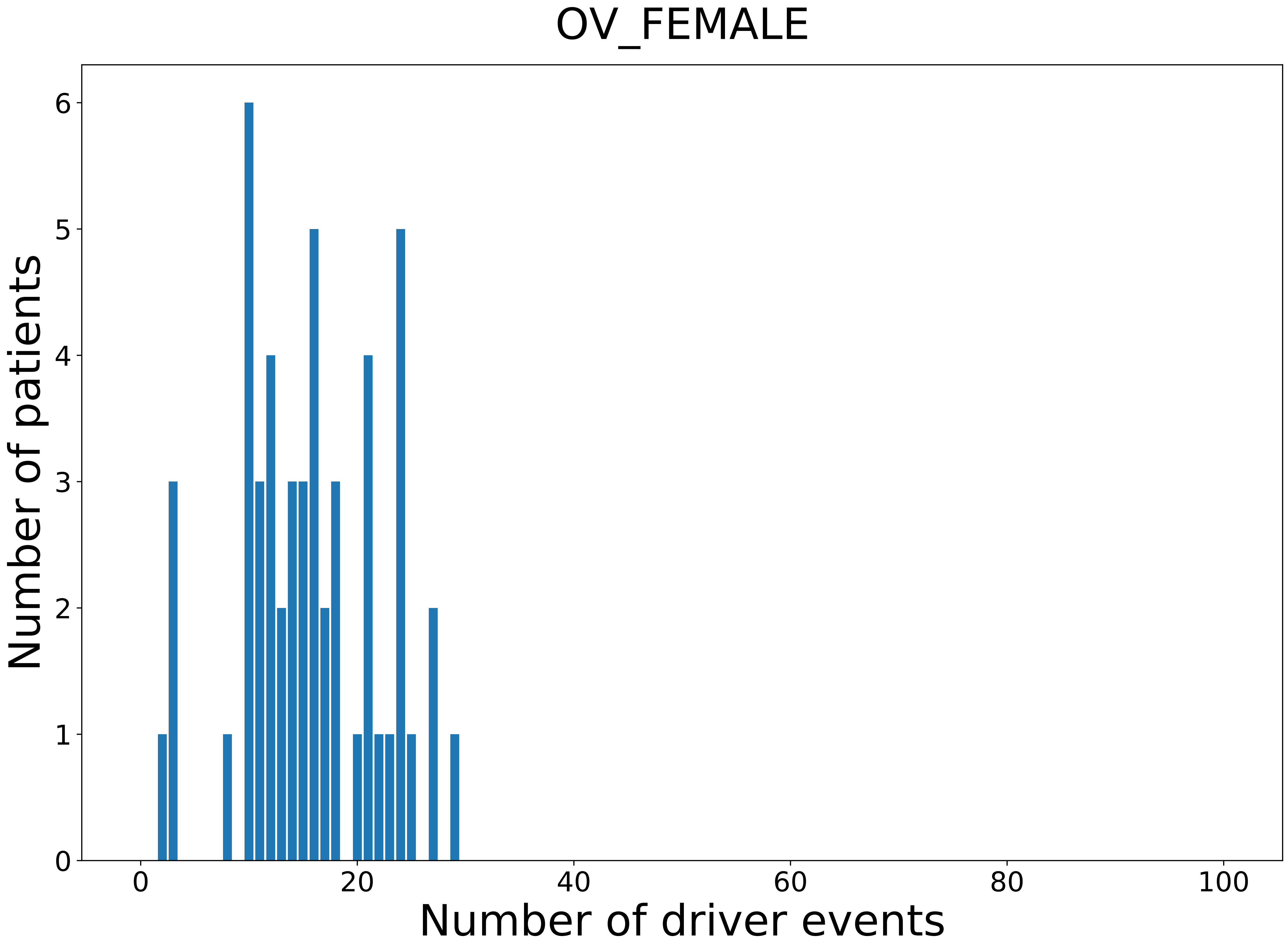

### 2021_8_16_14_9_PAAD.pdf

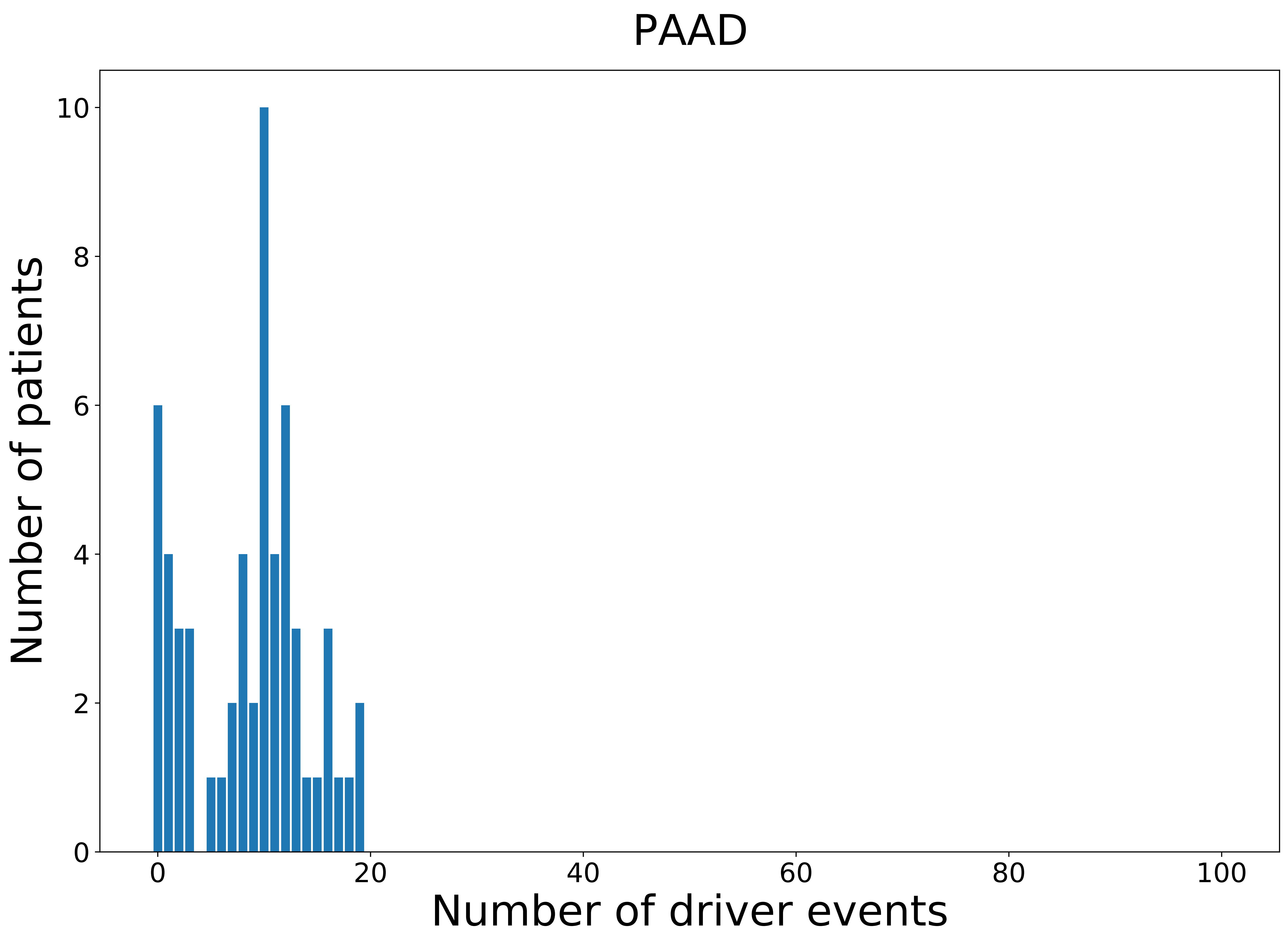

### 2021_8_16_14_9_PAAD_FEMALE.pdf

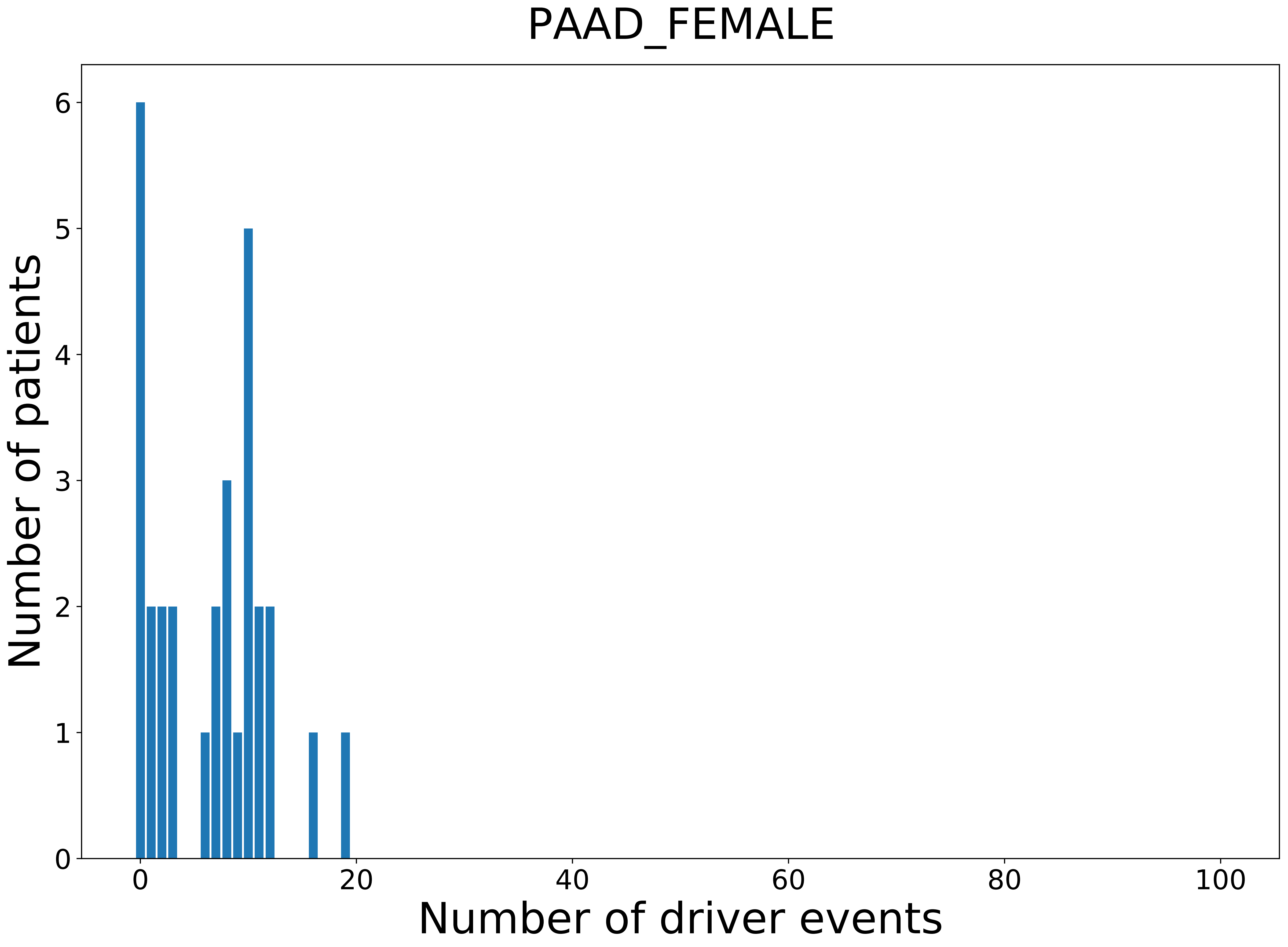

### 2021_8_16_14_9_PANCAN.pdf

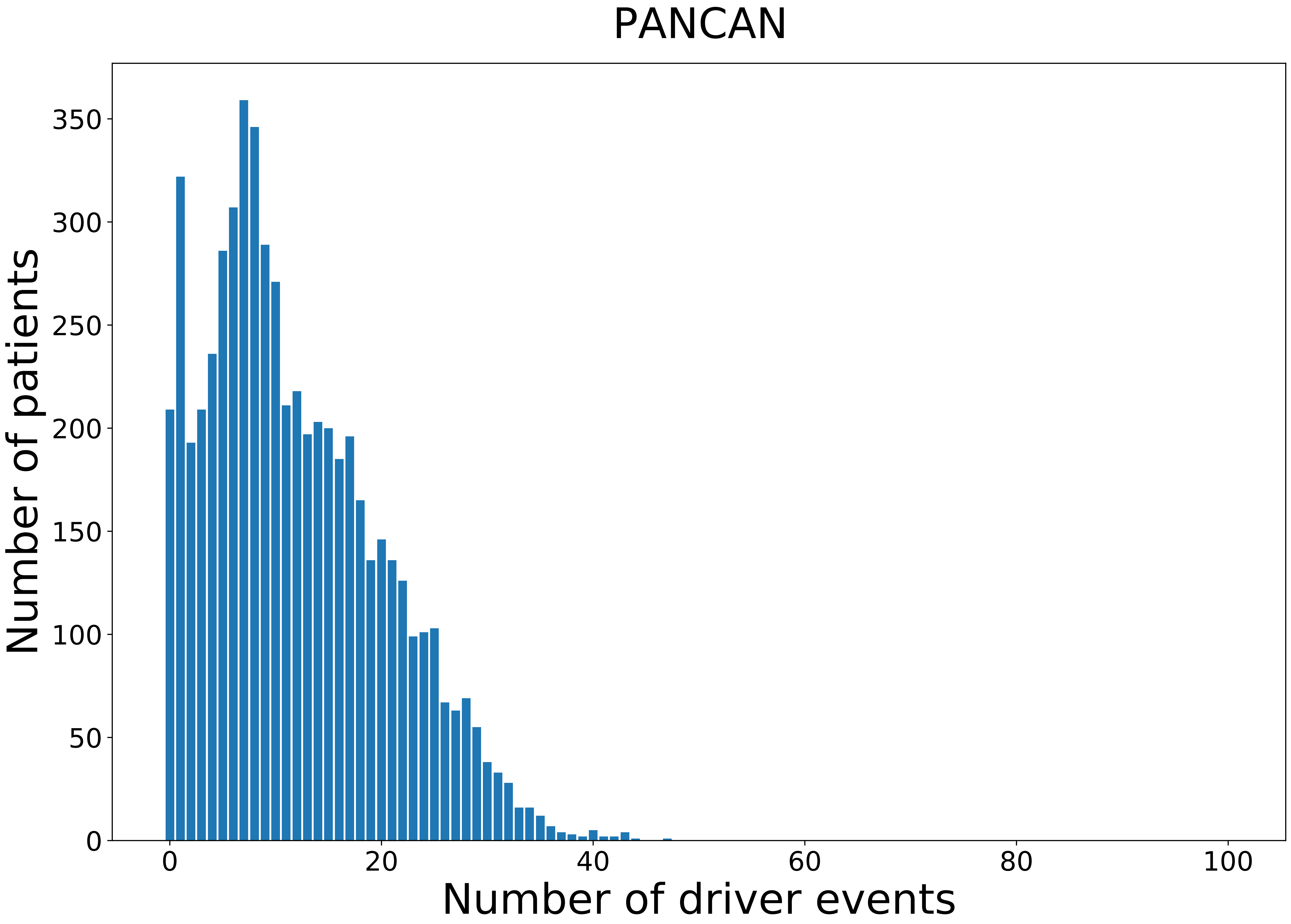

### 2021_8_16_14_9_PANCAN_FEMALE.pdf

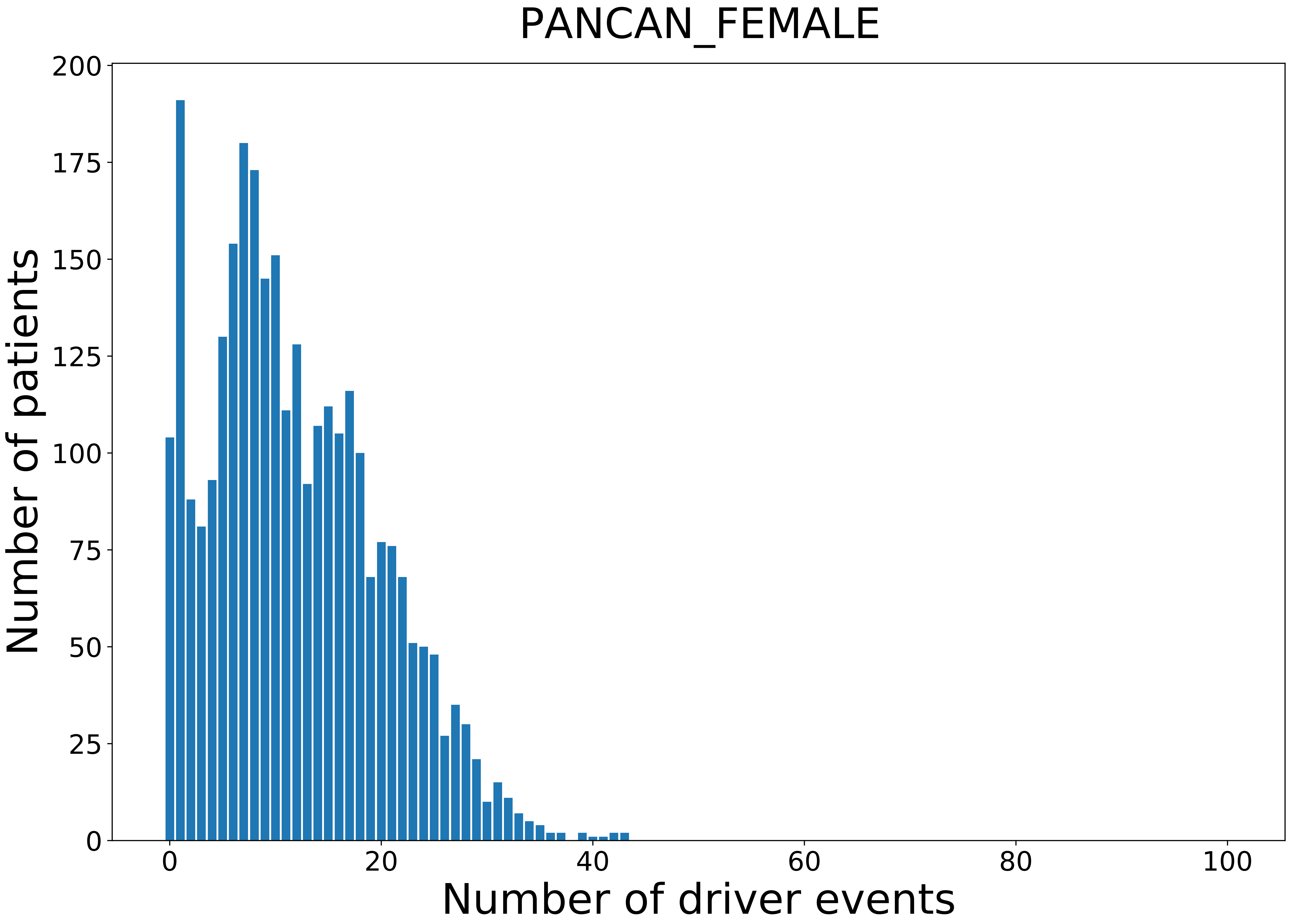

### 2021_8_16_14_9_PANCAN_MALE.pdf

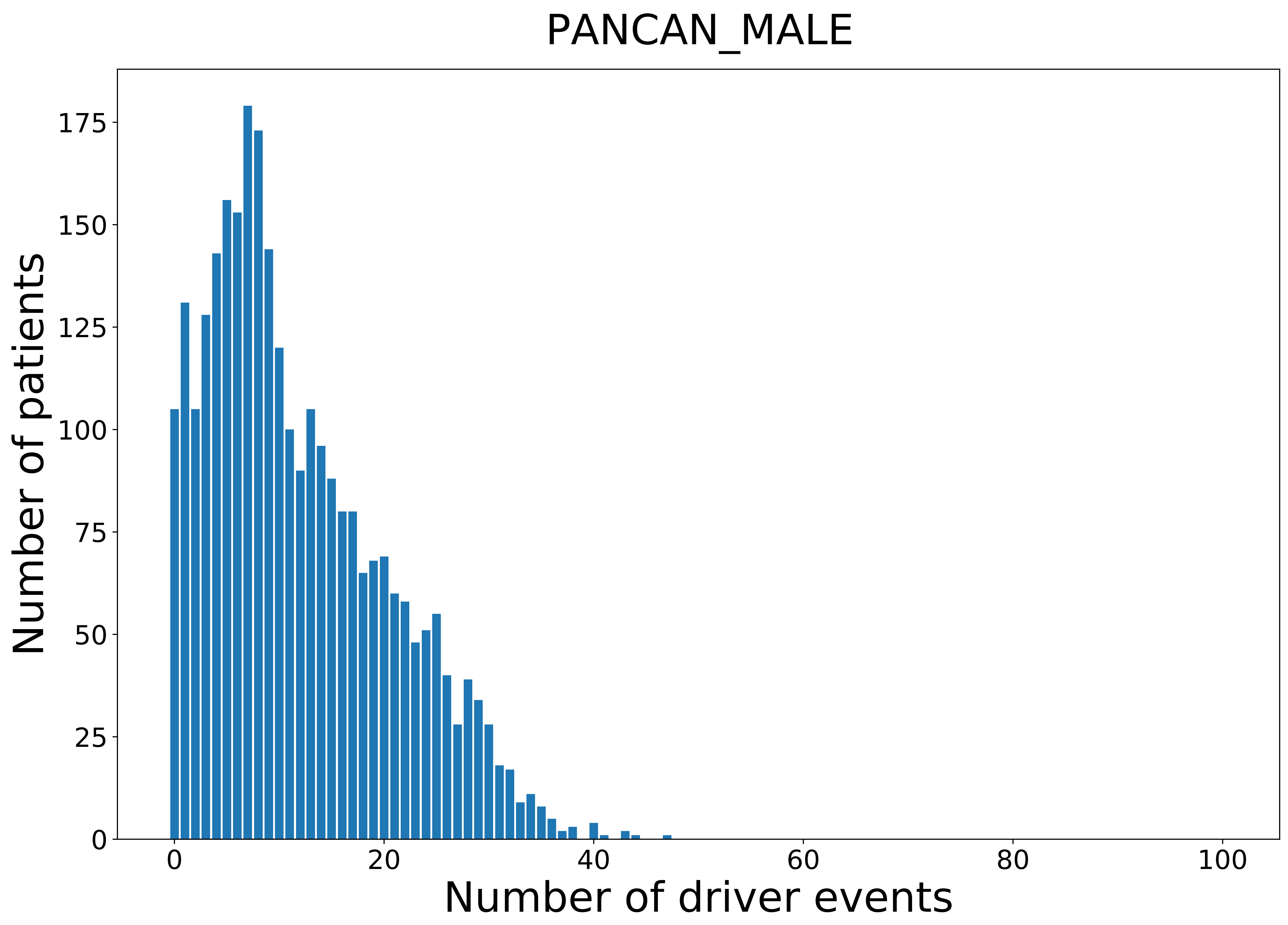

### 2021_8_16_14_9_PCPG.pdf

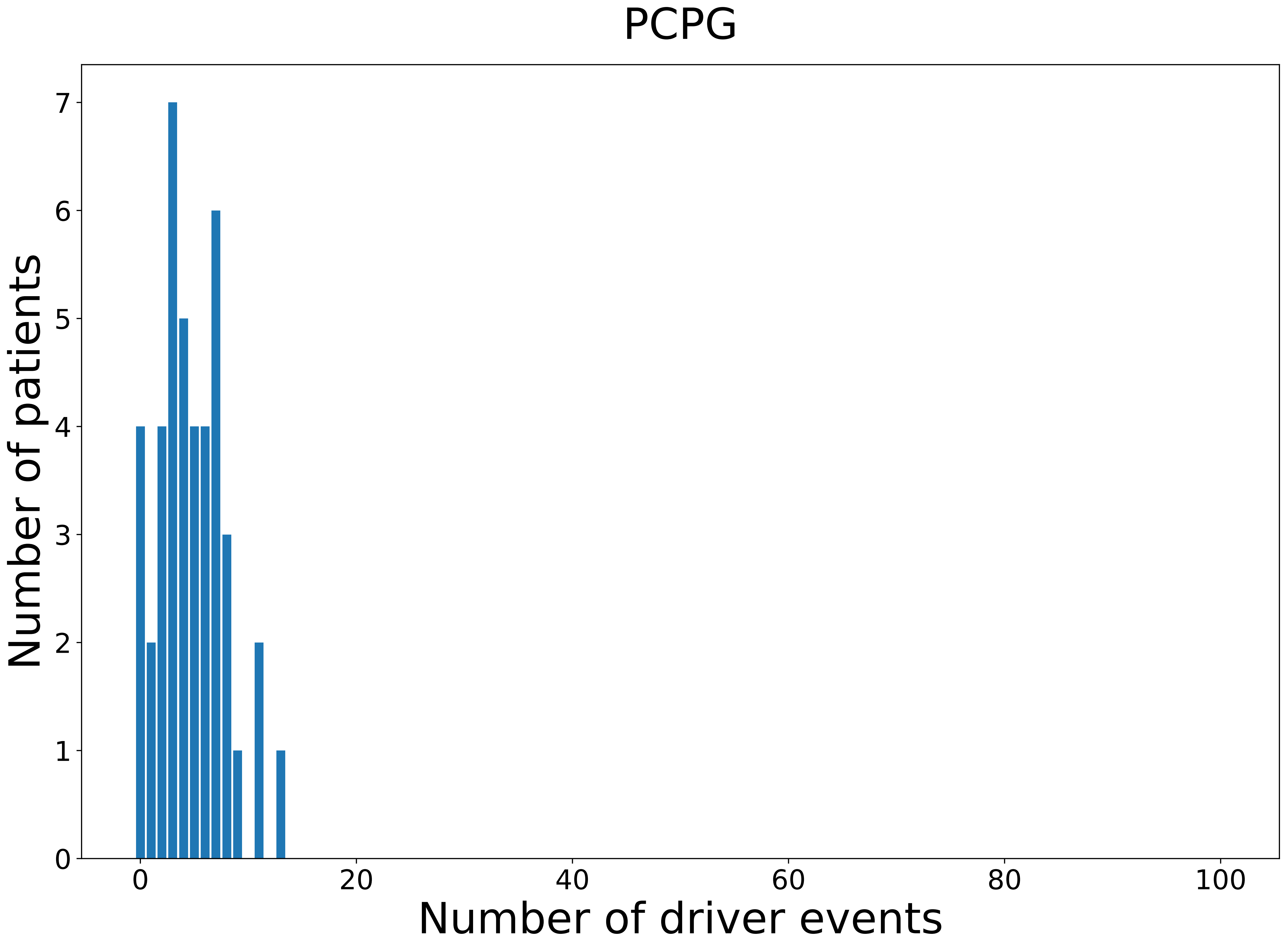

### 2021_8_16_14_9_PCPG_FEMALE.pdf

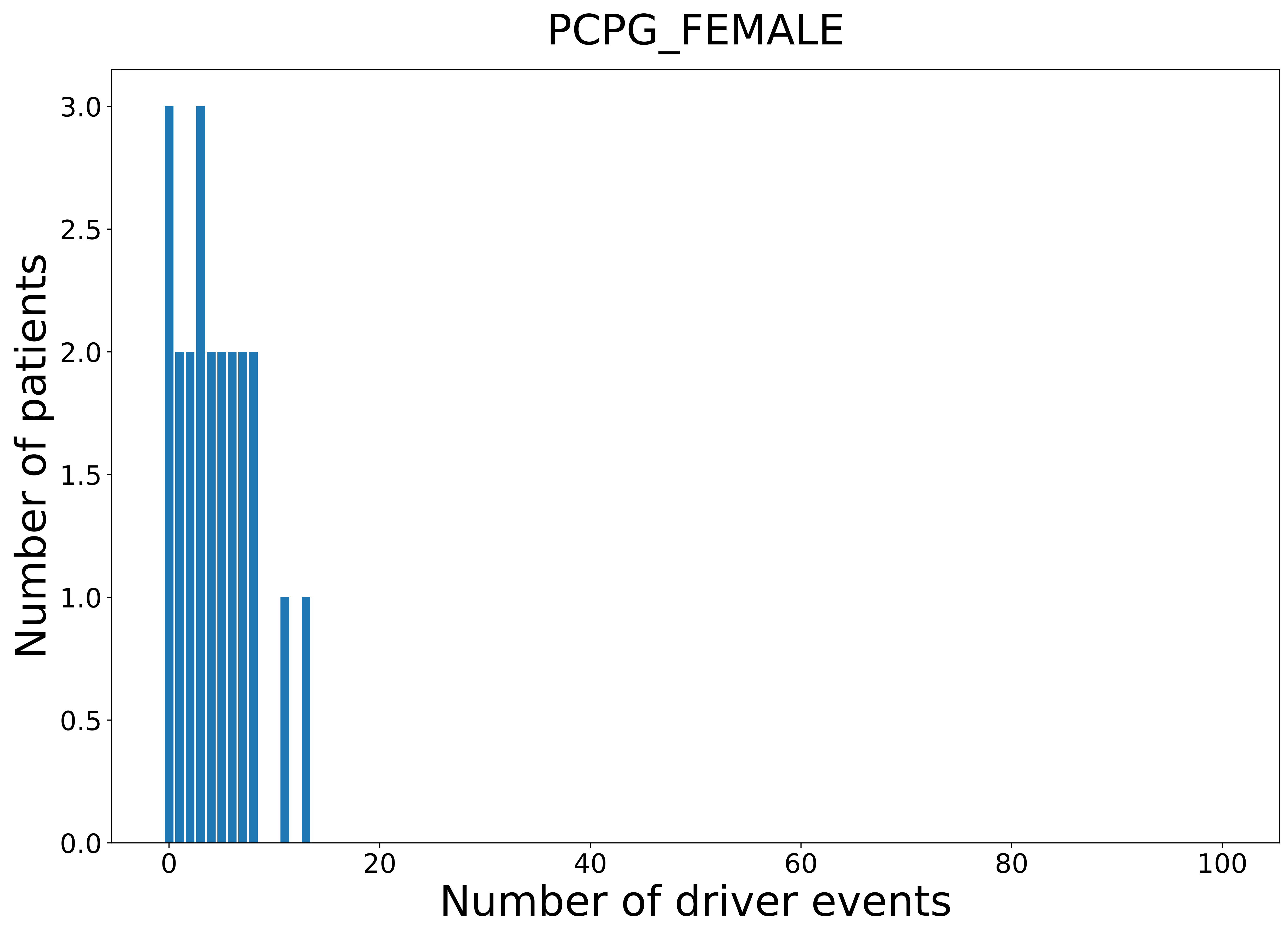

### 2021_8_16_14_9_PCPG_MALE.pdf

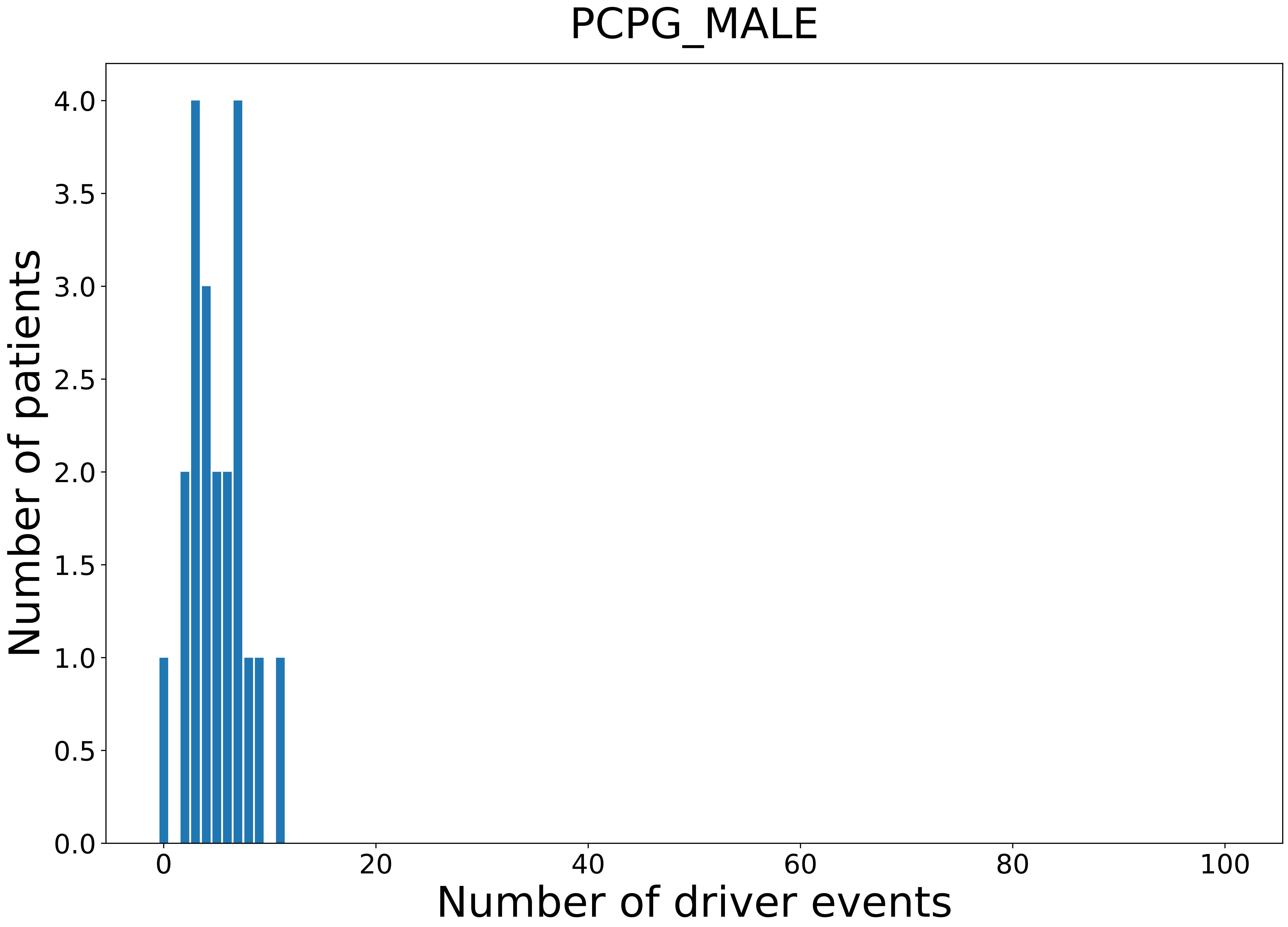

### 2021_8_16_14_9_PRAD.pdf

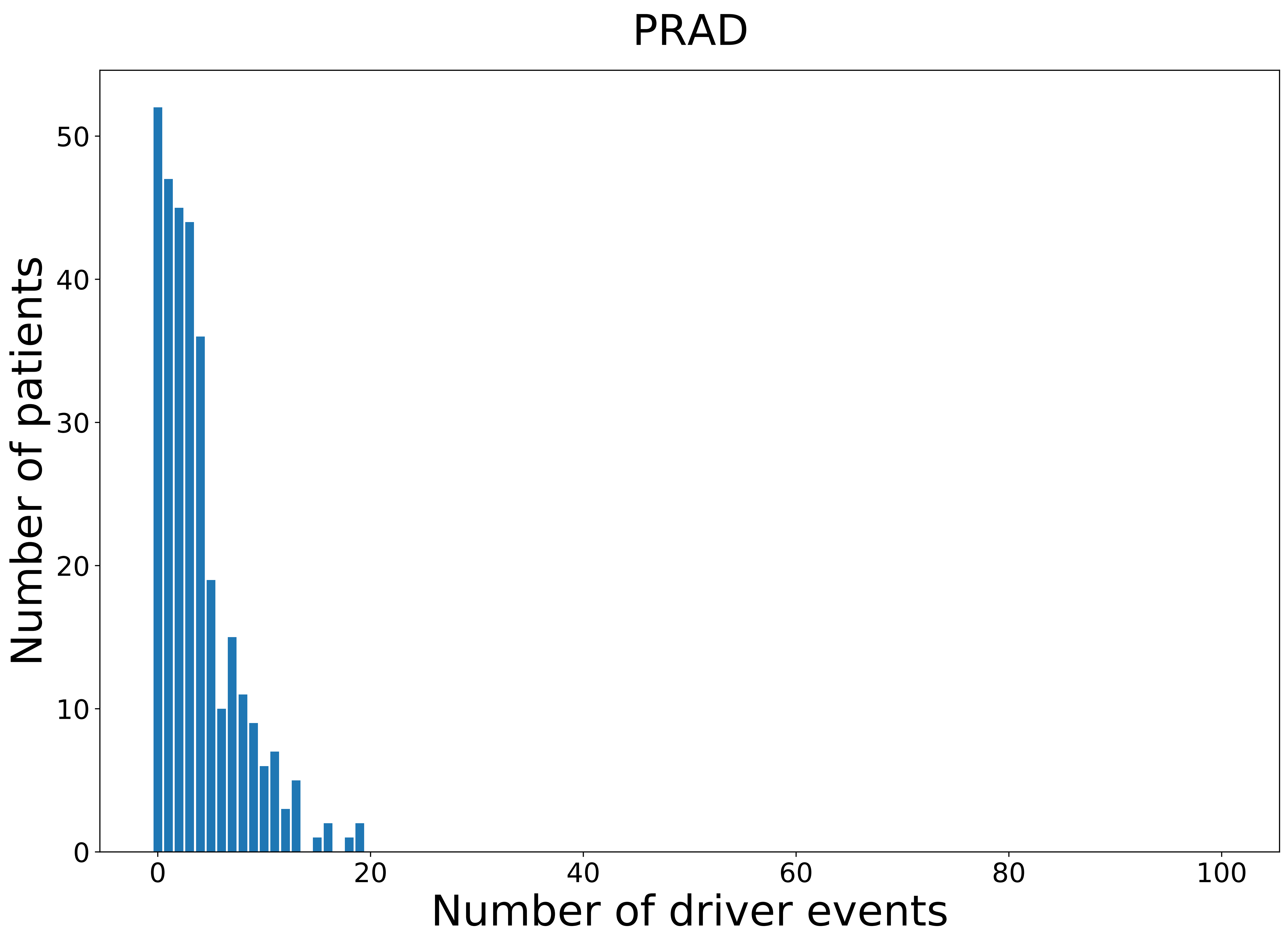

### 2021_8_16_14_9_READ.pdf

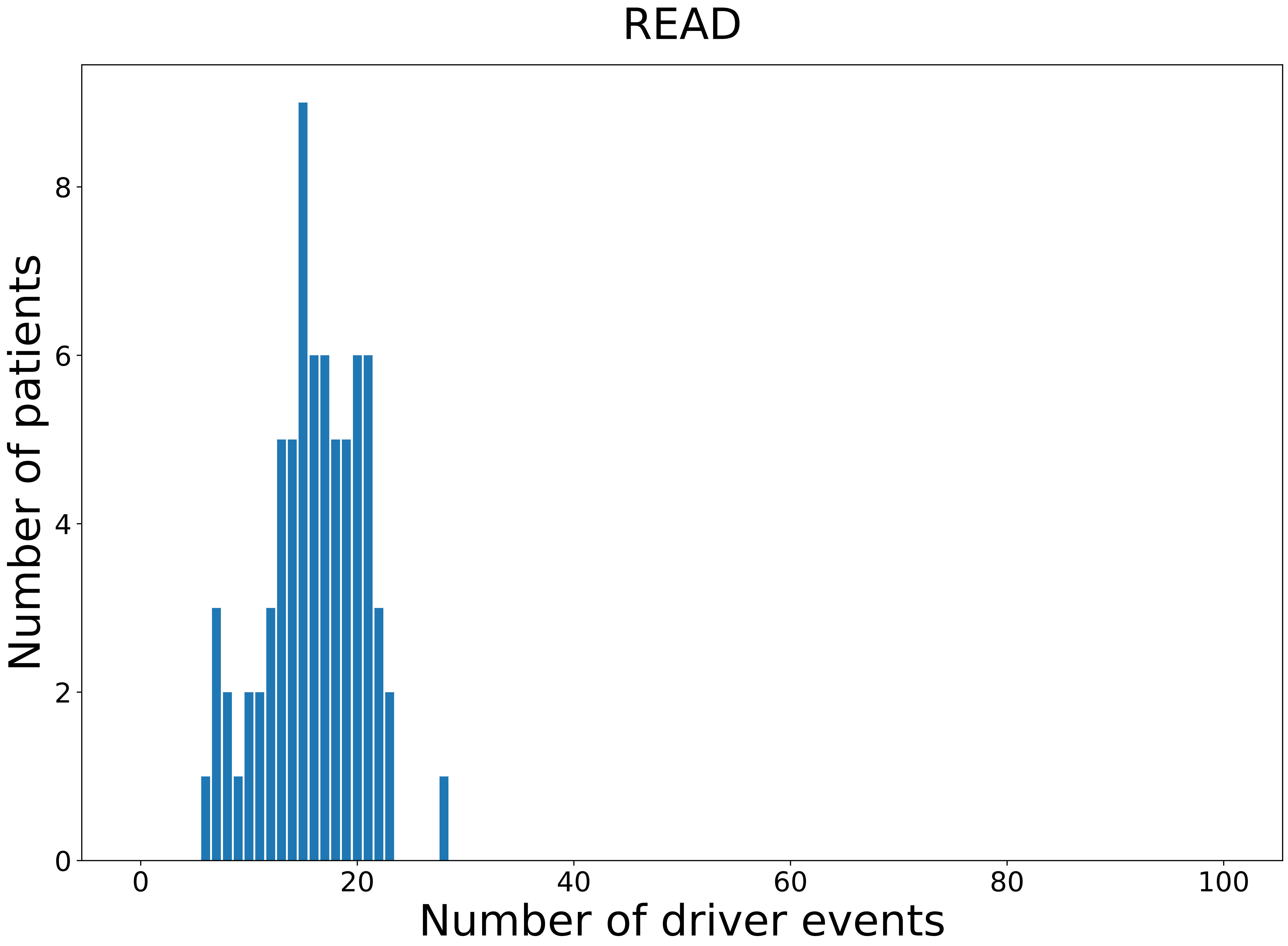

### 2021_8_16_14_9_READ_FEMALE.pdf

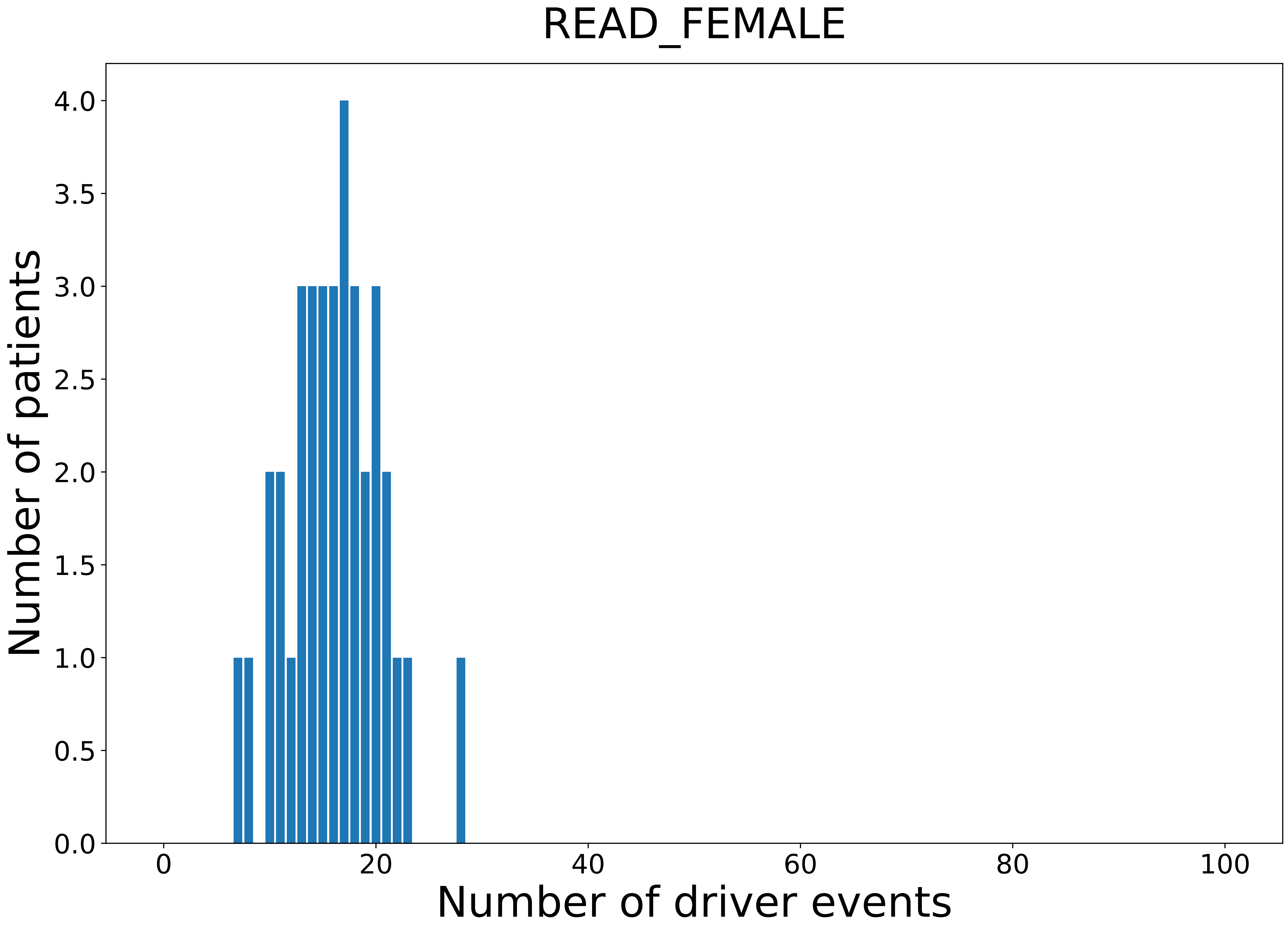

### 2021_8_16_14_9_READ_MALE.pdf

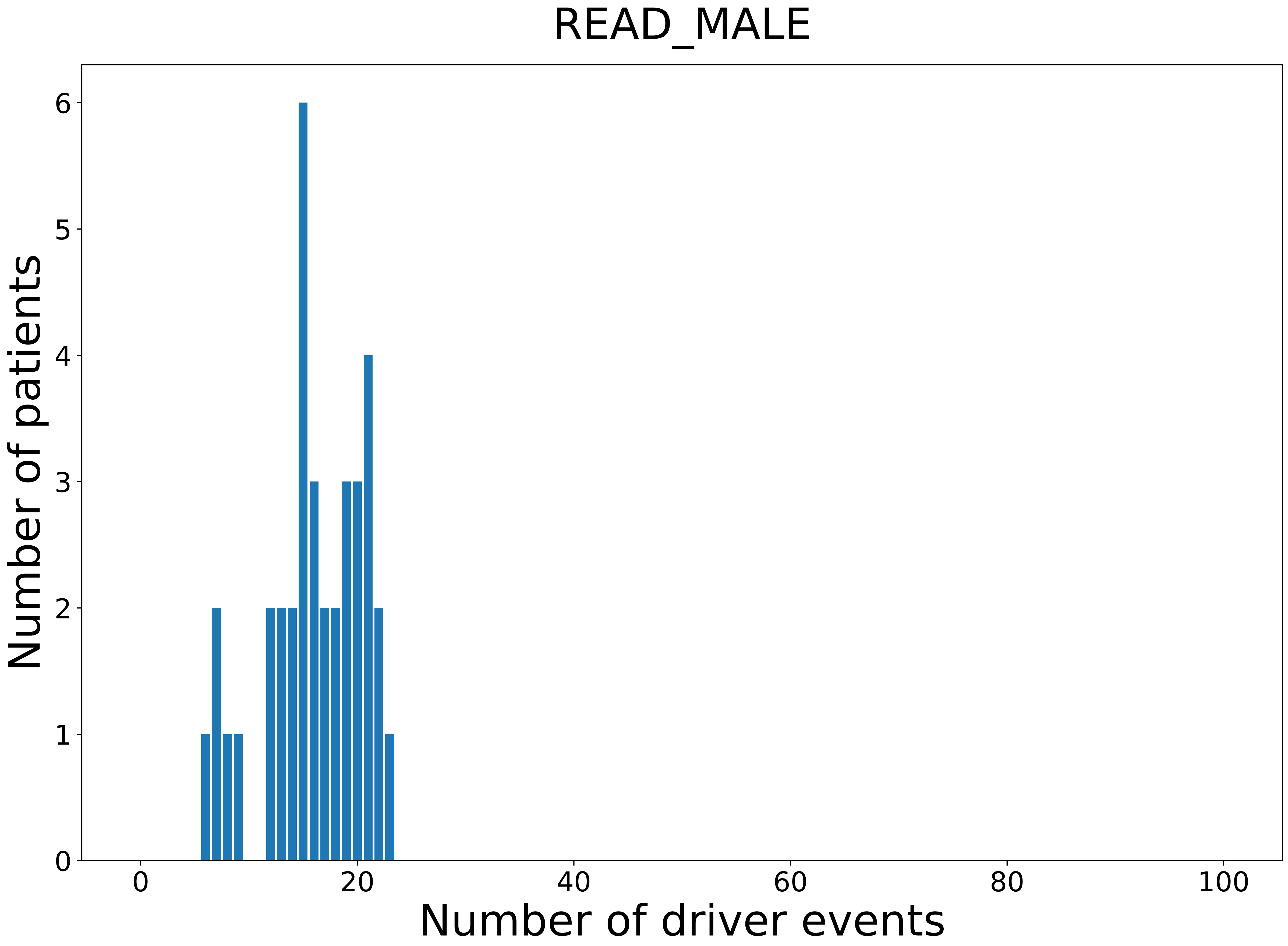

### 2021_8_16_14_9_SARC.pdf

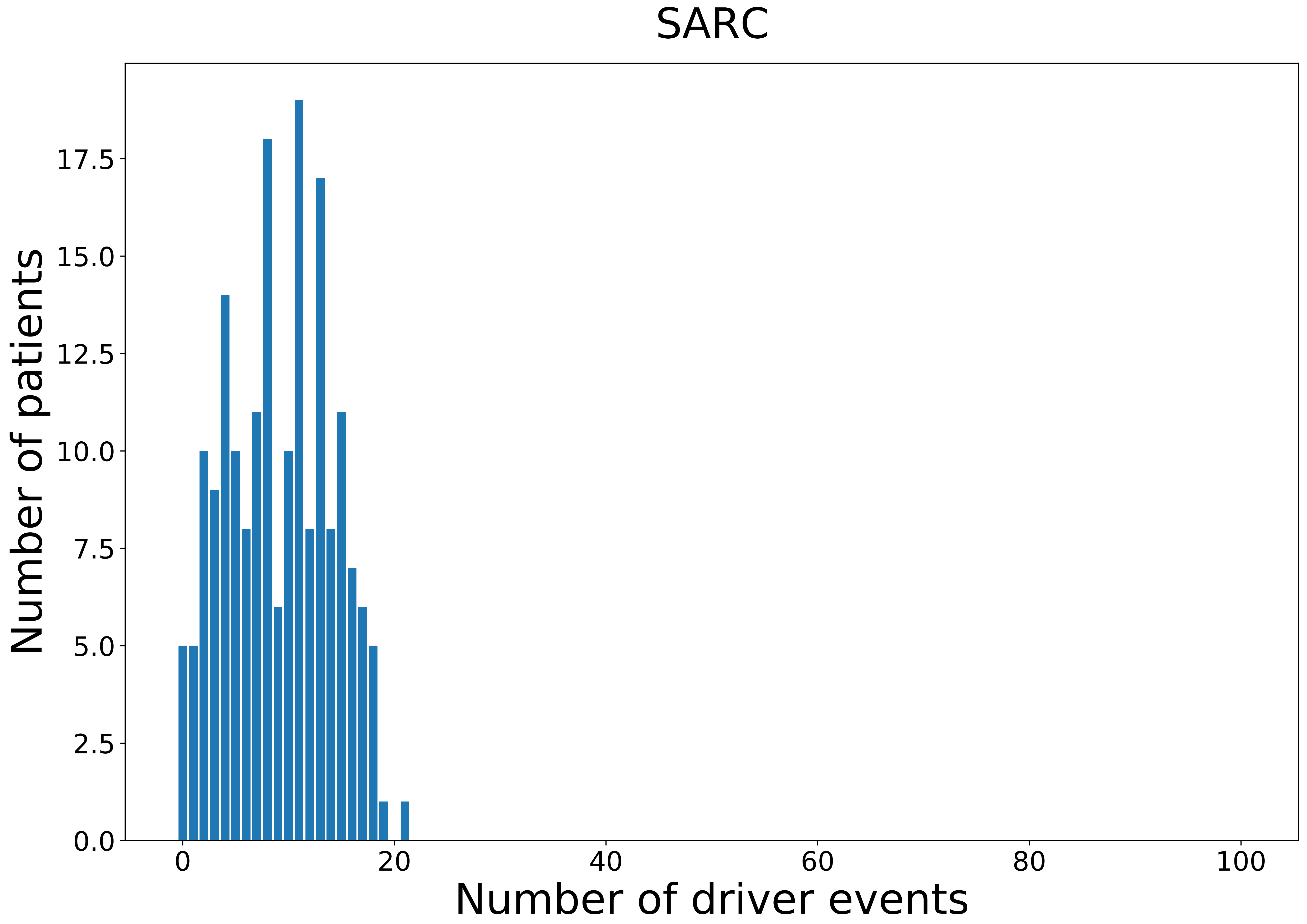

### 2021_8_16_14_9_SARC_FEMALE.pdf

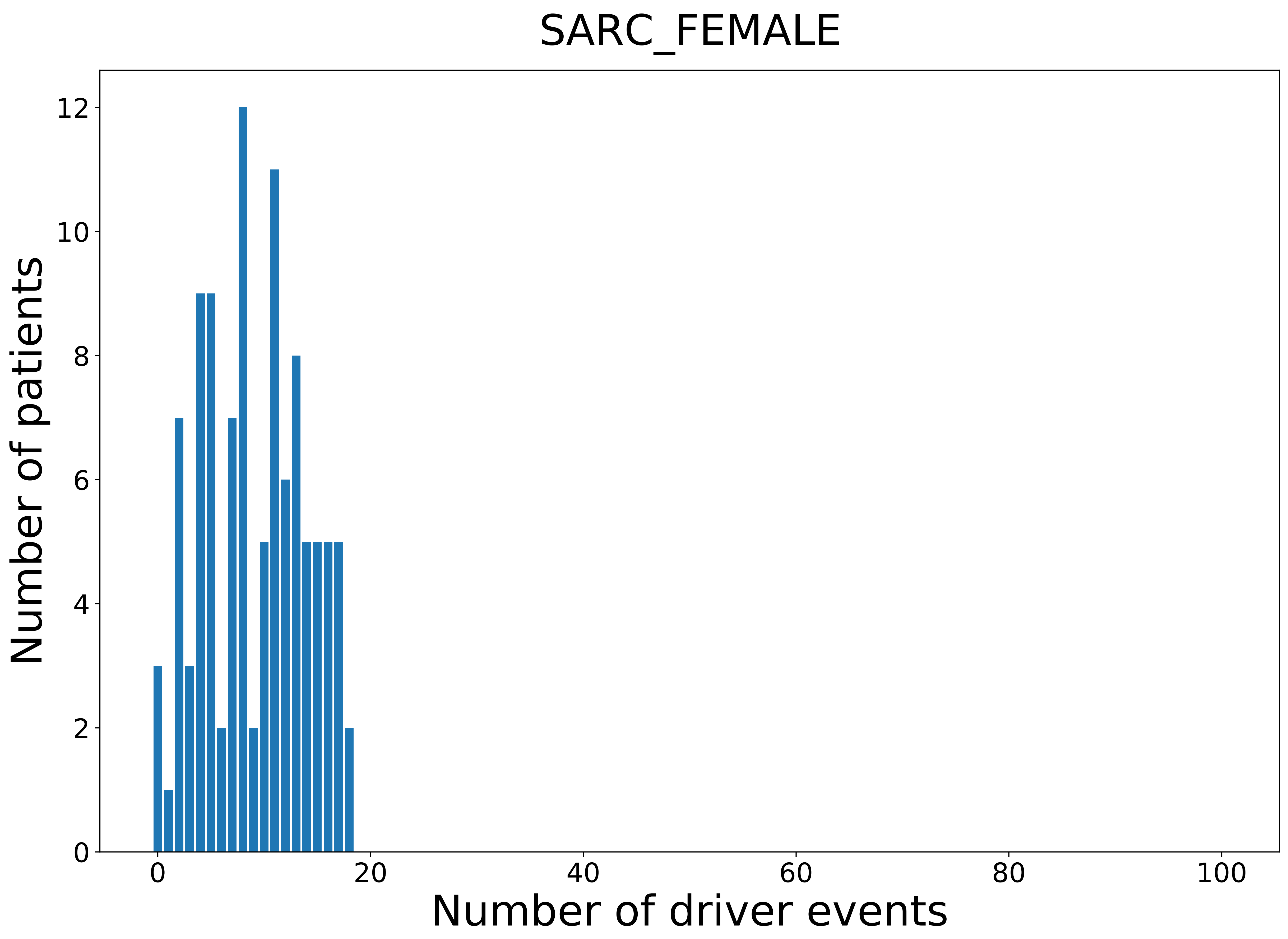

### 2021_8_16_14_9_SKCM_FEMALE.pdf

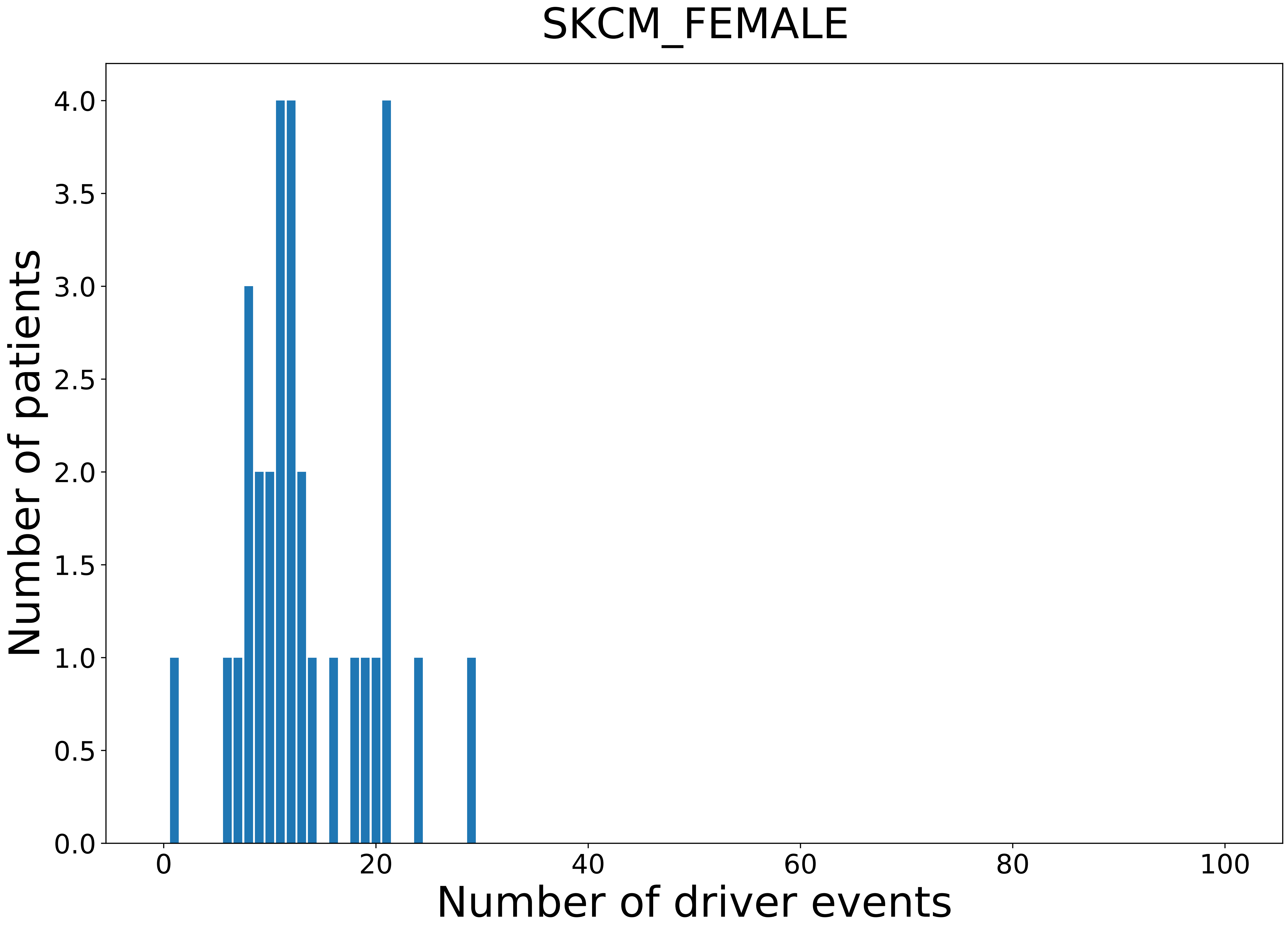

### 2021_8_16_14_9_STAD_FEMALE.pdf

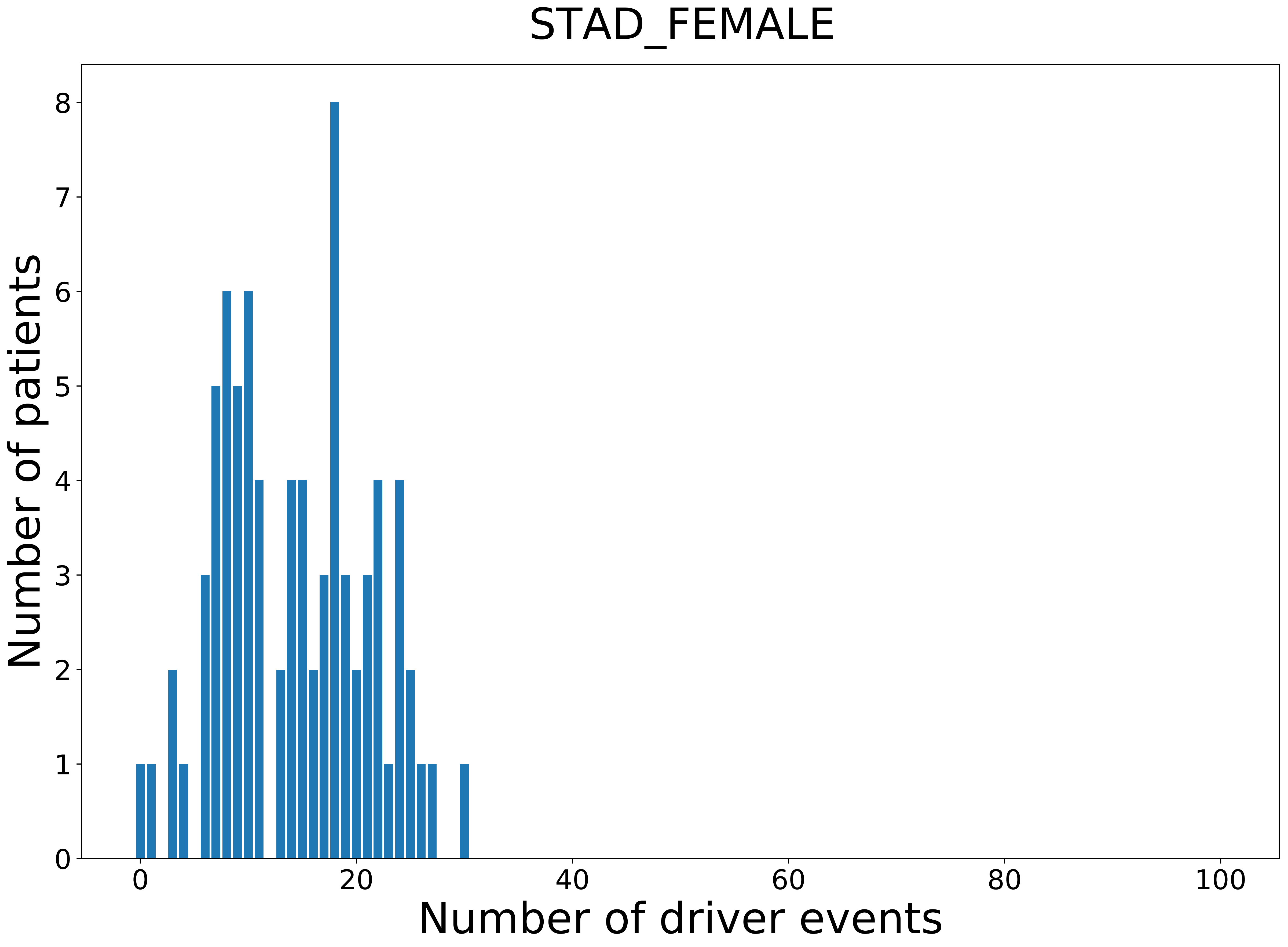

### 2021_8_16_14_9_STAD_MALE.pdf

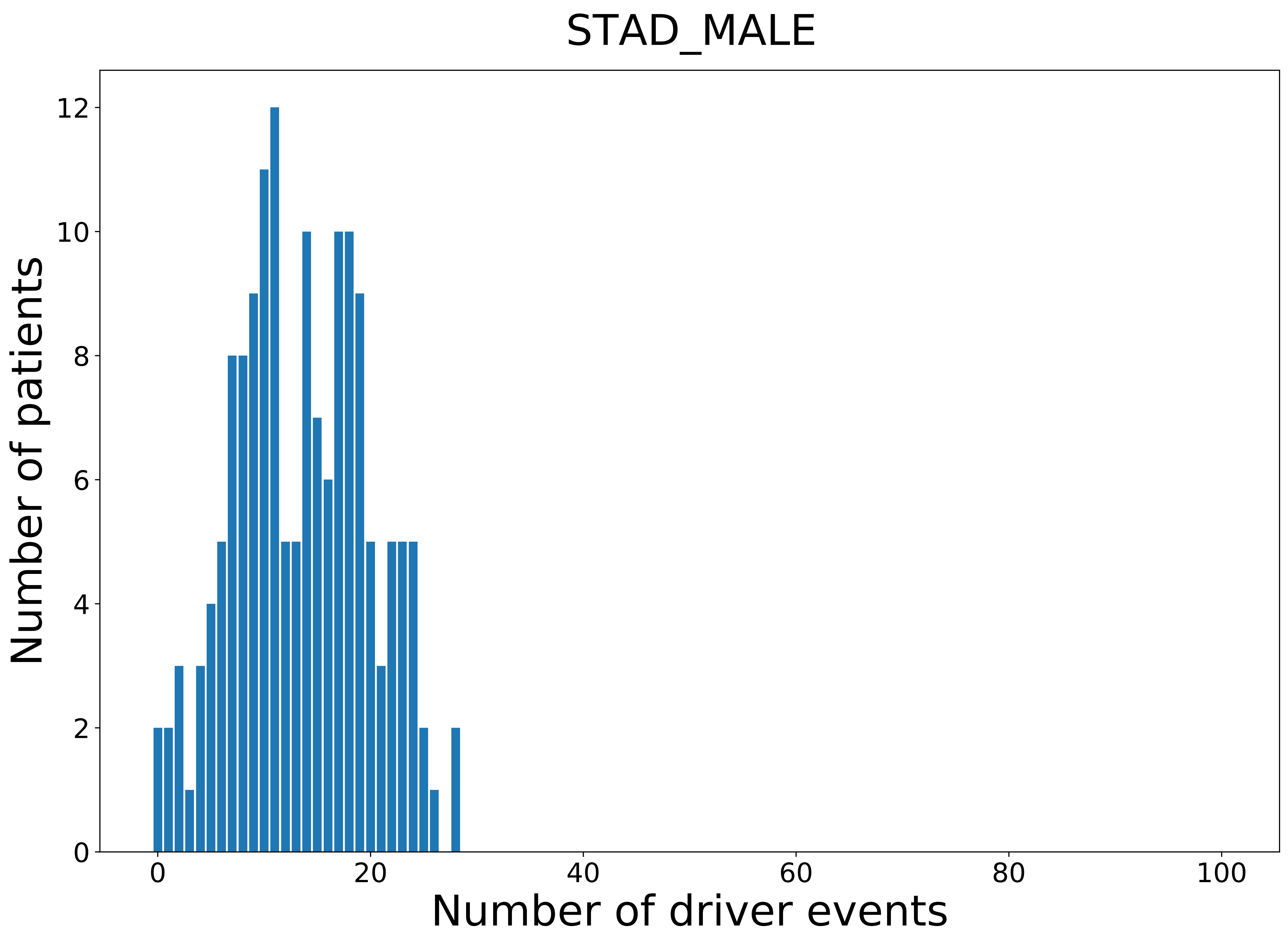

### 2021_8_16_14_9_TGCT.pdf

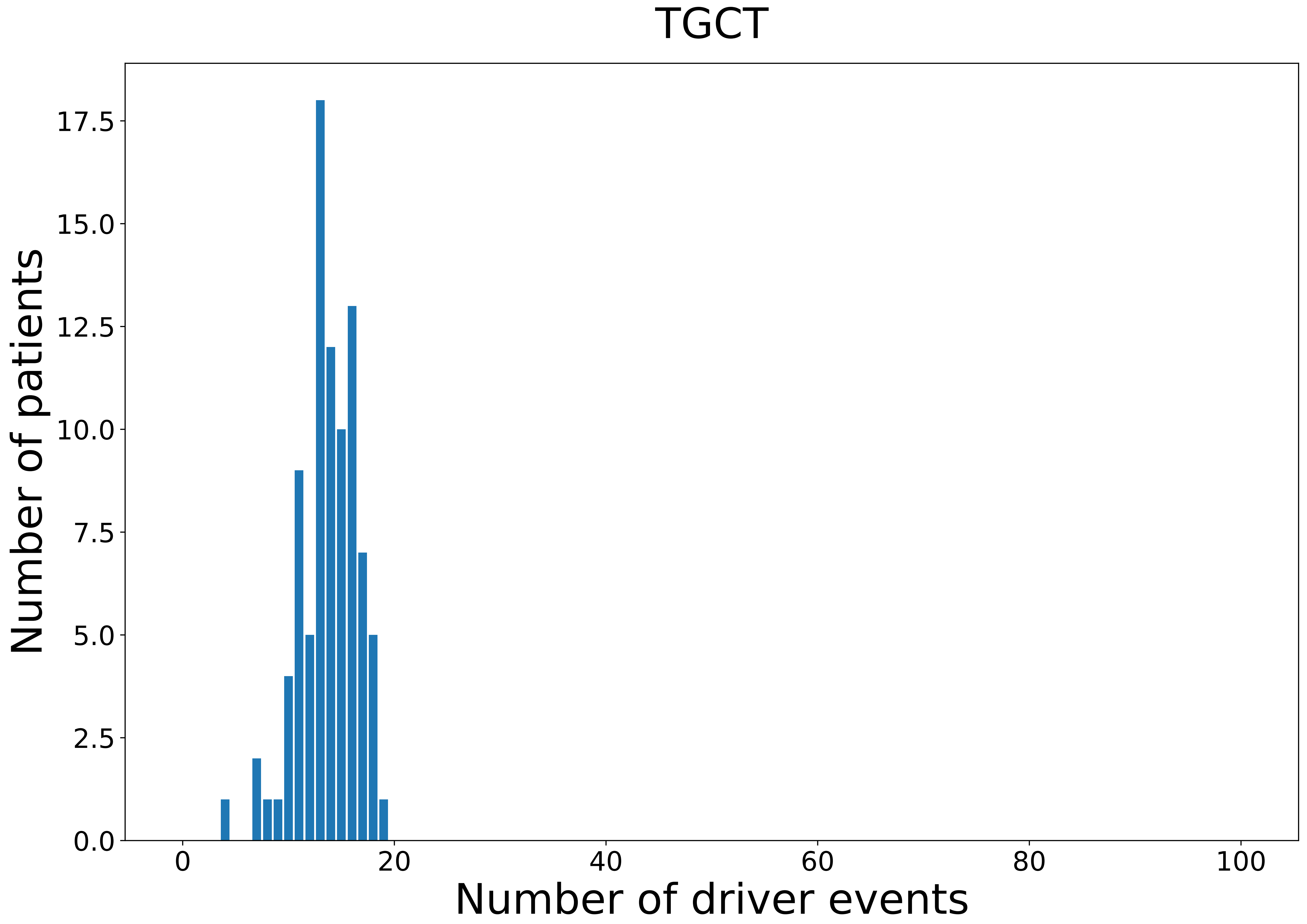

### 2021_8_16_14_9_THCA.pdf

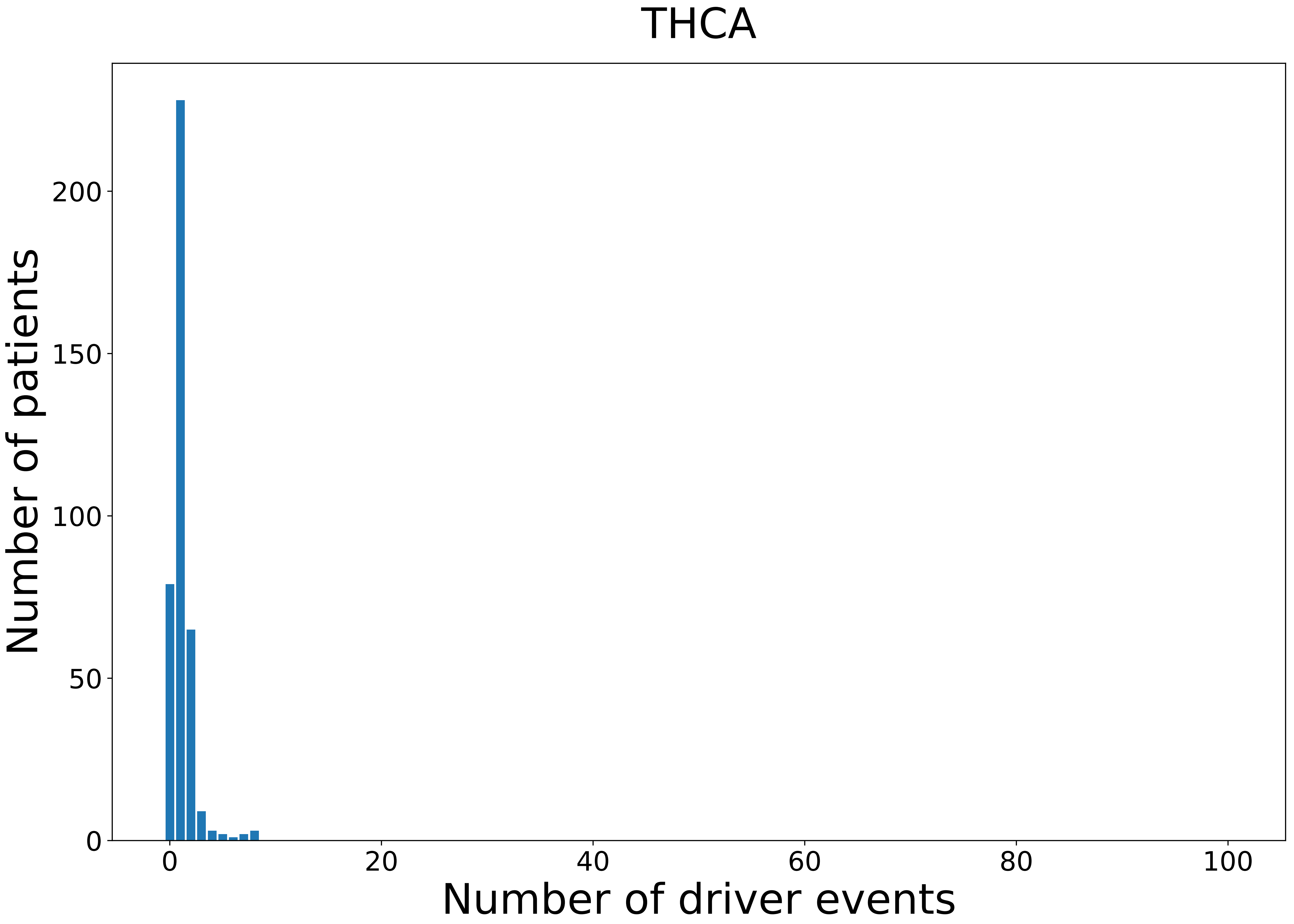

### 2021_8_16_14_9_THCA_FEMALE.pdf

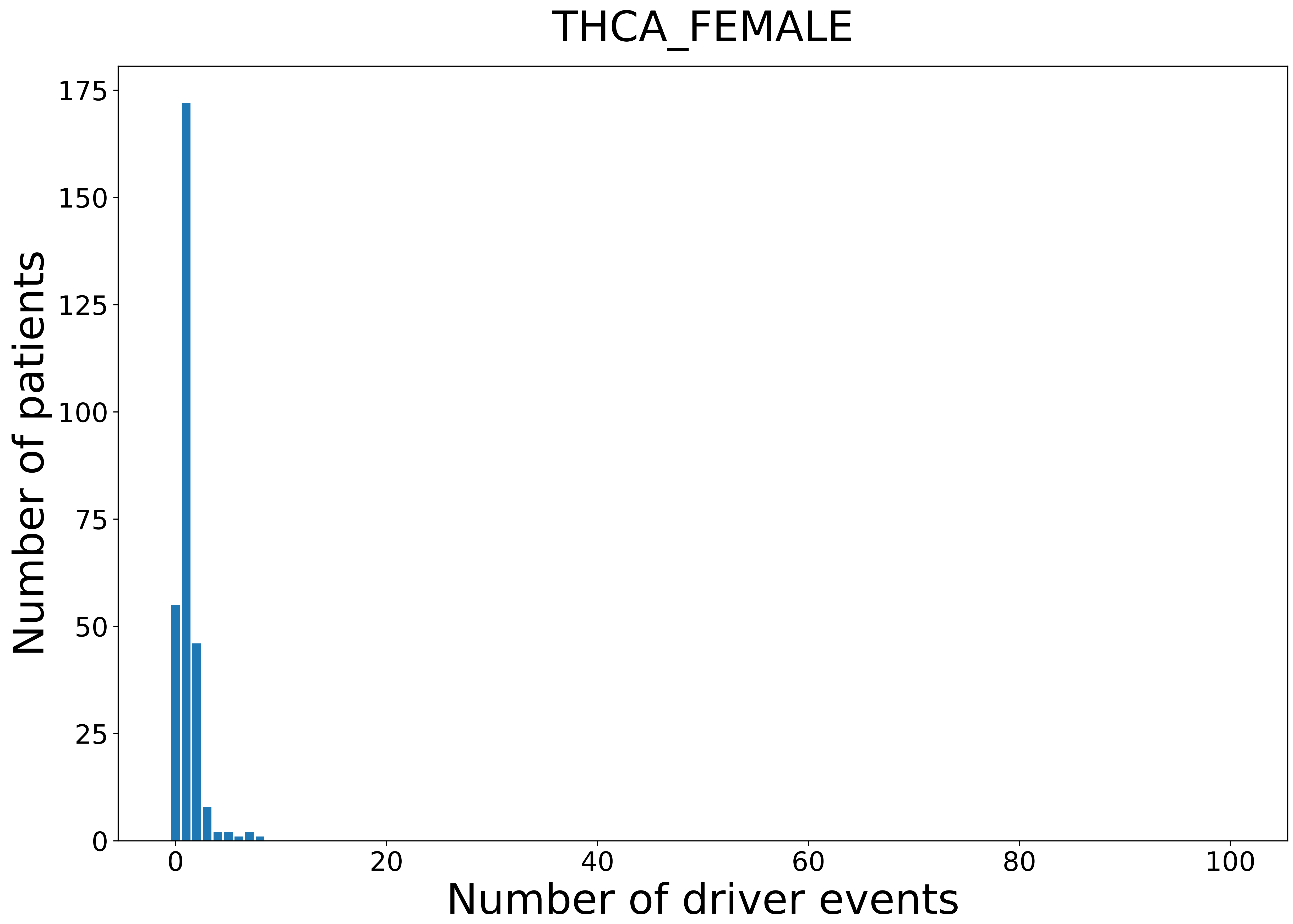

### 2021_8_16_14_9_THCA_MALE.pdf

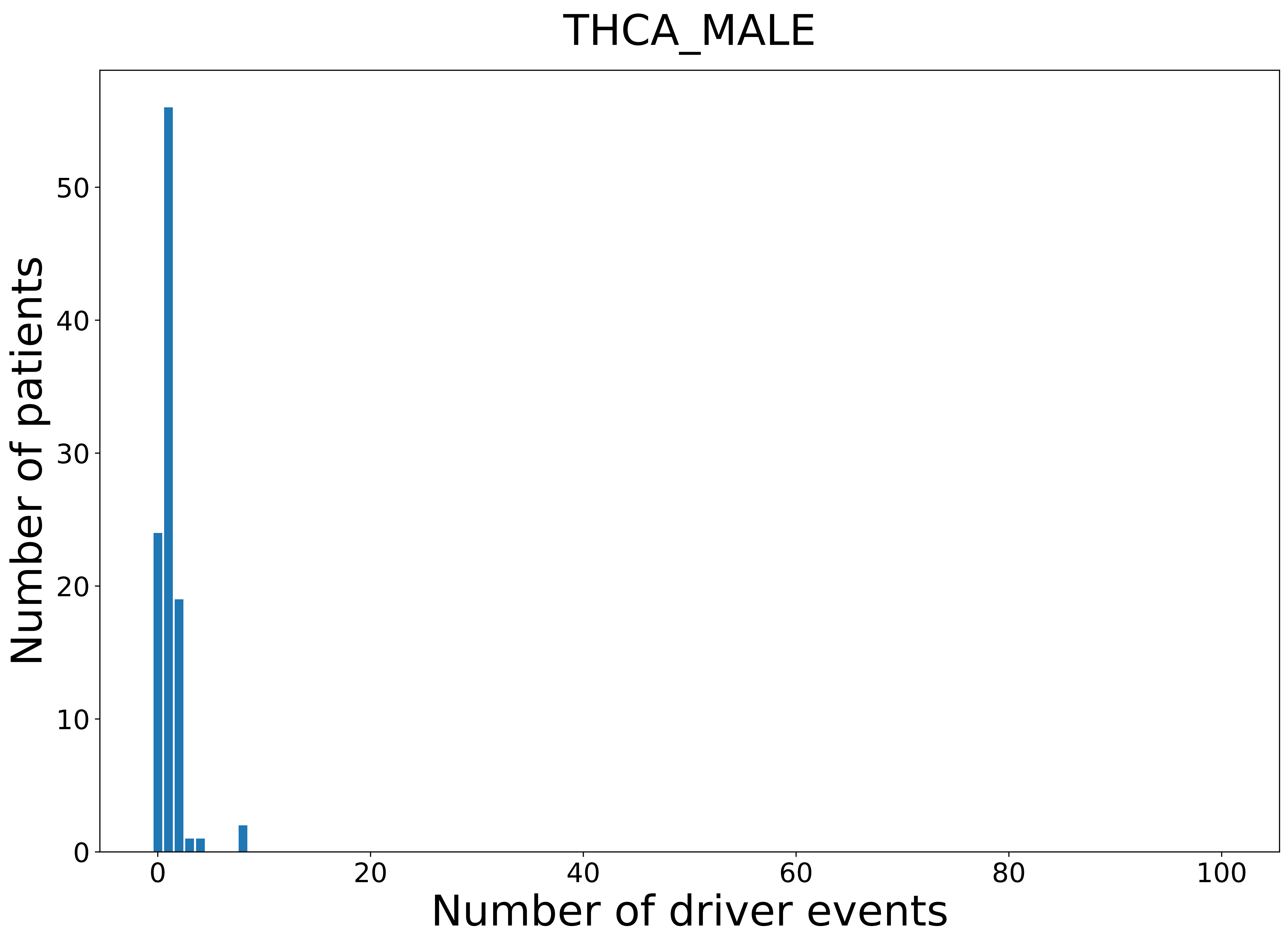

### 2021_8_16_14_9_THYM.pdf

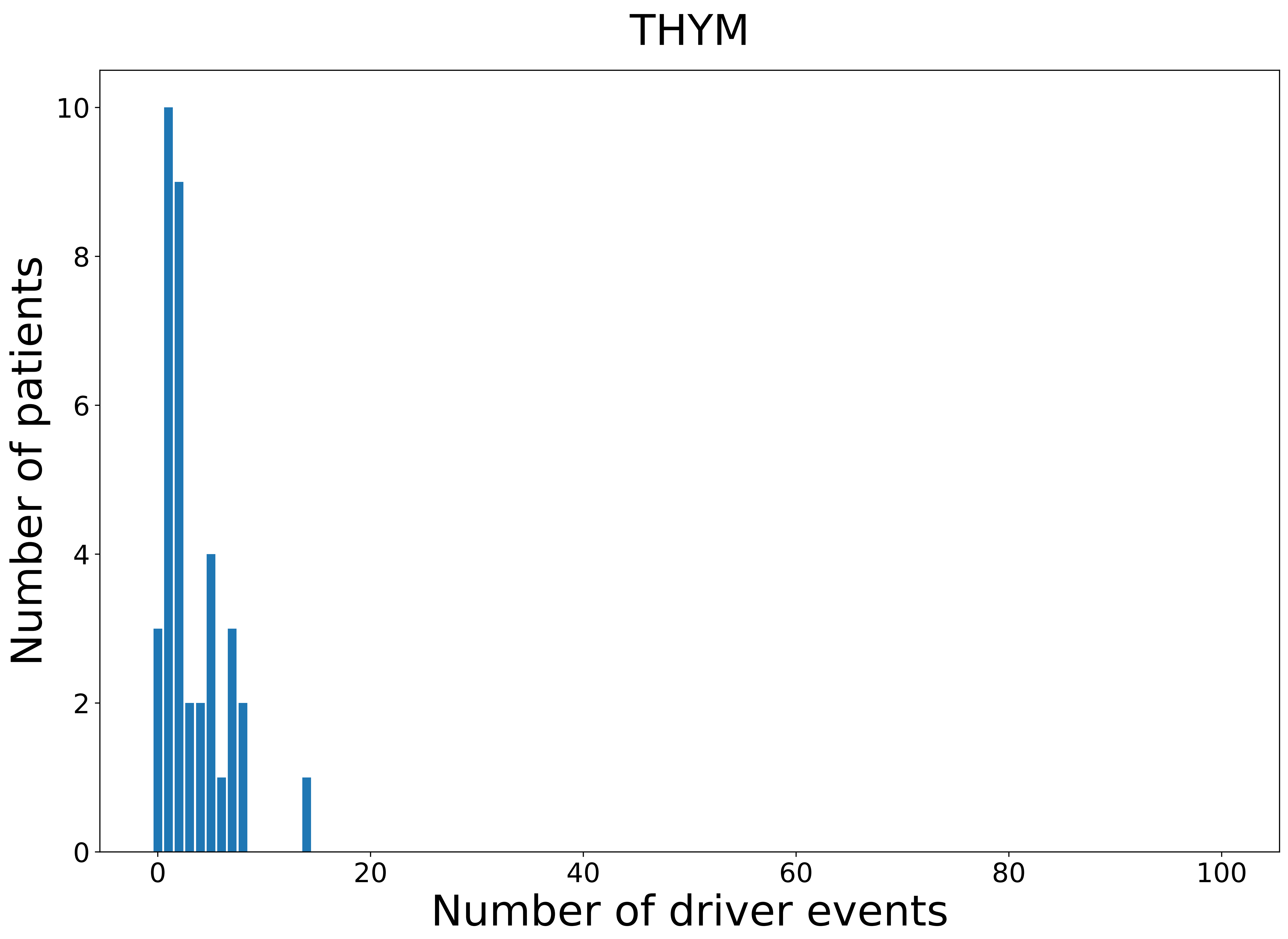

### 2021_8_16_14_9_THYM_FEMALE.pdf

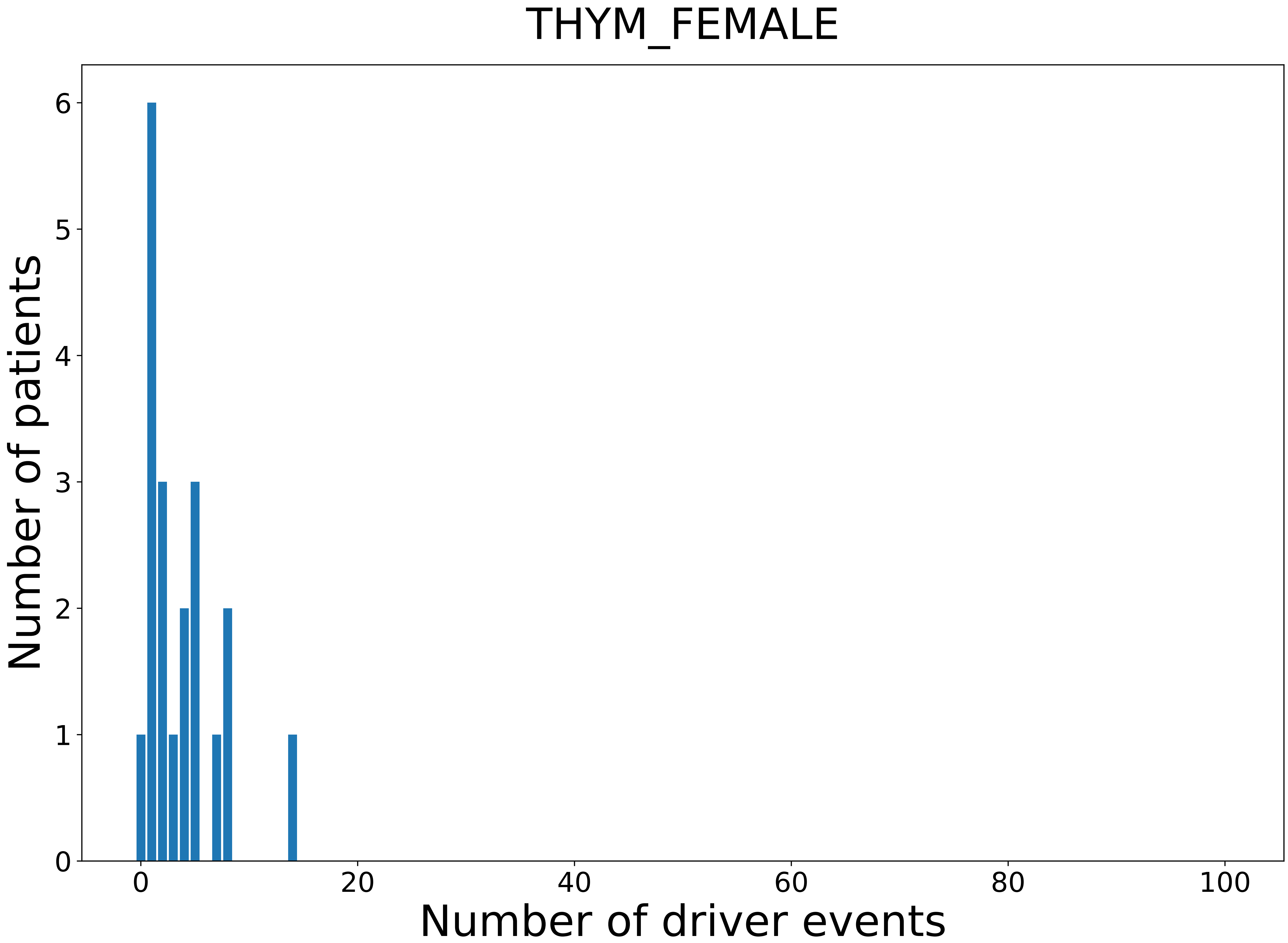

### 2021_8_16_14_9_THYM_MALE.pdf

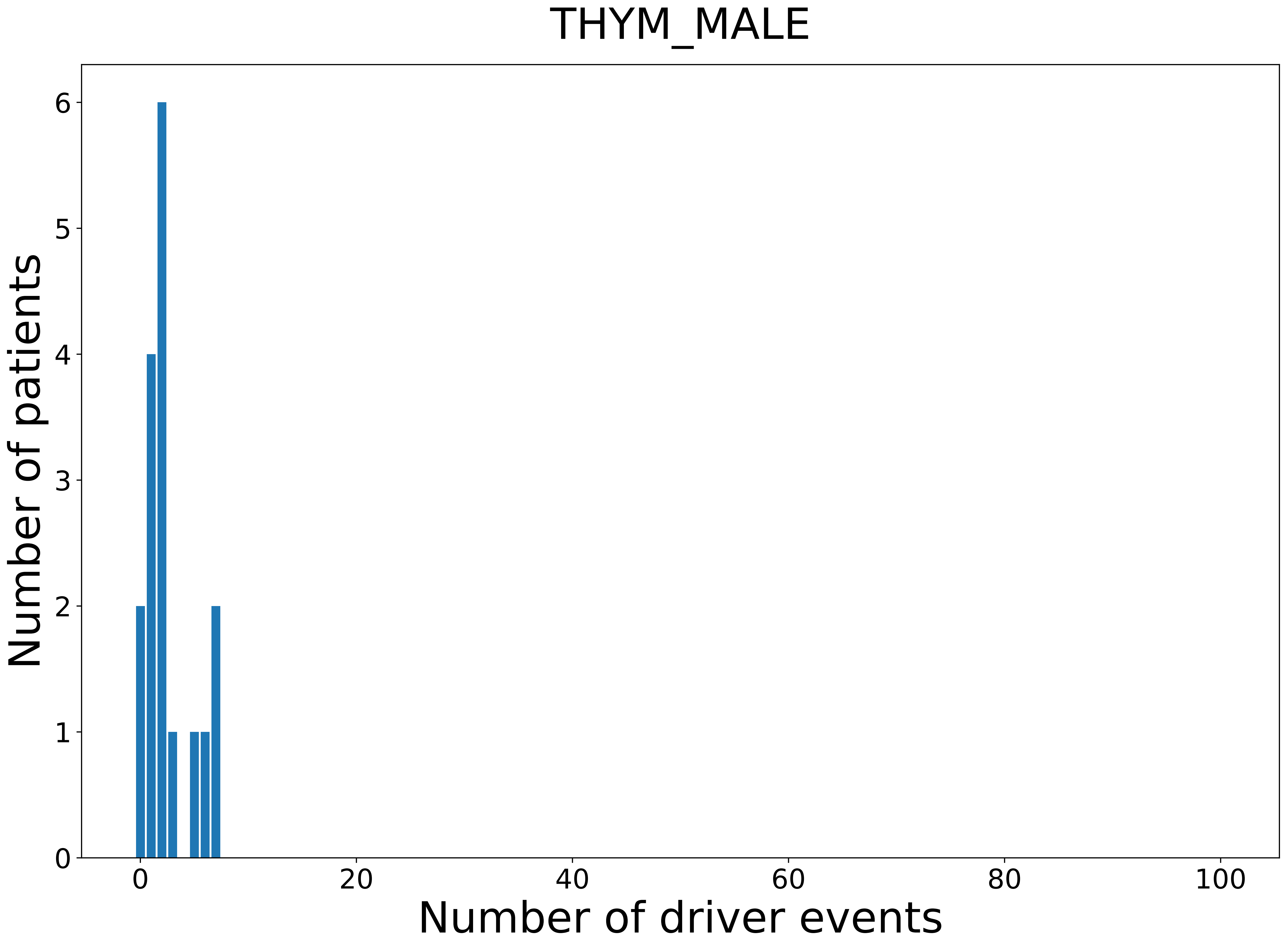

### 2021_8_16_14_9_UCEC_FEMALE.pdf

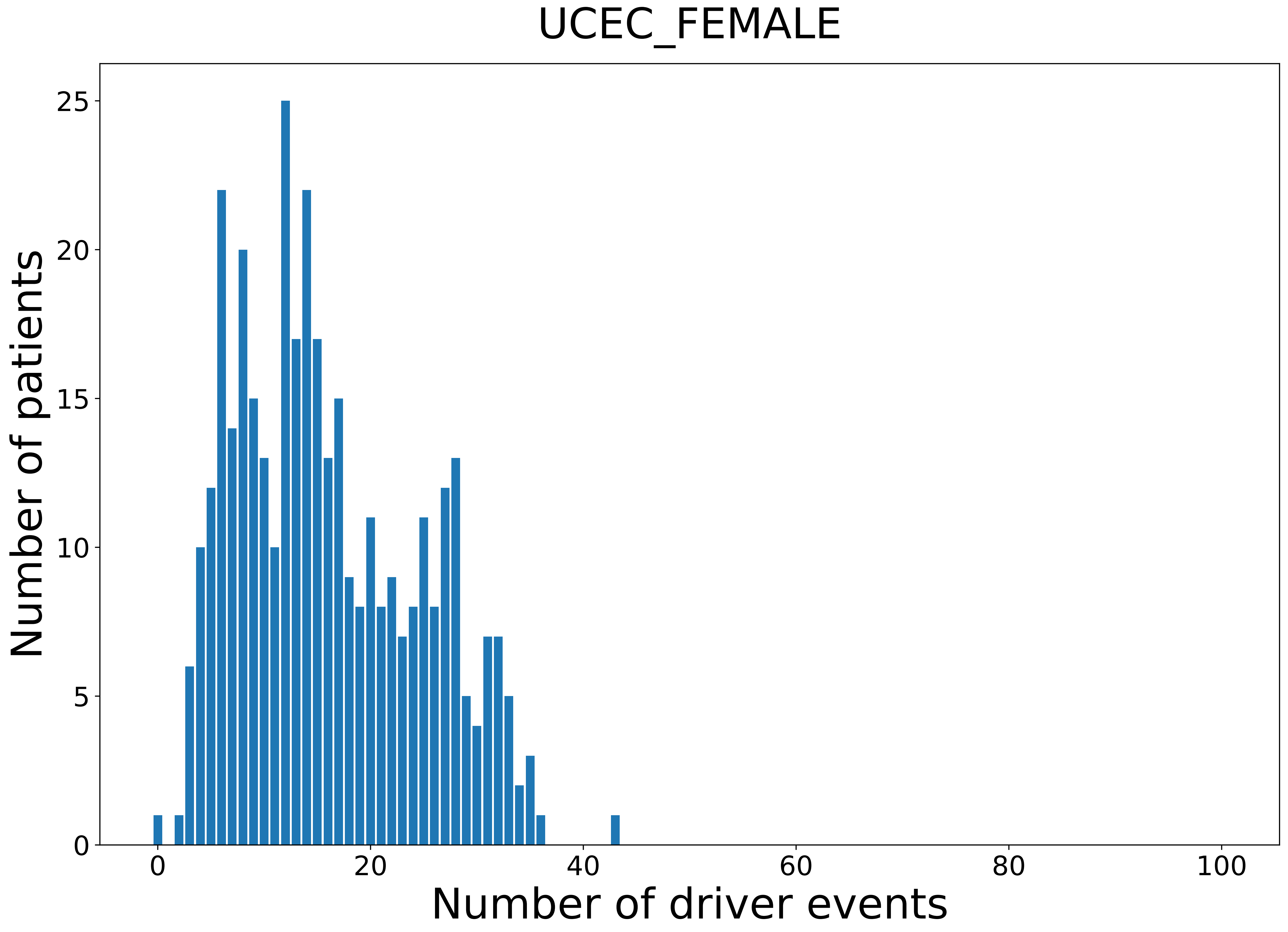

### 2021_8_16_14_9_UCS.pdf

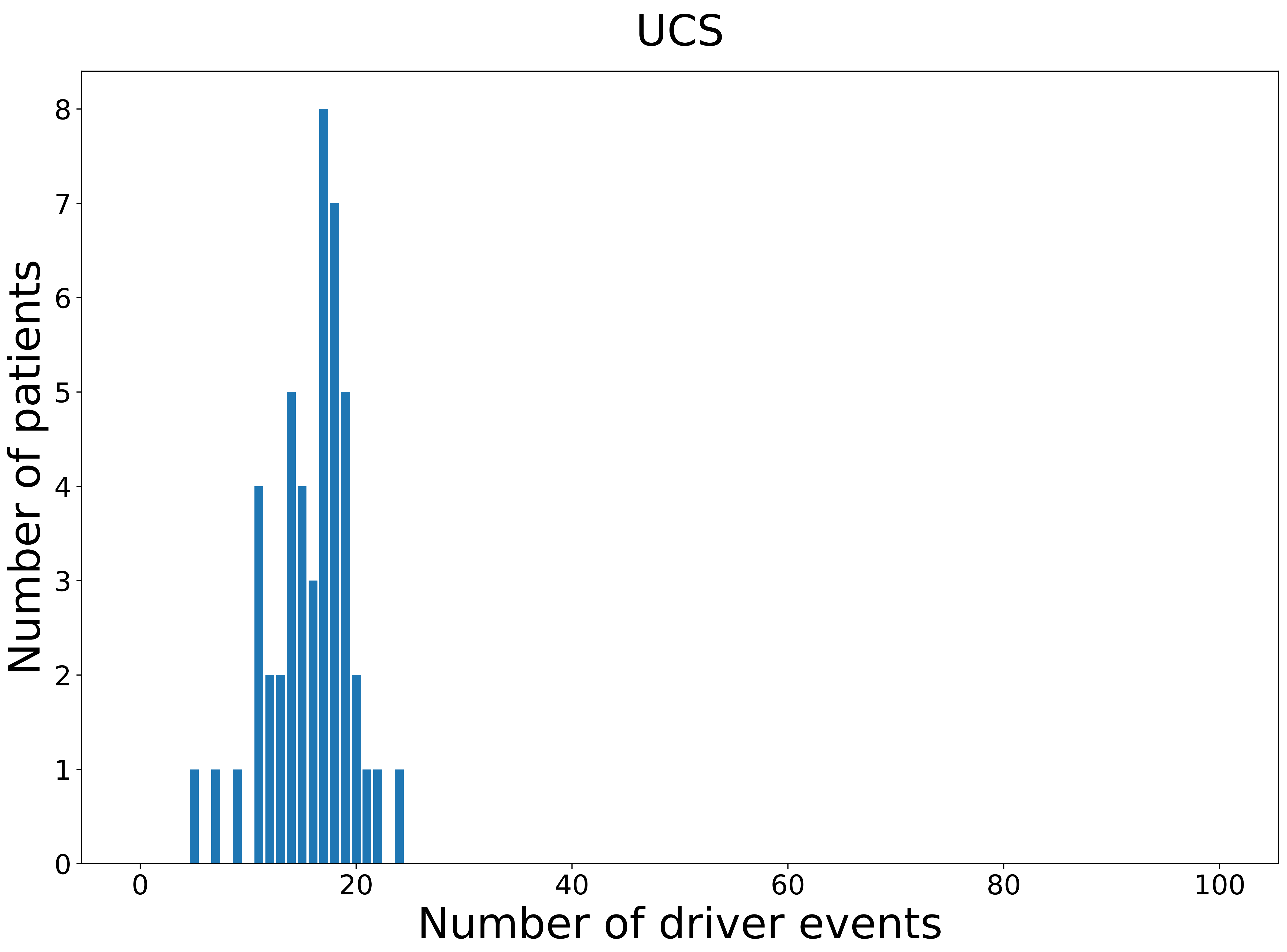

### 2021_8_16_14_9_UCS_FEMALE.pdf

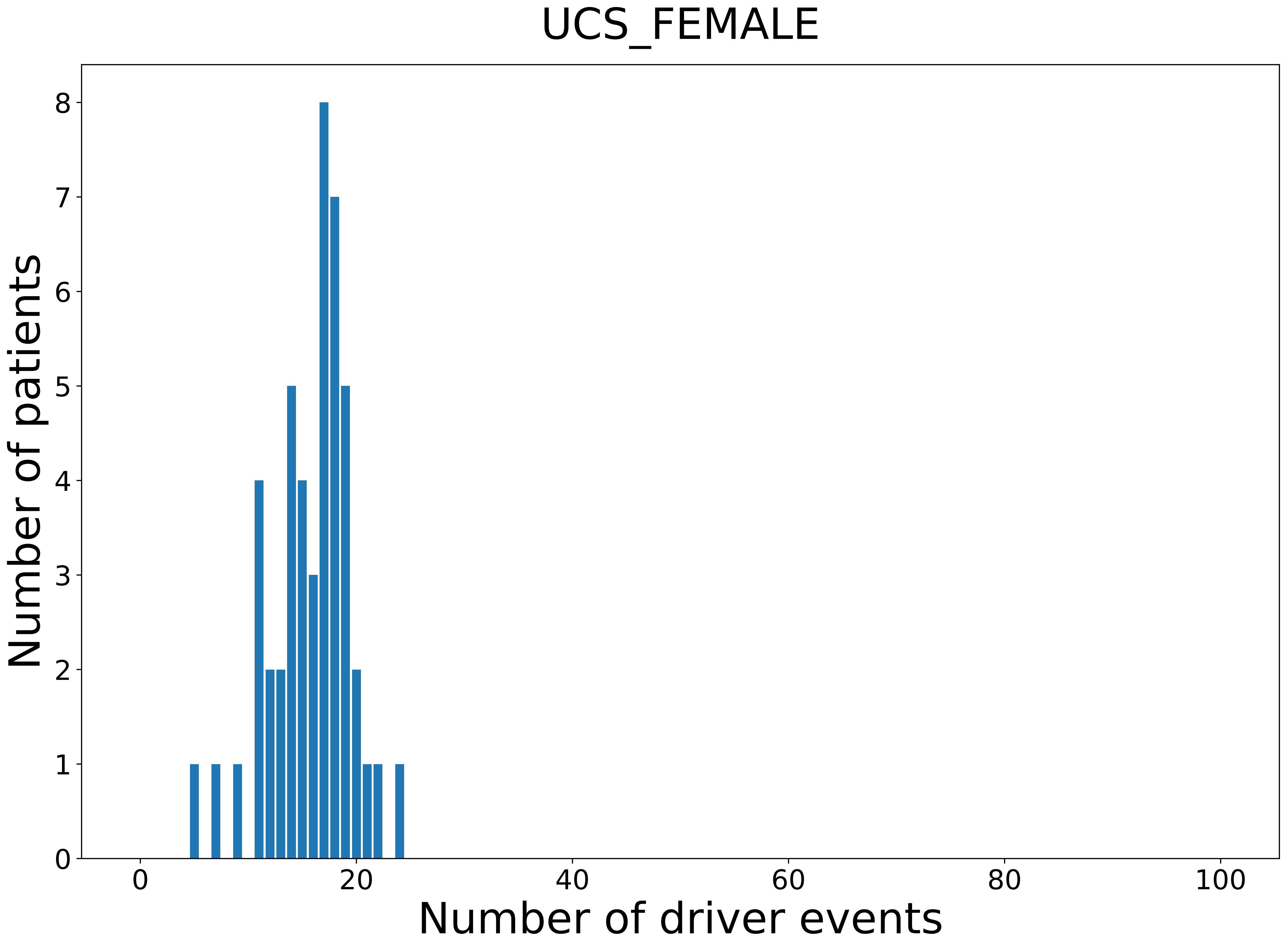

### 2021_8_16_14_9_UVM_MALE.pdf

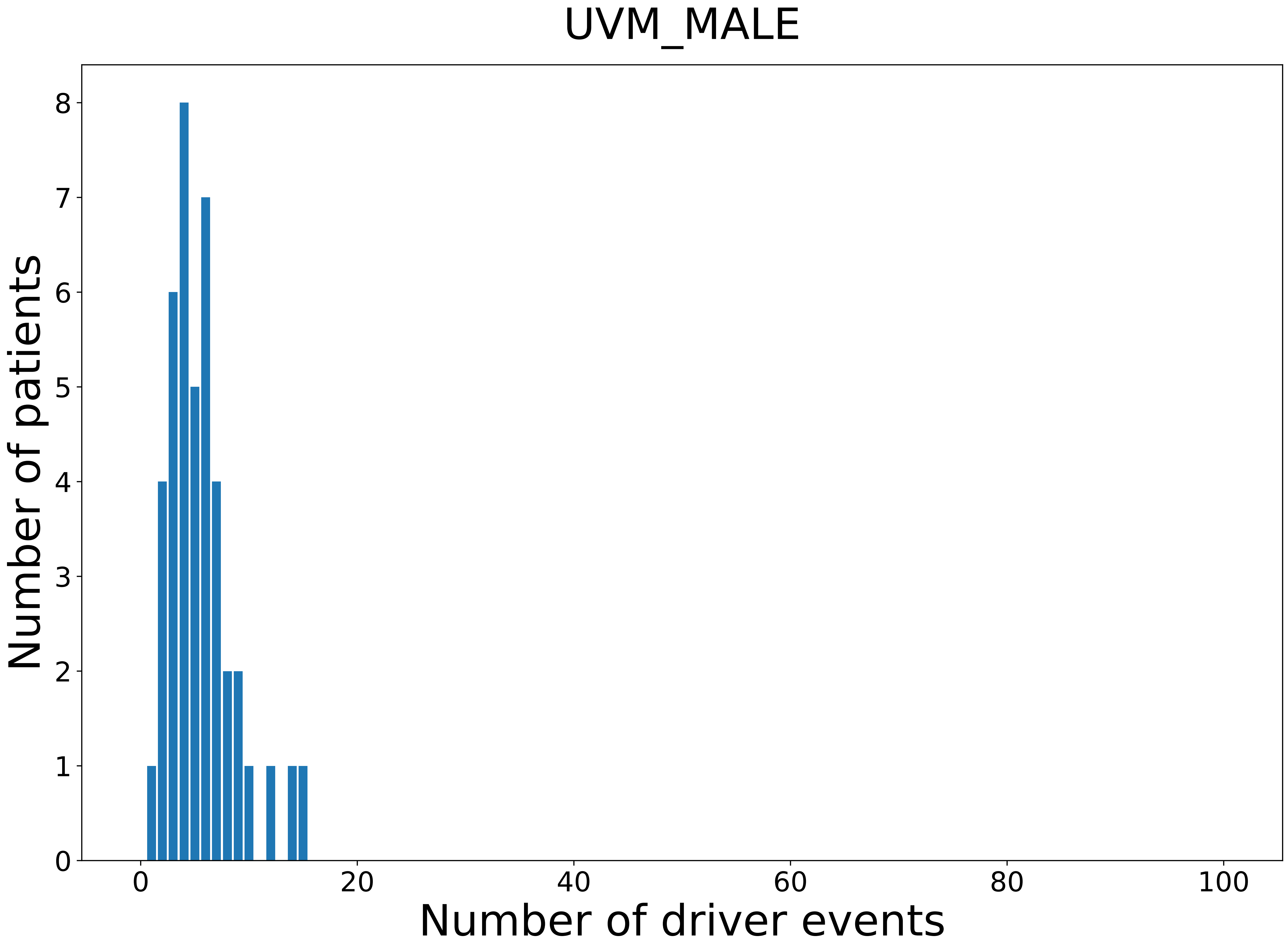
