## Additional file 5 for "Novel Driver Strength Index highlights important cancer genes in TCGA PanCanAtlas patients": Instructions.txt

This script requires Python 3.7 and internet connection

Always open terminal from the folder SNADRIF

First, run the following command to install required libraries:
pip3 install -r requirements.txt

To run the whole pipeline: python3 run.py

To run a particular step: python3 run_one_step.py N
where N is the step number

All downloaded data, intermediate and final results are located in the folder "data"
