## Additional file 6 for "Novel Driver Strength Index highlights important cancer genes in TCGA PanCanAtlas patients": step_14.py

```
import numpy as np
import matplotlib.pyplot as plt
import os
import matplotlib
import scipy.stats
import pandas as pd
from utils.utils import  sumVect, cancers, genders, rows,rows_genes
from collections import Counter
def step14(algo):
	print ("step 14 start")
	input_file = algo[:-4] +'/data/'+algo[:-4] + '_patients.tsv'
	input_file_genes = algo[:-4] +'/data_genes/'+algo[:-4] + '_patients_genes.tsv'
	colors_genes = {0:'lightcoral',1:'red',2:'maroon',3:'cyan',4:'c',5:'teal',6:'orange',7:'darkorange',8:'violet',9:'purple',10:'lightgreen'}
	algo_name = algo[:-4]
	try:
		os.mkdir(algo_name)
	except:
		pass
	try:
		os.mkdir(algo_name+"/cumulative histograms")
	except:
		pass
	try:
		os.mkdir(algo_name+"/data_genes")
	except:
		pass
	try:
		os.mkdir(algo_name+"/genes plots")
	except:
		pass
	try:
		os.mkdir(algo_name+"/genes plots/cohorts")
	except:
		pass

	header = "Cancer type"+"\t"+"\t".join(rows)+"\n"
	header_genes = "Cancer type"+"\t"+"\t".join(rows_genes)+"\n"

	total_events = {}
	male_events ={}
	female_events = {}
	male_events_genes ={}
	female_events_genes = {}
	total_events_genes = {}


	with open(input_file,'r') as inpt:
		inpt.readline().split("\t")
		
		for line in inpt:
			s = line.split("\t")
			if s[1] not in total_events.keys():
				total_events[s[1]] = [0 for i in range(11)]
				total_events[s[1]].append(set())
			total_events[s[1]][-1].add(s[0]) #add patient
			for i in range(11):
				total_events[s[1]][i] += int(s[5+i])

			###males
			if s[2] == "MALE":
				if s[1] not in male_events.keys():
					male_events[s[1]] = [0 for i in range(11)]
					male_events[s[1]].append(set())
				male_events[s[1]][-1].add(s[0]) #add patient
				for i in range(11):
					male_events[s[1]][i] += int(s[5+i])
			###females    
			if s[2] == "FEMALE":
				if s[1] not in female_events.keys():
					female_events[s[1]] = [0 for i in range(11)]
					female_events[s[1]].append(set())
				female_events[s[1]][-1].add(s[0]) #add patient
				for i in range(11):
					female_events[s[1]][i] += int(s[5+i])
					
	total = [0 for i in range(11)]
	all_p = set()
	for i in total_events.keys():
		total =sumVect(total_events[i][:-1],total)
		for j in total_events[i][-1]:
			all_p.add(j)
	total_events['PANCAN'] = total
	total_events['PANCAN'].append(all_p)

	total = [0 for i in range(11)]
	all_p = set()
	for i in male_events.keys():
		total =sumVect(male_events[i][:-1],total)
		for j in male_events[i][-1]:
			all_p.add(j)
	male_events['PANCAN'] = total
	male_events['PANCAN'].append(all_p)
	total = [0 for i in range(11)]
	all_p = set()
	for i in female_events.keys():
		total =sumVect(female_events[i][:-1],total)
		for j in female_events[i][-1]:
			all_p.add(j)
	female_events['PANCAN'] = total
	female_events['PANCAN'].append(all_p)

######################################################
	with open(input_file_genes,'r') as inpt:
		inpt.readline().split("\t")
		
		for line in inpt:
			s = line.split("\t")
			if s[1] not in total_events_genes.keys():
				total_events_genes[s[1]] = [[] for i in range(11)]
			for i in range(10):
				dd = s[5+i]
				if i == 9:
					dd = s[5+i][:-1]
				total_events_genes[s[1]][i].extend(dd.split(','))
				total_events_genes[s[1]][10].extend(dd.split(','))

			###males
			if s[2] == "MALE":
				if s[1] not in male_events_genes.keys():
					male_events_genes[s[1]] = [[] for i in range(11)]
				for i in range(10):
					dd = s[5+i]
					if i == 9:
						dd = s[5+i][:-1]
					male_events_genes[s[1]][i].extend(dd.split(','))
					male_events_genes[s[1]][10].extend(dd.split(','))
					
			###females    
			if s[2] == "FEMALE":
				if s[1] not in female_events_genes.keys():
					female_events_genes[s[1]] = [[] for i in range(11)]
				for i in range(10):
					dd = s[5+i]
					if i == 9:
						dd = s[5+i][:-1]
					female_events_genes[s[1]][i].extend(dd.split(','))
					female_events_genes[s[1]][10].extend(dd.split(','))
	
	tmp = [[] for i in range(11)]
	for i in female_events_genes.keys():
		for j in range(len(female_events_genes[i])):
			tmp[j].extend(female_events_genes[i][j])
	female_events_genes['PANCAN'] = tmp
	
	tmp = [[] for i in range(11)]
	for i in male_events_genes.keys():
		for j in range(len(male_events_genes[i])):
			tmp[j].extend(male_events_genes[i][j])
	male_events_genes['PANCAN'] = tmp

	tmp = [[] for i in range(11)]
	for i in total_events_genes.keys():
		for j in range(len(total_events_genes[i])):
			tmp[j].extend(total_events_genes[i][j])
	total_events_genes['PANCAN'] = tmp

	
	with open(algo_name+'/data/'+algo_name+"_distribution_cohorts.tsv",'w') as out:
		out.write(header)
		types_plot = {}
		for c in total_events.keys():
			out.write(c+"\t")
			num = len(total_events[c][-1])
			a = total_events[c][:-1]
			a = list(map(lambda x:x/num,a))
			types_plot[c] = a
			line = "\t".join(list(map(lambda x:str(x),a)))+"\n"
			out.write(line)
	
#############################
	with open(algo_name+'/data_genes/'+algo_name+"_distribution_cohorts_genes.tsv",'w') as out:
		out.write(header_genes)
		genders_plot_genes = {}
		for c in total_events_genes.keys():
			out.write(c+"\t")
			a = total_events_genes[c]#[:-1]
			a = [Counter(i).most_common() for i in a]
			line = []
			genders_plot_genes[c] = a
			for j in a:
				line.append(",".join([i[0]+":"+str(i[1])  for i in j if i[0] not in ["-",'']]))
			line = "\t".join(line)+"\n"
			out.write(line)
	
	x = total_events_genes.keys()
	data = genders_plot_genes
	for i in x:#gender/stage...
		fig, ax =  plt.subplots(4, 3, squeeze=False)
		
		for j in range(11):#тип мутации
			k = 0
			title = rows_genes[j]
			#fig, ax = plt.subplots()
			xx,yy = j//3,j%3
			if j==8:
				xx,yy = j//3+1,0
			elif j>8:
				xx,yy = j//3,j%3+1
			for driver in data[i][j]:#top 10 genes
				if k <10 and driver[0] not in ['','-']:
					ax[xx][yy].barh(driver[0], int(driver[1]),color = colors_genes[j])
					k+=1
			if k!=0:
				for ik in range(k,9):
					ax[xx][yy].barh(str(ik+100), 0,color = colors_genes[j],linewidth = 0.1)
					plt.setp(ax[xx][yy].get_yticklabels()[-1], visible=False)
			ax[xx][yy].set_xlabel('Number of events', fontdict = {'fontsize':18})
			ax[xx][yy].set_title(title,pad = 18, fontdict = {'fontsize':20})
			fig.set_figwidth(36)
			fig.set_figheight(36)
			fig.set_facecolor('floralwhite')
			ax[xx][yy].set_facecolor('seashell')
			ax[xx][yy].invert_yaxis()
			ax[xx][yy].tick_params(axis="y", labelsize=16)
			ax[xx][yy].tick_params(axis="x", labelsize=13)
			if k == 0:
				ax[xx][yy].set_yticks([])
				ax[xx][yy].set_yticks([], minor=True)
		fig.delaxes(ax[-2][-1])
		plt.savefig(algo_name+"/"+"/genes plots/cohorts/"+algo_name+"_"+"distribution_cohorts_"+i+".pdf",dpi = 600)
	with open(algo_name+'/data_genes/'+algo_name+"_distribution_cohorts_males_genes.tsv",'w') as out:
		out.write(header_genes)
		genders_plot_genes = {}
		for c in male_events_genes.keys():
			out.write(c+"\t")
			a = male_events_genes[c]#[:-1]
			a = [Counter(i).most_common() for i in a]
			line = []
			genders_plot_genes[c] = a
			for j in a:
				line.append(",".join([i[0]+":"+str(i[1])  for i in j if i[0] not in ["-",'']]))
			line = "\t".join(line)+"\n"
			out.write(line)
	x = male_events_genes.keys()
	data = genders_plot_genes

	for i in x:#gender/stage...
		fig, ax =  plt.subplots(4, 3, squeeze=False)
		
		for j in range(11):#тип мутации
			k = 0
			title = rows_genes[j]
			#fig, ax = plt.subplots()
			xx,yy = j//3,j%3
			if j==8:
				xx,yy = j//3+1,0
			elif j>8:
				xx,yy = j//3,j%3+1
			for driver in data[i][j]:#top 10 genes
				if k <10 and driver[0] not in ['','-']:
					ax[xx][yy].barh(driver[0], int(driver[1]),color = colors_genes[j])
					k+=1
			if k!=0:
				for ik in range(k,9):
					ax[xx][yy].barh(str(ik+100), 0,color = colors_genes[j],linewidth = 0.1)
					plt.setp(ax[xx][yy].get_yticklabels()[-1], visible=False)
			ax[xx][yy].set_xlabel('Number of events', fontdict = {'fontsize':18})
			ax[xx][yy].set_title(title,pad = 18, fontdict = {'fontsize':20})
			fig.set_figwidth(36)
			fig.set_figheight(36)
			fig.set_facecolor('floralwhite')
			ax[xx][yy].set_facecolor('seashell')
			ax[xx][yy].invert_yaxis()
			ax[xx][yy].tick_params(axis="y", labelsize=16)
			ax[xx][yy].tick_params(axis="x", labelsize=13)
			if k == 0:
				ax[xx][yy].set_yticks([])
				ax[xx][yy].set_yticks([], minor=True)
		fig.delaxes(ax[-2][-1])
		plt.savefig(algo_name+"/"+"/genes plots/cohorts/"+algo_name+"_"+"distribution_cohorts_males_"+i+".pdf",dpi = 600)
	with open(algo_name+'/data_genes/'+algo_name+"_distribution_cohorts_females_genes.tsv",'w') as out:
		out.write(header_genes)
		genders_plot_genes = {}
		for c in female_events_genes.keys():
			out.write(c+"\t")
			a = female_events_genes[c]#[:-1]
			a = [Counter(i).most_common() for i in a]
			genders_plot_genes[c] = a
			line = []
			for j in a:
				line.append(",".join([i[0]+":"+str(i[1])  for i in j if i[0] not in ["-",'']]))
			line = "\t".join(line)+"\n"
			out.write(line)
	x = female_events_genes.keys()
	data = genders_plot_genes

	for i in x:#gender/stage...
		fig, ax =  plt.subplots(4, 3, squeeze=False)
		
		for j in range(11):#тип мутации
			k = 0
			title = rows_genes[j]
			#fig, ax = plt.subplots()
			xx,yy = j//3,j%3
			if j==8:
				xx,yy = j//3+1,0
			elif j>8:
				xx,yy = j//3,j%3+1
			for driver in data[i][j]:#top 10 genes
				if k <10 and driver[0] not in ['','-']:
					ax[xx][yy].barh(driver[0], int(driver[1]),color = colors_genes[j])
					k+=1
			if k!=0:
				for ik in range(k,9):
					ax[xx][yy].barh(str(ik+100), 0,color = colors_genes[j],linewidth = 0.1)
					plt.setp(ax[xx][yy].get_yticklabels()[-1], visible=False)
			ax[xx][yy].set_xlabel('Number of events', fontdict = {'fontsize':18})
			ax[xx][yy].set_title(title,pad = 18, fontdict = {'fontsize':20})
			fig.set_figwidth(36)
			fig.set_figheight(36)
			fig.set_facecolor('floralwhite')
			ax[xx][yy].set_facecolor('seashell')
			ax[xx][yy].invert_yaxis()
			ax[xx][yy].tick_params(axis="y", labelsize=16)
			ax[xx][yy].tick_params(axis="x", labelsize=13)
			if k == 0:
				ax[xx][yy].set_yticks([])
				ax[xx][yy].set_yticks([], minor=True)
		fig.delaxes(ax[-2][-1])
		plt.savefig(algo_name+"/"+"/genes plots/cohorts/"+algo_name+"_"+"distribution_cohorts_females_"+i+".pdf",dpi = 600)
			
	x = total_events.keys()
	data = types_plot
	fig, ax = plt.subplots()
	title = 'Driver event distribution by cancer type'
	for i in x:
		ax.bar(i, data[i][0],color = 'lightcoral')
		bott = data[i][0]
		ax.bar(i, data[i][1], bottom = bott,color = 'red')
		bott+= data[i][1]
		ax.bar(i, data[i][2], bottom = bott,color = 'maroon')
		bott+= data[i][2]
		ax.bar(i, data[i][3], bottom = bott,color = 'cyan')
		bott+= data[i][3]
		ax.bar(i, data[i][4], bottom = bott,color = 'c')
		bott+= data[i][4]
		ax.bar(i, data[i][5], bottom = bott,color = 'teal')
		bott+= data[i][5]
		ax.bar(i, data[i][6], bottom = bott,color = 'orange')
		bott+= data[i][6]
		ax.bar(i, data[i][7], bottom = bott,color = 'darkorange')
		bott+= data[i][7]
		ax.bar(i, data[i][8], bottom = bott,color = 'violet')
		bott+= data[i][8]
		ax.bar(i, data[i][9], bottom = bott,color = 'purple')
	ax.set_ylabel('Average number of driver events', fontdict = {'fontsize':30})
	ax.set_xlabel('Cohorts', fontdict = {'fontsize':30})
	ax.set_title(title, pad = 18,fontdict = {'fontsize':30})
	plt.xticks(rotation = 90,fontsize = 19)
	plt.yticks(fontsize = 19)
	fig.set_figwidth(20)
	fig.set_figheight(16)
	fig.set_facecolor('floralwhite')
	ax.set_facecolor('seashell')
	plt.savefig(algo_name+"/"+"/cumulative histograms/"+algo_name+"_"+"distribution_cohorts"+".pdf",dpi = 600)

###MALES
	with open(algo_name+'/data/'+algo_name+"_distribution_cohorts_males.tsv",'w') as out:
		out.write(header)
		males_plot= {}
		for c in male_events.keys():
			out.write(c+"\t")
			num = len(male_events[c][-1])
			a = male_events[c][:-1]
			a = list(map(lambda x:x/num,a))
			males_plot[c] = a
			line = "\t".join(list(map(lambda x:str(x),a)))+"\n"
			out.write(line)
	x = male_events.keys()
	data = males_plot
	fig, ax = plt.subplots()
	title = 'Driver event distribution by cancer type in males'
	for i in x:
		ax.bar(i, data[i][0],color = 'lightcoral')
		bott = data[i][0]
		ax.bar(i, data[i][1], bottom = bott,color = 'red')
		bott+= data[i][1]
		ax.bar(i, data[i][2], bottom = bott,color = 'maroon')
		bott+= data[i][2]
		ax.bar(i, data[i][3], bottom = bott,color = 'cyan')
		bott+= data[i][3]
		ax.bar(i, data[i][4], bottom = bott,color = 'c')
		bott+= data[i][4]
		ax.bar(i, data[i][5], bottom = bott,color = 'teal')
		bott+= data[i][5]
		ax.bar(i, data[i][6], bottom = bott,color = 'orange')
		bott+= data[i][6]
		ax.bar(i, data[i][7], bottom = bott,color = 'darkorange')
		bott+= data[i][7]
		ax.bar(i, data[i][8], bottom = bott,color = 'violet')
		bott+= data[i][8]
		ax.bar(i, data[i][9], bottom = bott,color = 'purple')
	ax.set_ylabel('Average number of driver events', fontdict = {'fontsize':30})
	ax.set_xlabel('Cohorts', fontdict = {'fontsize':30})
	ax.set_title(title,pad = 18, fontdict = {'fontsize':30})
	plt.xticks(rotation = 90,fontsize = 19)
	plt.yticks(fontsize = 19)
	fig.set_figwidth(20)
	fig.set_figheight(16)
	fig.set_facecolor('floralwhite')
	ax.set_facecolor('seashell')
	plt.savefig(algo_name+"/"+"/cumulative histograms/"+algo_name+"_"+"distribution_cohorts_males"+".pdf",dpi = 600)

### FEMALES
	with open(algo_name+'/data/'+algo_name+"_distribution_cohorts_females.tsv",'w') as out:
		out.write(header)
		females_plot = {}
		for c in female_events.keys():
			out.write(c+"\t")
			num = len(female_events[c][-1])
			a = female_events[c][:-1]
			a = list(map(lambda x:x/num,a))
			females_plot[c] = a
			line = "\t".join(list(map(lambda x:str(x),a)))+"\n"
			out.write(line)

	x = female_events.keys()
	data = females_plot
	fig, ax = plt.subplots()
	title = 'Driver event distribution by cancer type in females'
	for i in x:
		ax.bar(i, data[i][0],color = 'lightcoral')
		bott = data[i][0]
		ax.bar(i, data[i][1], bottom = bott,color = 'red')
		bott+= data[i][1]
		ax.bar(i, data[i][2], bottom = bott,color = 'maroon')
		bott+= data[i][2]
		ax.bar(i, data[i][3], bottom = bott,color = 'cyan')
		bott+= data[i][3]
		ax.bar(i, data[i][4], bottom = bott,color = 'c')
		bott+= data[i][4]
		ax.bar(i, data[i][5], bottom = bott,color = 'teal')
		bott+= data[i][5]
		ax.bar(i, data[i][6], bottom = bott,color = 'orange')
		bott+= data[i][6]
		ax.bar(i, data[i][7], bottom = bott,color = 'darkorange')
		bott+= data[i][7]
		ax.bar(i, data[i][8], bottom = bott,color = 'violet')
		bott+= data[i][8]
		ax.bar(i, data[i][9], bottom = bott,color = 'purple')
	ax.set_ylabel('Average number of driver events', fontdict = {'fontsize':30})
	ax.set_xlabel('Cohorts', fontdict = {'fontsize':30})
	ax.set_title(title,pad = 18, fontdict = {'fontsize':30})
	plt.xticks(rotation = 90,fontsize = 19)
	plt.yticks(fontsize = 19)
	fig.set_figwidth(20)
	fig.set_figheight(16)
	fig.set_facecolor('floralwhite')
	ax.set_facecolor('seashell')
	plt.savefig(algo_name+"/"+"/cumulative histograms/"+algo_name+"_"+"distribution_cohorts_females"+".pdf",dpi = 600)


	print ("step 14 end")
```
