## Additional file 6 for "Novel Driver Strength Index highlights important cancer genes in TCGA PanCanAtlas patients": step_15.py

```
import os
import numpy as np
import matplotlib
import matplotlib.pyplot as plt
from utils.utils import  sumVect, cancers, genders, rows, sort,rows_genes
import shutil
from collections import Counter,defaultdict

def step15(algo):
	print ("step 15 start")
	input_file = algo[:-4] + '/data/'+algo[:-4] +'_patients.tsv'
	input_file_genes = algo[:-4] +'/data_genes/'+algo[:-4] + '_patients_genes.tsv'
	algo_name = algo[:-4]

	try:
		os.mkdir(algo_name)
	except:
		pass
	try:
		os.mkdir(algo_name+"/cumulative histograms")
	except:
		pass
	try:
		shutil.copyfile('legeng.png',algo_name+"/cumulative histograms/"+'legeng.png')
	except:
		pass
	try:
		shutil.copyfile('legend.png',algo_name+"/cumulative histograms/"+'legend.png')
	except:
		pass
	try:
		os.mkdir(algo_name+"/data_genes")
	except:
		pass
	try:
		os.mkdir(algo_name+"/genes plots")
	except:
		pass
	try:
		os.mkdir(algo_name+"/genes plots/detailed")
	except:
		pass
	try:
		os.mkdir(algo_name+"/genes plots/indices")
	except:
		pass
	colors_genes = {0:'lightcoral',1:'red',2:'maroon',3:'cyan',4:'c',5:'teal',6:'orange',7:'darkorange',8:'violet',9:'purple',10:'lightgreen'}
	header = "Total number of driver events in a patient"+"\t"+"\t".join(rows)+"\n"
	header_genes = "Total number of driver events in a patient"+"\t"+"\t".join(rows_genes)+"\n"
	header_genes_dsi = "\t".join(rows_genes)+"\n"
	total_events = {}
	male_events = {}
	female_events = {}
	brcd_total = {}
	male_events_genes ={}
	female_events_genes = {}
	total_events_genes = {}
	
	
#TOP
	def get_top(data,name):
		a2 = dict()
		with open("data/Homo_sapiens.gene_info",'r') as an:
			for line in an:
				synonims = line.split("\t")[4]
				synonims = synonims.split('|')
				for i in synonims:
					a2[i] = line.split("\t")[1]
				a2[line.split("\t")[2]] = line.split("\t")[1]
		top_ = defaultdict(list)
		for i in range(len(data)):
			for j in range(len(data[i])):
				top_[data[i][j][0]].append(data[i][j][1])
		top = {}
		for i in top_.keys():
			top[i] = max(top_[i])
		top = dict(sorted(top.items(), key=lambda item: item[1], reverse=True))
		with open(name,'w') as out:
			for k in top:
				if top[k]>=0.05:
					try:
						entr = a2[str(k)]
					except:
						entr = ''
					out.write(entr+'\t'+str(k)+'\t'+"{0:.5f}".format(top[k])+'\n')
		return top
#TOP
###########  >10 patients in class
	with open(algo_name+'/data_genes/'+algo_name+"_distribution_cohorts_genes.tsv",'r') as inpt:
		total_10_genes = [[] for i in range(11)]
		for line in inpt:
			s = line.split("\t")
			if s[0] == "PANCAN":
				for i in range(1,len(s)):
					k = s[i].split(",")
					if k!=['']:
						for j in k:
							if int(j.split(":")[1])>=10:
								total_10_genes[i-1].append(j.split(":")[0])
	with open(algo_name+'/data_genes/'+algo_name+"_distribution_cohorts_males_genes.tsv",'r') as inpt:
		male_10_genes = [[] for i in range(11)]
		for line in inpt:
			s = line.split("\t")
			if s[0] == "PANCAN":
				for i in range(1,len(s)):
					k = s[i].split(",")
					if k!=['']:
						for j in k:
							if int(j.split(":")[1])>=10:
								male_10_genes[i-1].append(j.split(":")[0])
	with open(algo_name+'/data_genes/'+algo_name+"_distribution_cohorts_females_genes.tsv",'r') as inpt:
		female_10_genes = [[] for i in range(11)]
		for line in inpt:
			s = line.split("\t")
			if s[0] == "PANCAN":
				for i in range(1,len(s)):
					k = s[i].split(",")
					if k!=['']:
						for j in k:
							if int(j.split(":")[1])>=10:
								female_10_genes[i-1].append(j.split(":")[0])
###########
	
	with open(input_file,'r') as inpt:
		inpt.readline().split("\t")
		for line in inpt:
			s = line.split("\t")
			s[-1] = s[-1][:-1]
			key = s[-1]
			
			if int(key)>=1 and int(key)<=100:
				brcd_total[s[0]] = key
				if key not in total_events.keys():
					total_events[key] = [0 for i in range(11)]
					total_events[key].append(set())
				total_events[key][-1].add(s[0]) #add patient
				for i in range(11):
					total_events[key][i] += int(s[5+i])
			###males
				if s[2] == "MALE":
					if key not in male_events.keys():
						male_events[key] = [0 for i in range(11)]
						male_events[key].append(set())
					male_events[key][-1].add(s[0]) #add patient
					for i in range(11):
						male_events[key][i] += int(s[5+i])
			###females
				if s[2] == "FEMALE":
					if key not in female_events.keys():
						female_events[key] = [0 for i in range(11)]
						female_events[key].append(set())
					female_events[key][-1].add(s[0]) #add patient
					for i in range(11):
						female_events[key][i] += int(s[5+i])
############################################
	with open(input_file_genes,'r') as inpt:
		inpt.readline().split("\t")
		for line in inpt:
			s = line.split("\t")
			if s[0] in brcd_total.keys():
				key = brcd_total[s[0]]
				if key not in total_events_genes.keys():
					total_events_genes[key] = [[] for i in range(11)]
				for i in range(10):
					dd = s[5+i]
					if i == 9:
						dd = s[5+i][:-1]
					total_events_genes[key][i].extend(dd.split(','))
					total_events_genes[key][10].extend(dd.split(','))

			###males
				if s[2] == "MALE":
					if key not in male_events_genes.keys():
						male_events_genes[key] = [[] for i in range(11)]

					for i in range(10):
						dd = s[5+i]
						if i == 9:
							dd = s[5+i][:-1]
						male_events_genes[key][i].extend(dd.split(','))
						male_events_genes[key][10].extend(dd.split(','))
			###females
				if s[2] == "FEMALE":
					if key not in female_events_genes.keys():
						female_events_genes[key] = [[] for i in range(11)]

					for i in range(10):
						dd = s[5+i]
						if i == 9:
							dd = s[5+i][:-1]
						female_events_genes[key][i].extend(dd.split(','))
						female_events_genes[key][10].extend(dd.split(','))
###############################################################


#################write
	with open(algo_name+'/data_genes/'+algo_name+"_distribution_events_detailed_genes.tsv",'w') as out:
		out.write(header_genes)
		genders_plot_genes = {}
		for_DSI = {}#!
		for c in sort(total_events_genes.keys()):
			out.write(c+"\t")
			a = total_events_genes[c]#[:-1]
			a = [Counter(i).most_common() for i in a]
			genders_plot_genes[c] = a
			for_DSI[c] = a#!
			line = []
			for j in a:
				line.append(",".join([i[0]+":"+str(i[1])  for i in j if i[0] not in ["-",'']]))
			line = "\t".join(line)+"\n"
			out.write(line)

############DSI
	with open(algo_name+'/data_genes/'+algo_name+"_distribution_events_detailed_genes_DSI.tsv",'w') as out, open(algo_name+'/data_genes/'+algo_name+"_distribution_events_detailed_genes_NDSI.tsv",'w') as out_ndsi:
		out.write(header_genes_dsi)
		#out.write('DSI'+"\t")
		
		out_ndsi.write(header_genes_dsi)
		#out_ndsi.write('NDSI'+"\t")
		
		genders_plot_DSI = []
		genders_plot_NDSI = []
		DSI_write = []
		NDSI_write = []
		for j in range(11):
			DSI = {}
			NDSI = {}
			DSI_for_NDSI = {}
			for c in sort(total_events_genes.keys()):
				i = int(c)
				p_i = len(total_events[c][-1])###!!!!!! [c][-1]
				for k in for_DSI[c][j]:#k=gene
					
					p_a_i = int(k[1])
					if k[0] not in ["-",'']:
						if k[0] not in DSI.keys(): DSI[k[0]] = 0
						DSI[k[0]]+=p_a_i/(i*p_i)
						if k[0] in total_10_genes[j]:
							if k[0] not in NDSI.keys(): 
								NDSI[k[0]] = 0
								DSI_for_NDSI[k[0]] = 0
							NDSI[k[0]]+=p_a_i/(p_i)
							DSI_for_NDSI[k[0]]+=p_a_i/(i*p_i)
						
						
			for g in NDSI.keys():
				NDSI[g] = DSI_for_NDSI[g]/NDSI[g]
			DSI = dict(sorted(DSI.items(), key=lambda item: item[1], reverse=True))
			NDSI = dict(sorted(NDSI.items(), key=lambda item: item[1], reverse=True))
			for jj in DSI:
				if jj not in ["-",'']:
					dsi_str = "{0:.5f}".format(DSI[jj])
					#out.write(jj+":"+str(dsi_str)+',')
			for jj in NDSI:
				if jj not in ["-",'']:
					Ndsi_str = "{0:.5f}".format(NDSI[jj])
					#out_ndsi.write(jj+":"+str(Ndsi_str)+',')
			#out.write("\t")
			#out_ndsi.write("\t")
			DSI_ = []
			NDSI_ = []
			for jj in DSI:
				DSI_.append((jj,DSI[jj]))
			for jj in NDSI:
				NDSI_.append((jj,NDSI[jj]))
			DSI_write.append(DSI_)
			NDSI_write.append(NDSI_)
			genders_plot_DSI.append(DSI_)
			genders_plot_NDSI.append(NDSI_)
		max_deep = max([len(DSI_write[k]) for k in range(len(DSI_write))])
		for i in range(max_deep):# rows
			stt =[]
			for j in range(len(DSI_write)):#columns
				if len(DSI_write[j])<i+1:
					ss = ''
				else:
					dsi_str = "{0:.5f}".format(DSI_write[j][i][1])
					ss = DSI_write[j][i][0]+':'+dsi_str
				stt.append(ss)
			out.write("\t".join(stt)+"\n")
		for i in range(max_deep):# rows
			stt =[]
			for j in range(len(NDSI_write)):#columns
				if len(NDSI_write[j])<i+1:
					ss = ''
				else:
					ndsi_str = "{0:.5f}".format(NDSI_write[j][i][1])
					ss = NDSI_write[j][i][0]+':'+ndsi_str
				stt.append(ss)
			out_ndsi.write("\t".join(stt)+"\n")

	fig, ax =  plt.subplots(4, 3, squeeze=False)
	data = genders_plot_DSI
	get_top(data,algo_name+'/data_genes/'+algo_name+"_distribution_events_detailed_genes_DSI_top.tsv")
	for j in range(11):#тип мутации
		k = 0
		title = rows_genes[j]
		#fig, ax = plt.subplots()
		xx,yy = j//3,j%3
		if j==8:
			xx,yy = j//3+1,0
		elif j>8:
			xx,yy = j//3,j%3+1
		for driver in data[j]:#top 10 genes
			if k <10 and driver[0] not in ['','-']:
				
				ax[xx][yy].barh(driver[0], float(driver[1]),color = colors_genes[j],linewidth = 0.1)
				k+=1
		if k!=0:
			for ik in range(k,9):
				ax[xx][yy].barh(str(ik+100), 0,color = colors_genes[j],linewidth = 0.1)
				plt.setp(ax[xx][yy].get_yticklabels()[-1], visible=False)
		ax[xx][yy].set_xlabel('DSI', fontdict = {'fontsize':18})
		ax[xx][yy].set_title(title,pad = 18, fontdict = {'fontsize':20})
		fig.set_figwidth(36)
		fig.set_figheight(36)
		fig.set_facecolor('floralwhite')
		ax[xx][yy].set_facecolor('seashell')
		ax[xx][yy].invert_yaxis()
		ax[xx][yy].tick_params(axis="y", labelsize=16)
		ax[xx][yy].tick_params(axis="x", labelsize=13)
		if k == 0:
			ax[xx][yy].set_yticks([])
			ax[xx][yy].set_yticks([], minor=True)
	fig.delaxes(ax[-2][-1])
	#fig.delaxes(ax[-1][-1])
	plt.savefig(algo_name+"/"+"/genes plots/indices/"+algo_name+"_"+"distribution_events_detailed_DSI"+".pdf",dpi = 600)
#NDSI
	fig, ax =  plt.subplots(4, 3, squeeze=False)
	data = genders_plot_NDSI
	get_top(data,algo_name+'/data_genes/'+algo_name+"_distribution_events_detailed_genes_NDSI_top.tsv")
	for j in range(11):#тип мутации
		k = 0
		title = rows_genes[j]
		#fig, ax = plt.subplots()
		xx,yy = j//3,j%3
		if j==8:
			xx,yy = j//3+1,0
		elif j>8:
			xx,yy = j//3,j%3+1
		for driver in data[j]:#top 10 genes
			if k <10 and driver[0] not in ['','-']:
				
				ax[xx][yy].barh(driver[0], float(driver[1]),color = colors_genes[j],linewidth = 0.1)
				k+=1
		if k!=0:
			for ik in range(k,9):
				ax[xx][yy].barh(str(ik+100), 0,color = colors_genes[j],linewidth = 0.1)
				plt.setp(ax[xx][yy].get_yticklabels()[-1], visible=False)
		ax[xx][yy].set_xlabel('NDSI', fontdict = {'fontsize':18})
		ax[xx][yy].set_title(title,pad = 18, fontdict = {'fontsize':20})
		fig.set_figwidth(36)
		fig.set_figheight(36)
		fig.set_facecolor('floralwhite')
		ax[xx][yy].set_facecolor('seashell')
		ax[xx][yy].invert_yaxis()
		ax[xx][yy].tick_params(axis="y", labelsize=16)
		ax[xx][yy].tick_params(axis="x", labelsize=13)
		if k == 0:
			ax[xx][yy].set_yticks([])
			ax[xx][yy].set_yticks([], minor=True)
	fig.delaxes(ax[-2][-1])
	#fig.delaxes(ax[-1][-1])
	plt.savefig(algo_name+"/"+"/genes plots/indices/"+algo_name+"_"+"distribution_events_detailed_NDSI"+".pdf",dpi = 600)
	
#######DSI
	x = genders_plot_genes.keys()
	data = genders_plot_genes
			
	for i in x:#gender/stage...
		fig, ax =  plt.subplots(4, 3, squeeze=False)
		
		for j in range(11):#тип мутации
			k = 0
			title = rows_genes[j]
			#fig, ax = plt.subplots()
			xx,yy = j//3,j%3
			if j==8:
				xx,yy = j//3+1,0
			elif j>8:
				xx,yy = j//3,j%3+1
			for driver in data[i][j]:#top 10 genes
				if k <10 and driver[0] not in ['','-']:
					ax[xx][yy].barh(driver[0], int(driver[1]),color = colors_genes[j],linewidth = 0.1)
					k+=1
			if k!=0:
				for ik in range(k,9):
					ax[xx][yy].barh(str(ik+100), 0,color = colors_genes[j],linewidth = 0.1)
					plt.setp(ax[xx][yy].get_yticklabels()[-1], visible=False)
			ax[xx][yy].set_xlabel('Number of events', fontdict = {'fontsize':18})
			ax[xx][yy].set_title(title,pad = 18, fontdict = {'fontsize':20})
			fig.set_figwidth(36)
			fig.set_figheight(36)
			fig.set_facecolor('floralwhite')
			ax[xx][yy].set_facecolor('seashell')
			ax[xx][yy].invert_yaxis()
			ax[xx][yy].tick_params(axis="y", labelsize=16)
			ax[xx][yy].tick_params(axis="x", labelsize=13)
			if k == 0:
				ax[xx][yy].set_yticks([])
				ax[xx][yy].set_yticks([], minor=True)
		fig.delaxes(ax[-2][-1])
		plt.savefig(algo_name+"/"+"/genes plots/detailed/"+algo_name+"_"+"distribution_events_detailed_"+i+".pdf",dpi = 600)
	with open(algo_name+'/data_genes/'+algo_name+"_distribution_events_detailed_males_genes.tsv",'w') as out:
		out.write(header_genes)
		genders_plot_genes = {}
		for_DSI = {}#!
		for c in sort(male_events_genes.keys()):
			out.write(c+"\t")
			a = male_events_genes[c]#[:-1]
			a = [Counter(i).most_common() for i in a]
			line = []
			genders_plot_genes[c] = a
			for_DSI[c] = a#!
			for j in a:
				line.append(",".join([i[0]+":"+str(i[1])  for i in j if i[0] not in ["-",'']]))
			line = "\t".join(line)+"\n"
			out.write(line)
	
############DSI
	with open(algo_name+'/data_genes/'+algo_name+"_distribution_events_detailed_males_genes_DSI.tsv",'w') as out, open(algo_name+'/data_genes/'+algo_name+"_distribution_events_detailed_males_genes_NDSI.tsv",'w') as out_ndsi:
		out.write(header_genes_dsi)
		#out.write('DSI'+"\t")
		
		out_ndsi.write(header_genes_dsi)
		#out_ndsi.write('NDSI'+"\t")
		
		genders_plot_DSI = []
		genders_plot_NDSI = []
		DSI_write = []
		NDSI_write = []
		for j in range(11):
			DSI = {}
			NDSI = {}
			DSI_for_NDSI = {}
			for c in sort(male_events_genes.keys()):
				i = int(c)
				p_i = len(male_events[c][-1])###!!!!!! [c][-1]
				for k in for_DSI[c][j]:#k=gene
					
					p_a_i = int(k[1])
					if k[0] not in ["-",'']:
						if k[0] not in DSI.keys(): DSI[k[0]] = 0
						DSI[k[0]]+=p_a_i/(i*p_i)
						if k[0] in male_10_genes[j]:
							if k[0] not in NDSI.keys(): 
								NDSI[k[0]] = 0
								DSI_for_NDSI[k[0]] = 0
							NDSI[k[0]]+=p_a_i/(p_i)
							DSI_for_NDSI[k[0]]+=p_a_i/(i*p_i)
						
						
			for g in NDSI.keys():
				NDSI[g] = DSI_for_NDSI[g]/NDSI[g]
			DSI = dict(sorted(DSI.items(), key=lambda item: item[1], reverse=True))
			NDSI = dict(sorted(NDSI.items(), key=lambda item: item[1], reverse=True))
			for jj in DSI:
				if jj not in ["-",'']:
					dsi_str = "{0:.5f}".format(DSI[jj])
					#out.write(jj+":"+str(dsi_str)+',')
			for jj in NDSI:
				if jj not in ["-",'']:
					Ndsi_str = "{0:.5f}".format(NDSI[jj])
					#out_ndsi.write(jj+":"+str(Ndsi_str)+',')
			#out.write("\t")
			#out_ndsi.write("\t")
			DSI_ = []
			NDSI_ = []
			for jj in DSI:
				DSI_.append((jj,DSI[jj]))
			for jj in NDSI:
				NDSI_.append((jj,NDSI[jj]))
			DSI_write.append(DSI_)
			NDSI_write.append(NDSI_)
			genders_plot_DSI.append(DSI_)
			genders_plot_NDSI.append(NDSI_)
		max_deep = max([len(DSI_write[k]) for k in range(len(DSI_write))])
		for i in range(max_deep):# rows
			stt =[]
			for j in range(len(DSI_write)):#columns
				if len(DSI_write[j])<i+1:
					ss = ''
				else:
					dsi_str = "{0:.5f}".format(DSI_write[j][i][1])
					ss = DSI_write[j][i][0]+':'+dsi_str
				stt.append(ss)
			out.write("\t".join(stt)+"\n")
		for i in range(max_deep):# rows
			stt =[]
			for j in range(len(NDSI_write)):#columns
				if len(NDSI_write[j])<i+1:
					ss = ''
				else:
					ndsi_str = "{0:.5f}".format(NDSI_write[j][i][1])
					ss = NDSI_write[j][i][0]+':'+ndsi_str
				stt.append(ss)
			out_ndsi.write("\t".join(stt)+"\n")
	
	fig, ax =  plt.subplots(4, 3, squeeze=False)
	data = genders_plot_DSI
	get_top(data,algo_name+'/data_genes/'+algo_name+"_distribution_events_detailed_males_genes_DSI_top.tsv")
	for j in range(11):#тип мутации
		k = 0
		title = rows_genes[j]
		#fig, ax = plt.subplots()
		xx,yy = j//3,j%3
		if j==8:
			xx,yy = j//3+1,0
		elif j>8:
			xx,yy = j//3,j%3+1
		for driver in data[j]:#top 10 genes
			if k <10 and driver[0] not in ['','-']:
				
				ax[xx][yy].barh(driver[0], float(driver[1]),color = colors_genes[j],linewidth = 0.1)
				k+=1
		if k!=0:
			for ik in range(k,9):
				ax[xx][yy].barh(str(ik+100), 0,color = colors_genes[j],linewidth = 0.1)
				plt.setp(ax[xx][yy].get_yticklabels()[-1], visible=False)
		ax[xx][yy].set_xlabel('DSI', fontdict = {'fontsize':18})
		ax[xx][yy].set_title(title,pad = 18, fontdict = {'fontsize':20})
		fig.set_figwidth(36)
		fig.set_figheight(36)
		fig.set_facecolor('floralwhite')
		ax[xx][yy].set_facecolor('seashell')
		ax[xx][yy].invert_yaxis()
		ax[xx][yy].tick_params(axis="y", labelsize=16)
		ax[xx][yy].tick_params(axis="x", labelsize=13)
		if k == 0:
			ax[xx][yy].set_yticks([])
			ax[xx][yy].set_yticks([], minor=True)
	fig.delaxes(ax[-2][-1])
	
	plt.savefig(algo_name+"/"+"/genes plots/indices/"+algo_name+"_"+"distribution_events_detailed_DSI_males"+".pdf",dpi = 600)
#NDSI
	fig, ax =  plt.subplots(4, 3, squeeze=False)
	data = genders_plot_NDSI
	get_top(data,algo_name+'/data_genes/'+algo_name+"_distribution_events_detailed_males_genes_NDSI_top.tsv")
	for j in range(11):#тип мутации
		k = 0
		title = rows_genes[j]
		#fig, ax = plt.subplots()
		xx,yy = j//3,j%3
		if j==8:
			xx,yy = j//3+1,0
		elif j>8:
			xx,yy = j//3,j%3+1
		for driver in data[j]:#top 10 genes
			if k <10 and driver[0] not in ['','-']:
				
				ax[xx][yy].barh(driver[0], float(driver[1]),color = colors_genes[j],linewidth = 0.1)
				k+=1
		if k!=0:
			for ik in range(k,9):
				ax[xx][yy].barh(str(ik+100), 0,color = colors_genes[j],linewidth = 0.1)
				plt.setp(ax[xx][yy].get_yticklabels()[-1], visible=False)
		ax[xx][yy].set_xlabel('NDSI', fontdict = {'fontsize':18})
		ax[xx][yy].set_title(title,pad = 18, fontdict = {'fontsize':20})
		fig.set_figwidth(36)
		fig.set_figheight(36)
		fig.set_facecolor('floralwhite')
		ax[xx][yy].set_facecolor('seashell')
		ax[xx][yy].invert_yaxis()
		ax[xx][yy].tick_params(axis="y", labelsize=16)
		ax[xx][yy].tick_params(axis="x", labelsize=13)
		if k == 0:
			ax[xx][yy].set_yticks([])
			ax[xx][yy].set_yticks([], minor=True)
	fig.delaxes(ax[-2][-1])
	
	plt.savefig(algo_name+"/"+"/genes plots/indices/"+algo_name+"_"+"distribution_events_detailed_NDSI_males"+".pdf",dpi = 600)
#######DSI
	
	x = genders_plot_genes.keys()
	data = genders_plot_genes

	for i in x:#gender/stage...
		fig, ax =  plt.subplots(4, 3, squeeze=False)
		
		for j in range(11):#тип мутации
			k = 0
			title = rows_genes[j]
			#fig, ax = plt.subplots()
			xx,yy = j//3,j%3
			if j==8:
				xx,yy = j//3+1,0
			elif j>8:
				xx,yy = j//3,j%3+1
			for driver in data[i][j]:#top 10 genes
				if k <10 and driver[0] not in ['','-']:
					ax[xx][yy].barh(driver[0], int(driver[1]),color = colors_genes[j],linewidth = 0.1)
					k+=1
			if k!=0:
				for ik in range(k,9):
					ax[xx][yy].barh(str(ik+100), 0,color = colors_genes[j],linewidth = 0.1)
					plt.setp(ax[xx][yy].get_yticklabels()[-1], visible=False)
			ax[xx][yy].set_xlabel('Number of events', fontdict = {'fontsize':18})
			ax[xx][yy].set_title(title,pad = 18, fontdict = {'fontsize':20})
			fig.set_figwidth(36)
			fig.set_figheight(36)
			fig.set_facecolor('floralwhite')
			ax[xx][yy].set_facecolor('seashell')
			ax[xx][yy].invert_yaxis()
			ax[xx][yy].tick_params(axis="y", labelsize=16)
			ax[xx][yy].tick_params(axis="x", labelsize=13)
			if k == 0:
				ax[xx][yy].set_yticks([])
				ax[xx][yy].set_yticks([], minor=True)
		fig.delaxes(ax[-2][-1])
		plt.savefig(algo_name+"/"+"/genes plots/detailed/"+algo_name+"_"+"distribution_events_detailed_males_"+i+".pdf",dpi = 600)
	with open(algo_name+'/data_genes/'+algo_name+"_distribution_events_detailed_females_genes.tsv",'w') as out:
		out.write(header_genes)
		genders_plot_genes = {}
		for_DSI = {}#!
		for c in sort(female_events_genes.keys()):
			out.write(c+"\t")
			a = female_events_genes[c]#[:-1]
			a = [Counter(i).most_common() for i in a]
			genders_plot_genes[c] = a
			line = []
			for_DSI[c] = a#!
			for j in a:
				line.append(",".join([i[0]+":"+str(i[1])  for i in j if i[0] not in ["-",'']]))
			line = "\t".join(line)+"\n"
			out.write(line)
			
###########DSI
	with open(algo_name+'/data_genes/'+algo_name+"_distribution_events_detailed_females_genes_DSI.tsv",'w') as out, open(algo_name+'/data_genes/'+algo_name+"_distribution_events_detailed_females_genes_NDSI.tsv",'w') as out_ndsi:
		out.write(header_genes_dsi)
		#out.write('DSI'+"\t")
		
		out_ndsi.write(header_genes_dsi)
		#out_ndsi.write('NDSI'+"\t")
		
		genders_plot_DSI = []
		genders_plot_NDSI = []
		DSI_write = []
		NDSI_write = []
		for j in range(11):
			DSI = {}
			NDSI = {}
			DSI_for_NDSI = {}
			for c in sort(female_events_genes.keys()):
				i = int(c)
				p_i = len(female_events[c][-1])###!!!!!! [c][-1]
				for k in for_DSI[c][j]:#k=gene
					
					p_a_i = int(k[1])
					if k[0] not in ["-",'']:
						if k[0] not in DSI.keys(): DSI[k[0]] = 0
						DSI[k[0]]+=p_a_i/(i*p_i)
						if k[0] in female_10_genes[j]:
							if k[0] not in NDSI.keys(): 
								NDSI[k[0]] = 0
								DSI_for_NDSI[k[0]] = 0
							NDSI[k[0]]+=p_a_i/(p_i)
							DSI_for_NDSI[k[0]]+=p_a_i/(i*p_i)
						
						
			for g in NDSI.keys():
				NDSI[g] = DSI_for_NDSI[g]/NDSI[g]
			DSI = dict(sorted(DSI.items(), key=lambda item: item[1], reverse=True))
			NDSI = dict(sorted(NDSI.items(), key=lambda item: item[1], reverse=True))
			for jj in DSI:
				if jj not in ["-",'']:
					dsi_str = "{0:.5f}".format(DSI[jj])
					#out.write(jj+":"+str(dsi_str)+',')
			for jj in NDSI:
				if jj not in ["-",'']:
					Ndsi_str = "{0:.5f}".format(NDSI[jj])
					#out_ndsi.write(jj+":"+str(Ndsi_str)+',')
			#out.write("\t")
			#out_ndsi.write("\t")
			DSI_ = []
			NDSI_ = []
			for jj in DSI:
				DSI_.append((jj,DSI[jj]))
			for jj in NDSI:
				NDSI_.append((jj,NDSI[jj]))
			DSI_write.append(DSI_)
			NDSI_write.append(NDSI_)
			genders_plot_DSI.append(DSI_)
			genders_plot_NDSI.append(NDSI_)
		max_deep = max([len(DSI_write[k]) for k in range(len(DSI_write))])
		for i in range(max_deep):# rows
			stt =[]
			for j in range(len(DSI_write)):#columns
				if len(DSI_write[j])<i+1:
					ss = ''
				else:
					dsi_str = "{0:.5f}".format(DSI_write[j][i][1])
					ss = DSI_write[j][i][0]+':'+dsi_str
				stt.append(ss)
			out.write("\t".join(stt)+"\n")
		for i in range(max_deep):# rows
			stt =[]
			for j in range(len(NDSI_write)):#columns
				if len(NDSI_write[j])<i+1:
					ss = ''
				else:
					ndsi_str = "{0:.5f}".format(NDSI_write[j][i][1])
					ss = NDSI_write[j][i][0]+':'+ndsi_str
				stt.append(ss)
			out_ndsi.write("\t".join(stt)+"\n")
	fig, ax =  plt.subplots(4, 3, squeeze=False)
	data = genders_plot_DSI
	get_top(data,algo_name+'/data_genes/'+algo_name+"_distribution_events_detailed_females_genes_DSI_top.tsv")
	for j in range(11):#тип мутации
		k = 0
		title = rows_genes[j]
		#fig, ax = plt.subplots()
		xx,yy = j//3,j%3
		if j==8:
			xx,yy = j//3+1,0
		elif j>8:
			xx,yy = j//3,j%3+1
		for driver in data[j]:#top 10 genes
			if k <10 and driver[0] not in ['','-']:
				
				ax[xx][yy].barh(driver[0], float(driver[1]),color = colors_genes[j],linewidth = 0.1)
				k+=1
		if k!=0:
			for ik in range(k,9):
				ax[xx][yy].barh(str(ik+100), 0,color = colors_genes[j],linewidth = 0.1)
				plt.setp(ax[xx][yy].get_yticklabels()[-1], visible=False)
		ax[xx][yy].set_xlabel('DSI', fontdict = {'fontsize':18})
		ax[xx][yy].set_title(title,pad = 18, fontdict = {'fontsize':20})
		fig.set_figwidth(36)
		fig.set_figheight(36)
		fig.set_facecolor('floralwhite')
		ax[xx][yy].set_facecolor('seashell')
		ax[xx][yy].invert_yaxis()
		ax[xx][yy].tick_params(axis="y", labelsize=16)
		ax[xx][yy].tick_params(axis="x", labelsize=13)
		if k == 0:
			ax[xx][yy].set_yticks([])
			ax[xx][yy].set_yticks([], minor=True)
	fig.delaxes(ax[-2][-1])

	plt.savefig(algo_name+"/"+"/genes plots/indices/"+algo_name+"_"+"distribution_events_detailed_DSI_females"+".pdf",dpi = 600)
			
#NDSI
	fig, ax =  plt.subplots(4, 3, squeeze=False)
	data = genders_plot_NDSI
	get_top(data,algo_name+'/data_genes/'+algo_name+"_distribution_events_detailed_females_genes_NDSI_top.tsv")
	for j in range(11):#тип мутации
		k = 0
		title = rows_genes[j]
		#fig, ax = plt.subplots()
		xx,yy = j//3,j%3
		if j==8:
			xx,yy = j//3+1,0
		elif j>8:
			xx,yy = j//3,j%3+1
		for driver in data[j]:#top 10 genes
			if k <10 and driver[0] not in ['','-']:
				
				ax[xx][yy].barh(driver[0], float(driver[1]),color = colors_genes[j],linewidth = 0.1)
				k+=1
		if k!=0:
			for ik in range(k,9):
				ax[xx][yy].barh(str(ik+100), 0,color = colors_genes[j],linewidth = 0.1)
				plt.setp(ax[xx][yy].get_yticklabels()[-1], visible=False)
		ax[xx][yy].set_xlabel('NDSI', fontdict = {'fontsize':18})
		ax[xx][yy].set_title(title,pad = 18, fontdict = {'fontsize':20})
		fig.set_figwidth(36)
		fig.set_figheight(36)
		fig.set_facecolor('floralwhite')
		ax[xx][yy].set_facecolor('seashell')
		ax[xx][yy].invert_yaxis()
		ax[xx][yy].tick_params(axis="y", labelsize=16)
		ax[xx][yy].tick_params(axis="x", labelsize=13)
		if k == 0:
			ax[xx][yy].set_yticks([])
			ax[xx][yy].set_yticks([], minor=True)
	fig.delaxes(ax[-2][-1])

	plt.savefig(algo_name+"/"+"/genes plots/indices/"+algo_name+"_"+"distribution_events_detailed_NDSI_females"+".pdf",dpi = 600)
	
#######DSI

	x = genders_plot_genes.keys()
	data = genders_plot_genes

	for i in x:#gender/stage...
		fig, ax =  plt.subplots(4, 3, squeeze=False)
		
		for j in range(11):#тип мутации
			k = 0
			title = rows_genes[j]
			#fig, ax = plt.subplots()
			xx,yy = j//3,j%3
			if j==8:
				xx,yy = j//3+1,0
			elif j>8:
				xx,yy = j//3,j%3+1
			for driver in data[i][j]:#top 10 genes
				if k <10 and driver[0] not in ['','-']:
					ax[xx][yy].barh(driver[0], int(driver[1]),color = colors_genes[j],linewidth = 0.1)
					k+=1
			if k!=0:
				for ik in range(k,9):
					ax[xx][yy].barh(str(ik+100), 0,color = colors_genes[j],linewidth = 0.1)
					plt.setp(ax[xx][yy].get_yticklabels()[-1], visible=False)
			ax[xx][yy].set_xlabel('Number of events', fontdict = {'fontsize':18})
			ax[xx][yy].set_title(title,pad = 18, fontdict = {'fontsize':20})
			fig.set_figwidth(36)
			fig.set_figheight(36)
			fig.set_facecolor('floralwhite')
			ax[xx][yy].set_facecolor('seashell')
			ax[xx][yy].invert_yaxis()
			ax[xx][yy].tick_params(axis="y", labelsize=16)
			ax[xx][yy].tick_params(axis="x", labelsize=13)
			if k == 0:
				ax[xx][yy].set_yticks([])
				ax[xx][yy].set_yticks([], minor=True)
		fig.delaxes(ax[-2][-1])
		plt.savefig(algo_name+"/"+"/genes plots/detailed/"+algo_name+"_"+"distribution_events_detailed_females_"+i+".pdf",dpi = 600)
#################write
	with open(algo_name+'/data/'+algo_name+"_distribution_events_detailed.tsv",'w') as out:
		out.write(header)
		types_plot = {}
		for c in sort(total_events.keys()):
			out.write(c+"\t")
			num = len(total_events[c][-1])
			a = total_events[c][:-1]
			a = list(map(lambda x:x/num,a))
			types_plot[c] = a
			line = "\t".join(list(map(lambda x:str(x),a)))+"\n"
			out.write(line)

	for i in range (1,101):
		if str(i) not in types_plot.keys():
			types_plot[str(i)] = [0.0 for  i in range(11)]
	data = types_plot
	x = [str(i) for i in range (1,101)]
	fig, ax = plt.subplots()
	title = 'Driver event distribution by total number of driver events per patient'
	for i in x:
		ax.bar(i, data[i][0],color = 'lightcoral')
		bott = data[i][0]
		ax.bar(i, data[i][1], bottom = bott,color = 'red')
		bott+= data[i][1]
		ax.bar(i, data[i][2], bottom = bott,color = 'maroon')
		bott+= data[i][2]
		ax.bar(i, data[i][3], bottom = bott,color = 'cyan')
		bott+= data[i][3]
		ax.bar(i, data[i][4], bottom = bott,color = 'c')
		bott+= data[i][4]
		ax.bar(i, data[i][5], bottom = bott,color = 'teal')
		bott+= data[i][5]
		ax.bar(i, data[i][6], bottom = bott,color = 'orange')
		bott+= data[i][6]
		ax.bar(i, data[i][7], bottom = bott,color = 'darkorange')
		bott+= data[i][7]
		ax.bar(i, data[i][8], bottom = bott,color = 'violet')
		bott+= data[i][8]
		ax.bar(i, data[i][9], bottom = bott,color = 'purple')
	ax.set_ylabel('Average number of driver events', fontdict = {'fontsize':30})
	ax.set_xlabel('Total number of driver events per patient', fontdict = {'fontsize':30})
	ax.set_title(title, pad = 18,fontdict = {'fontsize':30})
	plt.xticks(rotation = 90,fontsize = 14)
	plt.yticks(fontsize = 19)
	fig.set_figwidth(20)
	fig.set_figheight(16)
	fig.set_facecolor('floralwhite')
	ax.set_facecolor('seashell')
	plt.savefig(algo_name+"/"+"/cumulative histograms/"+algo_name+"_"+"distribution_events_detailed"+".pdf",dpi = 600)

	with open(algo_name+'/data/'+algo_name+"_distribution_events_detailed_males.tsv",'w') as out:
		out.write(header)
		males_plot= {}
		for c in sort(male_events.keys()):
			out.write(c+"\t")
			num = len(male_events[c][-1])
			a = male_events[c][:-1]
			a = list(map(lambda x:x/num,a))
			males_plot[c] = a
			line = "\t".join(list(map(lambda x:str(x),a)))+"\n"
			out.write(line)
	for i in range (1,101):
		if str(i) not in males_plot.keys():
			males_plot[str(i)] = [0.0 for  i in range(11)]		
	data = males_plot
	x = [str(i) for i in range (1,101)]
	fig, ax = plt.subplots()
	title = 'Driver event distribution by total number of driver events per patient in males'
	for i in x:
		ax.bar(i, data[i][0],color = 'lightcoral')
		bott = data[i][0]
		ax.bar(i, data[i][1], bottom = bott,color = 'red')
		bott+= data[i][1]
		ax.bar(i, data[i][2], bottom = bott,color = 'maroon')
		bott+= data[i][2]
		ax.bar(i, data[i][3], bottom = bott,color = 'cyan')
		bott+= data[i][3]
		ax.bar(i, data[i][4], bottom = bott,color = 'c')
		bott+= data[i][4]
		ax.bar(i, data[i][5], bottom = bott,color = 'teal')
		bott+= data[i][5]
		ax.bar(i, data[i][6], bottom = bott,color = 'orange')
		bott+= data[i][6]
		ax.bar(i, data[i][7], bottom = bott,color = 'darkorange')
		bott+= data[i][7]
		ax.bar(i, data[i][8], bottom = bott,color = 'violet')
		bott+= data[i][8]
		ax.bar(i, data[i][9], bottom = bott,color = 'purple')
	ax.set_ylabel('Average number of driver events', fontdict = {'fontsize':30})
	ax.set_xlabel('Total number of driver events per patient', fontdict = {'fontsize':30})
	ax.set_title(title, pad = 18,fontdict = {'fontsize':30})
	plt.xticks(rotation = 90,fontsize = 14)
	plt.yticks(fontsize = 19)
	fig.set_figwidth(20)
	fig.set_figheight(16)
	fig.set_facecolor('floralwhite')
	ax.set_facecolor('seashell')
	plt.savefig(algo_name+"/"+"/cumulative histograms/"+algo_name+"_"+"distribution_events_detailed_males"+".pdf",dpi = 600)

	with open(algo_name+'/data/'+algo_name+"_distribution_events_detailed_females.tsv",'w') as out:
		out.write(header)
		females_plot = {}
		for c in sort(female_events.keys()):
			out.write(c+"\t")
			num = len(female_events[c][-1])
			a = female_events[c][:-1]
			a = list(map(lambda x:x/num,a))
			females_plot[c] = a
			line = "\t".join(list(map(lambda x:str(x),a)))+"\n"
			out.write(line)
	for i in range (1,101):
		if str(i) not in females_plot.keys():
			females_plot[str(i)] = [0.0 for  i in range(11)]
	data = females_plot
	x = [str(i) for i in range (1,101)]
	fig, ax = plt.subplots()
	title = 'Driver event distribution by total number of driver events per patient in females'
	for i in x:
		ax.bar(i, data[i][0],color = 'lightcoral')
		bott = data[i][0]
		ax.bar(i, data[i][1], bottom = bott,color = 'red')
		bott+= data[i][1]
		ax.bar(i, data[i][2], bottom = bott,color = 'maroon')
		bott+= data[i][2]
		ax.bar(i, data[i][3], bottom = bott,color = 'cyan')
		bott+= data[i][3]
		ax.bar(i, data[i][4], bottom = bott,color = 'c')
		bott+= data[i][4]
		ax.bar(i, data[i][5], bottom = bott,color = 'teal')
		bott+= data[i][5]
		ax.bar(i, data[i][6], bottom = bott,color = 'orange')
		bott+= data[i][6]
		ax.bar(i, data[i][7], bottom = bott,color = 'darkorange')
		bott+= data[i][7]
		ax.bar(i, data[i][8], bottom = bott,color = 'violet')
		bott+= data[i][8]
		ax.bar(i, data[i][9], bottom = bott,color = 'purple')
	ax.set_ylabel('Average number of driver events', fontdict = {'fontsize':30})
	ax.set_xlabel('Total number of driver events per patient', fontdict = {'fontsize':30})
	ax.set_title(title,pad = 18, fontdict = {'fontsize':30})
	plt.xticks(rotation = 90,fontsize = 14)
	plt.yticks(fontsize = 19)
	fig.set_figwidth(20)
	fig.set_figheight(16)
	fig.set_facecolor('floralwhite')
	ax.set_facecolor('seashell')
	plt.savefig(algo_name+"/"+"/cumulative histograms/"+algo_name+"_"+"distribution_events_detailed_females"+".pdf",dpi = 600)
	print ("step 15 end")
```
